## Supplementary Figure 3 for "Fine-mapping a genome-wide meta-analysis of 98,374 migraine cases identifies 181 sets of candidate causal variants"

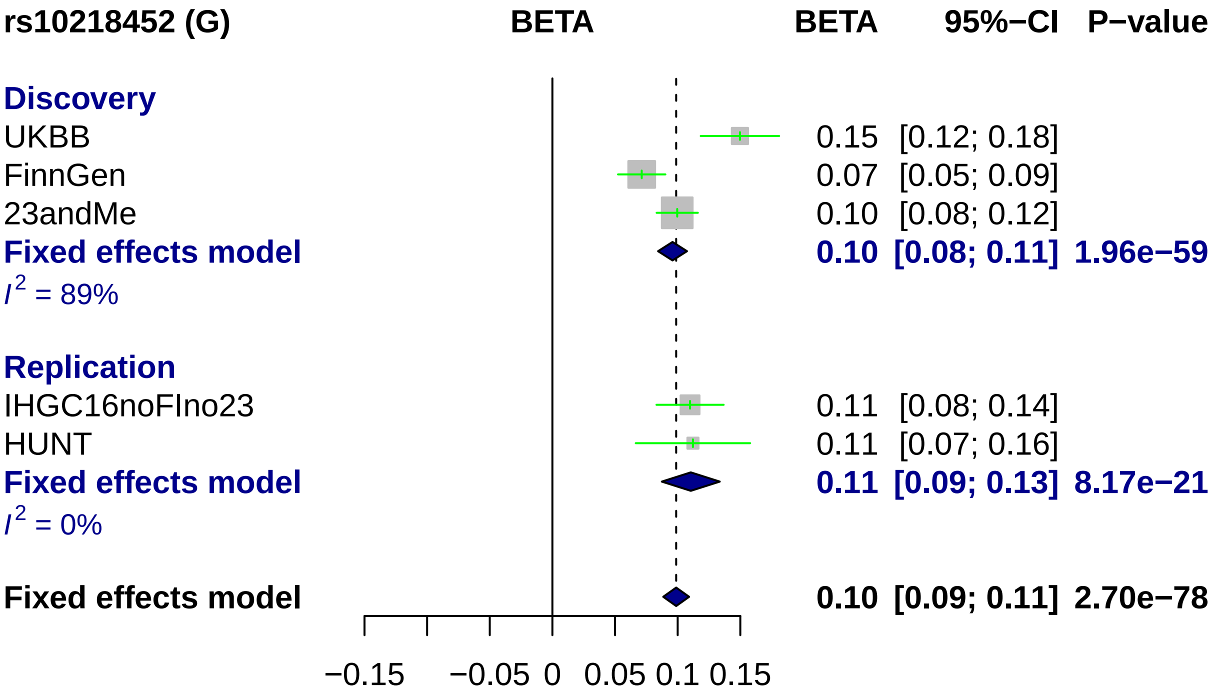
rs10218452 (G)
BETA
BETA
95%−CI
P−value
Discovery
UKBB
0.15
[0.12; 0.18]
FinnGen
0.07
[0.05; 0.09]
23andMe
0.10
[0.08; 0.12]
Fixed effects model
0.10
[0.08; 0.11]
1.96e−59
I2 = 89%
Replication
IHGC16noFIno23
0.11
[0.08; 0.14]
HUNT
0.11
[0.07; 0.16]
Fixed effects model
0.11
[0.09; 0.13]
8.17e−21
I2 = 0%
Fixed effects model
0.10
[0.09; 0.11]
2.70e−78
−0.15
−0.05
0
0.05
0.1
0.15

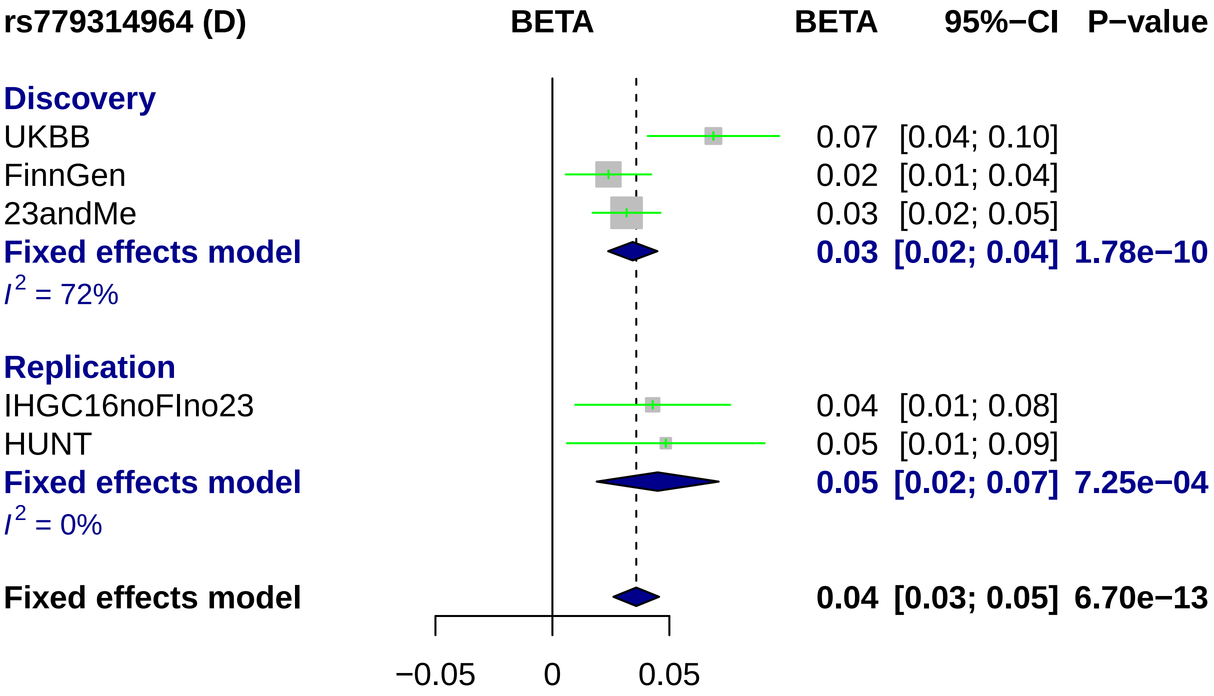
rs779314964 (D)
BETA
BETA
95%−CI
P−value
Discovery
UKBB
0.07 [0.04; 0.10]
FinnGen
0.02 [0.01; 0.04]
23andMe
0.03 [0.02; 0.05]
Fixed effects model
0.03 [0.02; 0.04] 1.78e−10
I2 = 72%
Replication
IHGC16noFIno23
0.04 [0.01; 0.08]
HUNT
0.05 [0.01; 0.09]
Fixed effects model
0.05 [0.02; 0.07] 7.25e−04
I2 = 0%
Fixed effects model
0.04 [0.03; 0.05] 6.70e−13
−0.05
0
0.05

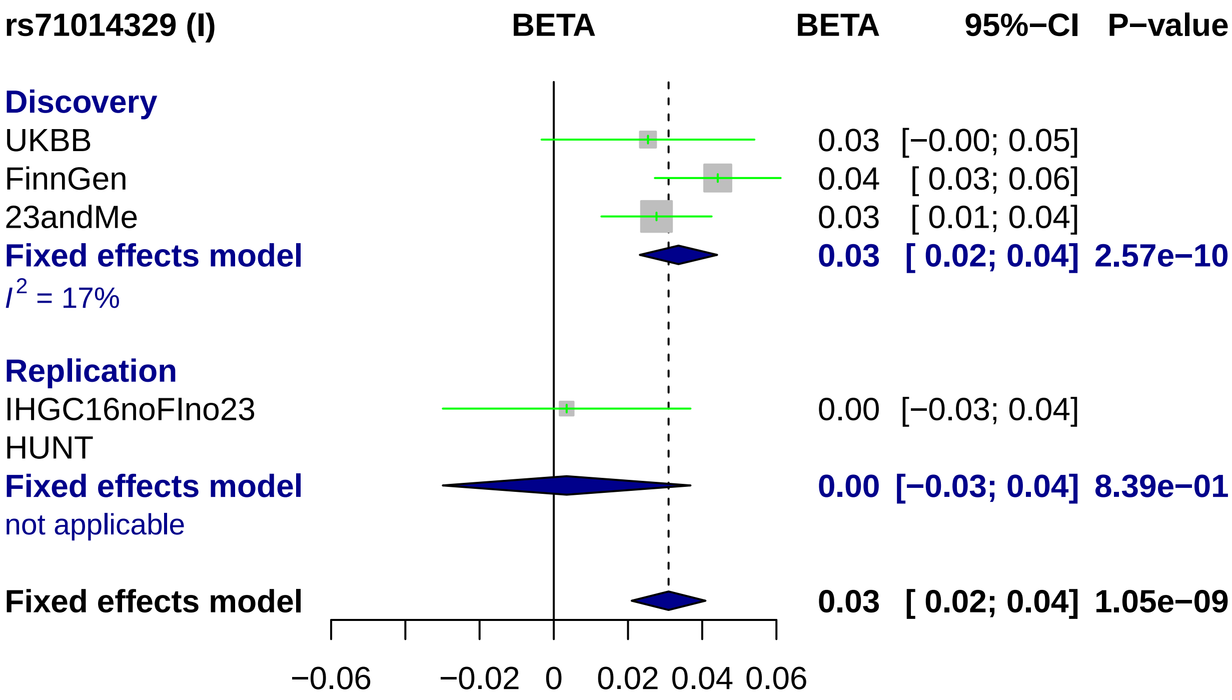
rs71014329 (I)
BETA
BETA
95%−CI
P−value
Discovery
UKBB
0.03
[−0.00; 0.05]
FinnGen
0.04
[ 0.03; 0.06]
23andMe
0.03
[ 0.01; 0.04]
Fixed effects model
0.03
[ 0.02; 0.04]
2.57e−10
I2 = 17%
Replication
IHGC16noFIno23
0.00
[−0.03; 0.04]
HUNT
Fixed effects model
0.00
[−0.03; 0.04]
8.39e−01
not applicable
Fixed effects model
0.03
[ 0.02; 0.04]
1.05e−09
−0.06
−0.02
0
0.02
0.04
0.06

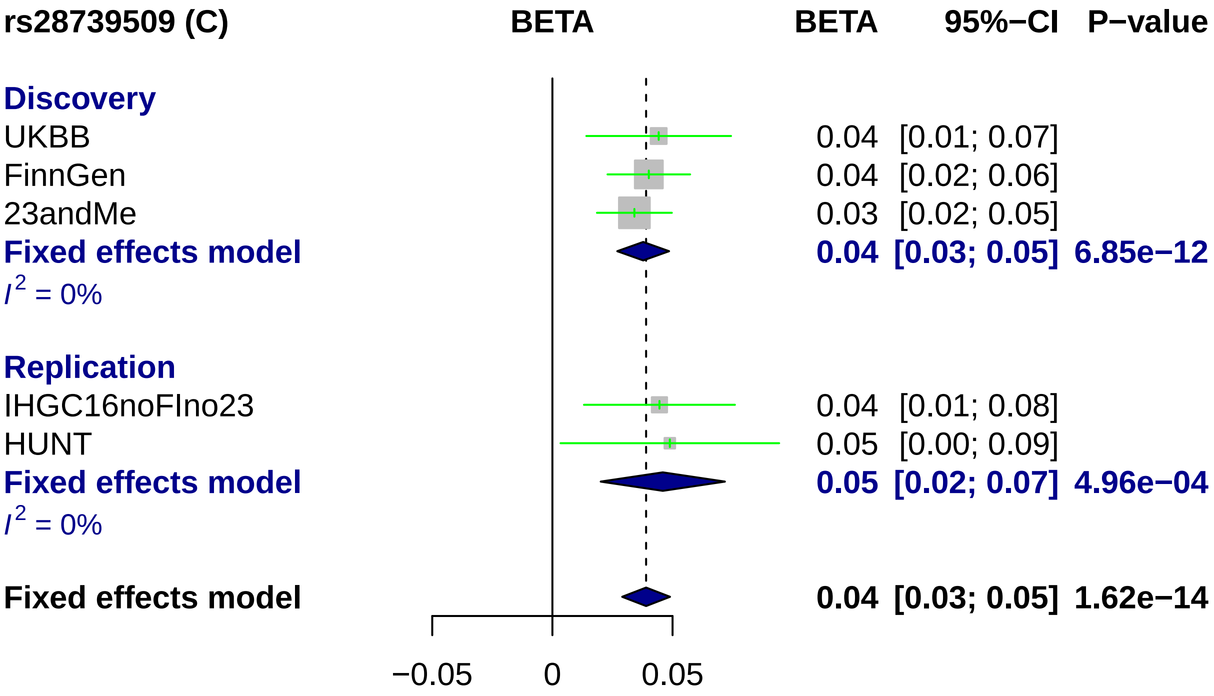
rs28739509 (C)
BETA
BETA
95%−CI
P−value
Discovery
UKBB
0.04
[0.01; 0.07]
FinnGen
0.04
[0.02; 0.06]
23andMe
0.03
[0.02; 0.05]
Fixed effects model
0.04
[0.03; 0.05]
6.85e−12
I2 = 0%
Replication
IHGC16noFIno23
0.04
[0.01; 0.08]
HUNT
0.05
[0.00; 0.09]
Fixed effects model
0.05
[0.02; 0.07]
4.96e−04
I2 = 0%
Fixed effects model
0.04
[0.03; 0.05]
1.62e−14
−0.05
0
0.05

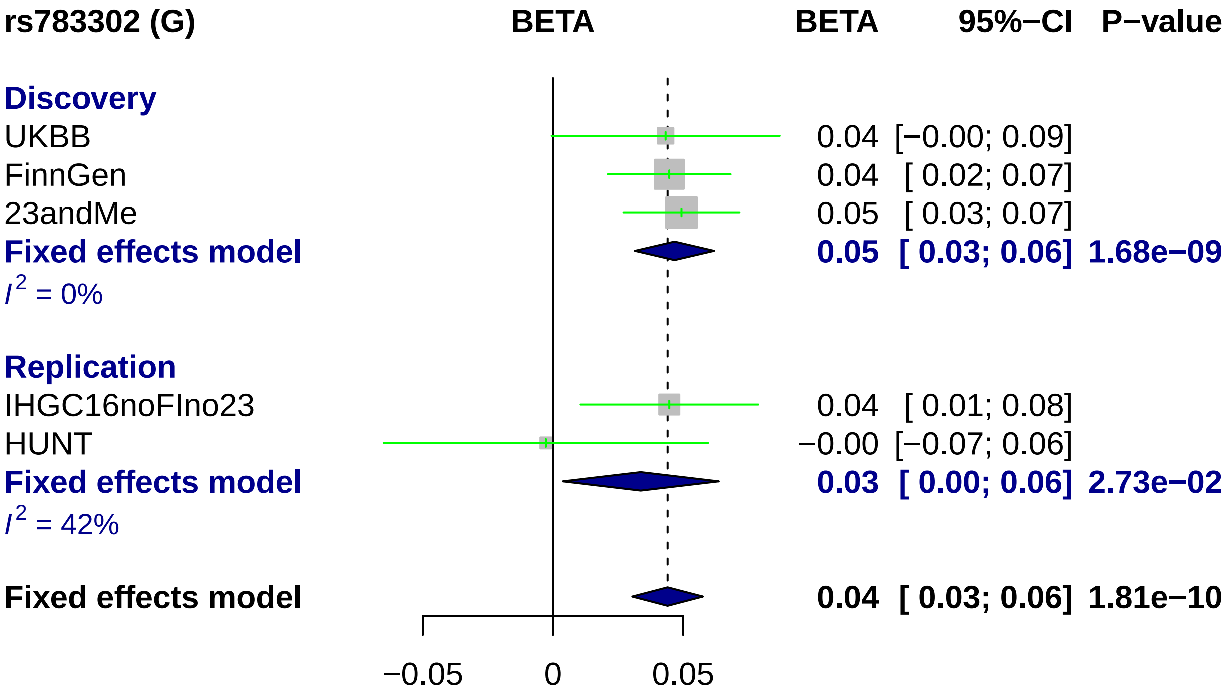
rs783302 (G)
BETA
BETA
95%−CI
P−value
Discovery
UKBB
0.04 [−0.00; 0.09]
FinnGen
0.04 [ 0.02; 0.07]
23andMe
0.05 [ 0.03; 0.07]
Fixed effects model
0.05 [ 0.03; 0.06] 1.68e−09
I2 = 0%
Replication
IHGC16noFIno23
0.04 [ 0.01; 0.08]
HUNT
−0.00 [−0.07; 0.06]
Fixed effects model
0.03 [ 0.00; 0.06] 2.73e−02
I2 = 42%
Fixed effects model
0.04 [ 0.03; 0.06] 1.81e−10
−0.05
0
0.05

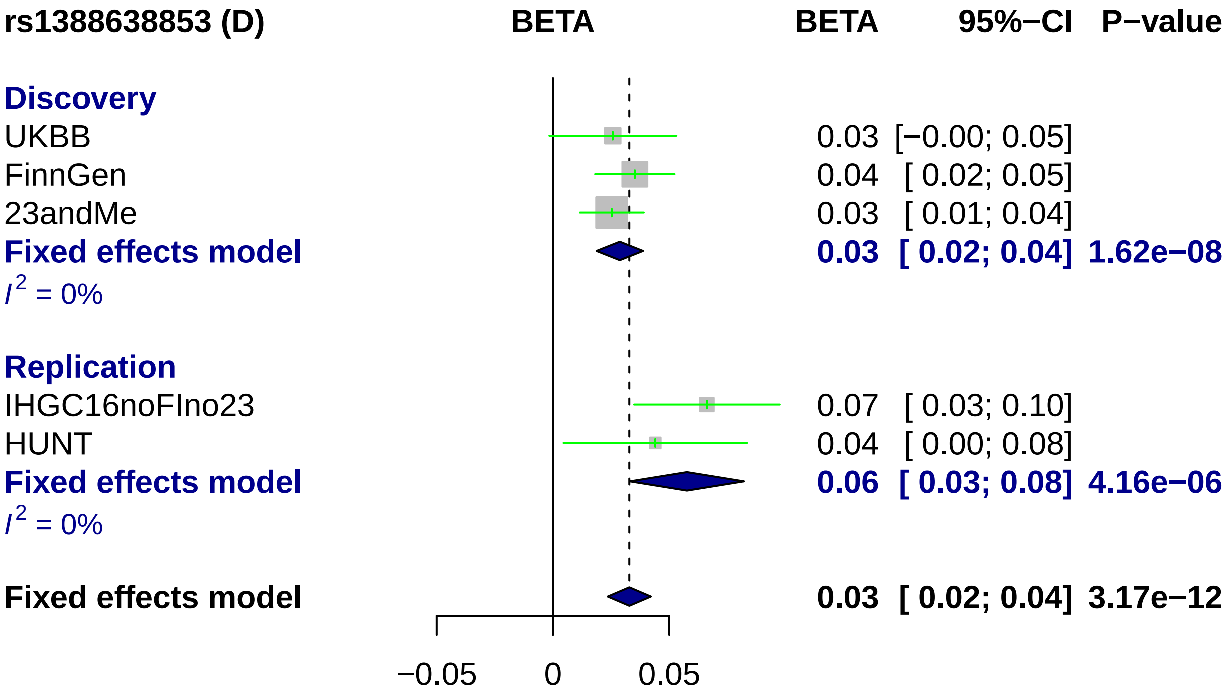
rs1388638853 (D)
BETA
BETA
95%−CI
P−value
Discovery
UKBB
0.03 [−0.00; 0.05]
FinnGen
0.04 [ 0.02; 0.05]
23andMe
0.03 [ 0.01; 0.04]
Fixed effects model
0.03 [ 0.02; 0.04] 1.62e−08
I2 = 0%
Replication
IHGC16noFIno23
0.07 [ 0.03; 0.10]
HUNT
0.04 [ 0.00; 0.08]
Fixed effects model
0.06 [ 0.03; 0.08] 4.16e−06
I2 = 0%
Fixed effects model
0.03 [ 0.02; 0.04] 3.17e−12
−0.05
0
0.05

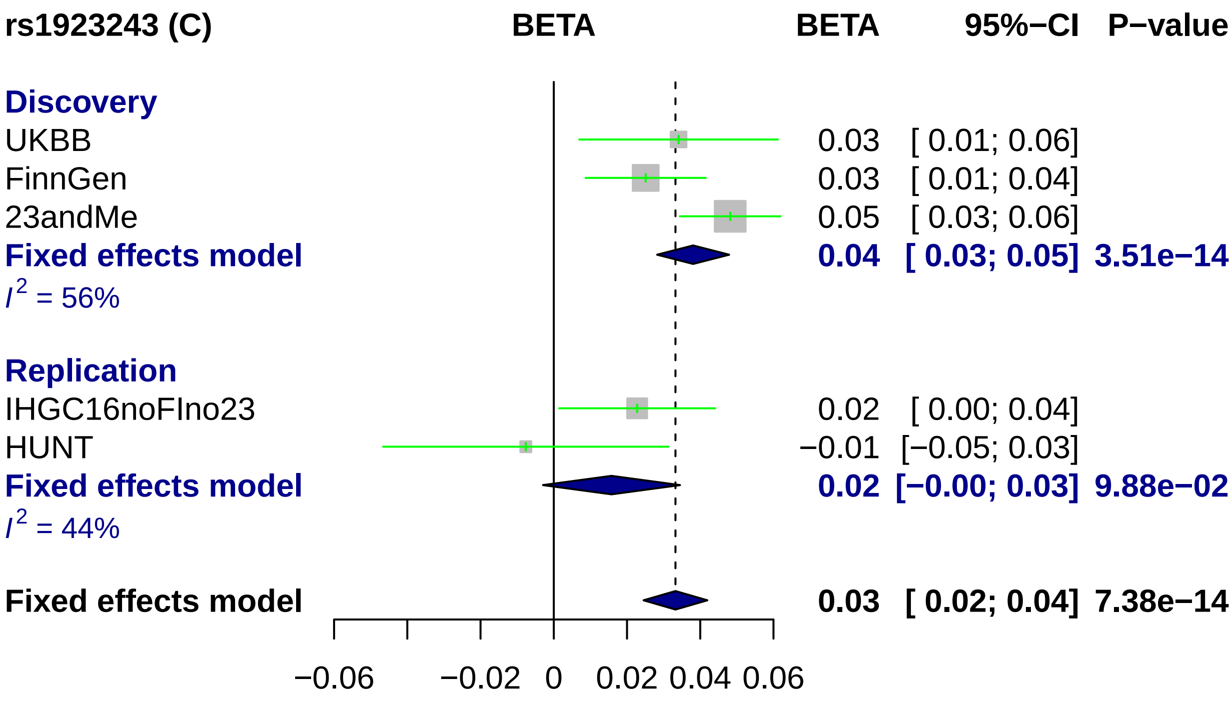

rs1923243 (C)
BETA
BETA
95%−CI
P−value
Discovery
UKBB
0.03 [ 0.01; 0.06]
FinnGen
0.03 [ 0.01; 0.04]
23andMe
0.05 [ 0.03; 0.06]
Fixed effects model
0.04 [ 0.03; 0.05] 3.51e−14
I2 = 56%
Replication
IHGC16noFIno23
0.02 [ 0.00; 0.04]
HUNT
−0.01 [−0.05; 0.03]
Fixed effects model
0.02 [−0.00; 0.03] 9.88e−02
I2 = 44%
Fixed effects model
0.03 [ 0.02; 0.04] 7.38e−14
−0.06
−0.02
0
0.02
0.04
0.06

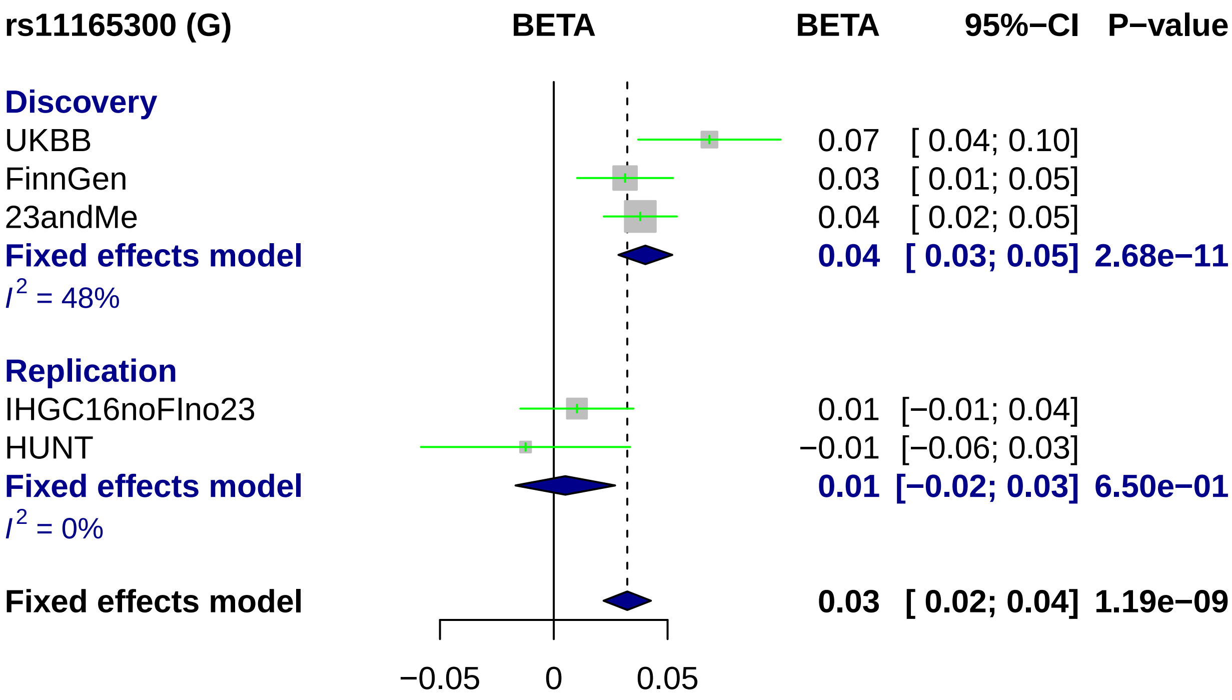
rs11165300 (G)
BETA
BETA
95%−CI
P−value
Discovery
UKBB 0.07 [ 0.04; 0.10]
FinnGen 0.03 [ 0.01; 0.05]
23andMe 0.04 [ 0.02; 0.05]
Fixed effects model 0.04 [ 0.03; 0.05] 2.68e−11
I2 = 48%
Replication
IHGC16noFIno23 0.01 [−0.01; 0.04]
HUNT −0.01 [−0.06; 0.03]
Fixed effects model 0.01 [−0.02; 0.03] 6.50e−01
I2 = 0%
Fixed effects model 0.03 [ 0.02; 0.04] 1.19e−09
−0.05
0
0.05

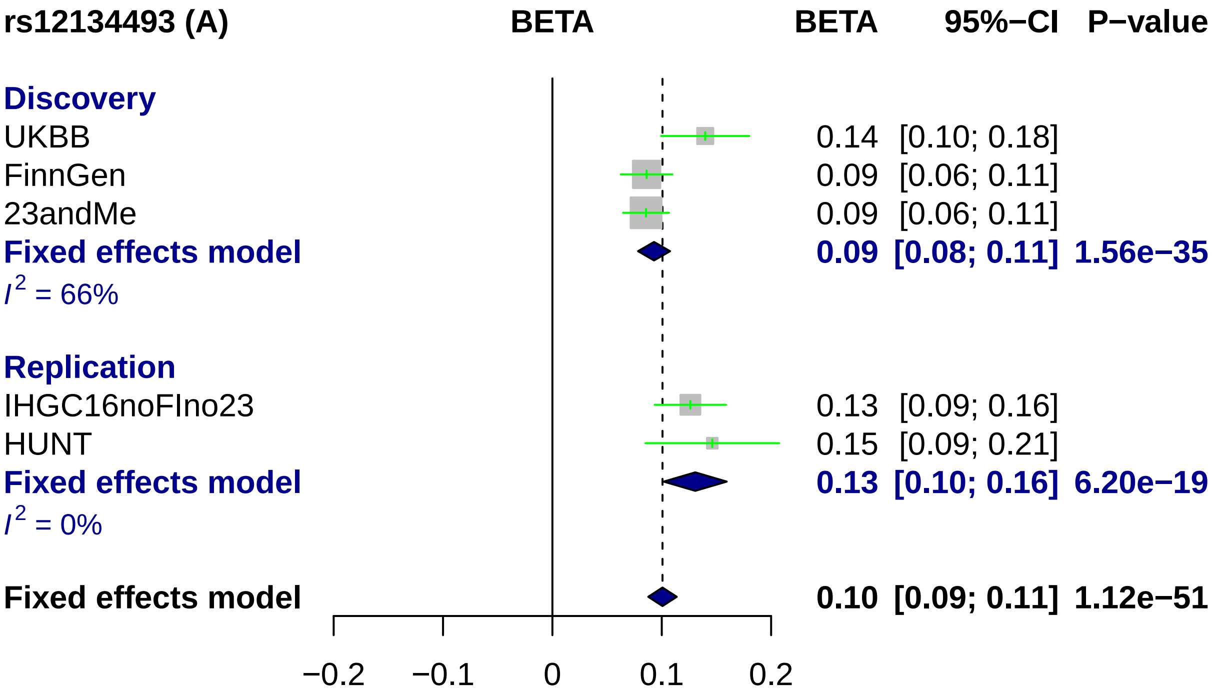

rs12134493 (A)
BETA
BETA
95%−CI
P−value
Discovery
UKBB
0.14 [0.10; 0.18]
FinnGen
0.09 [0.06; 0.11]
23andMe
0.09 [0.06; 0.11]
Fixed effects model
0.09 [0.08; 0.11] 1.56e−35
I2 = 66%
Replication
IHGC16noFIno23
0.13 [0.09; 0.16]
HUNT
0.15 [0.09; 0.21]
Fixed effects model
0.13 [0.10; 0.16] 6.20e−19
I2 = 0%
Fixed effects model
0.10 [0.09; 0.11] 1.12e−51
−0.2
−0.1
0
0.1
0.2

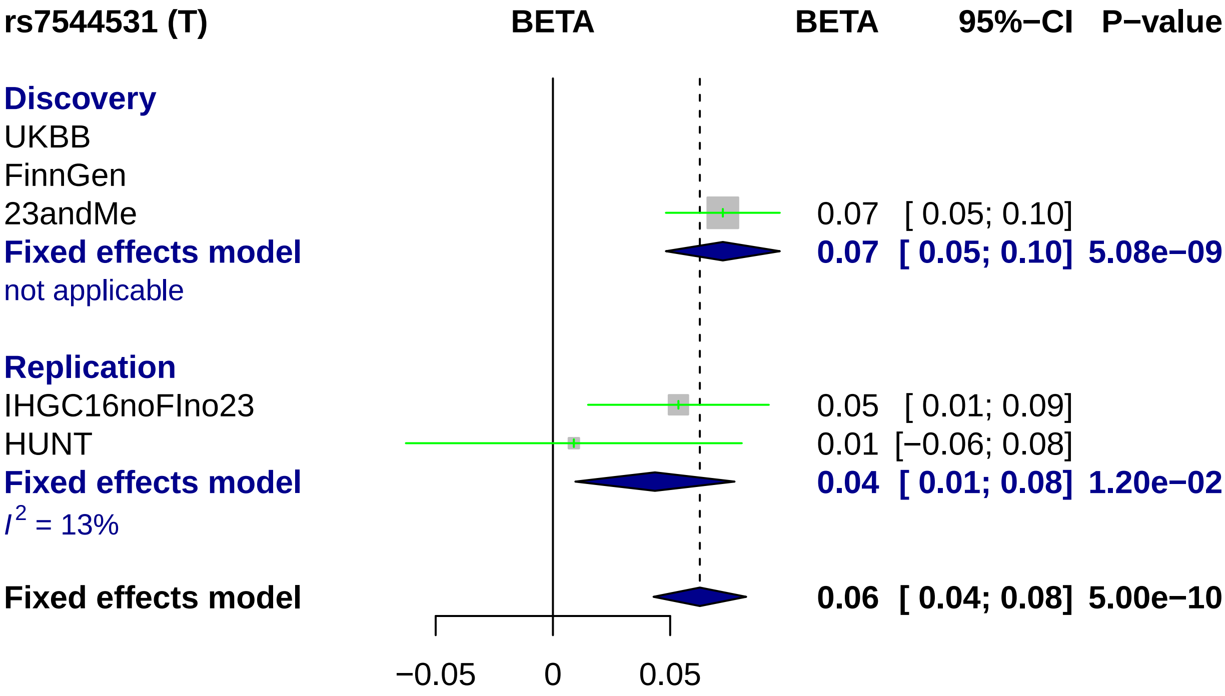

rs7544531 (T)
BETA
BETA
95%−CI
P−value
Discovery
UKBB
FinnGen
23andMe
0.07 [ 0.05; 0.10]
Fixed effects model
0.07 [ 0.05; 0.10] 5.08e−09
not applicable
Replication
IHGC16noFIno23
0.05 [ 0.01; 0.09]
HUNT
0.01 [−0.06; 0.08]
Fixed effects model
0.04 [ 0.01; 0.08] 1.20e−02
I2 = 13%
Fixed effects model
0.06 [ 0.04; 0.08] 5.00e−10
−0.05
0
0.05

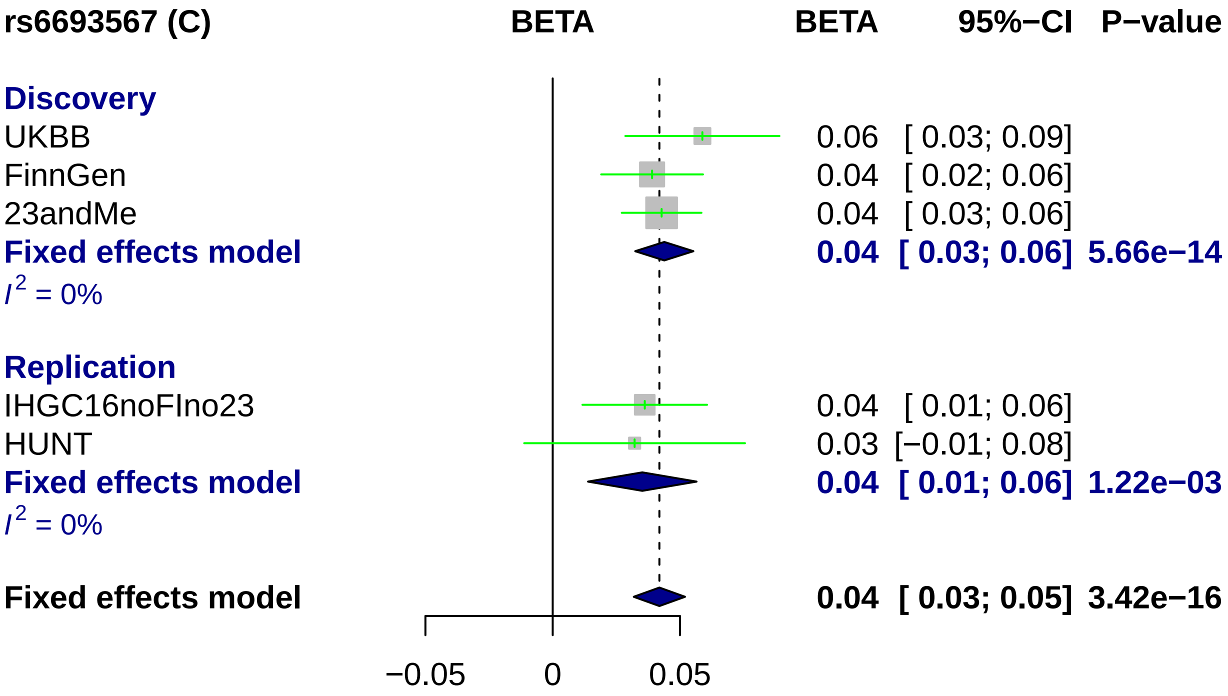

rs6693567 (C)
BETA
BETA
95%−CI
P−value
Discovery
UKBB
0.06 [ 0.03; 0.09]
FinnGen
0.04 [ 0.02; 0.06]
23andMe
0.04 [ 0.03; 0.06]
Fixed effects model
0.04 [ 0.03; 0.06] 5.66e−14
I2 = 0%
Replication
IHGC16noFIno23
0.04 [ 0.01; 0.06]
HUNT
0.03 [−0.01; 0.08]
Fixed effects model
0.04 [ 0.01; 0.06] 1.22e−03
I2 = 0%
Fixed effects model
0.04 [ 0.03; 0.05] 3.42e−16
−0.05
0
0.05

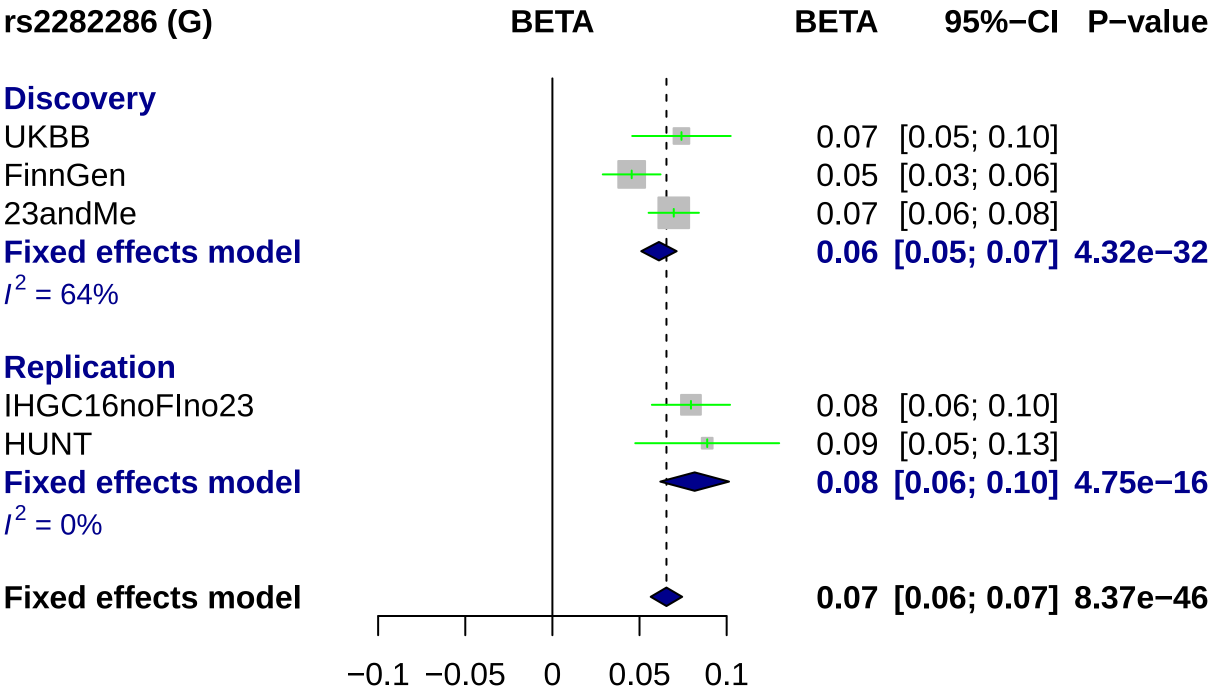
rs2282286 (G)
BETA
BETA
95%−CI
P−value
Discovery
UKBB
0.07 [0.05; 0.10]
FinnGen
0.05 [0.03; 0.06]
23andMe
0.07 [0.06; 0.08]
Fixed effects model
0.06 [0.05; 0.07] 4.32e−32
I2 = 64%
Replication
IHGC16noFIno23
0.08 [0.06; 0.10]
HUNT
0.09 [0.05; 0.13]
Fixed effects model
0.08 [0.06; 0.10] 4.75e−16
I2 = 0%
Fixed effects model
0.07 [0.06; 0.07] 8.37e−46
−0.1
−0.05
0
0.05
0.1

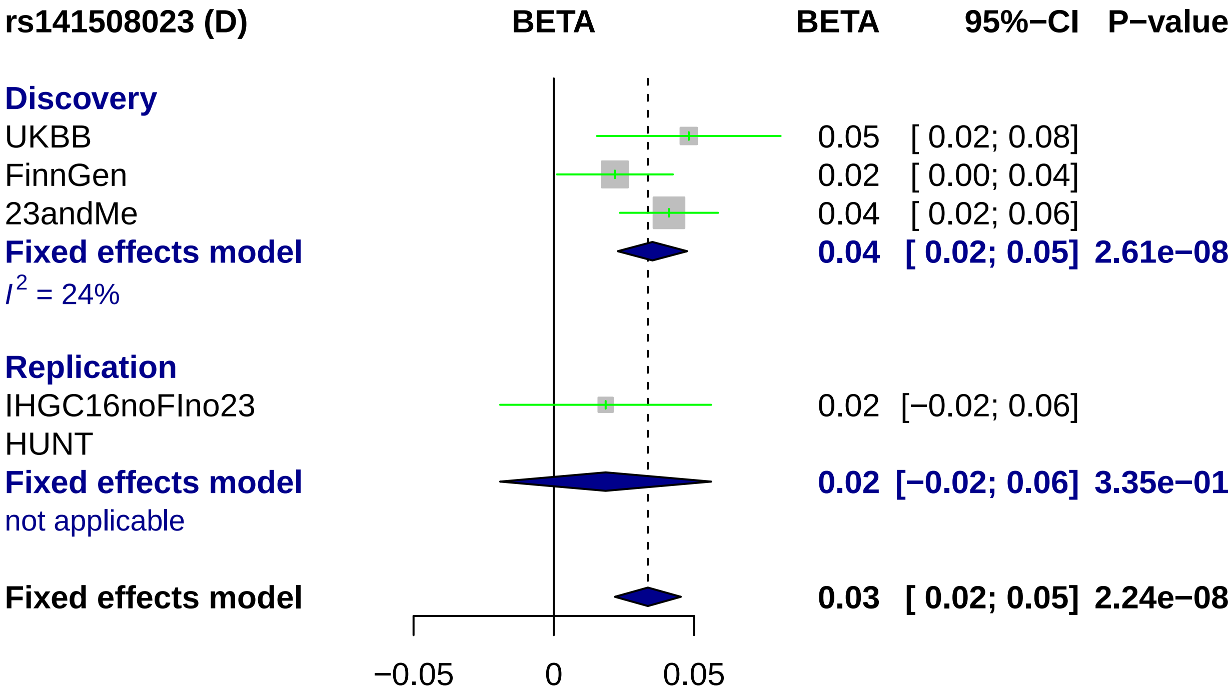
rs141508023 (D)
BETA
BETA
95%−CI
P−value
Discovery
UKBB
0.05 [ 0.02; 0.08]
FinnGen
0.02 [ 0.00; 0.04]
23andMe
0.04 [ 0.02; 0.06]
Fixed effects model
0.04 [ 0.02; 0.05] 2.61e−08
I^2 = 24%
Replication
IHGC16noFIno23
0.02 [−0.02; 0.06]
HUNT
Fixed effects model
0.02 [−0.02; 0.06] 3.35e−01
not applicable
Fixed effects model
0.03 [ 0.02; 0.05] 2.24e−08
−0.05
0
0.05

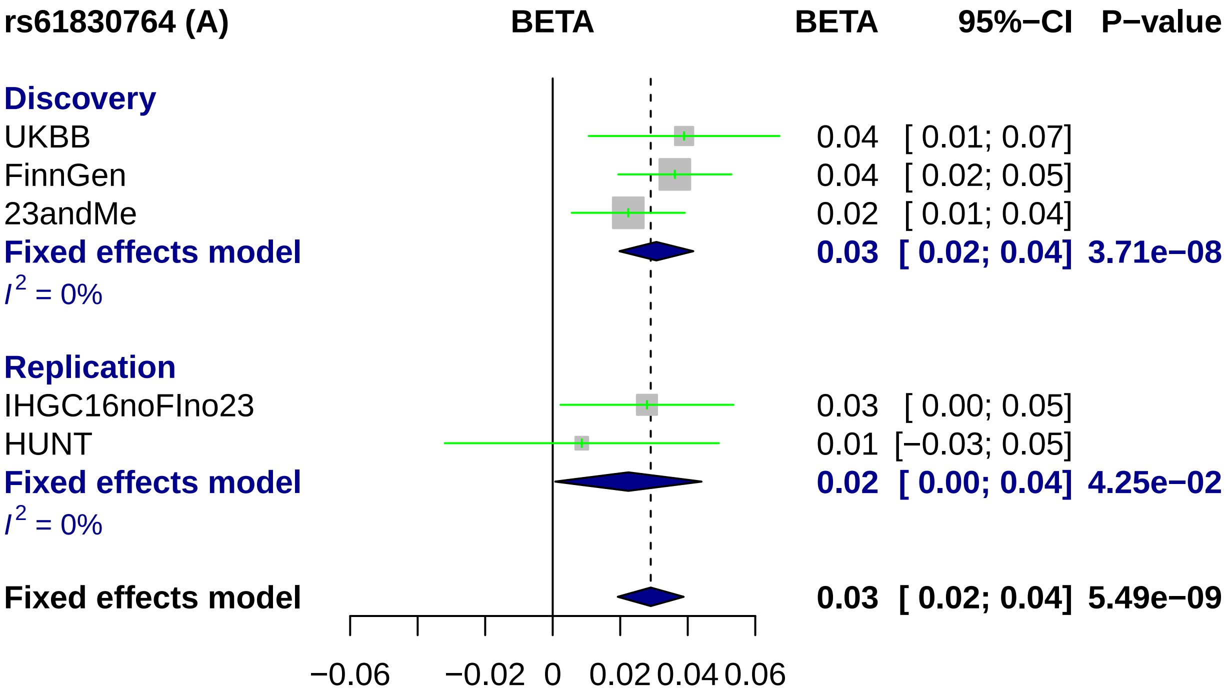

rs61830764 (A)
BETA
BETA
95%−CI
P−value
Discovery
UKBB
0.04 [ 0.01; 0.07]
FinnGen
0.04 [ 0.02; 0.05]
23andMe
0.02 [ 0.01; 0.04]
Fixed effects model
0.03 [ 0.02; 0.04] 3.71e−08
I2 = 0%
Replication
IHGC16noFIno23
0.03 [ 0.00; 0.05]
HUNT
0.01 [−0.03; 0.05]
Fixed effects model
0.02 [ 0.00; 0.04] 4.25e−02
I2 = 0%
Fixed effects model
0.03 [ 0.02; 0.04] 5.49e−09
−0.06
−0.02
0
0.02
0.04
0.06

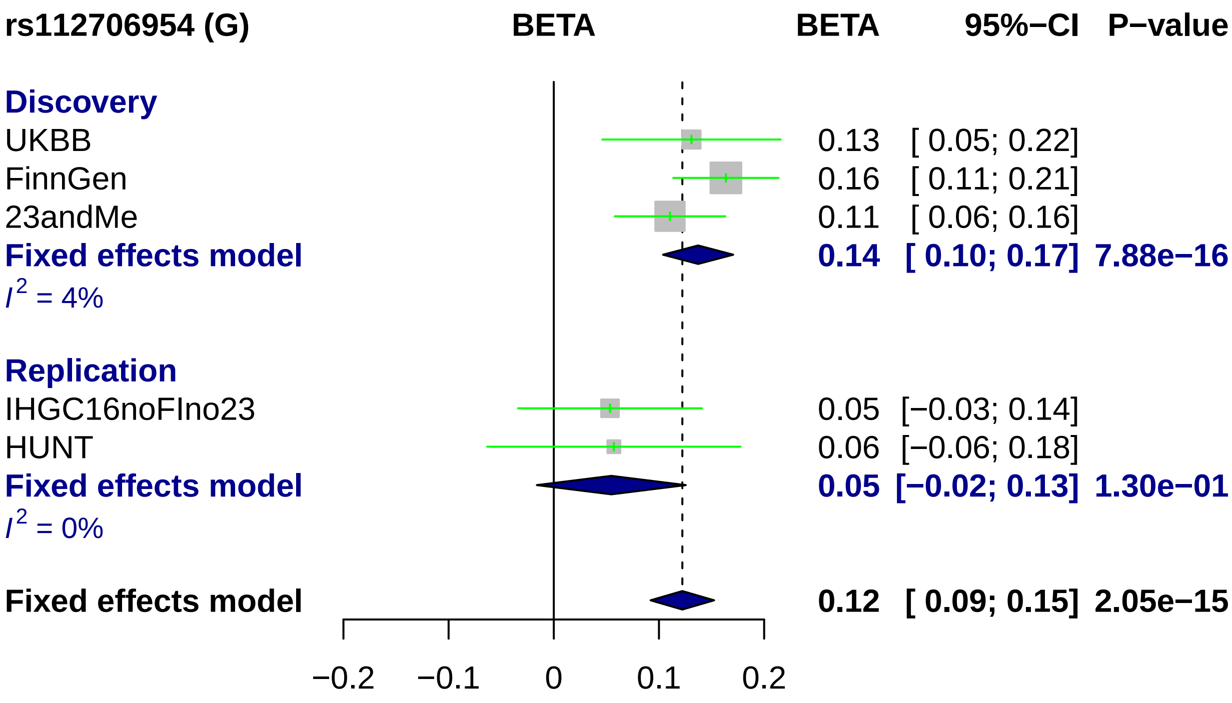

rs112706954 (G)
BETA
BETA
95%−CI
P−value
Discovery
UKBB
0.13
[ 0.05; 0.22]
FinnGen
0.16
[ 0.11; 0.21]
23andMe
0.11
[ 0.06; 0.16]
Fixed effects model
0.14
[ 0.10; 0.17]
7.88e−16
I2 = 4%
Replication
IHGC16noFIno23
0.05
[−0.03; 0.14]
HUNT
0.06
[−0.06; 0.18]
Fixed effects model
0.05
[−0.02; 0.13]
1.30e−01
I2 = 0%
Fixed effects model
0.12
[ 0.09; 0.15]
2.05e−15
−0.2
−0.1
0
0.1
0.2

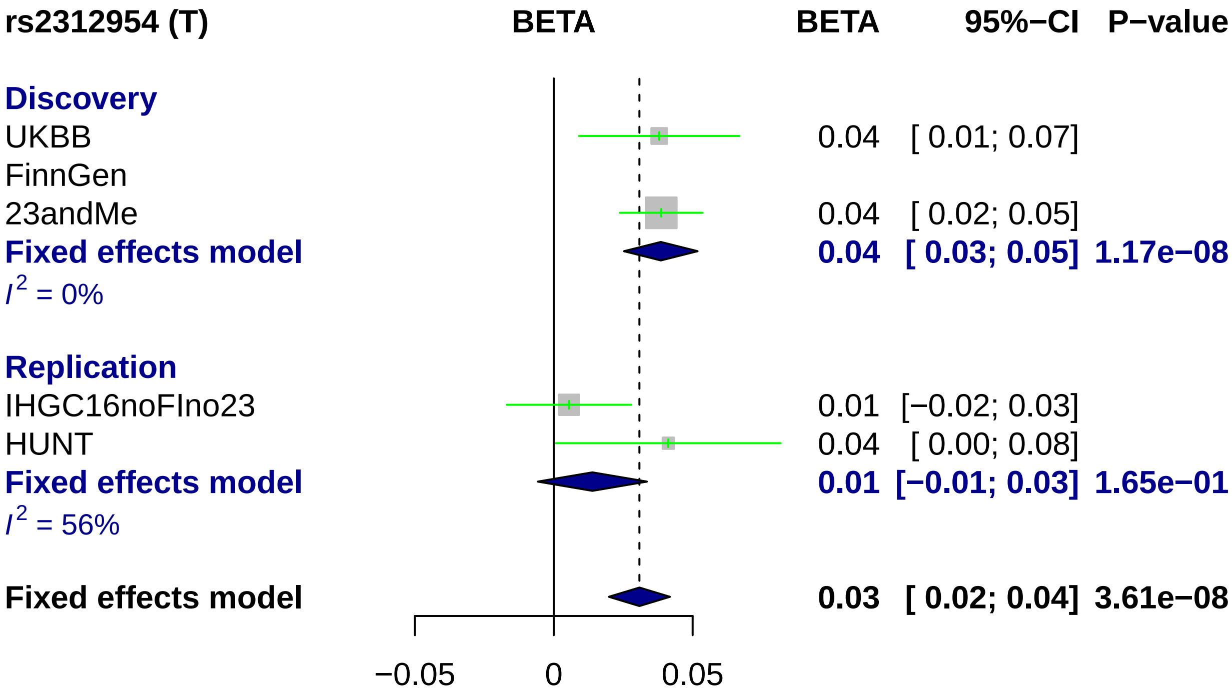
rs2312954 (T)
BETA
BETA
95%−CI
P−value
Discovery
UKBB
0.04
[ 0.01; 0.07]
FinnGen
23andMe
0.04
[ 0.02; 0.05]
Fixed effects model
0.04
[ 0.03; 0.05]
1.17e−08
$I^2$ = 0%
Replication
IHGC16noFIno23
0.01
[−0.02; 0.03]
HUNT
0.04
[ 0.00; 0.08]
Fixed effects model
0.01
[−0.01; 0.03]
1.65e−01
$I^2$ = 56%
Fixed effects model
0.03
[ 0.02; 0.04]
3.61e−08
−0.05
0
0.05

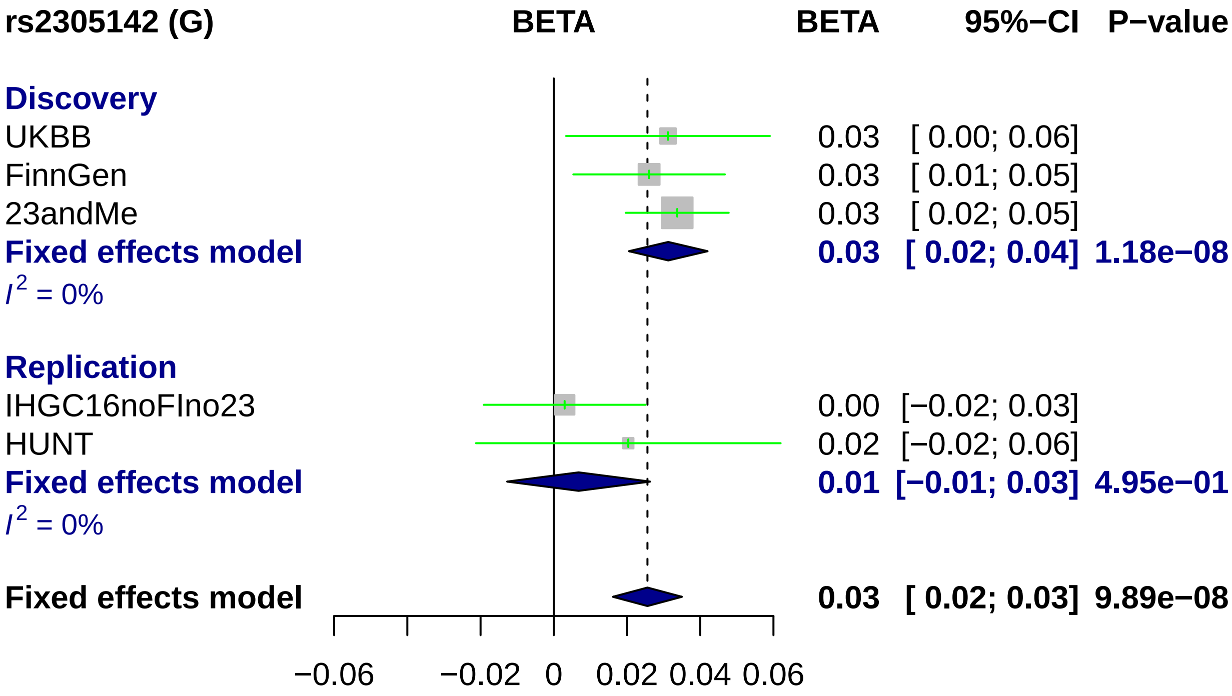
rs2305142 (G)
BETA
BETA
95%−CI
P−value
Discovery
UKBB
0.03
[ 0.00; 0.06]
FinnGen
0.03
[ 0.01; 0.05]
23andMe
0.03
[ 0.02; 0.05]
Fixed effects model
0.03
[ 0.02; 0.04]
1.18e−08
$I^2$ = 0%
Replication
IHGC16noFIno23
0.00
[−0.02; 0.03]
HUNT
0.02
[−0.02; 0.06]
Fixed effects model
0.01
[−0.01; 0.03]
4.95e−01
$I^2$ = 0%
Fixed effects model
0.03
[ 0.02; 0.03]
9.89e−08
−0.06
−0.02
0
0.02
0.04
0.06

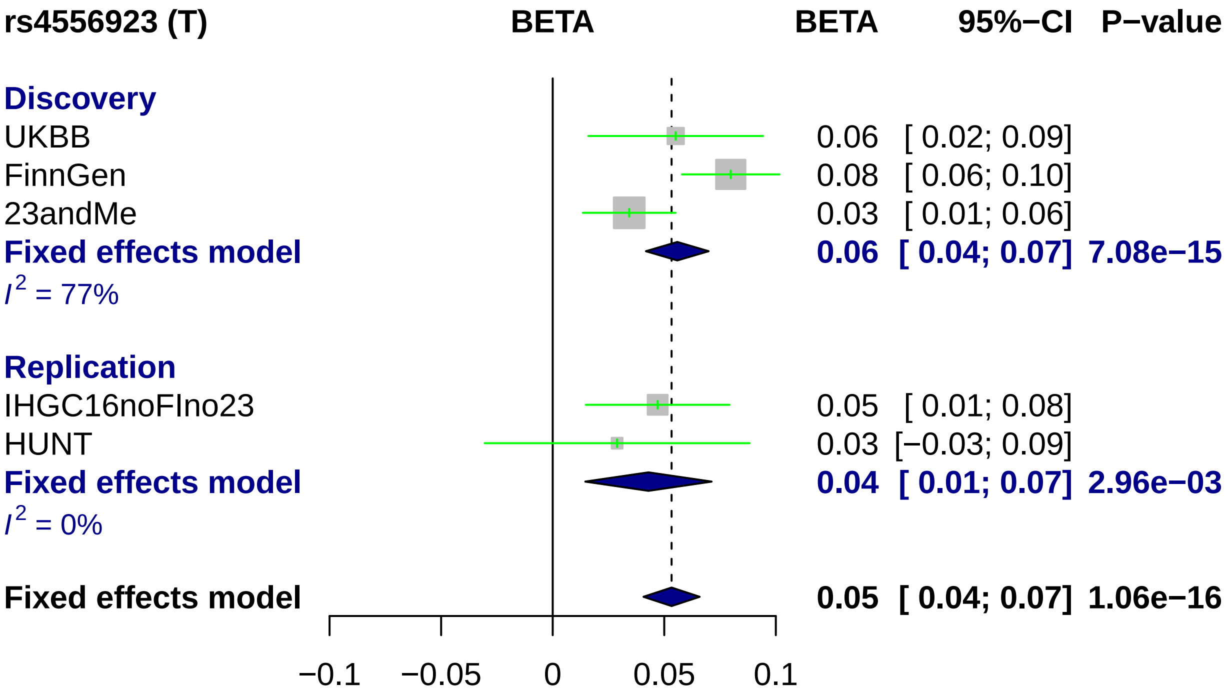
rs4556923 (T)
BETA
BETA
95%−CI
P−value
Discovery
UKBB
0.06 [ 0.02; 0.09]
FinnGen
0.08 [ 0.06; 0.10]
23andMe
0.03 [ 0.01; 0.06]
Fixed effects model
0.06 [ 0.04; 0.07] 7.08e−15
I2 = 77%
Replication
IHGC16noFIno23
0.05 [ 0.01; 0.08]
HUNT
0.03 [−0.03; 0.09]
Fixed effects model
0.04 [ 0.01; 0.07] 2.96e−03
I2 = 0%
Fixed effects model
0.05 [ 0.04; 0.07] 1.06e−16
−0.1
−0.05
0
0.05
0.1

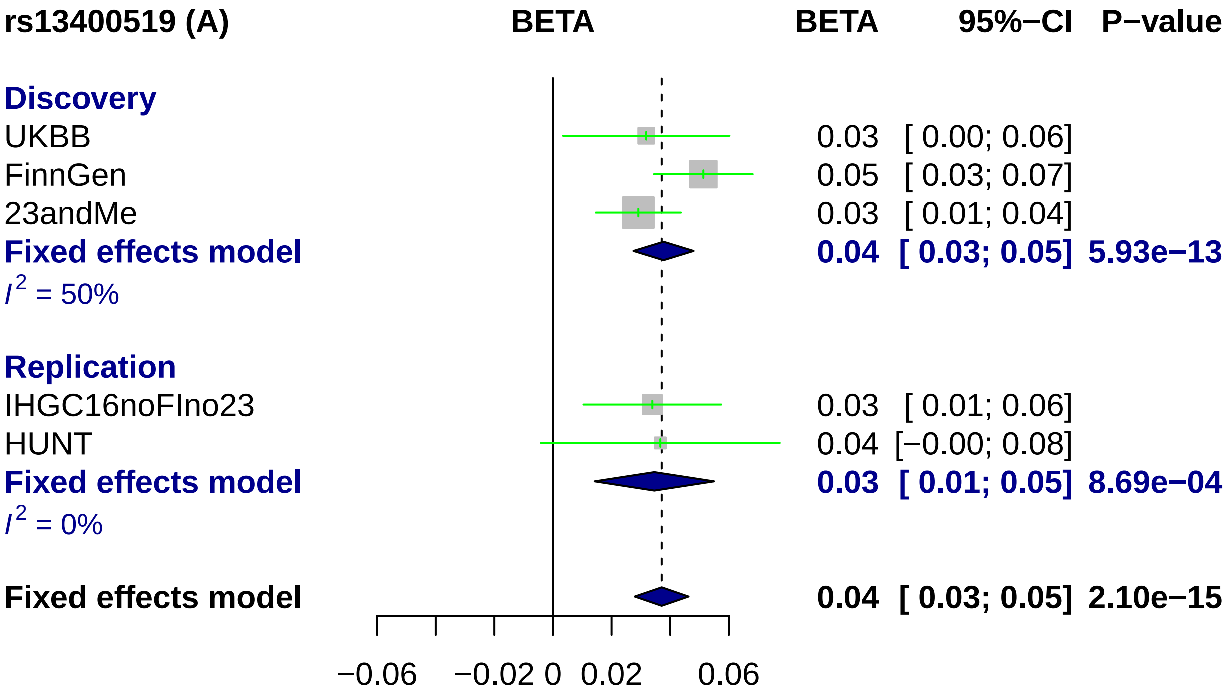
rs13400519 (A)
BETA
BETA
95%−CI
P−value
Discovery
UKBB
0.03 [ 0.00; 0.06]
FinnGen
0.05 [ 0.03; 0.07]
23andMe
0.03 [ 0.01; 0.04]
Fixed effects model
0.04 [ 0.03; 0.05] 5.93e−13
I2 = 50%
Replication
IHGC16noFIno23
0.03 [ 0.01; 0.06]
HUNT
0.04 [−0.00; 0.08]
Fixed effects model
0.03 [ 0.01; 0.05] 8.69e−04
I2 = 0%
Fixed effects model
0.04 [ 0.03; 0.05] 2.10e−15
−0.06
−0.02
0
0.02
0.06

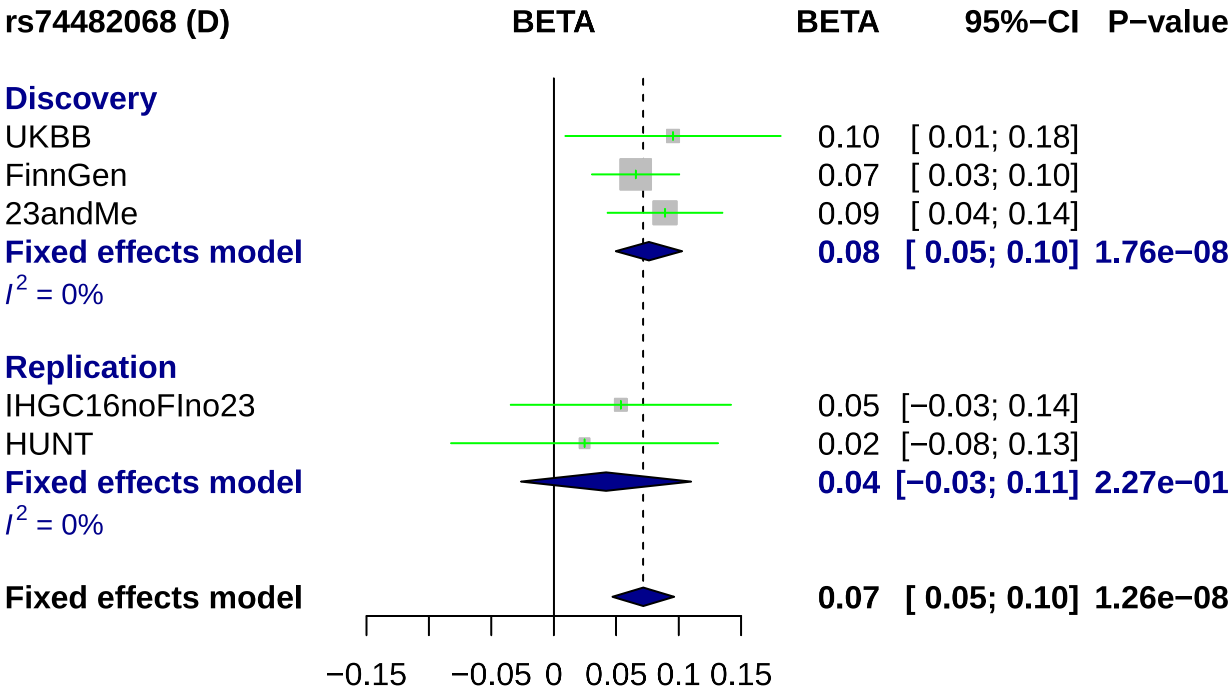
rs74482068 (D)
BETA
BETA 95%−CI P−value
Discovery
UKBB 0.10 [ 0.01; 0.18]
FinnGen 0.07 [ 0.03; 0.10]
23andMe 0.09 [ 0.04; 0.14]
Fixed effects model 0.08 [ 0.05; 0.10] 1.76e−08
I2 = 0%
Replication
IHGC16noFIno23 0.05 [−0.03; 0.14]
HUNT 0.02 [−0.08; 0.13]
Fixed effects model 0.04 [−0.03; 0.11] 2.27e−01
I2 = 0%
Fixed effects model 0.07 [ 0.05; 0.10] 1.26e−08
−0.15
−0.05
0
0.05
0.1
0.15

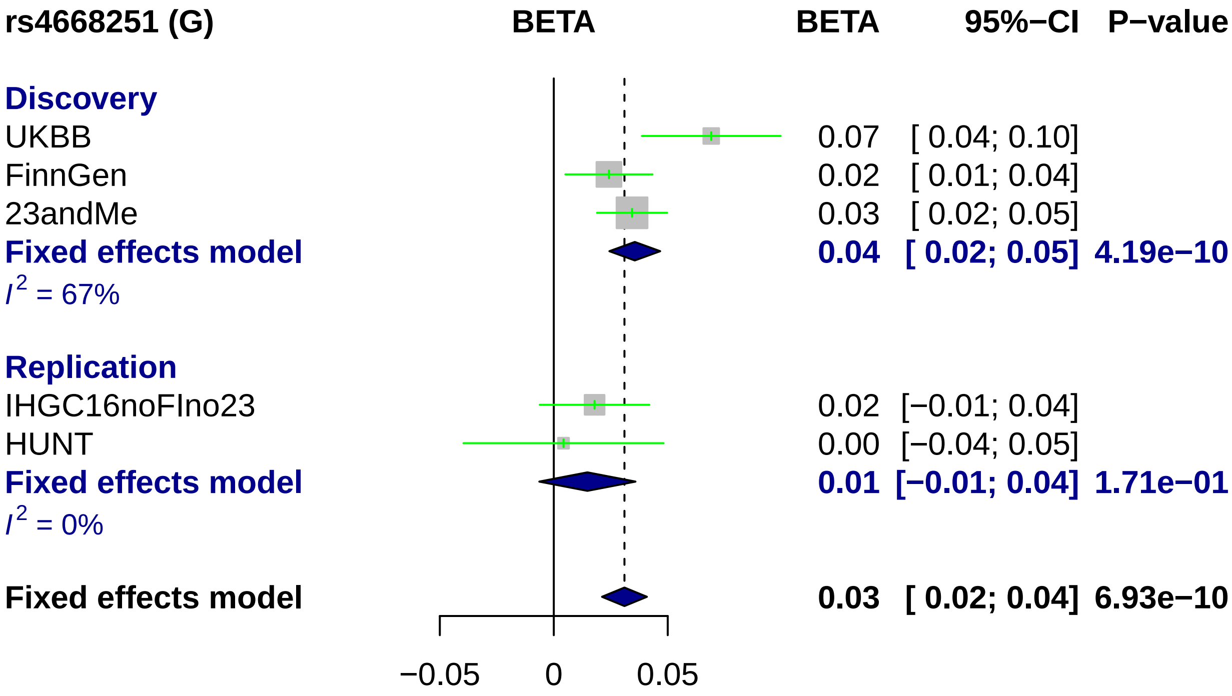
rs4668251 (G)
BETA
BETA
95%−CI
P−value
Discovery
UKBB
0.07
[ 0.04; 0.10]
FinnGen
0.02
[ 0.01; 0.04]
23andMe
0.03
[ 0.02; 0.05]
Fixed effects model
0.04
[ 0.02; 0.05]
4.19e−10
I2 = 67%
Replication
IHGC16noFIno23
0.02
[−0.01; 0.04]
HUNT
0.00
[−0.04; 0.05]
Fixed effects model
0.01
[−0.01; 0.04]
1.71e−01
I2 = 0%
Fixed effects model
0.03
[ 0.02; 0.04]
6.93e−10
−0.05
0
0.05

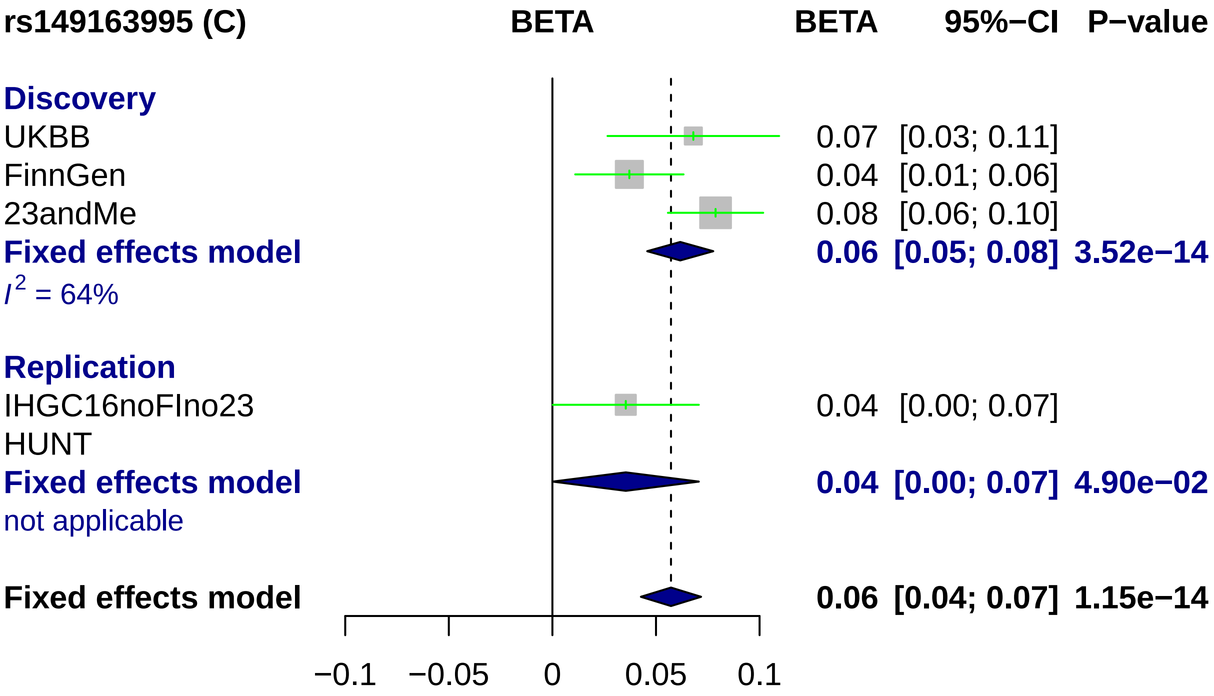
rs149163995 (C)
BETA
BETA
95%−CI
P−value
Discovery
UKBB
0.07 [0.03; 0.11]
FinnGen
0.04 [0.01; 0.06]
23andMe
0.08 [0.06; 0.10]
Fixed effects model
0.06 [0.05; 0.08] 3.52e−14
I2 = 64%
Replication
IHGC16noFIno23
0.04 [0.00; 0.07]
HUNT
Fixed effects model
0.04 [0.00; 0.07] 4.90e−02
not applicable
Fixed effects model
0.06 [0.04; 0.07] 1.15e−14
−0.1
−0.05
0
0.05
0.1

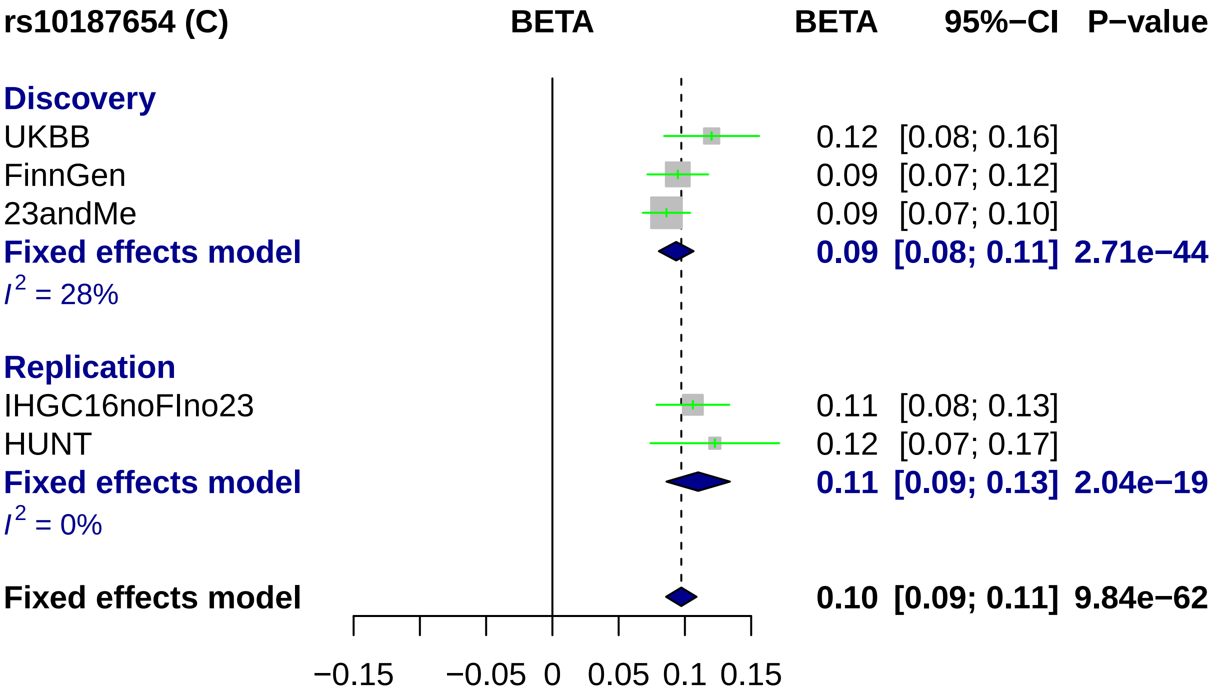
rs10187654 (C)
BETA
BETA
95%−CI
P−value
Discovery
UKBB
0.12 [0.08; 0.16]
FinnGen
0.09 [0.07; 0.12]
23andMe
0.09 [0.07; 0.10]
Fixed effects model
0.09 [0.08; 0.11] 2.71e−44
I2 = 28%
Replication
IHGC16noFIno23
0.11 [0.08; 0.13]
HUNT
0.12 [0.07; 0.17]
Fixed effects model
0.11 [0.09; 0.13] 2.04e−19
I2 = 0%
Fixed effects model
0.10 [0.09; 0.11] 9.84e−62
−0.15
−0.05
0
0.05
0.1
0.15

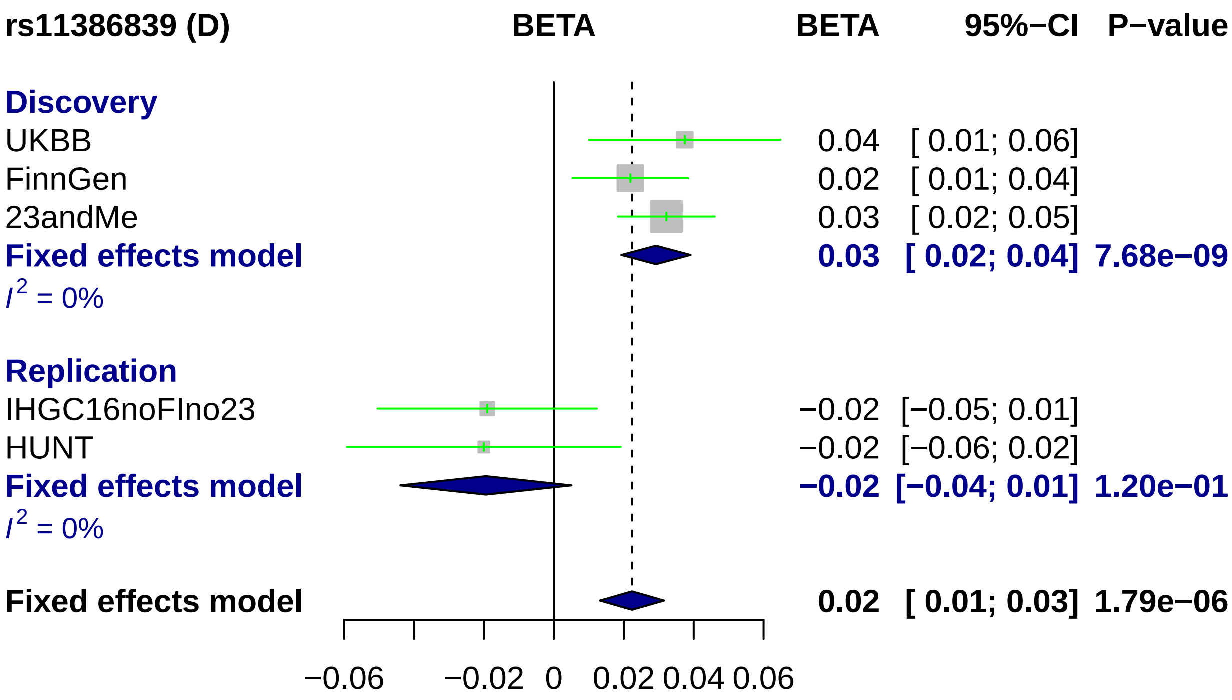
rs11386839 (D)
BETA
BETA
95%−CI
P−value
Discovery
UKBB
0.04
[ 0.01; 0.06]
FinnGen
0.02
[ 0.01; 0.04]
23andMe
0.03
[ 0.02; 0.05]
Fixed effects model
0.03
[ 0.02; 0.04]
7.68e−09
$I^2$ = 0%
Replication
IHGC16noFIno23
−0.02
[−0.05; 0.01]
HUNT
−0.02
[−0.06; 0.02]
Fixed effects model
−0.02
[−0.04; 0.01]
1.20e−01
$I^2$ = 0%
Fixed effects model
0.02
[ 0.01; 0.03]
1.79e−06
−0.06
−0.02
0
0.02
0.04
0.06

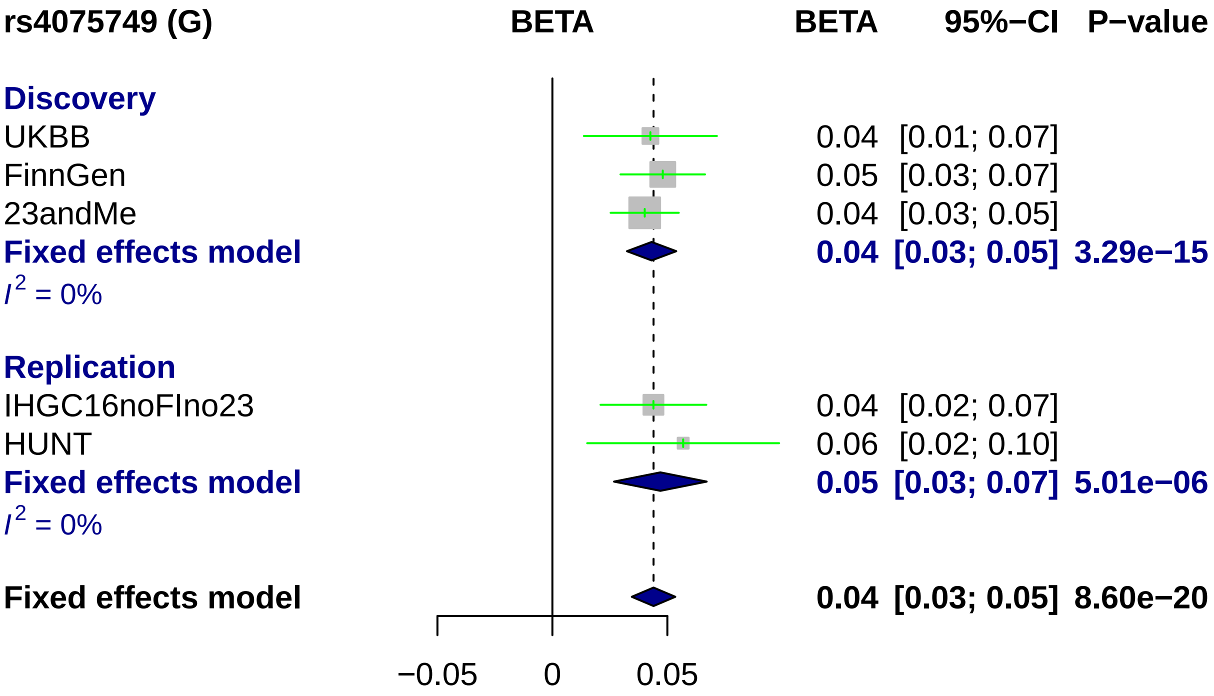

rs4075749 (G)
BETA
BETA
95%−CI
P−value
Discovery
UKBB
0.04 [0.01; 0.07]
FinnGen
0.05 [0.03; 0.07]
23andMe
0.04 [0.03; 0.05]
Fixed effects model
0.04 [0.03; 0.05] 3.29e−15
I^2 = 0%
Replication
IHGC16noFIno23
0.04 [0.02; 0.07]
HUNT
0.06 [0.02; 0.10]
Fixed effects model
0.05 [0.03; 0.07] 5.01e−06
I^2 = 0%
Fixed effects model
0.04 [0.03; 0.05] 8.60e−20
−0.05
0
0.05

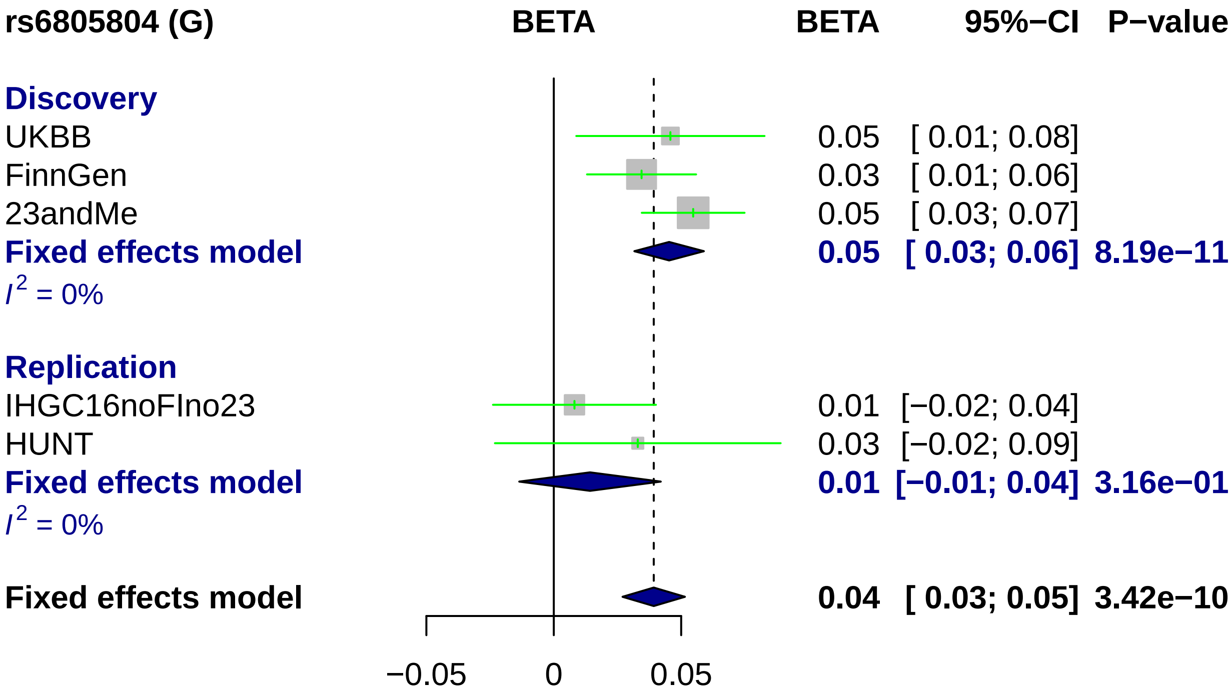
rs6805804 (G)
BETA
BETA
95%−CI
P−value
Discovery
UKBB
0.05
[ 0.01; 0.08]
FinnGen
0.03
[ 0.01; 0.06]
23andMe
0.05
[ 0.03; 0.07]
Fixed effects model
0.05
[ 0.03; 0.06]
8.19e−11
I2 = 0%
Replication
IHGC16noFIno23
0.01
[−0.02; 0.04]
HUNT
0.03
[−0.02; 0.09]
Fixed effects model
0.01
[−0.01; 0.04]
3.16e−01
I2 = 0%
Fixed effects model
0.04
[ 0.03; 0.05]
3.42e−10
−0.05
0
0.05

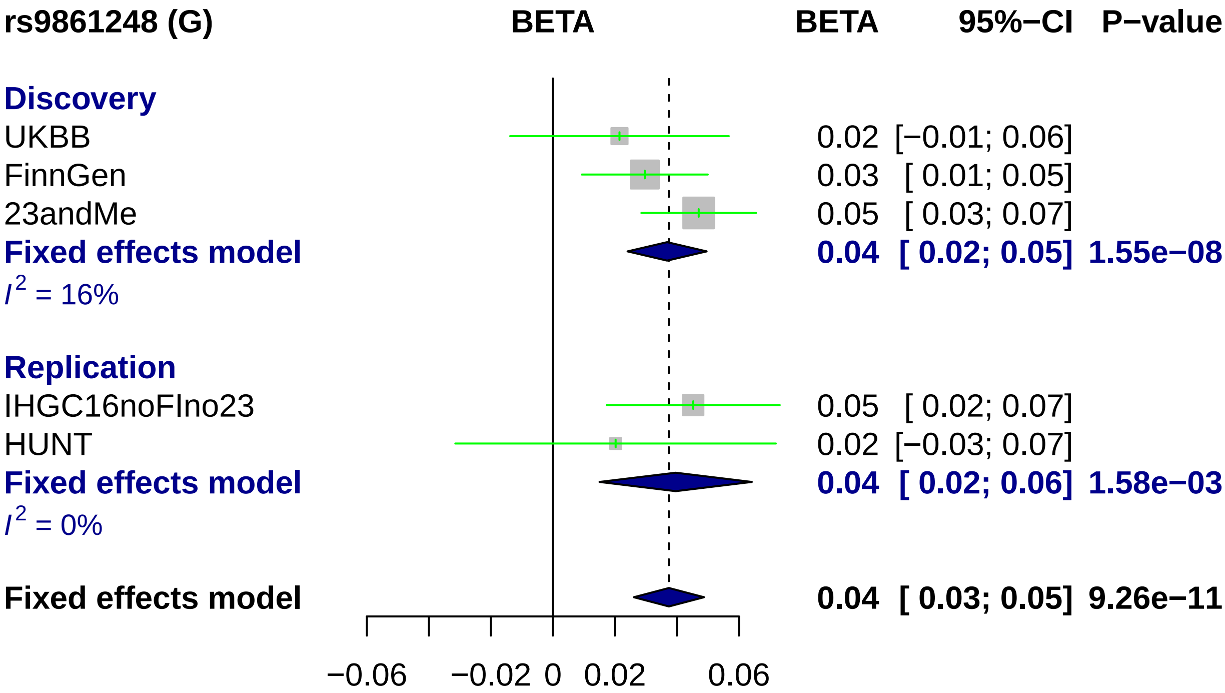
rs9861248 (G)
BETA
BETA
95%−CI
P−value
Discovery
UKBB
0.02 [−0.01; 0.06]
FinnGen
0.03 [ 0.01; 0.05]
23andMe
0.05 [ 0.03; 0.07]
Fixed effects model
0.04 [ 0.02; 0.05] 1.55e−08
I2 = 16%
Replication
IHGC16noFIno23
0.05 [ 0.02; 0.07]
HUNT
0.02 [−0.03; 0.07]
Fixed effects model
0.04 [ 0.02; 0.06] 1.58e−03
I2 = 0%
Fixed effects model
0.04 [ 0.03; 0.05] 9.26e−11
−0.06
−0.02
0
0.02
0.06

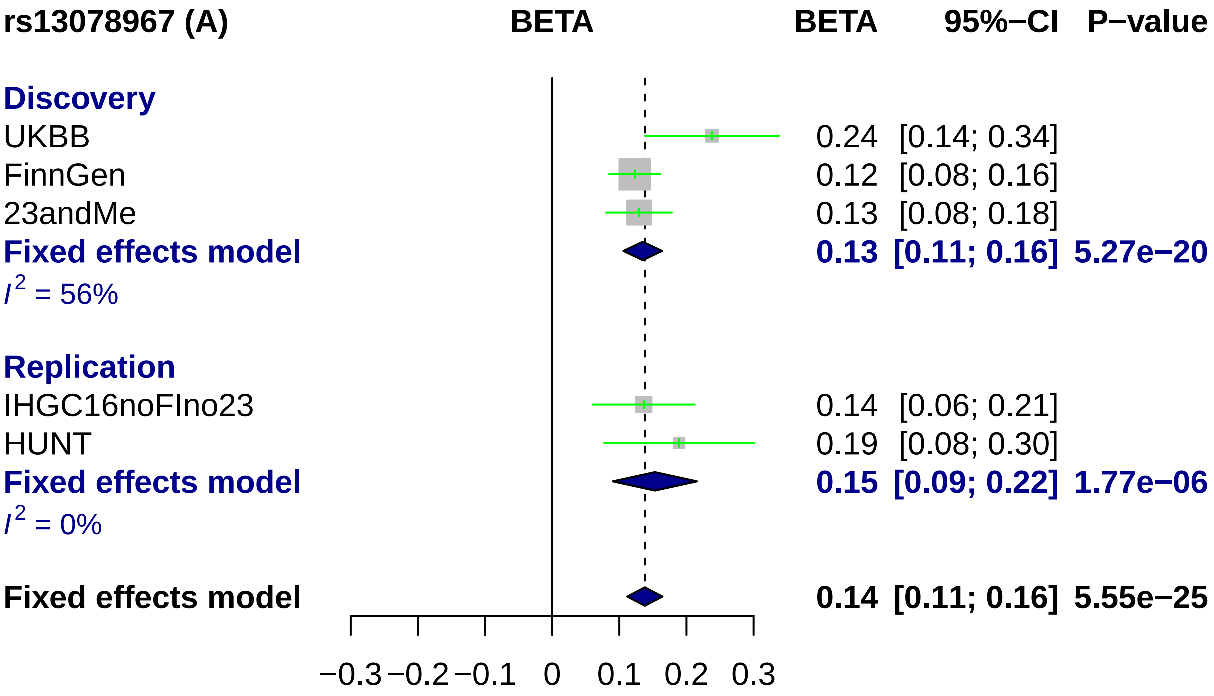

rs13078967 (A)
BETA
BETA
95%−CI
P−value
Discovery
UKBB
0.24 [0.14; 0.34]
FinnGen
0.12 [0.08; 0.16]
23andMe
0.13 [0.08; 0.18]
Fixed effects model
0.13 [0.11; 0.16] 5.27e−20
I2 = 56%
Replication
IHGC16noFIno23
0.14 [0.06; 0.21]
HUNT
0.19 [0.08; 0.30]
Fixed effects model
0.15 [0.09; 0.22] 1.77e−06
I2 = 0%
Fixed effects model
0.14 [0.11; 0.16] 5.55e−25
−0.3
−0.2
−0.1
0
0.1
0.2
0.3

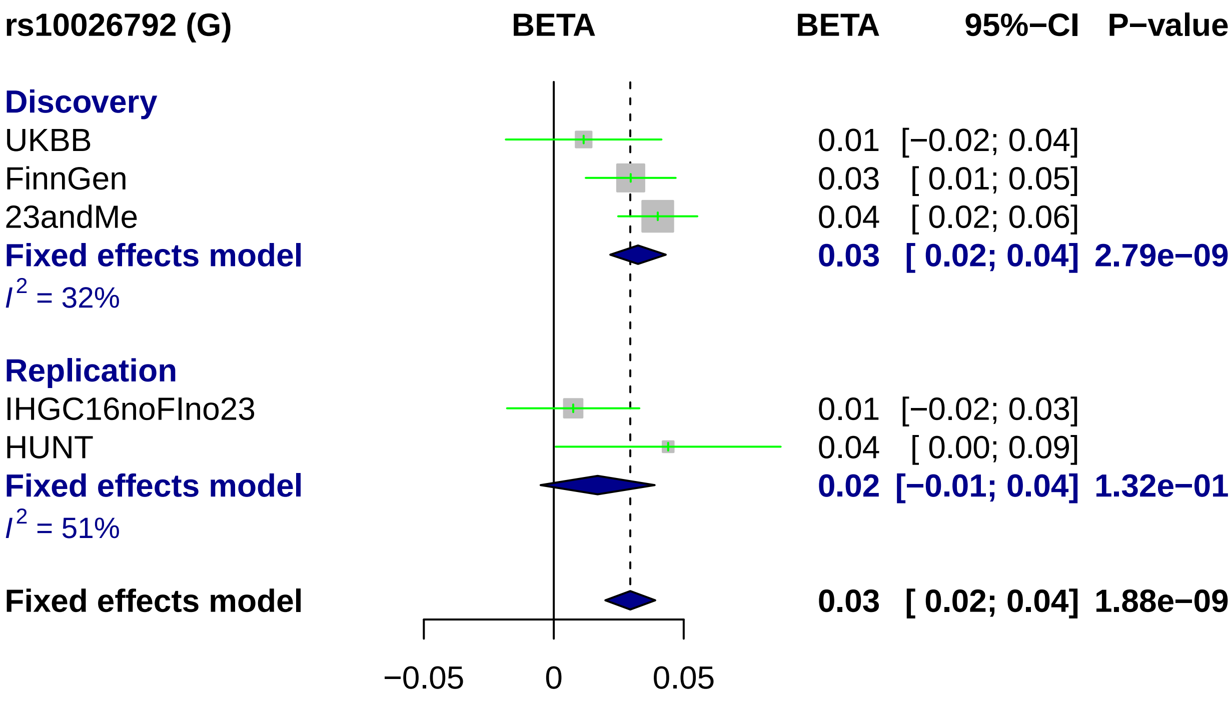
rs10026792 (G)
BETA
BETA
95%−CI
P−value
Discovery
UKBB
0.01 [−0.02; 0.04]
FinnGen
0.03 [ 0.01; 0.05]
23andMe
0.04 [ 0.02; 0.06]
Fixed effects model
0.03 [ 0.02; 0.04] 2.79e−09
I² = 32%
Replication
IHGC16noFIno23
0.01 [−0.02; 0.03]
HUNT
0.04 [ 0.00; 0.09]
Fixed effects model
0.02 [−0.01; 0.04] 1.32e−01
I² = 51%
Fixed effects model
0.03 [ 0.02; 0.04] 1.88e−09
−0.05
0
0.05

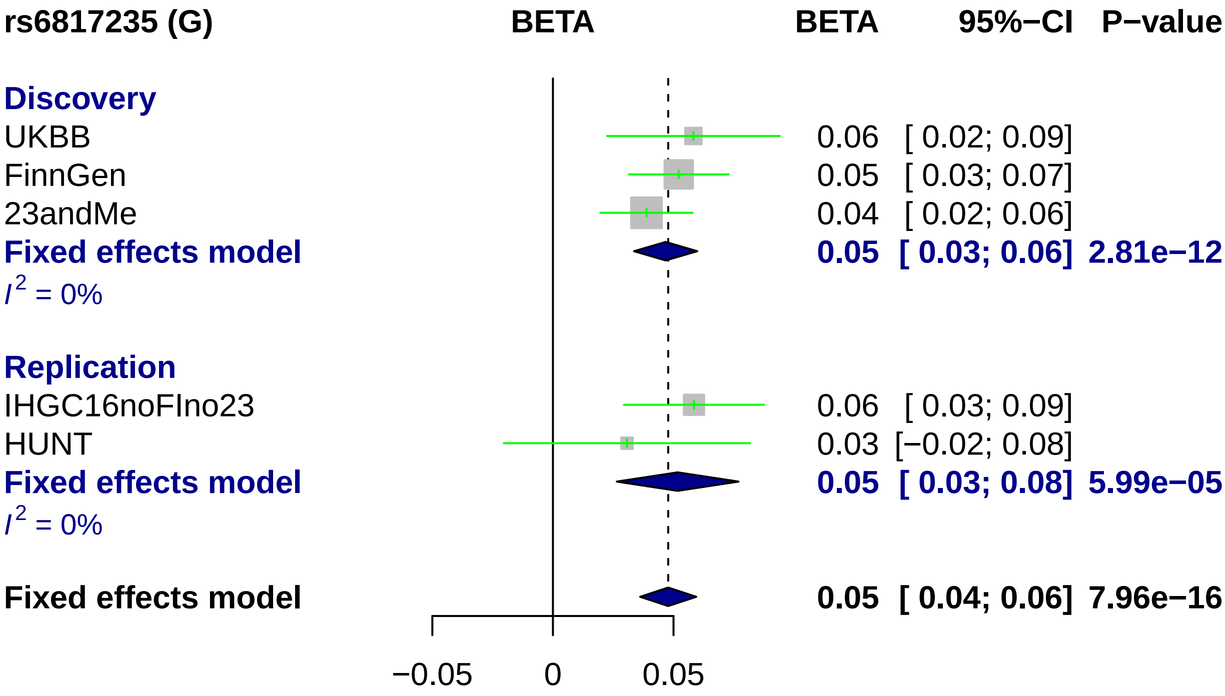
rs6817235 (G)
BETA
BETA
95%−CI
P−value
Discovery
UKBB
0.06 [ 0.02; 0.09]
FinnGen
0.05 [ 0.03; 0.07]
23andMe
0.04 [ 0.02; 0.06]
Fixed effects model
0.05 [ 0.03; 0.06] 2.81e−12
$I^2$ = 0%
Replication
IHGC16noFIno23
0.06 [ 0.03; 0.09]
HUNT
0.03 [−0.02; 0.08]
Fixed effects model
0.05 [ 0.03; 0.08] 5.99e−05
$I^2$ = 0%
Fixed effects model
0.05 [ 0.04; 0.06] 7.96e−16
−0.05
0
0.05

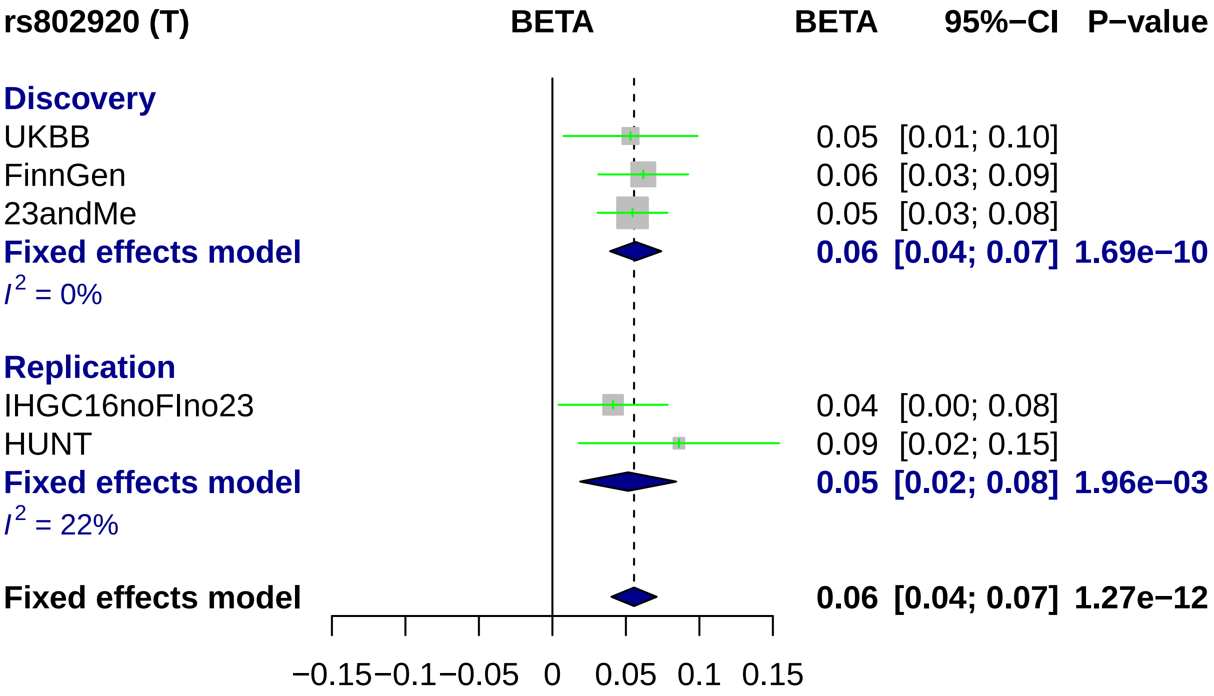

rs802920 (T)
BETA
BETA
95%−CI
P−value
Discovery
UKBB
0.05 [0.01; 0.10]
FinnGen
0.06 [0.03; 0.09]
23andMe
0.05 [0.03; 0.08]
Fixed effects model
0.06 [0.04; 0.07] 1.69e−10
I2 = 0%
Replication
IHGC16noFIno23
0.04 [0.00; 0.08]
HUNT
0.09 [0.02; 0.15]
Fixed effects model
0.05 [0.02; 0.08] 1.96e−03
I2 = 22%
Fixed effects model
0.06 [0.04; 0.07] 1.27e−12
−0.15 −0.1 −0.05 0 0.05 0.1 0.15

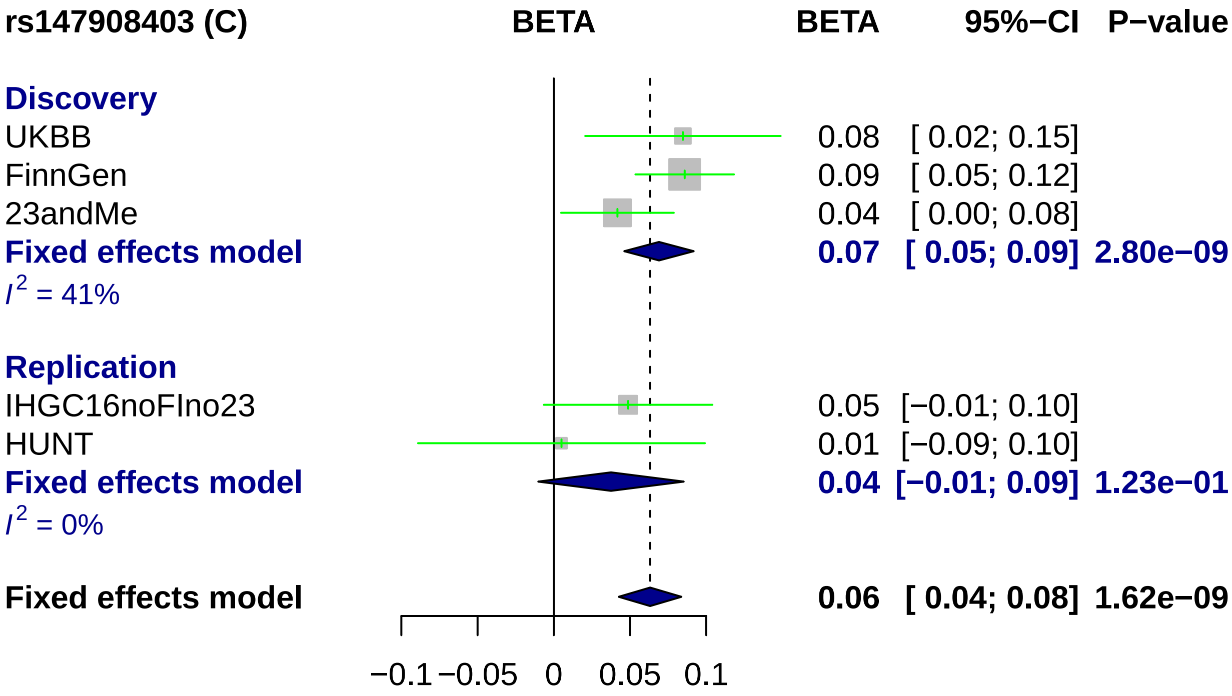
rs147908403 (C)
BETA
BETA
95%−CI
P−value
Discovery
UKBB
0.08
[ 0.02; 0.15]
FinnGen
0.09
[ 0.05; 0.12]
23andMe
0.04
[ 0.00; 0.08]
Fixed effects model
0.07
[ 0.05; 0.09]
2.80e−09
I2 = 41%
Replication
IHGC16noFIno23
0.05
[−0.01; 0.10]
HUNT
0.01
[−0.09; 0.10]
Fixed effects model
0.04
[−0.01; 0.09]
1.23e−01
I2 = 0%
Fixed effects model
0.06
[ 0.04; 0.08]
1.62e−09
−0.1
−0.05
0
0.05
0.1

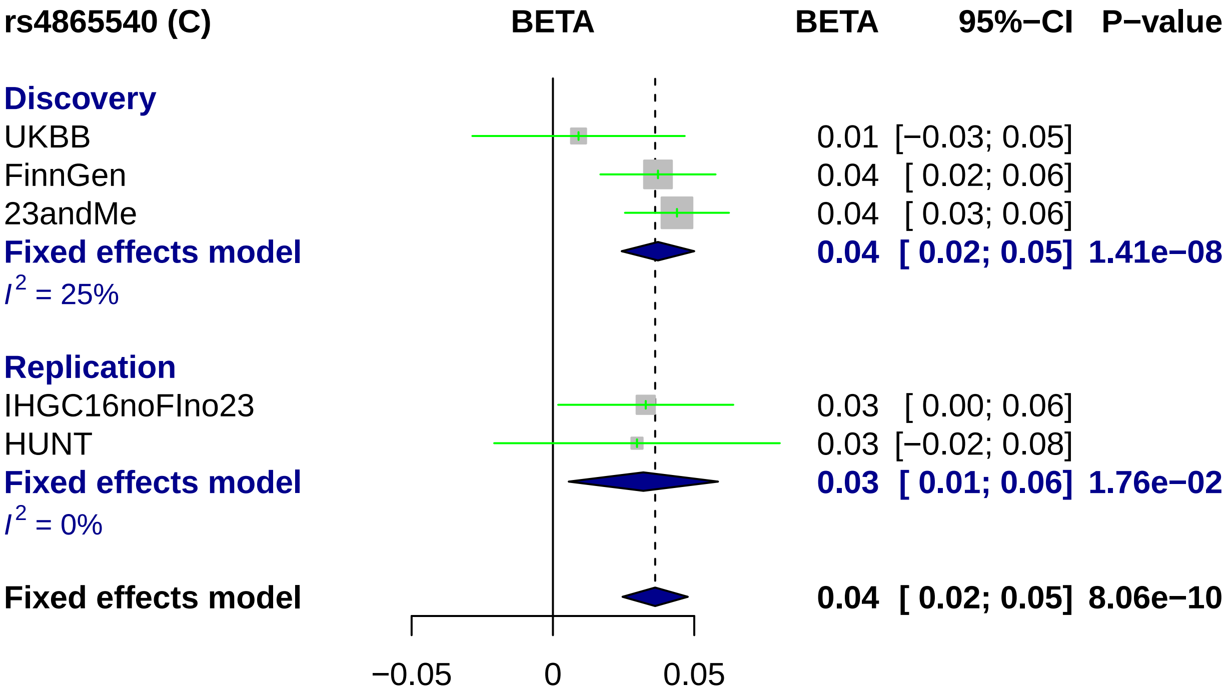
rs4865540 (C)
BETA
BETA 95%−CI P−value
Discovery
UKBB 0.01 [−0.03; 0.05]
FinnGen 0.04 [ 0.02; 0.06]
23andMe 0.04 [ 0.03; 0.06]
Fixed effects model 0.04 [ 0.02; 0.05] 1.41e−08
I2 = 25%
Replication
IHGC16noFIno23 0.03 [ 0.00; 0.06]
HUNT 0.03 [−0.02; 0.08]
Fixed effects model 0.03 [ 0.01; 0.06] 1.76e−02
I2 = 0%
Fixed effects model 0.04 [ 0.02; 0.05] 8.06e−10
−0.05
0
0.05

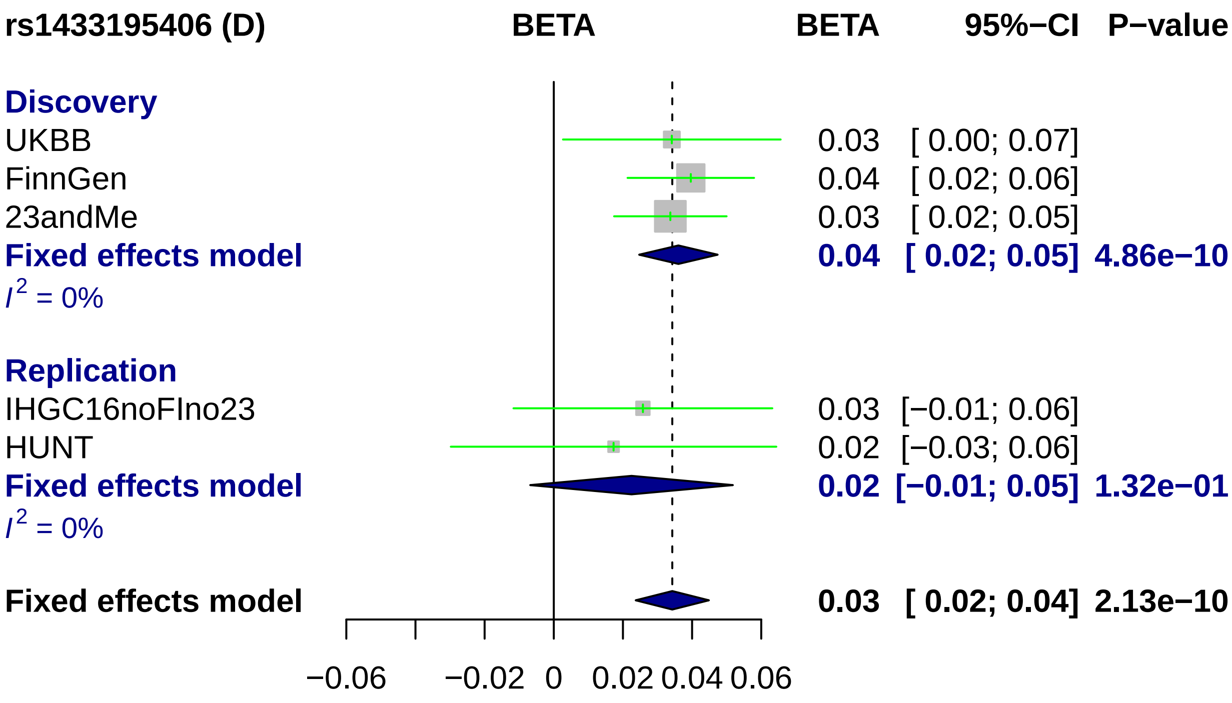
rs1433195406 (D)
BETA
BETA
95%−CI
P−value
Discovery
UKBB
0.03
[ 0.00; 0.07]
FinnGen
0.04
[ 0.02; 0.06]
23andMe
0.03
[ 0.02; 0.05]
Fixed effects model
0.04
[ 0.02; 0.05]
4.86e−10
I2 = 0%
Replication
IHGC16noFIno23
0.03
[−0.01; 0.06]
HUNT
0.02
[−0.03; 0.06]
Fixed effects model
0.02
[−0.01; 0.05]
1.32e−01
I2 = 0%
Fixed effects model
0.03
[ 0.02; 0.04]
2.13e−10
−0.06
−0.02
0
0.02
0.04
0.06

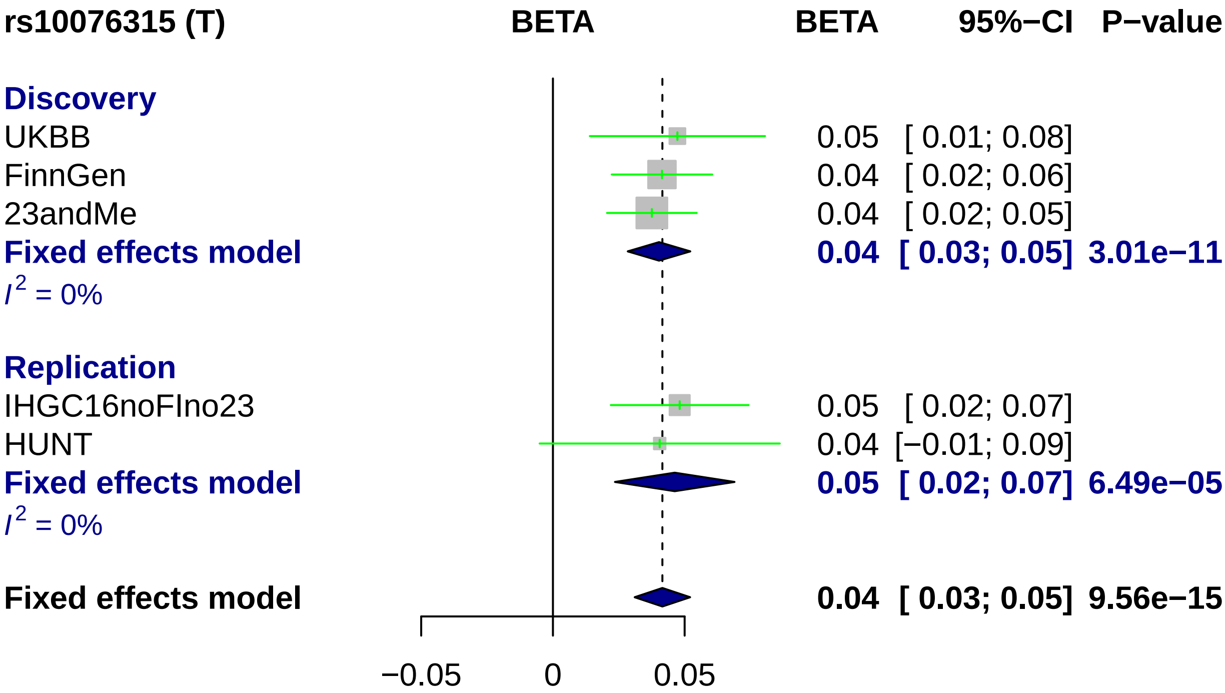
rs10076315 (T)
BETA
BETA
95%−CI
P−value
Discovery
UKBB
0.05
[ 0.01; 0.08]
FinnGen
0.04
[ 0.02; 0.06]
23andMe
0.04
[ 0.02; 0.05]
Fixed effects model
0.04
[ 0.03; 0.05]
3.01e−11
I2 = 0%
Replication
IHGC16noFIno23
0.05
[ 0.02; 0.07]
HUNT
0.04
[−0.01; 0.09]
Fixed effects model
0.05
[ 0.02; 0.07]
6.49e−05
I2 = 0%
Fixed effects model
0.04
[ 0.03; 0.05]
9.56e−15
−0.05
0
0.05

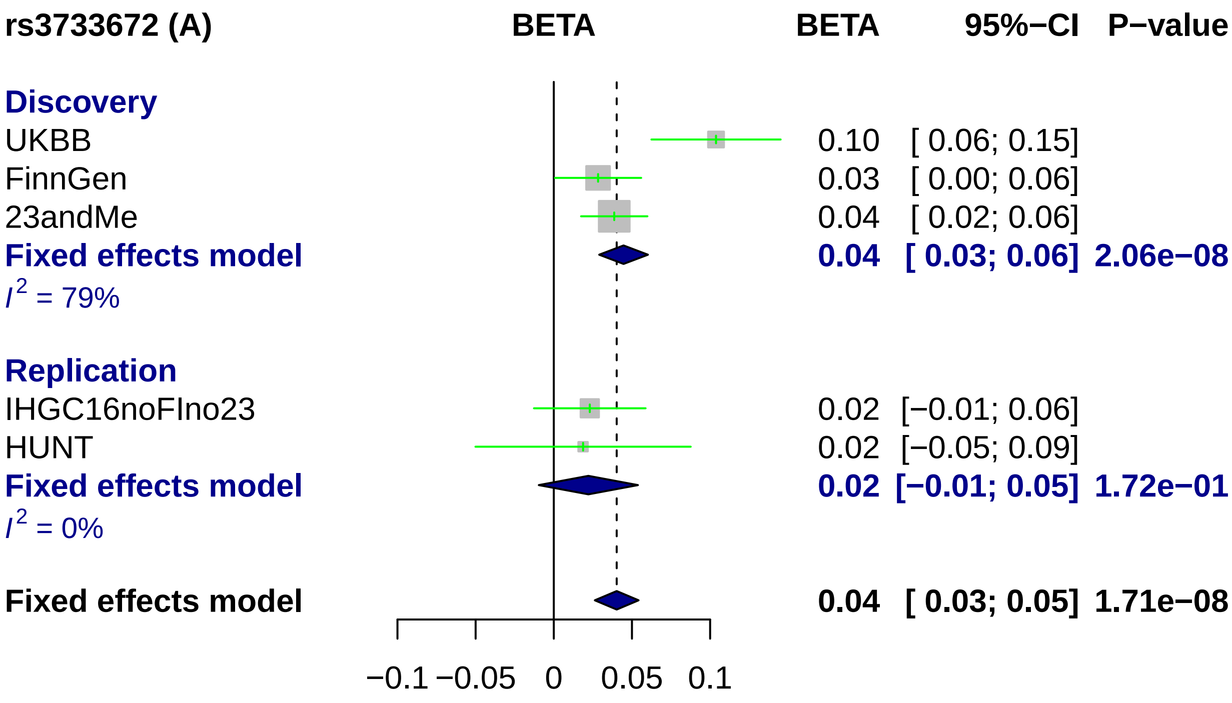
rs3733672 (A)
BETA
BETA
95%−CI
P−value
Discovery
UKBB
0.10
[ 0.06; 0.15]
FinnGen
0.03
[ 0.00; 0.06]
23andMe
0.04
[ 0.02; 0.06]
Fixed effects model
0.04
[ 0.03; 0.06]
2.06e−08
I2 = 79%
Replication
IHGC16noFIno23
0.02
[−0.01; 0.06]
HUNT
0.02
[−0.05; 0.09]
Fixed effects model
0.02
[−0.01; 0.05]
1.72e−01
I2 = 0%
Fixed effects model
0.04
[ 0.03; 0.05]
1.71e−08
−0.1
−0.05
0
0.05
0.1

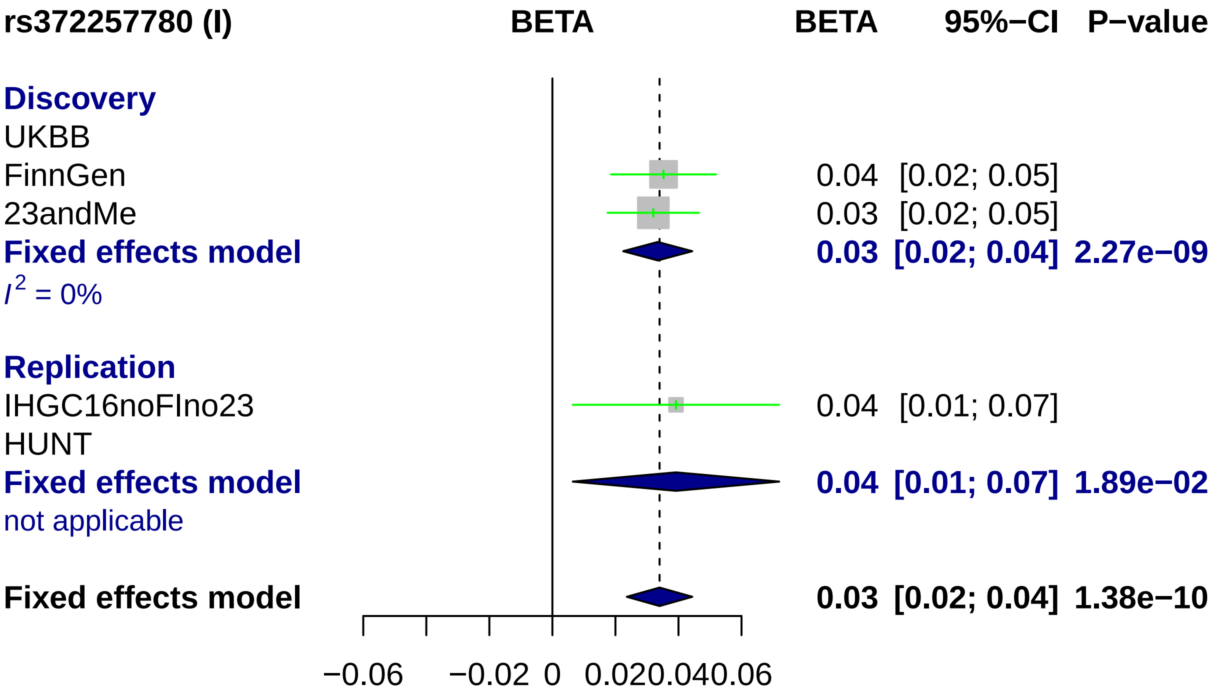

rs372257780 (I)
BETA
BETA
95%−CI
P−value
Discovery
UKBB
FinnGen
0.04 [0.02; 0.05]
23andMe
0.03 [0.02; 0.05]
Fixed effects model
0.03 [0.02; 0.04] 2.27e−09
$I^2$ = 0%
Replication
IHGC16noFIno23
0.04 [0.01; 0.07]
HUNT
Fixed effects model
0.04 [0.01; 0.07] 1.89e−02
not applicable
Fixed effects model
0.03 [0.02; 0.04] 1.38e−10
−0.06
−0.02
0
0.02
0.04
0.06

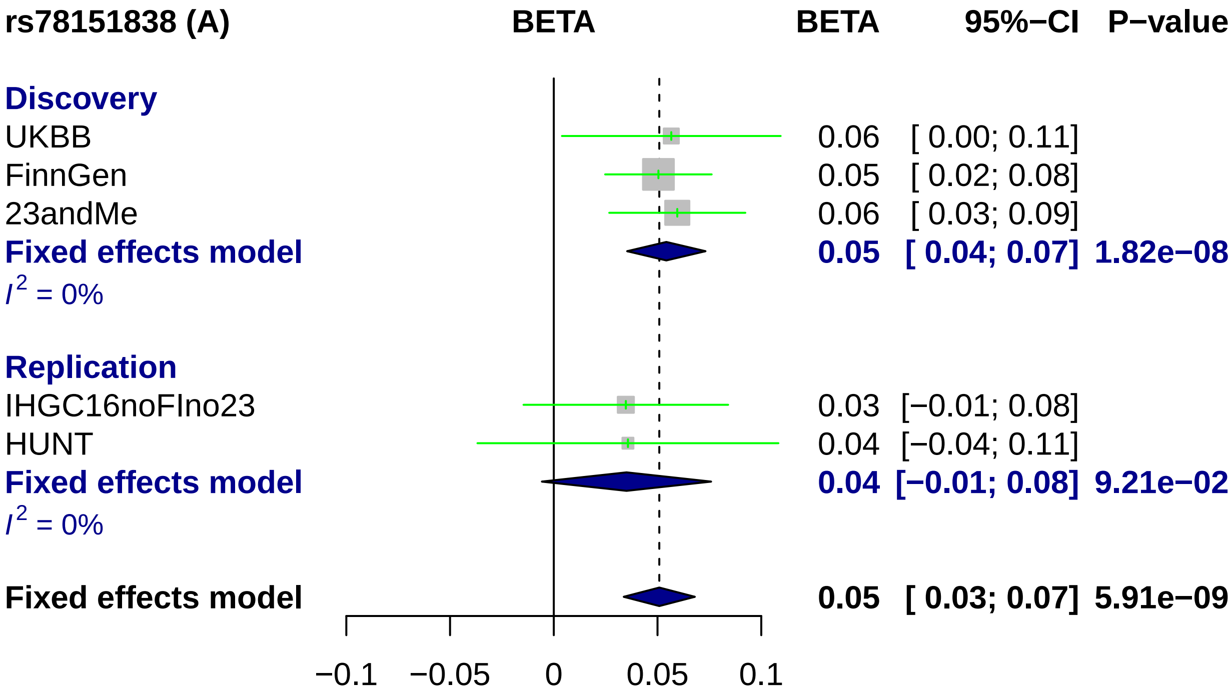
rs78151838 (A)
BETA
BETA
95%−CI
P−value
Discovery
UKBB 0.06 [ 0.00; 0.11]
FinnGen 0.05 [ 0.02; 0.08]
23andMe 0.06 [ 0.03; 0.09]
Fixed effects model 0.05 [ 0.04; 0.07] 1.82e−08
$I^2$ = 0%
Replication
IHGC16noFIno23 0.03 [−0.01; 0.08]
HUNT 0.04 [−0.04; 0.11]
Fixed effects model 0.04 [−0.01; 0.08] 9.21e−02
$I^2$ = 0%
Fixed effects model 0.05 [ 0.03; 0.07] 5.91e−09
−0.1
−0.05
0
0.05
0.1

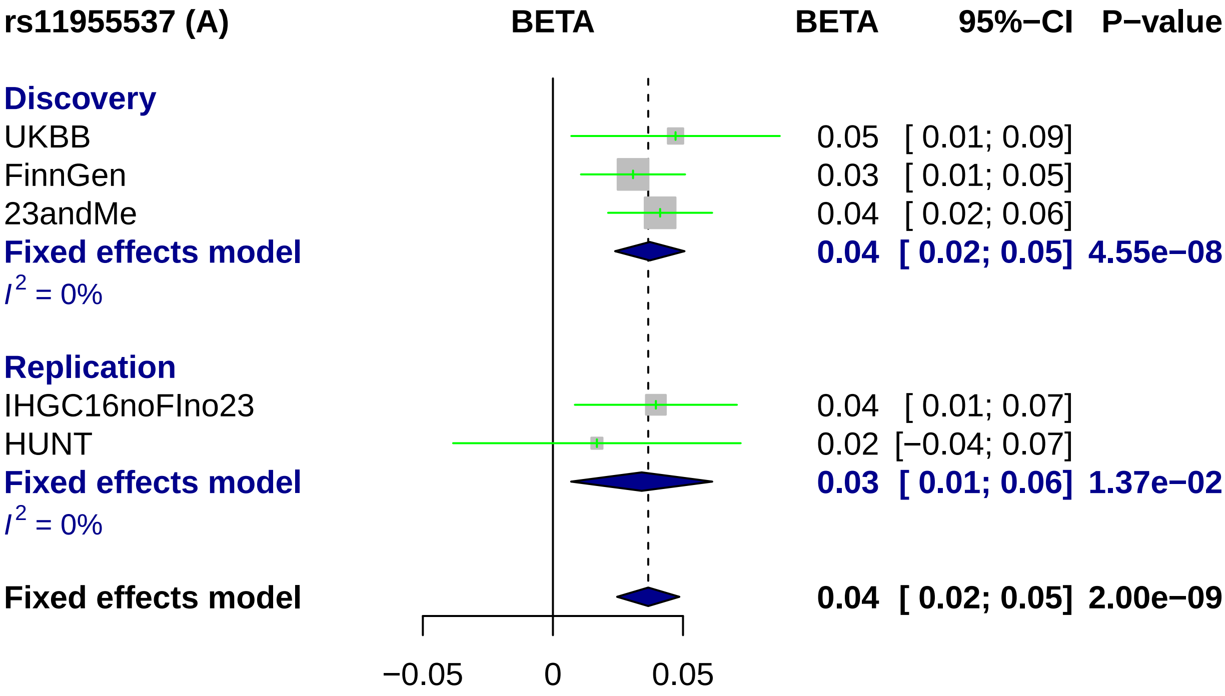
rs11955537 (A)
BETA
BETA
95%−CI
P−value
Discovery
UKBB
0.05 [ 0.01; 0.09]
FinnGen
0.03 [ 0.01; 0.05]
23andMe
0.04 [ 0.02; 0.06]
Fixed effects model
0.04 [ 0.02; 0.05] 4.55e−08
I2 = 0%
Replication
IHGC16noFIno23
0.04 [ 0.01; 0.07]
HUNT
0.02 [−0.04; 0.07]
Fixed effects model
0.03 [ 0.01; 0.06] 1.37e−02
I2 = 0%
Fixed effects model
0.04 [ 0.02; 0.05] 2.00e−09
−0.05
0
0.05

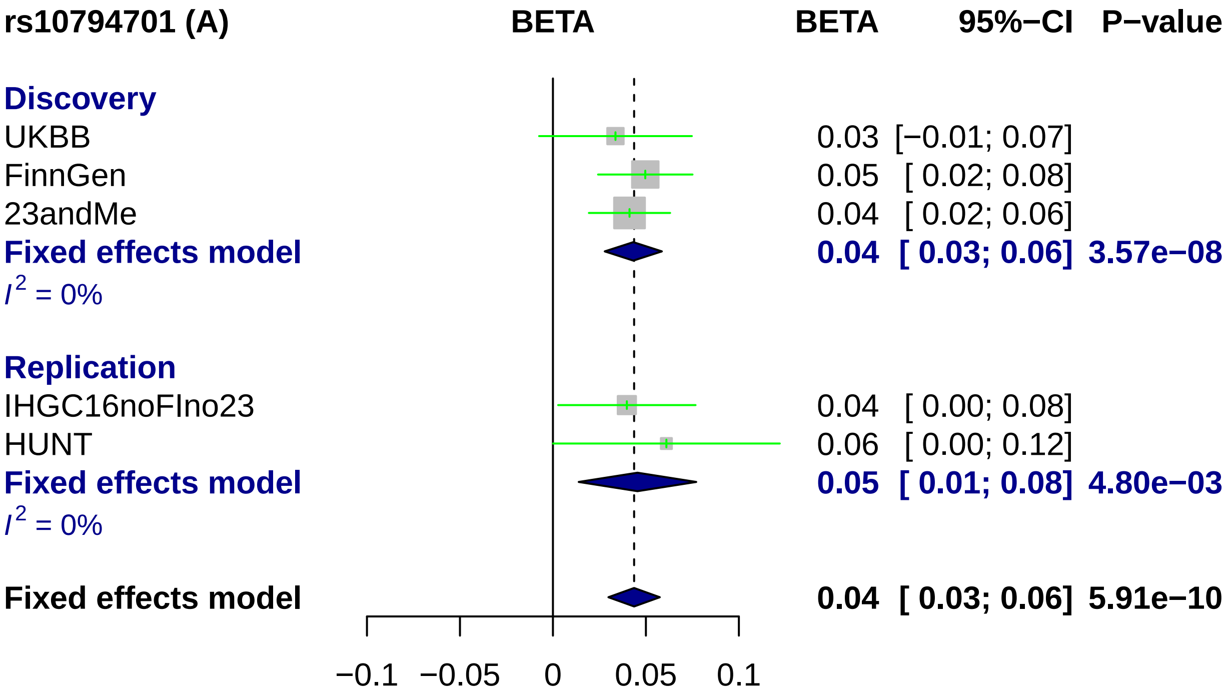
rs10794701 (A)
BETA
BETA
95%−CI
P−value
Discovery
UKBB
0.03 [−0.01; 0.07]
FinnGen
0.05 [ 0.02; 0.08]
23andMe
0.04 [ 0.02; 0.06]
Fixed effects model
0.04 [ 0.03; 0.06] 3.57e−08
I^2 = 0%
Replication
IHGC16noFIno23
0.04 [ 0.00; 0.08]
HUNT
0.06 [ 0.00; 0.12]
Fixed effects model
0.05 [ 0.01; 0.08] 4.80e−03
I^2 = 0%
Fixed effects model
0.04 [ 0.03; 0.06] 5.91e−10
−0.1
−0.05
0
0.05
0.1

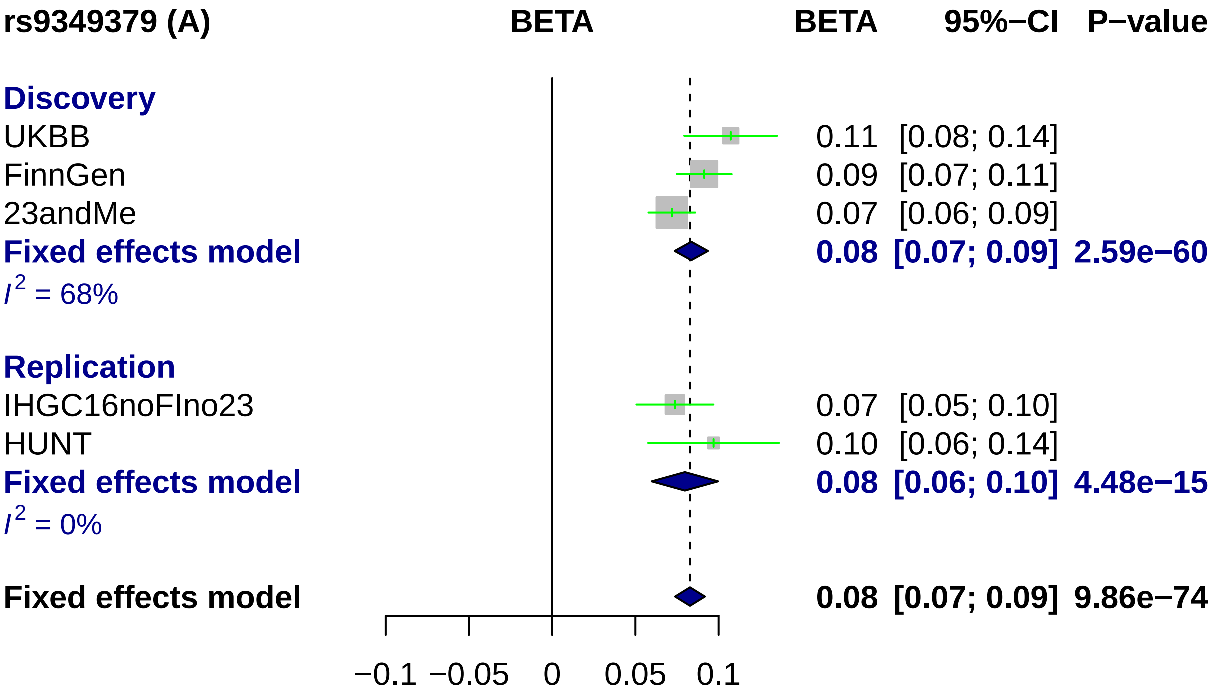

rs9349379 (A)
BETA
BETA
95%−CI
P−value
Discovery
UKBB
0.11
[0.08; 0.14]
FinnGen
0.09
[0.07; 0.11]
23andMe
0.07
[0.06; 0.09]
Fixed effects model
0.08
[0.07; 0.09]
2.59e−60
I2 = 68%
Replication
IHGC16noFIno23
0.07
[0.05; 0.10]
HUNT
0.10
[0.06; 0.14]
Fixed effects model
0.08
[0.06; 0.10]
4.48e−15
I2 = 0%
Fixed effects model
0.08
[0.07; 0.09]
9.86e−74
−0.1
−0.05
0
0.05
0.1

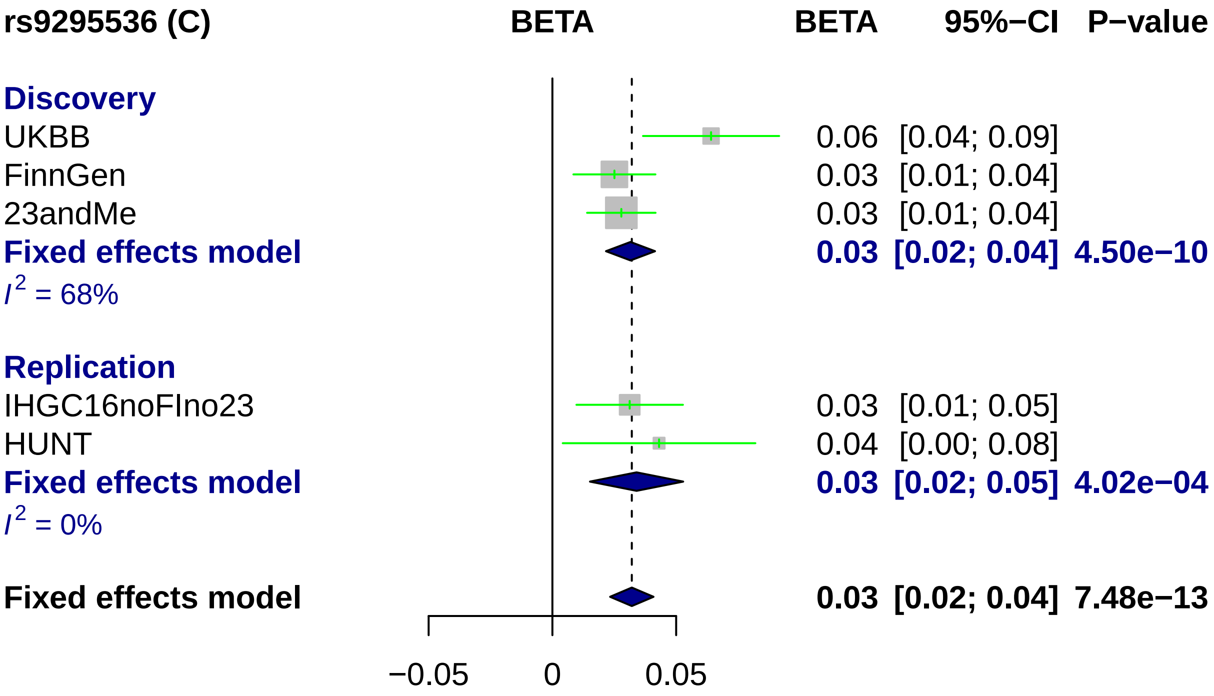
rs9295536 (C)
BETA
BETA 95%−CI P−value
Discovery
UKBB 0.06 [0.04; 0.09]
FinnGen 0.03 [0.01; 0.04]
23andMe 0.03 [0.01; 0.04]
Fixed effects model 0.03 [0.02; 0.04] 4.50e−10
$I^2$ = 68%
Replication
IHGC16noFIno23 0.03 [0.01; 0.05]
HUNT 0.04 [0.00; 0.08]
Fixed effects model 0.03 [0.02; 0.05] 4.02e−04
$I^2$ = 0%
Fixed effects model 0.03 [0.02; 0.04] 7.48e−13
−0.05
0
0.05

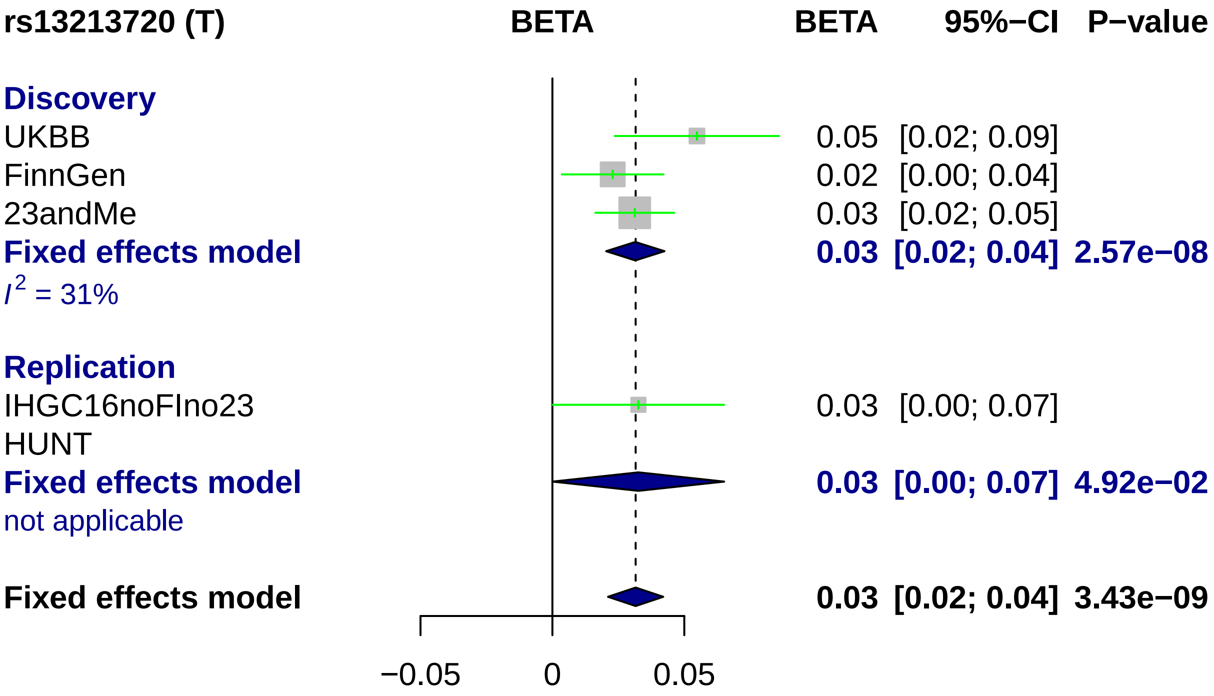

rs13213720 (T)
BETA
BETA
95%−CI
P−value
Discovery
UKBB
0.05 [0.02; 0.09]
FinnGen
0.02 [0.00; 0.04]
23andMe
0.03 [0.02; 0.05]
Fixed effects model
0.03 [0.02; 0.04] 2.57e−08
I2 = 31%
Replication
IHGC16noFIno23
0.03 [0.00; 0.07]
HUNT
Fixed effects model
0.03 [0.00; 0.07] 4.92e−02
not applicable
Fixed effects model
0.03 [0.02; 0.04] 3.43e−09
−0.05
0
0.05

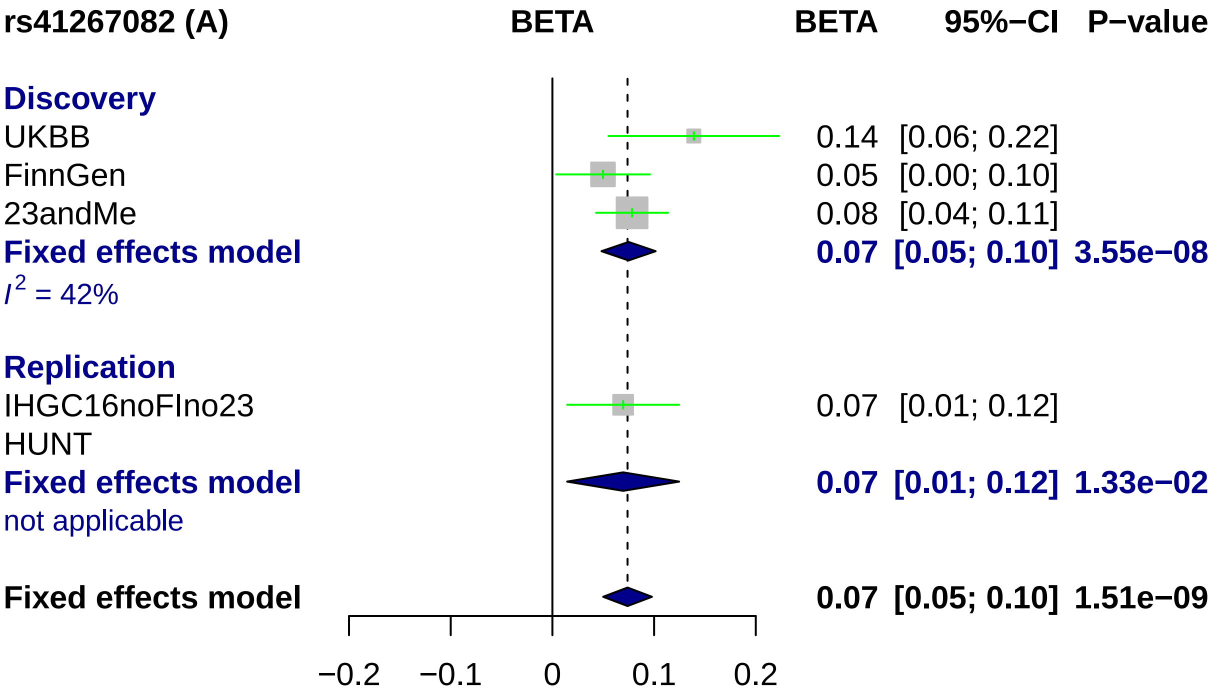
rs41267082 (A)
BETA
BETA
95%−CI
P−value
Discovery
UKBB 0.14 [0.06; 0.22]
FinnGen 0.05 [0.00; 0.10]
23andMe 0.08 [0.04; 0.11]
Fixed effects model 0.07 [0.05; 0.10] 3.55e−08
I2 = 42%
Replication
IHGC16noFIno23 0.07 [0.01; 0.12]
HUNT
Fixed effects model 0.07 [0.01; 0.12] 1.33e−02
not applicable
Fixed effects model 0.07 [0.05; 0.10] 1.51e−09
−0.2
−0.1
0
0.1
0.2

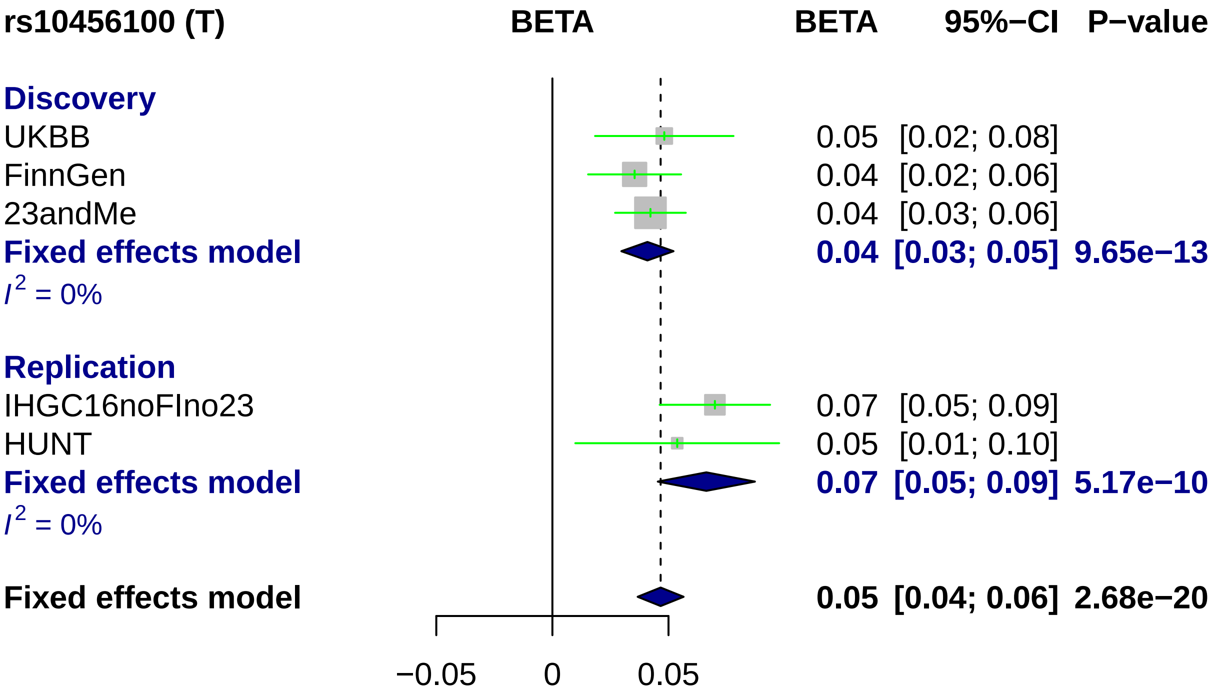
rs10456100 (T)
BETA
BETA
95%−CI
P−value
Discovery
UKBB
0.05
[0.02; 0.08]
FinnGen
0.04
[0.02; 0.06]
23andMe
0.04
[0.03; 0.06]
Fixed effects model
0.04
[0.03; 0.05]
9.65e−13
I2 = 0%
Replication
IHGC16noFIno23
0.07
[0.05; 0.09]
HUNT
0.05
[0.01; 0.10]
Fixed effects model
0.07
[0.05; 0.09]
5.17e−10
I2 = 0%
Fixed effects model
0.05
[0.04; 0.06]
2.68e−20
−0.05
0
0.05

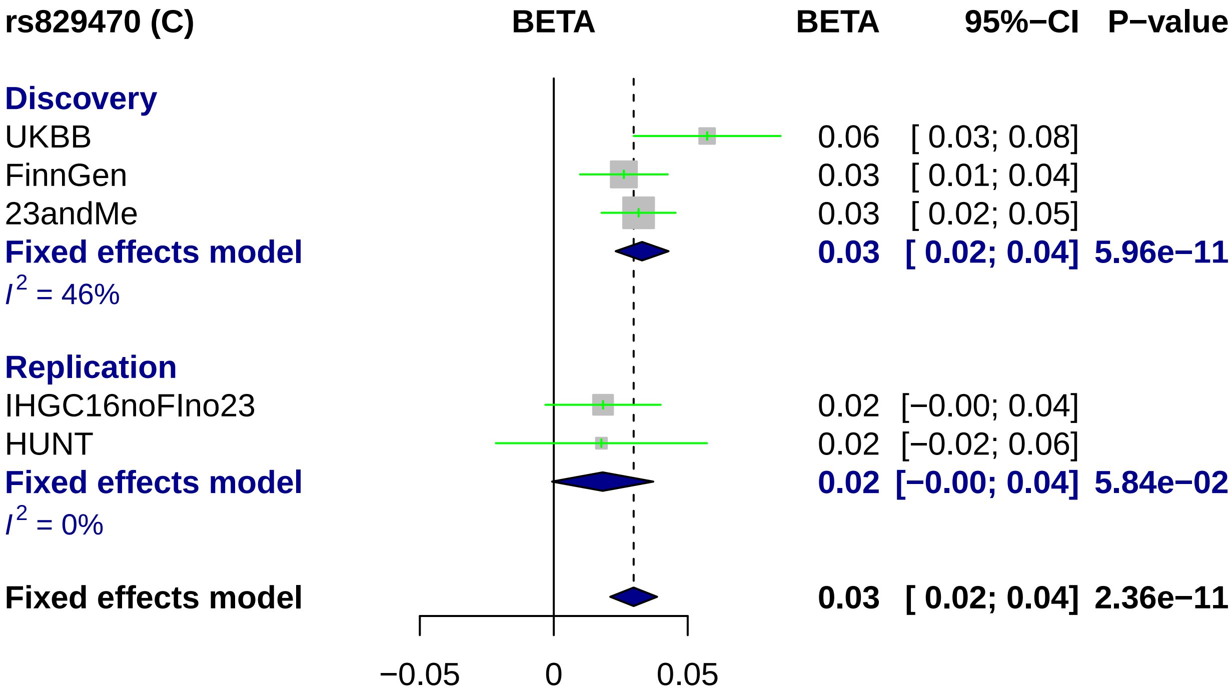
rs829470 (C)
BETA
BETA
95%−CI
P−value
Discovery
UKBB
0.06
[ 0.03; 0.08]
FinnGen
0.03
[ 0.01; 0.04]
23andMe
0.03
[ 0.02; 0.05]
Fixed effects model
0.03
[ 0.02; 0.04]
5.96e−11
I2 = 46%
Replication
IHGC16noFIno23
0.02
[−0.00; 0.04]
HUNT
0.02
[−0.02; 0.06]
Fixed effects model
0.02
[−0.00; 0.04]
5.84e−02
I2 = 0%
Fixed effects model
0.03
[ 0.02; 0.04]
2.36e−11
−0.05
0
0.05

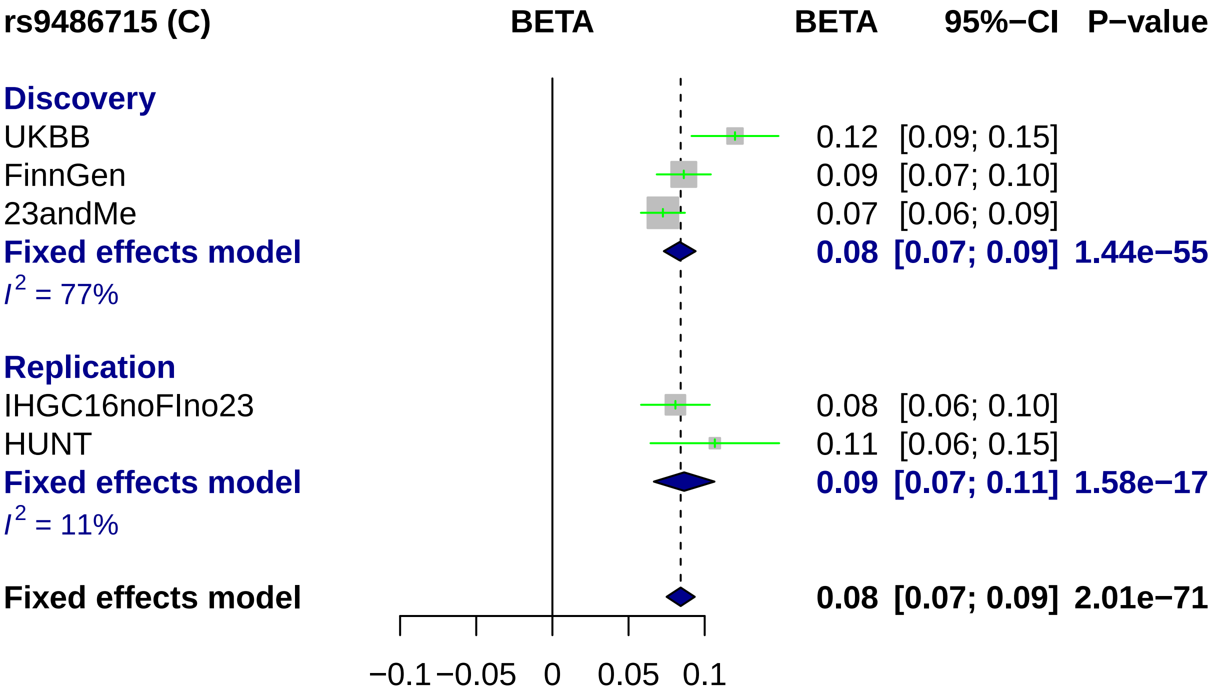
rs9486715 (C)
BETA
BETA 95%−CI P−value
Discovery
UKBB 0.12 [0.09; 0.15]
FinnGen 0.09 [0.07; 0.10]
23andMe 0.07 [0.06; 0.09]
Fixed effects model 0.08 [0.07; 0.09] 1.44e−55
I2 = 77%
Replication
IHGC16noFIno23 0.08 [0.06; 0.10]
HUNT 0.11 [0.06; 0.15]
Fixed effects model 0.09 [0.07; 0.11] 1.58e−17
I2 = 11%
Fixed effects model 0.08 [0.07; 0.09] 2.01e−71
−0.1
−0.05
0
0.05
0.1

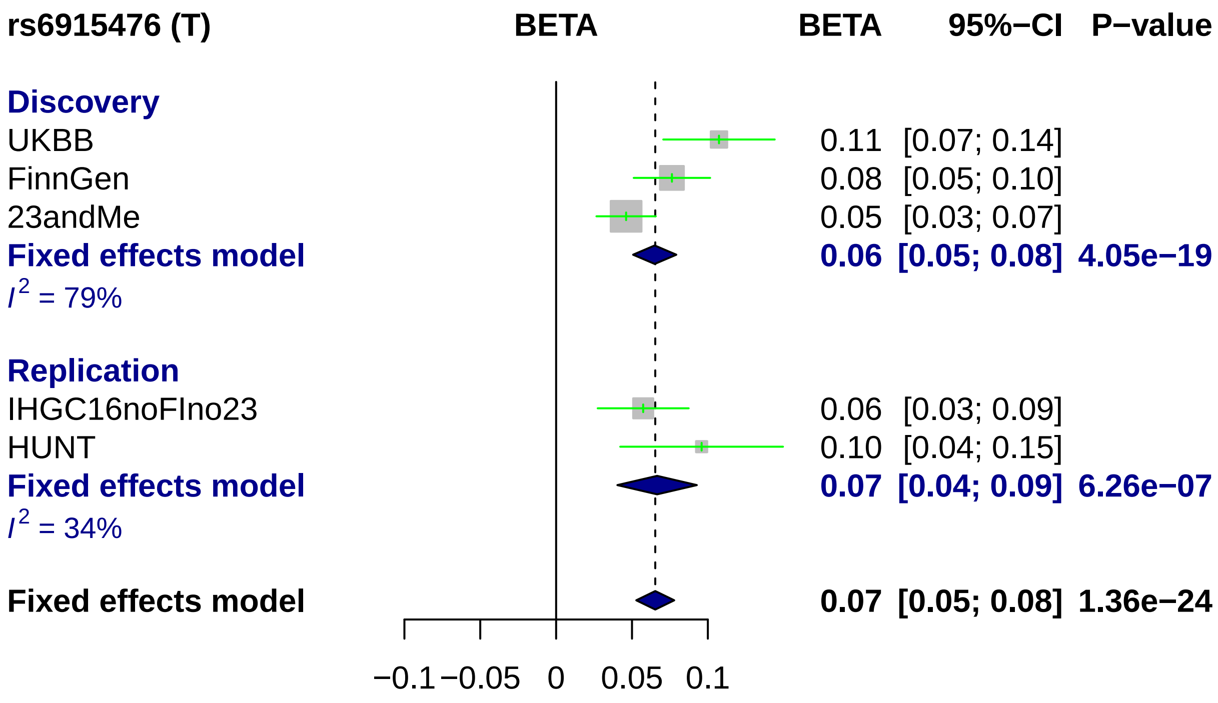
rs6915476 (T)
BETA
BETA
95%−CI
P−value
Discovery
UKBB
0.11 [0.07; 0.14]
FinnGen
0.08 [0.05; 0.10]
23andMe
0.05 [0.03; 0.07]
Fixed effects model
0.06 [0.05; 0.08] 4.05e−19
I2 = 79%
Replication
IHGC16noFIno23
0.06 [0.03; 0.09]
HUNT
0.10 [0.04; 0.15]
Fixed effects model
0.07 [0.04; 0.09] 6.26e−07
I2 = 34%
Fixed effects model
0.07 [0.05; 0.08] 1.36e−24
−0.1
−0.05
0
0.05
0.1

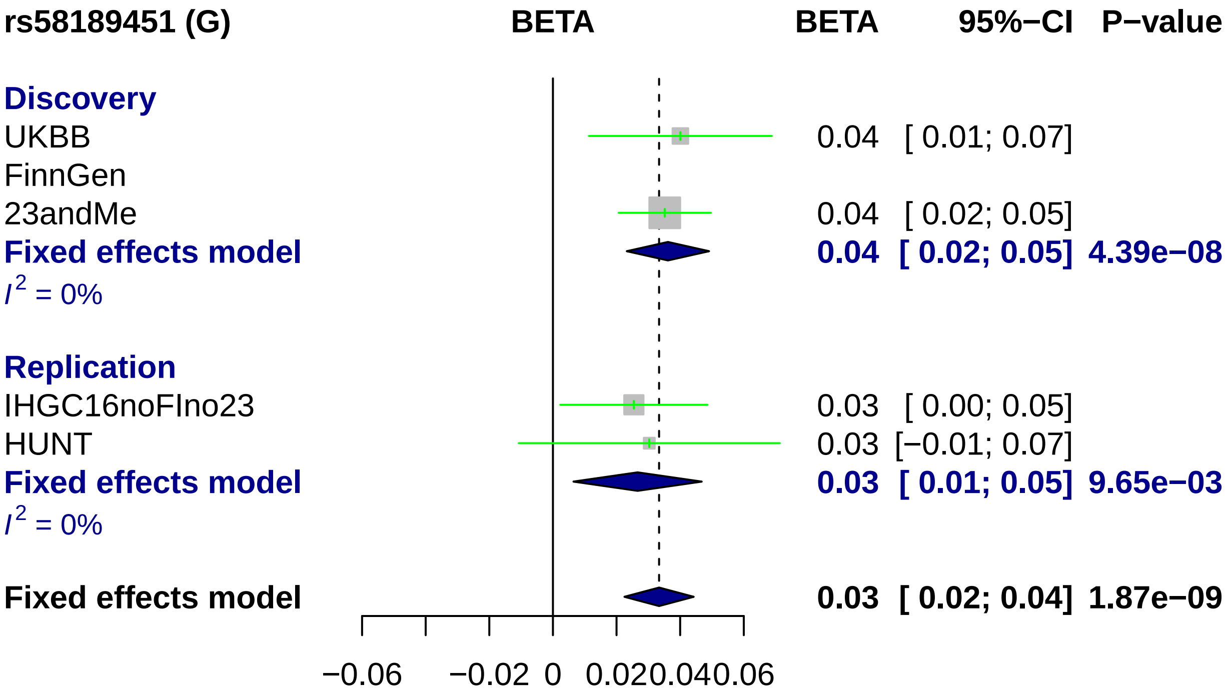
rs58189451 (G)
BETA
BETA
95%−CI
P−value
Discovery
UKBB
0.04 [ 0.01; 0.07]
FinnGen
23andMe
0.04 [ 0.02; 0.05]
Fixed effects model
0.04 [ 0.02; 0.05] 4.39e−08
I2 = 0%
Replication
IHGC16noFIno23
0.03 [ 0.00; 0.05]
HUNT
0.03 [−0.01; 0.07]
Fixed effects model
0.03 [ 0.01; 0.05] 9.65e−03
I2 = 0%
Fixed effects model
0.03 [ 0.02; 0.04] 1.87e−09
−0.06
−0.02
0
0.02
0.04
0.06

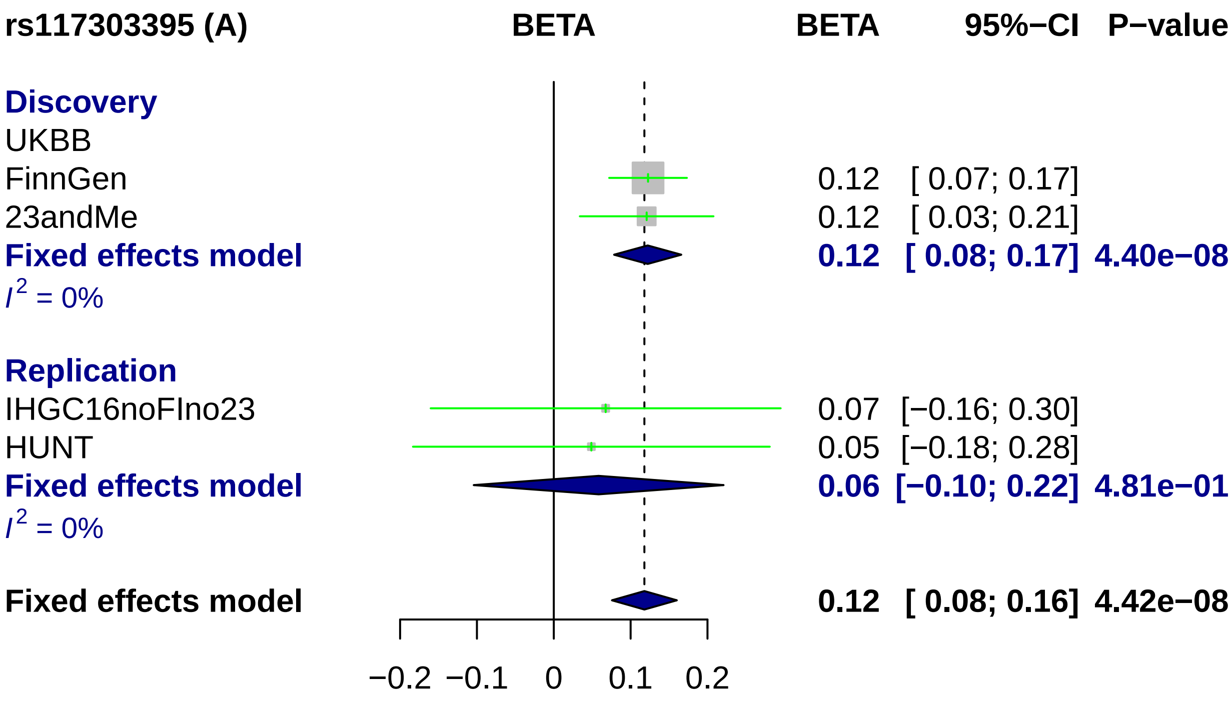
rs117303395 (A)
BETA
BETA
95%−CI
P−value
Discovery
UKBB
FinnGen
0.12 [ 0.07; 0.17]
23andMe
0.12 [ 0.03; 0.21]
Fixed effects model
0.12 [ 0.08; 0.17] 4.40e−08
I2 = 0%
Replication
IHGC16noFIno23
0.07 [−0.16; 0.30]
HUNT
0.05 [−0.18; 0.28]
Fixed effects model
0.06 [−0.10; 0.22] 4.81e−01
I2 = 0%
Fixed effects model
0.12 [ 0.08; 0.16] 4.42e−08
−0.2
−0.1
0
0.1
0.2

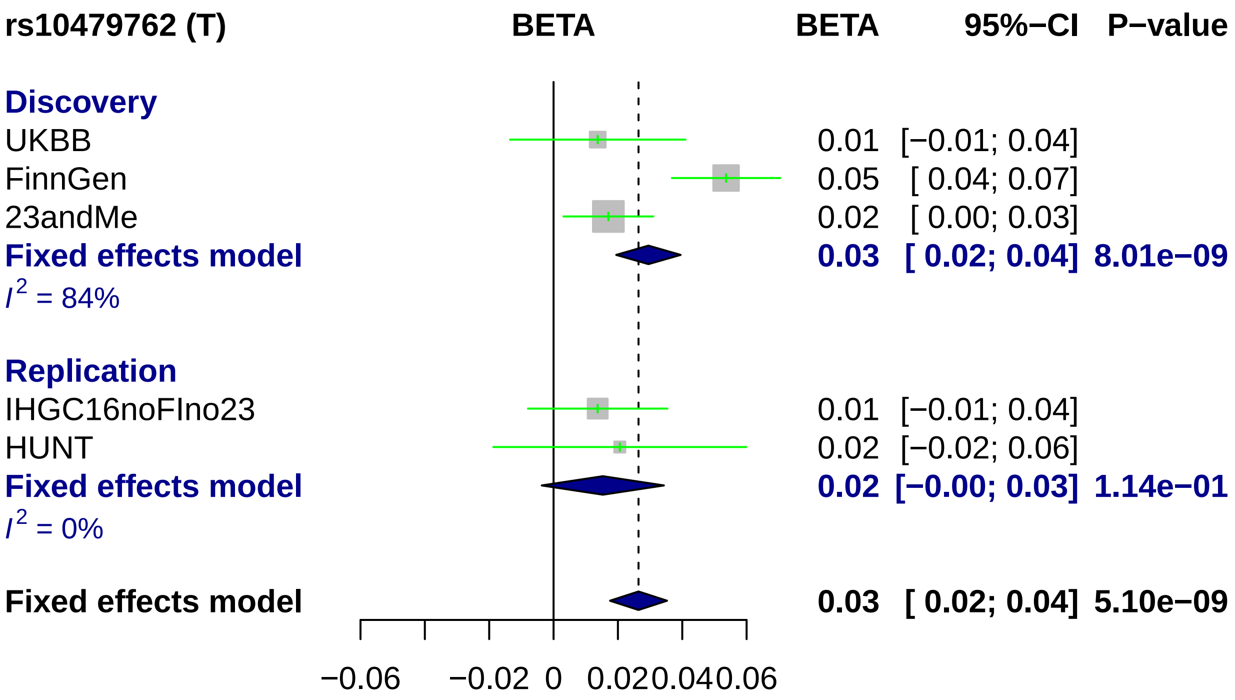
rs10479762 (T)
BETA
BETA 95%−CI P−value
Discovery
UKBB 0.01 [−0.01; 0.04]
FinnGen 0.05 [ 0.04; 0.07]
23andMe 0.02 [ 0.00; 0.03]
Fixed effects model 0.03 [ 0.02; 0.04] 8.01e−09
I2 = 84%
Replication
IHGC16noFIno23 0.01 [−0.01; 0.04]
HUNT 0.02 [−0.02; 0.06]
Fixed effects model 0.02 [−0.00; 0.03] 1.14e−01
I2 = 0%
Fixed effects model 0.03 [ 0.02; 0.04] 5.10e−09
−0.06 −0.02 0 0.02 0.04 0.06

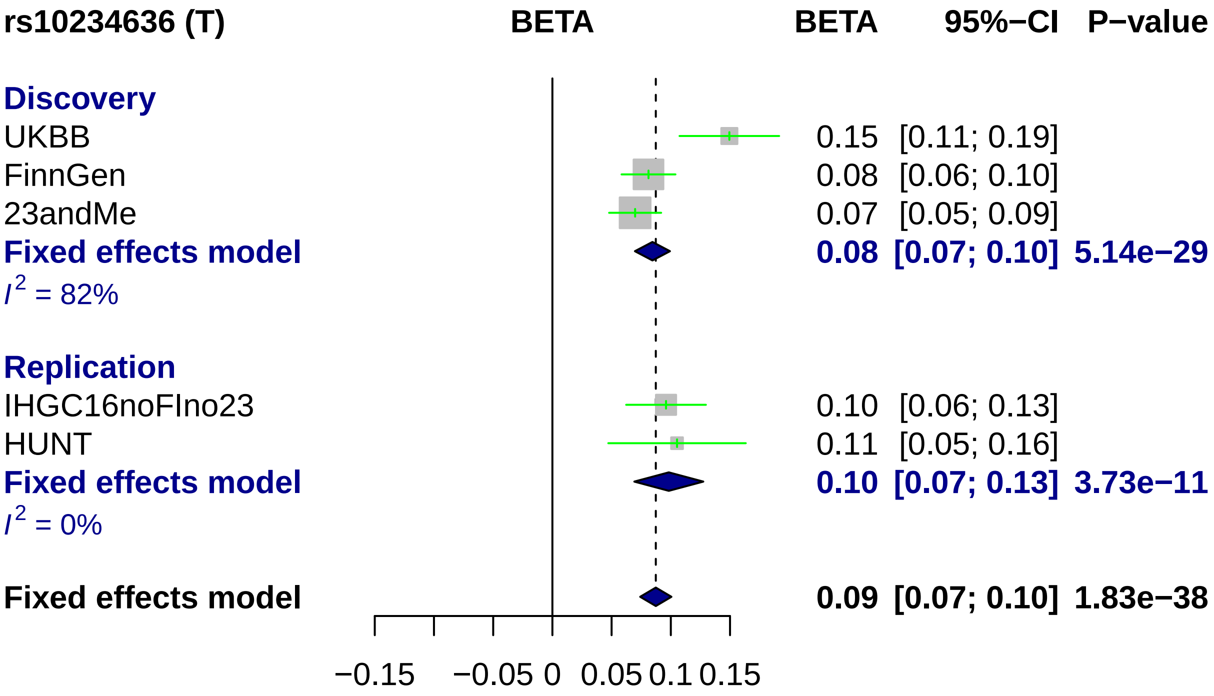

rs10234636 (T)
BETA
BETA 95%−CI P−value
Discovery
UKBB 0.15 [0.11; 0.19]
FinnGen 0.08 [0.06; 0.10]
23andMe 0.07 [0.05; 0.09]
Fixed effects model 0.08 [0.07; 0.10] 5.14e−29
I2 = 82%
Replication
IHGC16noFIno23 0.10 [0.06; 0.13]
HUNT 0.11 [0.05; 0.16]
Fixed effects model 0.10 [0.07; 0.13] 3.73e−11
I2 = 0%
Fixed effects model 0.09 [0.07; 0.10] 1.83e−38
−0.15
−0.05 0
0.05 0.1 0.15

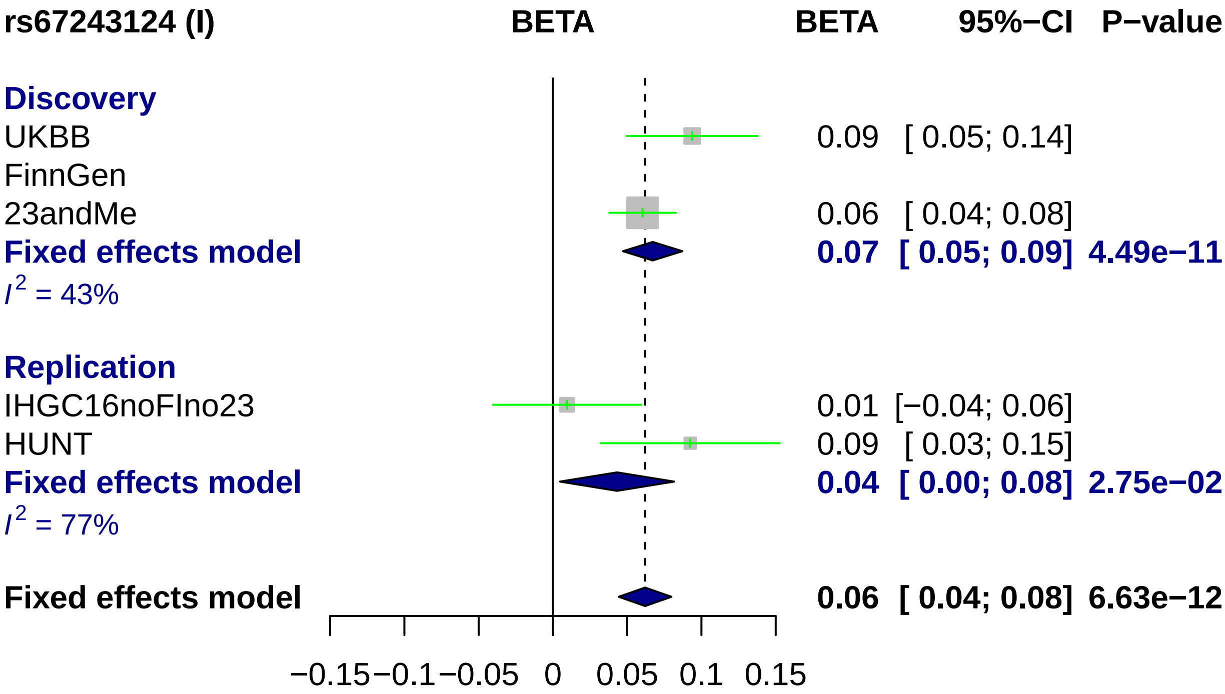
rs67243124 (I)
BETA
BETA
95%−CI
P−value
Discovery
UKBB
0.09 [ 0.05; 0.14]
FinnGen
23andMe
0.06 [ 0.04; 0.08]
Fixed effects model
0.07 [ 0.05; 0.09] 4.49e−11
I2 = 43%
Replication
IHGC16noFIno23
0.01 [−0.04; 0.06]
HUNT
0.09 [ 0.03; 0.15]
Fixed effects model
0.04 [ 0.00; 0.08] 2.75e−02
I2 = 77%
Fixed effects model
0.06 [ 0.04; 0.08] 6.63e−12
−0.15
−0.1
−0.05
0
0.05
0.1
0.15

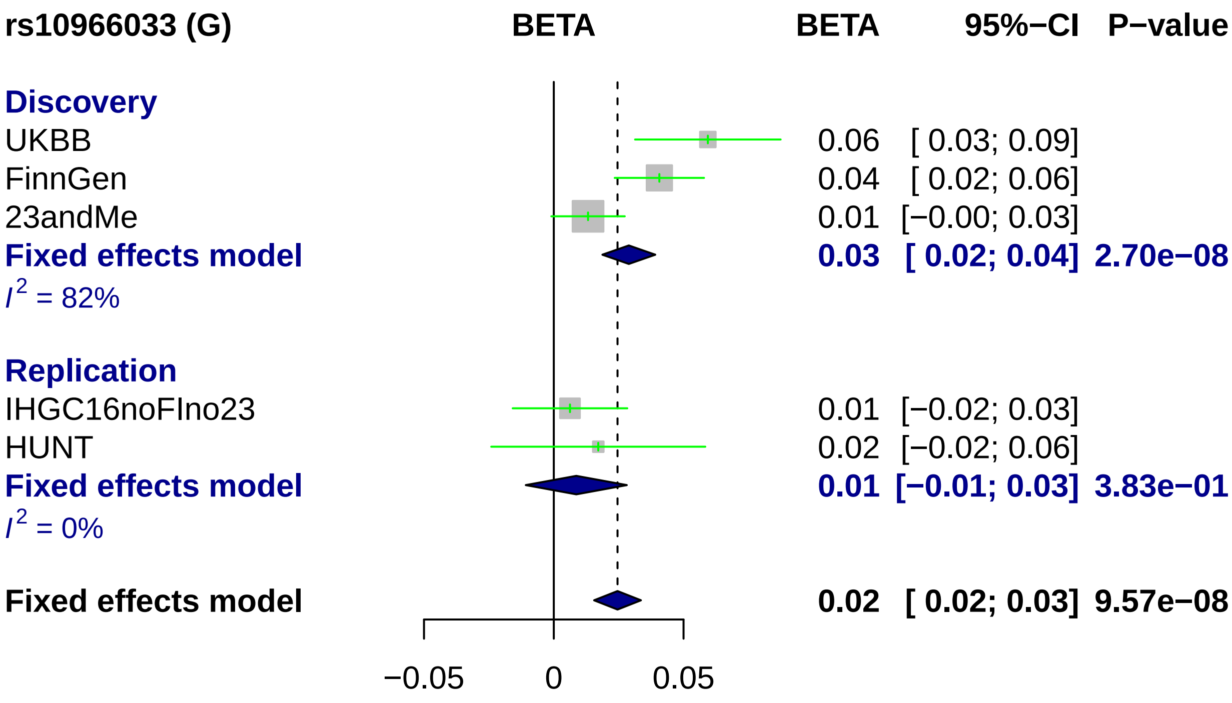
rs10966033 (G)
BETA
BETA
95%−CI
P−value
Discovery
UKBB
0.06
[ 0.03; 0.09]
FinnGen
0.04
[ 0.02; 0.06]
23andMe
0.01
[−0.00; 0.03]
Fixed effects model
0.03
[ 0.02; 0.04]
2.70e−08
I2 = 82%
Replication
IHGC16noFIno23
0.01
[−0.02; 0.03]
HUNT
0.02
[−0.02; 0.06]
Fixed effects model
0.01
[−0.01; 0.03]
3.83e−01
I2 = 0%
Fixed effects model
0.02
[ 0.02; 0.03]
9.57e−08
−0.05
0
0.05

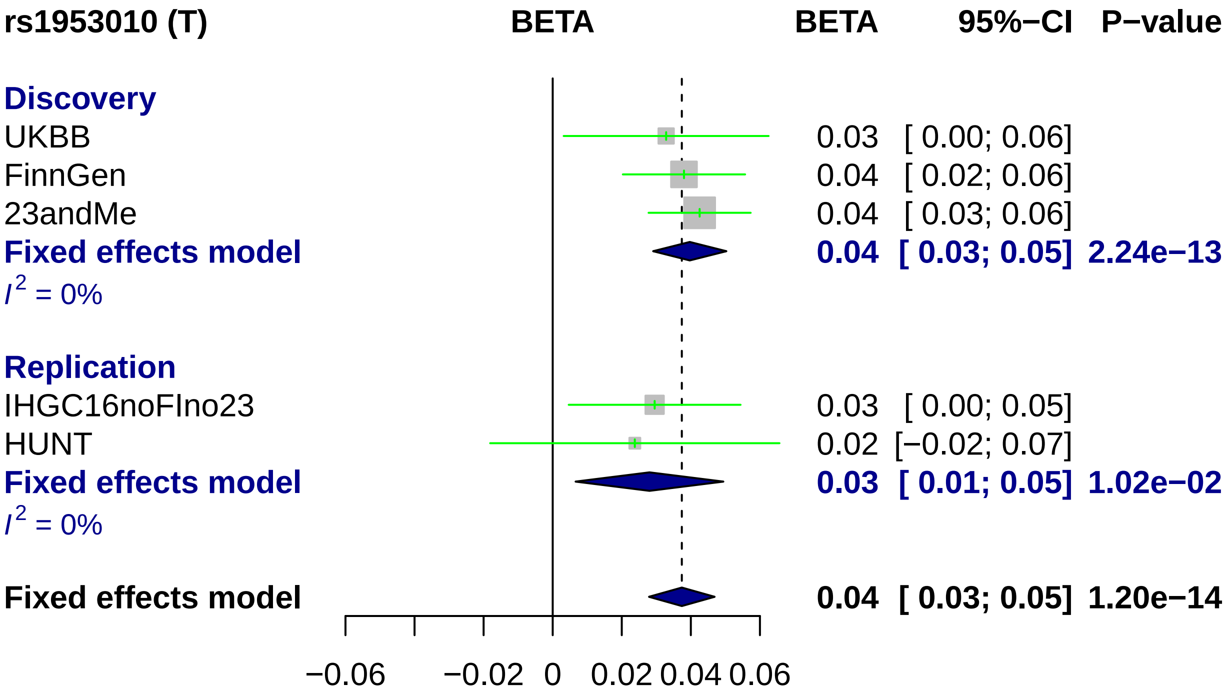
rs1953010 (T)
BETA
BETA
95%−CI
P−value
Discovery
UKBB
0.03 [ 0.00; 0.06]
FinnGen
0.04 [ 0.02; 0.06]
23andMe
0.04 [ 0.03; 0.06]
Fixed effects model
0.04 [ 0.03; 0.05] 2.24e−13
I2 = 0%
Replication
IHGC16noFIno23
0.03 [ 0.00; 0.05]
HUNT
0.02 [−0.02; 0.07]
Fixed effects model
0.03 [ 0.01; 0.05] 1.02e−02
I2 = 0%
Fixed effects model
0.04 [ 0.03; 0.05] 1.20e−14
−0.06
−0.02
0
0.02
0.04
0.06

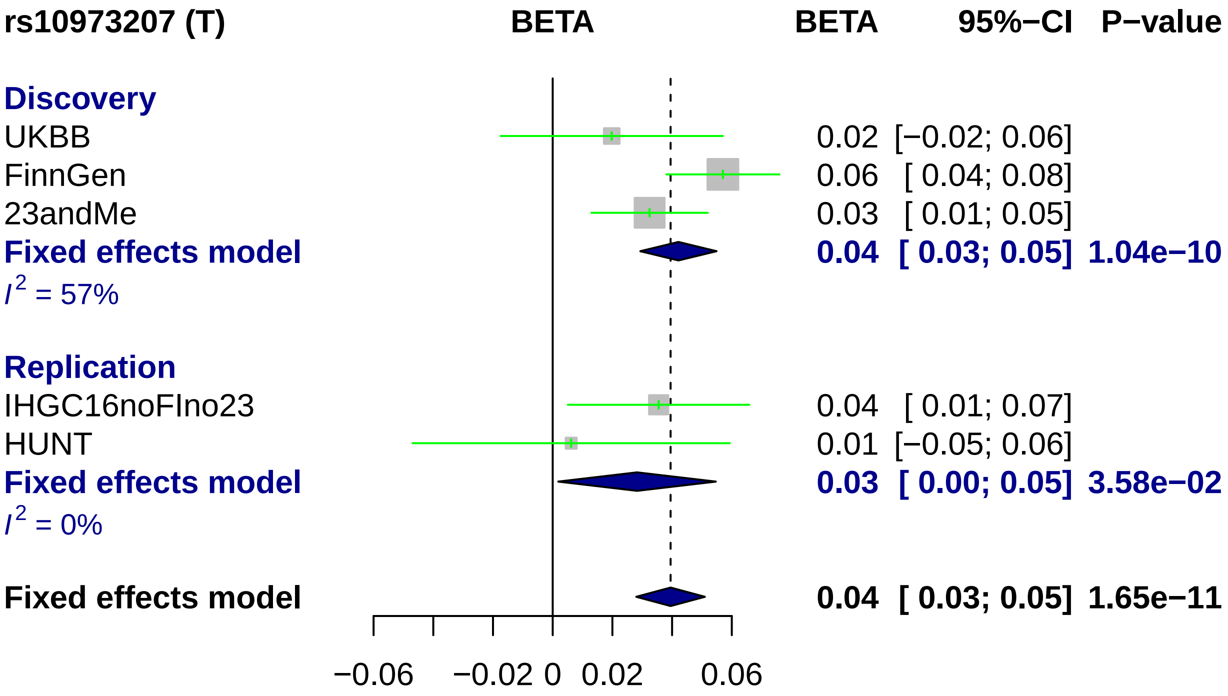

rs10973207 (T)
BETA
BETA
95%−CI
P−value
Discovery
UKBB
0.02 [−0.02; 0.06]
FinnGen
0.06 [ 0.04; 0.08]
23andMe
0.03 [ 0.01; 0.05]
Fixed effects model
0.04 [ 0.03; 0.05] 1.04e−10
I^2 = 57%
Replication
IHGC16noFIno23
0.04 [ 0.01; 0.07]
HUNT
0.01 [−0.05; 0.06]
Fixed effects model
0.03 [ 0.00; 0.05] 3.58e−02
I^2 = 0%
Fixed effects model
0.04 [ 0.03; 0.05] 1.65e−11
−0.06
−0.02
0
0.02
0.06

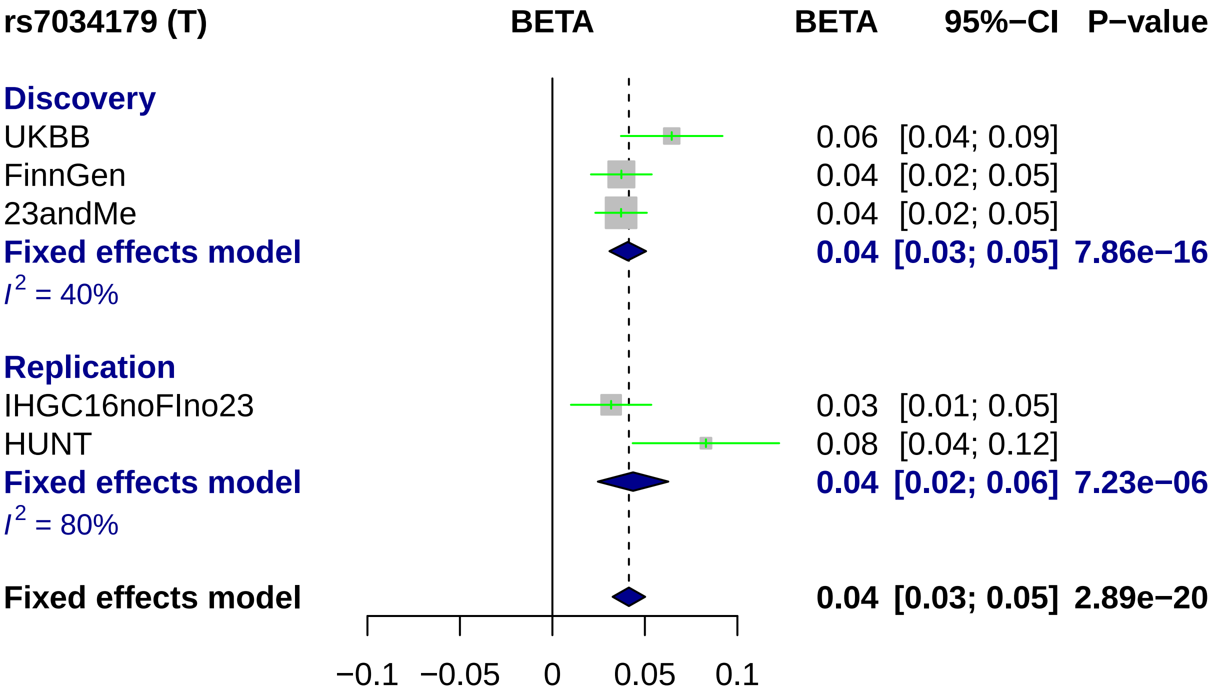
rs7034179 (T)
BETA
BETA
95%−CI
P−value
Discovery
UKBB
0.06 [0.04; 0.09]
FinnGen
0.04 [0.02; 0.05]
23andMe
0.04 [0.02; 0.05]
Fixed effects model
0.04 [0.03; 0.05] 7.86e−16
I2 = 40%
Replication
IHGC16noFIno23
0.03 [0.01; 0.05]
HUNT
0.08 [0.04; 0.12]
Fixed effects model
0.04 [0.02; 0.06] 7.23e−06
I2 = 80%
Fixed effects model
0.04 [0.03; 0.05] 2.89e−20
−0.1
−0.05
0
0.05
0.1

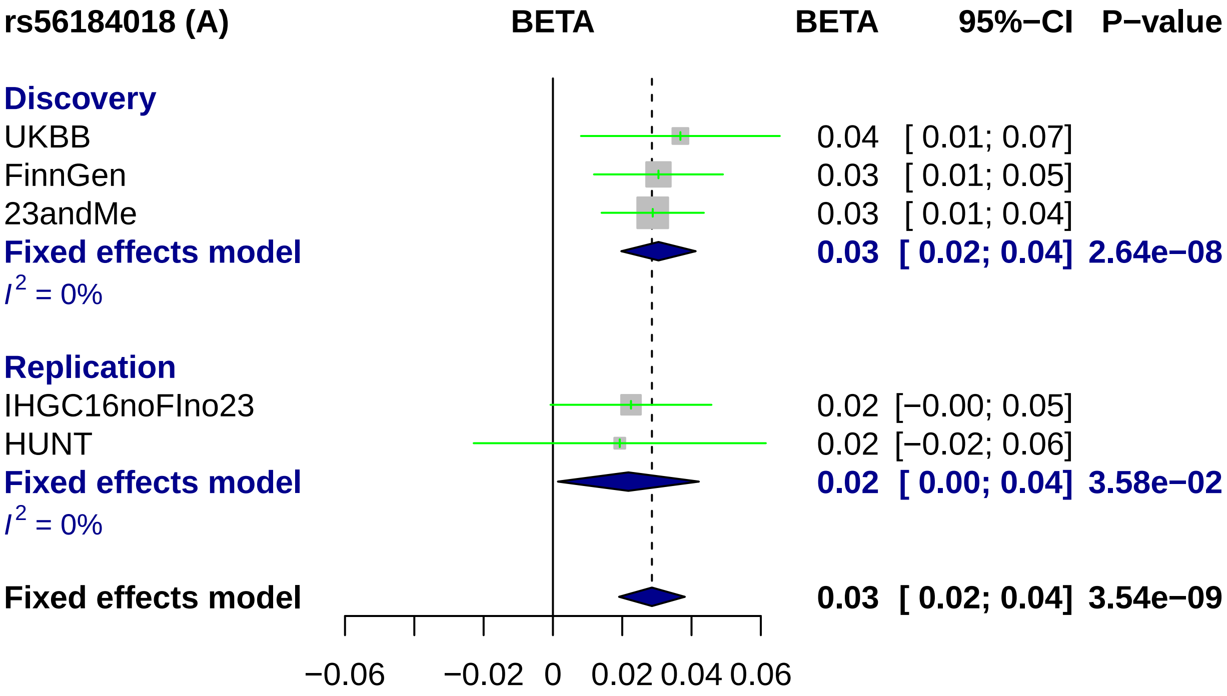
rs56184018 (A)
BETA
BETA
95%−CI
P−value
Discovery
UKBB
0.04 [ 0.01; 0.07]
FinnGen
0.03 [ 0.01; 0.05]
23andMe
0.03 [ 0.01; 0.04]
Fixed effects model
0.03 [ 0.02; 0.04] 2.64e−08
I 2 = 0%
Replication
IHGC16noFIno23
0.02 [−0.00; 0.05]
HUNT
0.02 [−0.02; 0.06]
Fixed effects model
0.02 [ 0.00; 0.04] 3.58e−02
I 2 = 0%
Fixed effects model
0.03 [ 0.02; 0.04] 3.54e−09
−0.06
−0.02
0
0.02
0.04
0.06

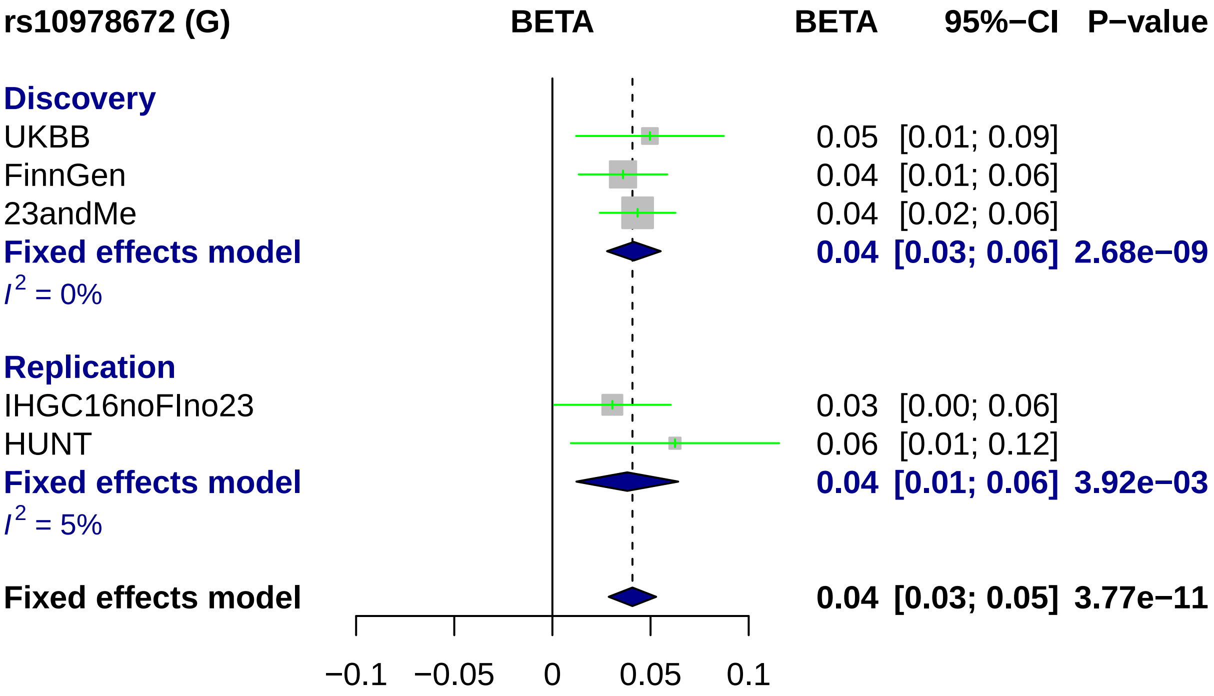
rs10978672 (G)
BETA
BETA
95%−CI
P−value
Discovery
UKBB
FinnGen
23andMe
Fixed effects model
I2 = 0%
Replication
IHGC16noFIno23
HUNT
Fixed effects model
I2 = 5%
Fixed effects model
0.05 [0.01; 0.09]
0.04 [0.01; 0.06]
0.04 [0.02; 0.06]
0.04 [0.03; 0.06] 2.68e−09
0.03 [0.00; 0.06]
0.06 [0.01; 0.12]
0.04 [0.01; 0.06] 3.92e−03
0.04 [0.03; 0.05] 3.77e−11
−0.1
−0.05
0
0.05
0.1

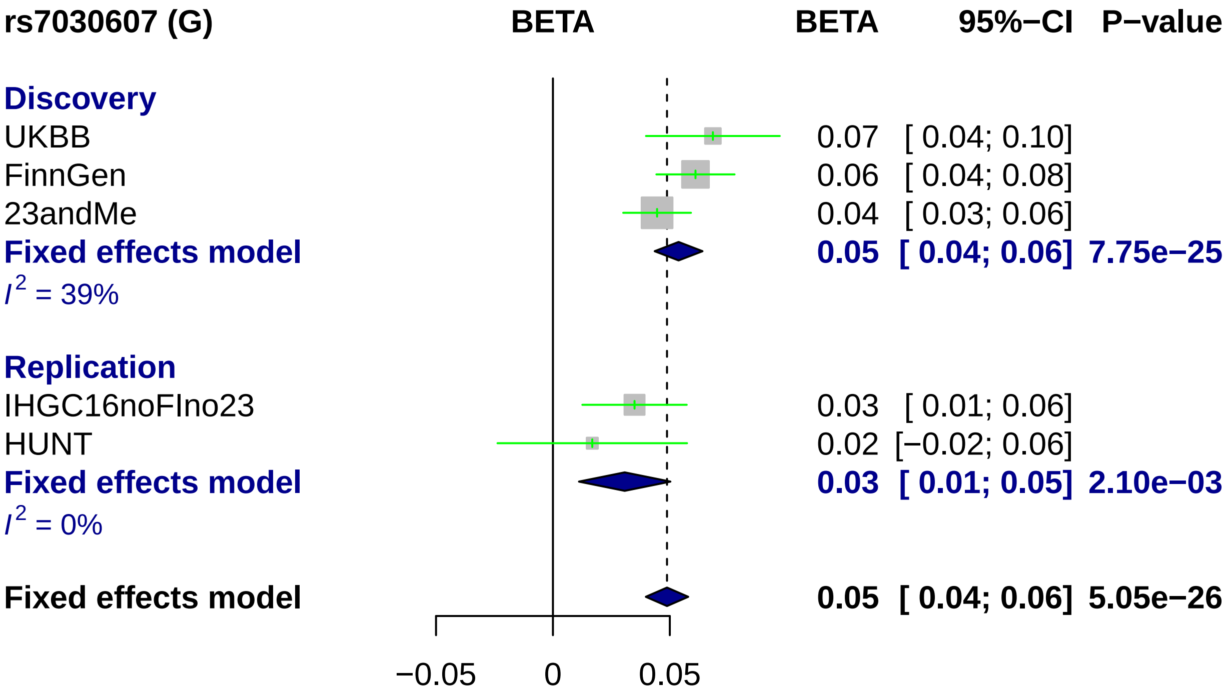
rs7030607 (G)
BETA
BETA
95%−CI
P−value
Discovery
UKBB
0.07 [ 0.04; 0.10]
FinnGen
0.06 [ 0.04; 0.08]
23andMe
0.04 [ 0.03; 0.06]
Fixed effects model
0.05 [ 0.04; 0.06] 7.75e−25
I^2 = 39%
Replication
IHGC16noFIno23
0.03 [ 0.01; 0.06]
HUNT
0.02 [−0.02; 0.06]
Fixed effects model
0.03 [ 0.01; 0.05] 2.10e−03
I^2 = 0%
Fixed effects model
0.05 [ 0.04; 0.06] 5.05e−26
−0.05
0
0.05

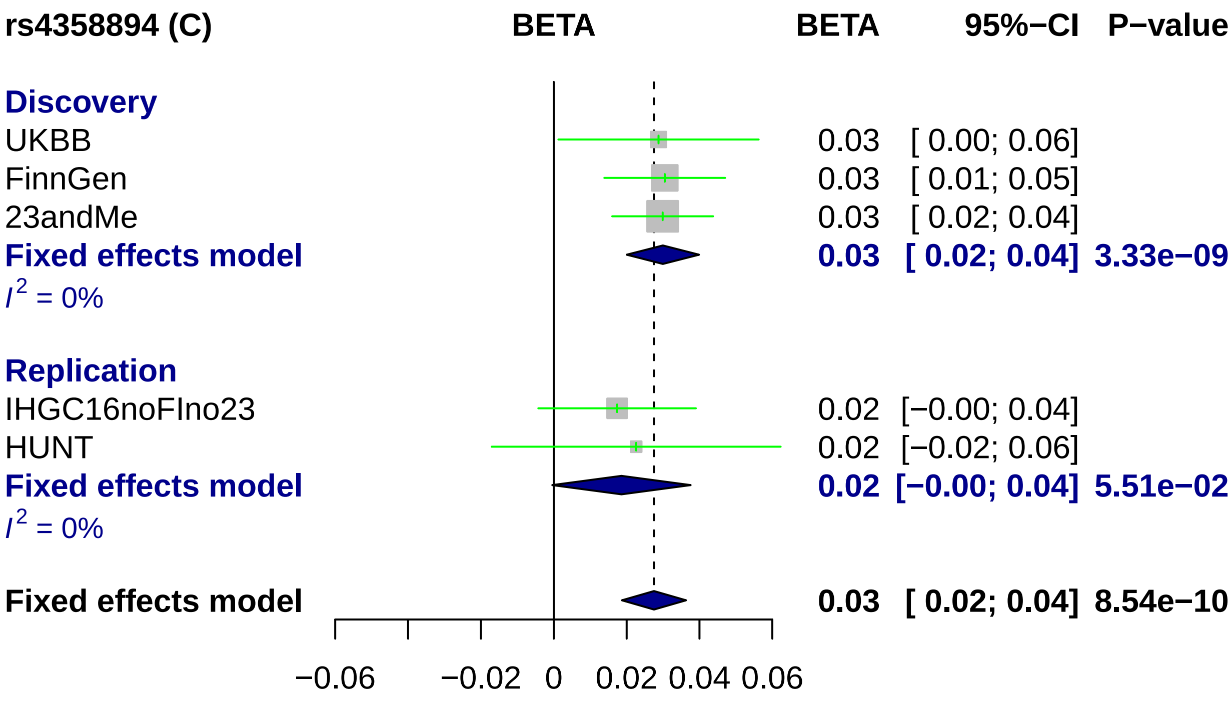
rs4358894 (C)
BETA
BETA
95%−CI
P−value
Discovery
UKBB
0.03 [ 0.00; 0.06]
FinnGen
0.03 [ 0.01; 0.05]
23andMe
0.03 [ 0.02; 0.04]
Fixed effects model
0.03 [ 0.02; 0.04] 3.33e−09
$I^2$ = 0%
Replication
IHGC16noFIno23
0.02 [−0.00; 0.04]
HUNT
0.02 [−0.02; 0.06]
Fixed effects model
0.02 [−0.00; 0.04] 5.51e−02
$I^2$ = 0%
Fixed effects model
0.03 [ 0.02; 0.04] 8.54e−10
−0.06
−0.02
0
0.02
0.04
0.06

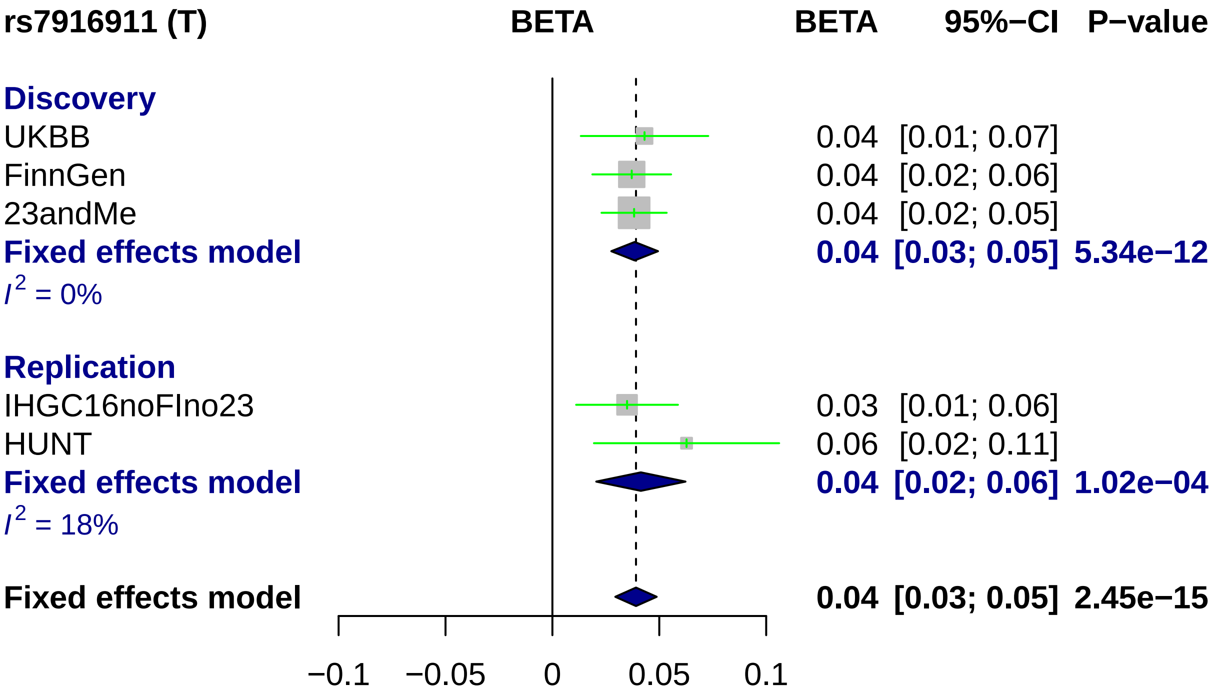

rs7916911 (T)
BETA
BETA
95%−CI
P−value
Discovery
UKBB
0.04 [0.01; 0.07]
FinnGen
0.04 [0.02; 0.06]
23andMe
0.04 [0.02; 0.05]
Fixed effects model
0.04 [0.03; 0.05] 5.34e−12
I2 = 0%
Replication
IHGC16noFIno23
0.03 [0.01; 0.06]
HUNT
0.06 [0.02; 0.11]
Fixed effects model
0.04 [0.02; 0.06] 1.02e−04
I2 = 18%
Fixed effects model
0.04 [0.03; 0.05] 2.45e−15
−0.1
−0.05
0
0.05
0.1

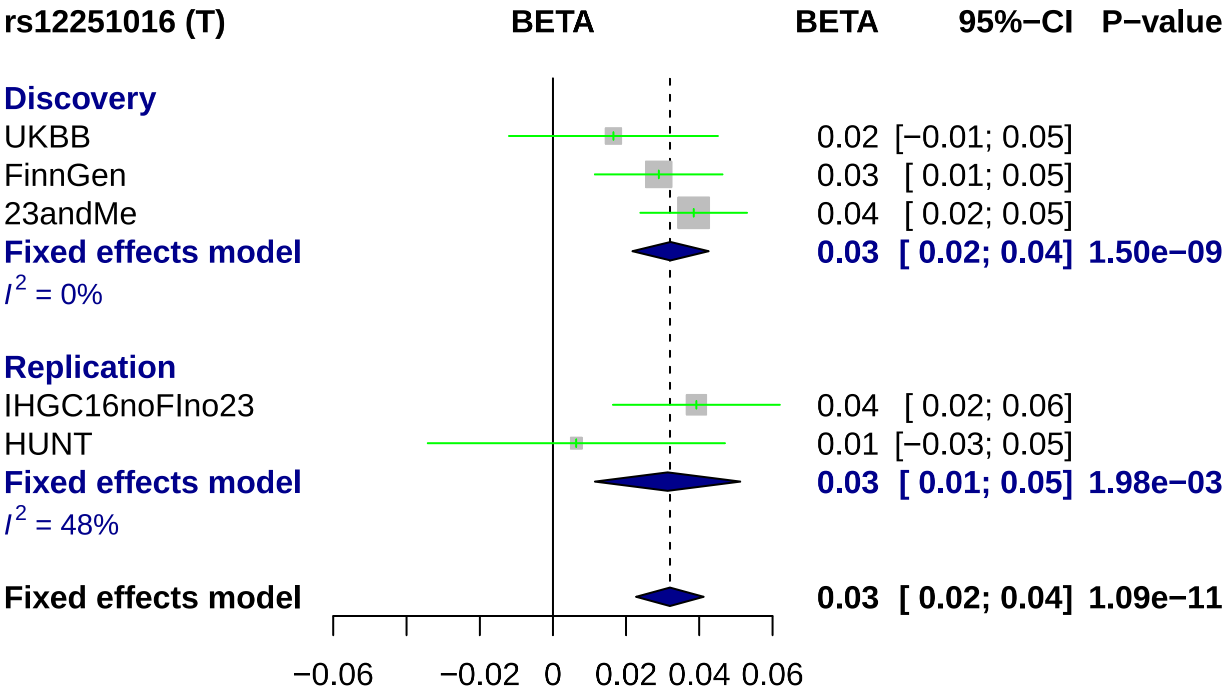
rs12251016 (T)
BETA
BETA
95%−CI
P−value
Discovery
UKBB
0.02 [−0.01; 0.05]
FinnGen
0.03 [ 0.01; 0.05]
23andMe
0.04 [ 0.02; 0.05]
Fixed effects model
0.03 [ 0.02; 0.04] 1.50e−09
I2 = 0%
Replication
IHGC16noFIno23
0.04 [ 0.02; 0.06]
HUNT
0.01 [−0.03; 0.05]
Fixed effects model
0.03 [ 0.01; 0.05] 1.98e−03
I2 = 48%
Fixed effects model
0.03 [ 0.02; 0.04] 1.09e−11
−0.06
−0.02
0
0.02
0.04
0.06

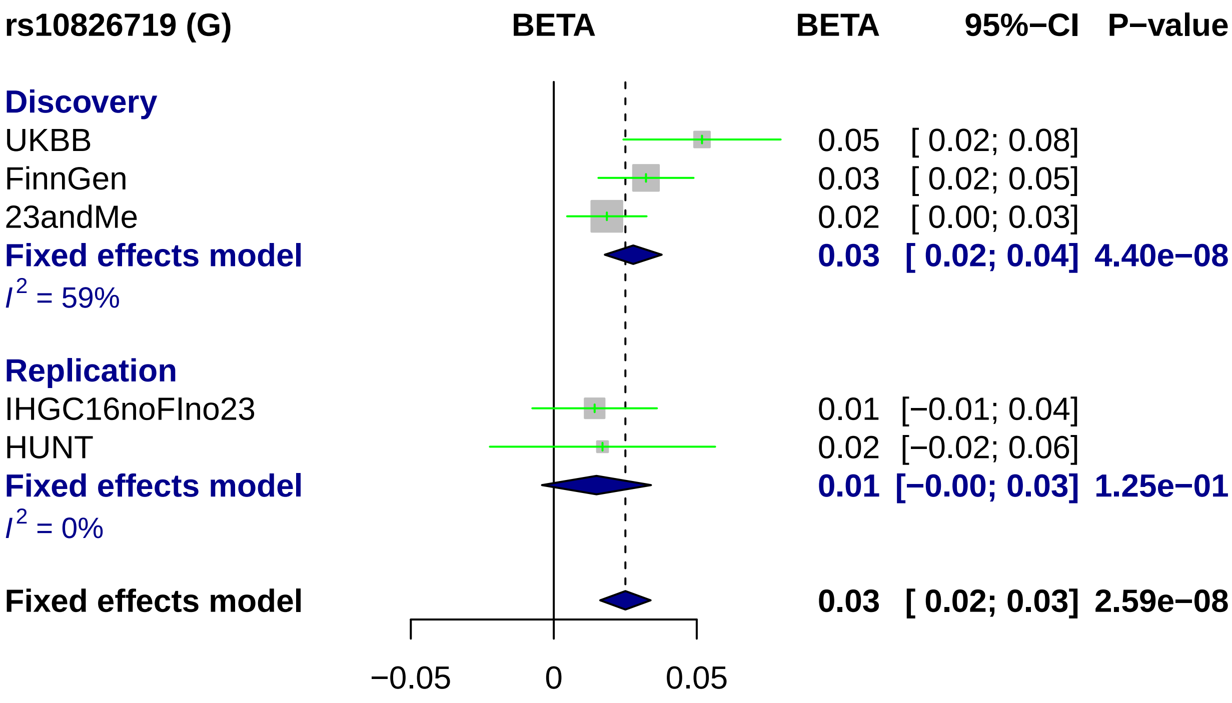
rs10826719 (G)
BETA
BETA 95%−CI P−value
Discovery
UKBB 0.05 [ 0.02; 0.08]
FinnGen 0.03 [ 0.02; 0.05]
23andMe 0.02 [ 0.00; 0.03]
Fixed effects model 0.03 [ 0.02; 0.04] 4.40e−08
I2 = 59%
Replication
IHGC16noFIno23 0.01 [−0.01; 0.04]
HUNT 0.02 [−0.02; 0.06]
Fixed effects model 0.01 [−0.00; 0.03] 1.25e−01
I2 = 0%
Fixed effects model 0.03 [ 0.02; 0.03] 2.59e−08
−0.05
0
0.05

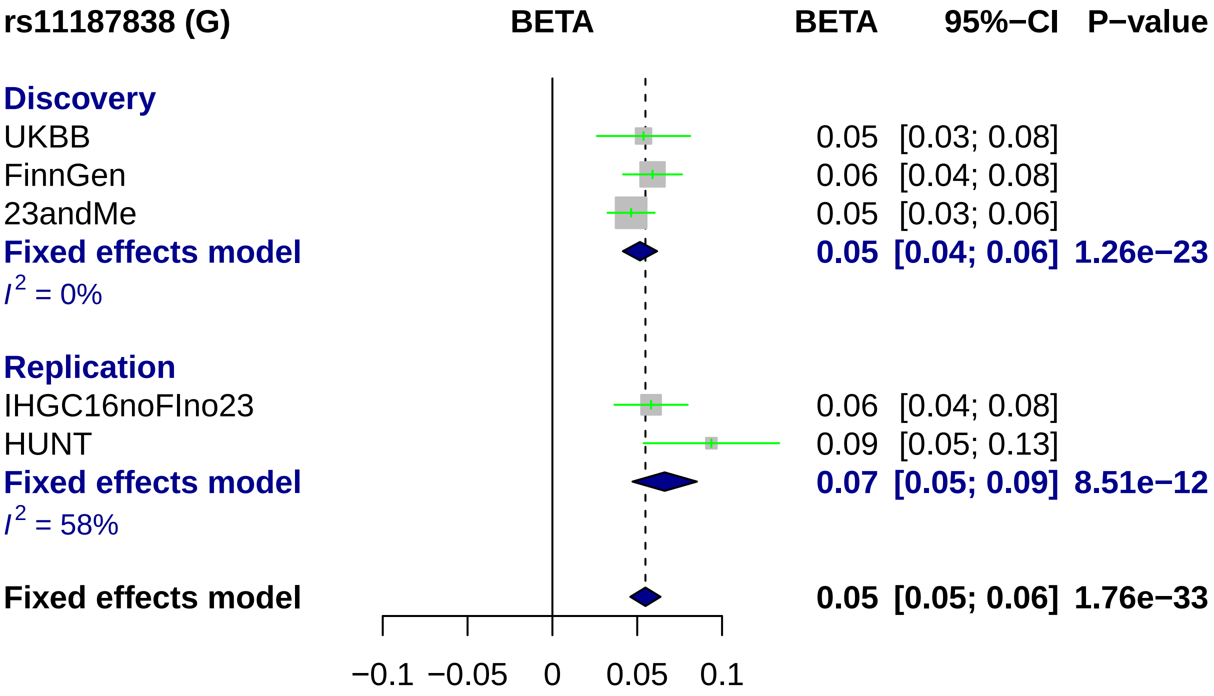
rs11187838 (G)
BETA
BETA
95%−CI
P−value
Discovery
UKBB
0.05 [0.03; 0.08]
FinnGen
0.06 [0.04; 0.08]
23andMe
0.05 [0.03; 0.06]
Fixed effects model
0.05 [0.04; 0.06] 1.26e−23
I2 = 0%
Replication
IHGC16noFIno23
0.06 [0.04; 0.08]
HUNT
0.09 [0.05; 0.13]
Fixed effects model
0.07 [0.05; 0.09] 8.51e−12
I2 = 58%
Fixed effects model
0.05 [0.05; 0.06] 1.76e−33
−0.1
−0.05
0
0.05
0.1

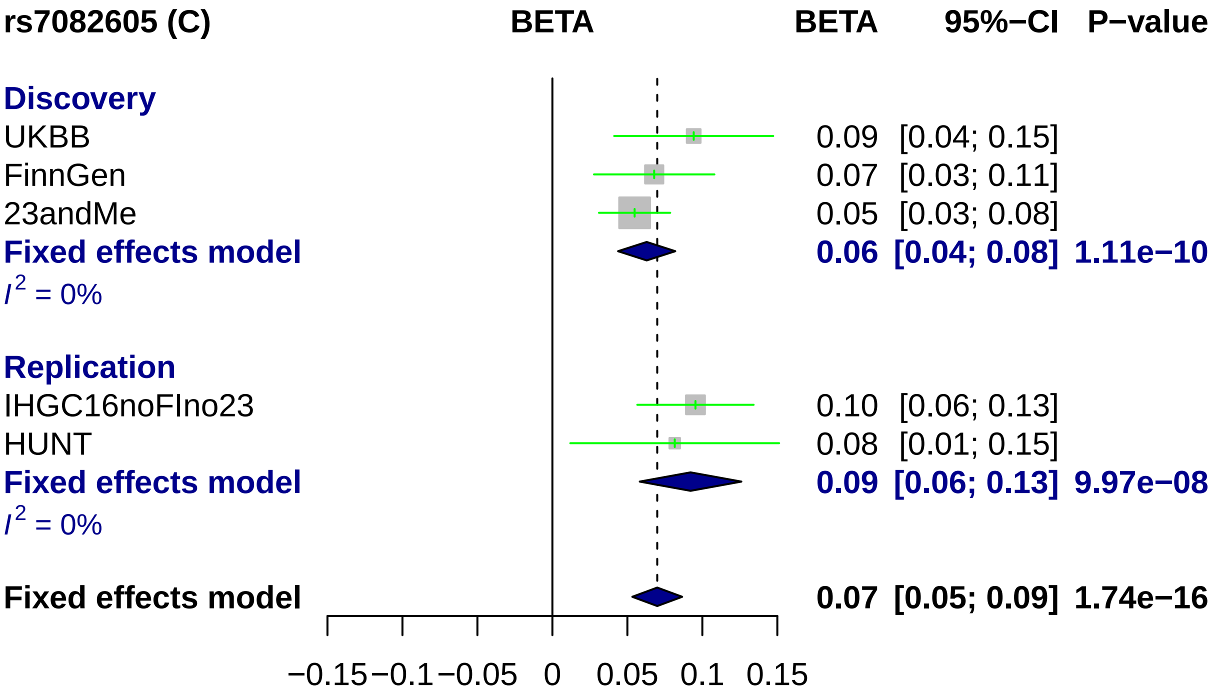

rs7082605 (C)
BETA
BETA
95%−CI
P−value
Discovery
UKBB 0.09 [0.04; 0.15]
FinnGen 0.07 [0.03; 0.11]
23andMe 0.05 [0.03; 0.08]
Fixed effects model 0.06 [0.04; 0.08] 1.11e−10
I2 = 0%
Replication
IHGC16noFIno23 0.10 [0.06; 0.13]
HUNT 0.08 [0.01; 0.15]
Fixed effects model 0.09 [0.06; 0.13] 9.97e−08
I2 = 0%
Fixed effects model 0.07 [0.05; 0.09] 1.74e−16
−0.15
−0.1
−0.05
0
0.05
0.1
0.15

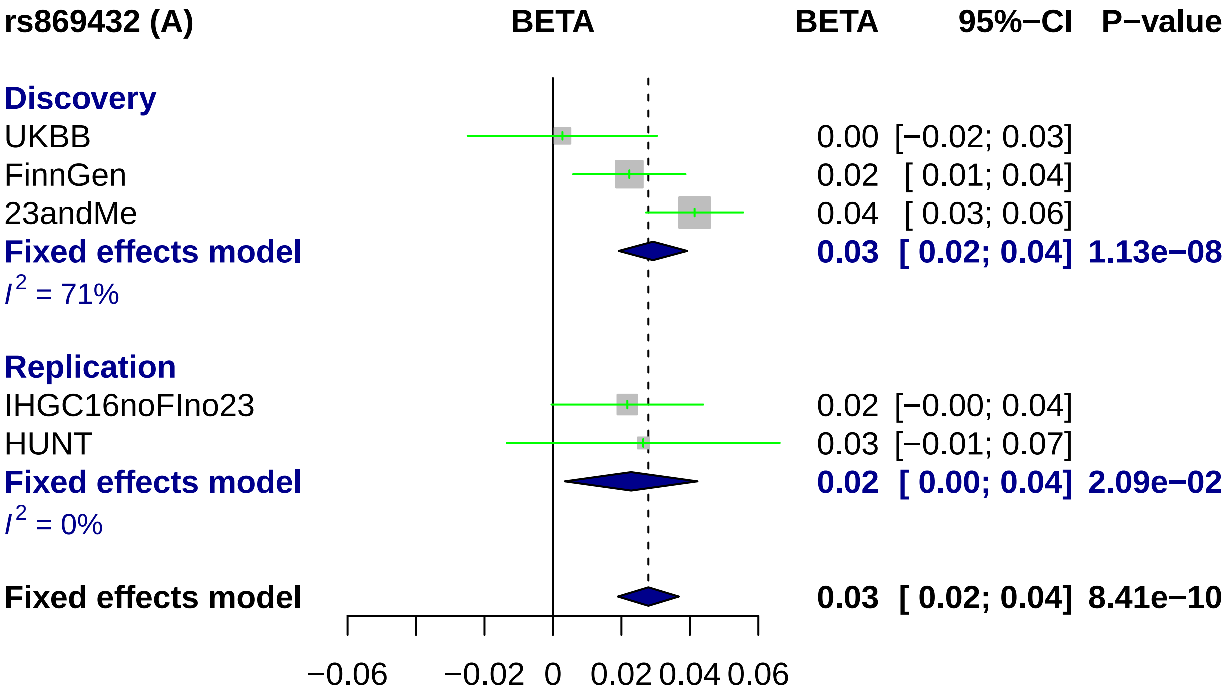
rs869432 (A)
BETA
BETA
95%−CI
P−value
Discovery
UKBB
0.00 [−0.02; 0.03]
FinnGen
0.02 [ 0.01; 0.04]
23andMe
0.04 [ 0.03; 0.06]
Fixed effects model
0.03 [ 0.02; 0.04] 1.13e−08
$I^2$ = 71%
Replication
IHGC16noFIno23
0.02 [−0.00; 0.04]
HUNT
0.03 [−0.01; 0.07]
Fixed effects model
0.02 [ 0.00; 0.04] 2.09e−02
$I^2$ = 0%
Fixed effects model
0.03 [ 0.02; 0.04] 8.41e−10
−0.06
−0.02
0
0.02
0.04
0.06

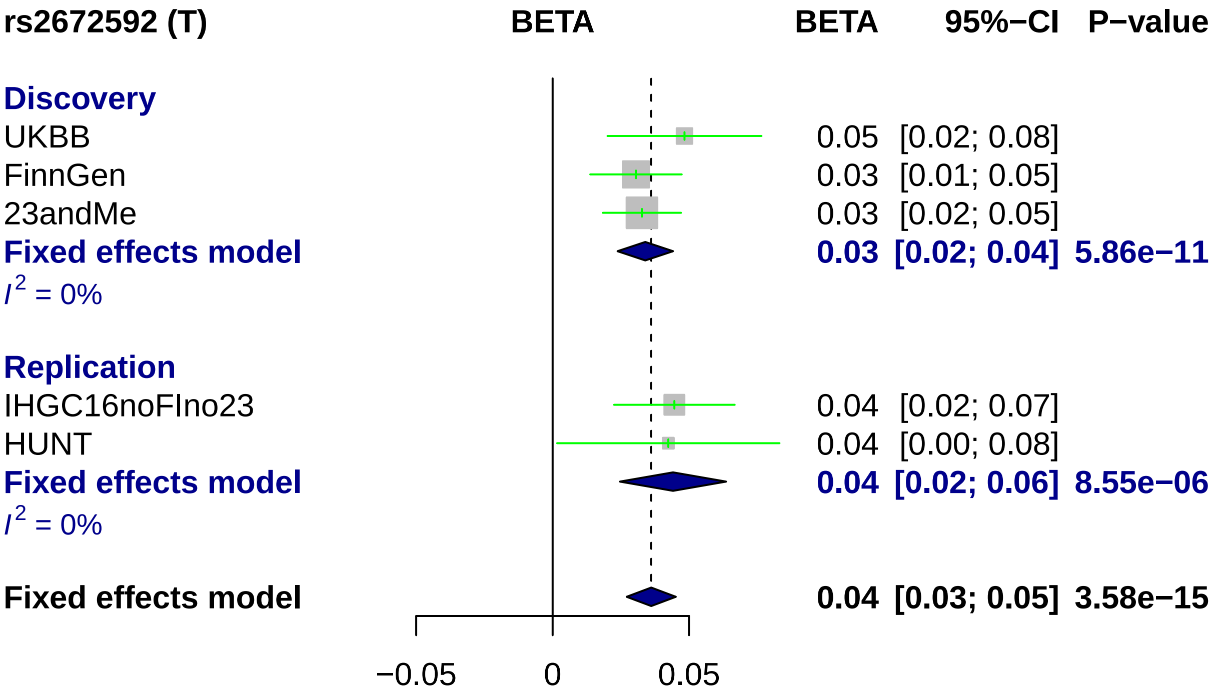

rs2672592 (T)
BETA
BETA
95%−CI
P−value
Discovery
UKBB
0.05 [0.02; 0.08]
FinnGen
0.03 [0.01; 0.05]
23andMe
0.03 [0.02; 0.05]
Fixed effects model
0.03 [0.02; 0.04] 5.86e−11
I2 = 0%
Replication
IHGC16noFIno23
0.04 [0.02; 0.07]
HUNT
0.04 [0.00; 0.08]
Fixed effects model
0.04 [0.02; 0.06] 8.55e−06
I2 = 0%
Fixed effects model
0.04 [0.03; 0.05] 3.58e−15
−0.05
0
0.05

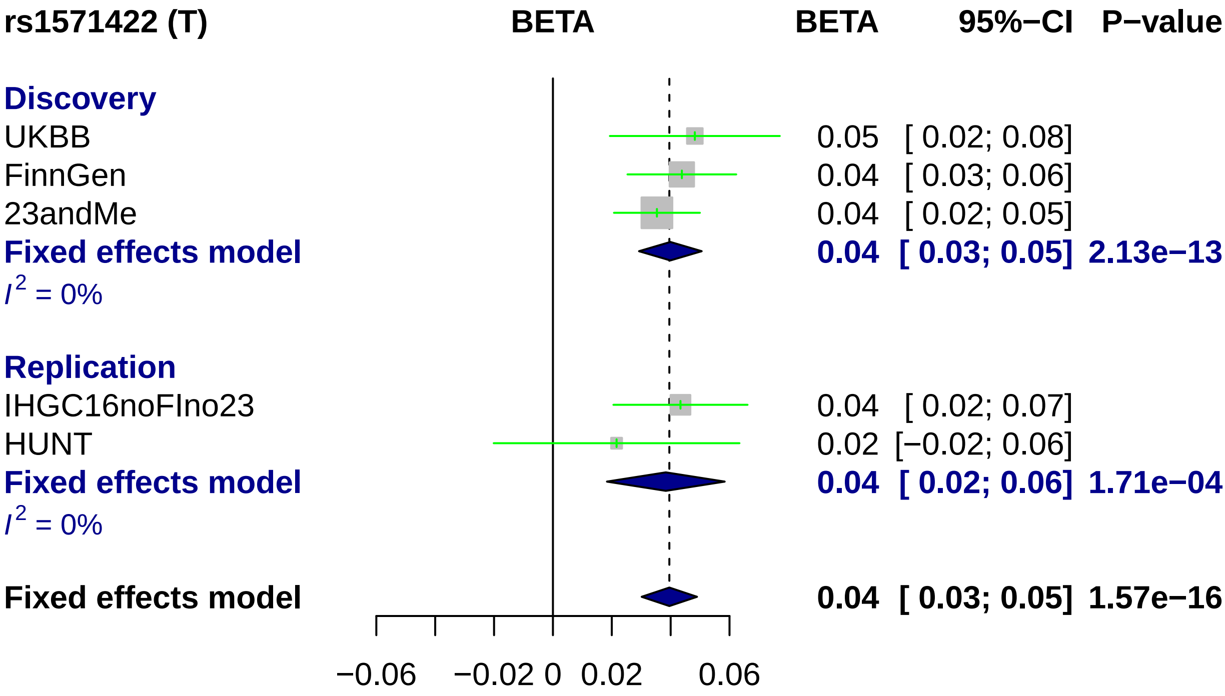
rs1571422 (T)
BETA
BETA
95%−CI
P−value
Discovery
UKBB
0.05 [ 0.02; 0.08]
FinnGen
0.04 [ 0.03; 0.06]
23andMe
0.04 [ 0.02; 0.05]
Fixed effects model
0.04 [ 0.03; 0.05] 2.13e−13
I2 = 0%
Replication
IHGC16noFIno23
0.04 [ 0.02; 0.07]
HUNT
0.02 [−0.02; 0.06]
Fixed effects model
0.04 [ 0.02; 0.06] 1.71e−04
I2 = 0%
Fixed effects model
0.04 [ 0.03; 0.05] 1.57e−16
−0.06
−0.02
0
0.02
0.06

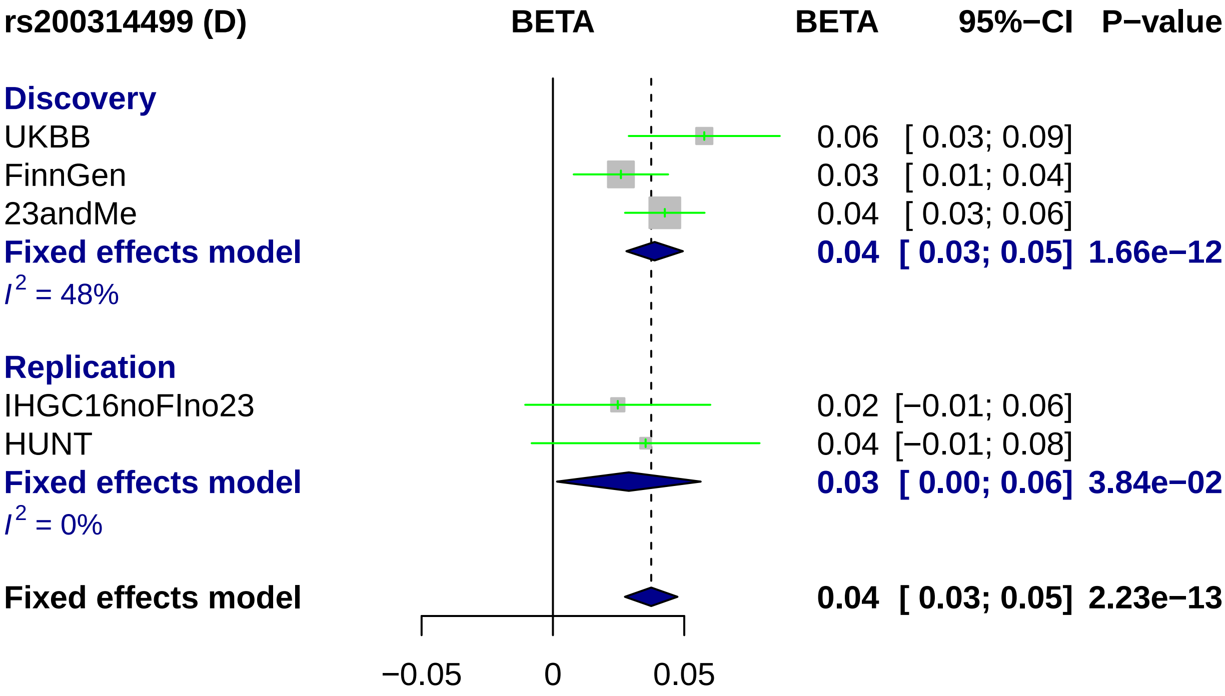
rs200314499 (D)
BETA
BETA
95%−CI
P−value
Discovery
UKBB
0.06 [ 0.03; 0.09]
FinnGen
0.03 [ 0.01; 0.04]
23andMe
0.04 [ 0.03; 0.06]
Fixed effects model
0.04 [ 0.03; 0.05] 1.66e−12
I2 = 48%
Replication
IHGC16noFIno23
0.02 [−0.01; 0.06]
HUNT
0.04 [−0.01; 0.08]
Fixed effects model
0.03 [ 0.00; 0.06] 3.84e−02
I2 = 0%
Fixed effects model
0.04 [ 0.03; 0.05] 2.23e−13
−0.05
0
0.05

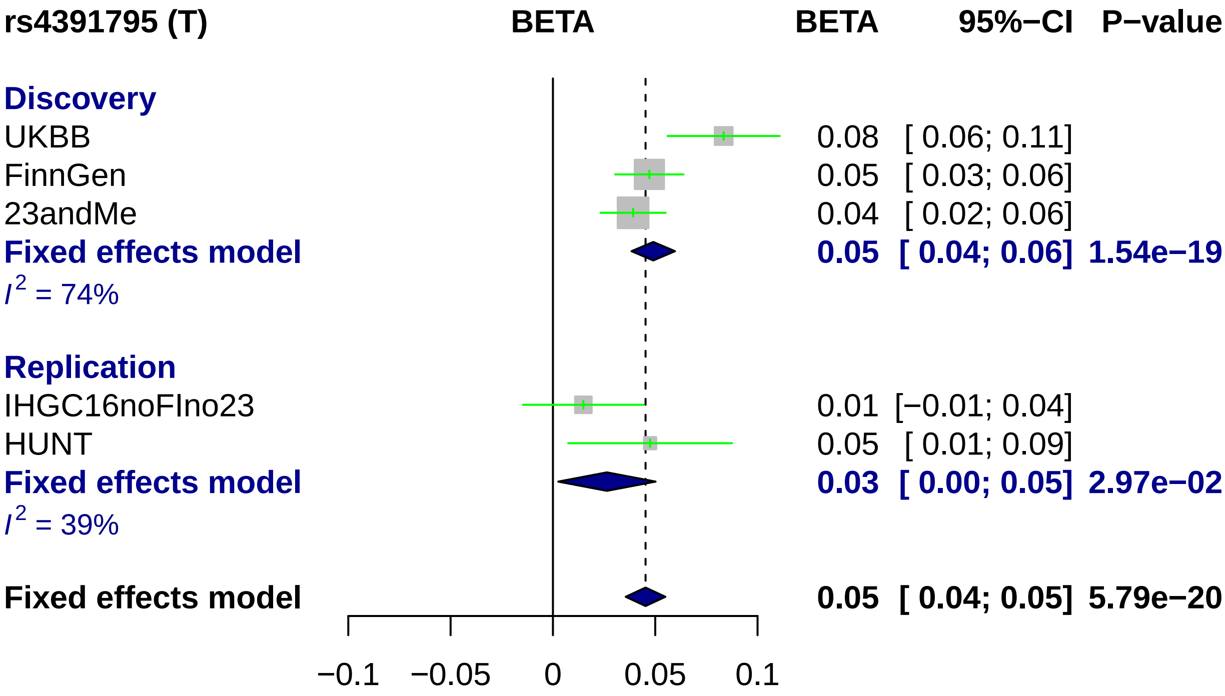
rs4391795 (T)
BETA
BETA
95%−CI
P−value
Discovery
UKBB 0.08 [ 0.06; 0.11]
FinnGen 0.05 [ 0.03; 0.06]
23andMe 0.04 [ 0.02; 0.06]
Fixed effects model 0.05 [ 0.04; 0.06] 1.54e−19
I2 = 74%
Replication
IHGC16noFIno23 0.01 [−0.01; 0.04]
HUNT 0.05 [ 0.01; 0.09]
Fixed effects model 0.03 [ 0.00; 0.05] 2.97e−02
I2 = 39%
Fixed effects model 0.05 [ 0.04; 0.05] 5.79e−20
−0.1
−0.05
0
0.05
0.1

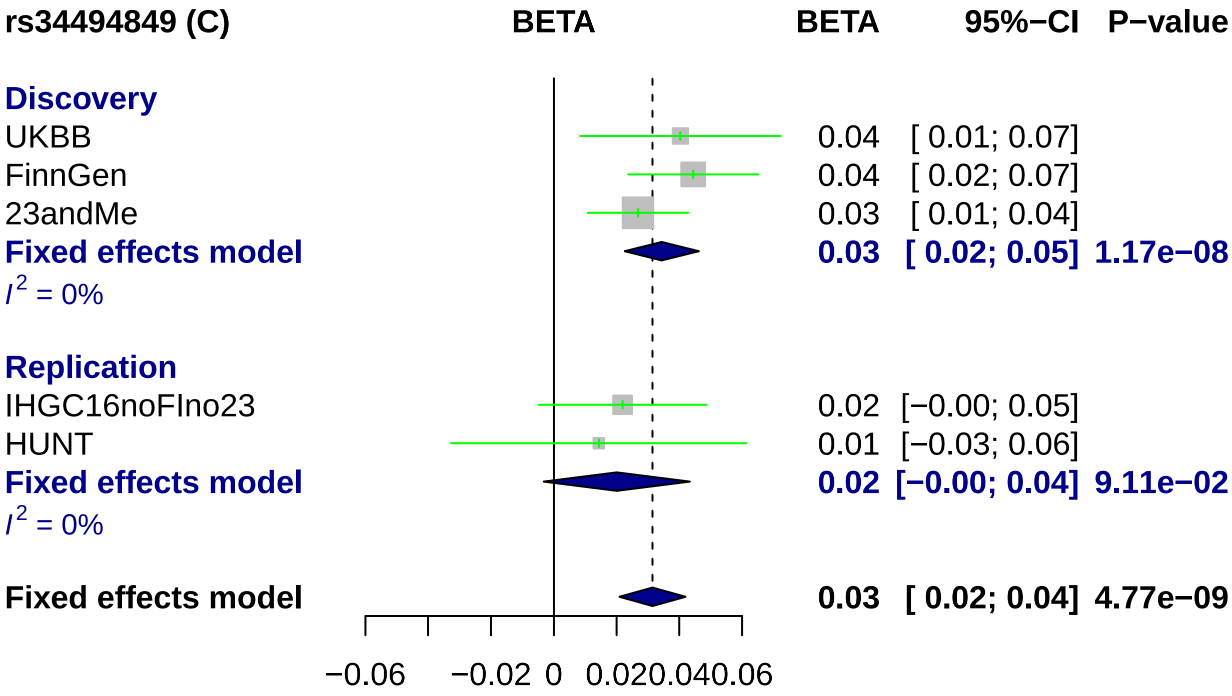
rs34494849 (C)
BETA
BETA
95%−CI
P−value
Discovery
UKBB
0.04
[ 0.01; 0.07]
FinnGen
0.04
[ 0.02; 0.07]
23andMe
0.03
[ 0.01; 0.04]
Fixed effects model
0.03
[ 0.02; 0.05]
1.17e−08
I2 = 0%
Replication
IHGC16noFIno23
0.02
[−0.00; 0.05]
HUNT
0.01
[−0.03; 0.06]
Fixed effects model
0.02
[−0.00; 0.04]
9.11e−02
I2 = 0%
Fixed effects model
0.03
[ 0.02; 0.04]
4.77e−09
−0.06
−0.02
0
0.02
0.04
0.06

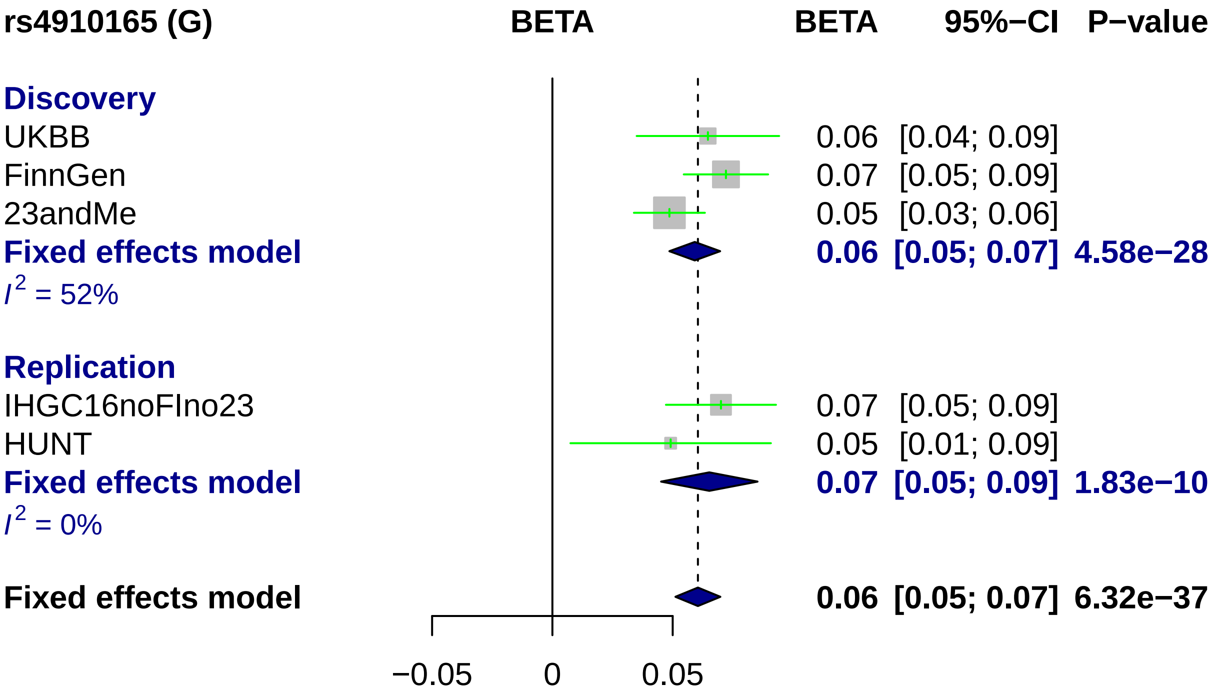

rs4910165 (G)
BETA
BETA
95%−CI
P−value
Discovery
UKBB
0.06 [0.04; 0.09]
FinnGen
0.07 [0.05; 0.09]
23andMe
0.05 [0.03; 0.06]
Fixed effects model
0.06 [0.05; 0.07] 4.58e−28
$I^2$ = 52%
Replication
IHGC16noFIno23
0.07 [0.05; 0.09]
HUNT
0.05 [0.01; 0.09]
Fixed effects model
0.07 [0.05; 0.09] 1.83e−10
$I^2$ = 0%
Fixed effects model
0.06 [0.05; 0.07] 6.32e−37
−0.05
0
0.05

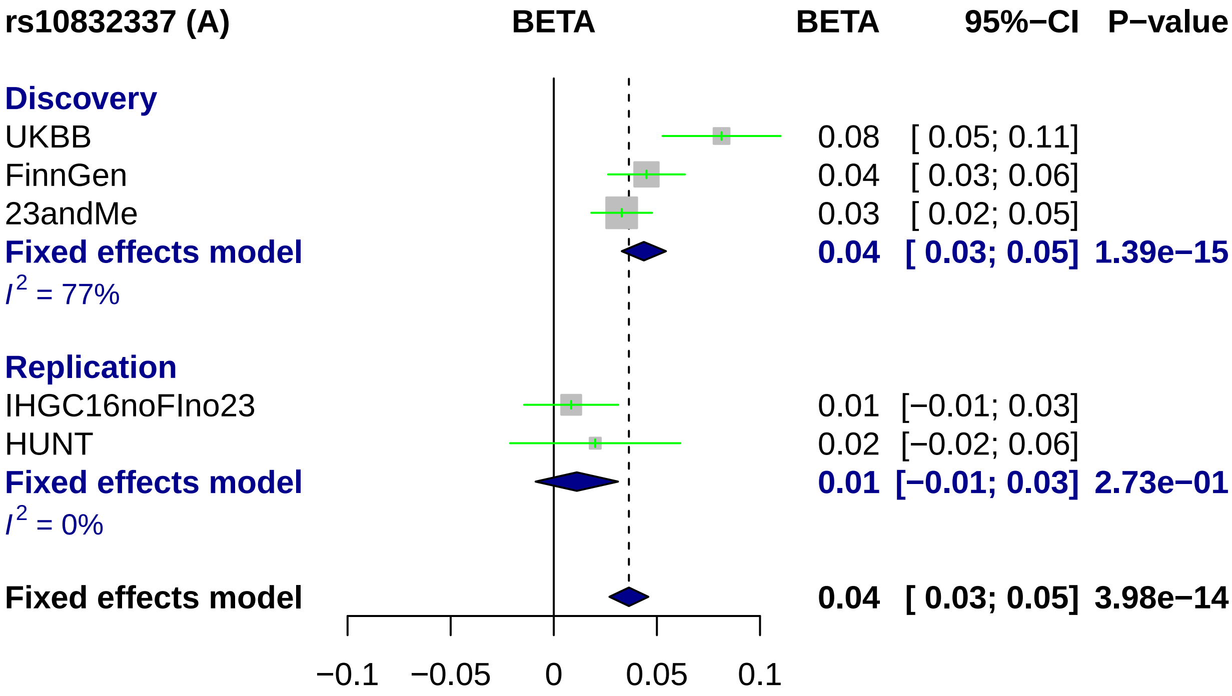
rs10832337 (A)
BETA
BETA
95%−CI
P−value
Discovery
UKBB
0.08 [ 0.05; 0.11]
FinnGen
0.04 [ 0.03; 0.06]
23andMe
0.03 [ 0.02; 0.05]
Fixed effects model
0.04 [ 0.03; 0.05] 1.39e−15
I2 = 77%
Replication
IHGC16noFIno23
0.01 [−0.01; 0.03]
HUNT
0.02 [−0.02; 0.06]
Fixed effects model
0.01 [−0.01; 0.03] 2.73e−01
I2 = 0%
Fixed effects model
0.04 [ 0.03; 0.05] 3.98e−14
−0.1
−0.05
0
0.05
0.1

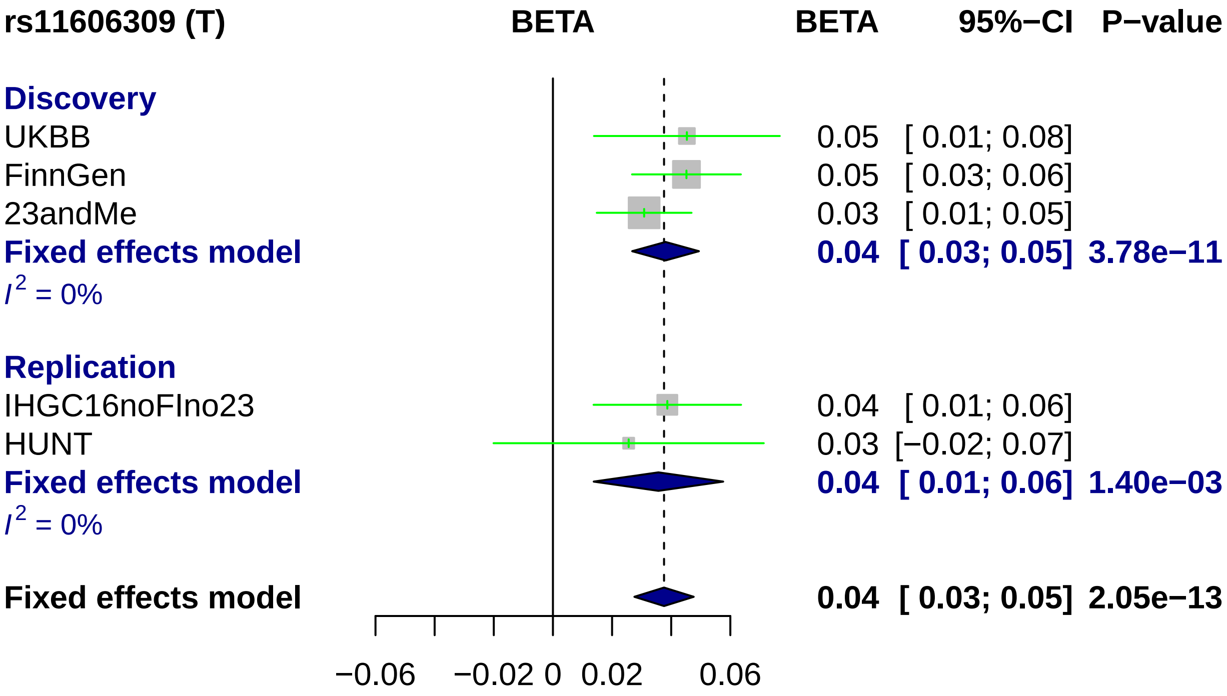
rs11606309 (T)
BETA
BETA
95%−CI
P−value
Discovery
UKBB
0.05 [ 0.01; 0.08]
FinnGen
0.05 [ 0.03; 0.06]
23andMe
0.03 [ 0.01; 0.05]
Fixed effects model
0.04 [ 0.03; 0.05] 3.78e−11
I2 = 0%
Replication
IHGC16noFIno23
0.04 [ 0.01; 0.06]
HUNT
0.03 [−0.02; 0.07]
Fixed effects model
0.04 [ 0.01; 0.06] 1.40e−03
I2 = 0%
Fixed effects model
0.04 [ 0.03; 0.05] 2.05e−13
−0.06
−0.02
0
0.02
0.06

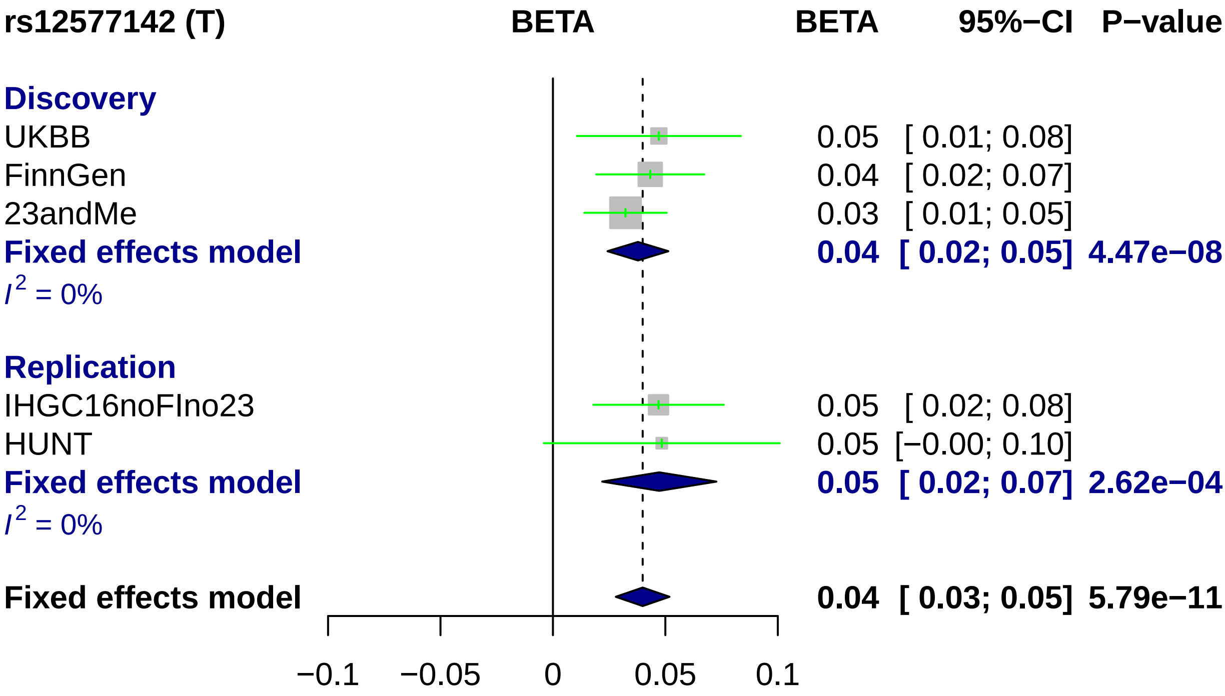
rs12577142 (T)
BETA
BETA
95%−CI
P−value
Discovery
UKBB
0.05
[ 0.01; 0.08]
FinnGen
0.04
[ 0.02; 0.07]
23andMe
0.03
[ 0.01; 0.05]
Fixed effects model
0.04
[ 0.02; 0.05]
4.47e−08
I2 = 0%
Replication
IHGC16noFIno23
0.05
[ 0.02; 0.08]
HUNT
0.05
[−0.00; 0.10]
Fixed effects model
0.05
[ 0.02; 0.07]
2.62e−04
I2 = 0%
Fixed effects model
0.04
[ 0.03; 0.05]
5.79e−11
−0.1
−0.05
0
0.05
0.1

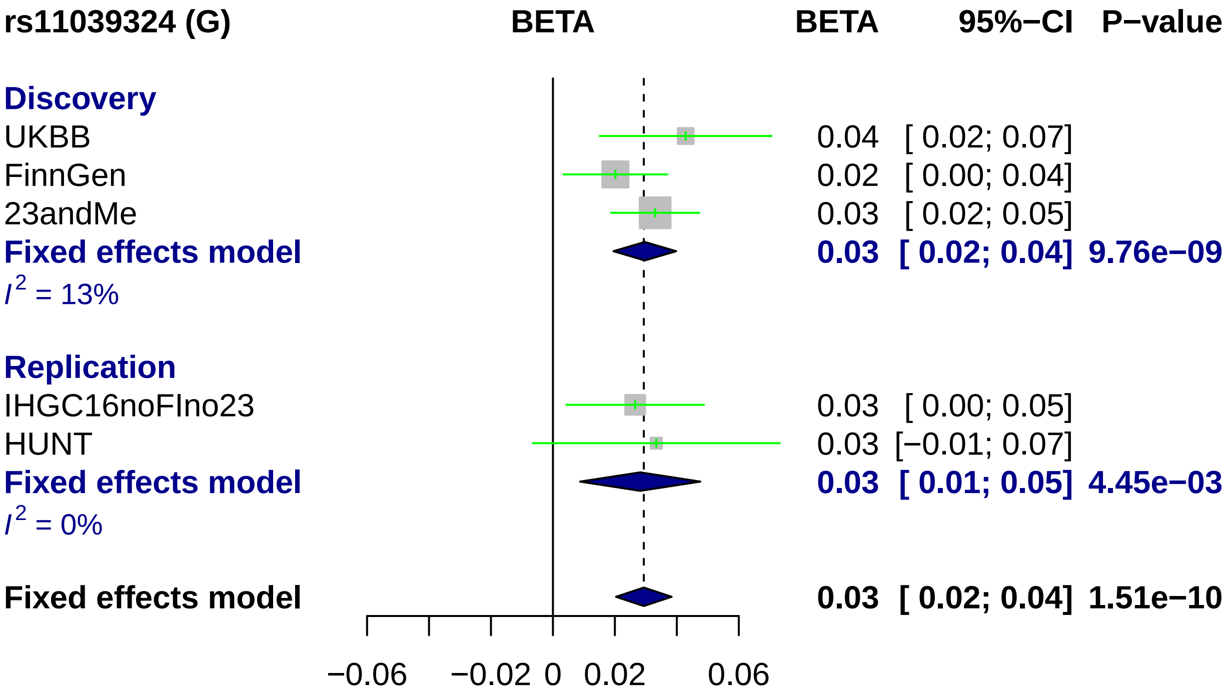
rs11039324 (G)
BETA
BETA
95%−CI
P−value
Discovery
UKBB
0.04 [ 0.02; 0.07]
FinnGen
0.02 [ 0.00; 0.04]
23andMe
0.03 [ 0.02; 0.05]
Fixed effects model
0.03 [ 0.02; 0.04] 9.76e−09
I2 = 13%
Replication
IHGC16noFIno23
0.03 [ 0.00; 0.05]
HUNT
0.03 [−0.01; 0.07]
Fixed effects model
0.03 [ 0.01; 0.05] 4.45e−03
I2 = 0%
Fixed effects model
0.03 [ 0.02; 0.04] 1.51e−10
−0.06
−0.02
0
0.02
0.06

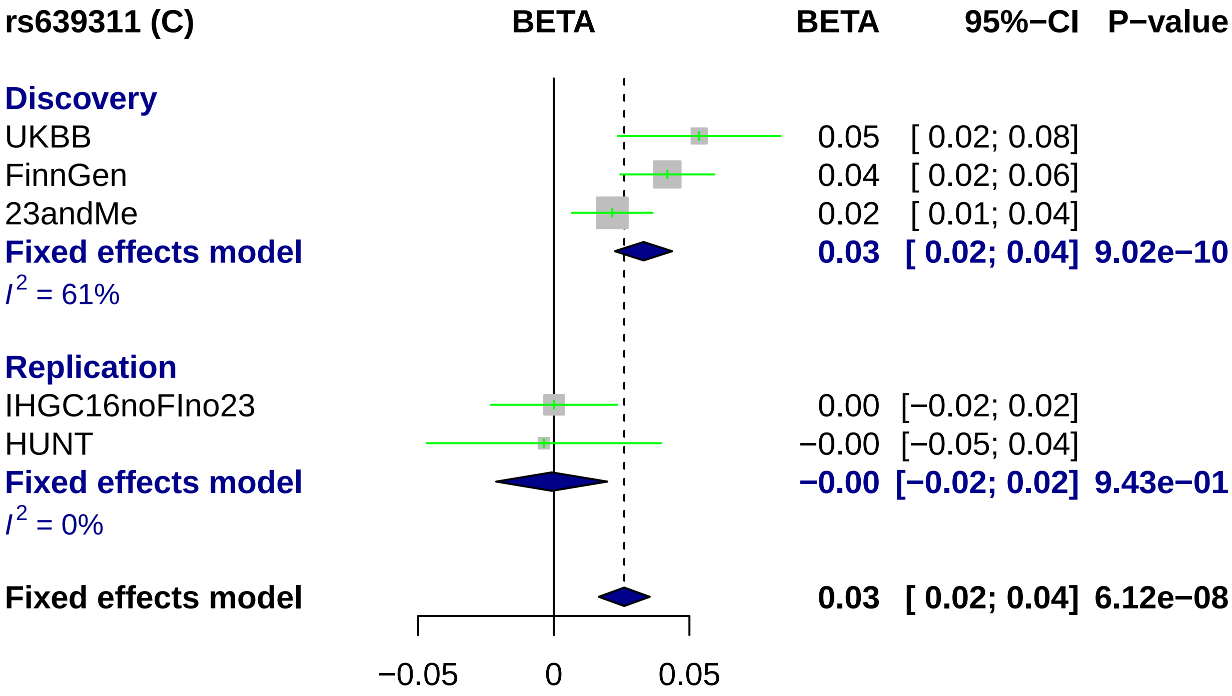
rs639311 (C)
BETA
BETA
95%−CI
P−value
Discovery
UKBB
0.05
[ 0.02; 0.08]
FinnGen
0.04
[ 0.02; 0.06]
23andMe
0.02
[ 0.01; 0.04]
Fixed effects model
0.03
[ 0.02; 0.04]
9.02e−10
I^2 = 61%
Replication
IHGC16noFIno23
0.00
[−0.02; 0.02]
HUNT
−0.00
[−0.05; 0.04]
Fixed effects model
−0.00
[−0.02; 0.02]
9.43e−01
I^2 = 0%
Fixed effects model
0.03
[ 0.02; 0.04]
6.12e−08
−0.05
0
0.05

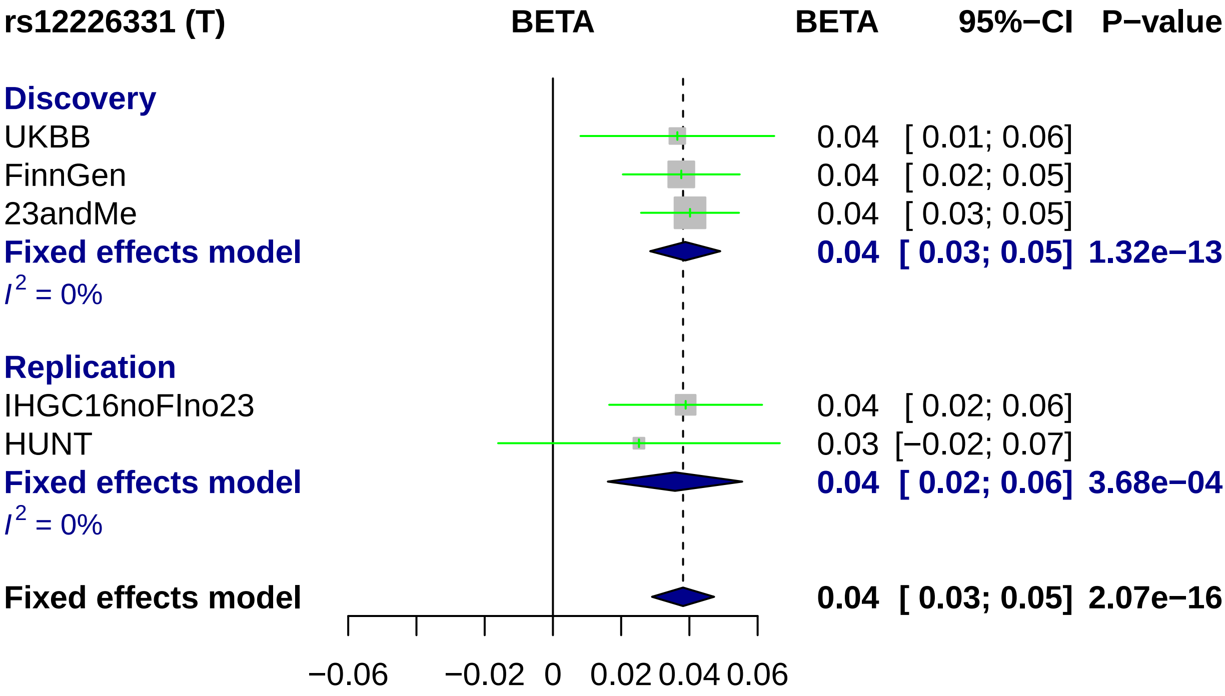
rs12226331 (T)
BETA
BETA
95%−CI
P−value
Discovery
UKBB 0.04 [ 0.01; 0.06]
FinnGen 0.04 [ 0.02; 0.05]
23andMe 0.04 [ 0.03; 0.05]
Fixed effects model 0.04 [ 0.03; 0.05] 1.32e−13
I2 = 0%
Replication
IHGC16noFIno23 0.04 [ 0.02; 0.06]
HUNT 0.03 [−0.02; 0.07]
Fixed effects model 0.04 [ 0.02; 0.06] 3.68e−04
I2 = 0%
Fixed effects model 0.04 [ 0.03; 0.05] 2.07e−16
−0.06
−0.02
0
0.02
0.04
0.06

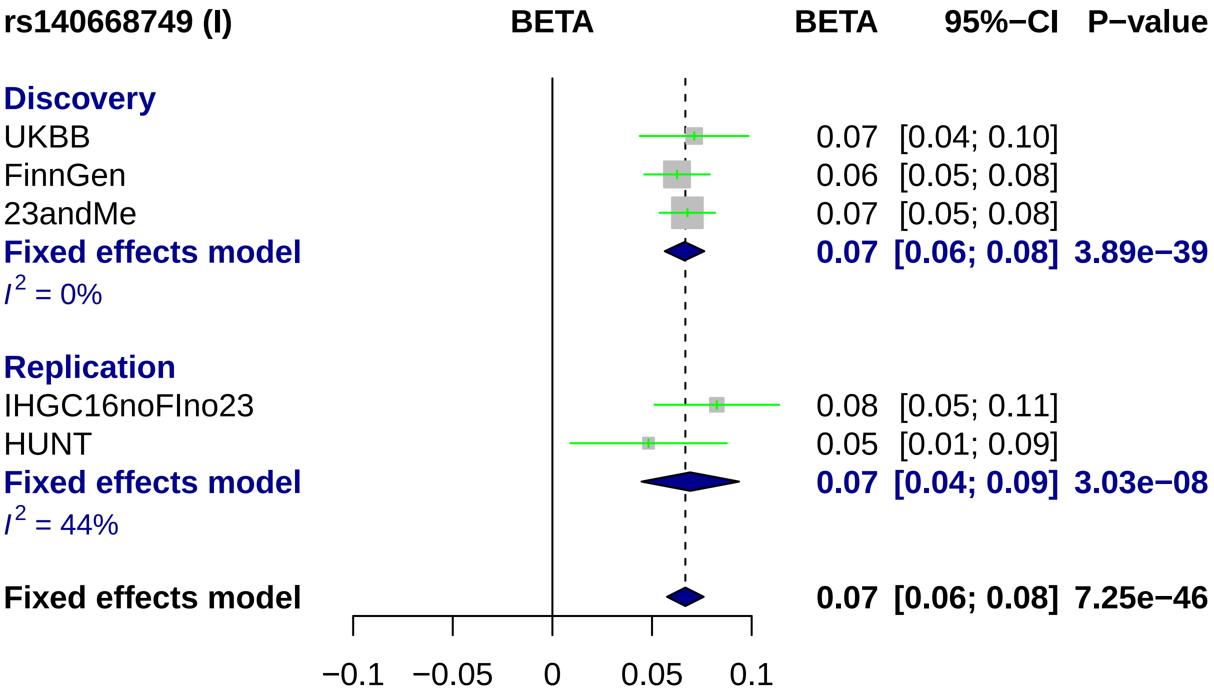
rs140668749 (I)
BETA
BETA
95%−CI
P−value
Discovery
UKBB
0.07 [0.04; 0.10]
FinnGen
0.06 [0.05; 0.08]
23andMe
0.07 [0.05; 0.08]
Fixed effects model
0.07 [0.06; 0.08] 3.89e−39
I^2 = 0%
Replication
IHGC16noFIno23
0.08 [0.05; 0.11]
HUNT
0.05 [0.01; 0.09]
Fixed effects model
0.07 [0.04; 0.09] 3.03e−08
I^2 = 44%
Fixed effects model
0.07 [0.06; 0.08] 7.25e−46
−0.1
−0.05
0
0.05
0.1

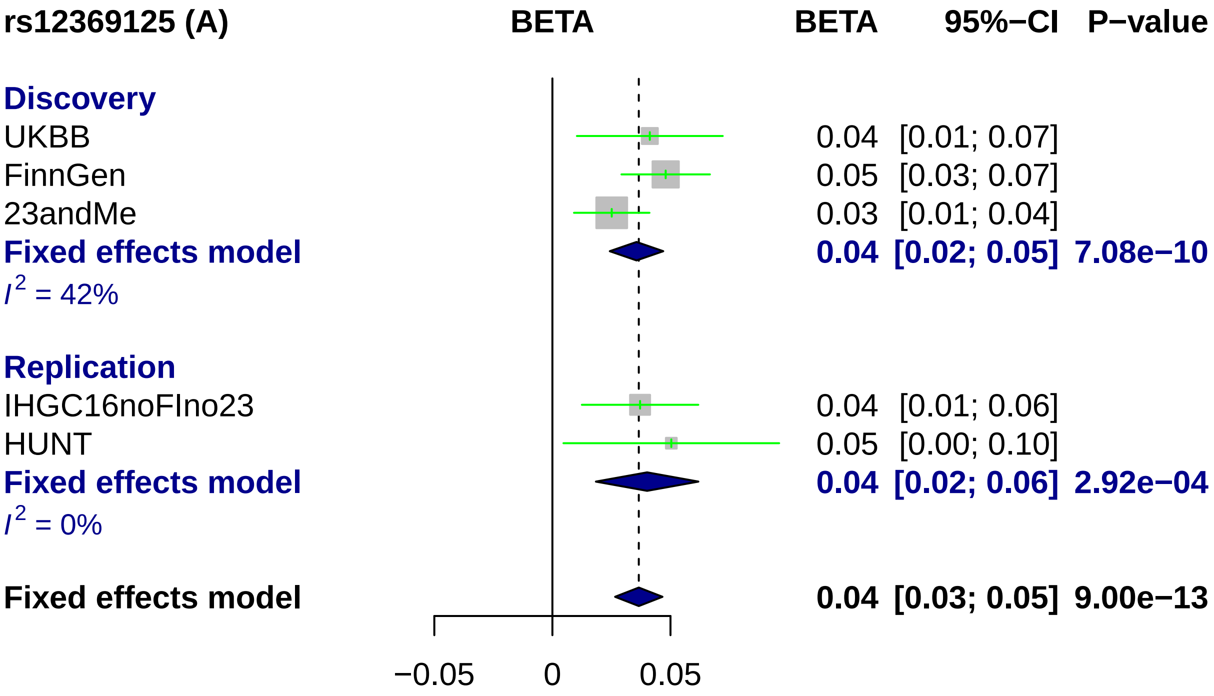

rs12369125 (A)
BETA
BETA
95%−CI
P−value
Discovery
UKBB
0.04
[0.01; 0.07]
FinnGen
0.05
[0.03; 0.07]
23andMe
0.03
[0.01; 0.04]
Fixed effects model
0.04
[0.02; 0.05]
7.08e−10
I2 = 42%
Replication
IHGC16noFIno23
0.04
[0.01; 0.06]
HUNT
0.05
[0.00; 0.10]
Fixed effects model
0.04
[0.02; 0.06]
2.92e−04
I2 = 0%
Fixed effects model
0.04
[0.03; 0.05]
9.00e−13
−0.05
0
0.05

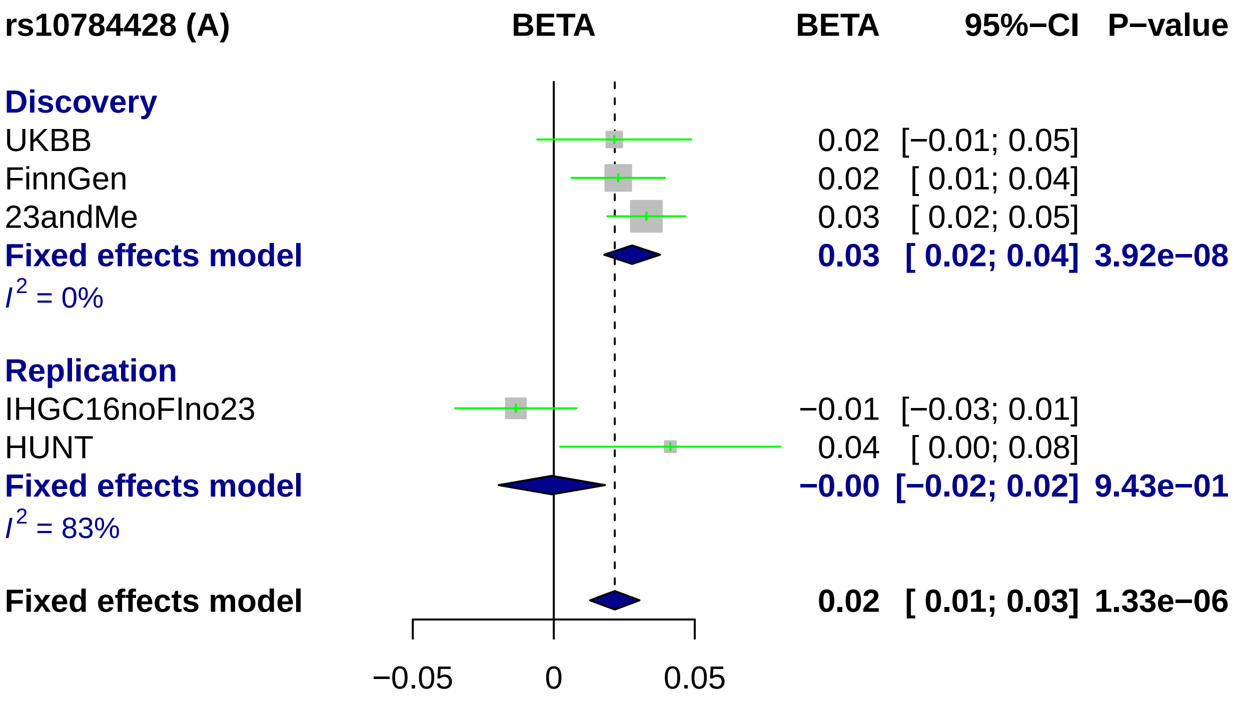
rs10784428 (A)
BETA
BETA
95%−CI
P−value
Discovery
UKBB
0.02 [−0.01; 0.05]
FinnGen
0.02 [ 0.01; 0.04]
23andMe
0.03 [ 0.02; 0.05]
Fixed effects model
0.03 [ 0.02; 0.04] 3.92e−08
$I^2$ = 0%
Replication
IHGC16noFIno23
−0.01 [−0.03; 0.01]
HUNT
0.04 [ 0.00; 0.08]
Fixed effects model
−0.00 [−0.02; 0.02] 9.43e−01
$I^2$ = 83%
Fixed effects model
0.02 [ 0.01; 0.03] 1.33e−06
−0.05
0
0.05

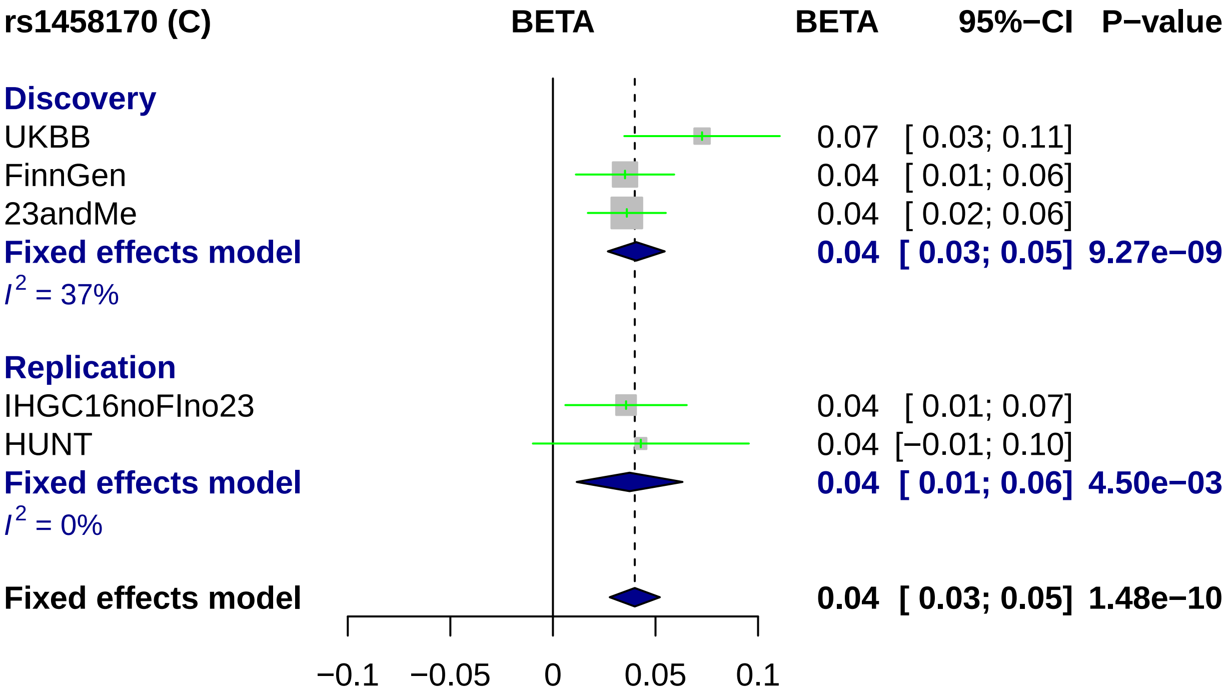
rs1458170 (C)
BETA
BETA
95%−CI
P−value
Discovery
UKBB
0.07 [ 0.03; 0.11]
FinnGen
0.04 [ 0.01; 0.06]
23andMe
0.04 [ 0.02; 0.06]
Fixed effects model
0.04 [ 0.03; 0.05] 9.27e−09
I2 = 37%
Replication
IHGC16noFIno23
0.04 [ 0.01; 0.07]
HUNT
0.04 [−0.01; 0.10]
Fixed effects model
0.04 [ 0.01; 0.06] 4.50e−03
I2 = 0%
Fixed effects model
0.04 [ 0.03; 0.05] 1.48e−10
−0.1
−0.05
0
0.05
0.1

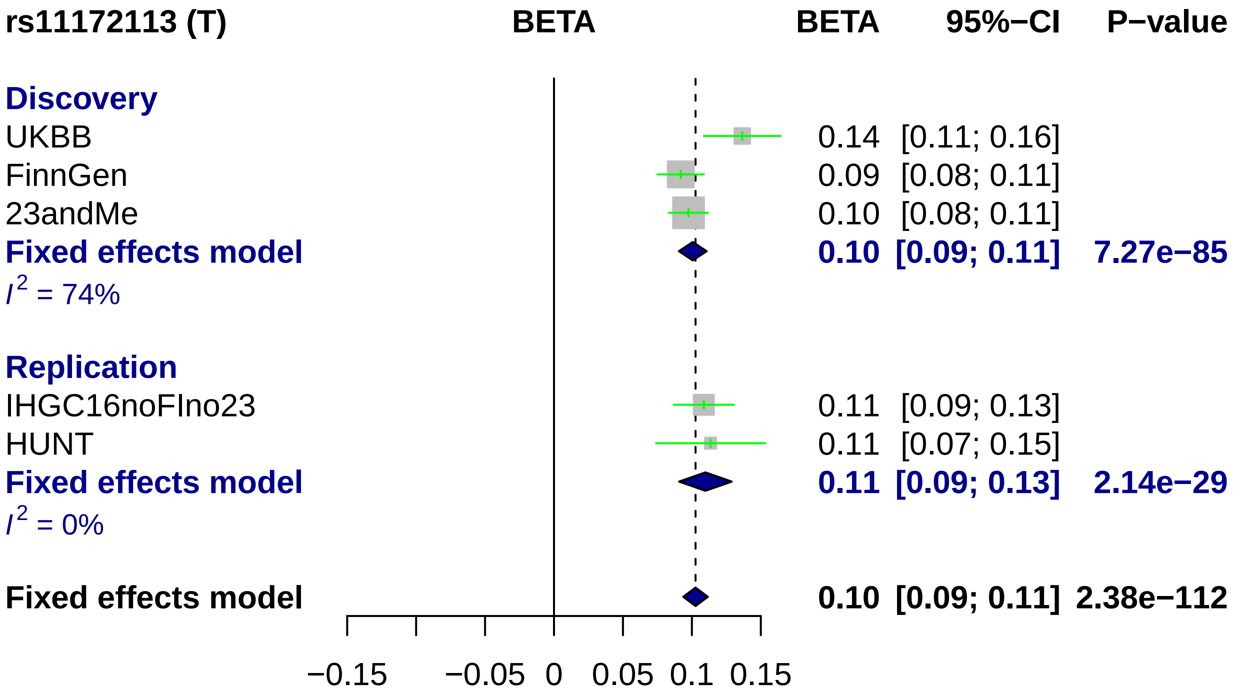
rs11172113 (T)
BETA
BETA
95%−CI
P−value
Discovery
UKBB
0.14 [0.11; 0.16]
FinnGen
0.09 [0.08; 0.11]
23andMe
0.10 [0.08; 0.11]
Fixed effects model
0.10 [0.09; 0.11] 7.27e−85
I2 = 74%
Replication
IHGC16noFIno23
0.11 [0.09; 0.13]
HUNT
0.11 [0.07; 0.15]
Fixed effects model
0.11 [0.09; 0.13] 2.14e−29
I2 = 0%
Fixed effects model
0.10 [0.09; 0.11] 2.38e−112
−0.15
−0.05
0
0.05
0.1
0.15

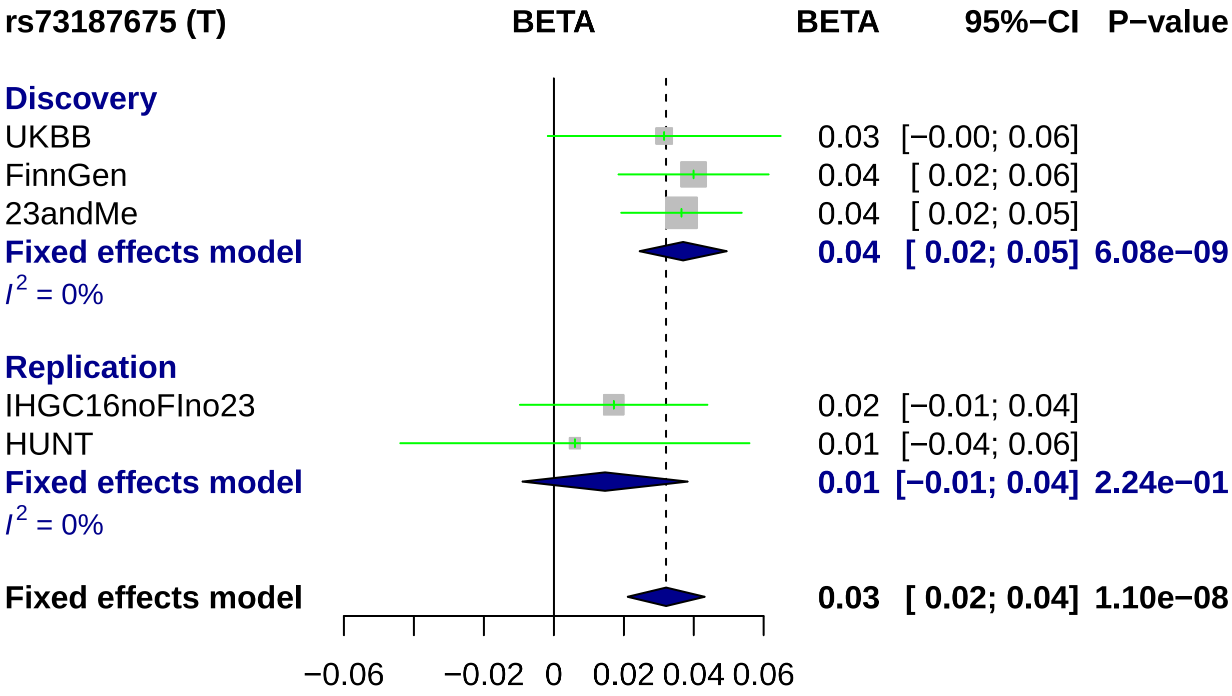
rs73187675 (T)
BETA
BETA
95%−CI
P−value
Discovery
UKBB
0.03 [−0.00; 0.06]
FinnGen
0.04 [ 0.02; 0.06]
23andMe
0.04 [ 0.02; 0.05]
Fixed effects model
0.04 [ 0.02; 0.05] 6.08e−09
I^2 = 0%
Replication
IHGC16noFIno23
0.02 [−0.01; 0.04]
HUNT
0.01 [−0.04; 0.06]
Fixed effects model
0.01 [−0.01; 0.04] 2.24e−01
I^2 = 0%
Fixed effects model
0.03 [ 0.02; 0.04] 1.10e−08
−0.06
−0.02
0
0.02
0.04
0.06

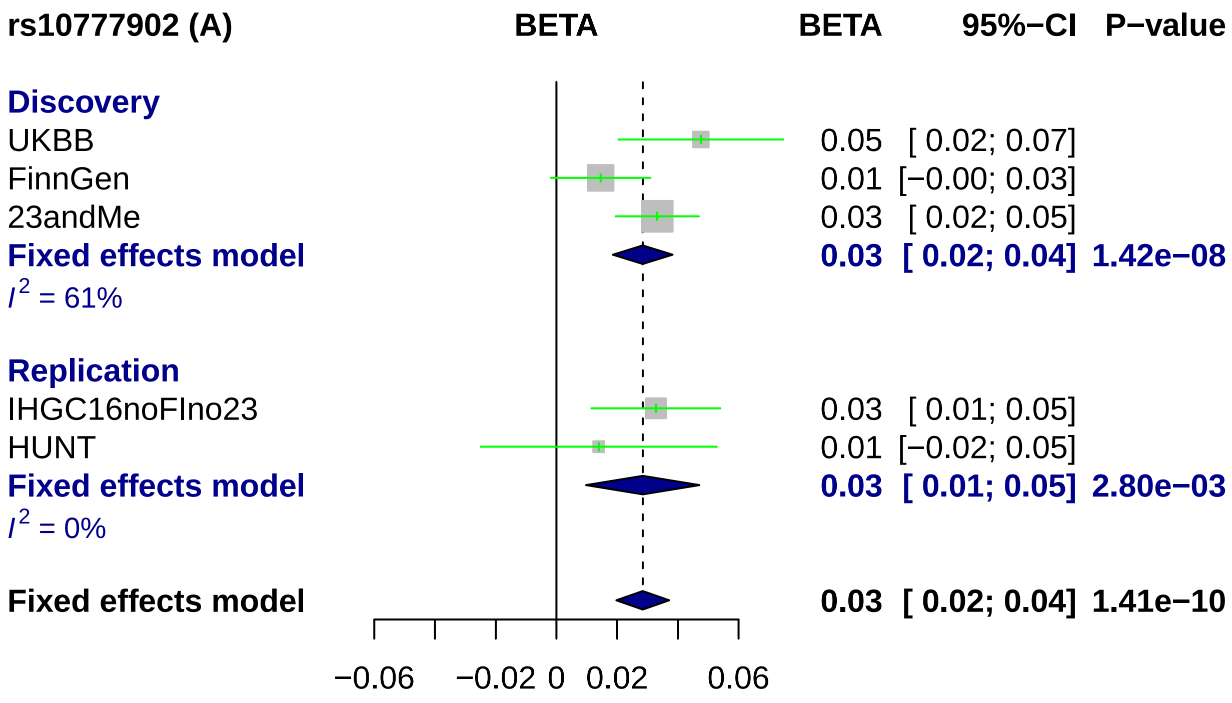
rs10777902 (A)
BETA
BETA
95%−CI
P−value
Discovery
UKBB
FinnGen
23andMe
Fixed effects model
$I^2$ = 61%
Replication
IHGC16noFIno23
HUNT
Fixed effects model
$I^2$ = 0%
Fixed effects model
0.05 [ 0.02; 0.07]
0.01 [−0.00; 0.03]
0.03 [ 0.02; 0.05]
0.03 [ 0.02; 0.04] 1.42e−08
0.03 [ 0.01; 0.05]
0.01 [−0.02; 0.05]
0.03 [ 0.01; 0.05] 2.80e−03
0.03 [ 0.02; 0.04] 1.41e−10
−0.06
−0.02
0
0.02
0.06

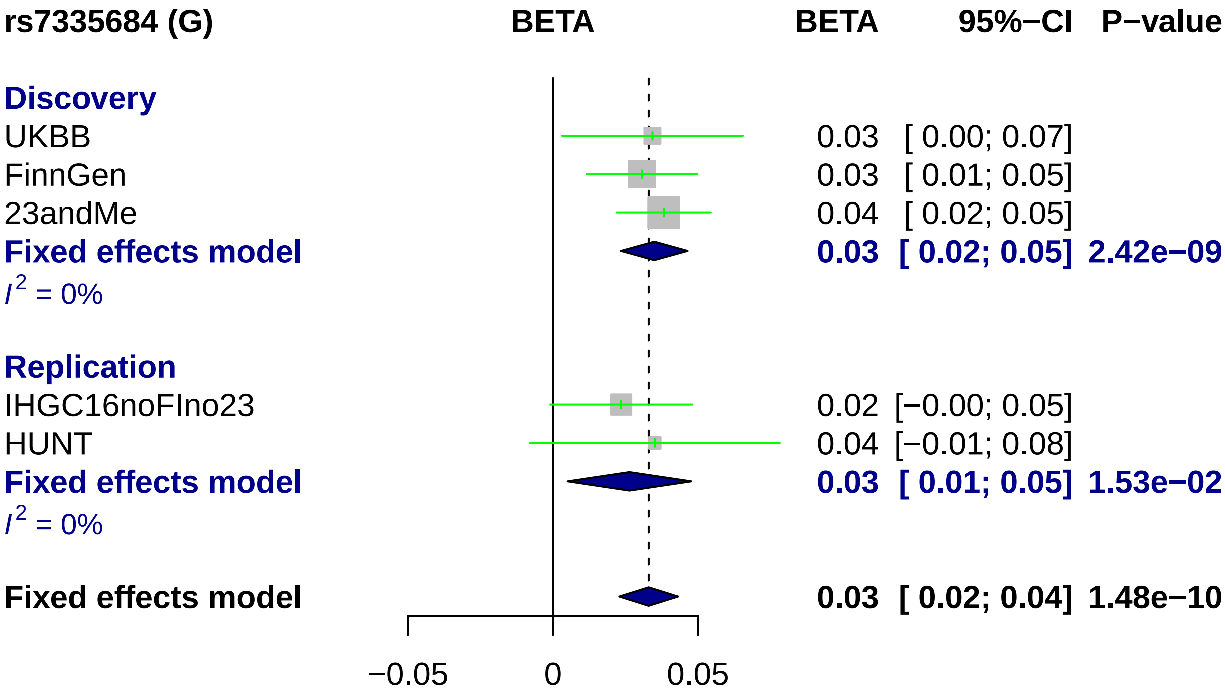
rs7335684 (G)
BETA
BETA
95%−CI
P−value
Discovery
UKBB
0.03
[ 0.00; 0.07]
FinnGen
0.03
[ 0.01; 0.05]
23andMe
0.04
[ 0.02; 0.05]
Fixed effects model
0.03
[ 0.02; 0.05]
2.42e−09
I2 = 0%
Replication
IHGC16noFIno23
0.02
[−0.00; 0.05]
HUNT
0.04
[−0.01; 0.08]
Fixed effects model
0.03
[ 0.01; 0.05]
1.53e−02
I2 = 0%
Fixed effects model
0.03
[ 0.02; 0.04]
1.48e−10
−0.05
0
0.05

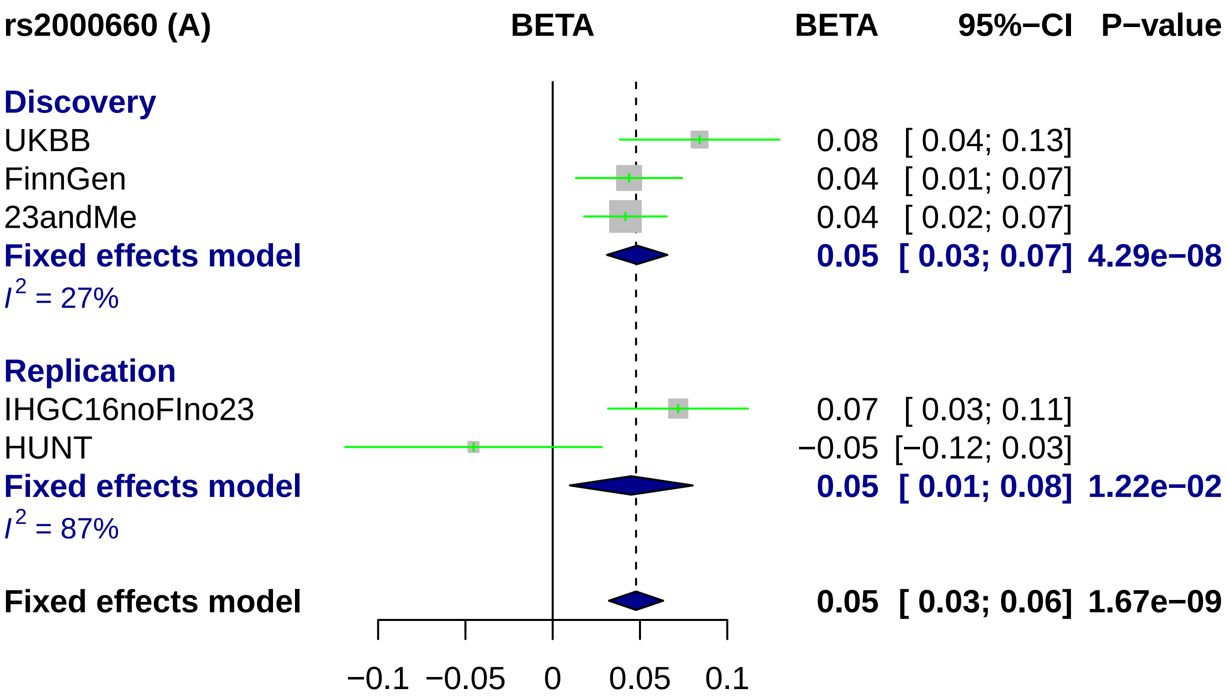
rs2000660 (A)
BETA
BETA
95%−CI
P−value
Discovery
UKBB
0.08
[ 0.04; 0.13]
FinnGen
0.04
[ 0.01; 0.07]
23andMe
0.04
[ 0.02; 0.07]
Fixed effects model
0.05
[ 0.03; 0.07]
4.29e−08
I2 = 27%
Replication
IHGC16noFIno23
0.07
[ 0.03; 0.11]
HUNT
−0.05
[−0.12; 0.03]
Fixed effects model
0.05
[ 0.01; 0.08]
1.22e−02
I2 = 87%
Fixed effects model
0.05
[ 0.03; 0.06]
1.67e−09
−0.1
−0.05
0
0.05
0.1

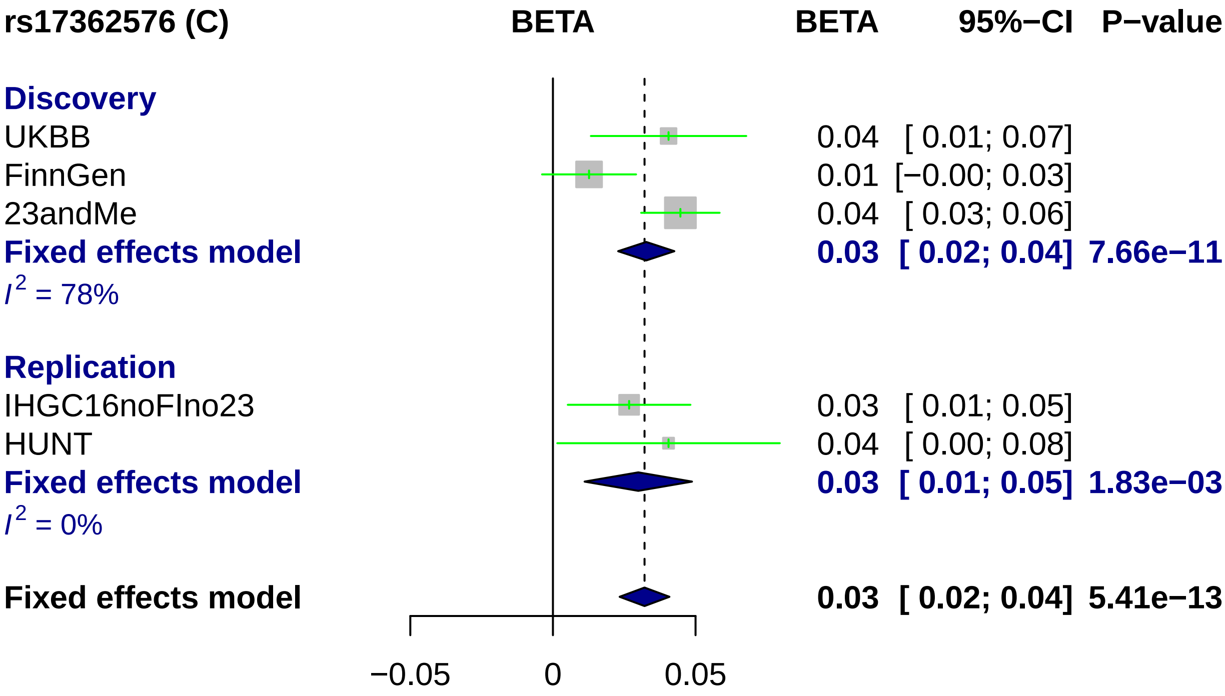
rs17362576 (C)
BETA
BETA
95%−CI
P−value
Discovery
UKBB
0.04 [ 0.01; 0.07]
FinnGen
0.01 [−0.00; 0.03]
23andMe
0.04 [ 0.03; 0.06]
Fixed effects model
0.03 [ 0.02; 0.04] 7.66e−11
I2 = 78%
Replication
IHGC16noFIno23
0.03 [ 0.01; 0.05]
HUNT
0.04 [ 0.00; 0.08]
Fixed effects model
0.03 [ 0.01; 0.05] 1.83e−03
I2 = 0%
Fixed effects model
0.03 [ 0.02; 0.04] 5.41e−13
−0.05
0
0.05

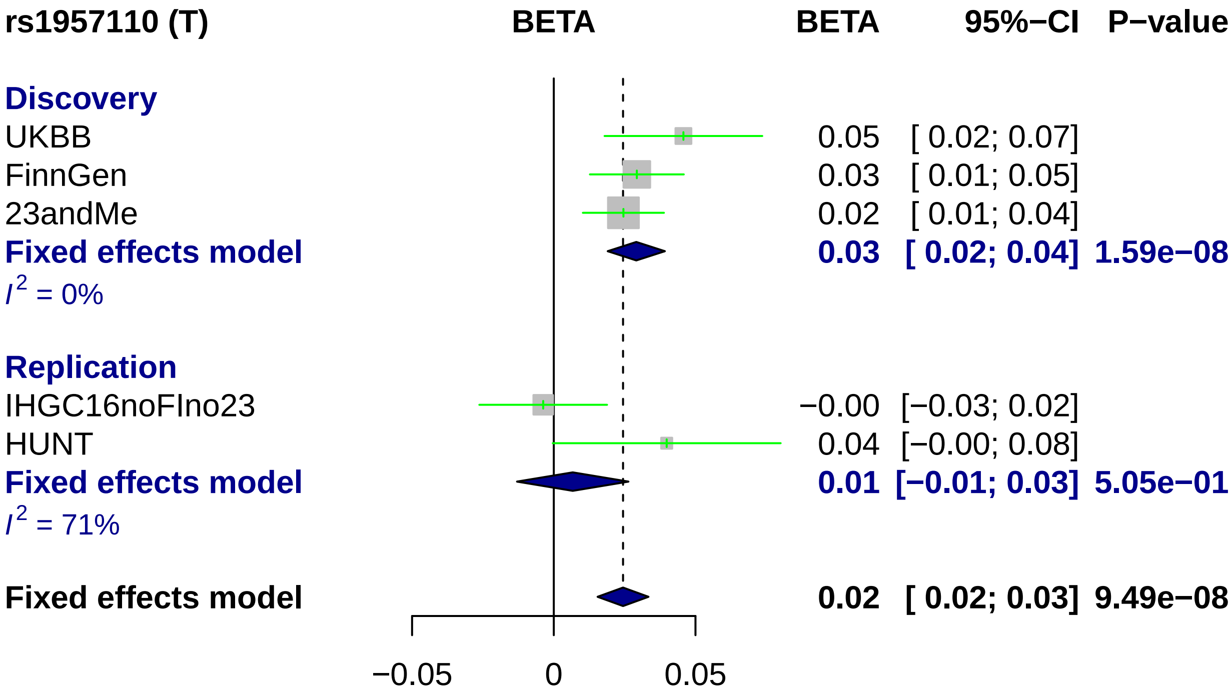
rs1957110 (T)
BETA
BETA
95%−CI
P−value
Discovery
UKBB 0.05 [ 0.02; 0.07]
FinnGen 0.03 [ 0.01; 0.05]
23andMe 0.02 [ 0.01; 0.04]
Fixed effects model 0.03 [ 0.02; 0.04] 1.59e−08
I2 = 0%
Replication
IHGC16noFIno23 −0.00 [−0.03; 0.02]
HUNT 0.04 [−0.00; 0.08]
Fixed effects model 0.01 [−0.01; 0.03] 5.05e−01
I2 = 71%
Fixed effects model 0.02 [ 0.02; 0.03] 9.49e−08
−0.05
0
0.05

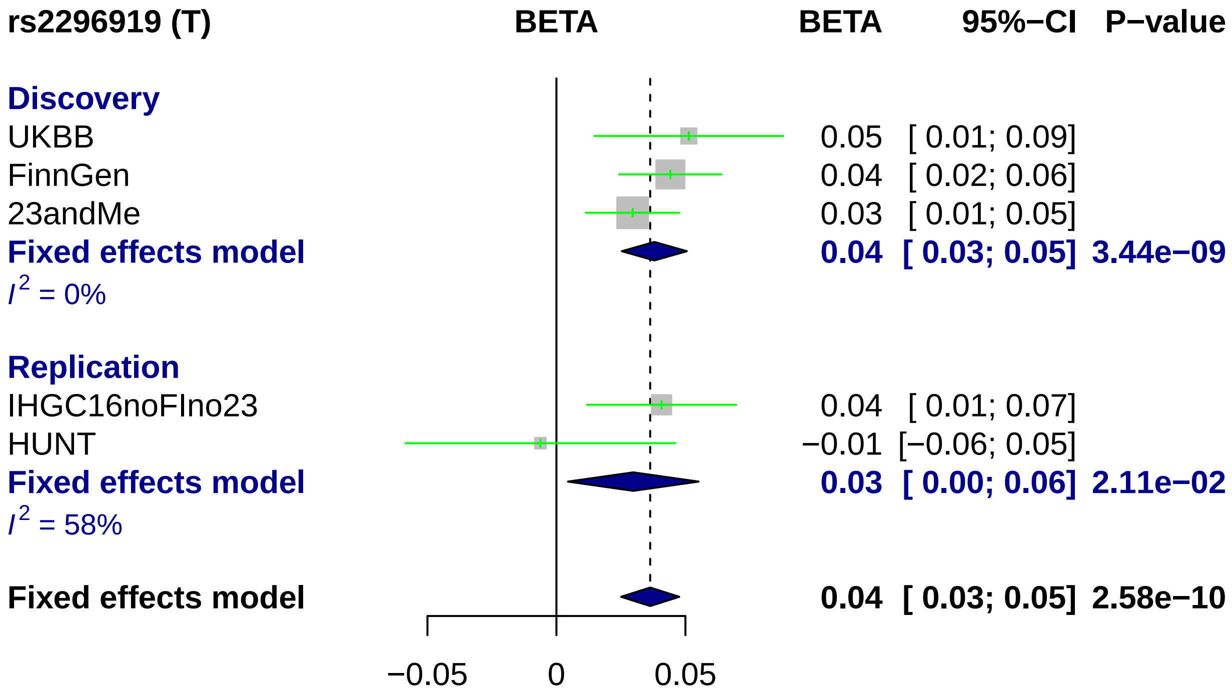

rs2296919 (T)
BETA
BETA
95%−CI
P−value
Discovery
UKBB
0.05 [ 0.01; 0.09]
FinnGen
0.04 [ 0.02; 0.06]
23andMe
0.03 [ 0.01; 0.05]
Fixed effects model
0.04 [ 0.03; 0.05] 3.44e−09
$I^2$ = 0%
Replication
IHGC16noFIno23
0.04 [ 0.01; 0.07]
HUNT
−0.01 [−0.06; 0.05]
Fixed effects model
0.03 [ 0.00; 0.06] 2.11e−02
$I^2$ = 58%
Fixed effects model
0.04 [ 0.03; 0.05] 2.58e−10
−0.05
0
0.05

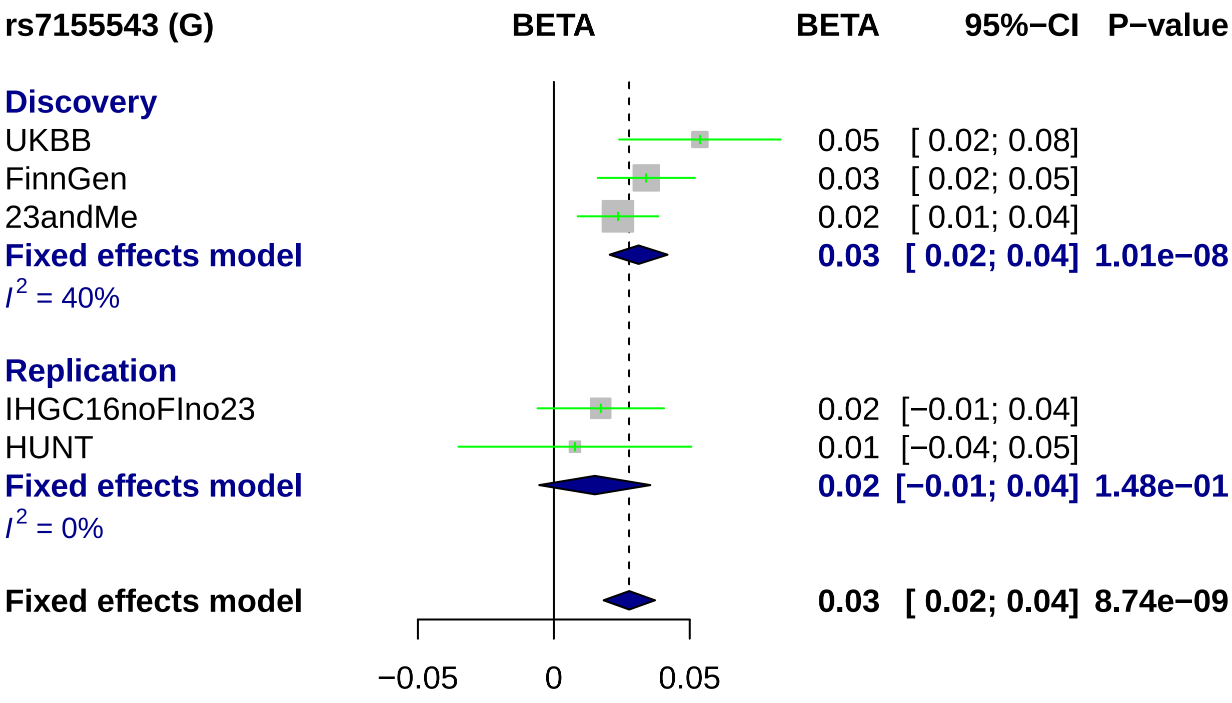

rs7155543 (G)
BETA
BETA
95%−CI
P−value
Discovery
UKBB
0.05 [ 0.02; 0.08]
FinnGen
0.03 [ 0.02; 0.05]
23andMe
0.02 [ 0.01; 0.04]
Fixed effects model
0.03 [ 0.02; 0.04] 1.01e−08
I2 = 40%
Replication
IHGC16noFIno23
0.02 [−0.01; 0.04]
HUNT
0.01 [−0.04; 0.05]
Fixed effects model
0.02 [−0.01; 0.04] 1.48e−01
I2 = 0%
Fixed effects model
0.03 [ 0.02; 0.04] 8.74e−09
−0.05
0
0.05

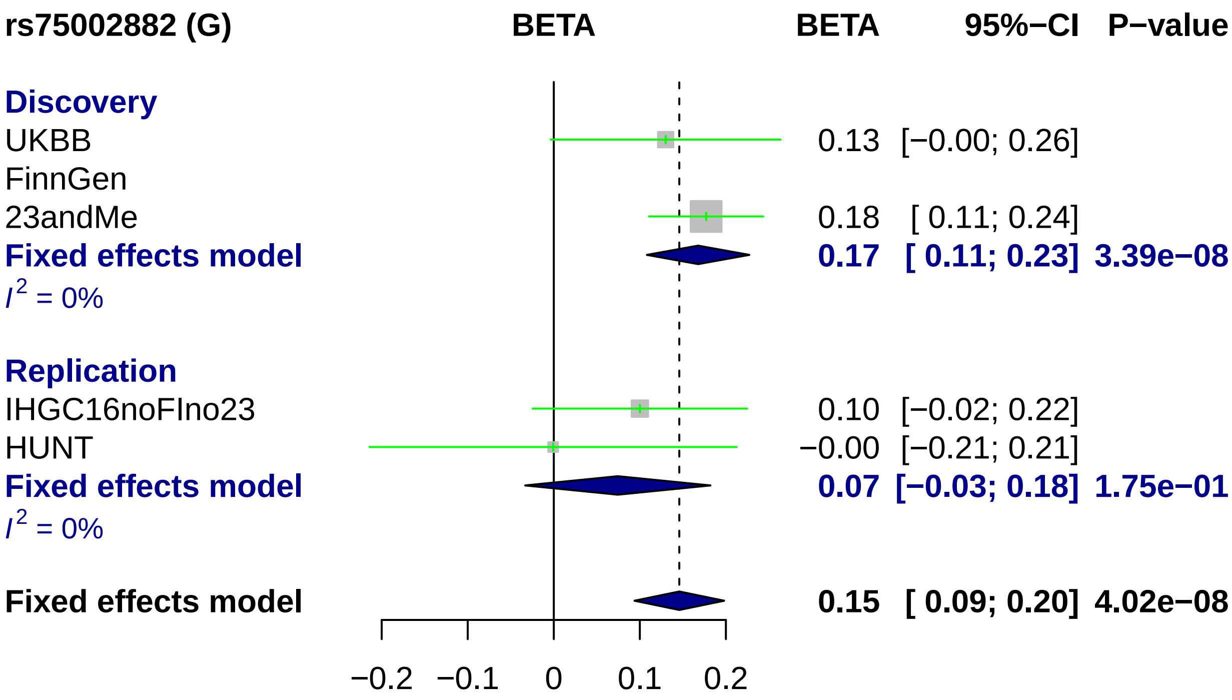
rs75002882 (G)
BETA
BETA
95%−CI
P−value
Discovery
UKBB
0.13 [−0.00; 0.26]
FinnGen
23andMe
0.18 [ 0.11; 0.24]
Fixed effects model
0.17 [ 0.11; 0.23] 3.39e−08
$I^2$ = 0%
Replication
IHGC16noFIno23
0.10 [−0.02; 0.22]
HUNT
−0.00 [−0.21; 0.21]
Fixed effects model
0.07 [−0.03; 0.18] 1.75e−01
$I^2$ = 0%
Fixed effects model
0.15 [ 0.09; 0.20] 4.02e−08
−0.2
−0.1
0
0.1
0.2

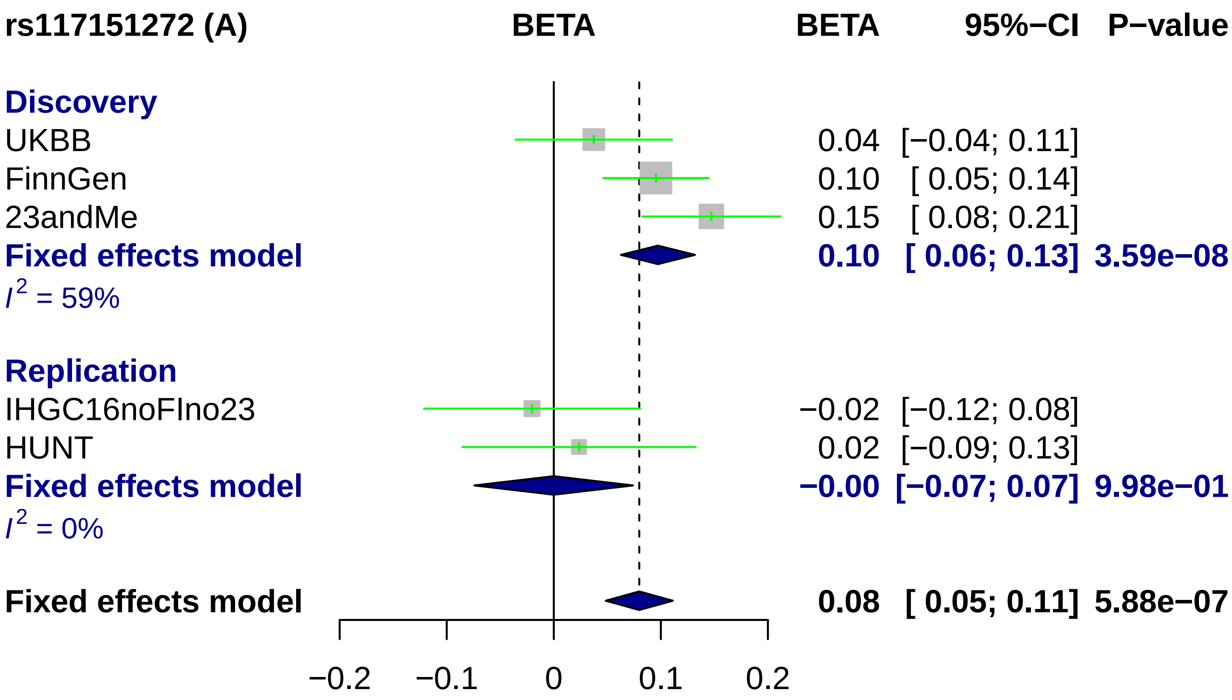
rs117151272 (A)
BETA
BETA
95%−CI
P−value
Discovery
UKBB
0.04
[−0.04; 0.11]
FinnGen
0.10
[ 0.05; 0.14]
23andMe
0.15
[ 0.08; 0.21]
Fixed effects model
0.10
[ 0.06; 0.13]
3.59e−08
I2 = 59%
Replication
IHGC16noFIno23
−0.02
[−0.12; 0.08]
HUNT
0.02
[−0.09; 0.13]
Fixed effects model
−0.00
[−0.07; 0.07]
9.98e−01
I2 = 0%
Fixed effects model
0.08
[ 0.05; 0.11]
5.88e−07
−0.2
−0.1
0
0.1
0.2

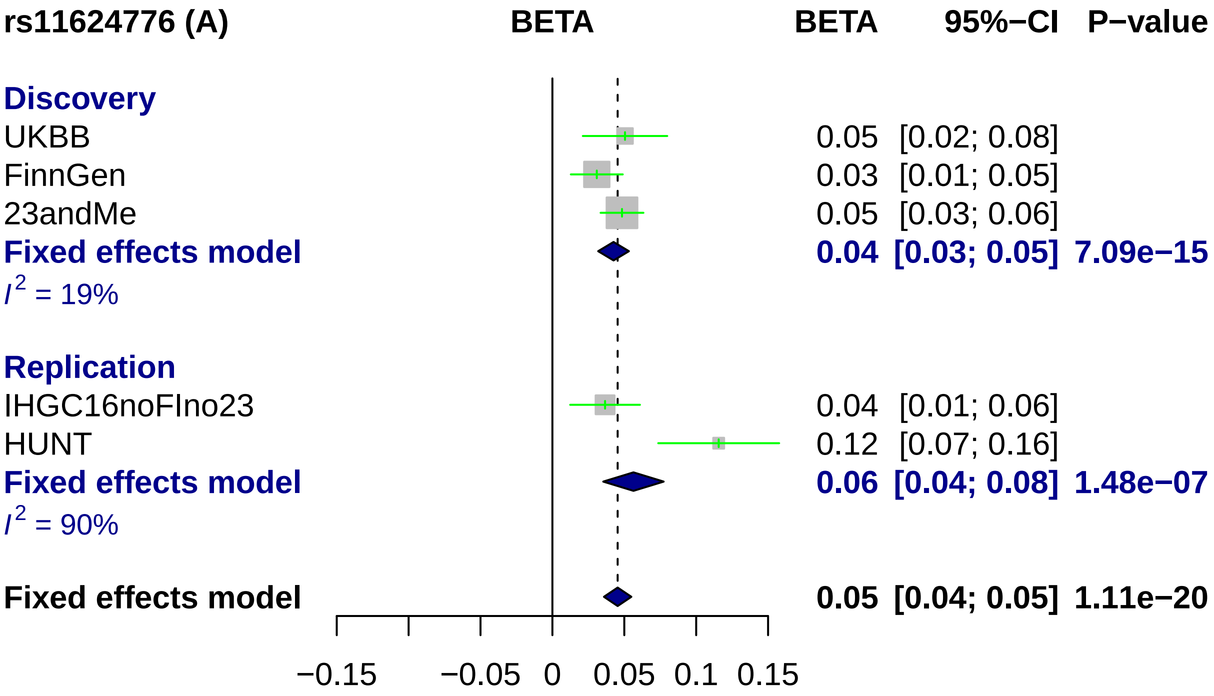
rs11624776 (A)
BETA
BETA
95%−CI
P−value
Discovery
UKBB
0.05 [0.02; 0.08]
FinnGen
0.03 [0.01; 0.05]
23andMe
0.05 [0.03; 0.06]
Fixed effects model
0.04 [0.03; 0.05] 7.09e−15
$I^2$ = 19%
Replication
IHGC16noFIno23
0.04 [0.01; 0.06]
HUNT
0.12 [0.07; 0.16]
Fixed effects model
0.06 [0.04; 0.08] 1.48e−07
$I^2$ = 90%
Fixed effects model
0.05 [0.04; 0.05] 1.11e−20
−0.15
−0.05
0
0.05
0.1
0.15

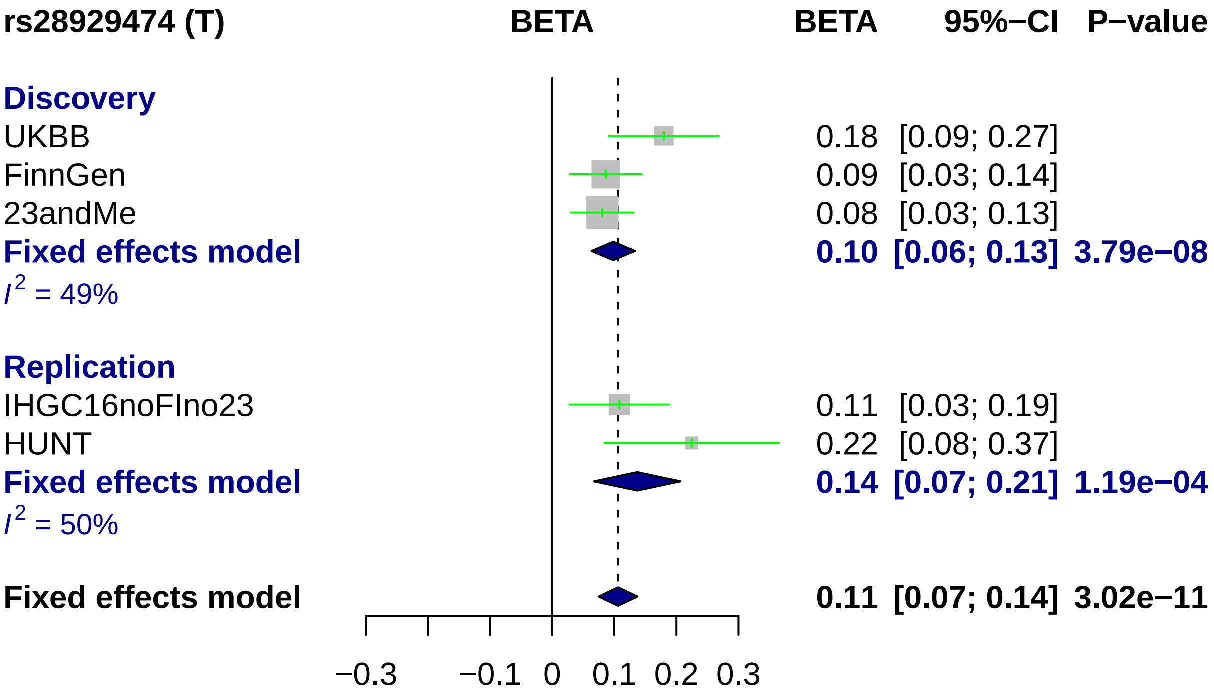
rs28929474 (T)
BETA
BETA
95%−CI
P−value
Discovery
UKBB
0.18 [0.09; 0.27]
FinnGen
0.09 [0.03; 0.14]
23andMe
0.08 [0.03; 0.13]
Fixed effects model
0.10 [0.06; 0.13] 3.79e−08
I2 = 49%
Replication
IHGC16noFIno23
0.11 [0.03; 0.19]
HUNT
0.22 [0.08; 0.37]
Fixed effects model
0.14 [0.07; 0.21] 1.19e−04
I2 = 50%
Fixed effects model
0.11 [0.07; 0.14] 3.02e−11
−0.3
−0.1
0
0.1
0.2
0.3

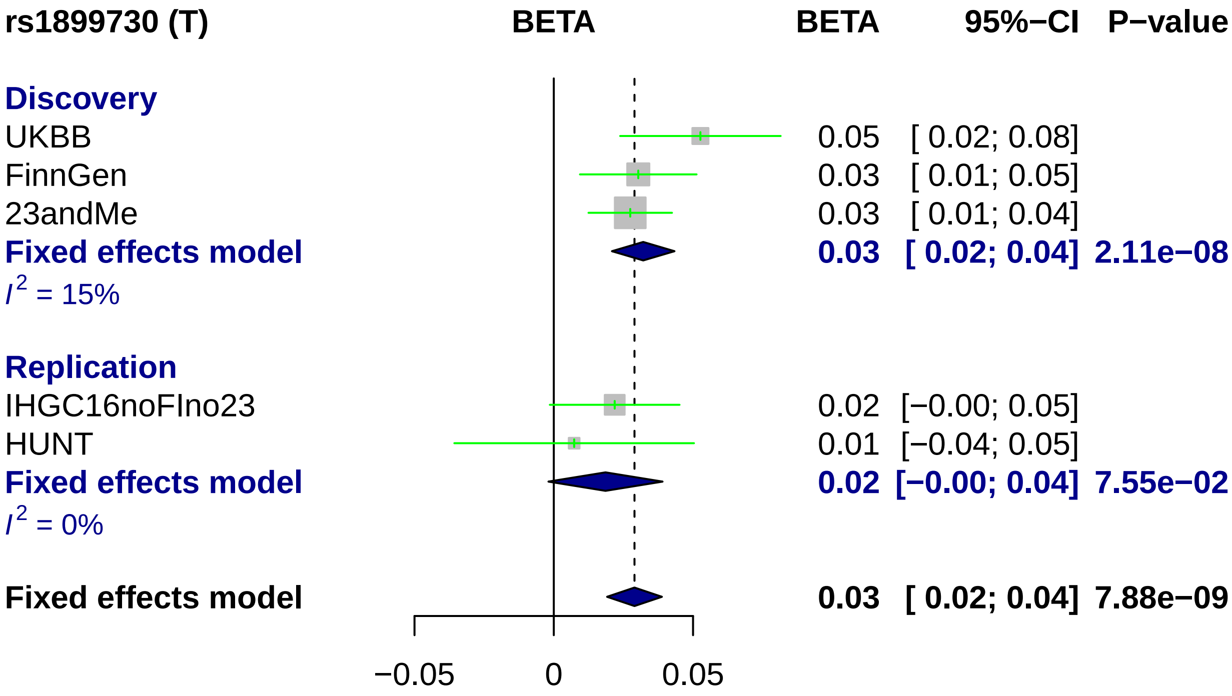

rs1899730 (T)
BETA
BETA
95%−CI
P−value
Discovery
UKBB
0.05
[ 0.02; 0.08]
FinnGen
0.03
[ 0.01; 0.05]
23andMe
0.03
[ 0.01; 0.04]
Fixed effects model
0.03
[ 0.02; 0.04]
2.11e−08
I2 = 15%
Replication
IHGC16noFIno23
0.02
[−0.00; 0.05]
HUNT
0.01
[−0.04; 0.05]
Fixed effects model
0.02
[−0.00; 0.04]
7.55e−02
I2 = 0%
Fixed effects model
0.03
[ 0.02; 0.04]
7.88e−09
−0.05
0
0.05

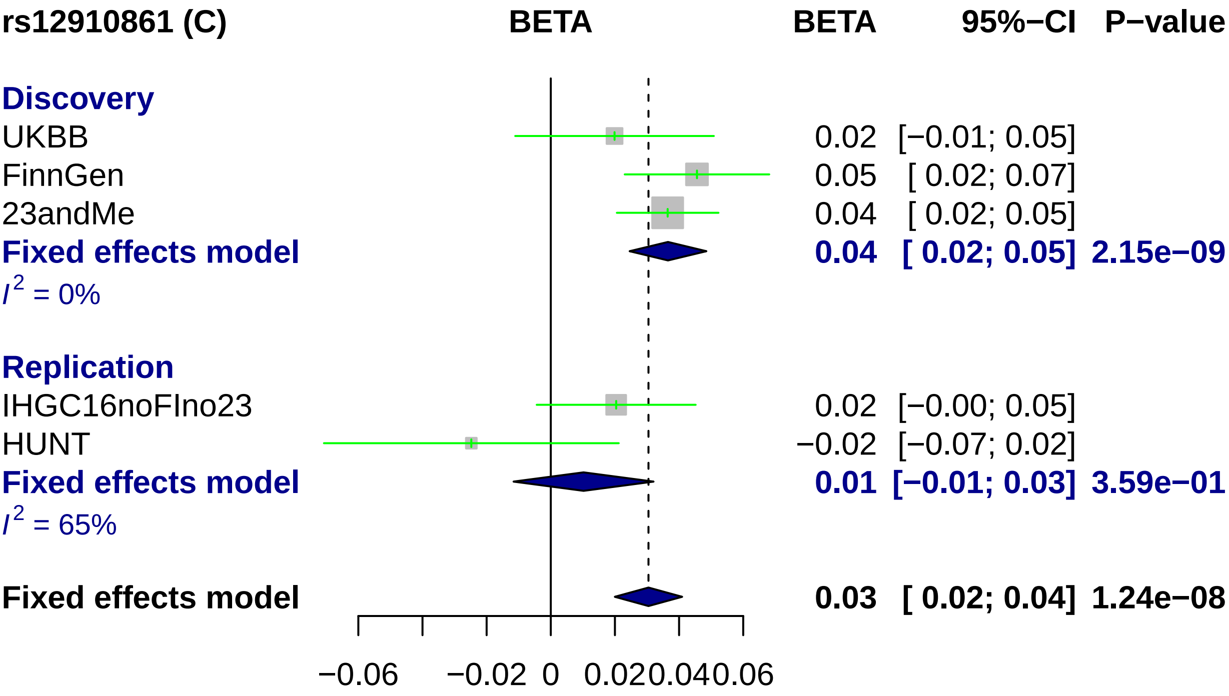
rs12910861 (C)
BETA
BETA
95%−CI
P−value
Discovery
UKBB
0.02 [−0.01; 0.05]
FinnGen
0.05 [ 0.02; 0.07]
23andMe
0.04 [ 0.02; 0.05]
Fixed effects model
0.04 [ 0.02; 0.05] 2.15e−09
I2 = 0%
Replication
IHGC16noFIno23
0.02 [−0.00; 0.05]
HUNT
−0.02 [−0.07; 0.02]
Fixed effects model
0.01 [−0.01; 0.03] 3.59e−01
I2 = 65%
Fixed effects model
0.03 [ 0.02; 0.04] 1.24e−08
−0.06
−0.02
0
0.02
0.04
0.06

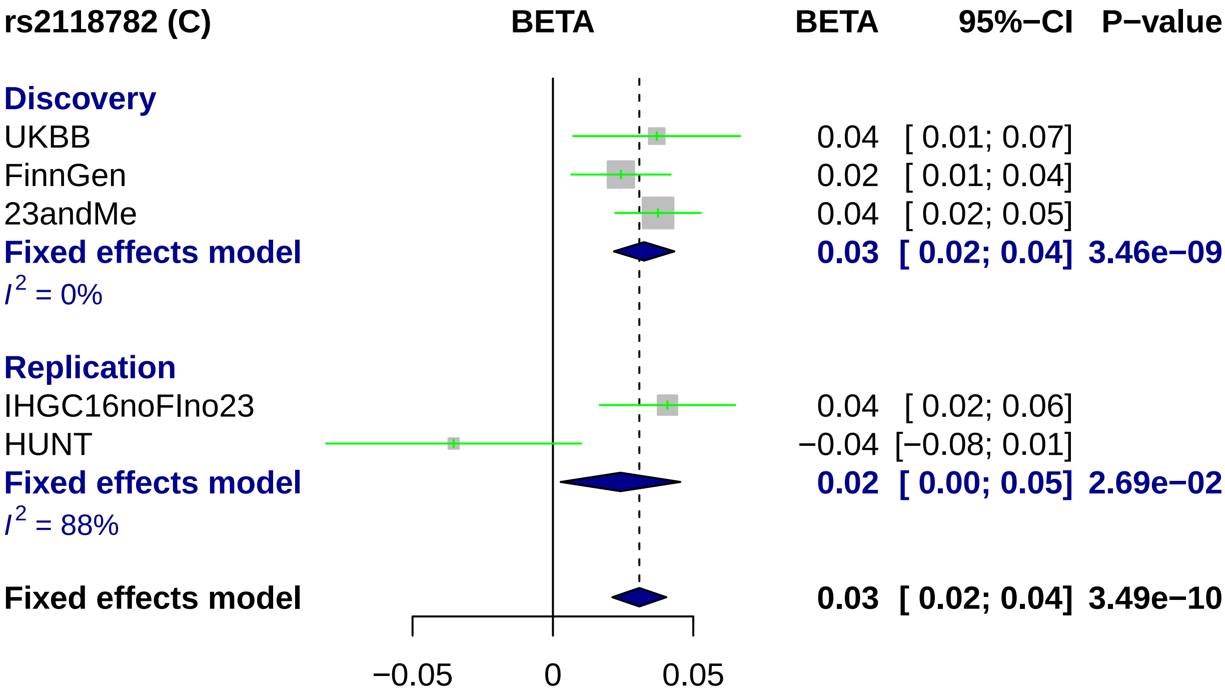
rs2118782 (C)
BETA
BETA
95%−CI
P−value
Discovery
UKBB
0.04 [ 0.01; 0.07]
FinnGen
0.02 [ 0.01; 0.04]
23andMe
0.04 [ 0.02; 0.05]
Fixed effects model
0.03 [ 0.02; 0.04] 3.46e−09
I^2 = 0%
Replication
IHGC16noFIno23
0.04 [ 0.02; 0.06]
HUNT
−0.04 [−0.08; 0.01]
Fixed effects model
0.02 [ 0.00; 0.05] 2.69e−02
I^2 = 88%
Fixed effects model
0.03 [ 0.02; 0.04] 3.49e−10
−0.05
0
0.05

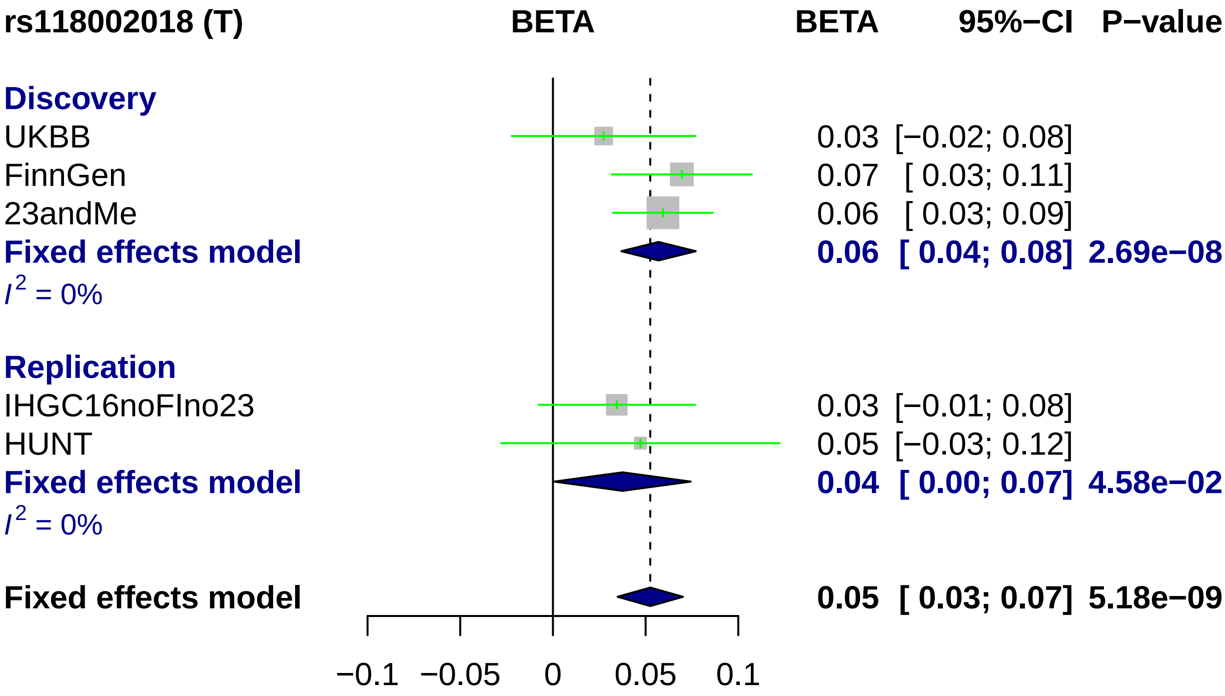
rs118002018 (T)
BETA
BETA
95%−CI
P−value
Discovery
UKBB
0.03 [−0.02; 0.08]
FinnGen
0.07 [ 0.03; 0.11]
23andMe
0.06 [ 0.03; 0.09]
Fixed effects model
0.06 [ 0.04; 0.08] 2.69e−08
I^2 = 0%
Replication
IHGC16noFIno23
0.03 [−0.01; 0.08]
HUNT
0.05 [−0.03; 0.12]
Fixed effects model
0.04 [ 0.00; 0.07] 4.58e−02
I^2 = 0%
Fixed effects model
0.05 [ 0.03; 0.07] 5.18e−09
−0.1
−0.05
0
0.05
0.1

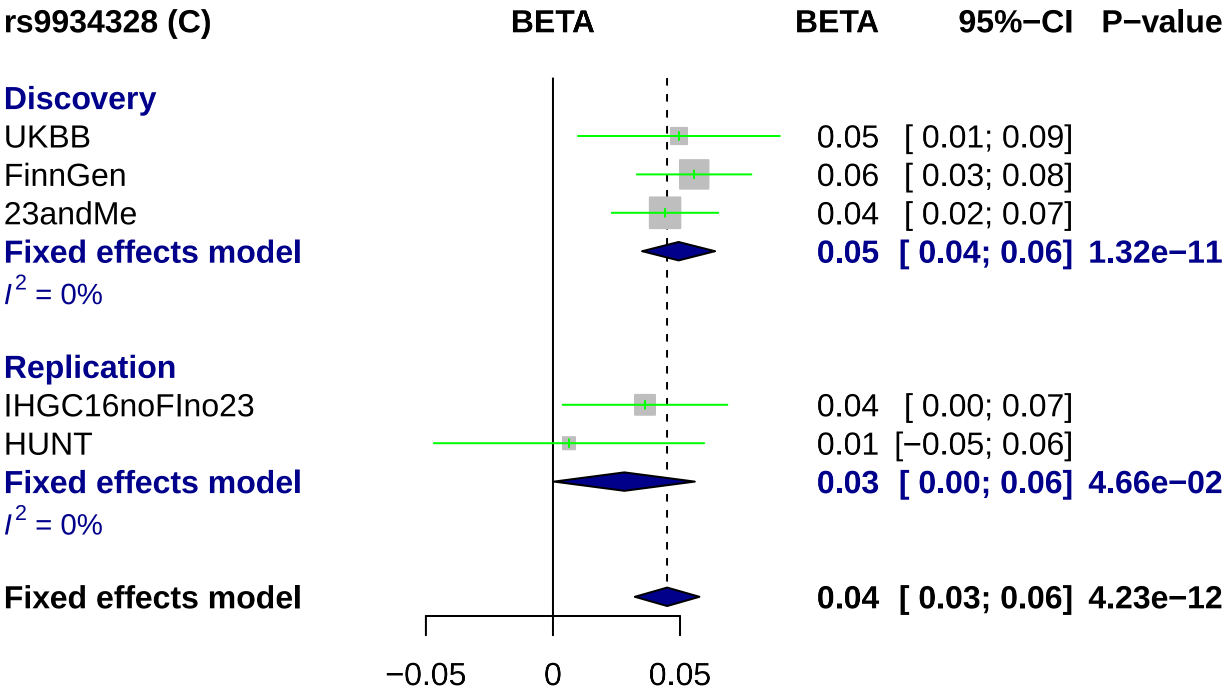
rs9934328 (C)
BETA
BETA
95%−CI
P−value
Discovery
UKBB
0.05 [ 0.01; 0.09]
FinnGen
0.06 [ 0.03; 0.08]
23andMe
0.04 [ 0.02; 0.07]
Fixed effects model
0.05 [ 0.04; 0.06] 1.32e−11
$I^2$ = 0%
Replication
IHGC16noFIno23
0.04 [ 0.00; 0.07]
HUNT
0.01 [−0.05; 0.06]
Fixed effects model
0.03 [ 0.00; 0.06] 4.66e−02
$I^2$ = 0%
Fixed effects model
0.04 [ 0.03; 0.06] 4.23e−12
−0.05
0
0.05

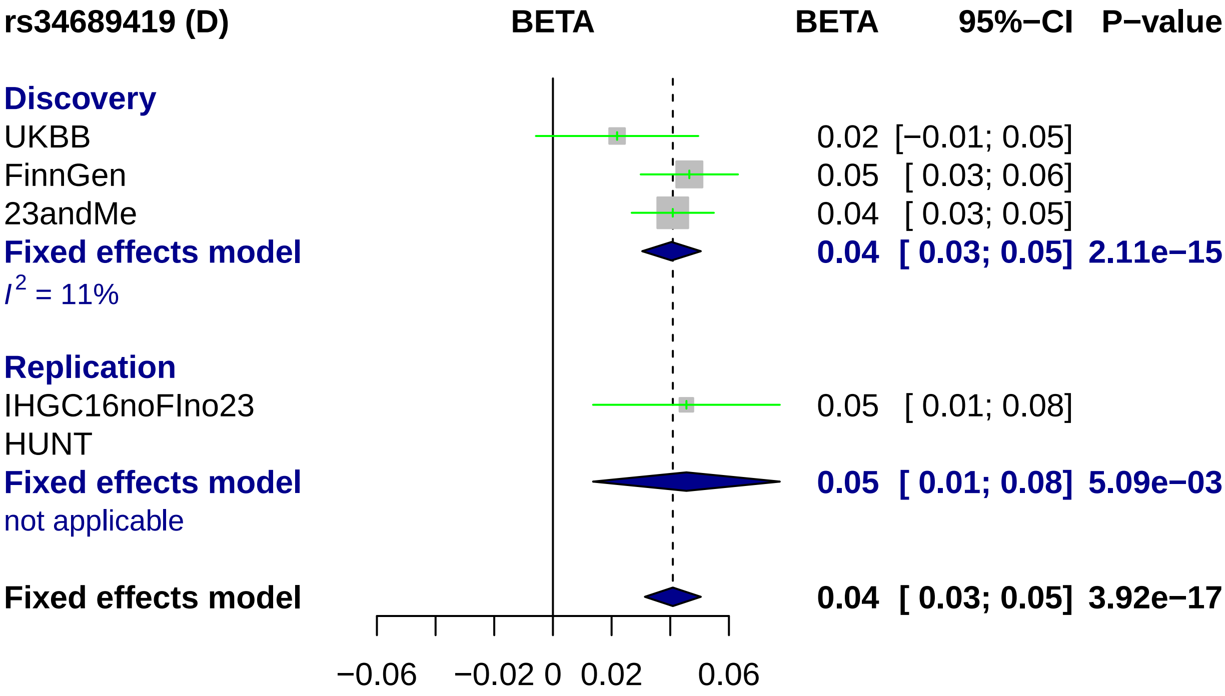
rs34689419 (D)
BETA
BETA
95%−CI
P−value
Discovery
UKBB 0.02 [−0.01; 0.05]
FinnGen 0.05 [ 0.03; 0.06]
23andMe 0.04 [ 0.03; 0.05]
Fixed effects model 0.04 [ 0.03; 0.05] 2.11e−15
I2 = 11%
Replication
IHGC16noFIno23 0.05 [ 0.01; 0.08]
HUNT
Fixed effects model 0.05 [ 0.01; 0.08] 5.09e−03
not applicable
Fixed effects model 0.04 [ 0.03; 0.05] 3.92e−17
−0.06
−0.02
0
0.02
0.06

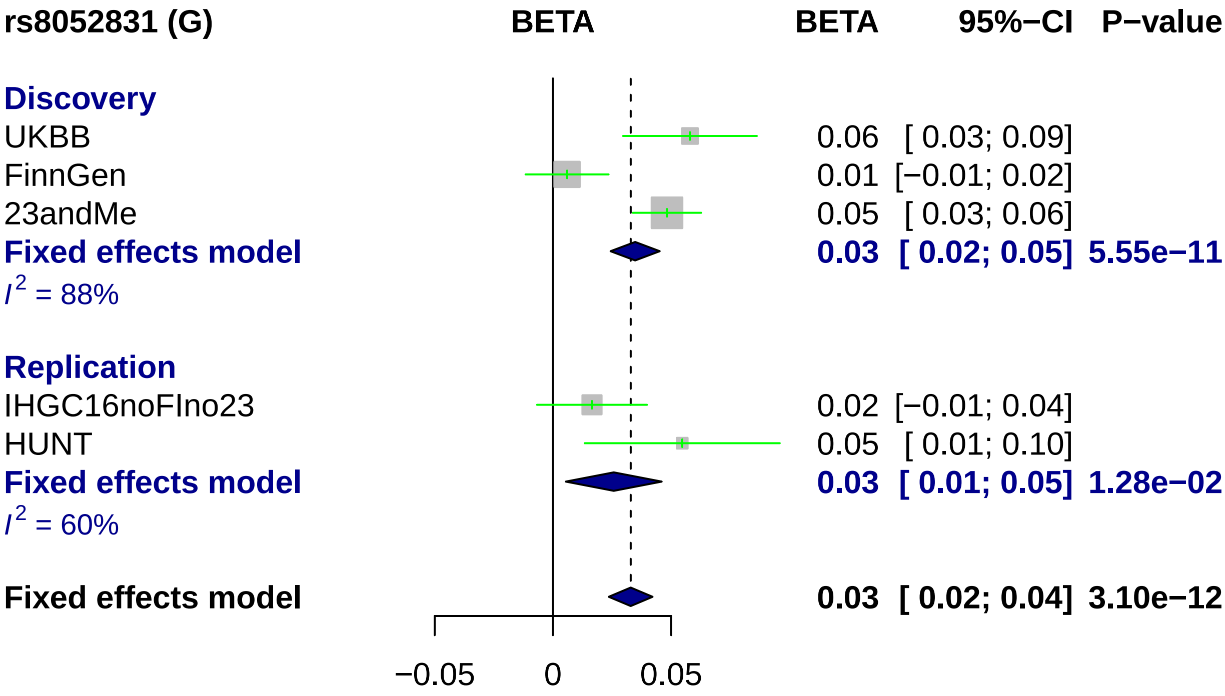
rs8052831 (G)
BETA
BETA
95%−CI
P−value
Discovery
UKBB
0.06
[ 0.03; 0.09]
FinnGen
0.01
[−0.01; 0.02]
23andMe
0.05
[ 0.03; 0.06]
Fixed effects model
0.03
[ 0.02; 0.05]
5.55e−11
$I^2$ = 88%
Replication
IHGC16noFIno23
0.02
[−0.01; 0.04]
HUNT
0.05
[ 0.01; 0.10]
Fixed effects model
0.03
[ 0.01; 0.05]
1.28e−02
$I^2$ = 60%
Fixed effects model
0.03
[ 0.02; 0.04]
3.10e−12
−0.05
0
0.05

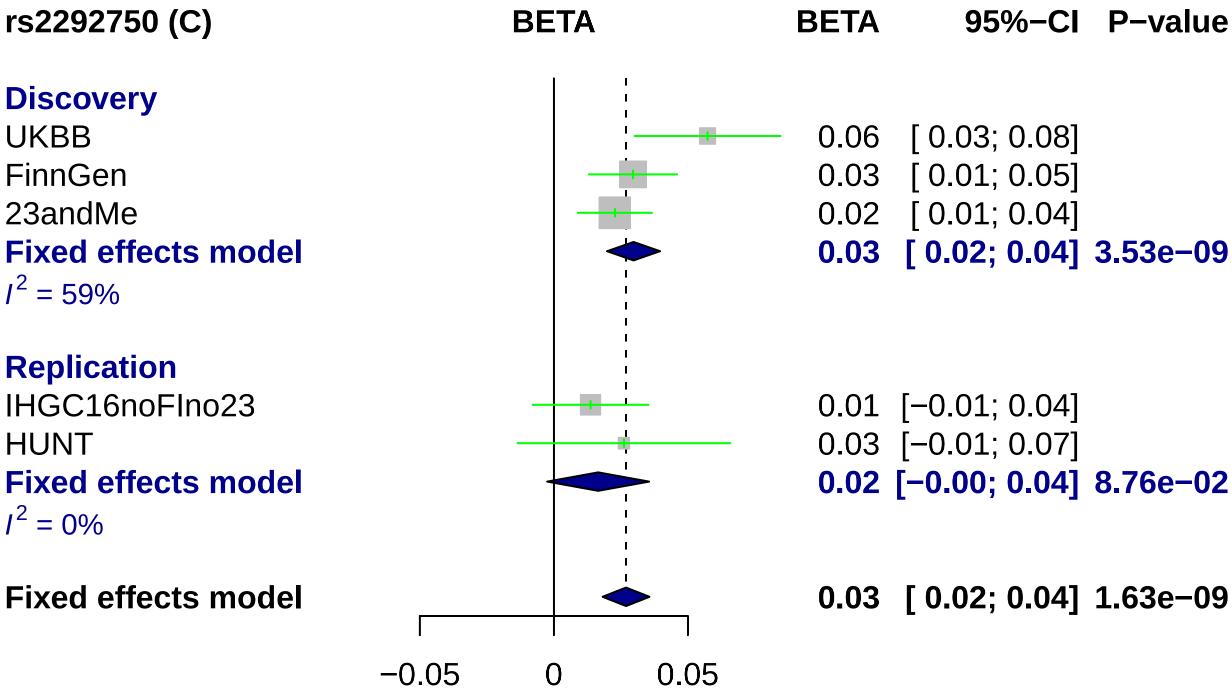
rs2292750 (C)
BETA
BETA
95%−CI
P−value
Discovery
UKBB
0.06
[ 0.03; 0.08]
FinnGen
0.03
[ 0.01; 0.05]
23andMe
0.02
[ 0.01; 0.04]
Fixed effects model
0.03
[ 0.02; 0.04]
3.53e−09
I2 = 59%
Replication
IHGC16noFIno23
0.01
[−0.01; 0.04]
HUNT
0.03
[−0.01; 0.07]
Fixed effects model
0.02
[−0.00; 0.04]
8.76e−02
I2 = 0%
Fixed effects model
0.03
[ 0.02; 0.04]
1.63e−09
−0.05
0
0.05

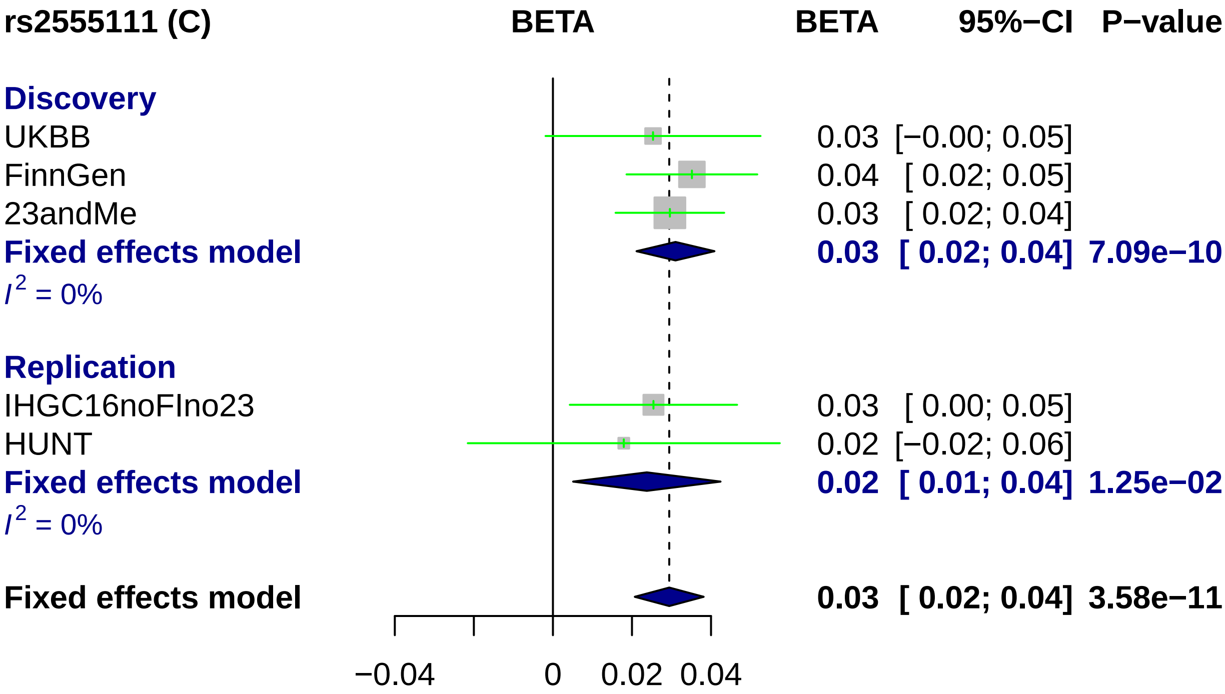
rs2555111 (C)
BETA
BETA 95%−CI P−value
Discovery
UKBB 0.03 [−0.00; 0.05]
FinnGen 0.04 [ 0.02; 0.05]
23andMe 0.03 [ 0.02; 0.04]
Fixed effects model 0.03 [ 0.02; 0.04] 7.09e−10
I2 = 0%
Replication
IHGC16noFIno23 0.03 [ 0.00; 0.05]
HUNT 0.02 [−0.02; 0.06]
Fixed effects model 0.02 [ 0.01; 0.04] 1.25e−02
I2 = 0%
Fixed effects model 0.03 [ 0.02; 0.04] 3.58e−11
−0.04
0
0.02
0.04

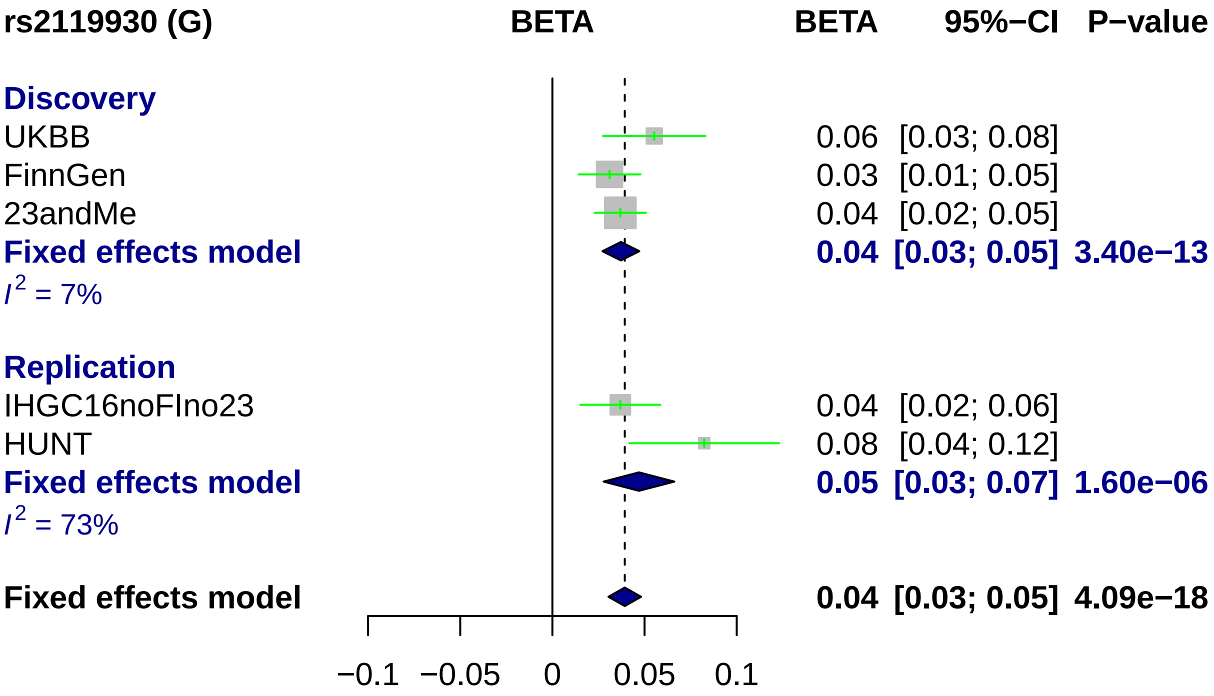

rs2119930 (G)
BETA
BETA
95%−CI
P−value
Discovery
UKBB 0.06 [0.03; 0.08]
FinnGen 0.03 [0.01; 0.05]
23andMe 0.04 [0.02; 0.05]
Fixed effects model 0.04 [0.03; 0.05] 3.40e−13
$I^2$ = 7%
Replication
IHGC16noFIno23 0.04 [0.02; 0.06]
HUNT 0.08 [0.04; 0.12]
Fixed effects model 0.05 [0.03; 0.07] 1.60e−06
$I^2$ = 73%
Fixed effects model 0.04 [0.03; 0.05] 4.09e−18
−0.1
−0.05
0
0.05
0.1

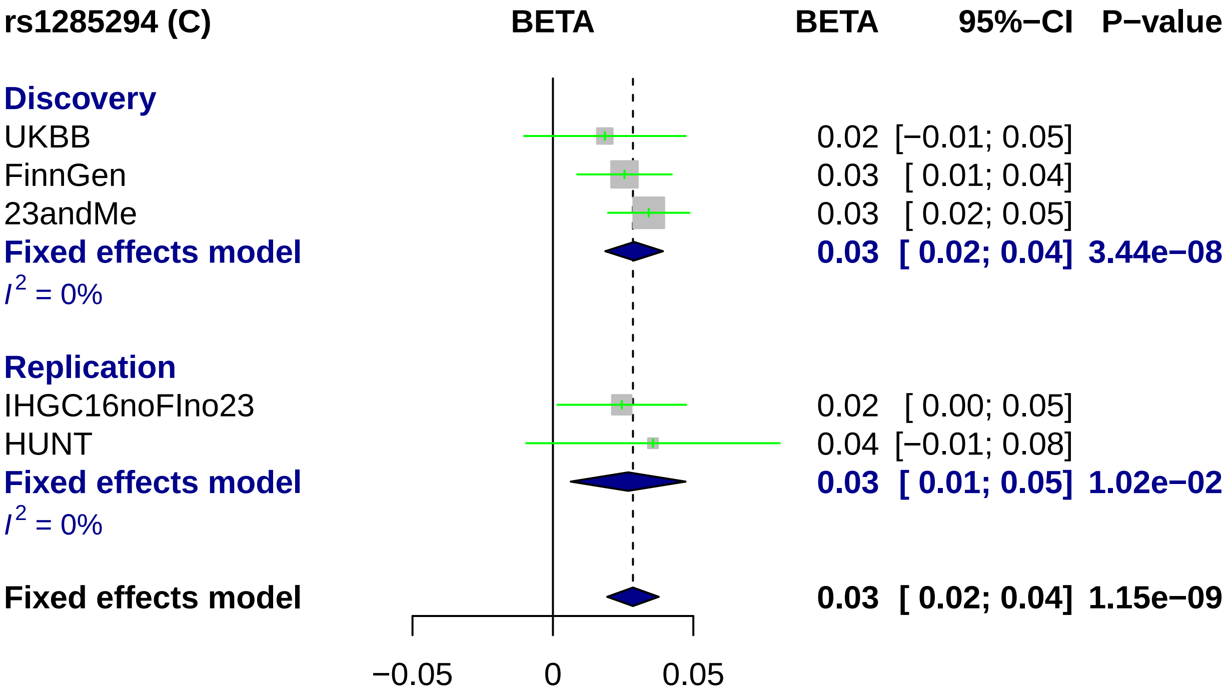
rs1285294 (C)
BETA
BETA
95%−CI
P−value
Discovery
UKBB
0.02
[−0.01; 0.05]
FinnGen
0.03
[ 0.01; 0.04]
23andMe
0.03
[ 0.02; 0.05]
Fixed effects model
0.03
[ 0.02; 0.04]
3.44e−08
I^2 = 0%
Replication
IHGC16noFIno23
0.02
[ 0.00; 0.05]
HUNT
0.04
[−0.01; 0.08]
Fixed effects model
0.03
[ 0.01; 0.05]
1.02e−02
I^2 = 0%
Fixed effects model
0.03
[ 0.02; 0.04]
1.15e−09
−0.05
0
0.05

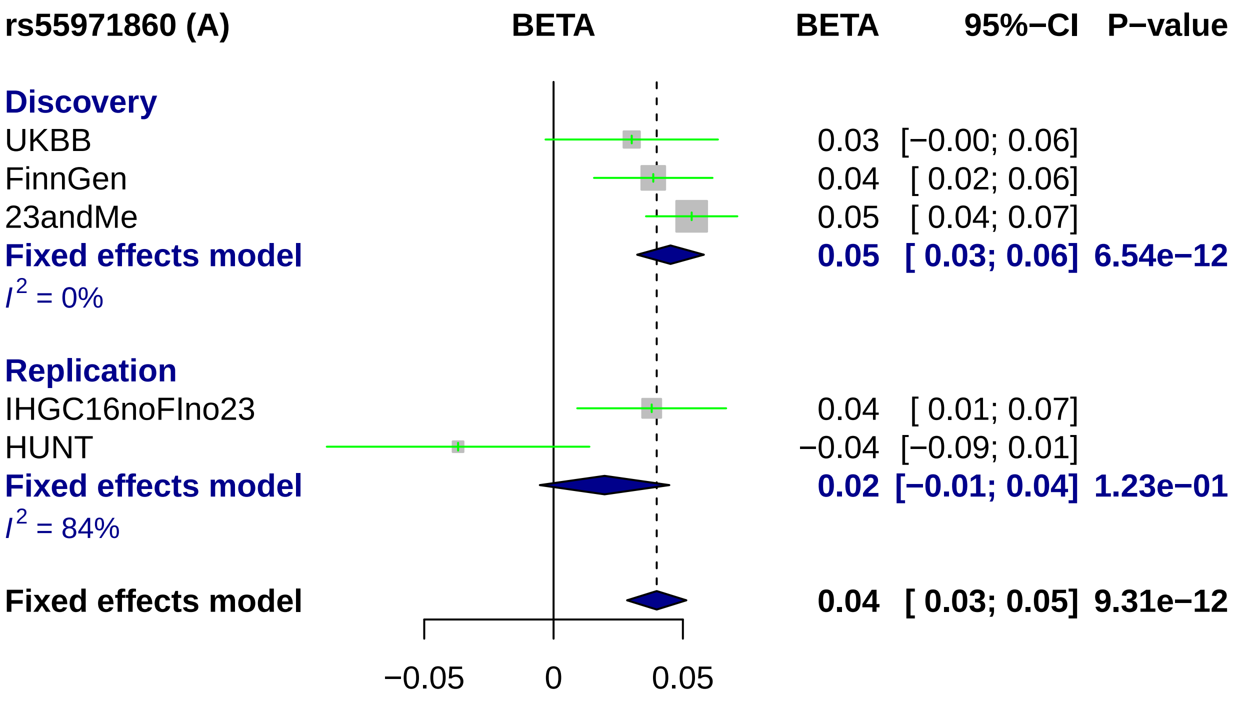
rs55971860 (A)
BETA
BETA
95%−CI
P−value
Discovery
UKBB
0.03 [−0.00; 0.06]
FinnGen
0.04 [ 0.02; 0.06]
23andMe
0.05 [ 0.04; 0.07]
Fixed effects model
0.05 [ 0.03; 0.06] 6.54e−12
I2 = 0%
Replication
IHGC16noFIno23
0.04 [ 0.01; 0.07]
HUNT
−0.04 [−0.09; 0.01]
Fixed effects model
0.02 [−0.01; 0.04] 1.23e−01
I2 = 84%
Fixed effects model
0.04 [ 0.03; 0.05] 9.31e−12
−0.05
0
0.05

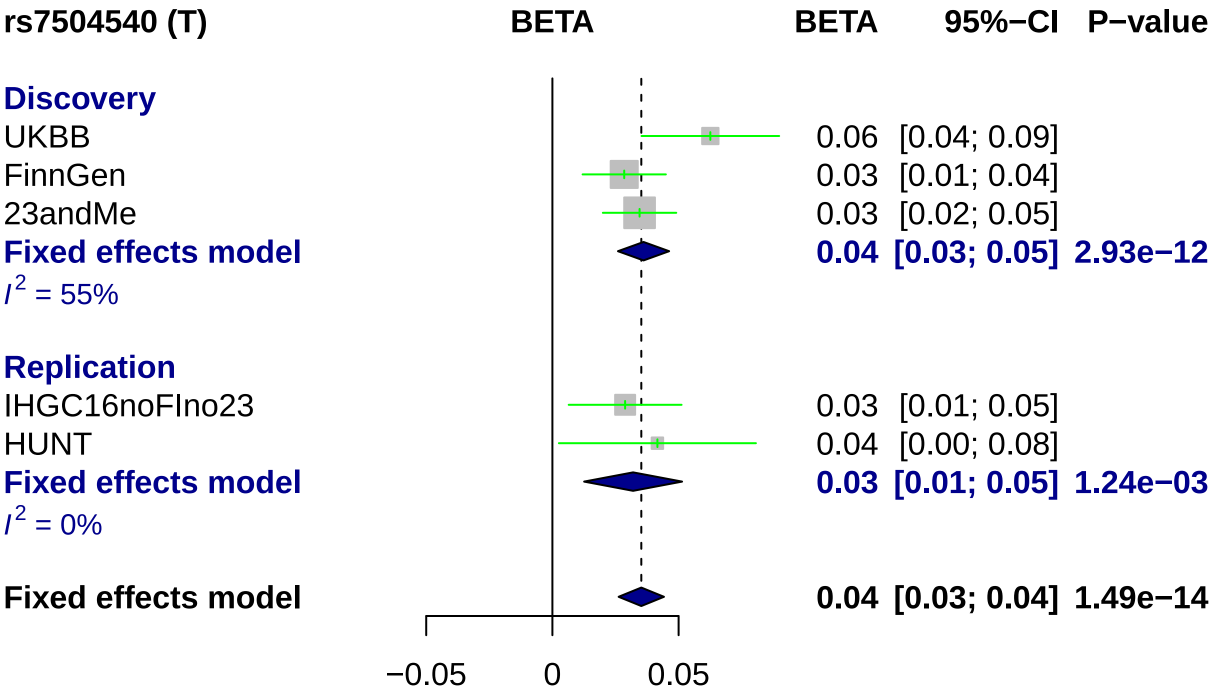
rs7504540 (T)
BETA
BETA
95%−CI
P−value
Discovery
UKBB
0.06
[0.04; 0.09]
FinnGen
0.03
[0.01; 0.04]
23andMe
0.03
[0.02; 0.05]
Fixed effects model
0.04
[0.03; 0.05]
2.93e−12
$I^2$ = 55%
Replication
IHGC16noFIno23
0.03
[0.01; 0.05]
HUNT
0.04
[0.00; 0.08]
Fixed effects model
0.03
[0.01; 0.05]
1.24e−03
$I^2$ = 0%
Fixed effects model
0.04
[0.03; 0.04]
1.49e−14
−0.05
0
0.05

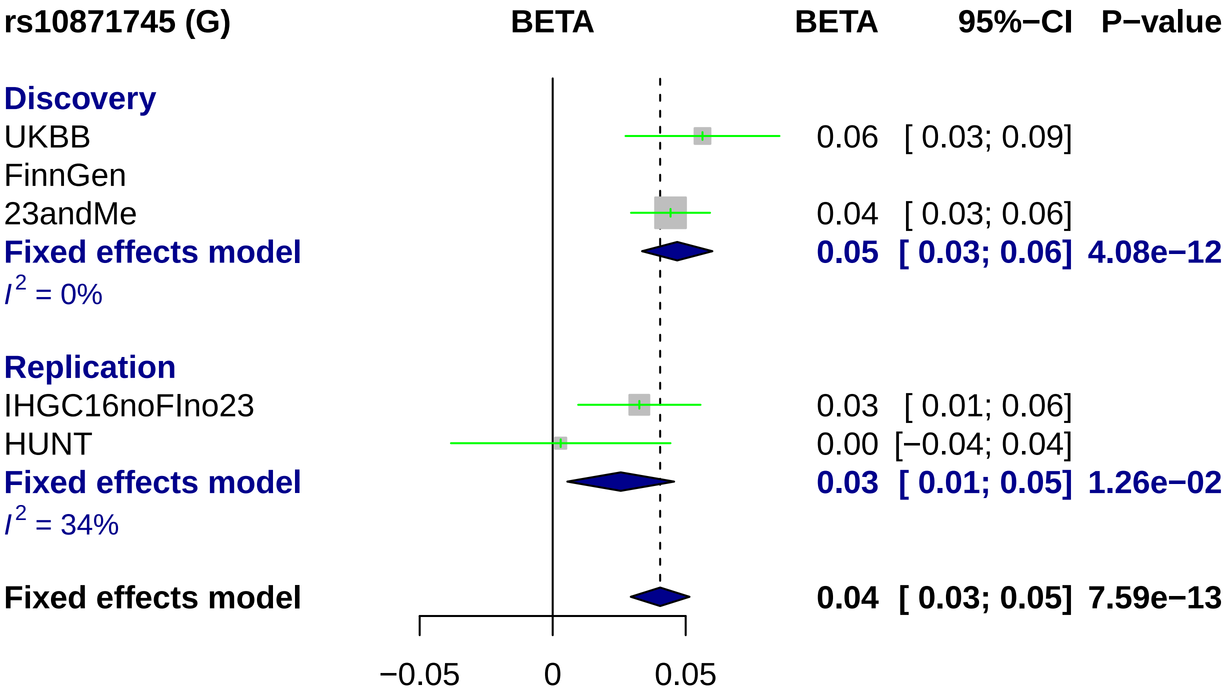

rs10871745 (G)
BETA
BETA
95%−CI
P−value
Discovery
UKBB
0.06 [ 0.03; 0.09]
FinnGen
23andMe
0.04 [ 0.03; 0.06]
Fixed effects model
0.05 [ 0.03; 0.06] 4.08e−12
I2 = 0%
Replication
IHGC16noFIno23
0.03 [ 0.01; 0.06]
HUNT
0.00 [−0.04; 0.04]
Fixed effects model
0.03 [ 0.01; 0.05] 1.26e−02
I2 = 34%
Fixed effects model
0.04 [ 0.03; 0.05] 7.59e−13
−0.05
0
0.05

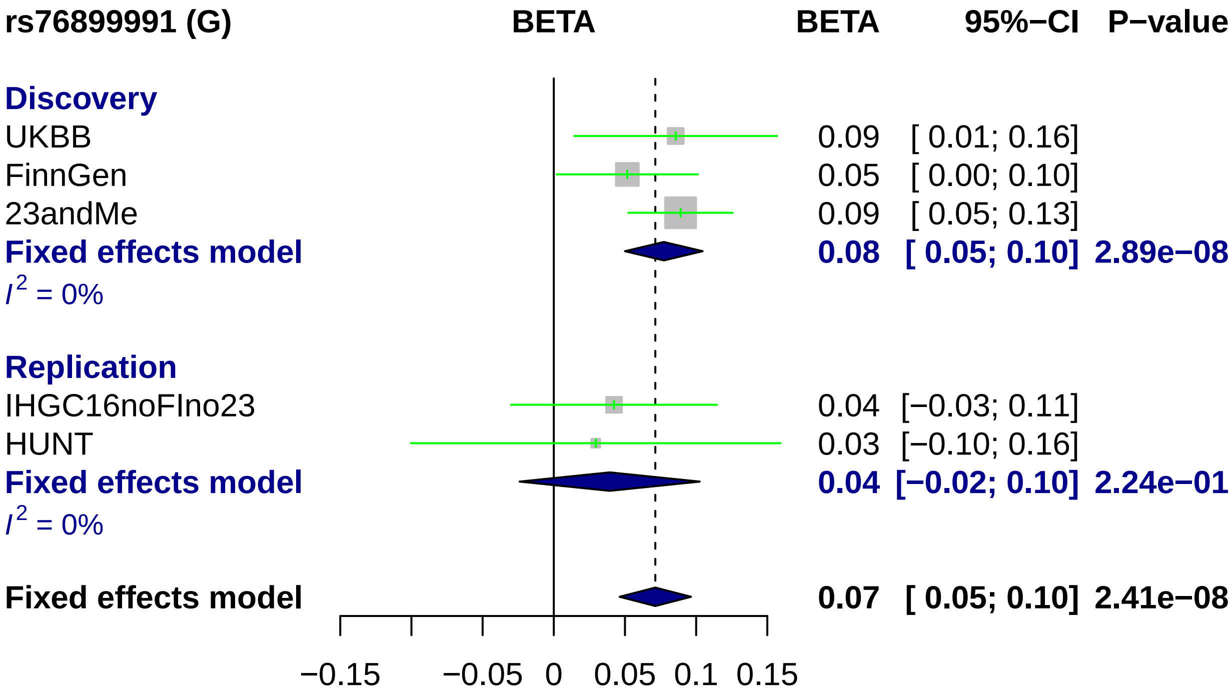
rs76899991 (G)
BETA
BETA
95%−CI
P−value
Discovery
UKBB
0.09
[ 0.01; 0.16]
FinnGen
0.05
[ 0.00; 0.10]
23andMe
0.09
[ 0.05; 0.13]
Fixed effects model
0.08
[ 0.05; 0.10]
2.89e−08
$I^2$ = 0%
Replication
IHGC16noFIno23
0.04
[−0.03; 0.11]
HUNT
0.03
[−0.10; 0.16]
Fixed effects model
0.04
[−0.02; 0.10]
2.24e−01
$I^2$ = 0%
Fixed effects model
0.07
[ 0.05; 0.10]
2.41e−08
−0.15
−0.05
0
0.05
0.1
0.15

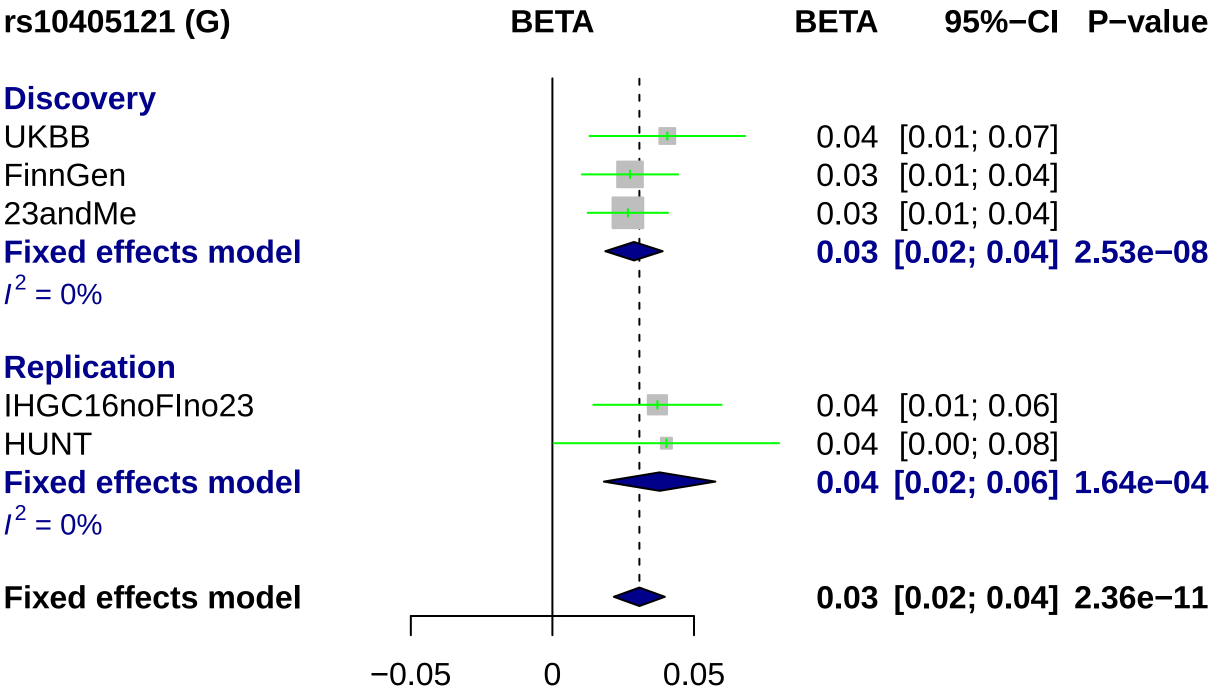

rs10405121 (G)
BETA
BETA
95%−CI
P−value
Discovery
UKBB
0.04 [0.01; 0.07]
FinnGen
0.03 [0.01; 0.04]
23andMe
0.03 [0.01; 0.04]
Fixed effects model
0.03 [0.02; 0.04] 2.53e−08
I2 = 0%
Replication
IHGC16noFIno23
0.04 [0.01; 0.06]
HUNT
0.04 [0.00; 0.08]
Fixed effects model
0.04 [0.02; 0.06] 1.64e−04
I2 = 0%
Fixed effects model
0.03 [0.02; 0.04] 2.36e−11
−0.05
0
0.05

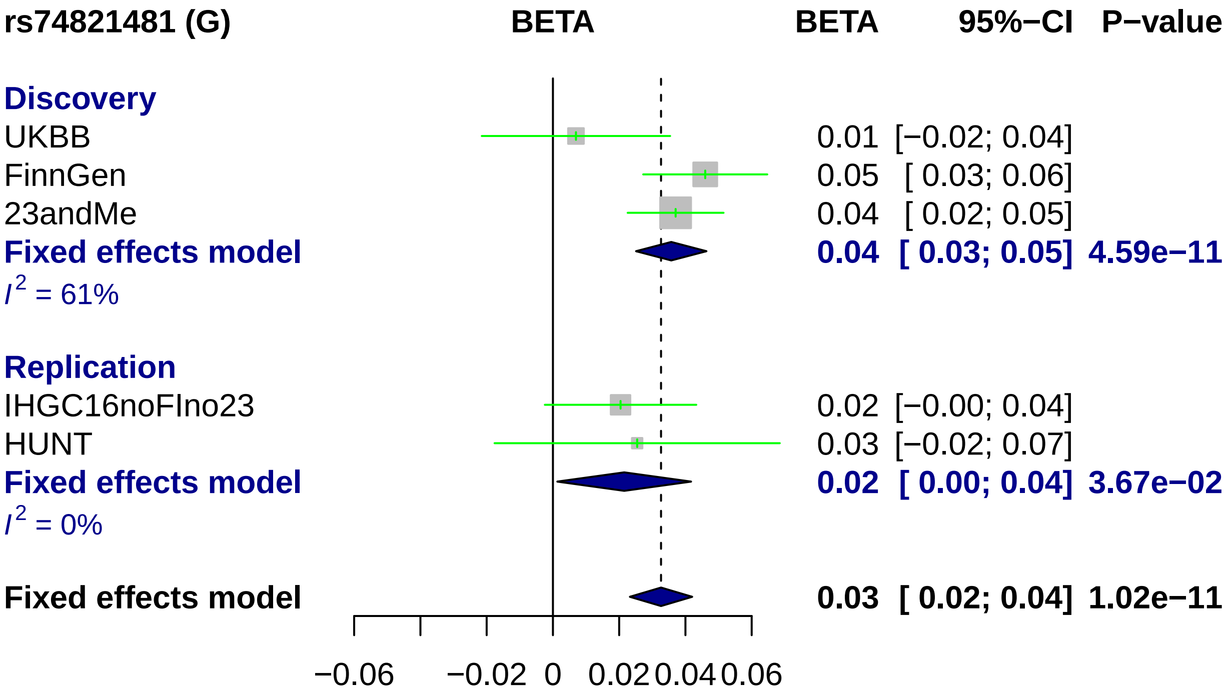

rs74821481 (G)
BETA
BETA
95%−CI
P−value
Discovery
UKBB
0.01 [−0.02; 0.04]
FinnGen
0.05 [ 0.03; 0.06]
23andMe
0.04 [ 0.02; 0.05]
Fixed effects model
0.04 [ 0.03; 0.05] 4.59e−11
$I^2$ = 61%
Replication
IHGC16noFIno23
0.02 [−0.00; 0.04]
HUNT
0.03 [−0.02; 0.07]
Fixed effects model
0.02 [ 0.00; 0.04] 3.67e−02
$I^2$ = 0%
Fixed effects model
0.03 [ 0.02; 0.04] 1.02e−11
−0.06
−0.02
0
0.02
0.04
0.06

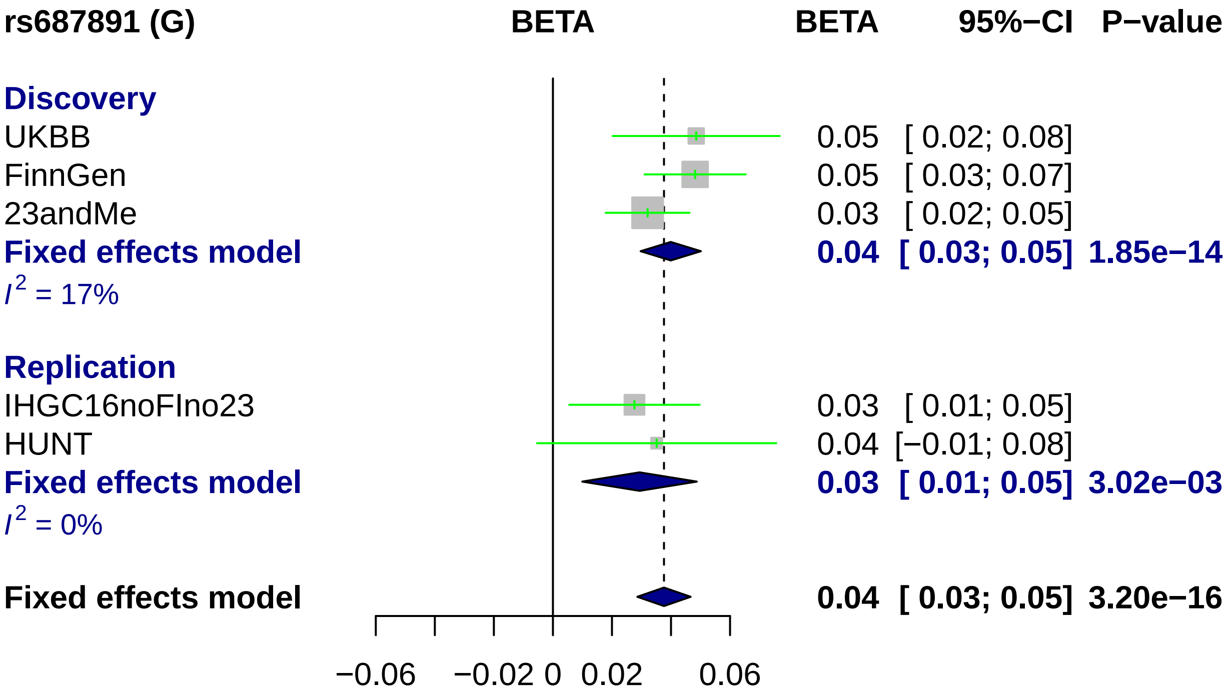
rs687891 (G)
BETA
BETA
95%−CI
P−value
Discovery
UKBB
0.05 [ 0.02; 0.08]
FinnGen
0.05 [ 0.03; 0.07]
23andMe
0.03 [ 0.02; 0.05]
Fixed effects model
0.04 [ 0.03; 0.05] 1.85e−14
I2 = 17%
Replication
IHGC16noFIno23
0.03 [ 0.01; 0.05]
HUNT
0.04 [−0.01; 0.08]
Fixed effects model
0.03 [ 0.01; 0.05] 3.02e−03
I2 = 0%
Fixed effects model
0.04 [ 0.03; 0.05] 3.20e−16
−0.06
−0.02
0
0.02
0.06

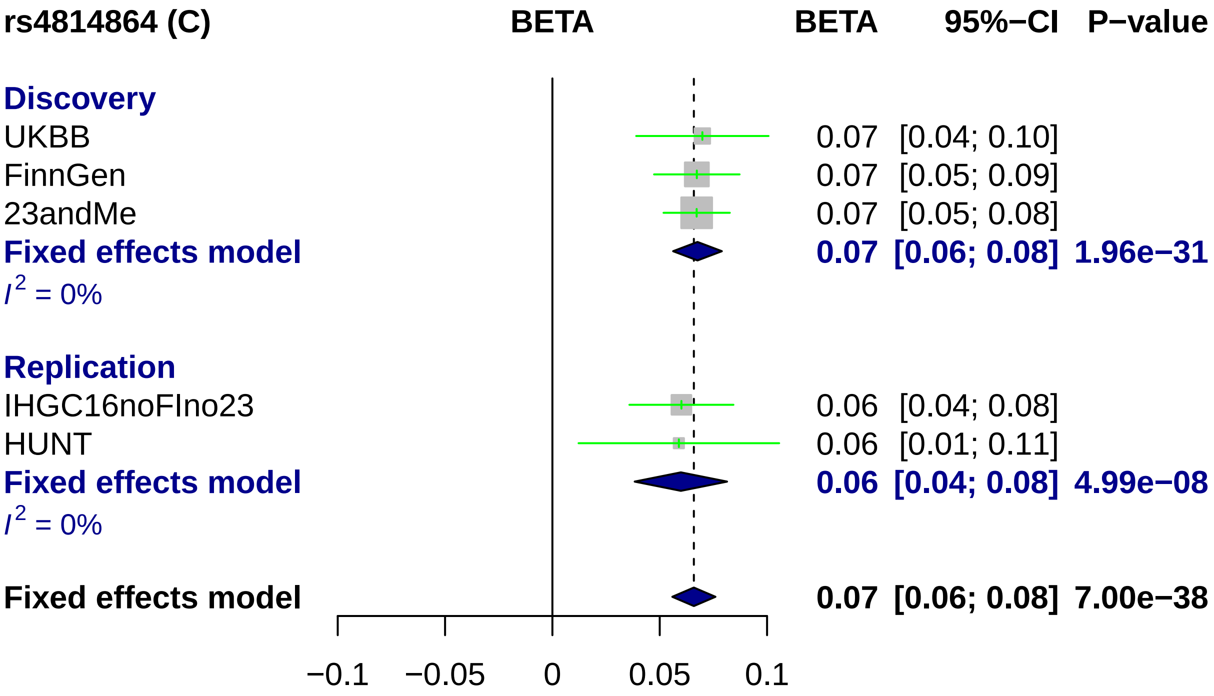
rs4814864 (C)
BETA
BETA
95%−CI
P−value
Discovery
UKBB
0.07 [0.04; 0.10]
FinnGen
0.07 [0.05; 0.09]
23andMe
0.07 [0.05; 0.08]
Fixed effects model
0.07 [0.06; 0.08] 1.96e−31
I^2 = 0%
Replication
IHGC16noFIno23
0.06 [0.04; 0.08]
HUNT
0.06 [0.01; 0.11]
Fixed effects model
0.06 [0.04; 0.08] 4.99e−08
I^2 = 0%
Fixed effects model
0.07 [0.06; 0.08] 7.00e−38
−0.1
−0.05
0
0.05
0.1

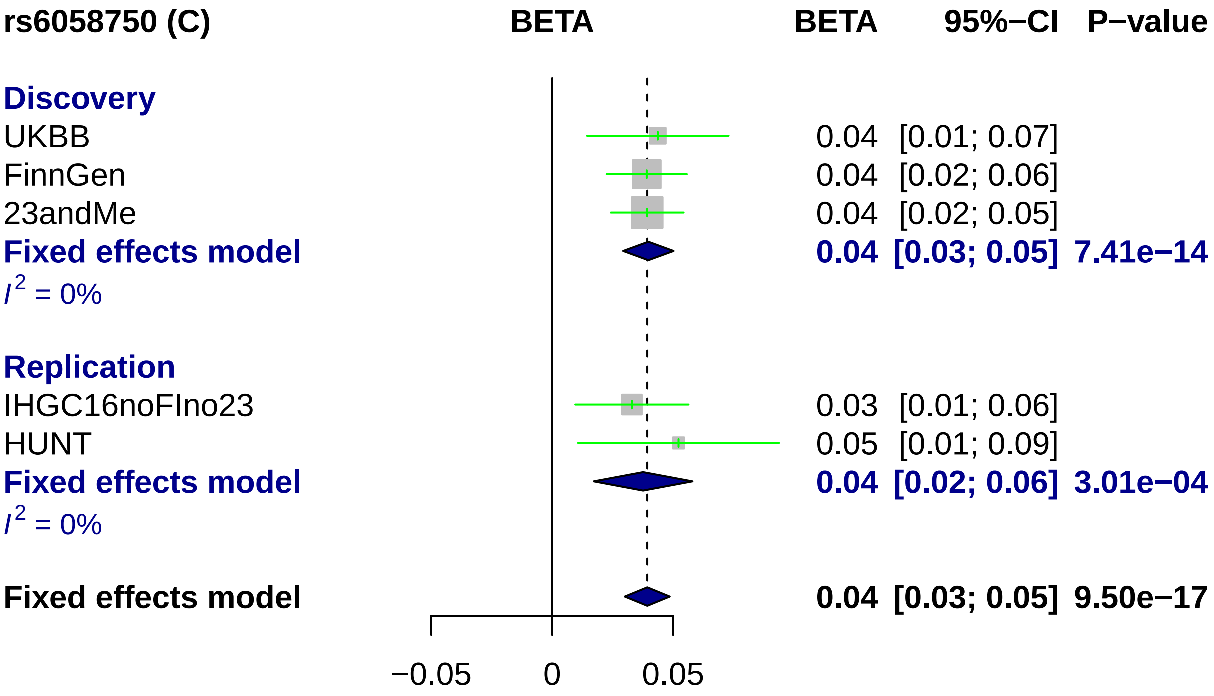
rs6058750 (C)
BETA
BETA
95%−CI
P−value
Discovery
UKBB
0.04 [0.01; 0.07]
FinnGen
0.04 [0.02; 0.06]
23andMe
0.04 [0.02; 0.05]
Fixed effects model
0.04 [0.03; 0.05] 7.41e−14
I2 = 0%
Replication
IHGC16noFIno23
0.03 [0.01; 0.06]
HUNT
0.05 [0.01; 0.09]
Fixed effects model
0.04 [0.02; 0.06] 3.01e−04
I2 = 0%
Fixed effects model
0.04 [0.03; 0.05] 9.50e−17
−0.05
0
0.05

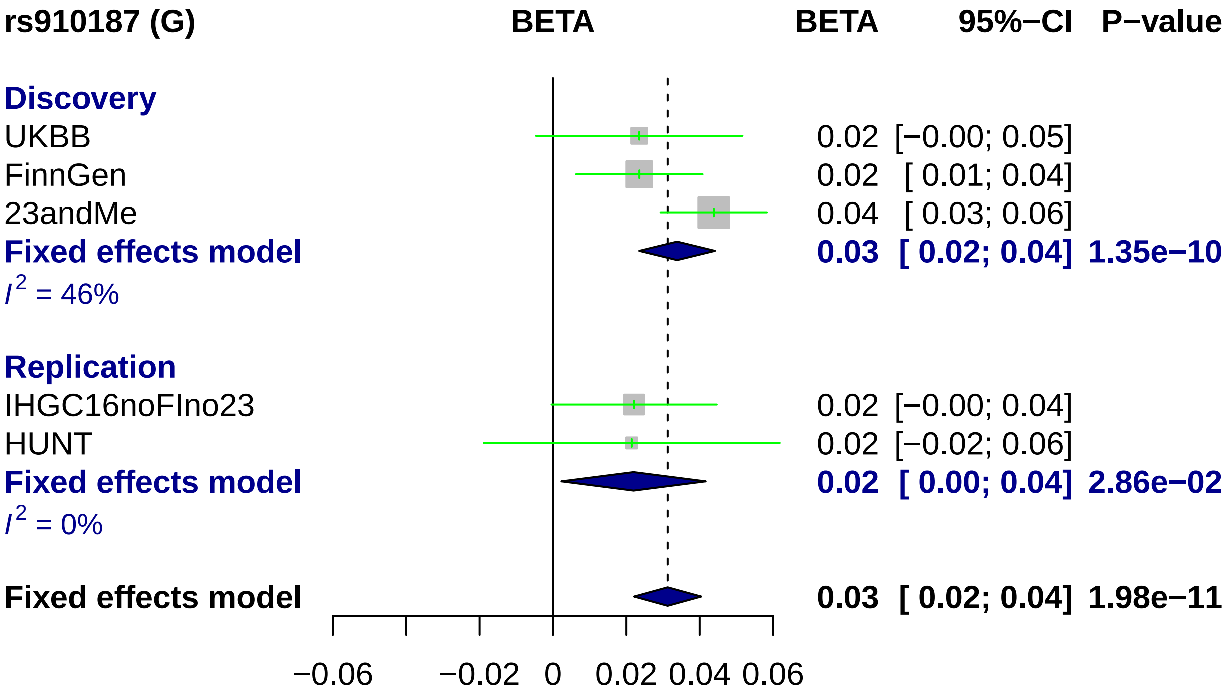
rs910187 (G)
BETA
BETA
95%−CI
P−value
Discovery
UKBB
0.02 [−0.00; 0.05]
FinnGen
0.02 [ 0.01; 0.04]
23andMe
0.04 [ 0.03; 0.06]
Fixed effects model
0.03 [ 0.02; 0.04] 1.35e−10
I2 = 46%
Replication
IHGC16noFIno23
0.02 [−0.00; 0.04]
HUNT
0.02 [−0.02; 0.06]
Fixed effects model
0.02 [ 0.00; 0.04] 2.86e−02
I2 = 0%
Fixed effects model
0.03 [ 0.02; 0.04] 1.98e−11
−0.06
−0.02
0
0.02
0.04
0.06

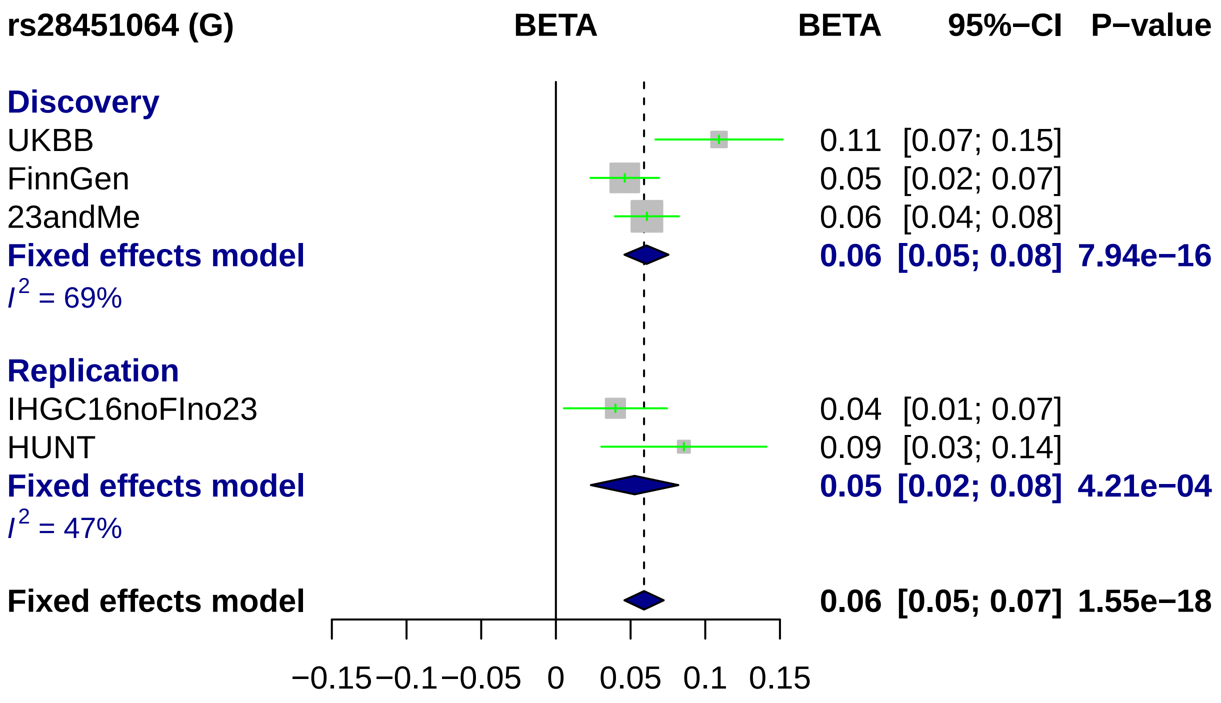
rs28451064 (G)
BETA
BETA
95%−CI
P−value
Discovery
UKBB
0.11 [0.07; 0.15]
FinnGen
0.05 [0.02; 0.07]
23andMe
0.06 [0.04; 0.08]
Fixed effects model
0.06 [0.05; 0.08] 7.94e−16
I2 = 69%
Replication
IHGC16noFIno23
0.04 [0.01; 0.07]
HUNT
0.09 [0.03; 0.14]
Fixed effects model
0.05 [0.02; 0.08] 4.21e−04
I2 = 47%
Fixed effects model
0.06 [0.05; 0.07] 1.55e−18
−0.15
−0.1
−0.05
0
0.05
0.1
0.15

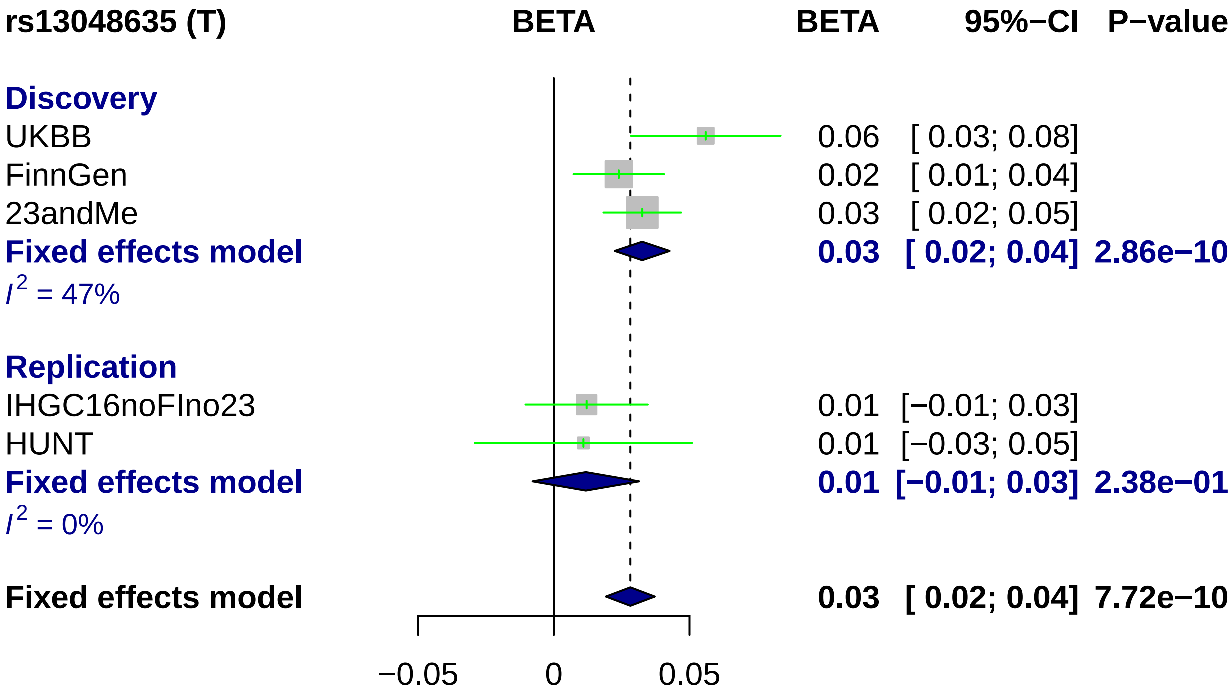
rs13048635 (T)
BETA
BETA
95%−CI
P−value
Discovery
UKBB
0.06
[ 0.03; 0.08]
FinnGen
0.02
[ 0.01; 0.04]
23andMe
0.03
[ 0.02; 0.05]
Fixed effects model
0.03
[ 0.02; 0.04]
2.86e−10
I2 = 47%
Replication
IHGC16noFIno23
0.01
[−0.01; 0.03]
HUNT
0.01
[−0.03; 0.05]
Fixed effects model
0.01
[−0.01; 0.03]
2.38e−01
I2 = 0%
Fixed effects model
0.03
[ 0.02; 0.04]
7.72e−10
−0.05
0
0.05

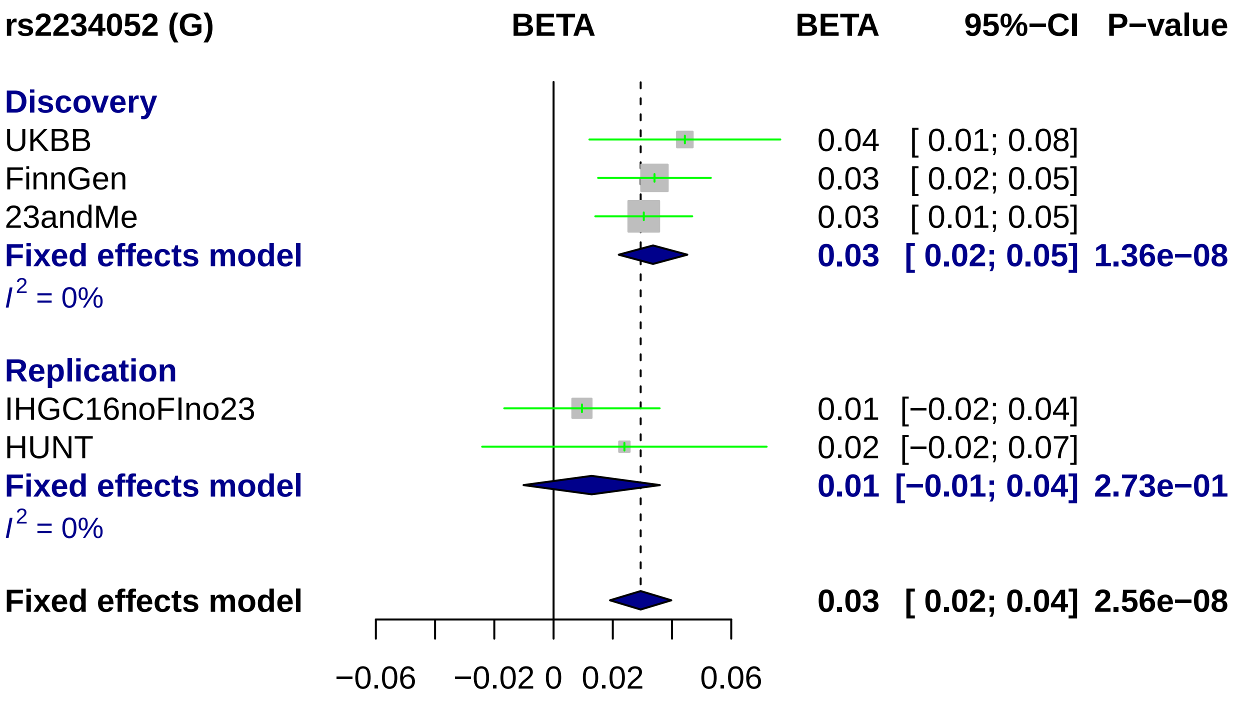

rs2234052 (G)
BETA
BETA
95%−CI
P−value
Discovery
UKBB
0.04
[ 0.01; 0.08]
FinnGen
0.03
[ 0.02; 0.05]
23andMe
0.03
[ 0.01; 0.05]
Fixed effects model
0.03
[ 0.02; 0.05]
1.36e−08
I2 = 0%
Replication
IHGC16noFIno23
0.01
[−0.02; 0.04]
HUNT
0.02
[−0.02; 0.07]
Fixed effects model
0.01
[−0.01; 0.04]
2.73e−01
I2 = 0%
Fixed effects model
0.03
[ 0.02; 0.04]
2.56e−08
−0.06
−0.02
0
0.02
0.06

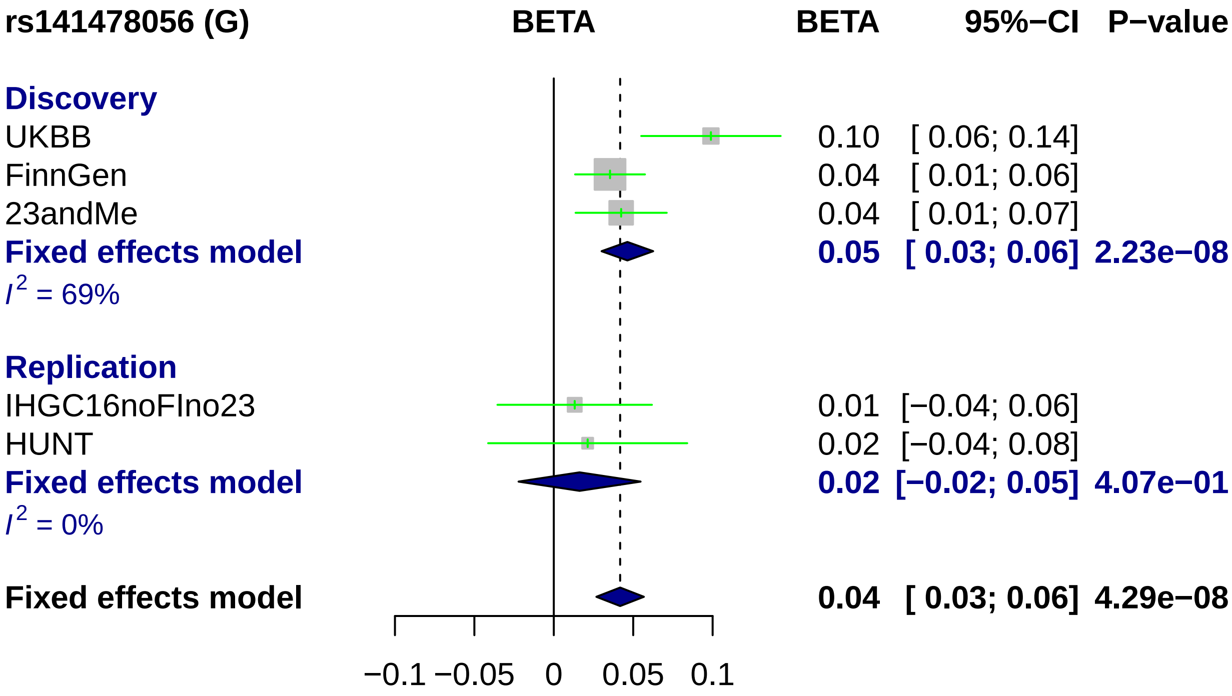
rs141478056 (G)
BETA
BETA
95%−CI
P−value
Discovery
UKBB
0.10
[ 0.06; 0.14]
FinnGen
0.04
[ 0.01; 0.06]
23andMe
0.04
[ 0.01; 0.07]
Fixed effects model
0.05
[ 0.03; 0.06]
2.23e−08
I2 = 69%
Replication
IHGC16noFIno23
0.01
[−0.04; 0.06]
HUNT
0.02
[−0.04; 0.08]
Fixed effects model
0.02
[−0.02; 0.05]
4.07e−01
I2 = 0%
Fixed effects model
0.04
[ 0.03; 0.06]
4.29e−08
−0.1
−0.05
0
0.05
0.1

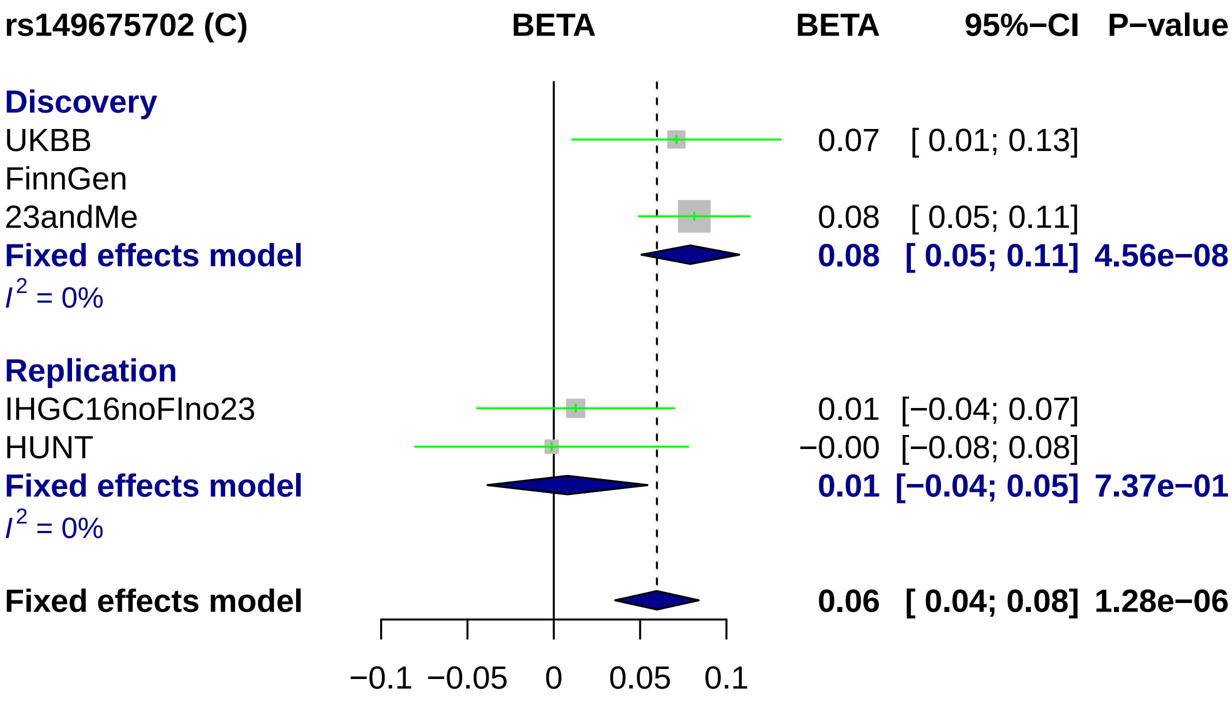
rs149675702 (C)
BETA
BETA
95%−CI
P−value
Discovery
UKBB
0.07
[ 0.01; 0.13]
FinnGen
23andMe
0.08
[ 0.05; 0.11]
Fixed effects model
0.08
[ 0.05; 0.11]
4.56e−08
I^2 = 0%
Replication
IHGC16noFIno23
0.01
[−0.04; 0.07]
HUNT
−0.00
[−0.08; 0.08]
Fixed effects model
0.01
[−0.04; 0.05]
7.37e−01
I^2 = 0%
Fixed effects model
0.06
[ 0.04; 0.08]
1.28e−06
−0.1
−0.05
0
0.05
0.1
