## Supplementary Figure 4 for "Fine-mapping a genome-wide meta-analysis of 98,374 migraine cases identifies 181 sets of candidate causal variants"

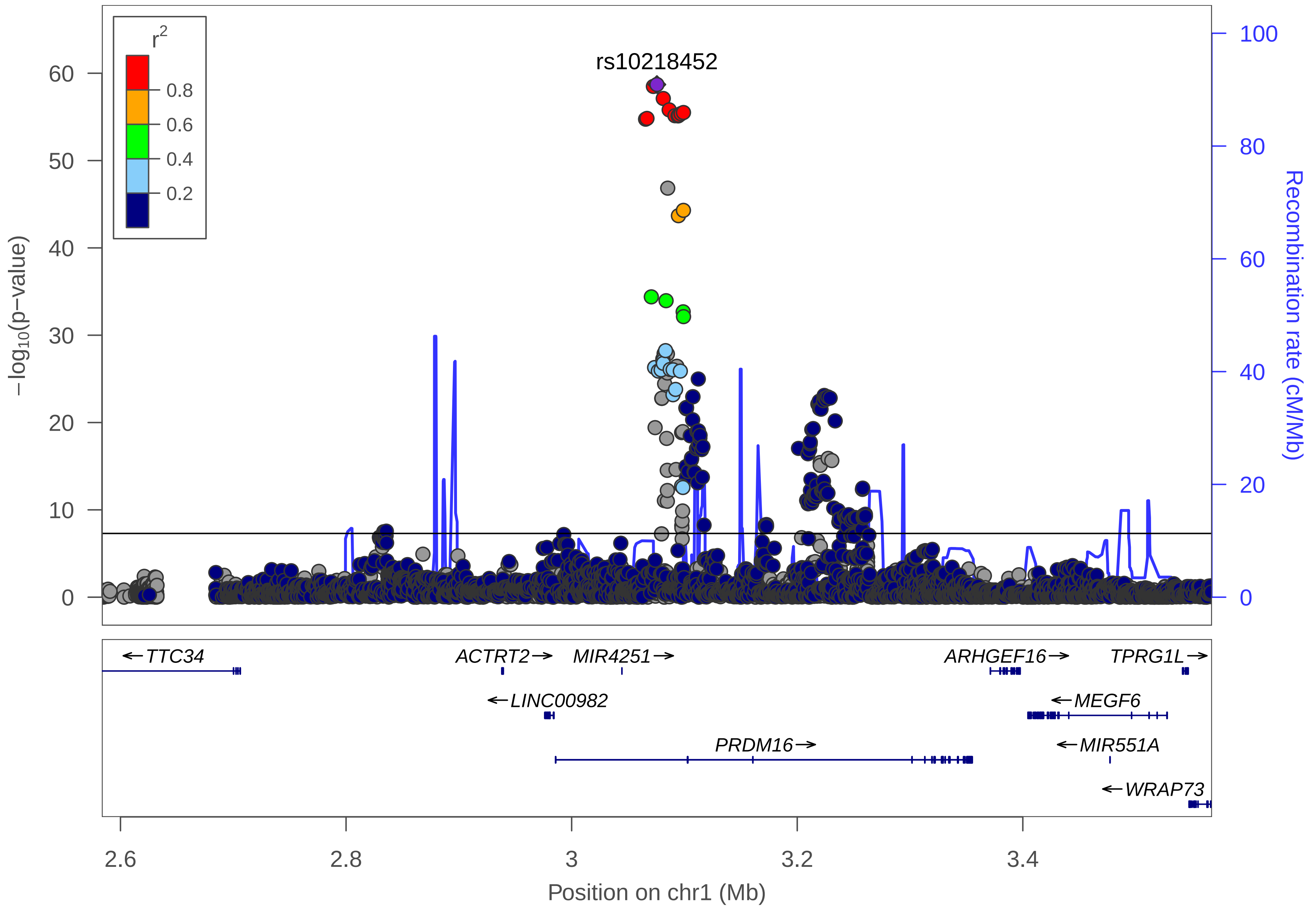

r2
0.8
0.6
0.4
0.2
rs10218452
−log10(p−value)
0
10
20
30
40
50
60
Recombination rate (cM/Mb)
0
20
40
60
80
100
TTC34
ACTRT2
MIR4251
ARHGEF16
TPRG1L
LINC00982
MEGF6
PRDM16
MIR551A
WRAP73
2.6
2.8
3
3.2
3.4
Position on chr1 (Mb)

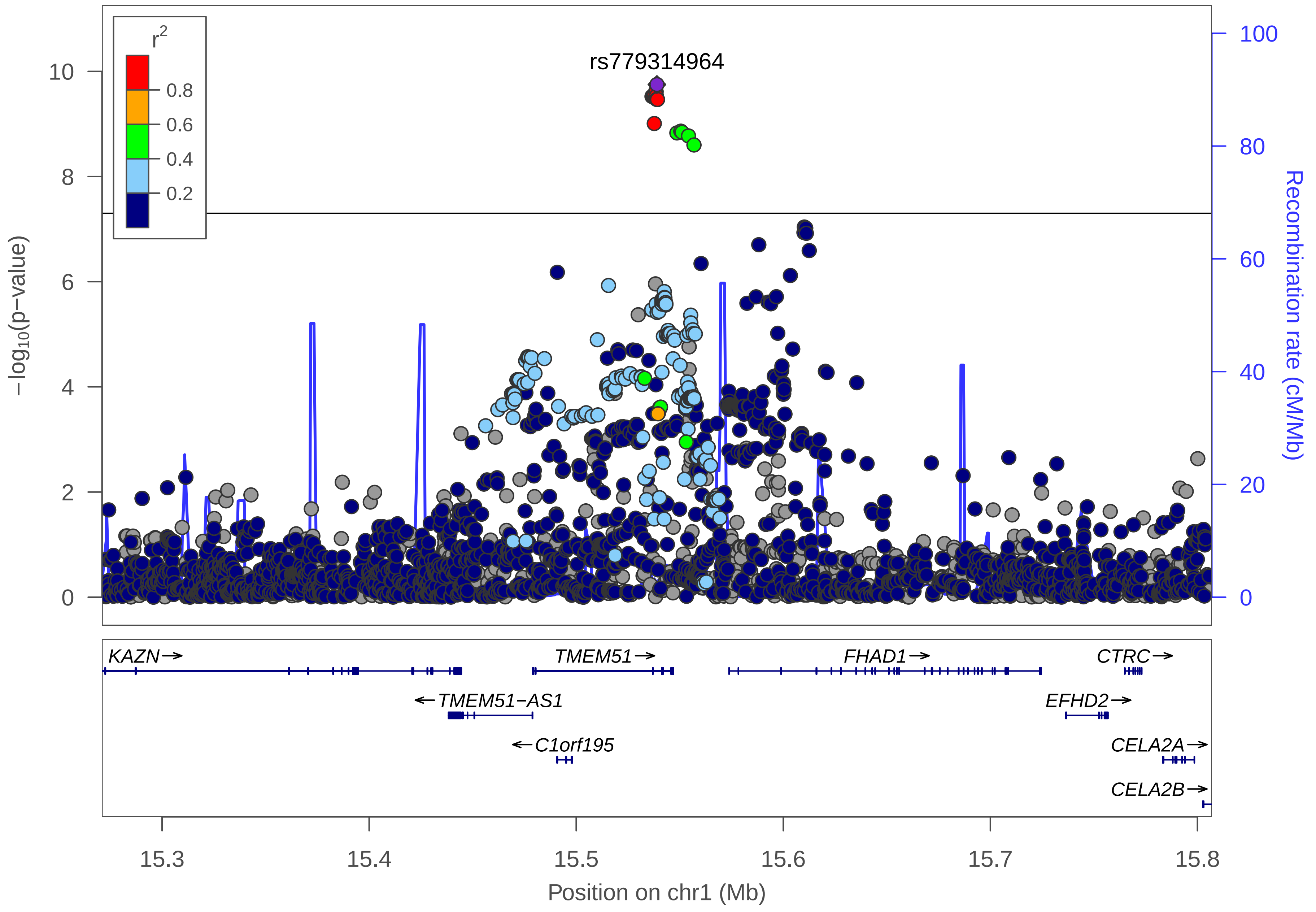

r2
0.8
0.6
0.4
0.2
rs779314964
10
8
6
4
2
0
−log10(p−value)
100
80
60
40
20
0
Recombination rate (cM/Mb)
KAZN→
TMEM51→
FHAD1→
CTRC→
←TMEM51−AS1
EFHD2→
←C1orf195
CELA2A→
CELA2B→
15.3
15.4
15.5
15.6
15.7
15.8
Position on chr1 (Mb)

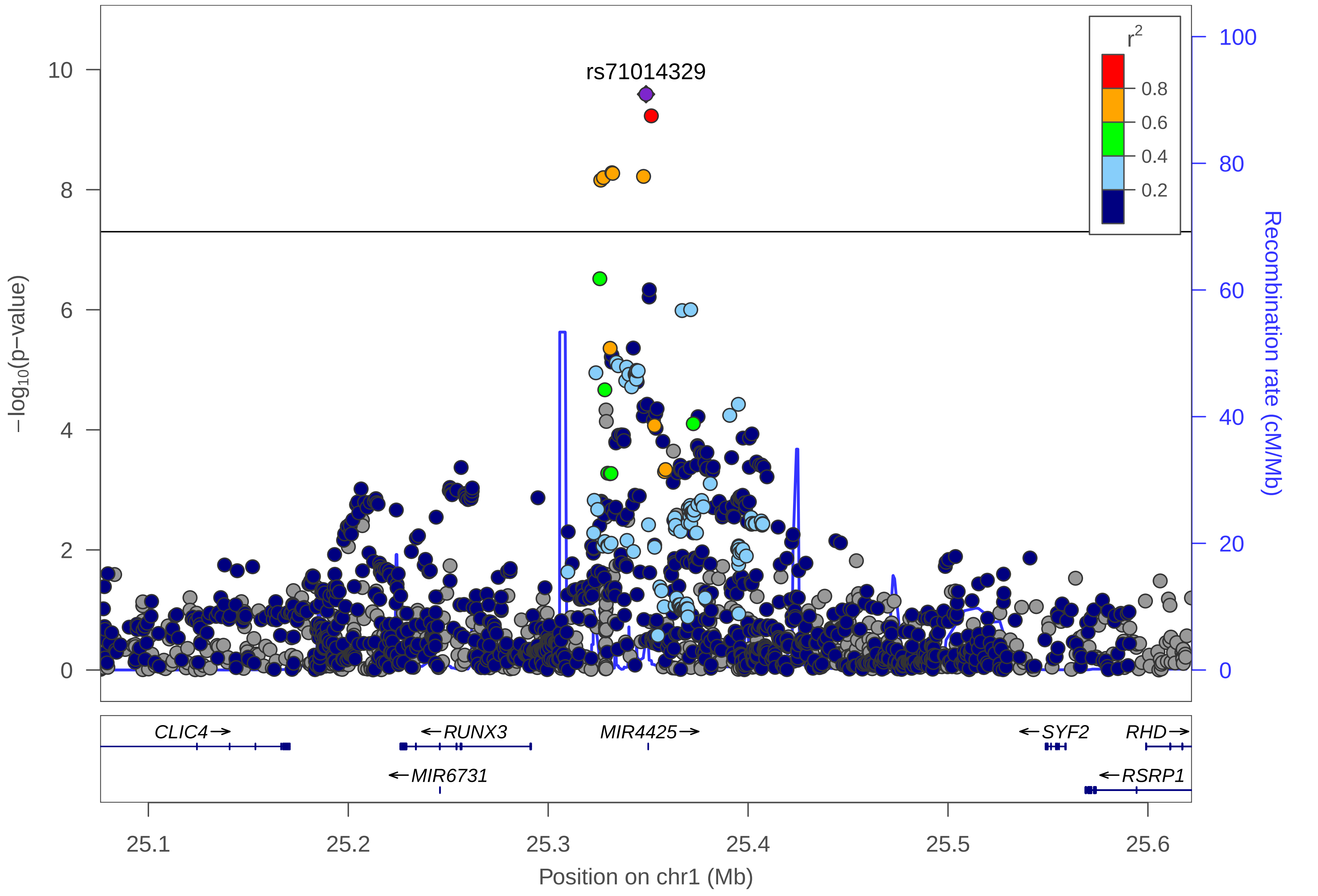

rs71014329
r2
0.8
0.6
0.4
0.2
−log10(p−value)
Recombination rate (cM/Mb)
0
2
4
6
8
10
0
20
40
60
80
100
CLIC4→
←RUNX3
MIR4425→
←SYF2
RHD→
←MIR6731
←RSRP1
25.1
25.2
25.3
25.4
25.5
25.6
Position on chr1 (Mb)

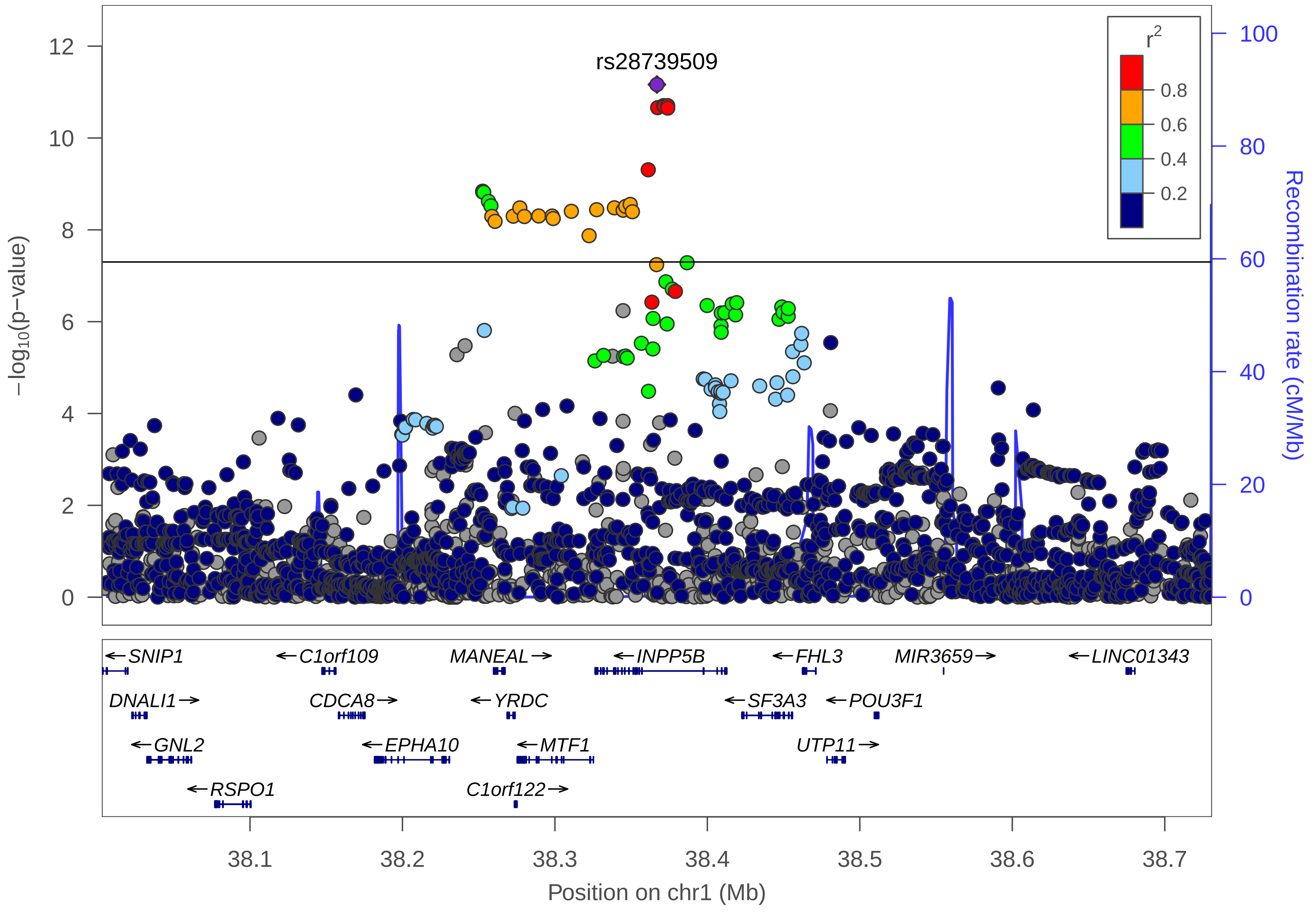
rs28739509
r2
0.8
0.6
0.4
0.2
−log10(p−value)
Recombination rate (cM/Mb)
SNIP1
C1orf109
MANEAL
INPP5B
FHL3
MIR3659
LINC01343
DNALI1
CDCA8
YRDC
SF3A3
POU3F1
GNL2
EPHA10
MTF1
UTP11
RSPO1
C1orf122
38.1
38.2
38.3
38.4
38.5
38.6
38.7
Position on chr1 (Mb)

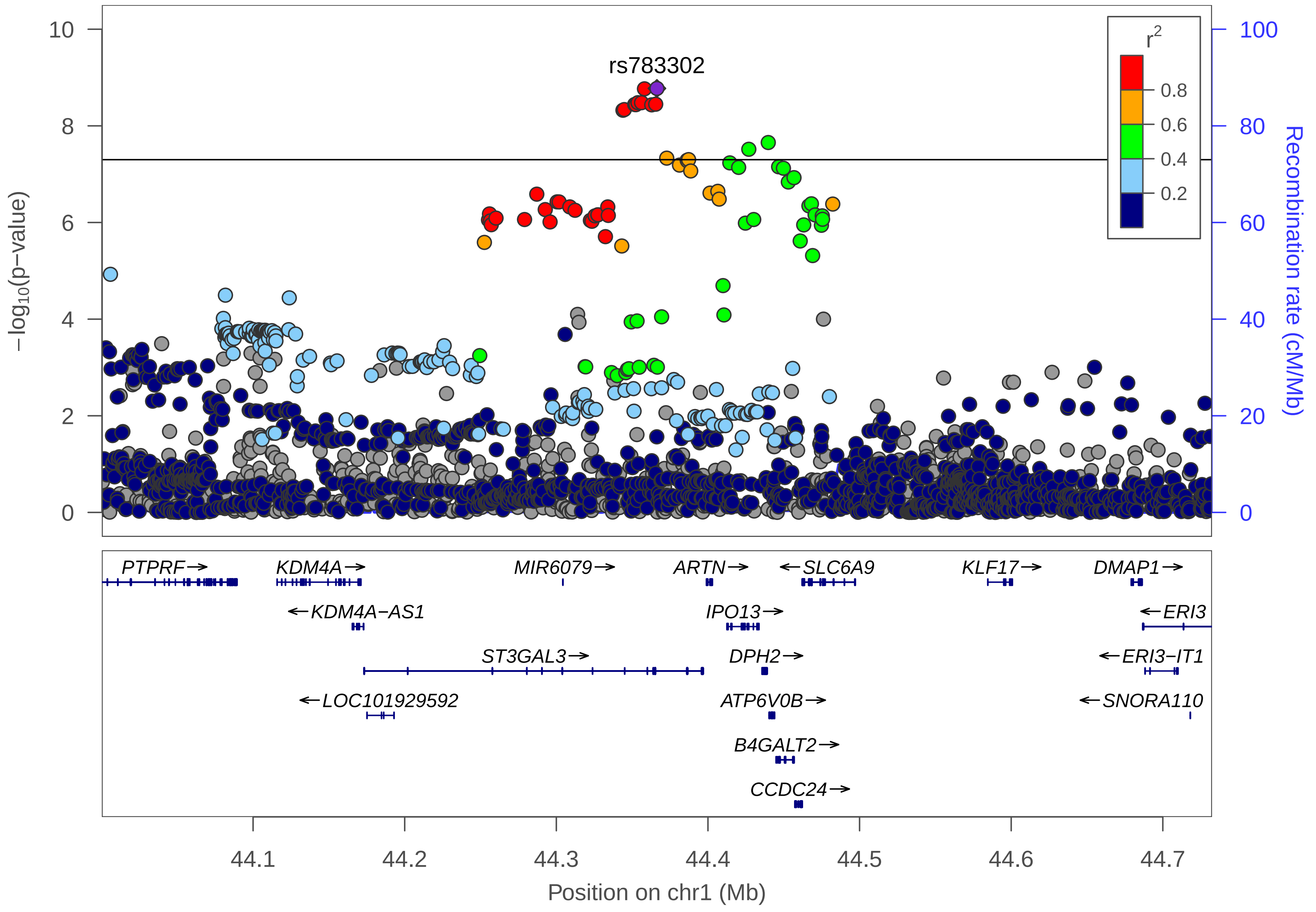

rs783302
r²
0.8
0.6
0.4
0.2
−log10(p−value)
Recombination rate (cM/Mb)
PTPRF→
KDM4A→
MIR6079→
ARTN→
←SLC6A9
KLF17→
DMAP1→
←KDM4A−AS1
IPO13→
←ERI3
ST3GAL3→
DPH2→
←ERI3−IT1
←LOC101929592
ATP6V0B→
←SNORA110
B4GALT2→
CCDC24→
44.1
44.2
44.3
44.4
44.5
44.6
44.7
Position on chr1 (Mb)

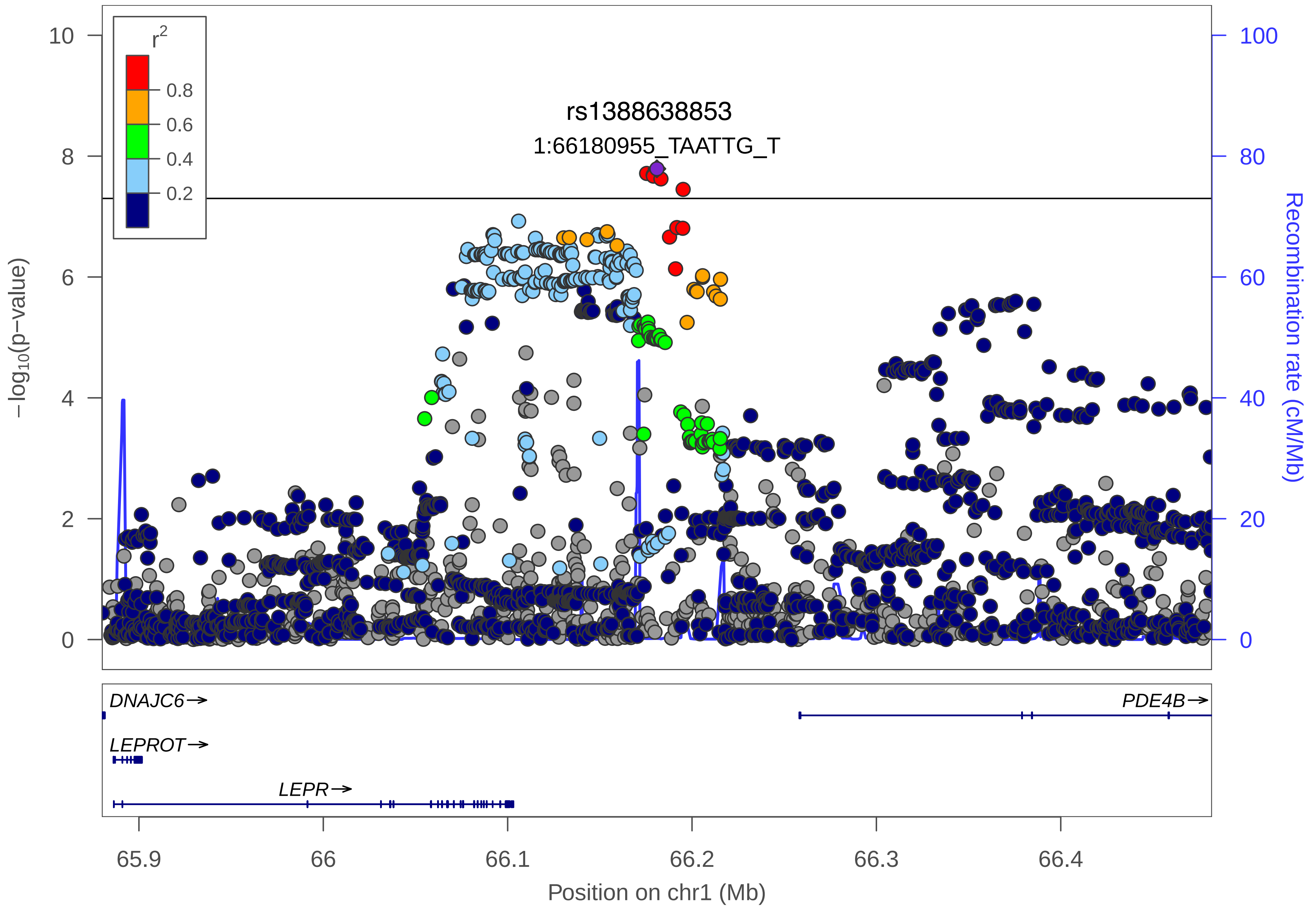

r2
0.8
0.6
0.4
0.2
rs1388638853
1:66180955_TAATTG_T
−log10(p−value)
Recombination rate (cM/Mb)
10
8
6
4
2
0
100
80
60
40
20
0
DNAJC6→
PDE4B→
LEPROT→
LEPR→
65.9
66
66.1
66.2
66.3
66.4
Position on chr1 (Mb)

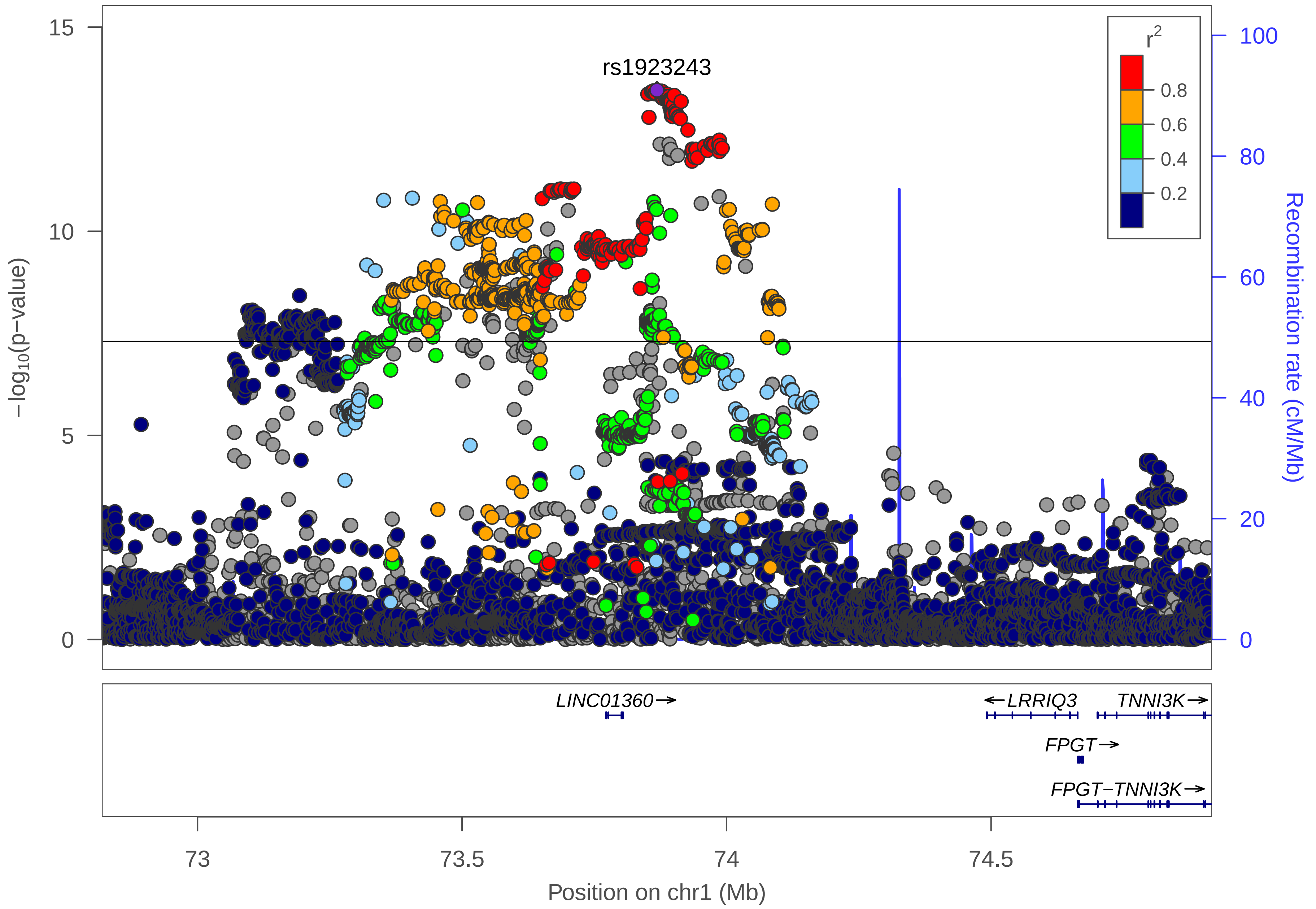

rs1923243
r^2
0.8
0.6
0.4
0.2
15
10
5
0
−log10(p−value)
100
80
60
40
20
0
Recombination rate (cM/Mb)
LINC01360→
←LRRIQ3
TNNI3K→
FPGT→
FPGT−TNNI3K→
73
73.5
74
74.5
Position on chr1 (Mb)

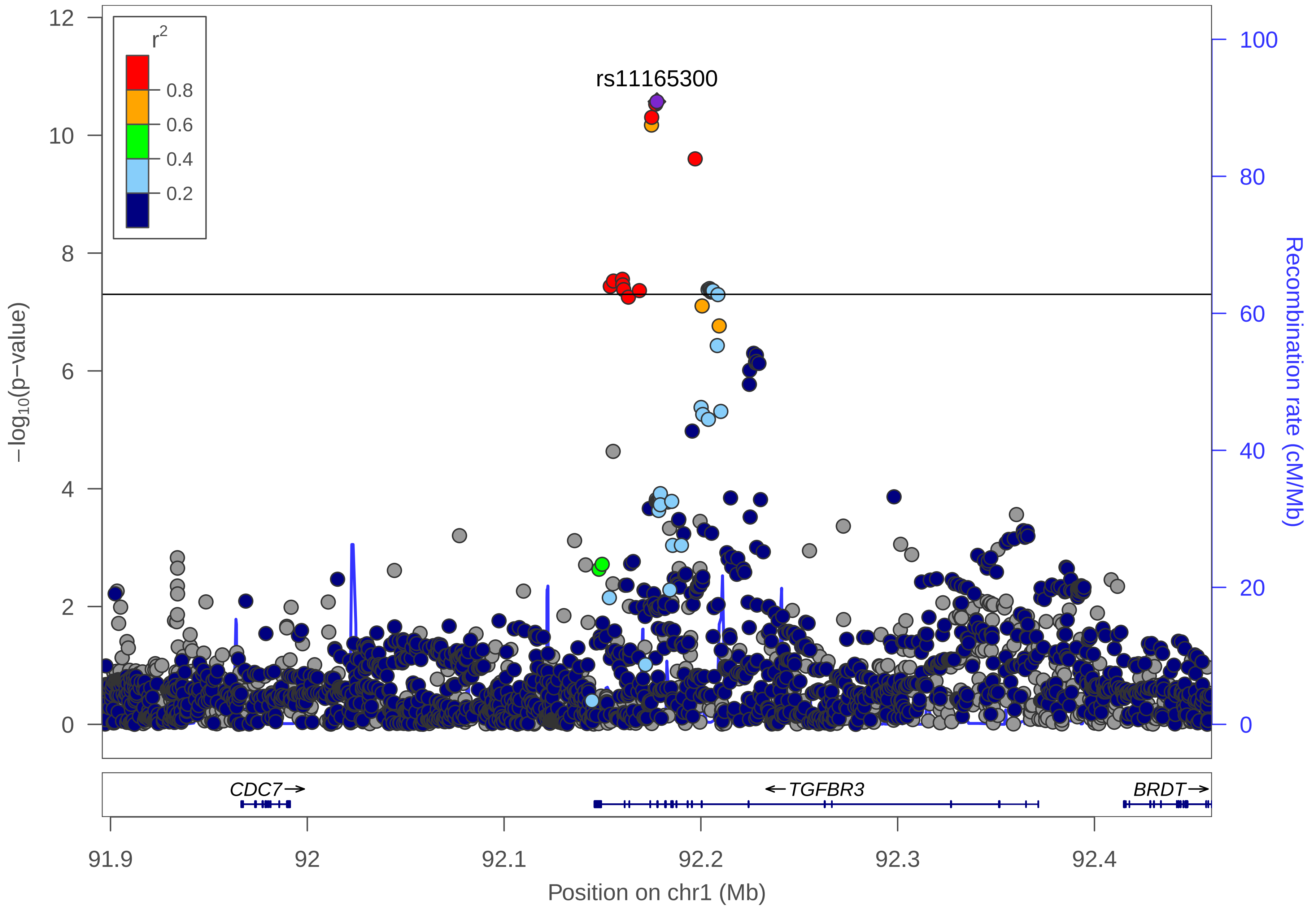

r2
0.8
0.6
0.4
0.2
rs11165300
−log10(p−value)
Recombination rate (cM/Mb)
12
10
8
6
4
2
0
100
80
60
40
20
0
CDC7→
←TGFBR3
BRDT→
91.9
92
92.1
92.2
92.3
92.4
Position on chr1 (Mb)

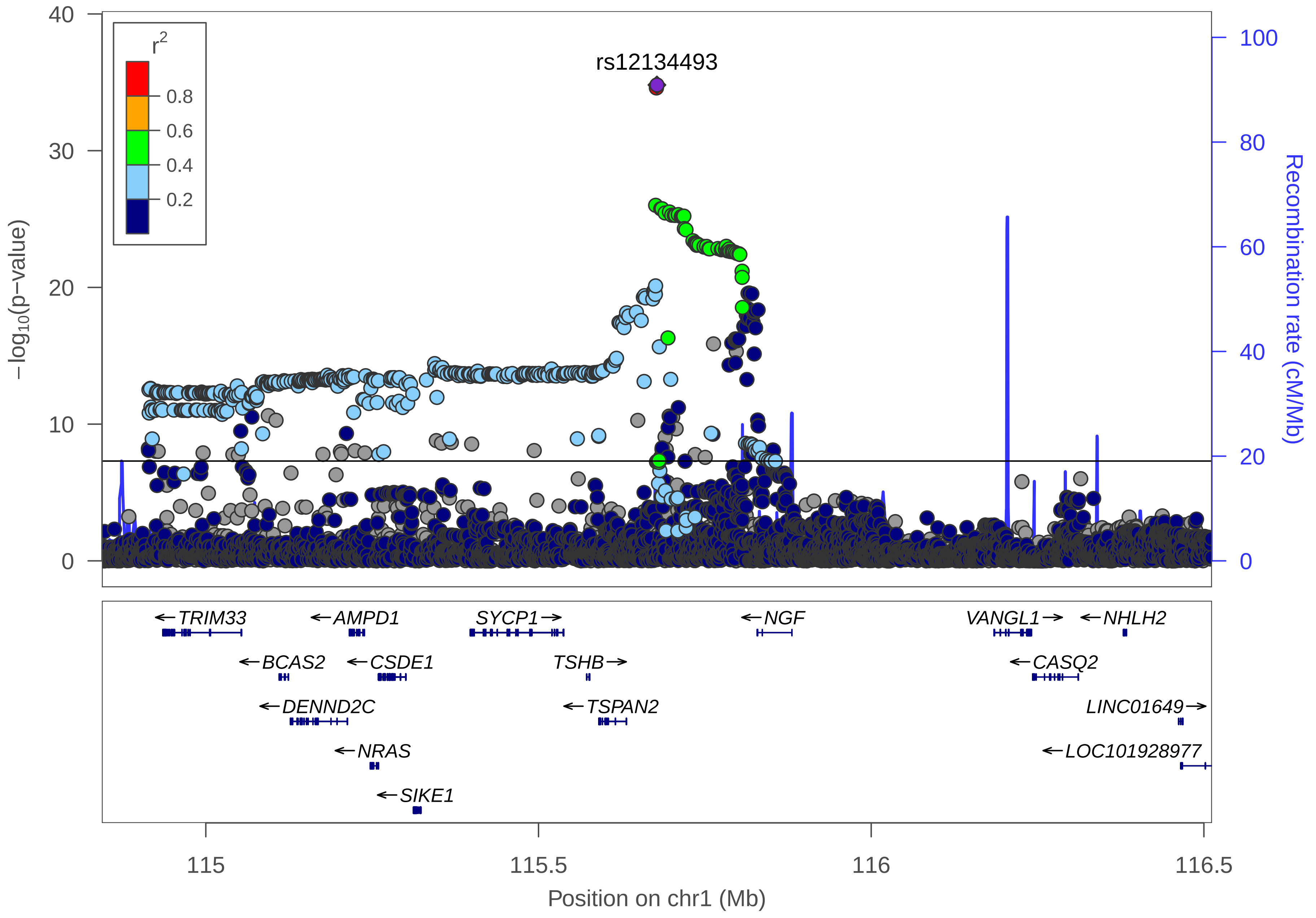

rs12134493
r2
0.8
0.6
0.4
0.2
−log10(p−value)
Recombination rate (cM/Mb)
0
10
20
30
40
0
20
40
60
80
100
TRIM33
AMPD1
SYCP1
NGF
VANGL1
NHLH2
BCAS2
CSDE1
TSHB
CASQ2
DENND2C
TSPAN2
LINC01649
NRAS
LOC101928977
SIKE1
115
115.5
116
116.5
Position on chr1 (Mb)

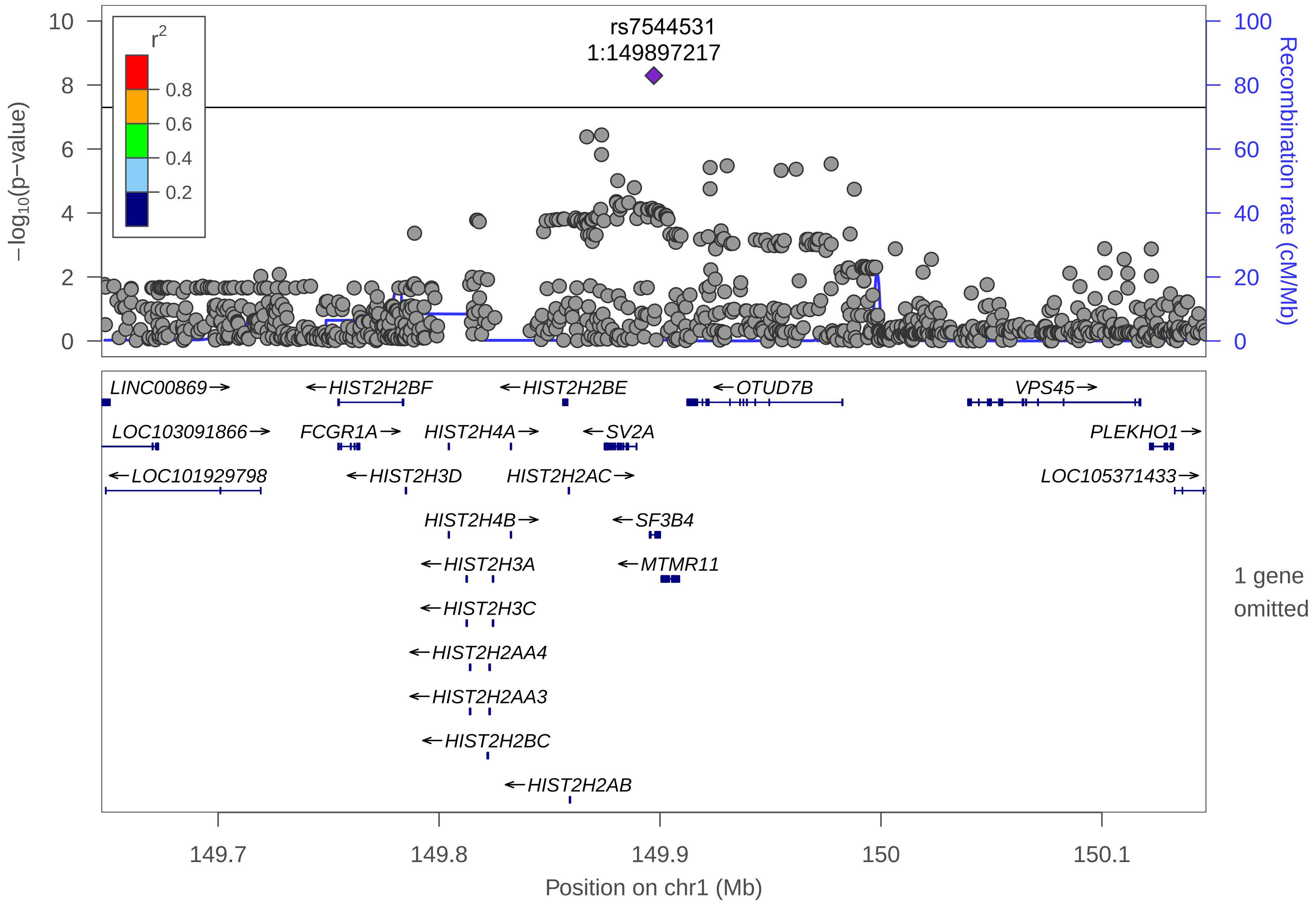

rs7544531
1:149897217
r2
0.8
0.6
0.4
0.2
−log10(p−value)
Recombination rate (cM/Mb)
10
8
6
4
2
0
100
80
60
40
20
0
LINC00869→
←HIST2H2BF
←HIST2H2BE
←OTUD7B
VPS45→
LOC103091866→
FCGR1A→
HIST2H4A→
←SV2A
PLEKHO1→
←LOC101929798
←HIST2H3D
HIST2H2AC→
LOC105371433→
HIST2H4B→
←SF3B4
←HIST2H3A
←MTMR11
1 gene omitted
←HIST2H3C
←HIST2H2AA4
←HIST2H2AA3
←HIST2H2BC
←HIST2H2AB
149.7
149.8
149.9
150
150.1
Position on chr1 (Mb)

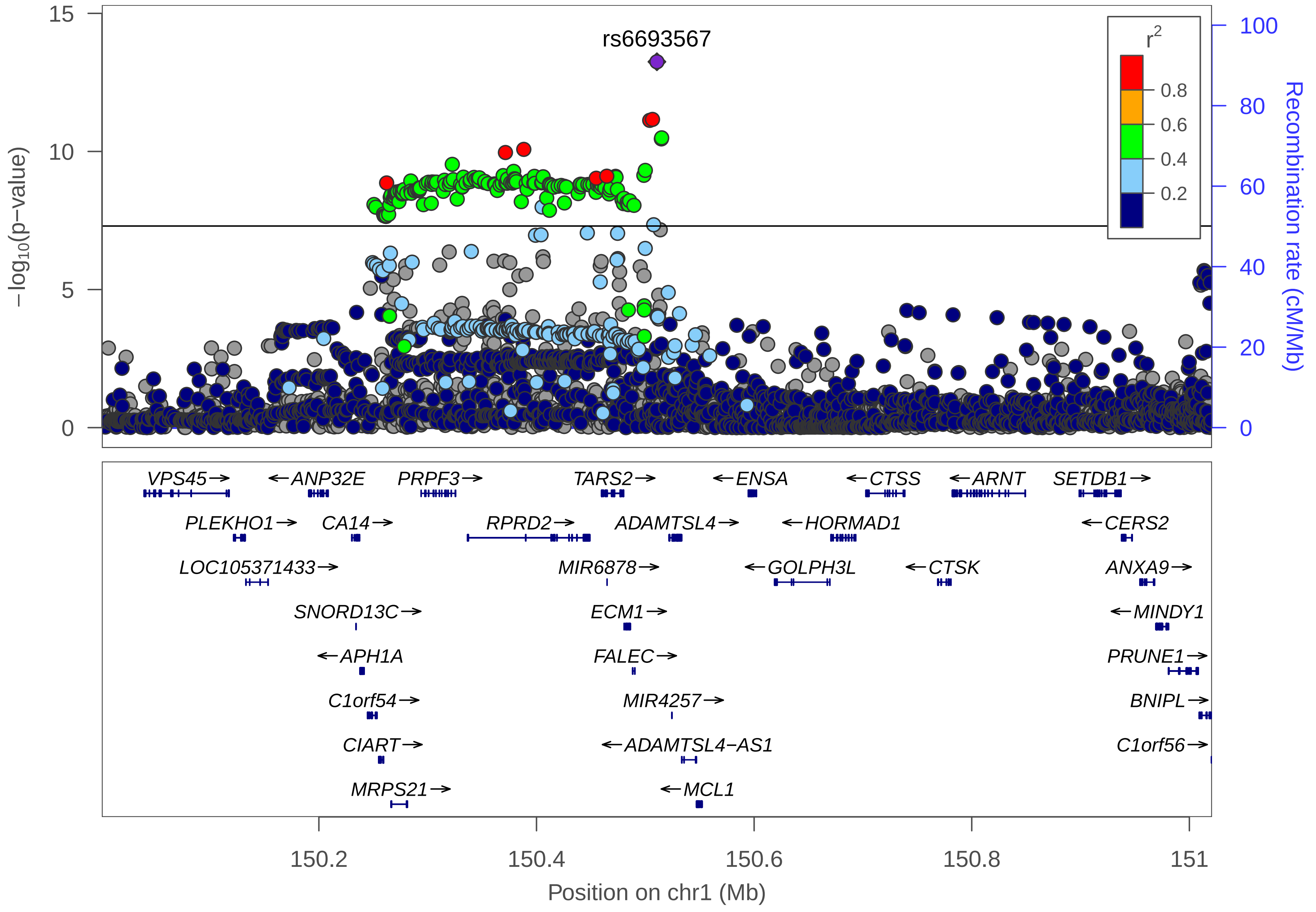
rs6693567
r2
0.8
0.6
0.4
0.2
−log10(p−value)
Recombination rate (cM/Mb)
VPS45→
←ANP32E
PRPF3→
TARS2→
←ENSA
←CTSS
←ARNT
SETDB1→
PLEKHO1→
CA14→
RPRD2→
ADAMTSL4→
←HORMAD1
←CERS2
LOC105371433→
MIR6878→
←GOLPH3L
←CTSK
ANXA9→
SNORD13C→
ECM1→
←MINDY1
←APH1A
FALEC→
PRUNE1→
C1orf54→
MIR4257→
BNIPL→
CIART→
←ADAMTSL4−AS1
C1orf56→
MRPS21→
←MCL1
150.2
150.4
150.6
150.8
151
Position on chr1 (Mb)

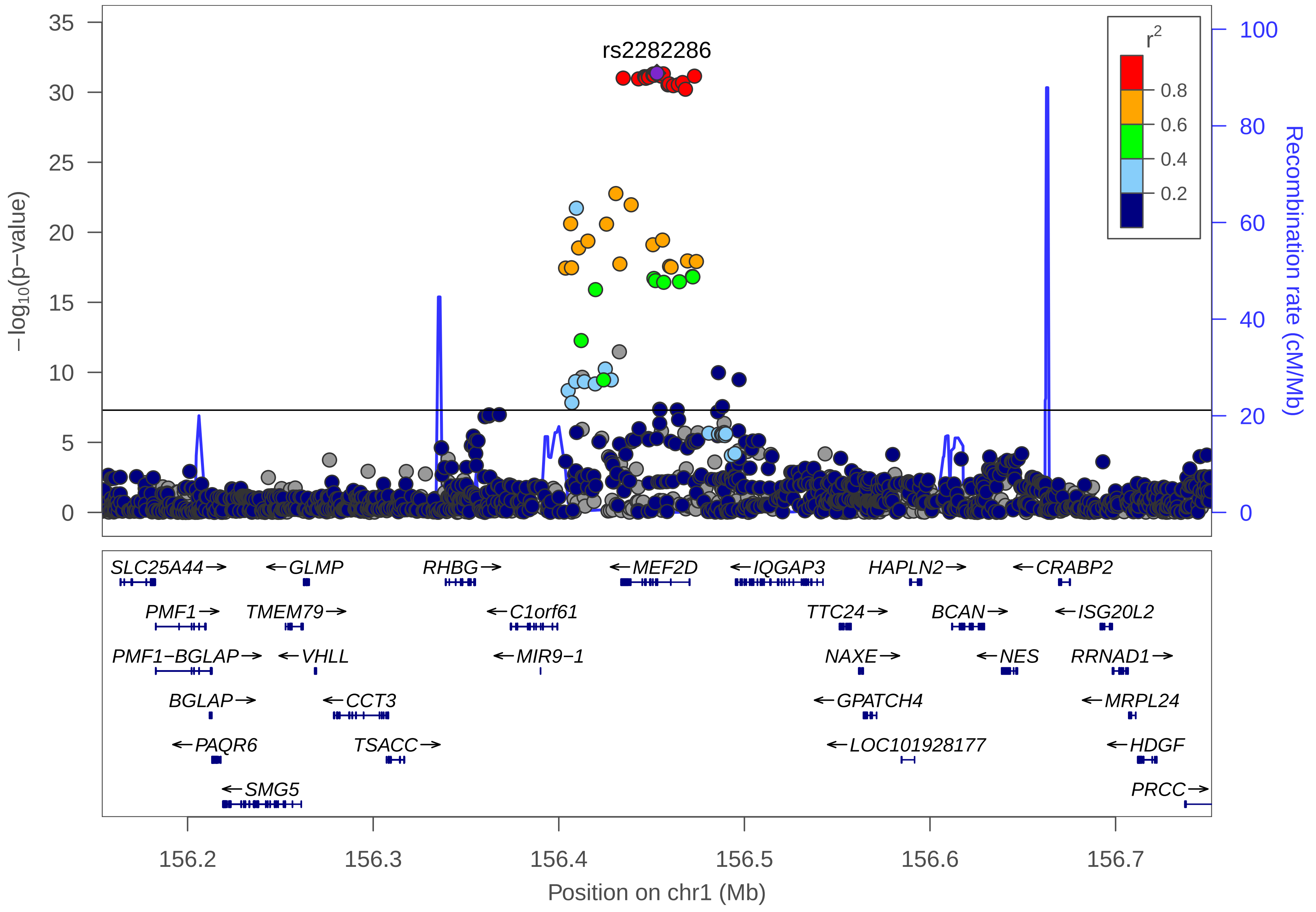

rs2282286
r2
0.8
0.6
0.4
0.2
35
30
25
20
15
10
5
0
−log10(p−value)
100
80
60
40
20
0
Recombination rate (cM/Mb)
SLC25A44→
←GLMP
RHBG→
←MEF2D
←IQGAP3
HAPLN2→
←CRABP2
PMF1→
TMEM79→
←C1orf61
TTC24→
BCAN→
←ISG20L2
PMF1−BGLAP→
←VHLL
←MIR9−1
NAXE→
←NES
RRNAD1→
BGLAP→
←CCT3
←GPATCH4
←MRPL24
←PAQR6
TSACC→
←LOC101928177
←HDGF
←SMG5
PRCC→
156.2
156.3
156.4
156.5
156.6
156.7
Position on chr1 (Mb)

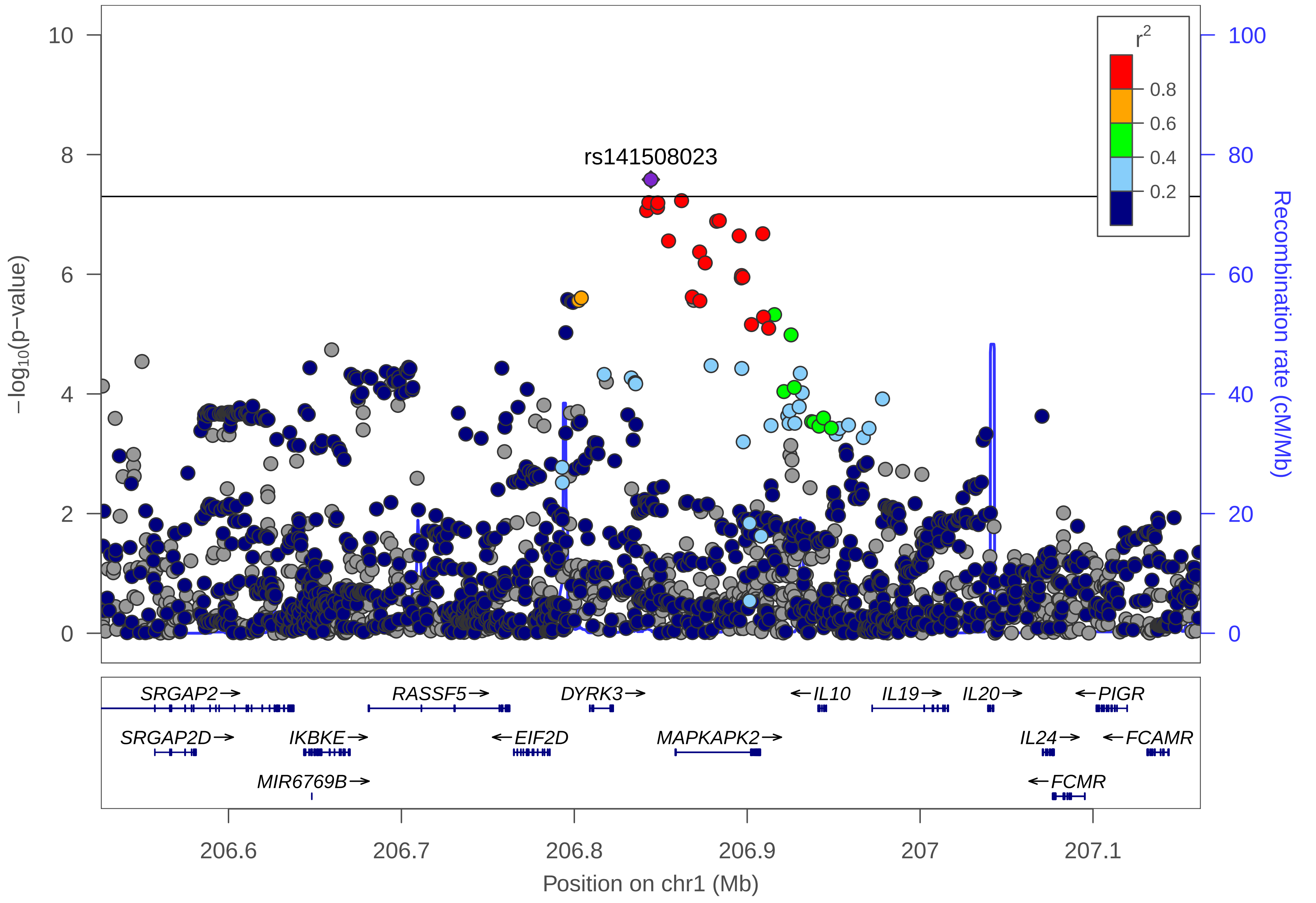
rs141508023
r2
0.8
0.6
0.4
0.2
−log10(p−value)
Recombination rate (cM/Mb)
10
8
6
4
2
0
100
80
60
40
20
0
SRGAP2→
RASSF5→
DYRK3→
←IL10
IL19→
IL20→
←PIGR
SRGAP2D→
IKBKE→
←EIF2D
MAPKAPK2→
IL24→
←FCAMR
MIR6769B→
←FCMR
206.6
206.7
206.8
206.9
207
207.1
Position on chr1 (Mb)

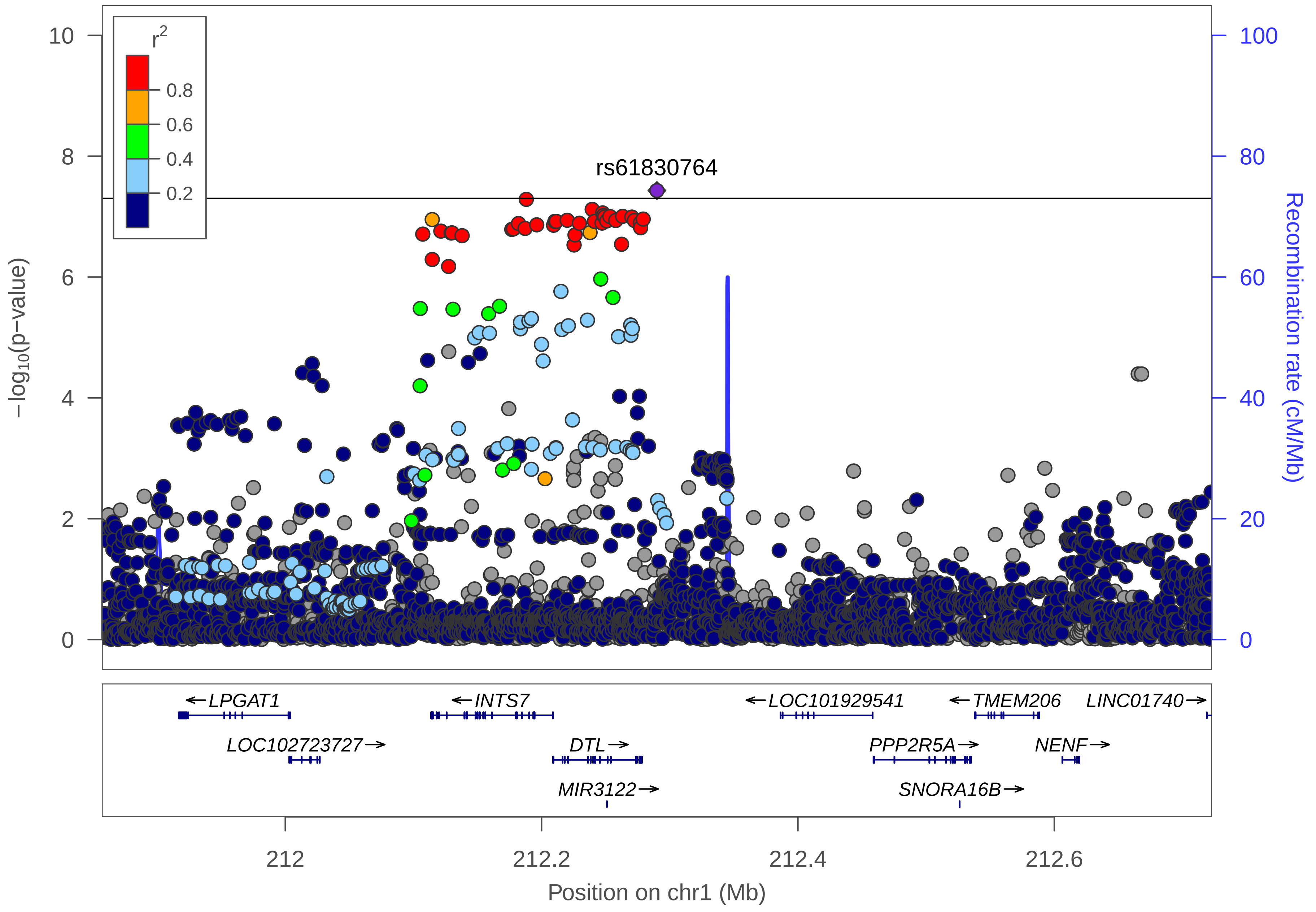

r2
0.8
0.6
0.4
0.2
rs61830764
10
8
6
4
2
0
100
80
60
40
20
0
−log10(p−value)
Recombination rate (cM/Mb)
←LPGAT1
←INTS7
←LOC101929541
←TMEM206
LINC01740→
LOC102723727→
DTL→
PPP2R5A→
NENF→
MIR3122→
SNORA16B→
212
212.2
212.4
212.6
Position on chr1 (Mb)

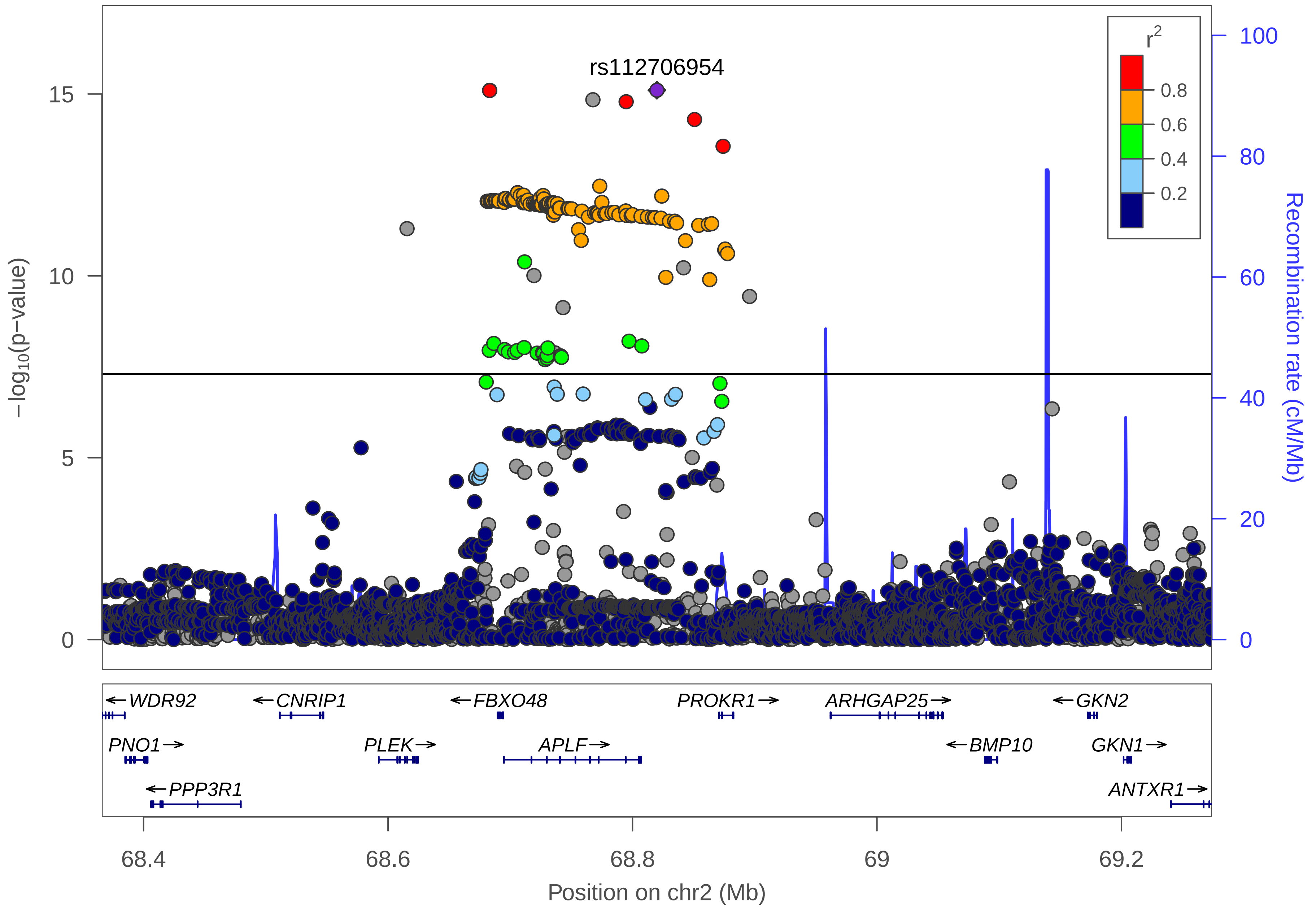
rs112706954
r2
0.8
0.6
0.4
0.2
15
10
5
0
−log10(p−value)
100
80
60
40
20
0
Recombination rate (cM/Mb)
WDR92
CNRIP1
FBXO48
PROKR1
ARHGAP25
GKN2
PNO1
PLEK
APLF
BMP10
GKN1
PPP3R1
ANTXR1
68.4
68.6
68.8
69
69.2
Position on chr2 (Mb)

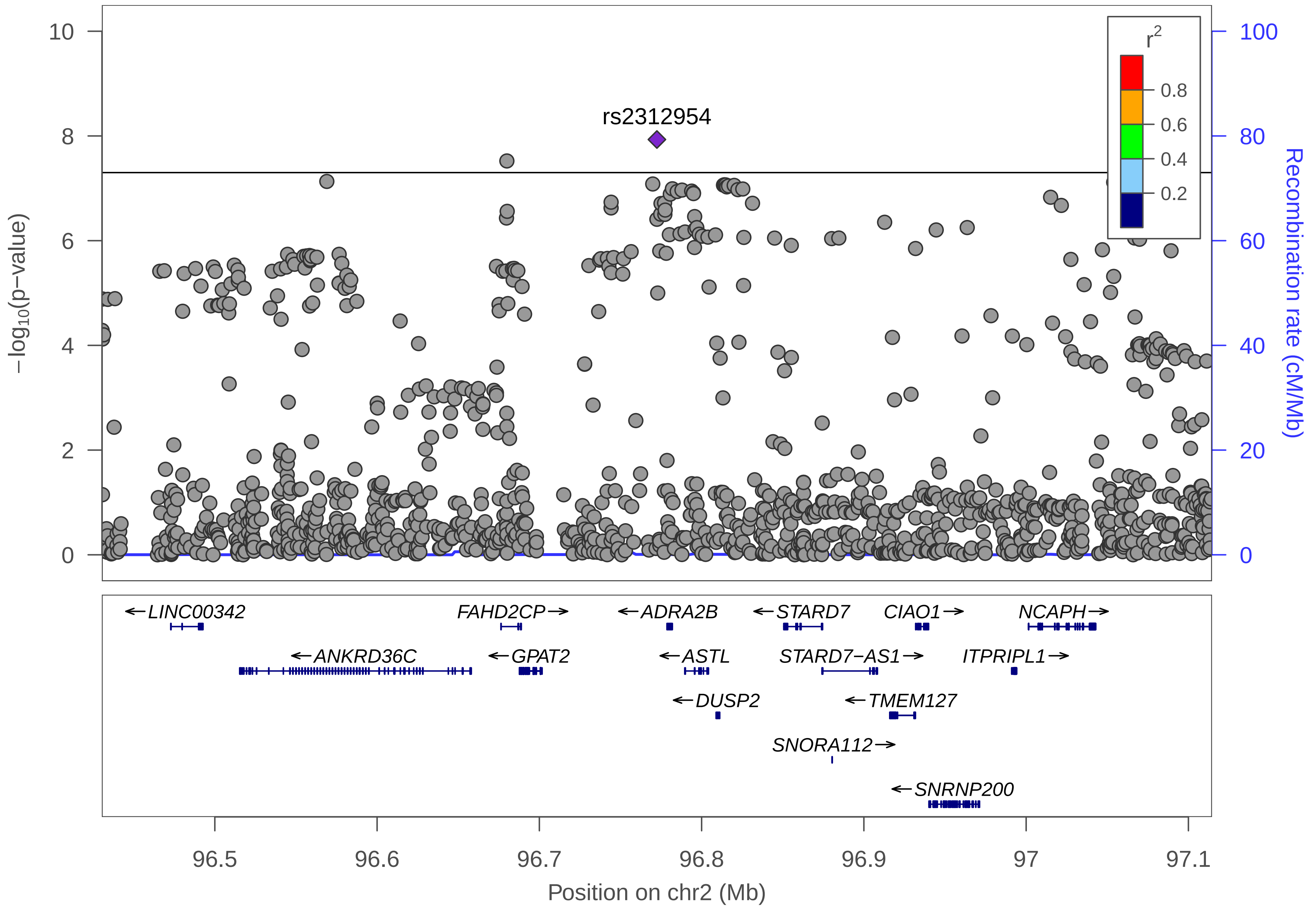

rs2312954
−log10(p−value)
Recombination rate (cM/Mb)
r2
0.8
0.6
0.4
0.2
10
8
6
4
2
0
100
80
60
40
20
0
←LINC00342
FAHD2CP→
←ADRA2B
←STARD7
CIAO1→
NCAPH→
←ANKRD36C
←GPAT2
←ASTL
STARD7−AS1→
ITPRIPL1→
←DUSP2
←TMEM127
SNORA112→
←SNRNP200
96.5
96.6
96.7
96.8
96.9
97
97.1
Position on chr2 (Mb)

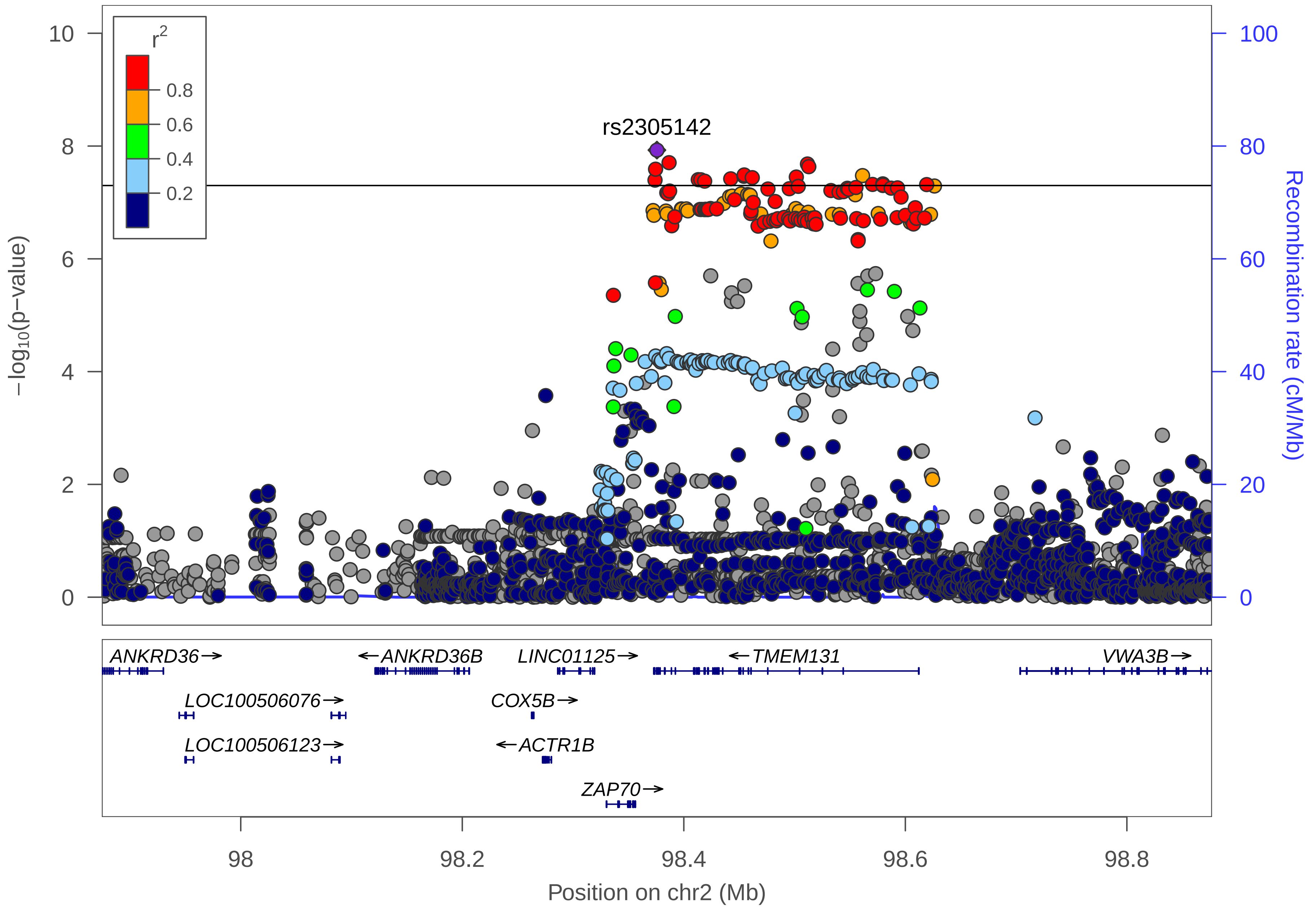

r2
0.8
0.6
0.4
0.2
rs2305142
10
8
6
4
2
0
100
80
60
40
20
0
−log10(p−value)
Recombination rate (cM/Mb)
ANKRD36→
←ANKRD36B
LINC01125→
←TMEM131
VWA3B→
LOC100506076→
COX5B→
LOC100506123→
←ACTR1B
ZAP70→
98
98.2
98.4
98.6
98.8
Position on chr2 (Mb)

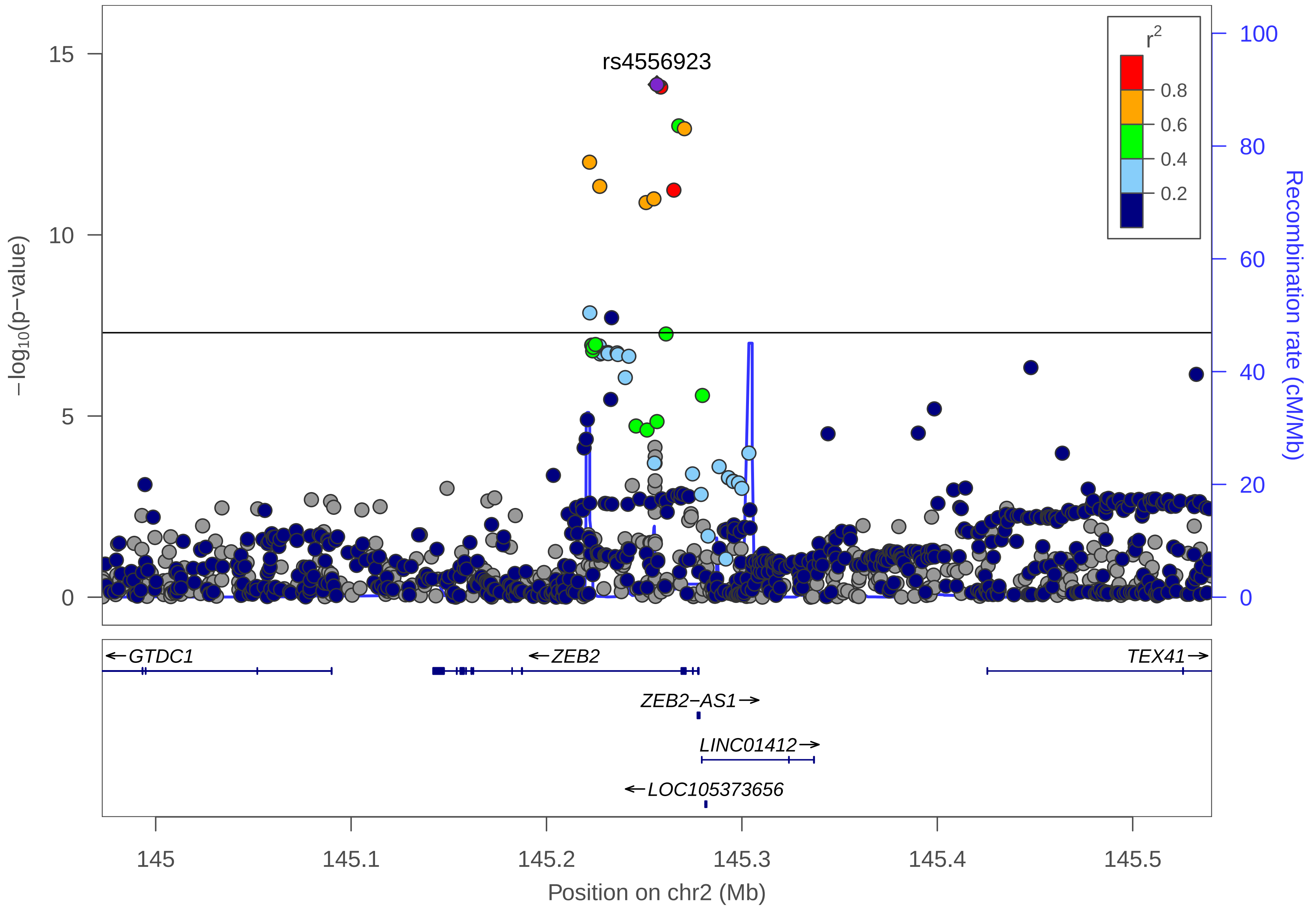

rs4556923
r2
0.8
0.6
0.4
0.2
15
10
5
0
−log10(p−value)
100
80
60
40
20
0
Recombination rate (cM/Mb)
←GTDC1
←ZEB2
TEX41→
ZEB2−AS1→
LINC01412→
←LOC105373656
145
145.1
145.2
145.3
145.4
145.5
Position on chr2 (Mb)

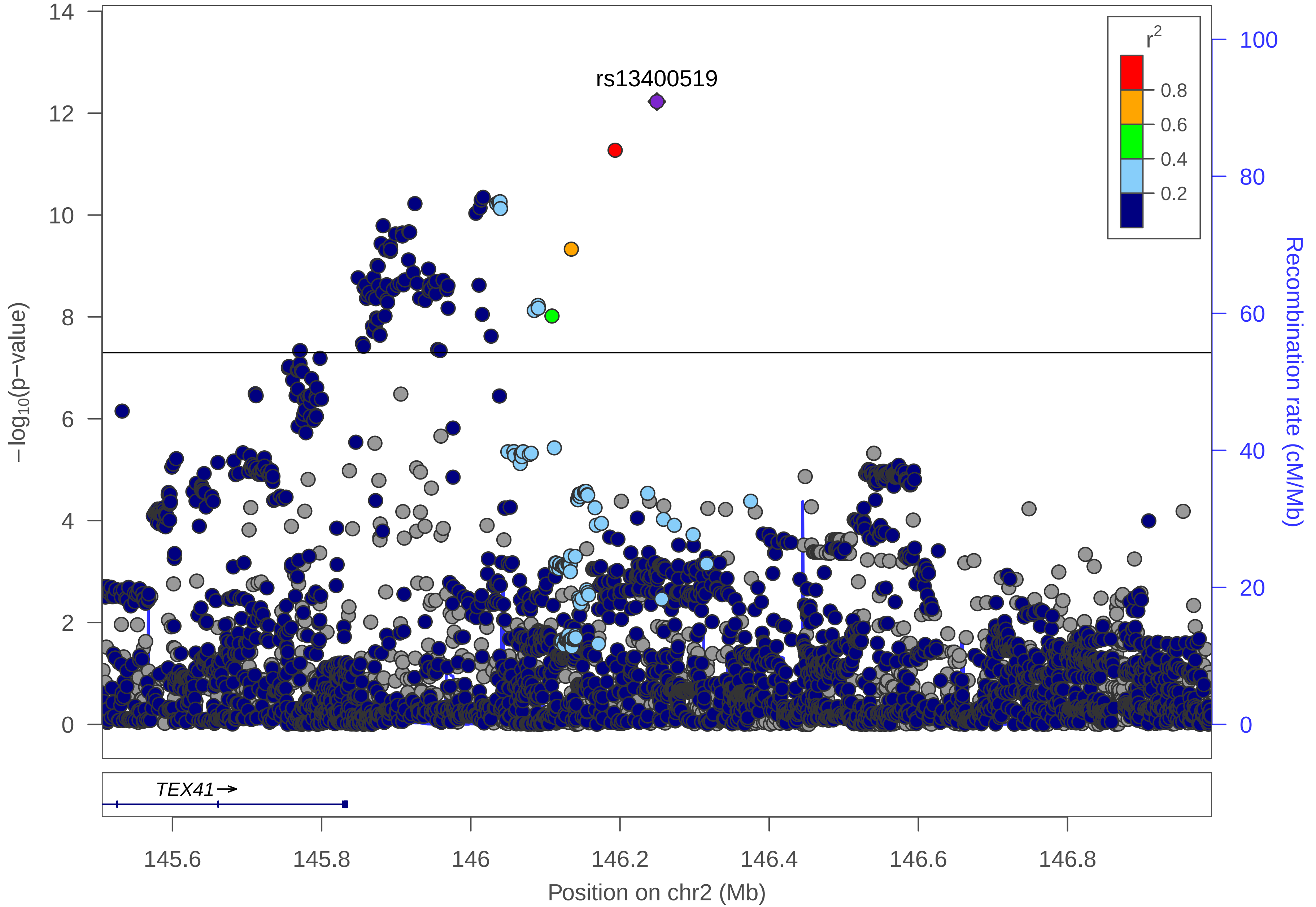

rs13400519
r2
0.8
0.6
0.4
0.2
14
12
10
8
6
4
2
0
−log10(p−value)
100
80
60
40
20
0
Recombination rate (cM/Mb)
TEX41→
145.6
145.8
146
146.2
146.4
146.6
146.8
Position on chr2 (Mb)

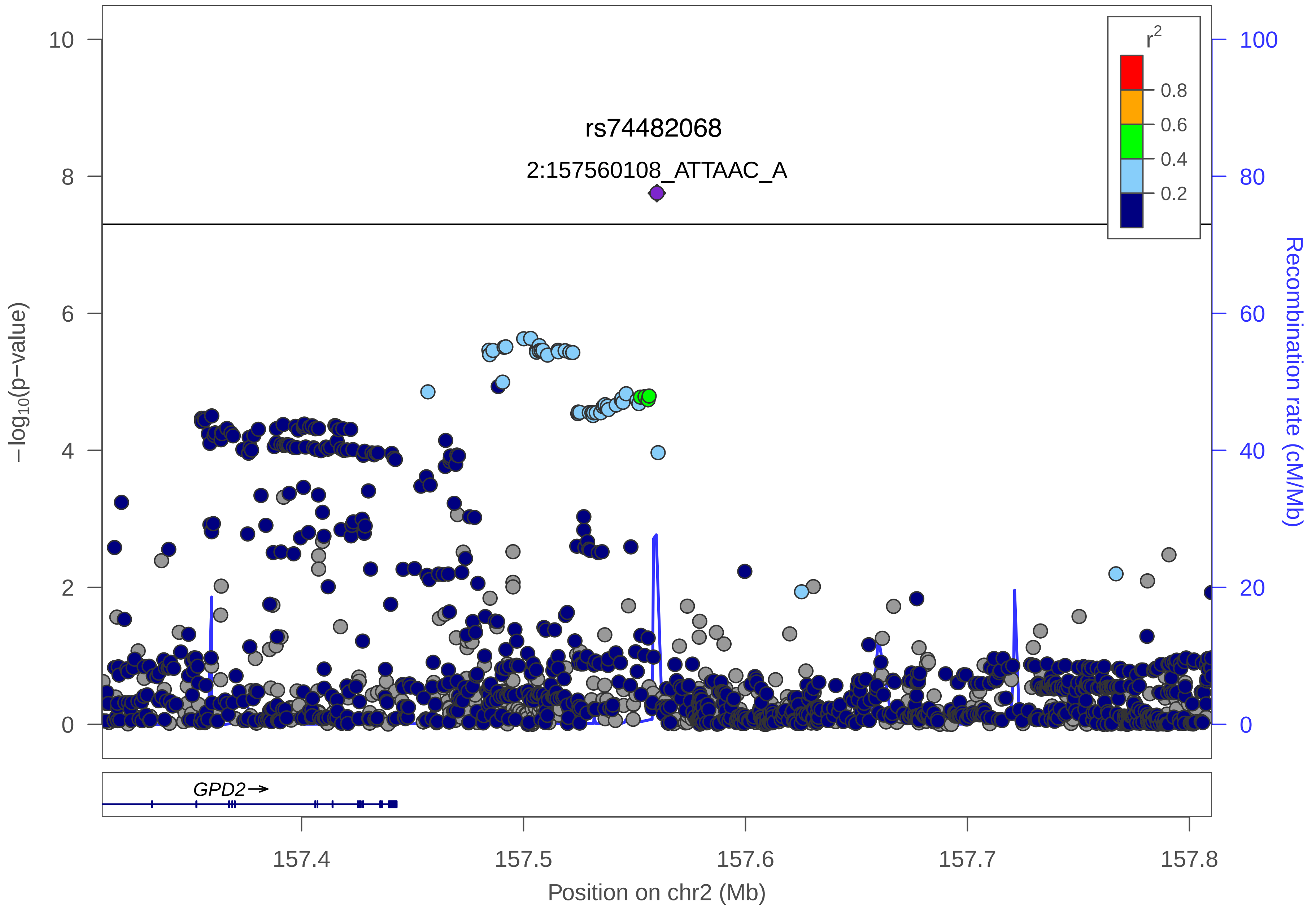
rs74482068
2:157560108_ATTAAC_A
r²
0.8
0.6
0.4
0.2
−log10(p−value)
Recombination rate (cM/Mb)
GPD2→
157.4
157.5
157.6
157.7
157.8
Position on chr2 (Mb)

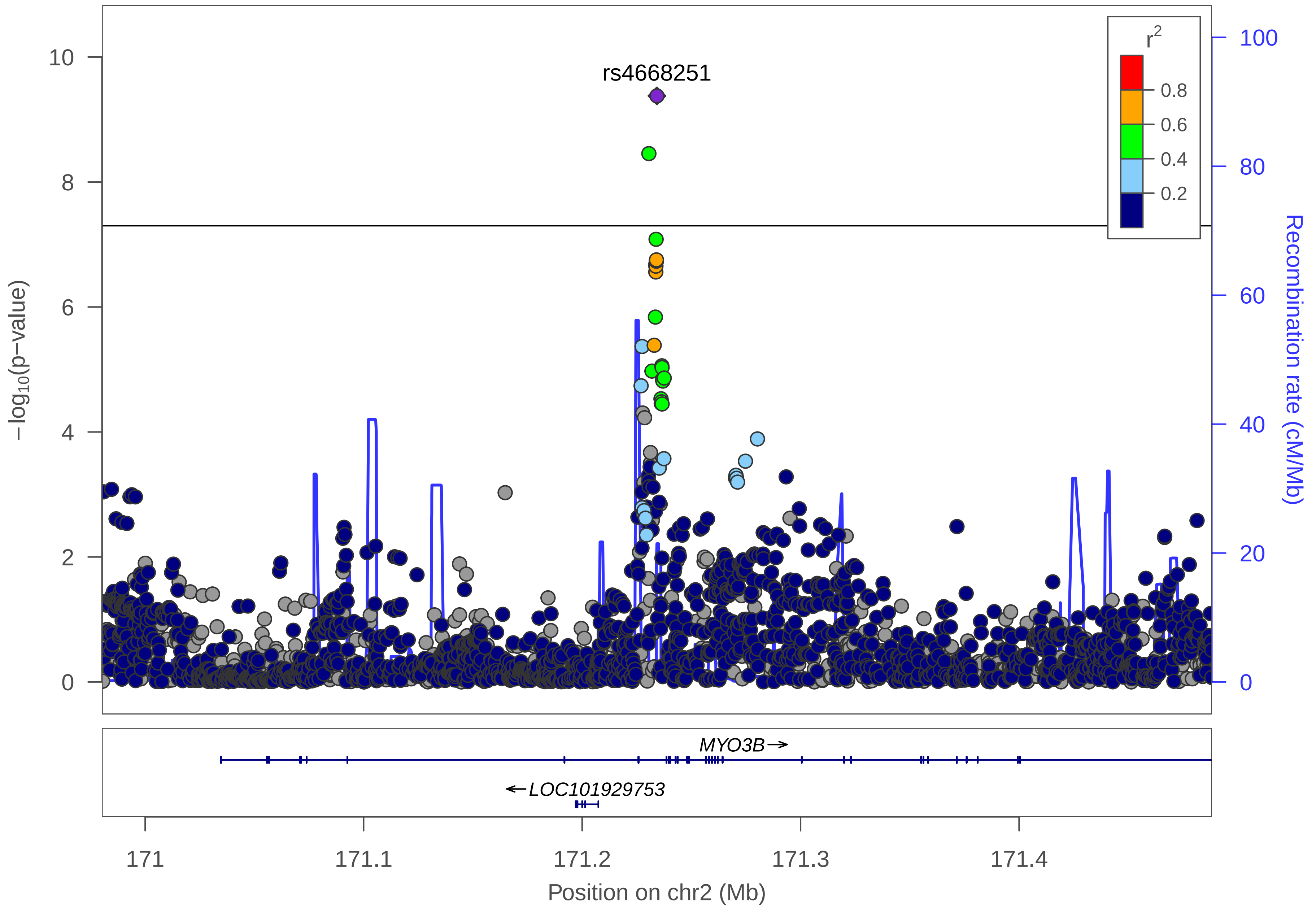

rs4668251
r2
0.8
0.6
0.4
0.2
−log10(p−value)
Recombination rate (cM/Mb)
MYO3B→
←LOC101929753
171
171.1
171.2
171.3
171.4
Position on chr2 (Mb)

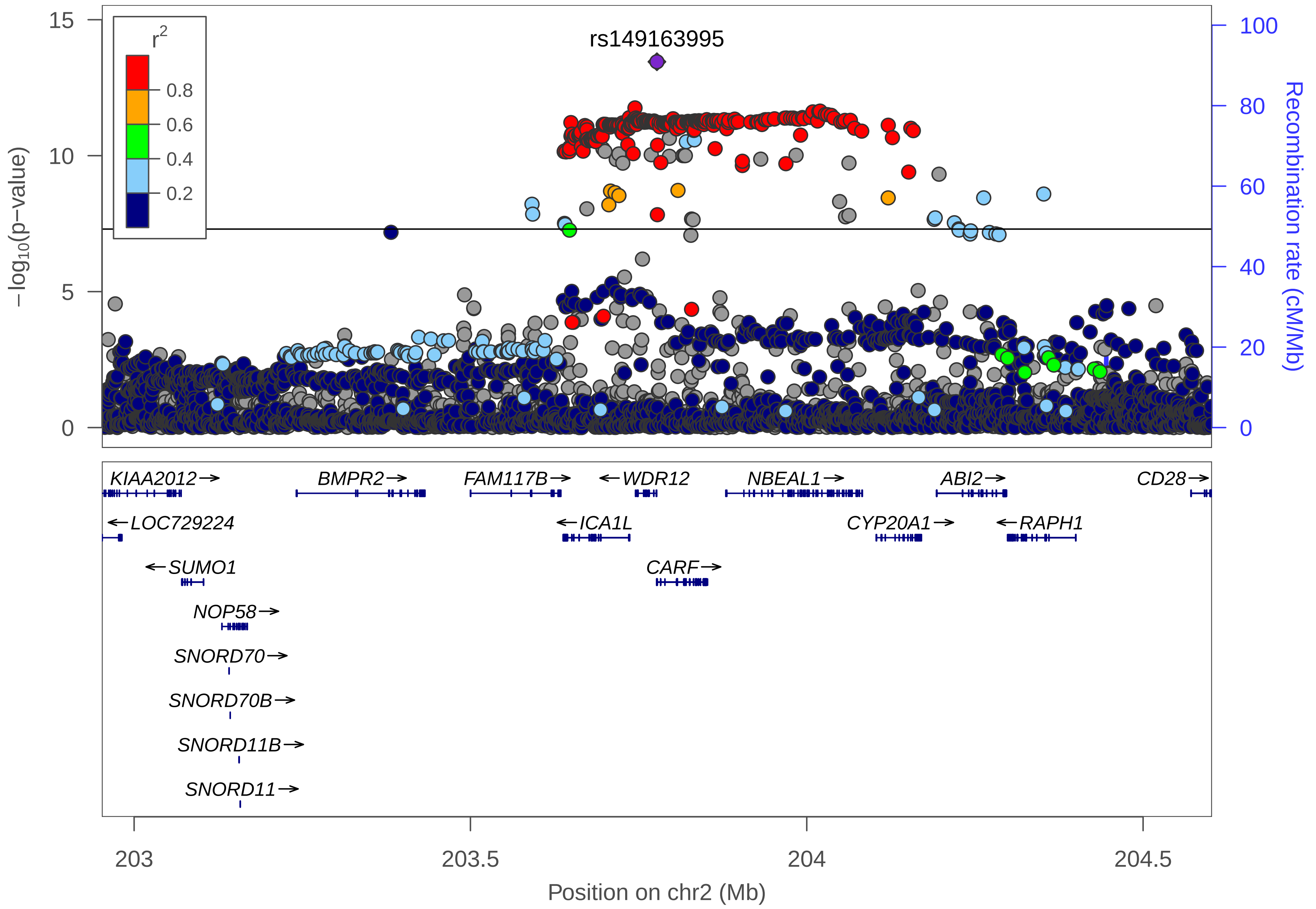

rs149163995
r2
0.8
0.6
0.4
0.2
−log10(p−value)
Recombination rate (cM/Mb)
KIAA2012→
BMPR2→
FAM117B→
←WDR12
NBEAL1→
ABI2→
CD28→
←LOC729224
←ICA1L
CYP20A1→
←RAPH1
←SUMO1
CARF→
NOP58→
SNORD70→
SNORD70B→
SNORD11B→
SNORD11→
203
203.5
204
204.5
Position on chr2 (Mb)

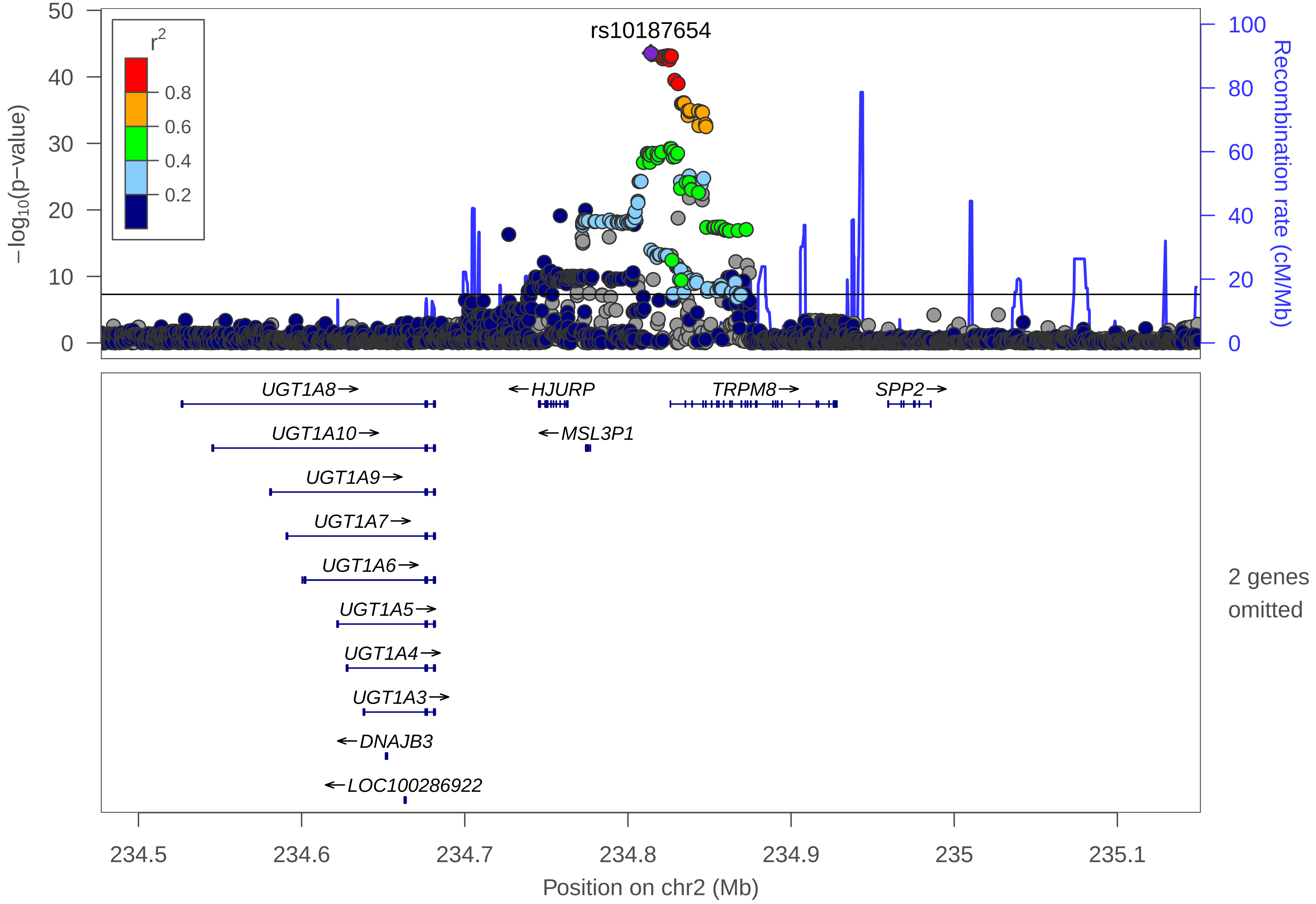

rs10187654
r2
0.8
0.6
0.4
0.2
−log10(p−value)
0
10
20
30
40
50
Recombination rate (cM/Mb)
0
20
40
60
80
100
UGT1A8→
←HJURP
TRPM8→
SPP2→
UGT1A10→
←MSL3P1
UGT1A9→
UGT1A7→
UGT1A6→
UGT1A5→
UGT1A4→
UGT1A3→
←DNAJB3
←LOC100286922
2 genes omitted
234.5
234.6
234.7
234.8
234.9
235
235.1
Position on chr2 (Mb)

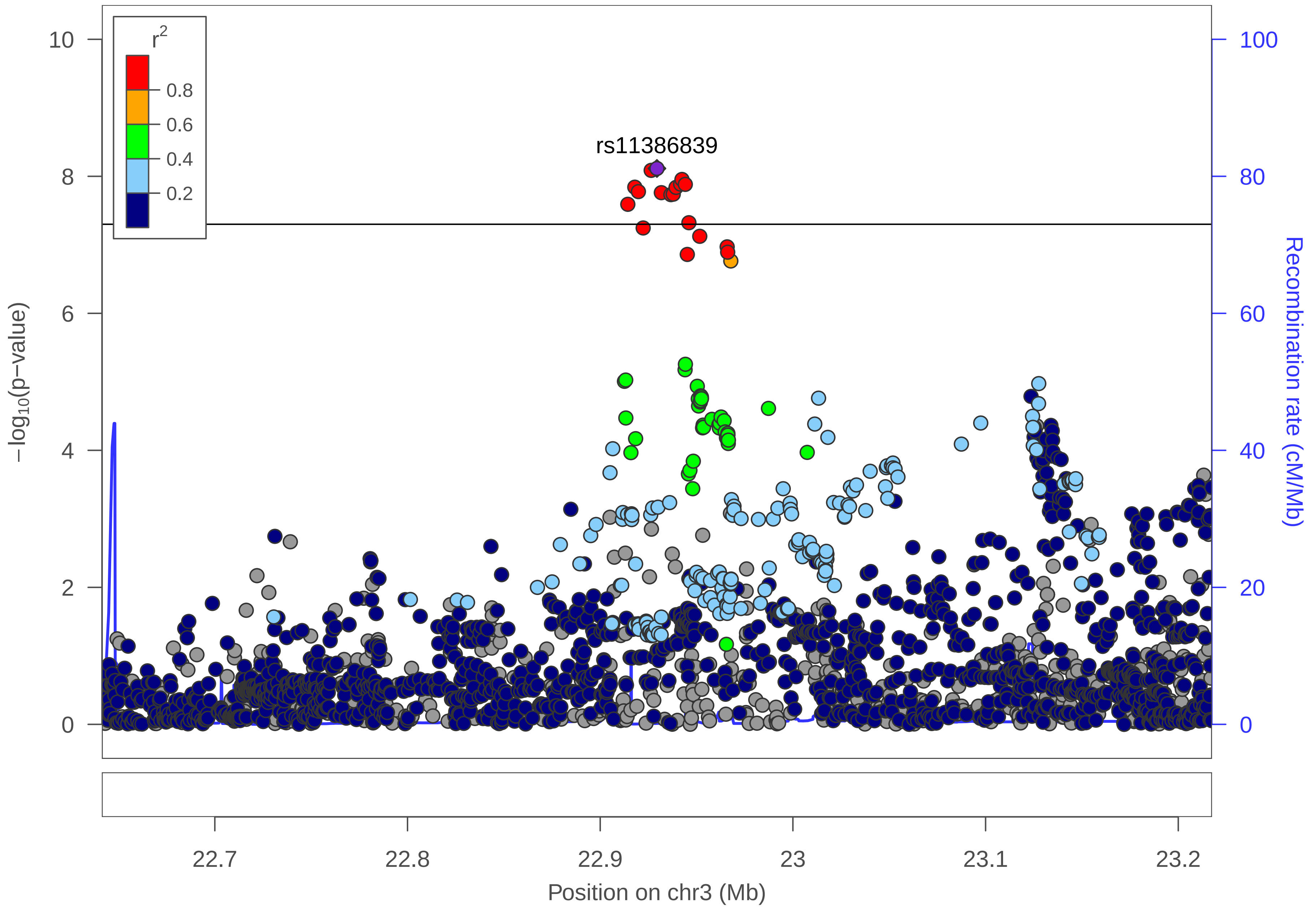

r2
0.8
0.6
0.4
0.2
rs11386839
−log10(p−value)
Recombination rate (cM/Mb)
0
2
4
6
8
10
0
20
40
60
80
100
22.7
22.8
22.9
23
23.1
23.2
Position on chr3 (Mb)

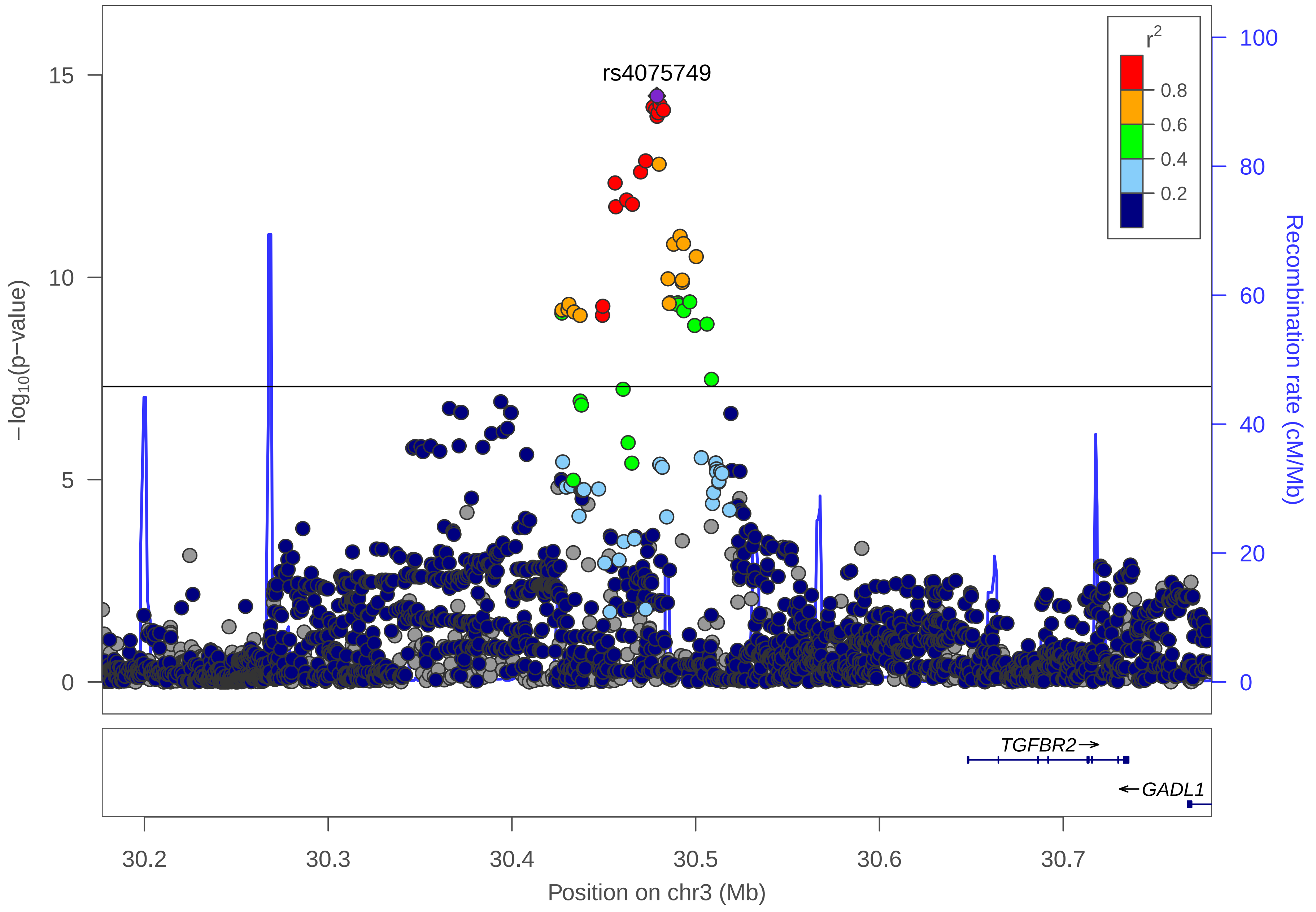

rs4075749
r2
0.8
0.6
0.4
0.2
15
10
5
0
−log10(p−value)
100
80
60
40
20
0
Recombination rate (cM/Mb)
TGFBR2→
←GADL1
30.2
30.3
30.4
30.5
30.6
30.7
Position on chr3 (Mb)

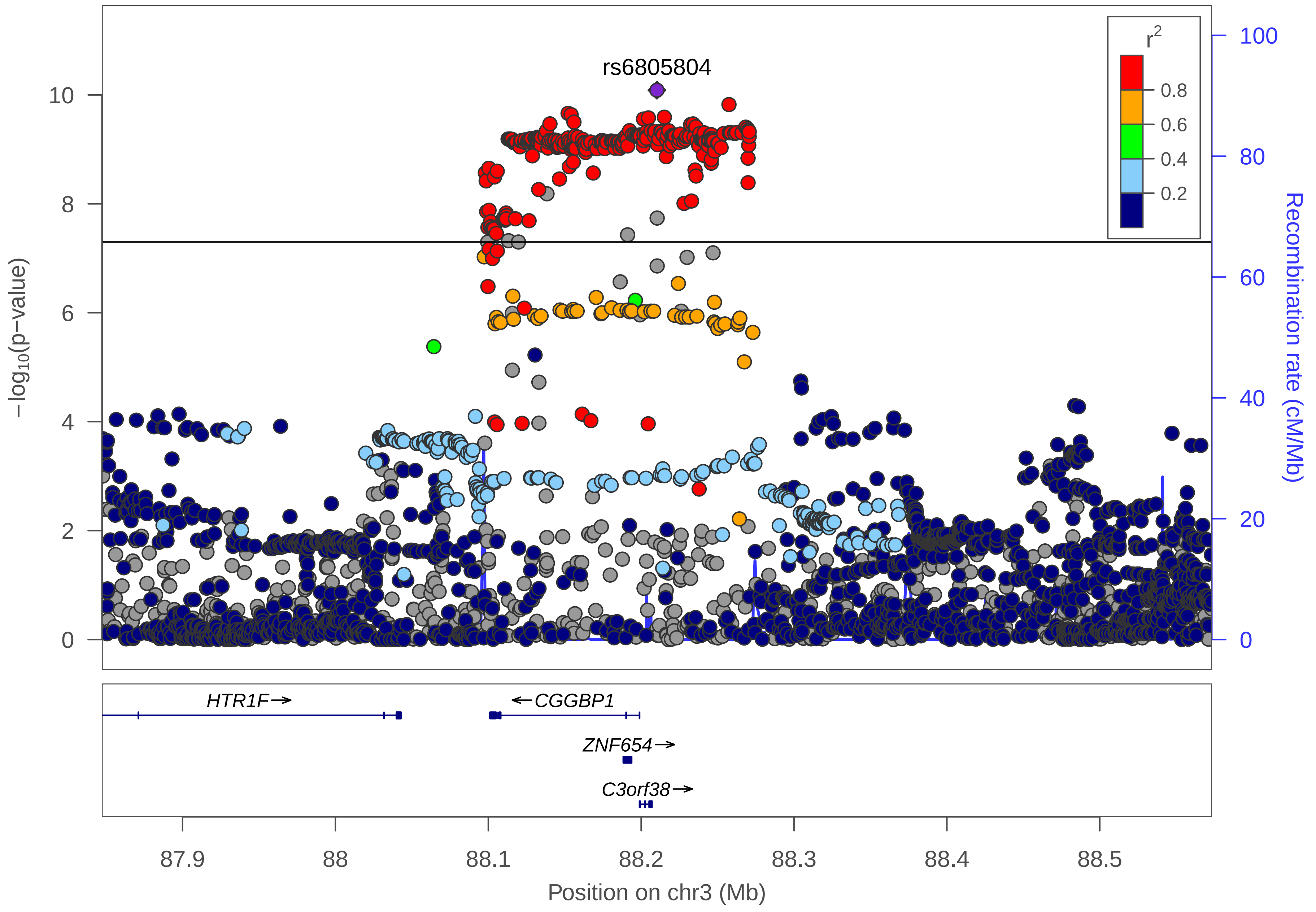

rs6805804
r2
0.8
0.6
0.4
0.2
−log10(p−value)
Recombination rate (cM/Mb)
HTR1F→
←CGGBP1
ZNF654→
C3orf38→
87.9
88
88.1
88.2
88.3
88.4
88.5
Position on chr3 (Mb)

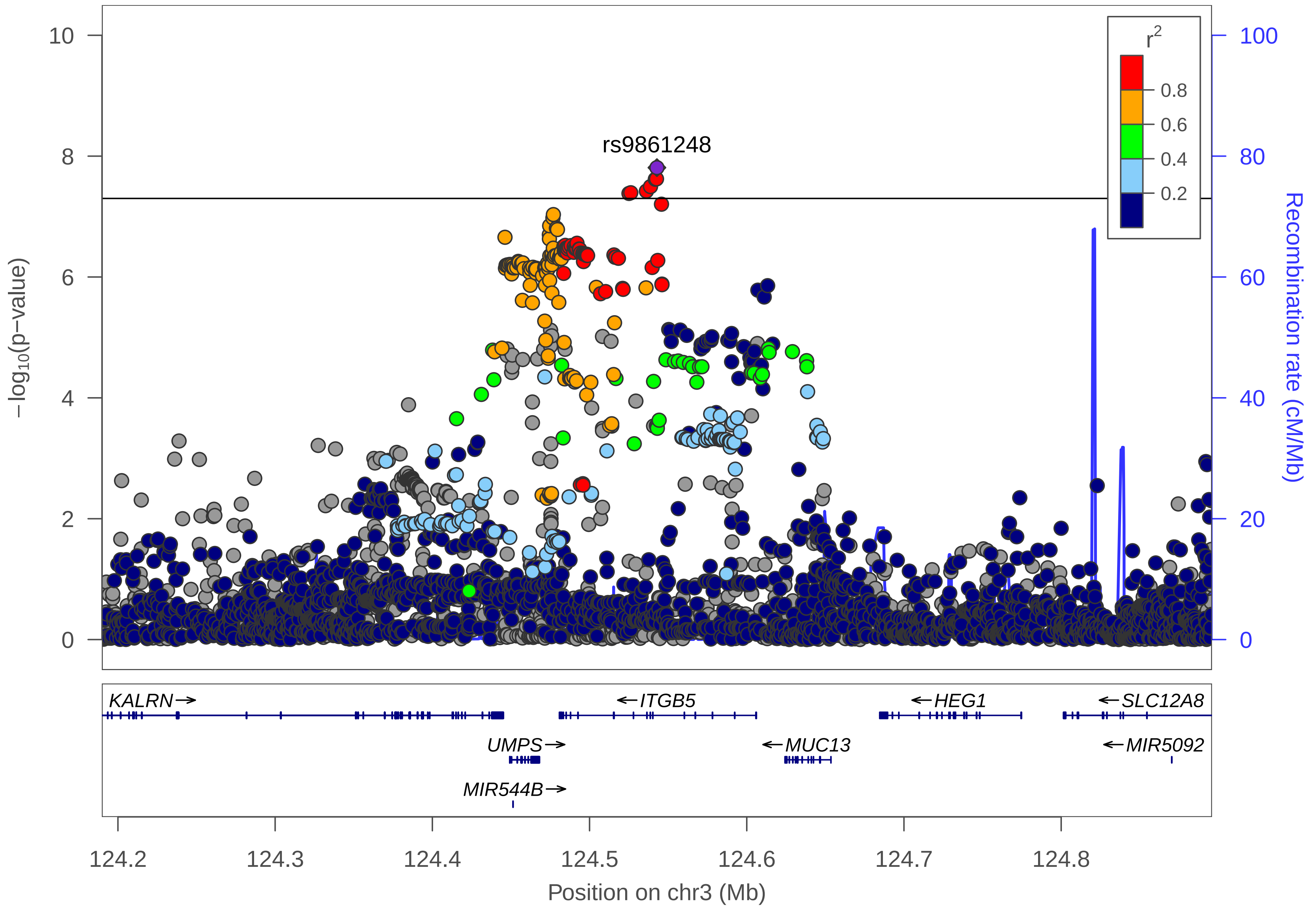
rs9861248
r2
0.8
0.6
0.4
0.2
10
8
6
4
2
0
100
80
60
40
20
0
−log10(p−value)
Recombination rate (cM/Mb)
KALRN→
←ITGB5
←HEG1
←SLC12A8
UMPS→
←MUC13
←MIR5092
MIR544B→
124.2
124.3
124.4
124.5
124.6
124.7
124.8
Position on chr3 (Mb)

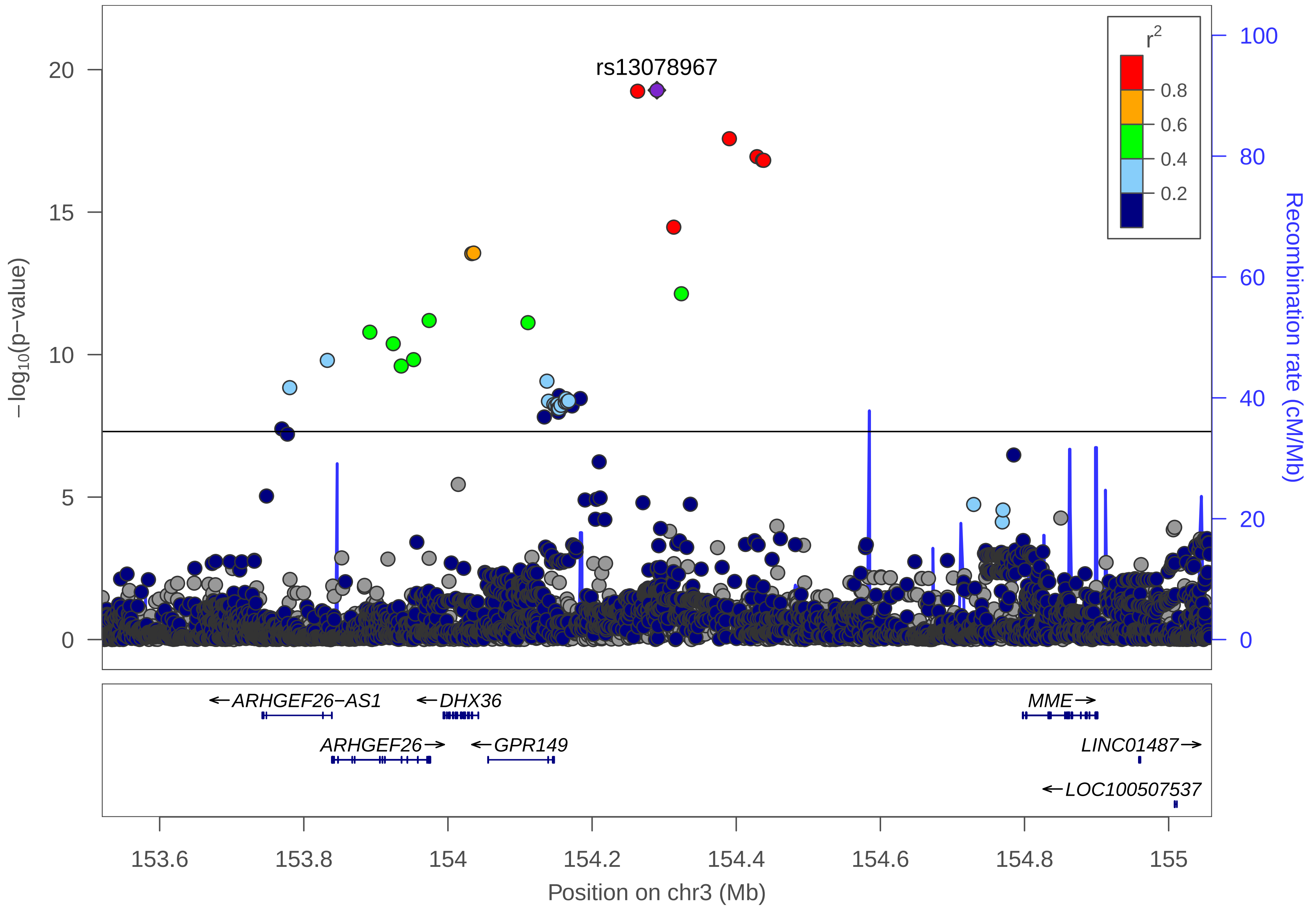
rs13078967
r2
0.8
0.6
0.4
0.2
−log10(p−value)
Recombination rate (cM/Mb)
ARHGEF26−AS1
DHX36
MME
ARHGEF26
GPR149
LINC01487
LOC100507537
153.6
153.8
154
154.2
154.4
154.6
154.8
155
Position on chr3 (Mb)

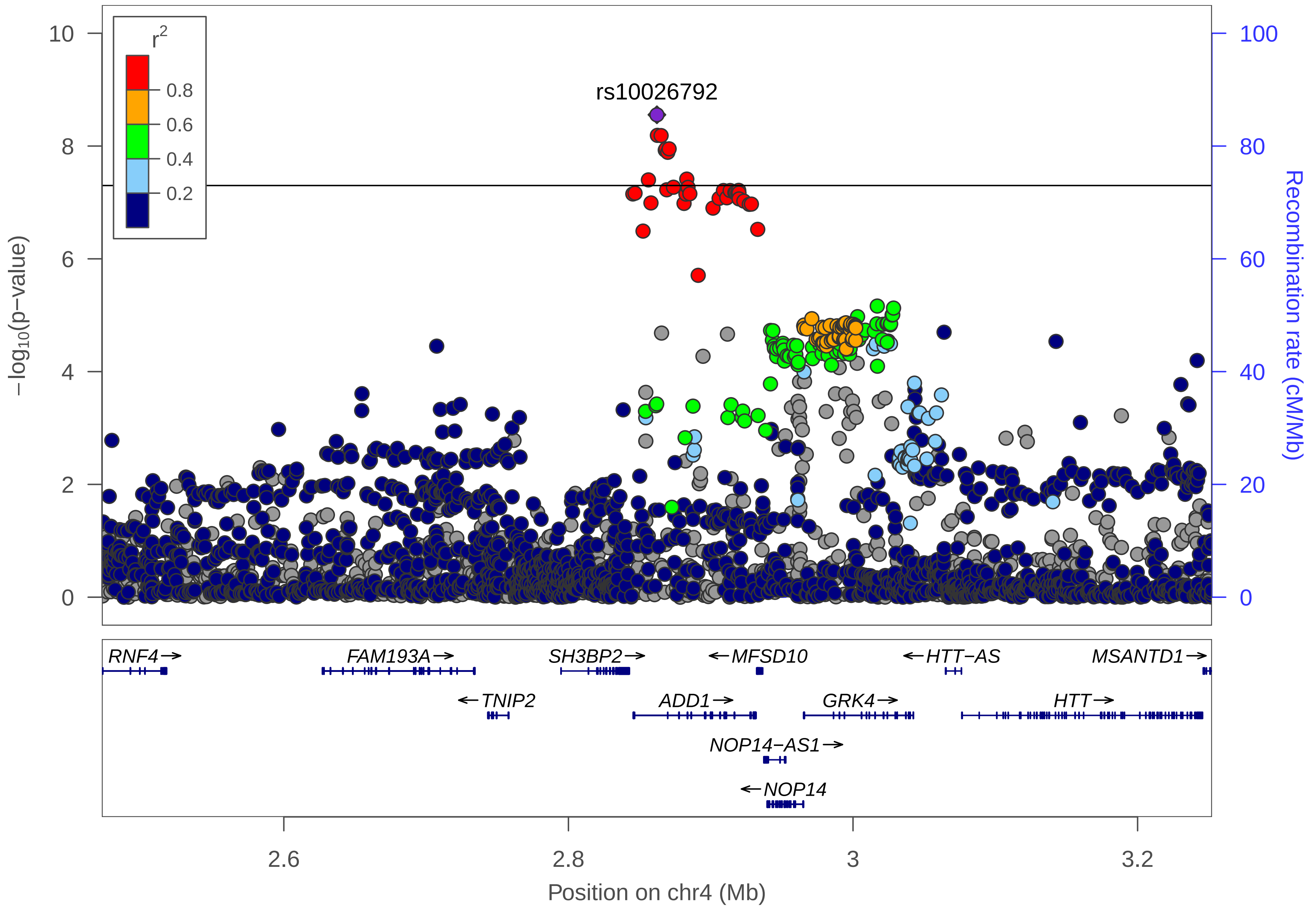

r2
0.8
0.6
0.4
0.2
rs10026792
10
8
6
4
2
0
100
80
60
40
20
0
−log10(p−value)
Recombination rate (cM/Mb)
RNF4→
FAM193A→
SH3BP2→
←MFSD10
←HTT−AS
MSANTD1→
←TNIP2
ADD1→
GRK4→
HTT→
NOP14−AS1→
←NOP14
2.6
2.8
3
3.2
Position on chr4 (Mb)

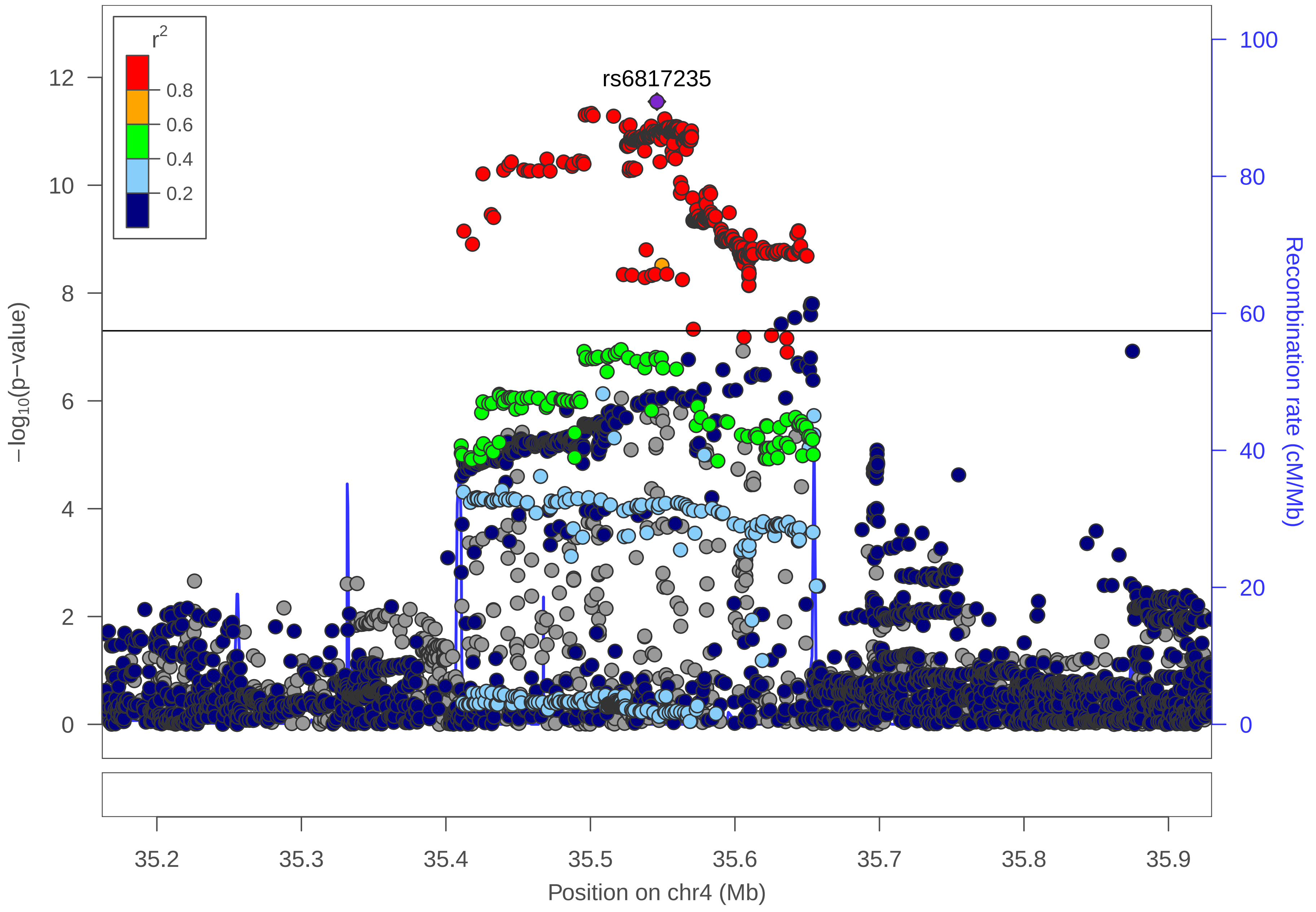

r2
0.8
0.6
0.4
0.2
rs6817235
12
10
8
6
4
2
0
−log10(p−value)
100
80
60
40
20
0
Recombination rate (cM/Mb)
35.2
35.3
35.4
35.5
35.6
35.7
35.8
35.9
Position on chr4 (Mb)

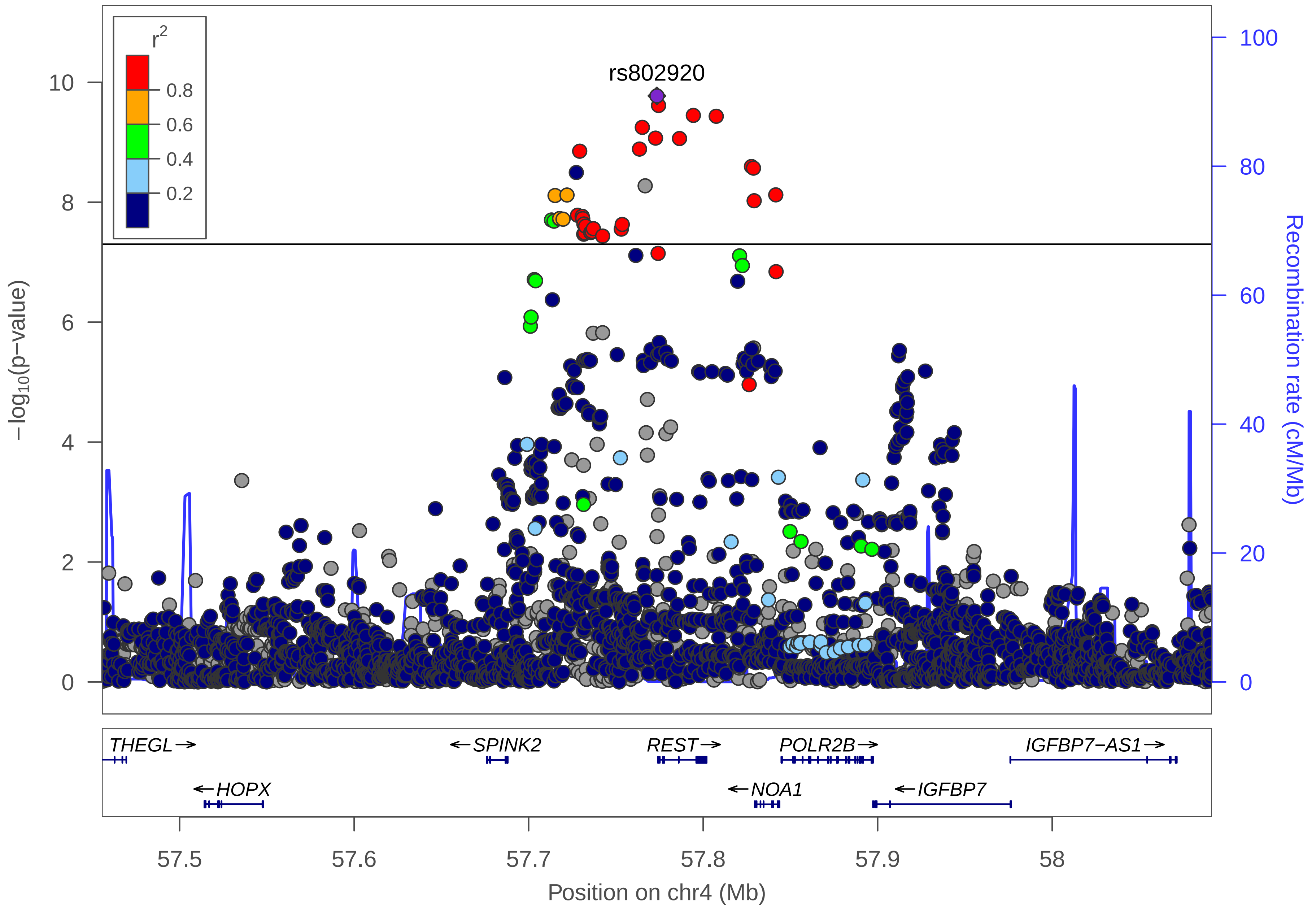

r^2
0.8
0.6
0.4
0.2
rs802920
10
8
6
4
2
0
−log10(p−value)
100
80
60
40
20
0
Recombination rate (cM/Mb)
THEGL→
←SPINK2
REST→
POLR2B→
IGFBP7−AS1→
←HOPX
←NOA1
←IGFBP7
57.5
57.6
57.7
57.8
57.9
58
Position on chr4 (Mb)

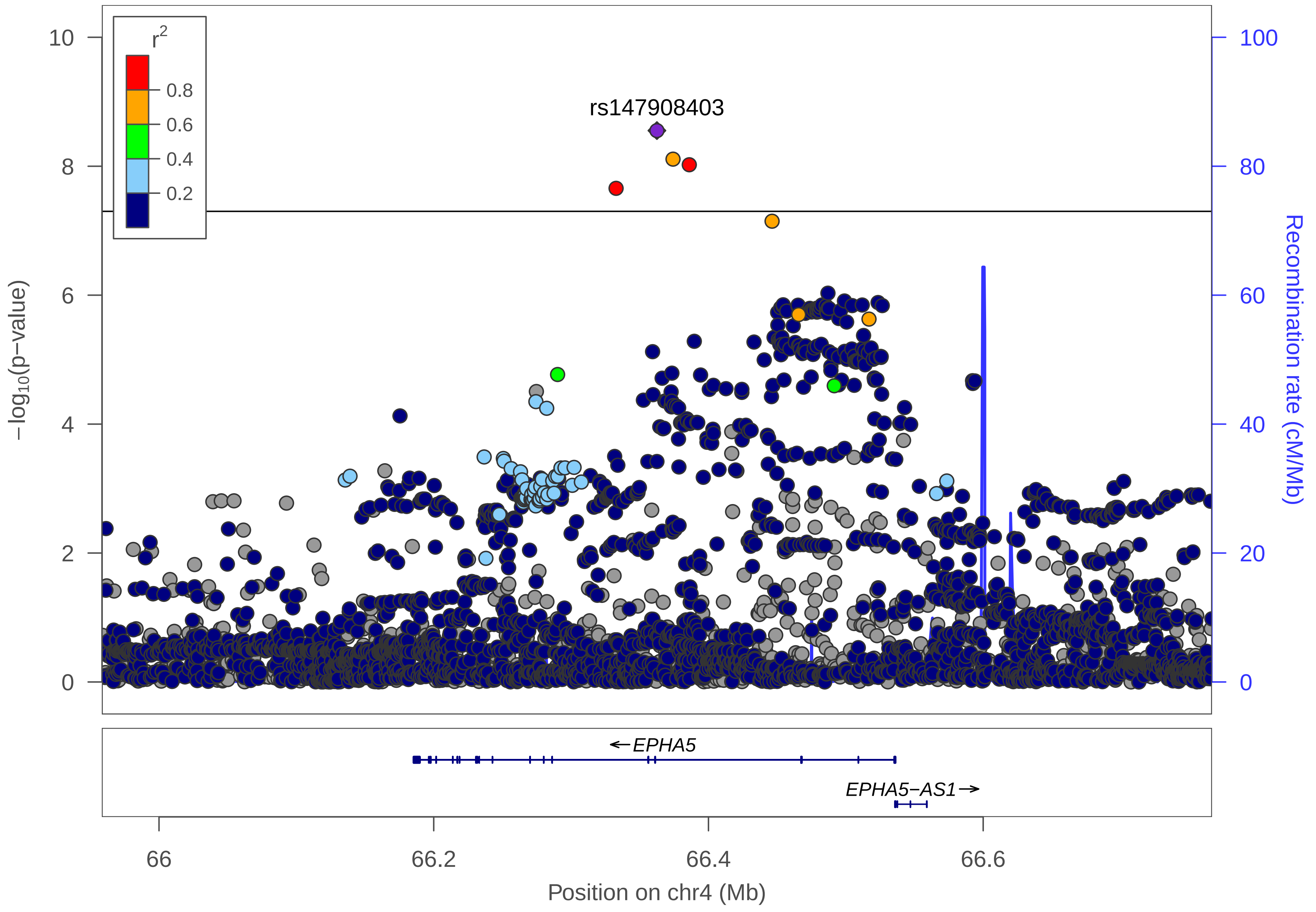

r2
0.8
0.6
0.4
0.2
rs147908403
−log10(p−value)
Recombination rate (cM/Mb)
10
8
6
4
2
0
100
80
60
40
20
0
←EPHA5
EPHA5−AS1→
66
66.2
66.4
66.6
Position on chr4 (Mb)

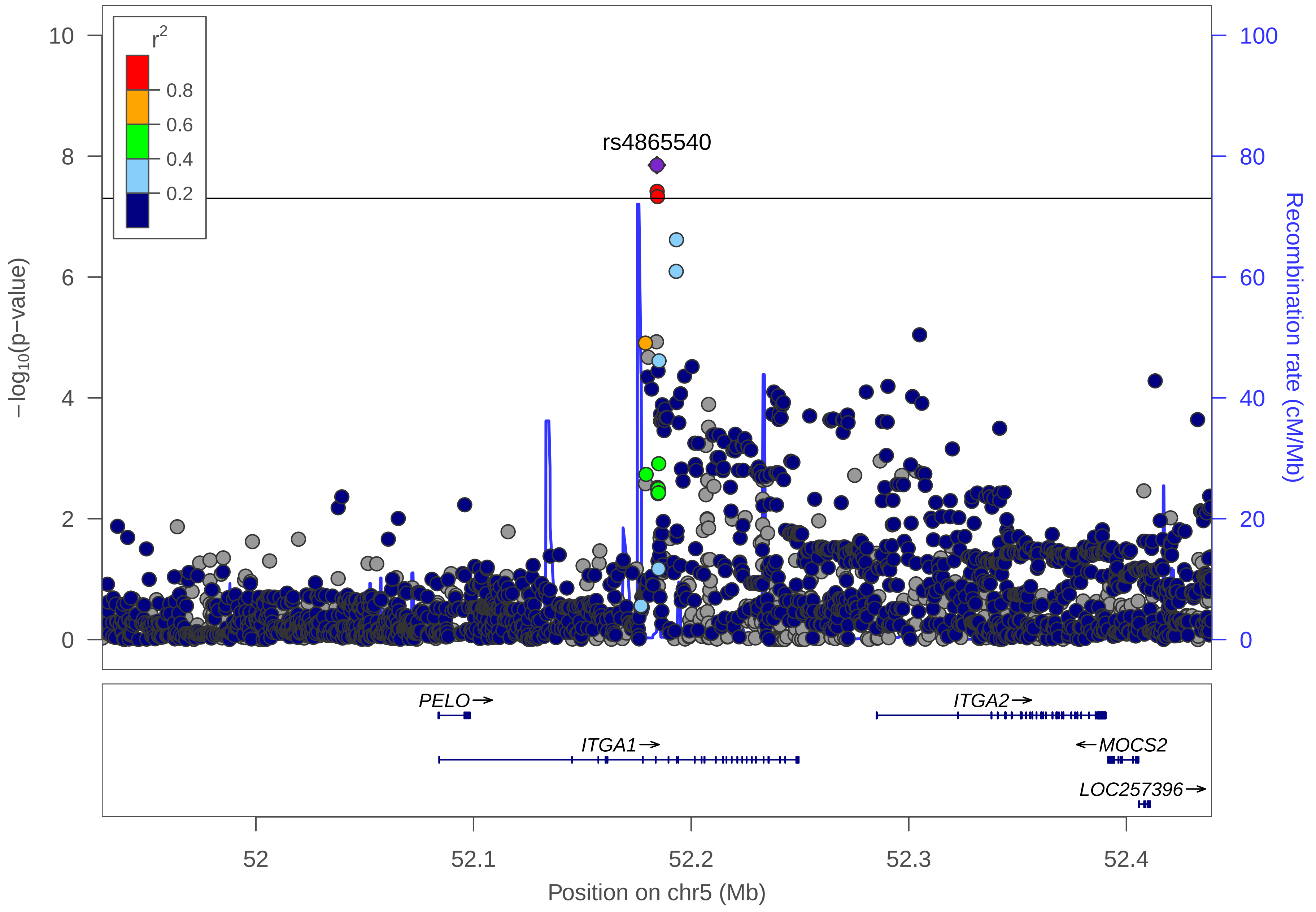

r2
0.8
0.6
0.4
0.2
rs4865540
−log10(p−value)
Recombination rate (cM/Mb)
10
8
6
4
2
0
100
80
60
40
20
0
PELO→
ITGA2→
ITGA1→
←MOCS2
LOC257396→
52
52.1
52.2
52.3
52.4
Position on chr5 (Mb)

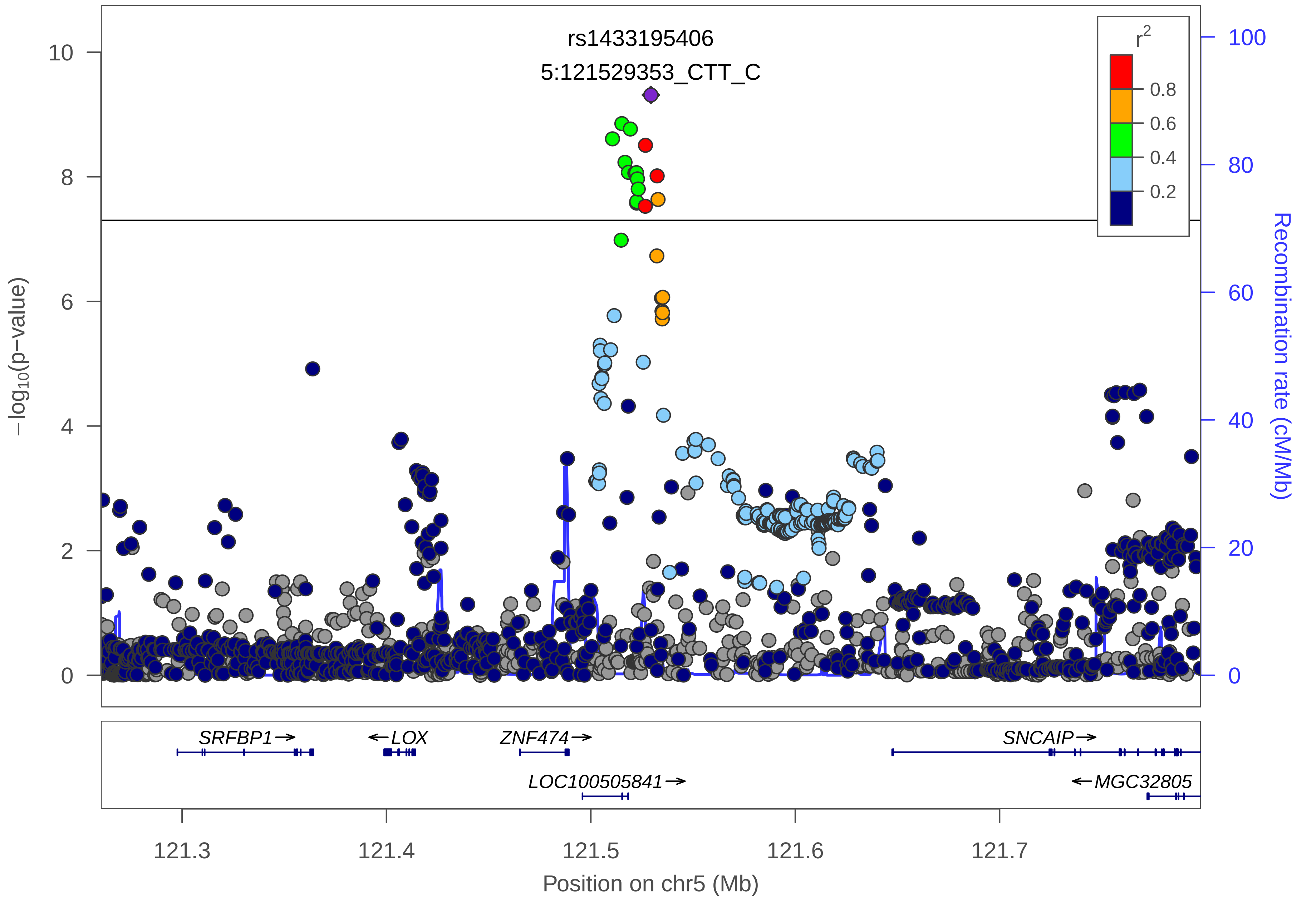

rs1433195406
5:121529353_CTT_C
r2
0.8
0.6
0.4
0.2
−log10(p−value)
Recombination rate (cM/Mb)
10
8
6
4
2
0
100
80
60
40
20
0
SRFBP1→
←LOX
ZNF474→
SNCAIP→
LOC100505841→
←MGC32805
121.3
121.4
121.5
121.6
121.7
Position on chr5 (Mb)

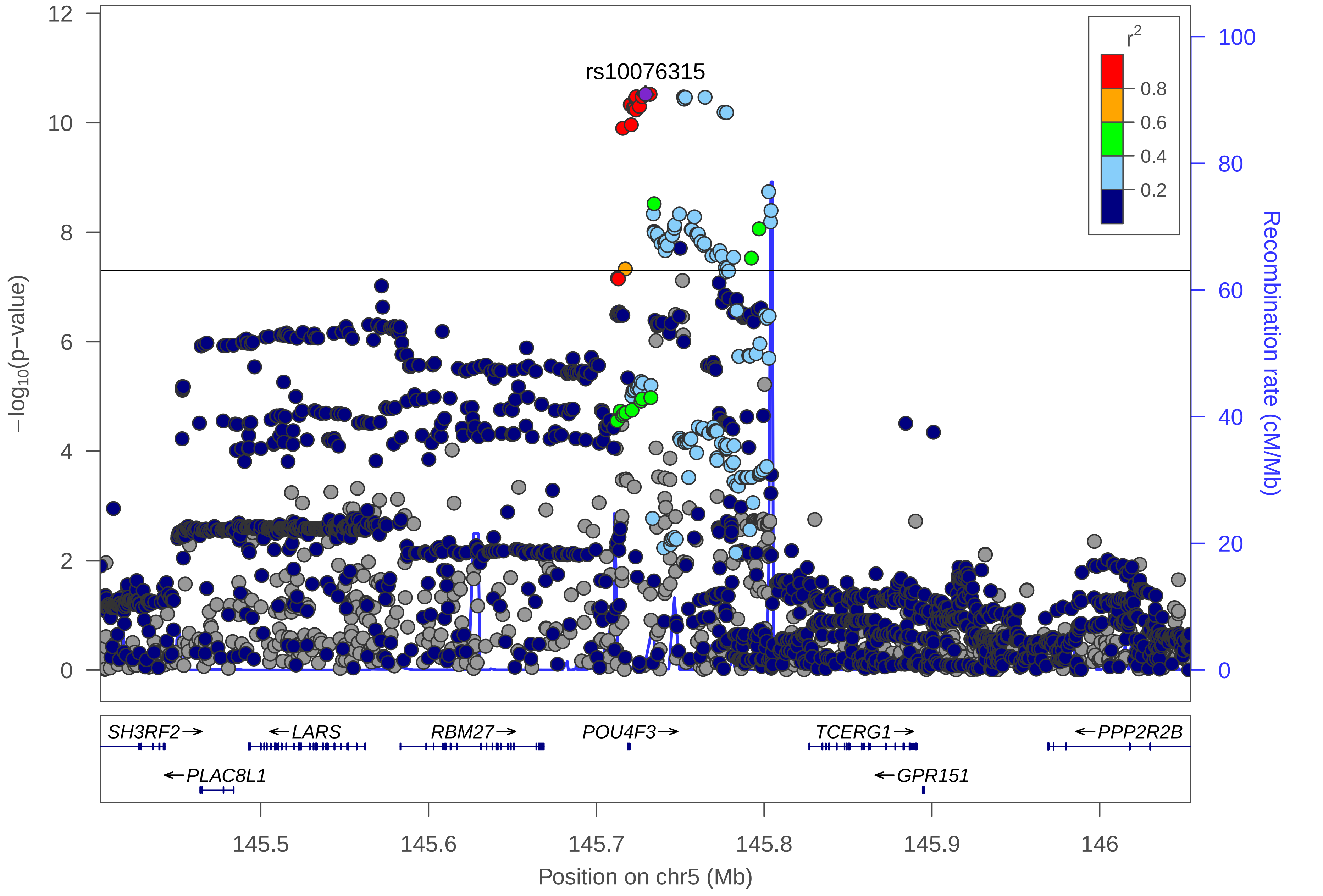
rs10076315
r2
0.8
0.6
0.4
0.2
−log10(p−value)
Recombination rate (cM/Mb)
12
10
8
6
4
2
0
100
80
60
40
20
0
SH3RF2→
←LARS
RBM27→
POU4F3→
TCERG1→
←PPP2R2B
←PLAC8L1
←GPR151
145.5
145.6
145.7
145.8
145.9
146
Position on chr5 (Mb)

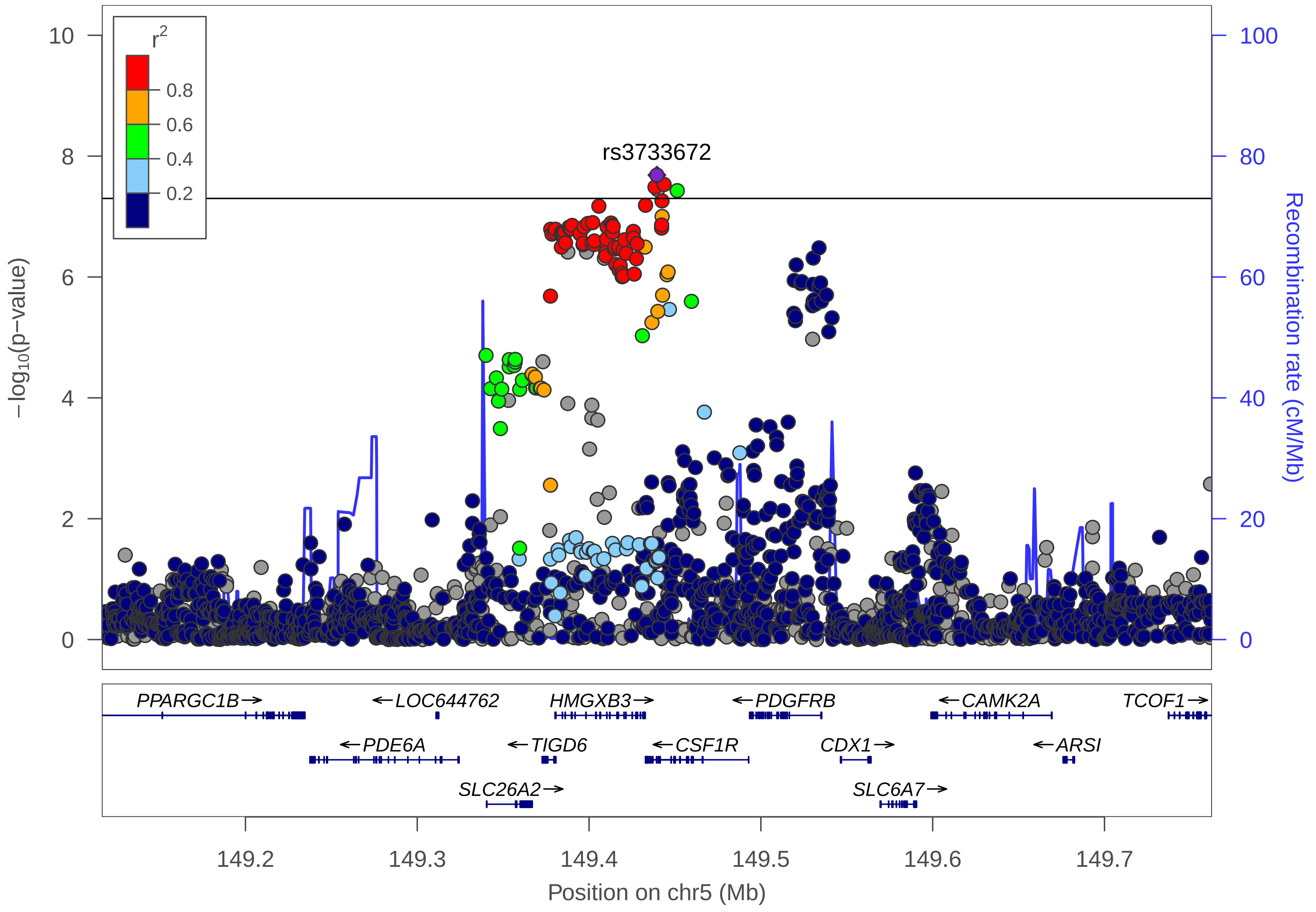
r2
0.8
0.6
0.4
0.2
rs3733672
10
8
6
4
2
0
−log10(p−value)
100
80
60
40
20
0
Recombination rate (cM/Mb)
PPARGC1B→
←LOC644762
HMGXB3→
←PDGFRB
←CAMK2A
TCOF1→
←PDE6A
←TIGD6
←CSF1R
CDX1→
←ARSI
SLC26A2→
SLC6A7→
149.2
149.3
149.4
149.5
149.6
149.7
Position on chr5 (Mb)

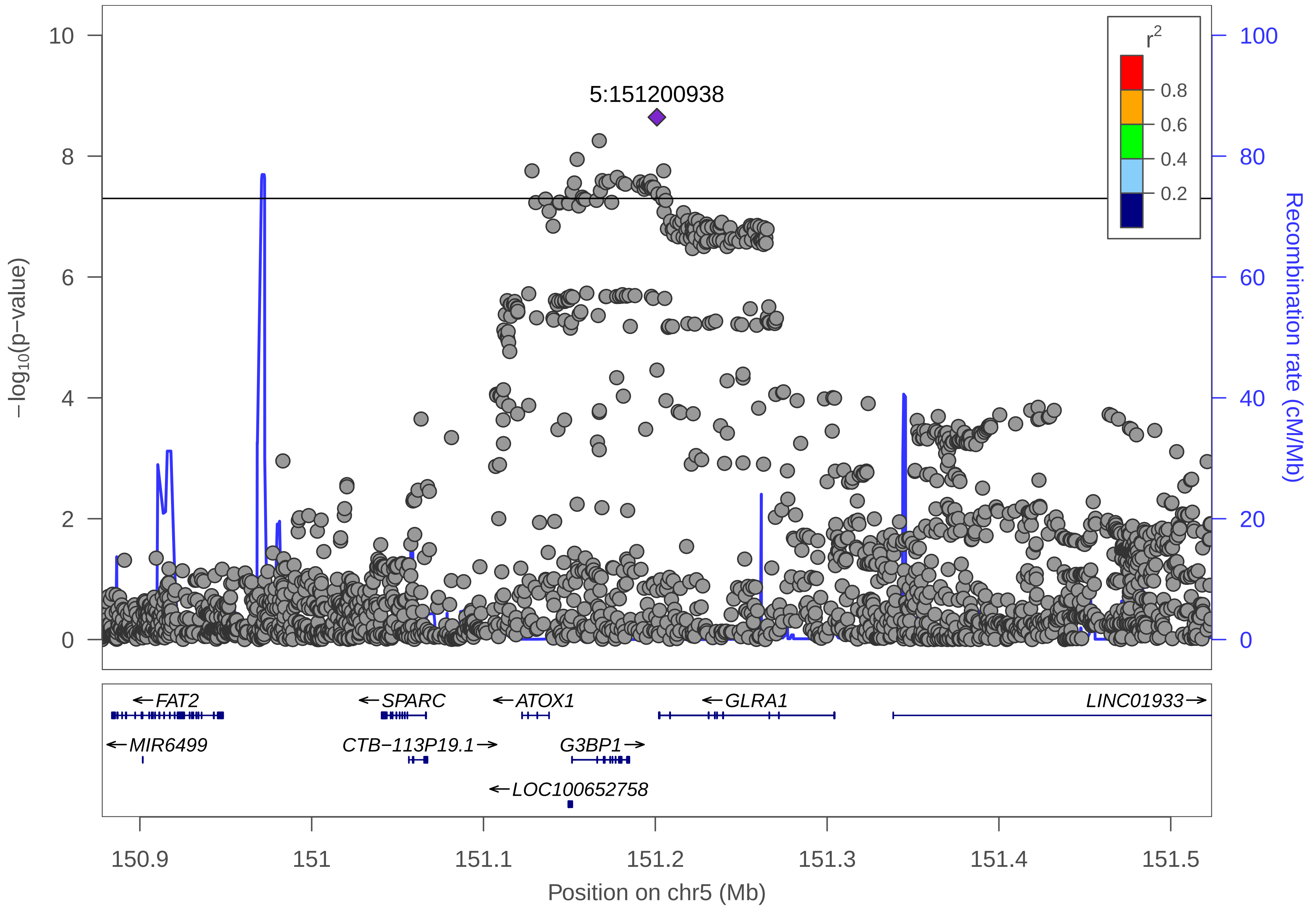

5:151200938
r2
0.8
0.6
0.4
0.2
−log10(p−value)
Recombination rate (cM/Mb)
0
2
4
6
8
10
0
20
40
60
80
100
FAT2
SPARC
ATOX1
GLRA1
LINC01933
MIR6499
CTB−113P19.1
G3BP1
LOC100652758
150.9
151
151.1
151.2
151.3
151.4
151.5
Position on chr5 (Mb)

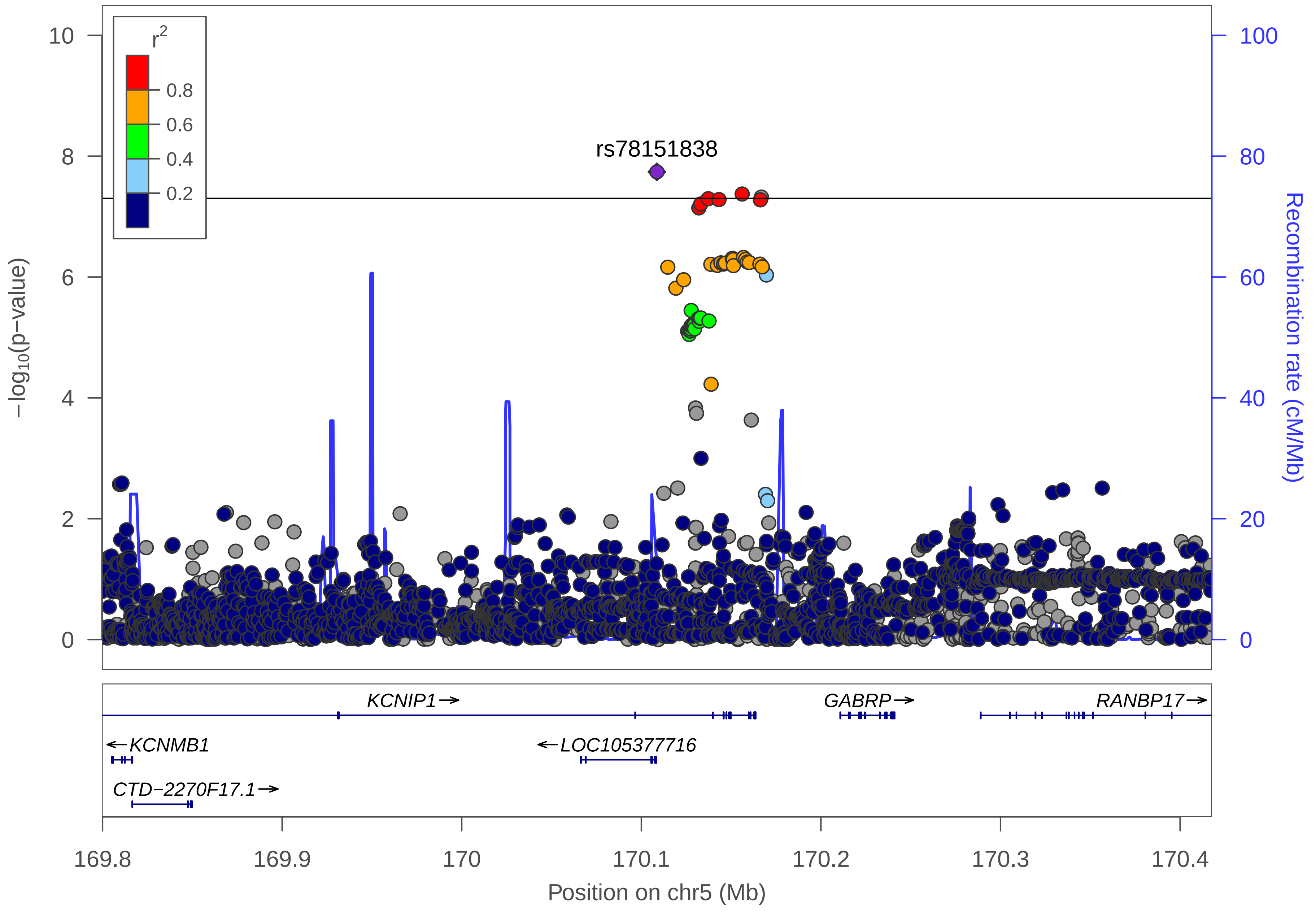

r²
0.8
0.6
0.4
0.2
rs78151838
10
8
6
4
2
0
−log10(p−value)
100
80
60
40
20
0
Recombination rate (cM/Mb)
KCNIP1→
GABRP→
RANBP17→
←KCNMB1
←LOC105377716
CTD−2270F17.1→
169.8
169.9
170
170.1
170.2
170.3
170.4
Position on chr5 (Mb)

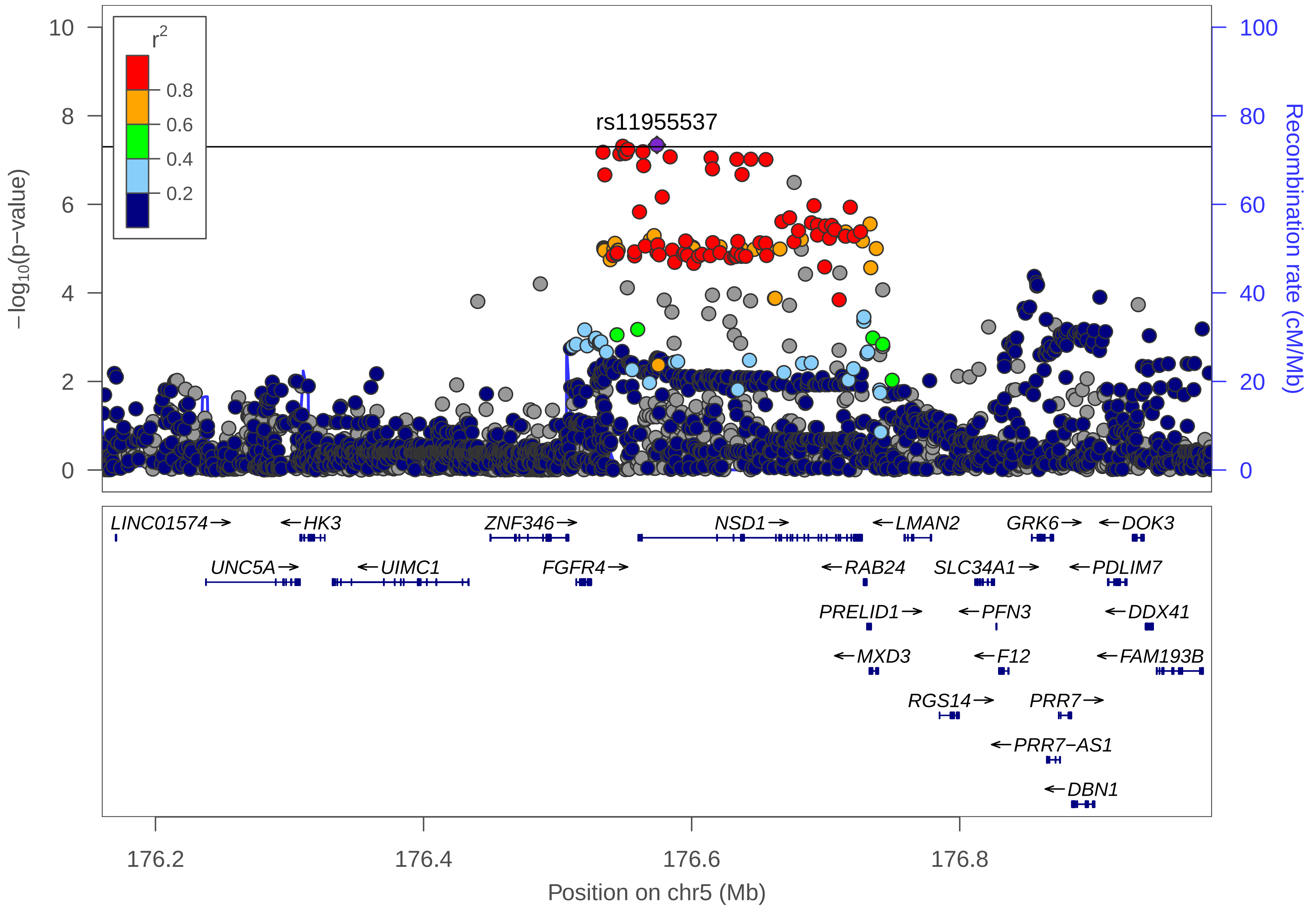

rs11955537
−log10(p−value)
Recombination rate (cM/Mb)
r2
0.8
0.6
0.4
0.2
0
2
4
6
8
10
0
20
40
60
80
100
LINC01574→
←HK3
ZNF346→
NSD1→
←LMAN2
GRK6→
←DOK3
UNC5A→
←UIMC1
FGFR4→
←RAB24
SLC34A1→
←PDLIM7
PRELID1→
←PFN3
←DDX41
←MXD3
←F12
←FAM193B
RGS14→
PRR7→
←PRR7−AS1
←DBN1
176.2
176.4
176.6
176.8
Position on chr5 (Mb)

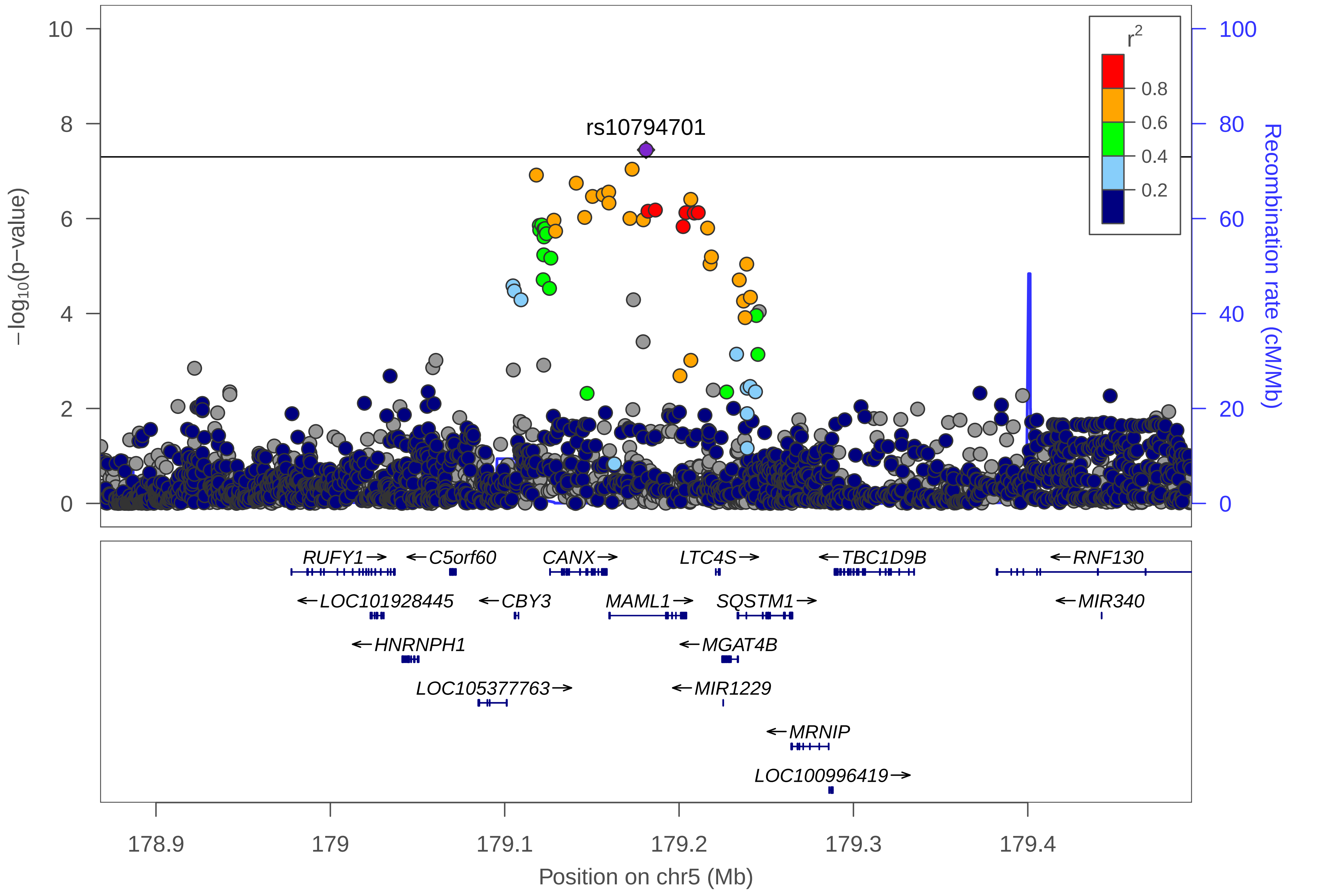

rs10794701
r2
0.8
0.6
0.4
0.2
−log10(p−value)
Recombination rate (cM/Mb)
0
2
4
6
8
10
0
20
40
60
80
100
RUFY1→
←C5orf60
CANX→
LTC4S→
←TBC1D9B
←RNF130
←LOC101928445
←CBY3
MAML1→
SQSTM1→
←MIR340
←HNRNPH1
←MGAT4B
LOC105377763→
←MIR1229
←MRNIP
LOC100996419→
178.9
179
179.1
179.2
179.3
179.4
Position on chr5 (Mb)

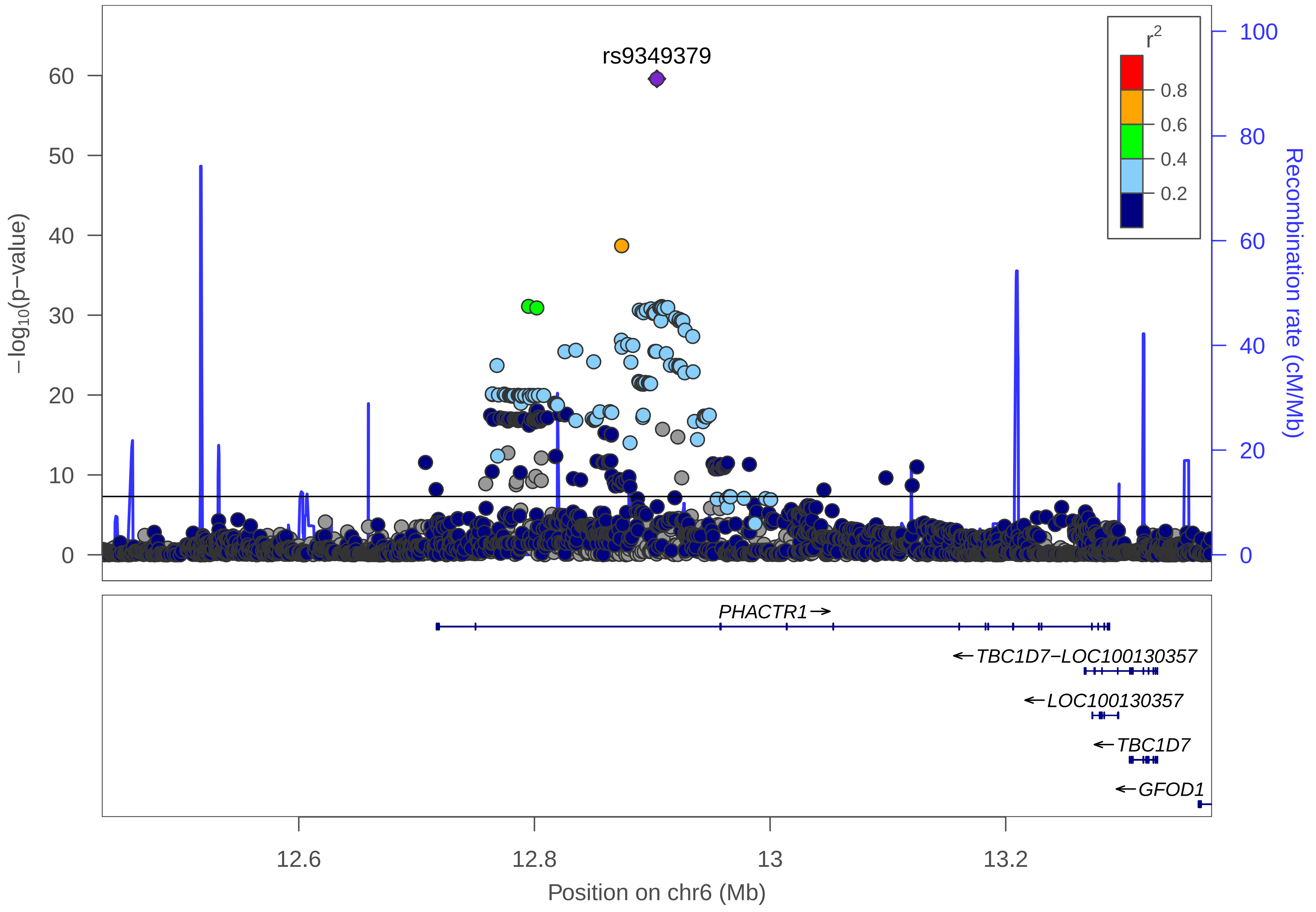

rs9349379
r2
0.8
0.6
0.4
0.2
−log10(p−value)
0
10
20
30
40
50
60
Recombination rate (cM/Mb)
0
20
40
60
80
100
PHACTR1
TBC1D7−LOC100130357
LOC100130357
TBC1D7
GFOD1
12.6
12.8
13
13.2
Position on chr6 (Mb)

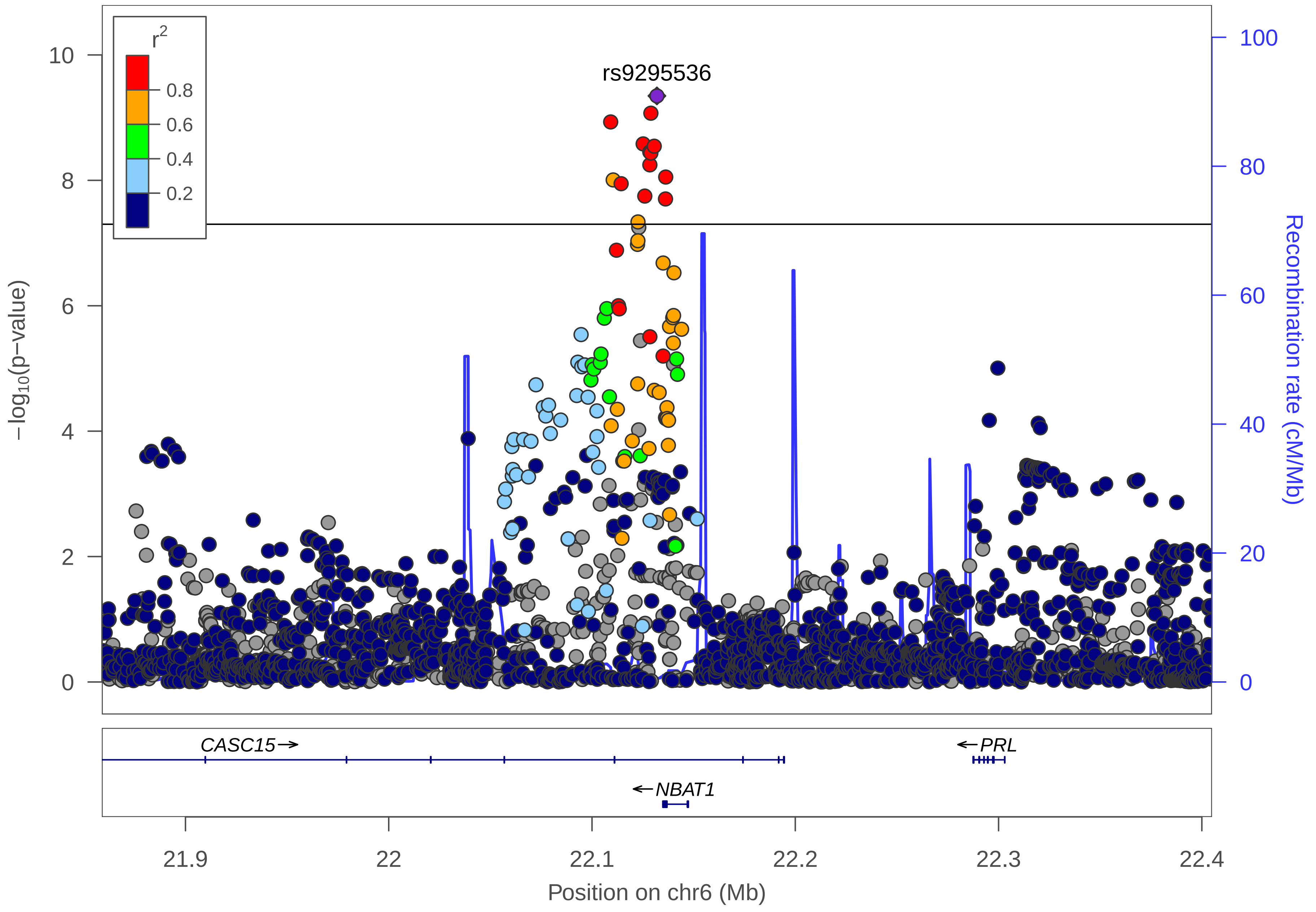

r2
0.8
0.6
0.4
0.2
rs9295536
−log10(p−value)
Recombination rate (cM/Mb)
10
8
6
4
2
0
100
80
60
40
20
0
CASC15→
←PRL
←NBAT1
21.9
22
22.1
22.2
22.3
22.4
Position on chr6 (Mb)

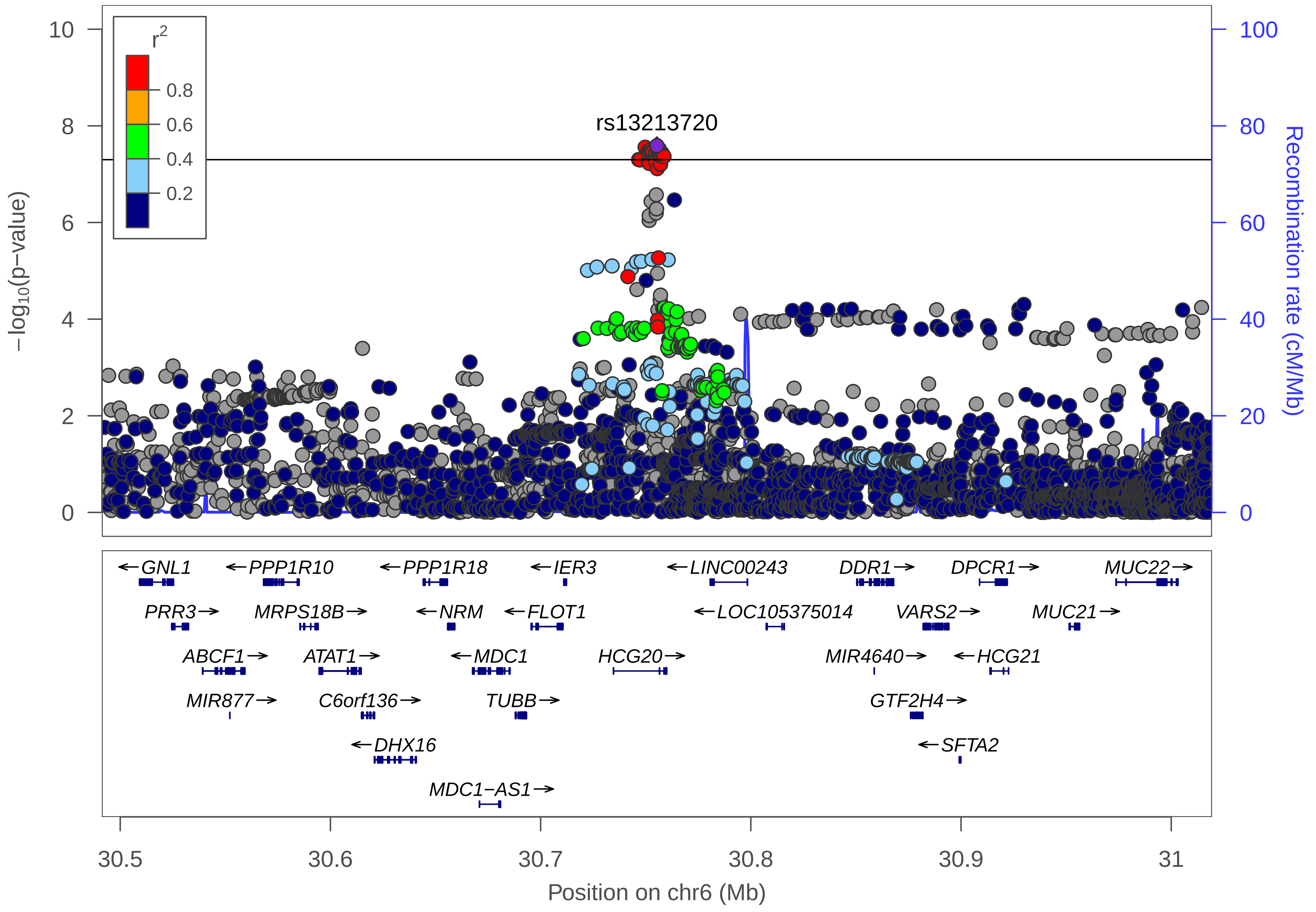

rs13213720
r2
0.8
0.6
0.4
0.2
−log10(p−value)
Recombination rate (cM/Mb)
10
8
6
4
2
0
100
80
60
40
20
0
←GNL1
←PPP1R10
←PPP1R18
←IER3
←LINC00243
DDR1→
DPCR1→
MUC22→
PRR3→
MRPS18B→
←NRM
←FLOT1
←LOC105375014
VARS2→
MUC21→
ABCF1→
ATAT1→
←MDC1
HCG20→
MIR4640→
←HCG21
MIR877→
C6orf136→
TUBB→
GTF2H4→
←DHX16
←SFTA2
MDC1−AS1→
30.5
30.6
30.7
30.8
30.9
31
Position on chr6 (Mb)

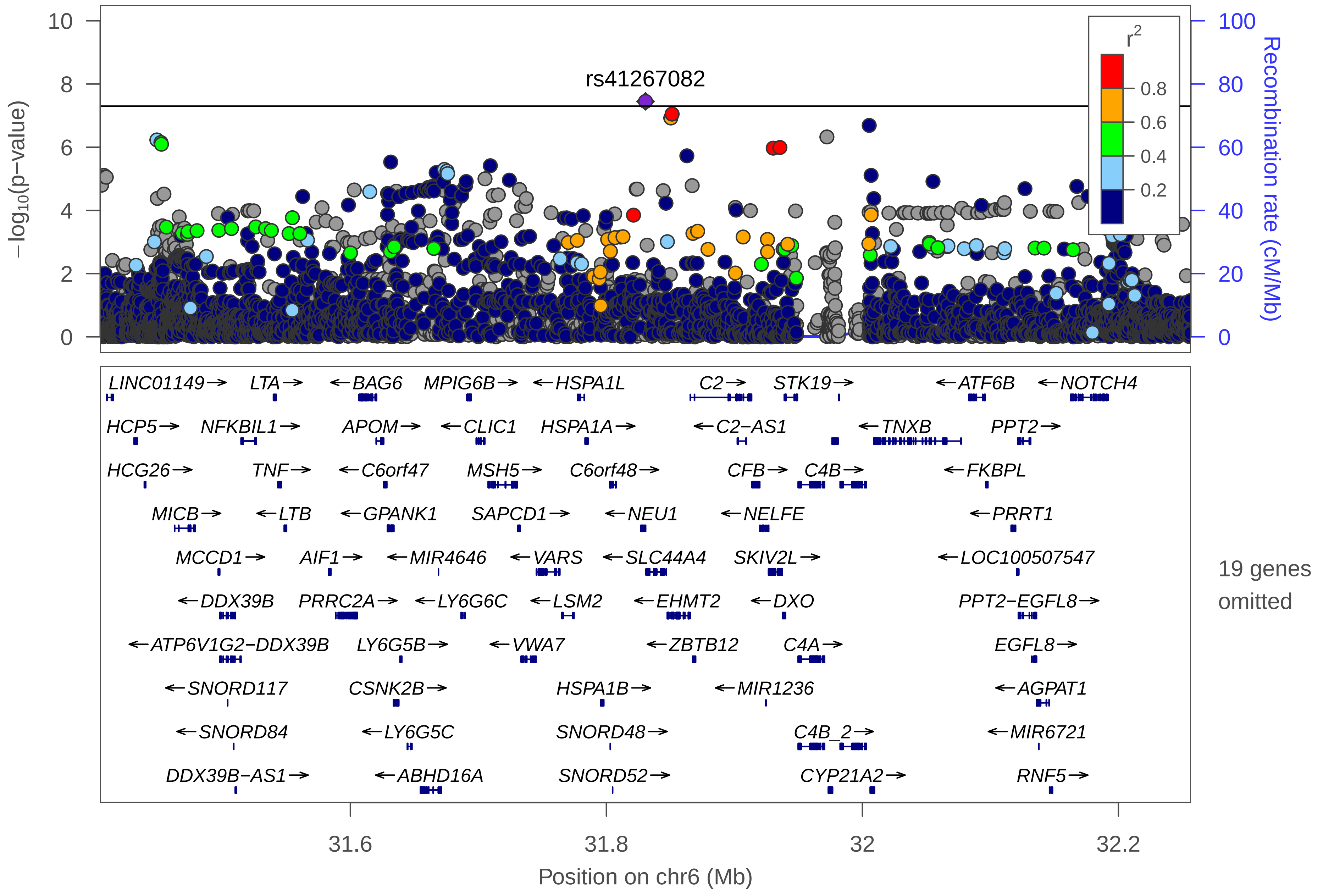

rs41267082
r2
0.8
0.6
0.4
0.2
−log10(p−value)
Recombination rate (cM/Mb)
LINC01149→ LTA→ ←BAG6 MPIG6B→ ←HSPA1L C2→ STK19→ ←ATF6B ←NOTCH4
HCP5→ NFKBIL1→ APOM→ ←CLIC1 HSPA1A→ ←C2−AS1 ←TNXB PPT2→
HCG26→ TNF→ ←C6orf47 MSH5→ C6orf48→ CFB→ C4B→ ←FKBPL
MICB→ ←LTB ←GPANK1 SAPCD1→ ←NEU1 ←NELFE ←PRRT1
MCCD1→ AIF1→ ←MIR4646 ←VARS ←SLC44A4 SKIV2L→ ←LOC100507547
←DDX39B PRRC2A→ ←LY6G6C ←LSM2 ←EHMT2 ←DXO PPT2−EGFL8→
←ATP6V1G2−DDX39B LY6G5B→ ←VWA7 ←ZBTB12 C4A→ EGFL8→
←SNORD117 CSNK2B→ HSPA1B→ ←MIR1236 ←AGPAT1
←SNORD84 ←LY6G5C SNORD48→ C4B_2→ ←MIR6721
DDX39B−AS1→ ←ABHD16A SNORD52→ CYP21A2→ RNF5→
19 genes omitted
31.6
31.8
32
32.2
Position on chr6 (Mb)

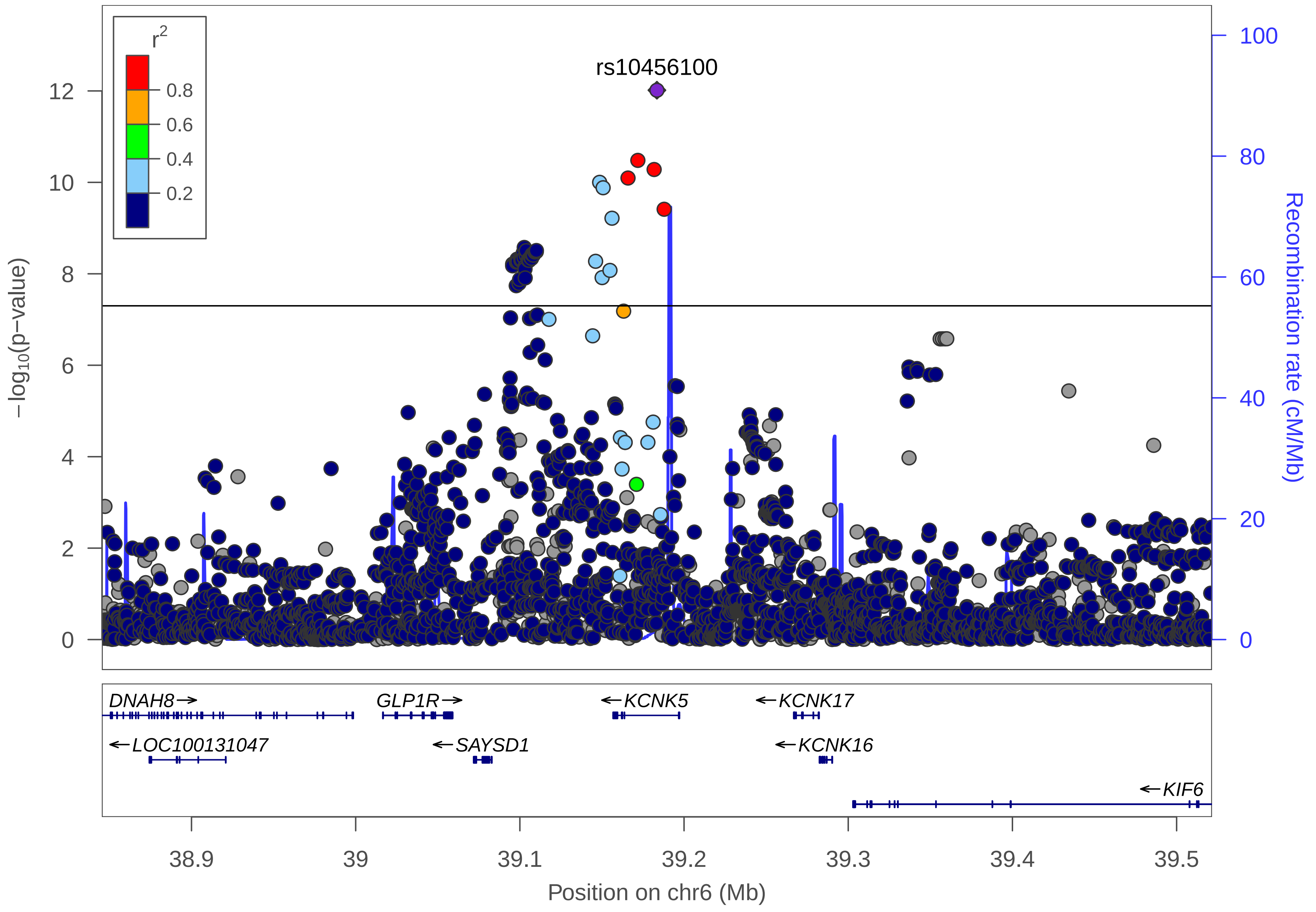

r2
0.8
0.6
0.4
0.2
rs10456100
−log10(p−value)
Recombination rate (cM/Mb)
0
2
4
6
8
10
12
20
40
60
80
100
DNAH8→
GLP1R→
←KCNK5
←KCNK17
←LOC100131047
←SAYSD1
←KCNK16
←KIF6
38.9
39
39.1
39.2
39.3
39.4
39.5
Position on chr6 (Mb)

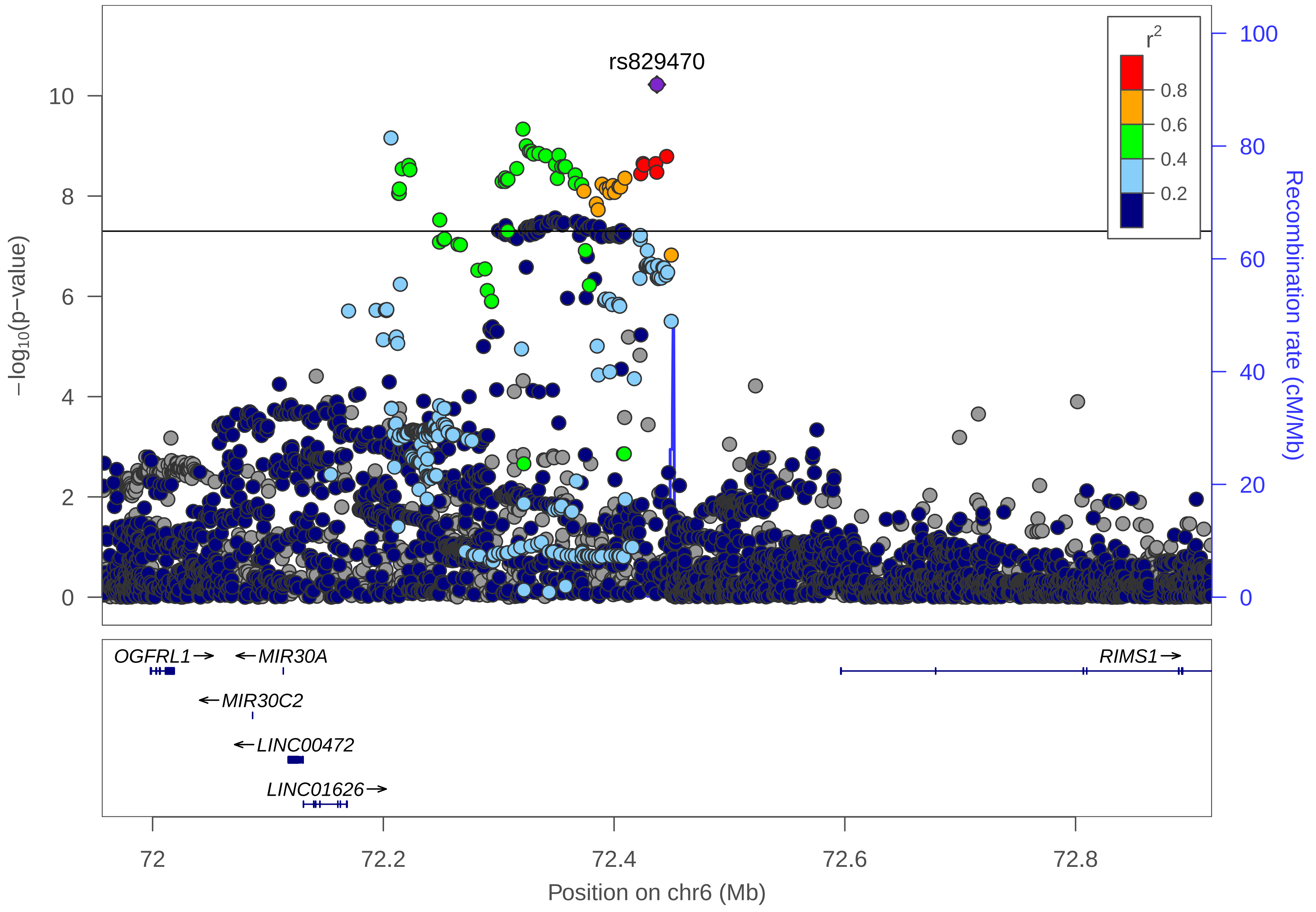

rs829470
r²
0.8
0.6
0.4
0.2
−log10(p−value)
0
2
4
6
8
10
Recombination rate (cM/Mb)
0
20
40
60
80
100
OGFRL1→
←MIR30A
←MIR30C2
←LINC00472
LINC01626→
RIMS1→
72
72.2
72.4
72.6
72.8
Position on chr6 (Mb)

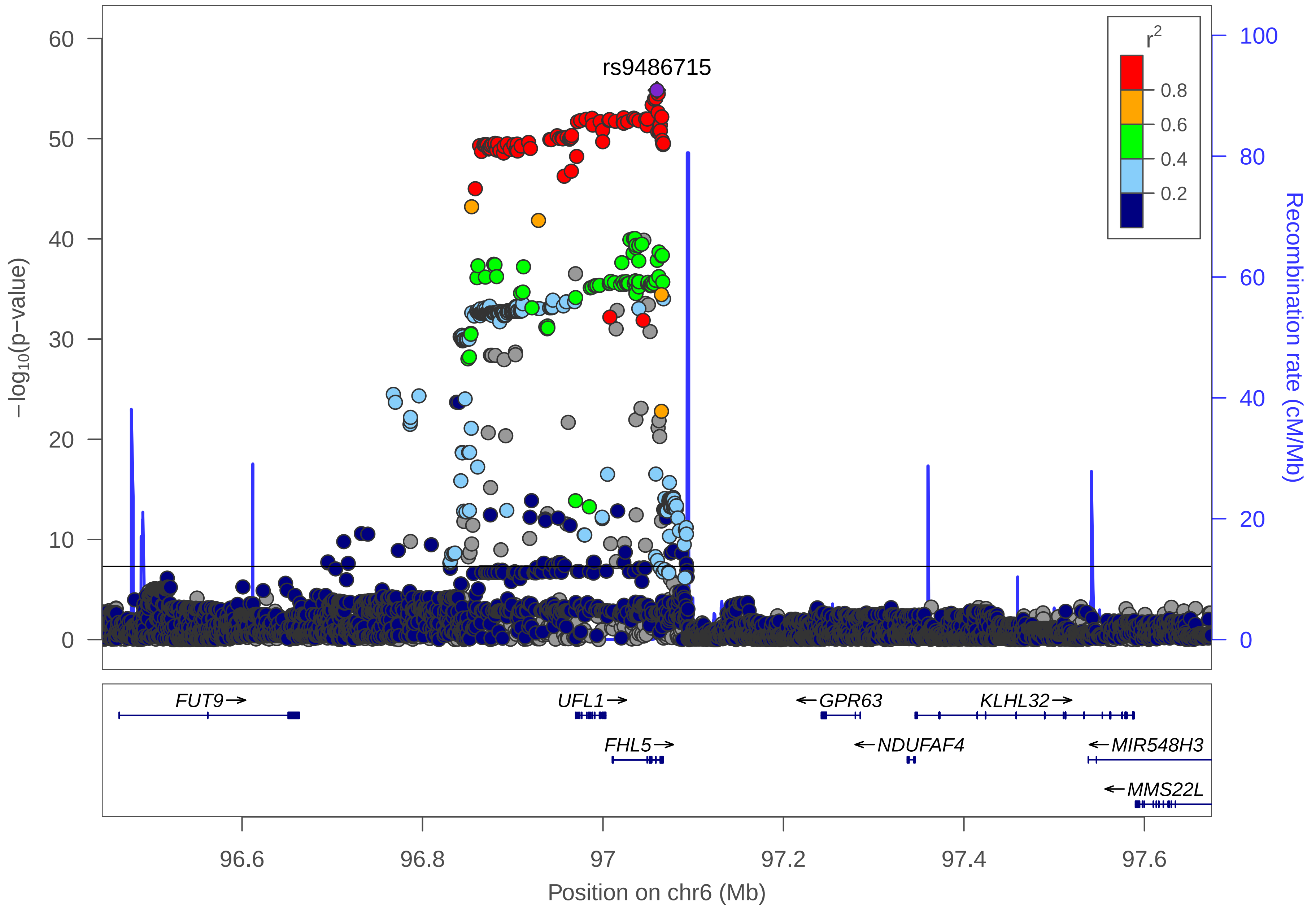

rs9486715
r2
0.8
0.6
0.4
0.2
−log10(p−value)
60
50
40
30
20
10
0
Recombination rate (cM/Mb)
100
80
60
40
20
0
FUT9→
UFL1→
←GPR63
KLHL32→
FHL5→
←NDUFAF4
←MIR548H3
←MMS22L
96.6
96.8
97
97.2
97.4
97.6
Position on chr6 (Mb)

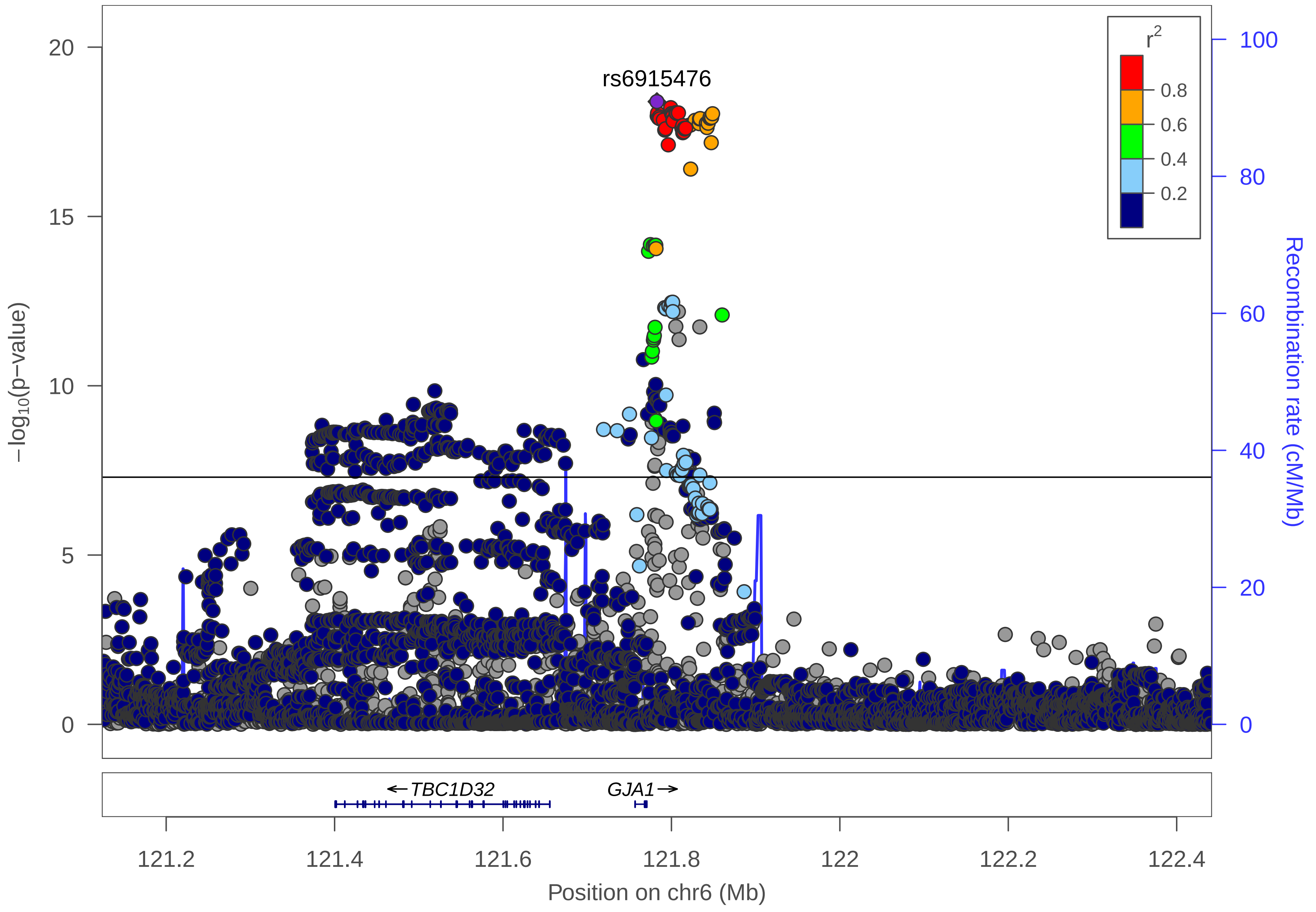

rs6915476
r2
0.8
0.6
0.4
0.2
20
15
10
5
0
−log10(p−value)
100
80
60
40
20
0
Recombination rate (cM/Mb)
←TBC1D32
GJA1→
121.2
121.4
121.6
121.8
122
122.2
122.4
Position on chr6 (Mb)

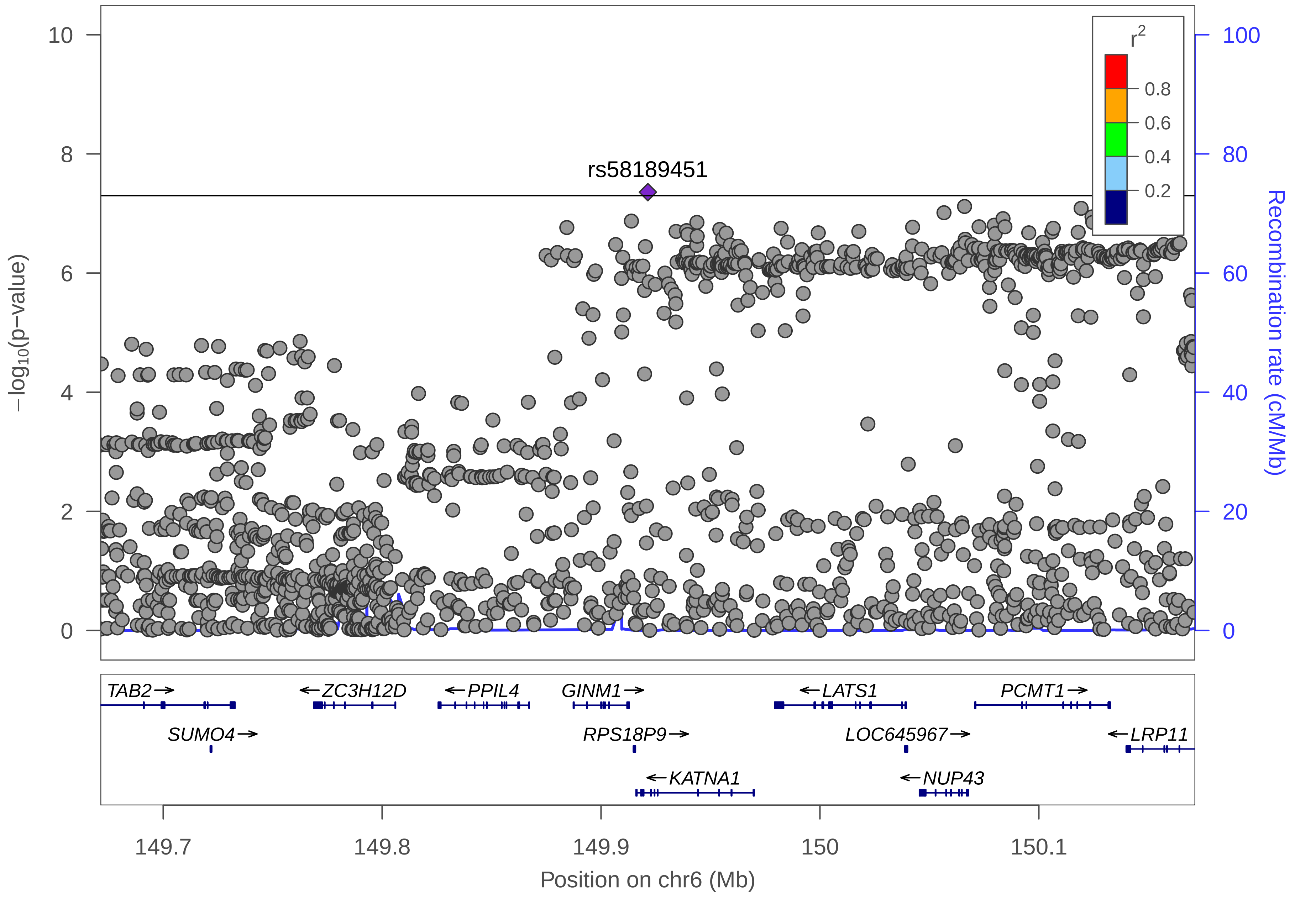

rs58189451
r2
0.8
0.6
0.4
0.2
−log10(p−value)
Recombination rate (cM/Mb)
10
8
6
4
2
0
100
80
60
40
20
0
TAB2
ZC3H12D
PPIL4
GINM1
LATS1
PCMT1
SUMO4
RPS18P9
LOC645967
LRP11
KATNA1
NUP43
149.7
149.8
149.9
150
150.1
Position on chr6 (Mb)

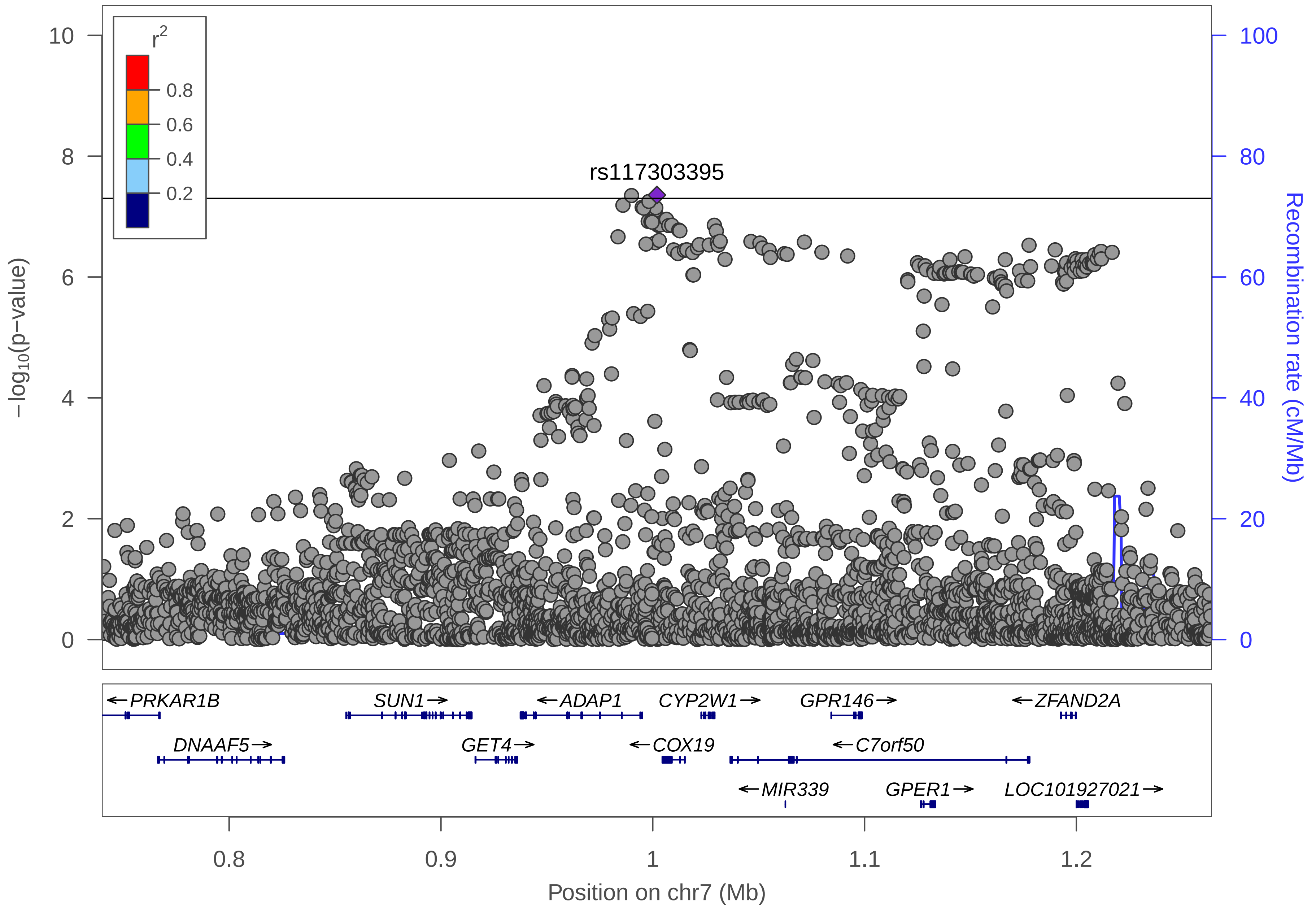

r2
0.8
0.6
0.4
0.2
10
8
6
4
2
0
−log10(p−value)
100
80
60
40
20
0
Recombination rate (cM/Mb)
rs117303395
PRKAR1B
SUN1
ADAP1
CYP2W1
GPR146
ZFAND2A
DNAAF5
GET4
COX19
C7orf50
MIR339
GPER1
LOC101927021
0.8
0.9
1
1.1
1.2
Position on chr7 (Mb)

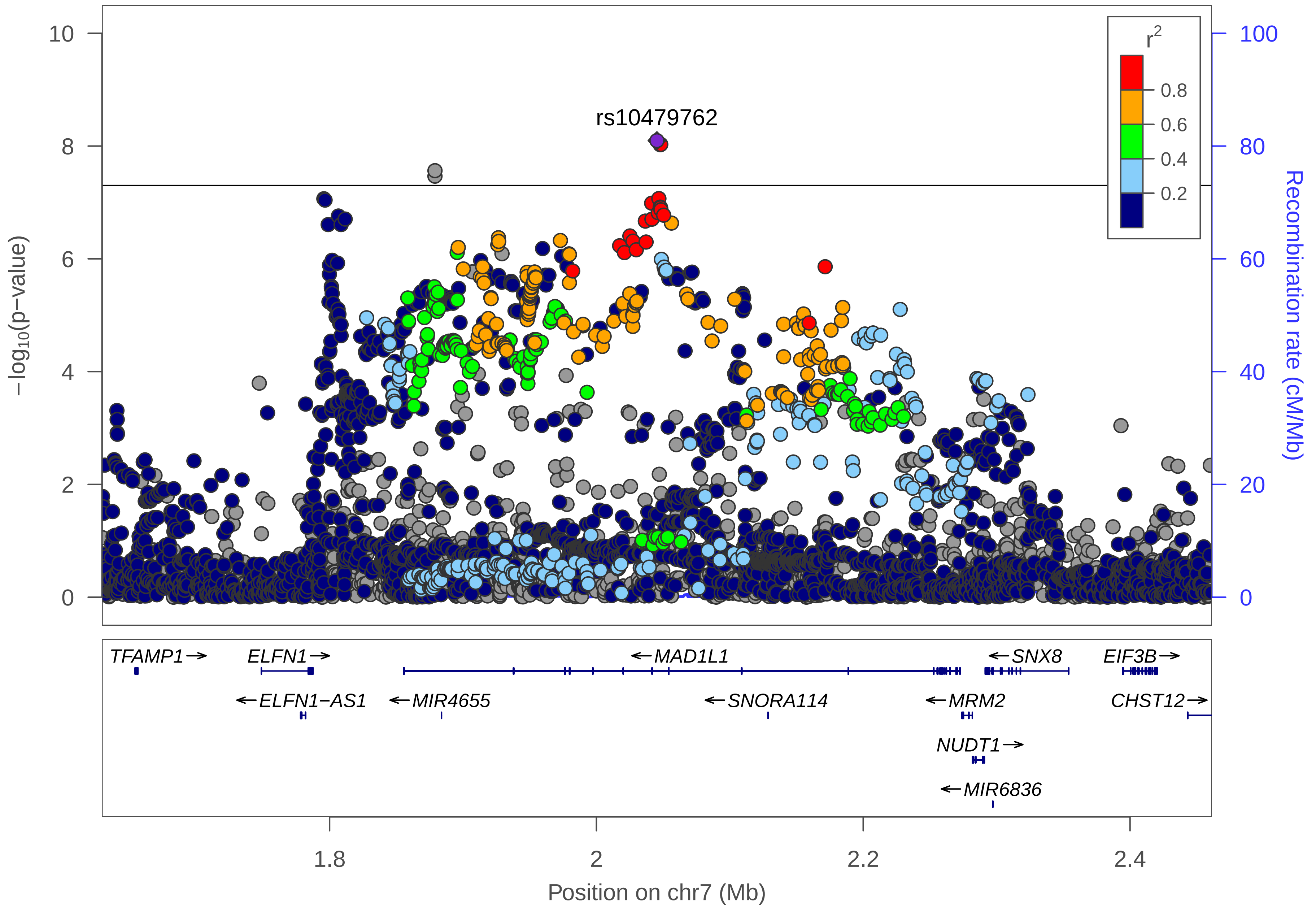

rs10479762
r2
0.8
0.6
0.4
0.2
10
8
6
4
2
0
100
80
60
40
20
0
−log10(p−value)
Recombination rate (cM/Mb)
TFAMP1→
ELFN1→
←MAD1L1
←SNX8
EIF3B→
←ELFN1−AS1
←MIR4655
←SNORA114
←MRM2
CHST12→
NUDT1→
←MIR6836
1.8
2
2.2
2.4
Position on chr7 (Mb)

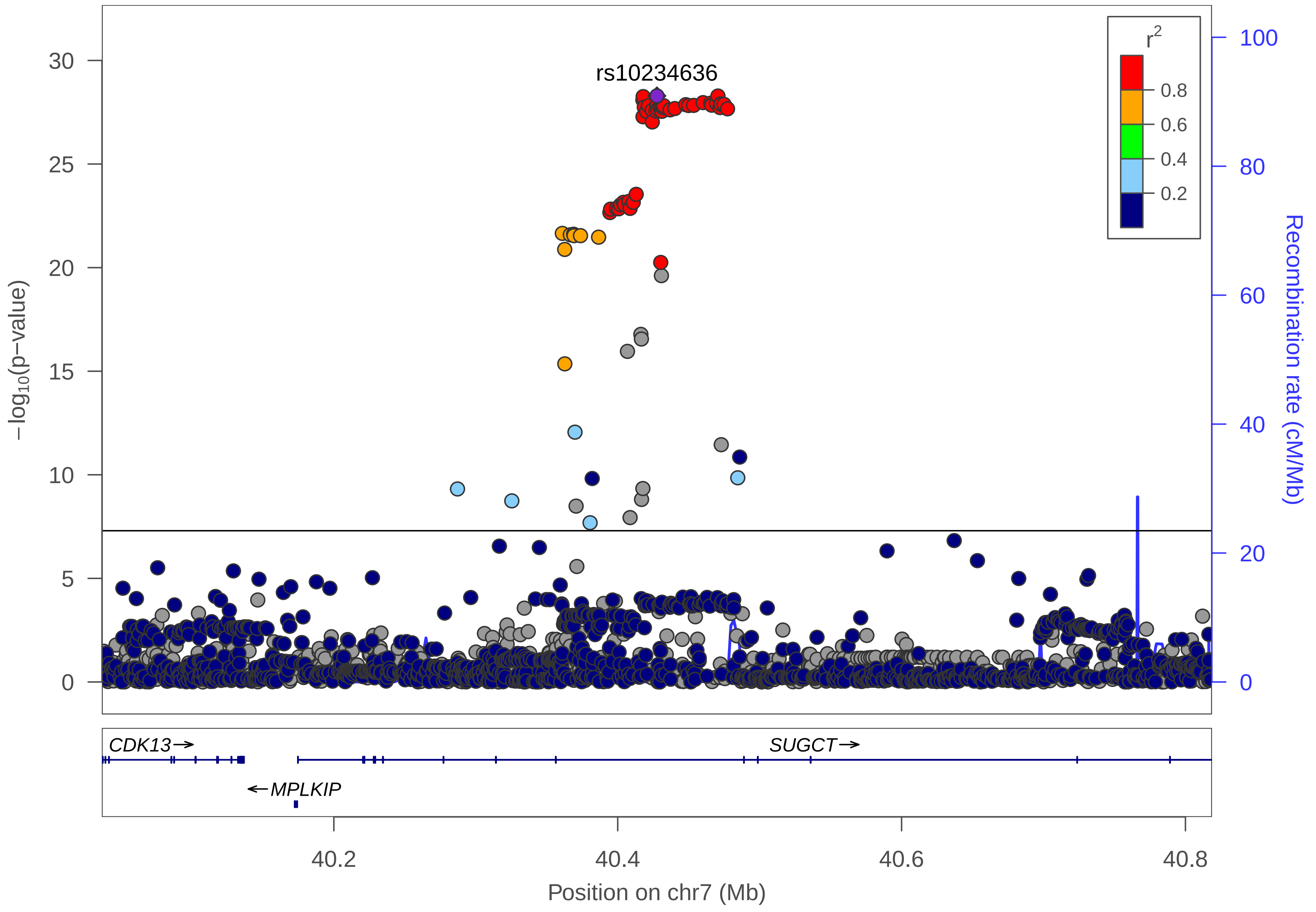

rs10234636
r2
0.8
0.6
0.4
0.2
−log10(p−value)
Recombination rate (cM/Mb)
CDK13→
SUGCT→
←MPLKIP
40.2
40.4
40.6
40.8
Position on chr7 (Mb)

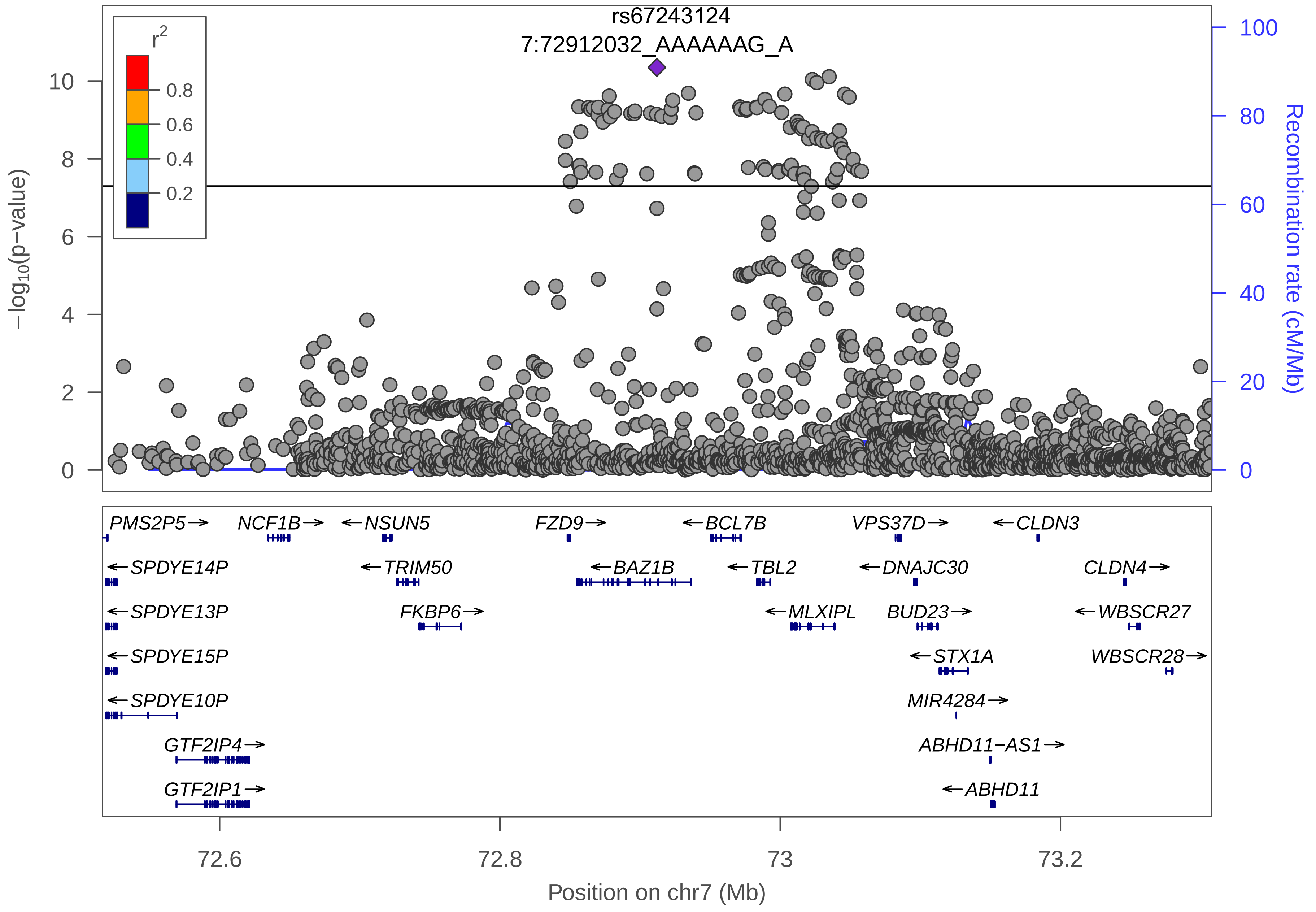

rs67243124
7:72912032_AAAAAAG_A
r2
0.8
0.6
0.4
0.2
−log10(p−value)
Recombination rate (cM/Mb)
PMS2P5
NCF1B
NSUN5
FZD9
BCL7B
VPS37D
CLDN3
SPDYE14P
TRIM50
BAZ1B
TBL2
DNAJC30
CLDN4
SPDYE13P
FKBP6
MLXIPL
BUD23
WBSCR27
SPDYE15P
STX1A
WBSCR28
SPDYE10P
MIR4284
GTF2IP4
ABHD11−AS1
GTF2IP1
ABHD11
72.6
72.8
73
73.2
Position on chr7 (Mb)

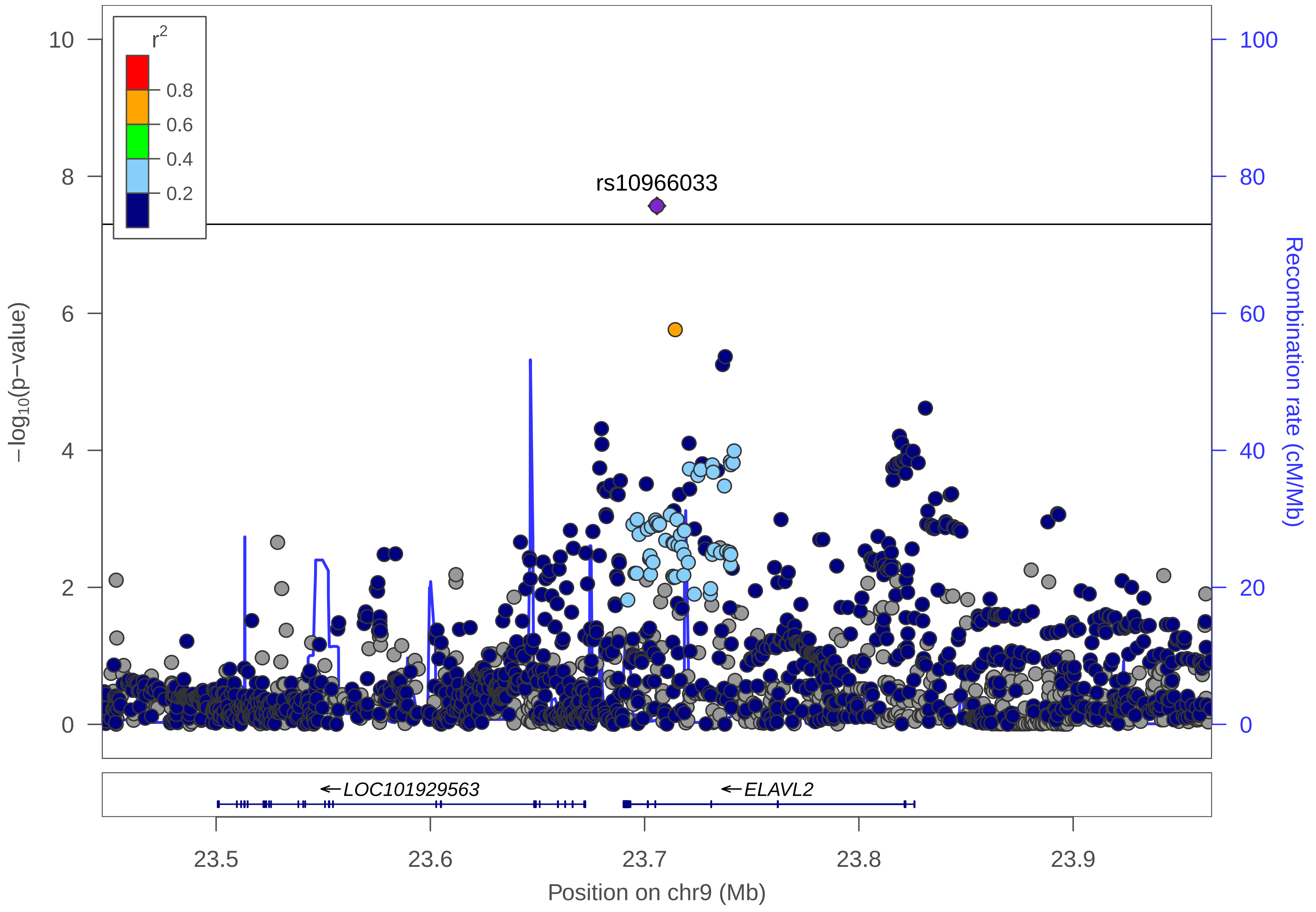

r2
0.8
0.6
0.4
0.2
rs10966033
−log10(p−value)
Recombination rate (cM/Mb)
10
8
6
4
2
0
100
80
60
40
20
0
←LOC101929563
←ELAVL2
23.5
23.6
23.7
23.8
23.9
Position on chr9 (Mb)

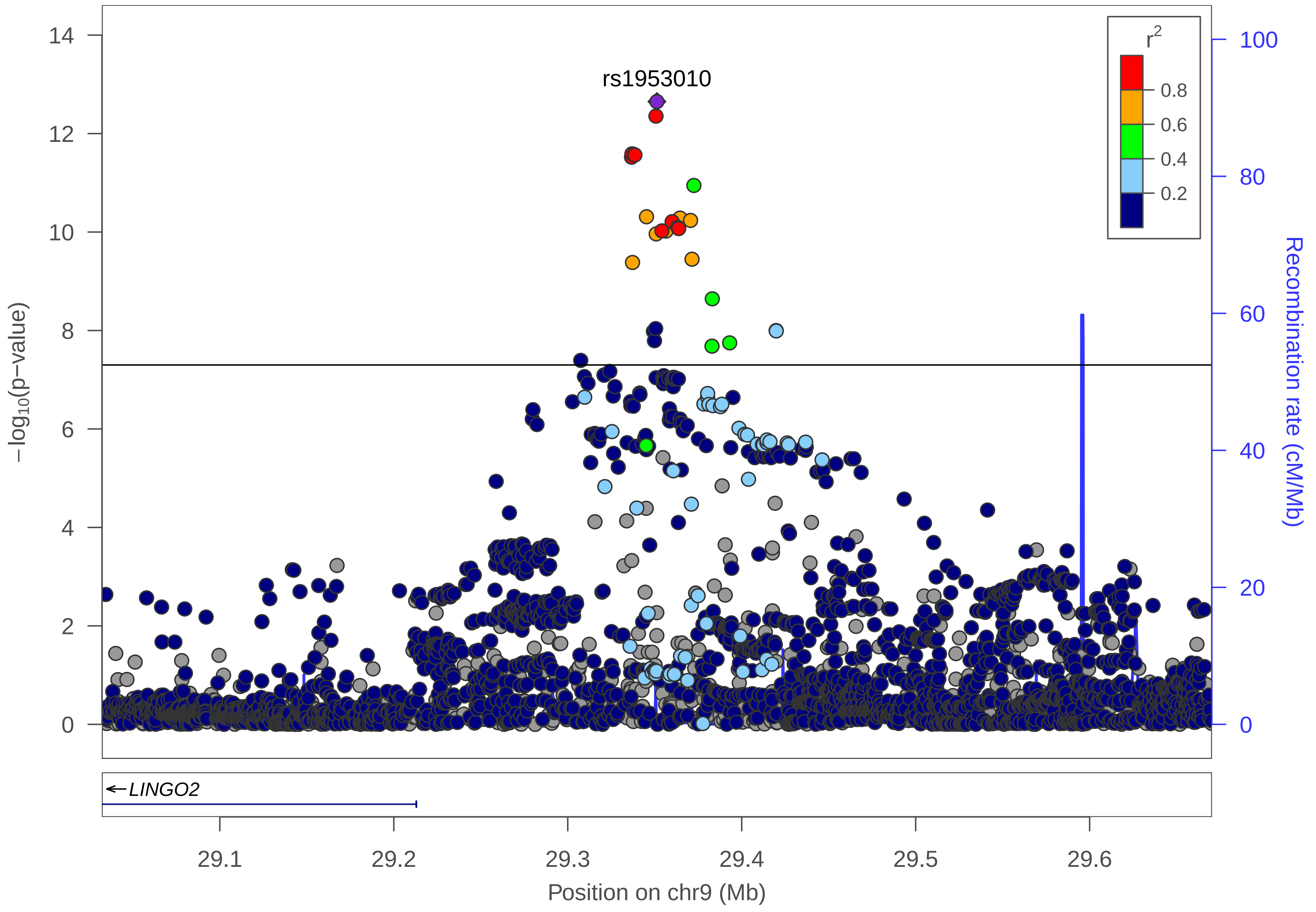

rs1953010
r²
0.8
0.6
0.4
0.2
−log10(p−value)
14
12
10
8
6
4
2
0
100
80
60
40
20
0
Recombination rate (cM/Mb)
LINGO2
29.1
29.2
29.3
29.4
29.5
29.6
Position on chr9 (Mb)

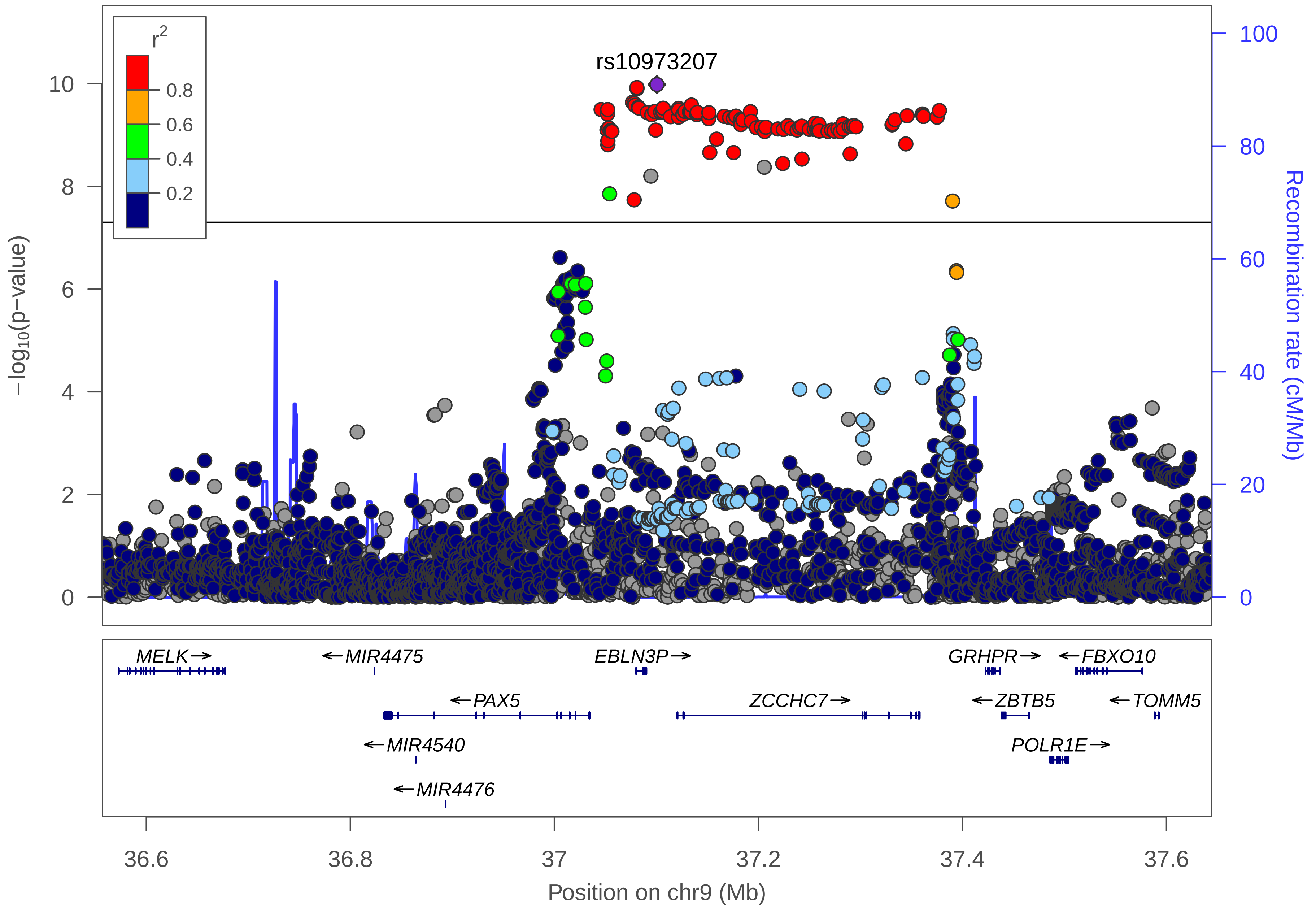

rs10973207
r2
0.8
0.6
0.4
0.2
−log10(p−value)
10
8
6
4
2
0
Recombination rate (cM/Mb)
100
80
60
40
20
0
MELK→
←MIR4475
EBLN3P→
GRHPR→
←FBXO10
←PAX5
ZCCHC7→
←ZBTB5
←TOMM5
←MIR4540
POLR1E→
←MIR4476
36.6
36.8
37
37.2
37.4
37.6
Position on chr9 (Mb)

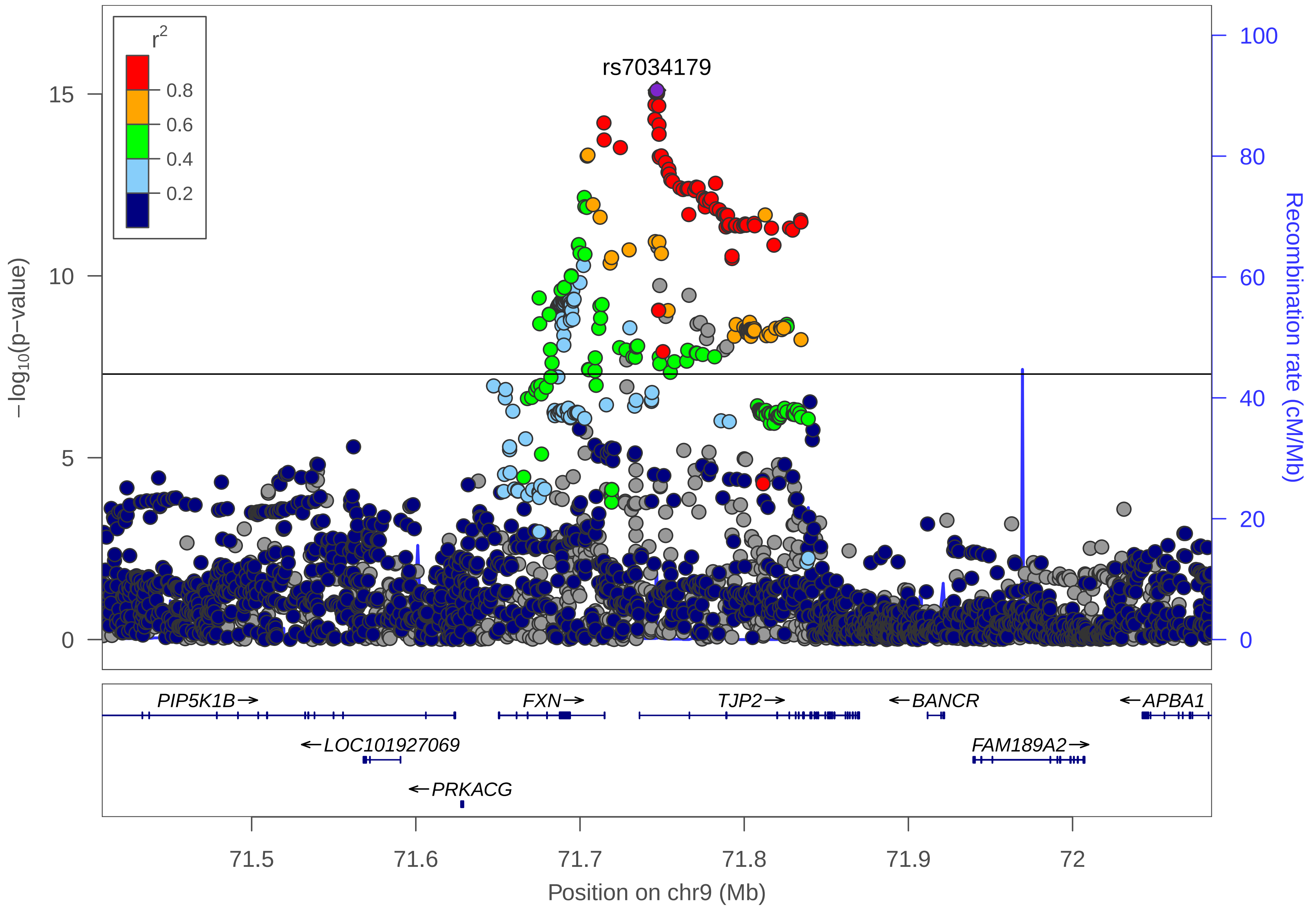
r2
0.8
0.6
0.4
0.2
rs7034179
15
10
5
0
−log10(p−value)
100
80
60
40
20
0
Recombination rate (cM/Mb)
PIP5K1B→
FXN→
TJP2→
←BANCR
←APBA1
←LOC101927069
FAM189A2→
←PRKACG
71.5
71.6
71.7
71.8
71.9
72
Position on chr9 (Mb)

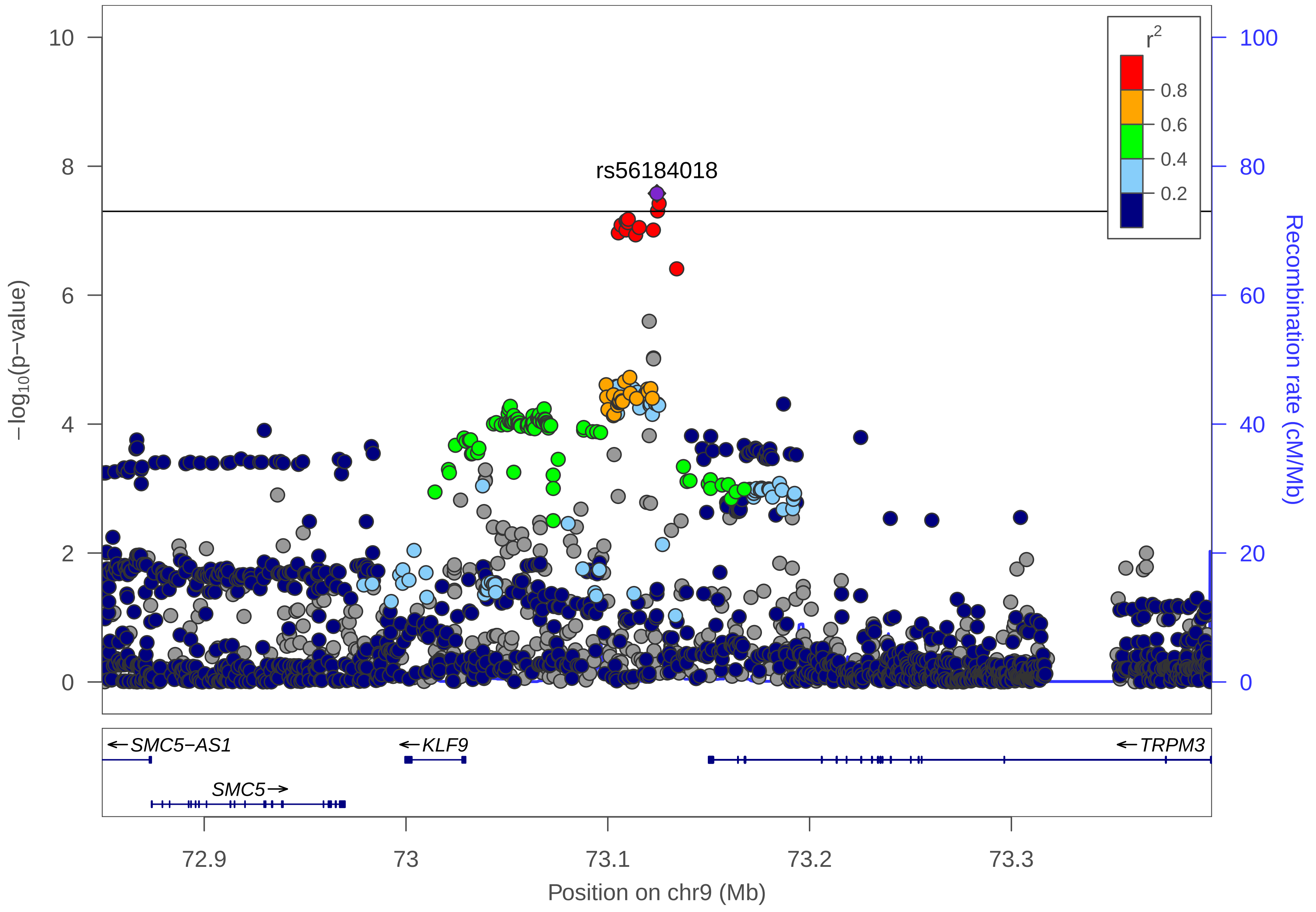

rs56184018
r²
0.8
0.6
0.4
0.2
−log10(p−value)
Recombination rate (cM/Mb)
0
2
4
6
8
10
0
20
40
60
80
100
←SMC5−AS1
←KLF9
←TRPM3
SMC5→
72.9
73
73.1
73.2
73.3
Position on chr9 (Mb)

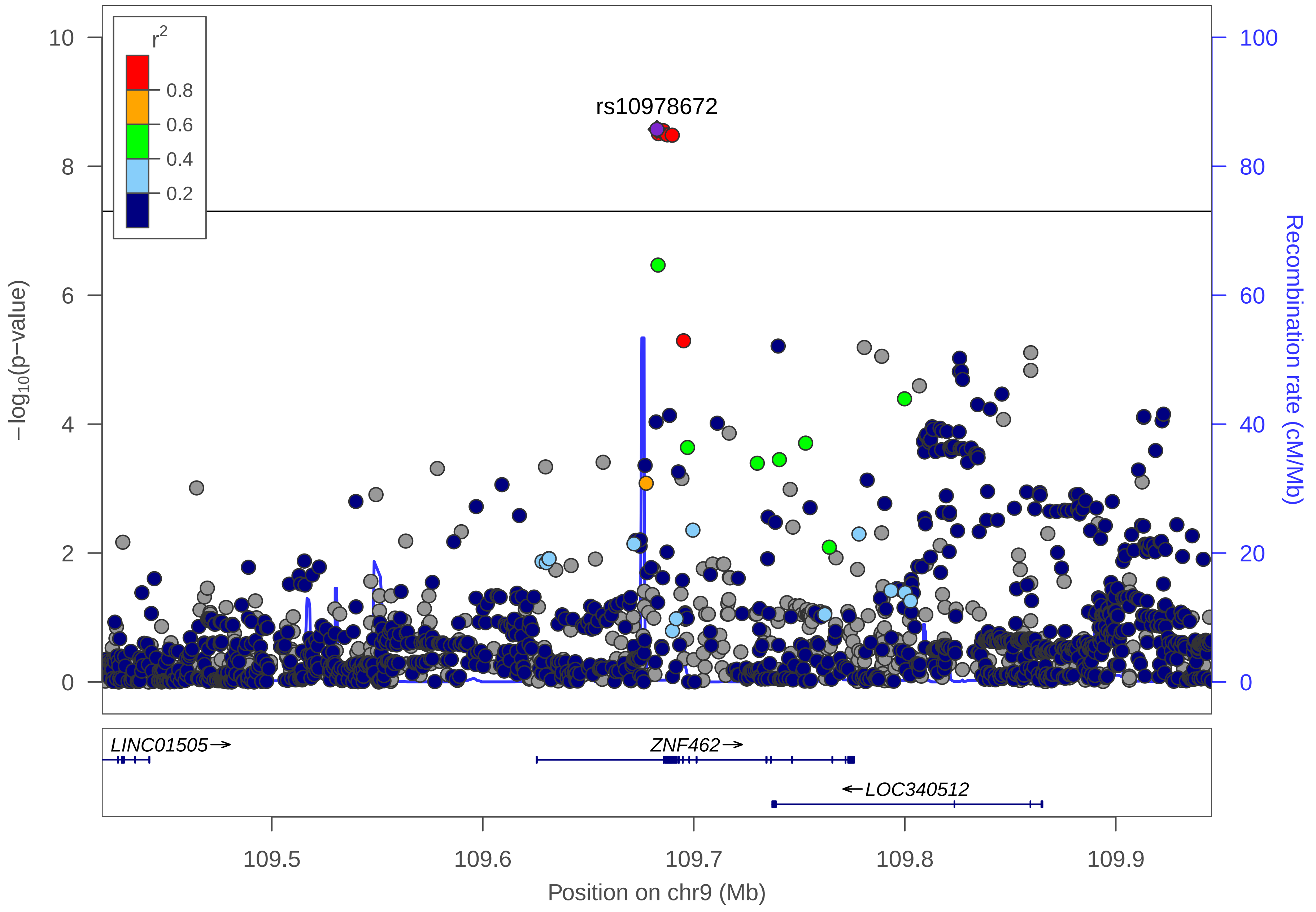
r2
0.8
0.6
0.4
0.2
rs10978672
−log10(p−value)
Recombination rate (cM/Mb)
10
8
6
4
2
0
100
80
60
40
20
0
LINC01505→
ZNF462→
←LOC340512
109.5
109.6
109.7
109.8
109.9
Position on chr9 (Mb)

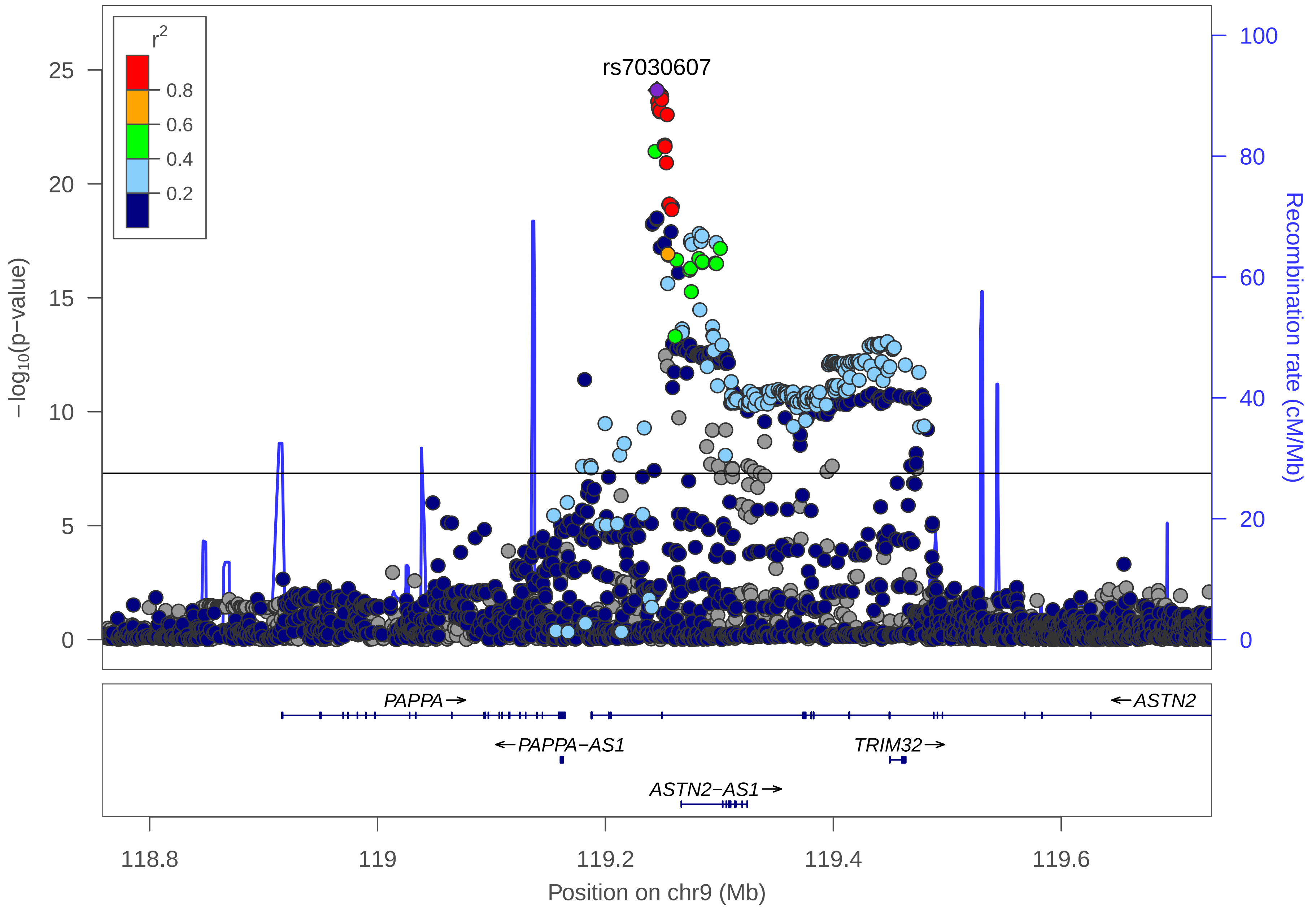

r2
0.8
0.6
0.4
0.2
rs7030607
25
20
15
10
5
0
−log10(p−value)
100
80
60
40
20
0
Recombination rate (cM/Mb)
PAPPA→
←ASTN2
←PAPPA−AS1
TRIM32→
ASTN2−AS1→
118.8
119
119.2
119.4
119.6
Position on chr9 (Mb)

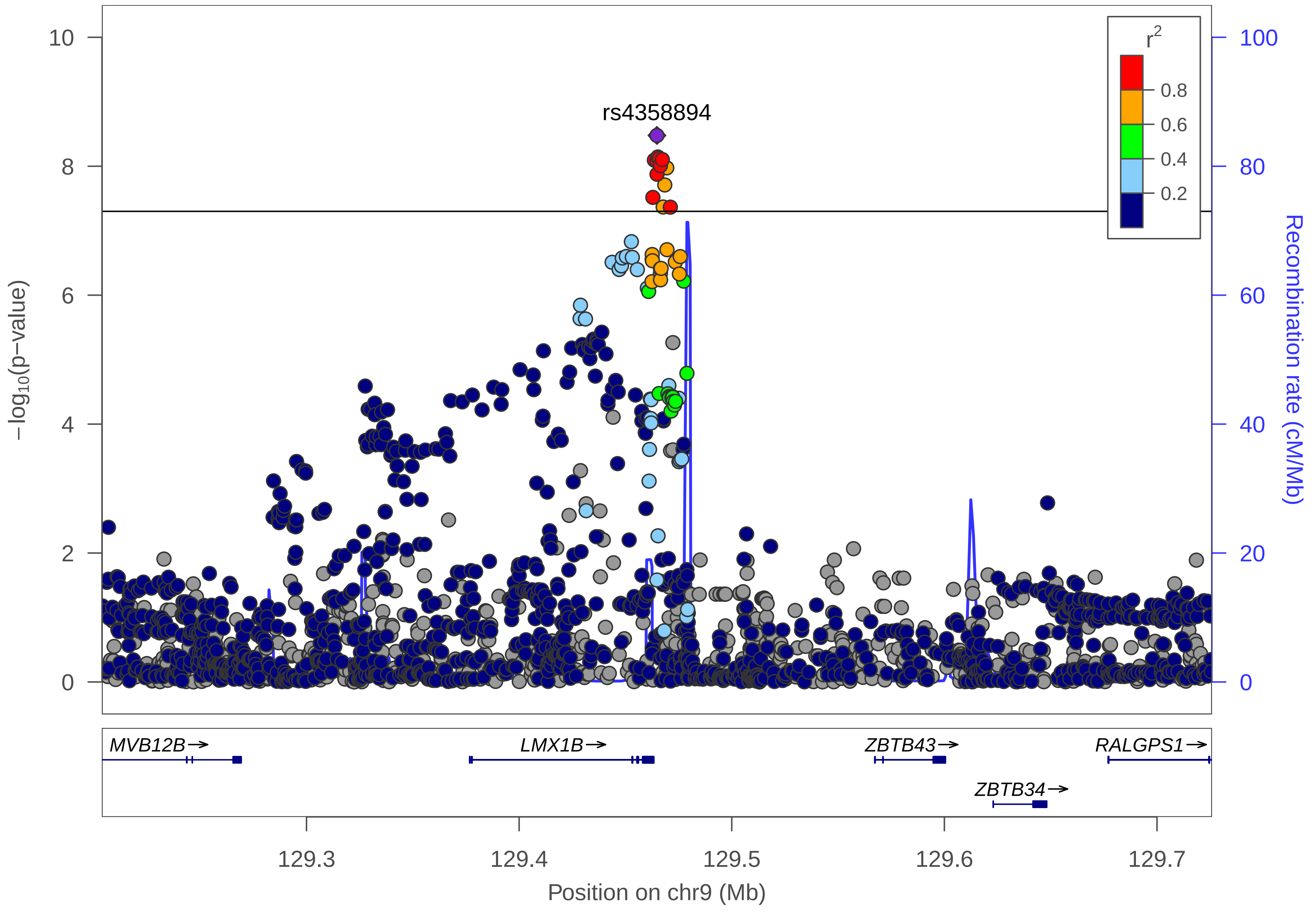

rs4358894
r2
0.8
0.6
0.4
0.2
10
8
6
4
2
0
100
80
60
40
20
0
−log10(p−value)
Recombination rate (cM/Mb)
MVB12B→
LMX1B→
ZBTB43→
ZBTB34→
RALGPS1→
129.3
129.4
129.5
129.6
129.7
Position on chr9 (Mb)

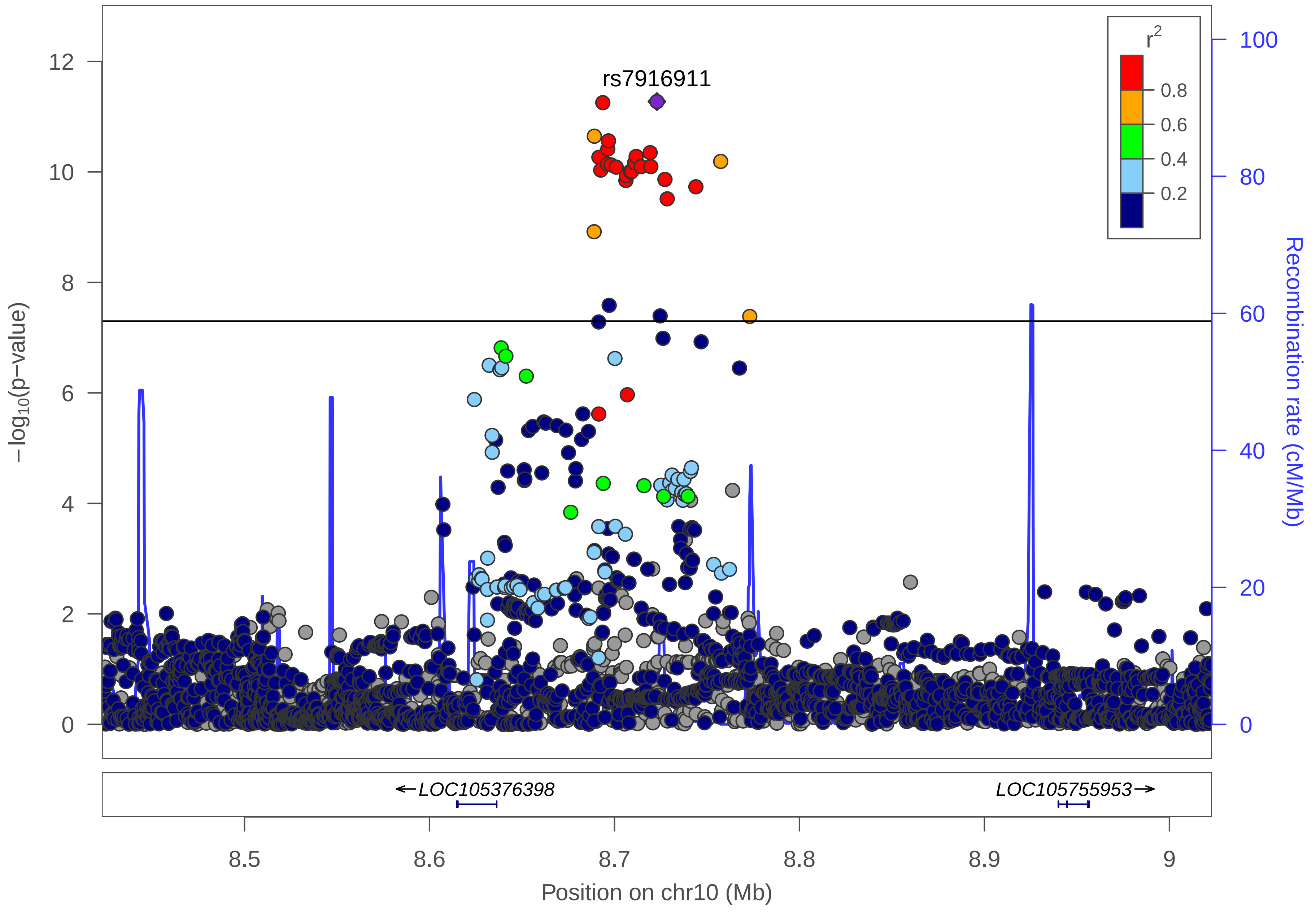

rs7916911
r2
0.8
0.6
0.4
0.2
12
10
8
6
4
2
0
−log10(p−value)
100
80
60
40
20
0
Recombination rate (cM/Mb)
←LOC105376398
LOC105755953→
8.5
8.6
8.7
8.8
8.9
9
Position on chr10 (Mb)

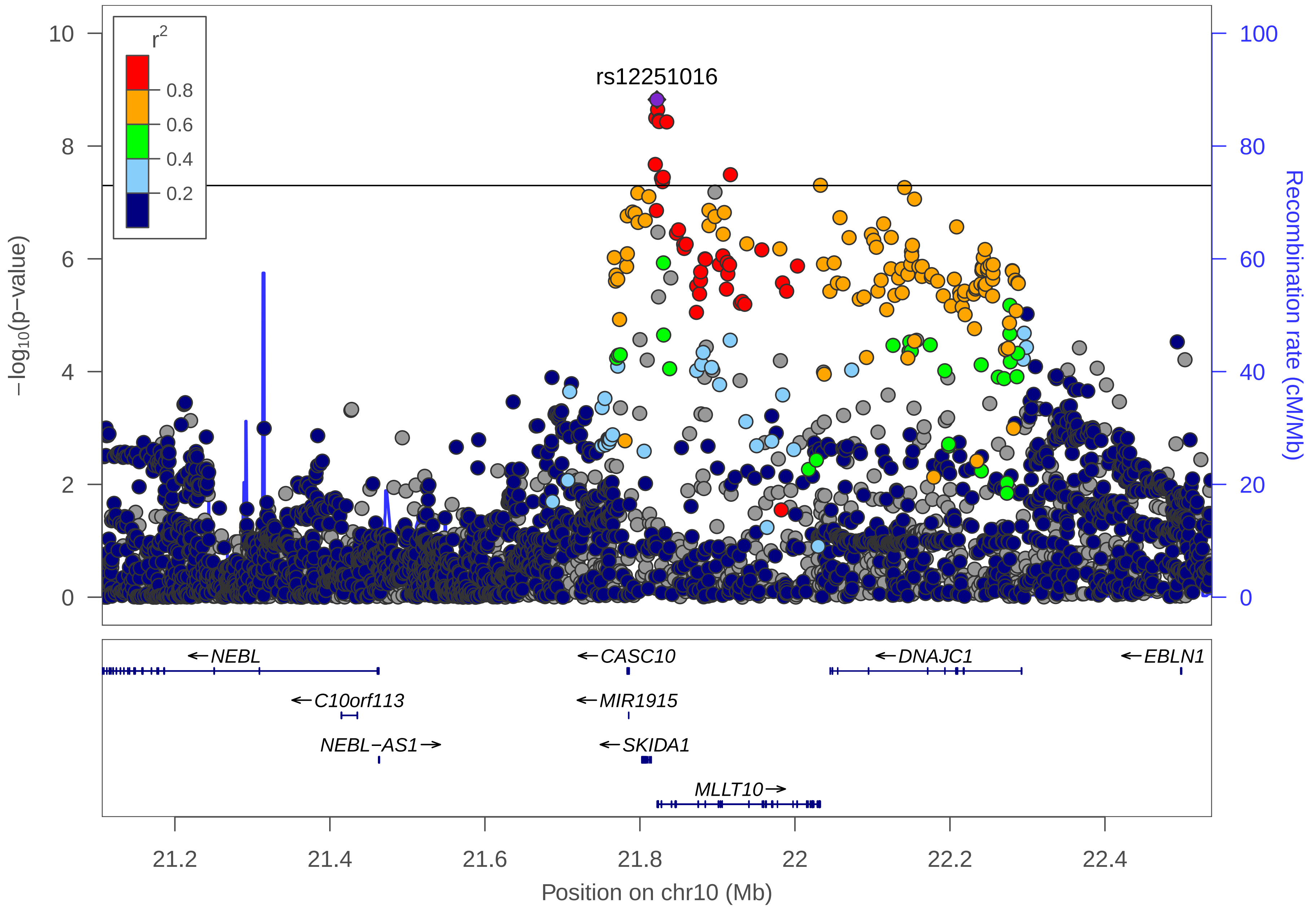

rs12251016
r2
0.8
0.6
0.4
0.2
−log10(p−value)
Recombination rate (cM/Mb)
0
2
4
6
8
10
0
20
40
60
80
100
←NEBL
←C10orf113
NEBL−AS1→
←CASC10
←MIR1915
←SKIDA1
MLLT10→
←DNAJC1
←EBLN1
21.2
21.4
21.6
21.8
22
22.2
22.4
Position on chr10 (Mb)

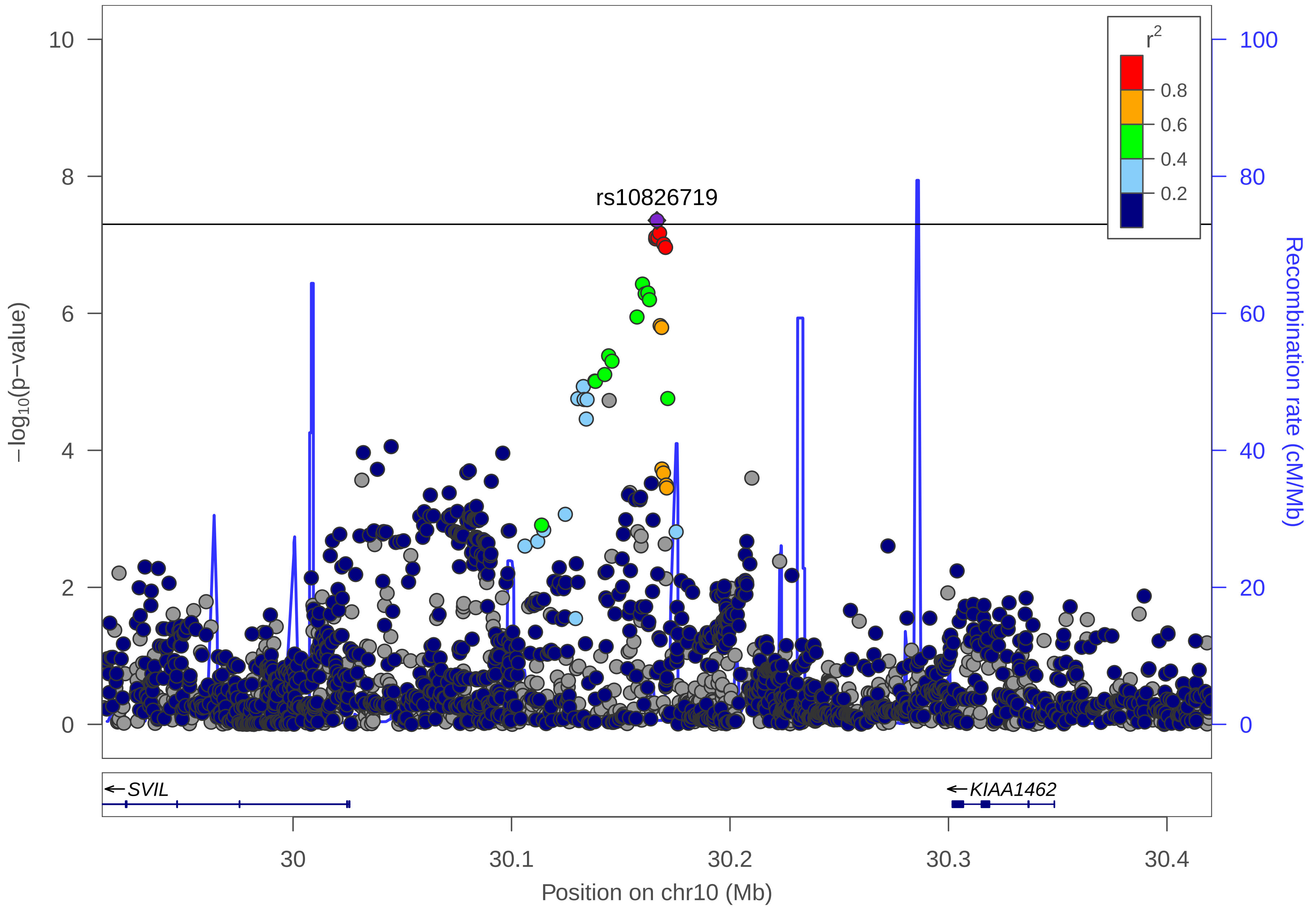

rs10826719
r²
0.8
0.6
0.4
0.2
10
8
6
4
2
0
−log10(p−value)
100
80
60
40
20
0
Recombination rate (cM/Mb)
←SVIL
←KIAA1462
30
30.1
30.2
30.3
30.4
Position on chr10 (Mb)

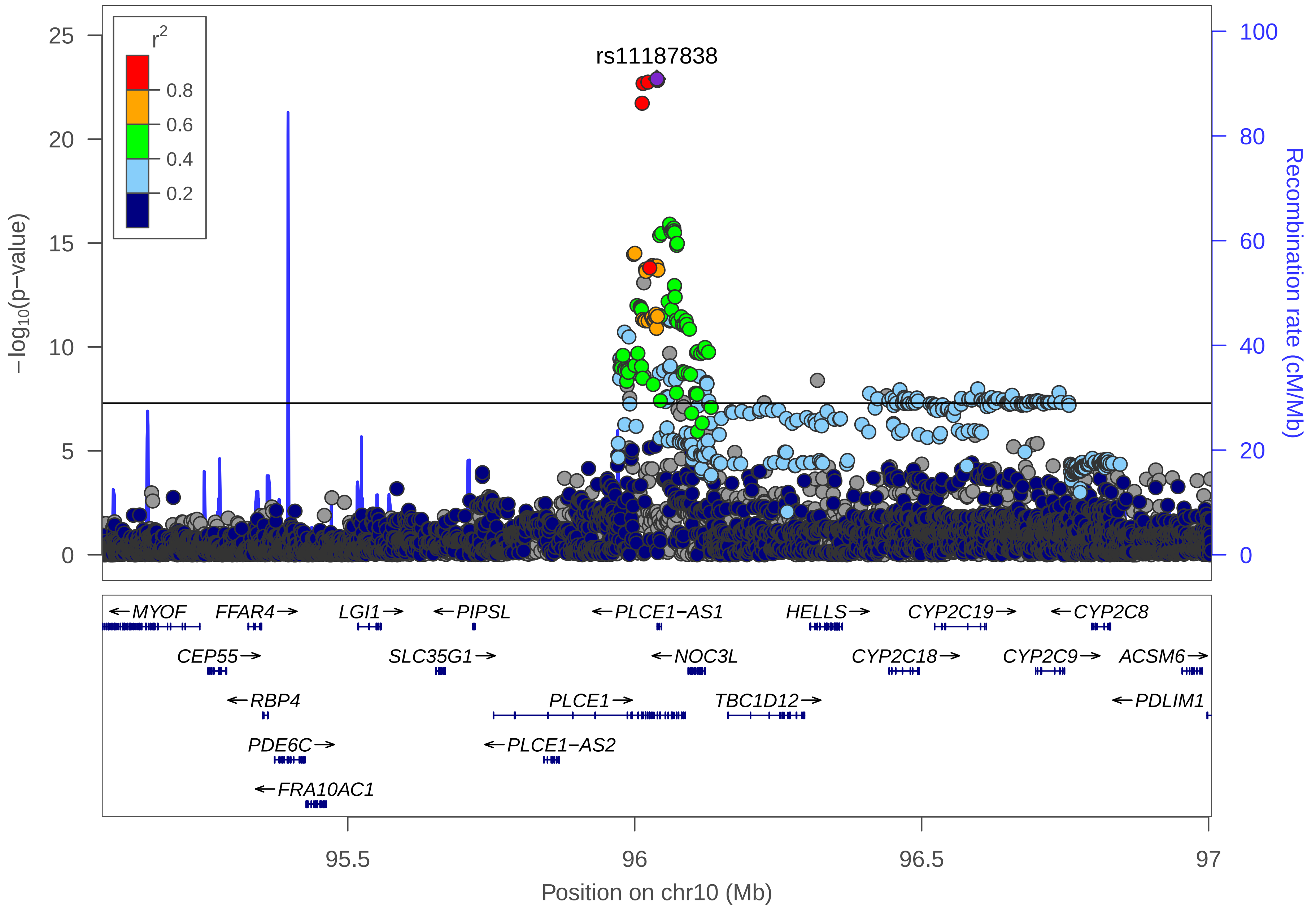

r2
0.8
0.6
0.4
0.2
rs11187838
−log10(p−value)
Recombination rate (cM/Mb)
0
5
10
15
20
25
0
20
40
60
80
100
←MYOF
FFAR4→
LGI1→
←PIPSL
←PLCE1−AS1
HELLS→
CYP2C19→
←CYP2C8
CEP55→
SLC35G1→
←NOC3L
CYP2C18→
CYP2C9→
ACSM6→
←RBP4
PLCE1→
TBC1D12→
←PDLIM1
PDE6C→
←PLCE1−AS2
←FRA10AC1
95.5
96
96.5
97
Position on chr10 (Mb)

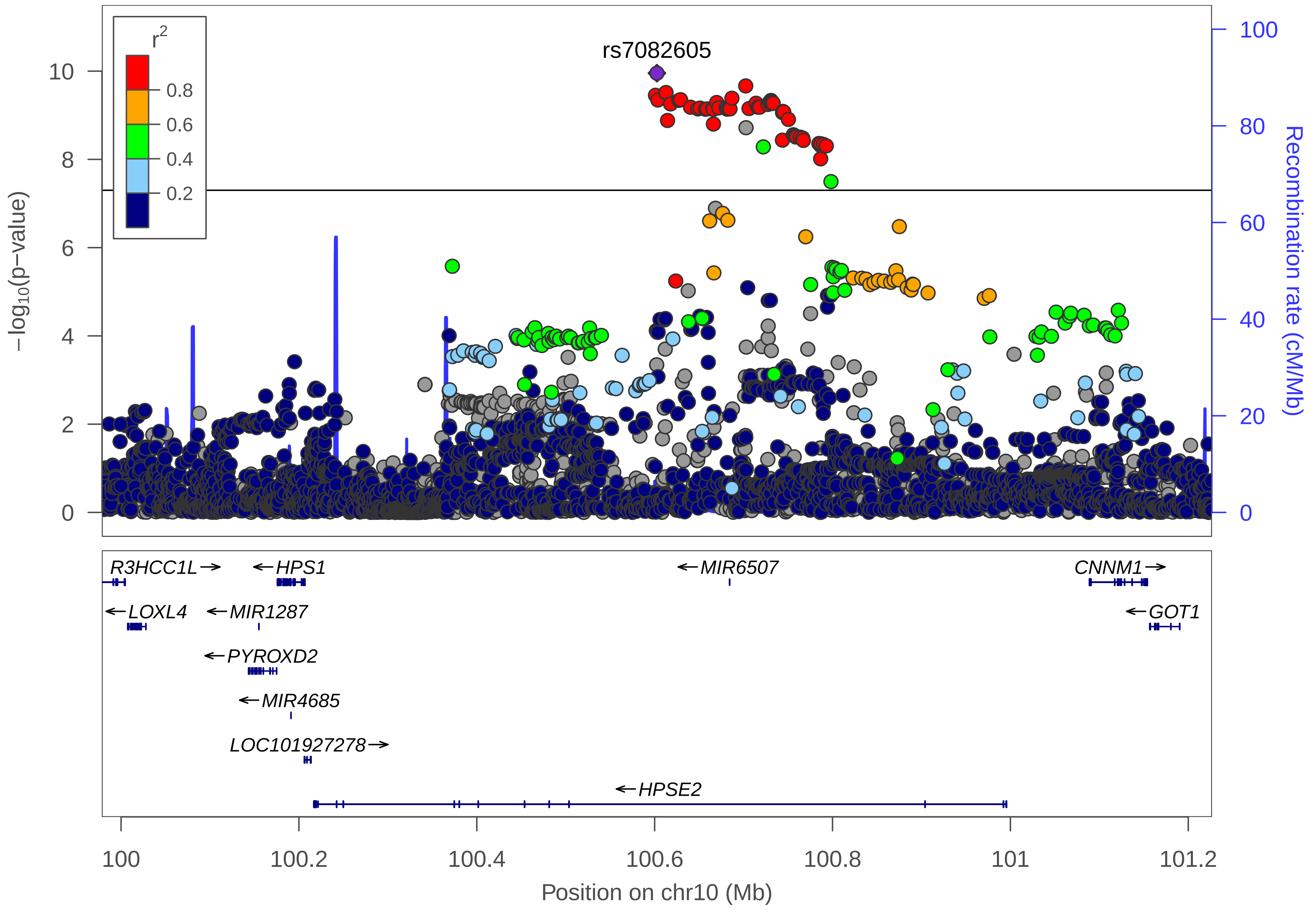

rs7082605
r2
0.8
0.6
0.4
0.2
10
8
6
4
2
0
−log10(p−value)
100
80
60
40
20
0
Recombination rate (cM/Mb)
R3HCC1L→
←HPS1
←MIR6507
CNNM1→
←LOXL4
←MIR1287
←GOT1
←PYROXD2
←MIR4685
LOC101927278→
←HPSE2
100
100.2
100.4
100.6
100.8
101
101.2
Position on chr10 (Mb)

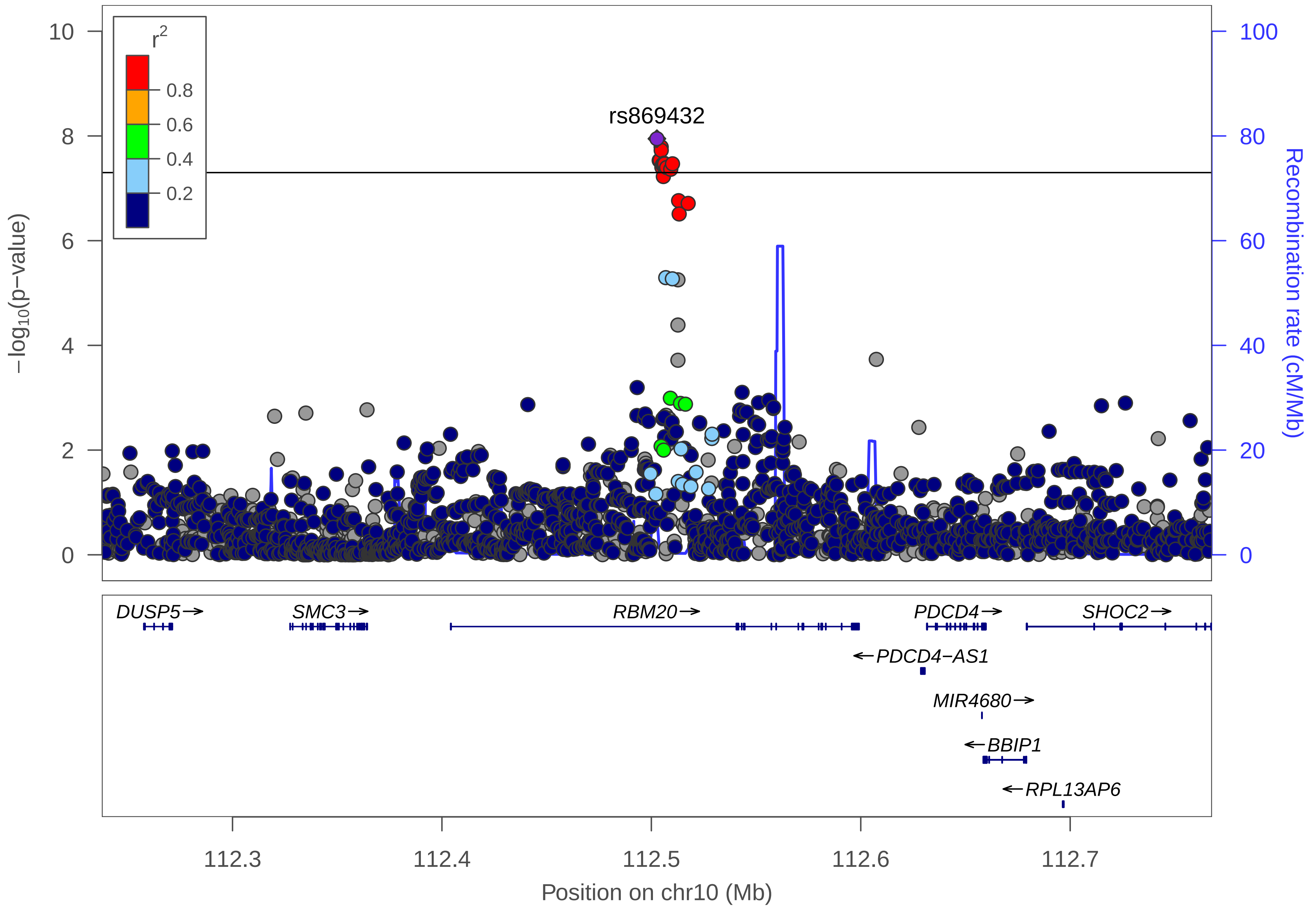

r2
0.8
0.6
0.4
0.2
rs869432
−log10(p−value)
Recombination rate (cM/Mb)
DUSP5→
SMC3→
RBM20→
PDCD4→
SHOC2→
←PDCD4−AS1
MIR4680→
←BBIP1
←RPL13AP6
112.3
112.4
112.5
112.6
112.7
Position on chr10 (Mb)

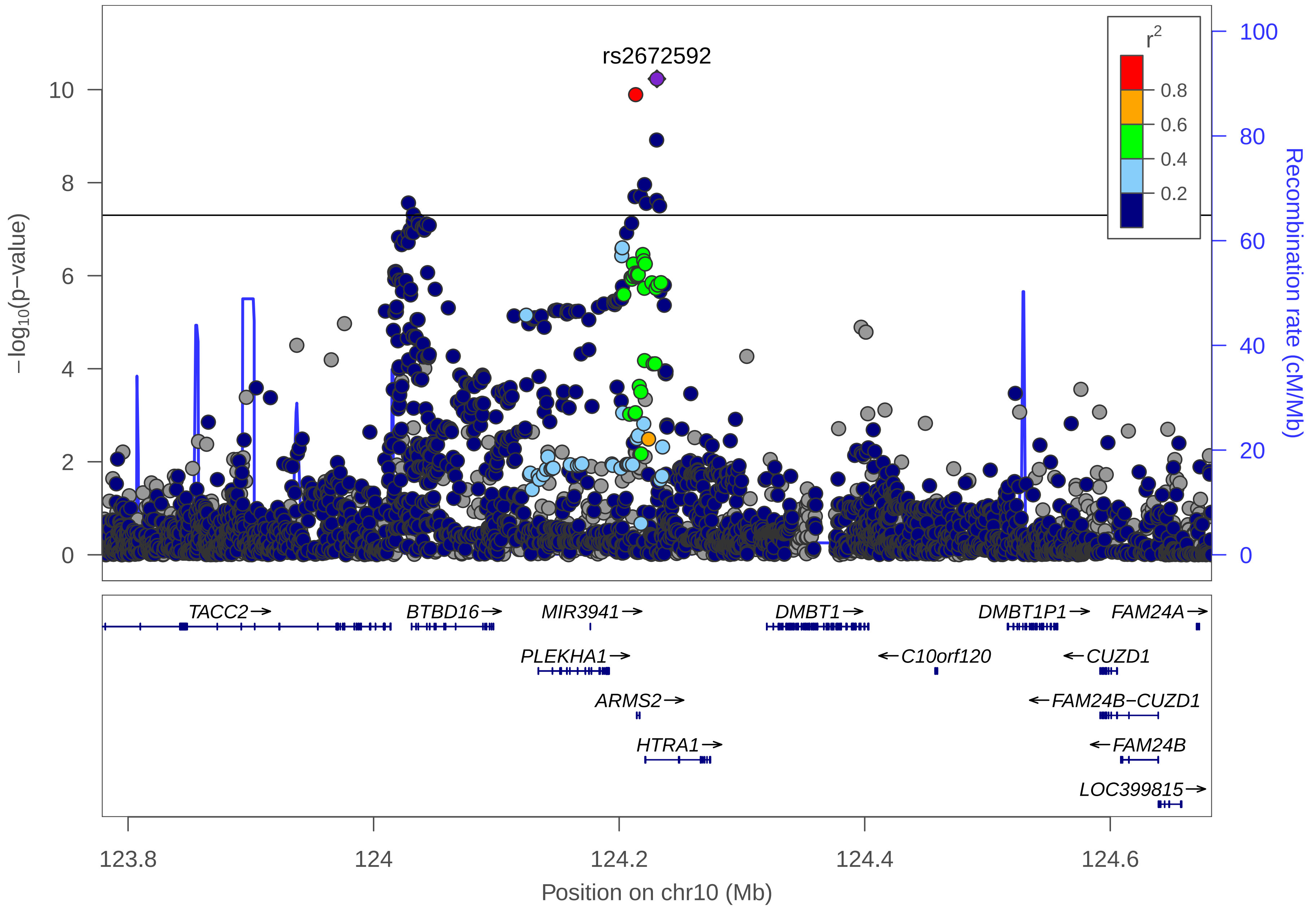

rs2672592
r2
0.8
0.6
0.4
0.2
−log10(p−value)
Recombination rate (cM/Mb)
0
2
4
6
8
10
0
20
40
60
80
100
TACC2→
BTBD16→
MIR3941→
DMBT1→
DMBT1P1→
FAM24A→
PLEKHA1→
←C10orf120
←CUZD1
ARMS2→
←FAM24B−CUZD1
HTRA1→
←FAM24B
LOC399815→
123.8
124
124.2
124.4
124.6
Position on chr10 (Mb)

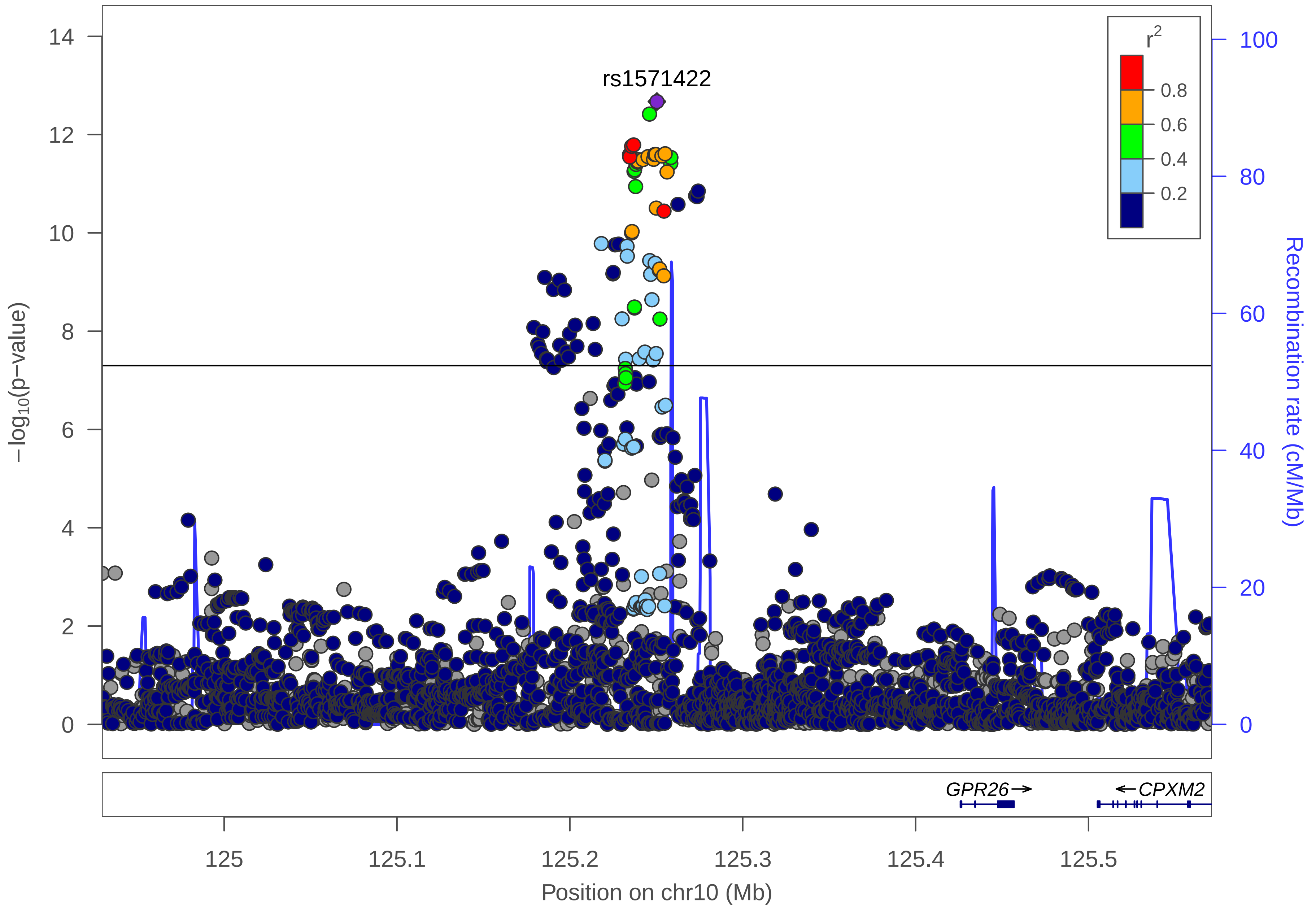

rs1571422
r2
0.8
0.6
0.4
0.2
−log10(p−value)
0
2
4
6
8
10
12
14
Recombination rate (cM/Mb)
0
20
40
60
80
100
GPR26
CPXM2
125
125.1
125.2
125.3
125.4
125.5
Position on chr10 (Mb)

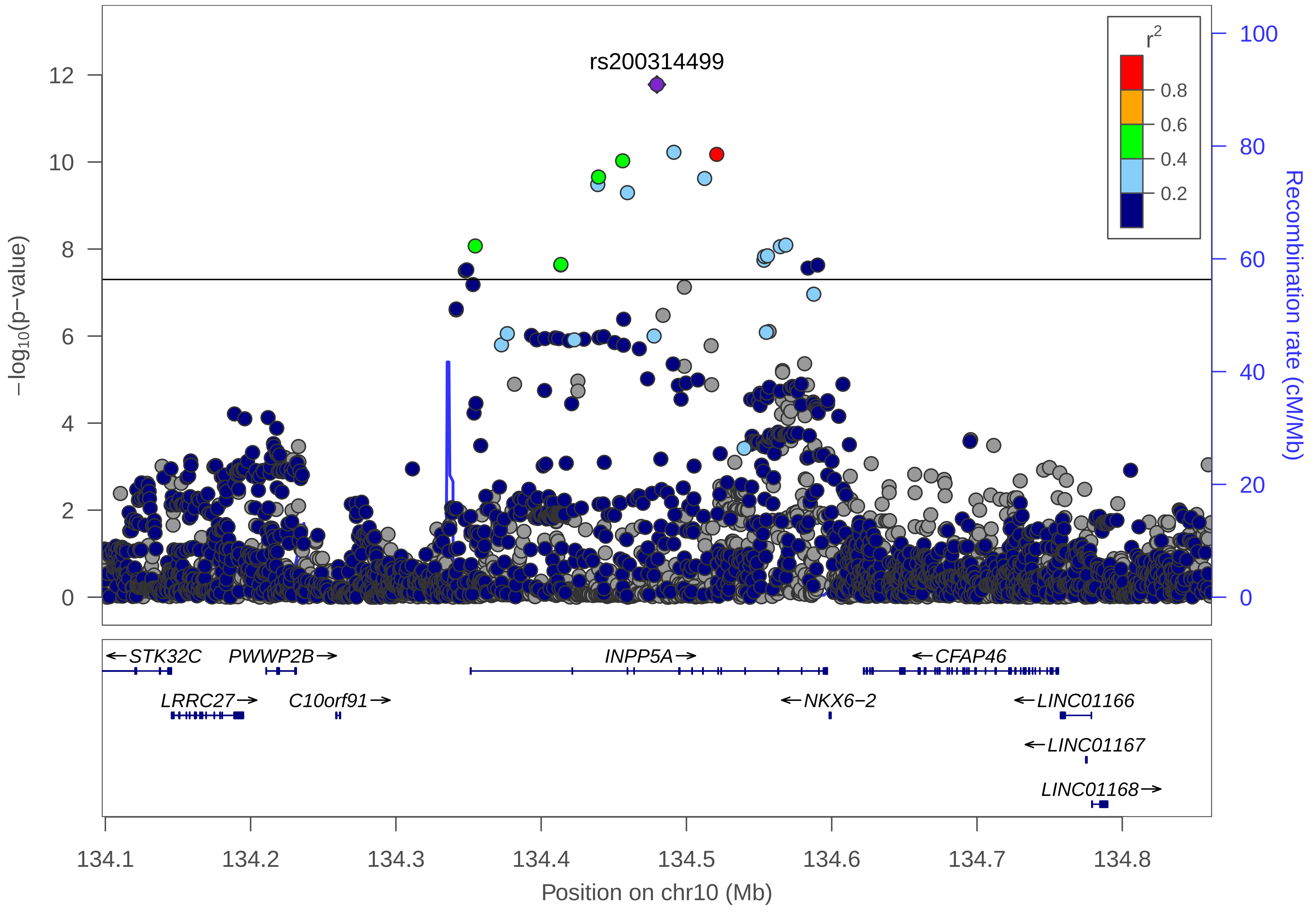

rs200314499
r2
0.8
0.6
0.4
0.2
−log10(p−value)
Recombination rate (cM/Mb)
0
2
4
6
8
10
12
0
20
40
60
80
100
STK32C
PWWP2B
INPP5A
CFAP46
LRRC27
C10orf91
NKX6−2
LINC01166
LINC01167
LINC01168
134.1
134.2
134.3
134.4
134.5
134.6
134.7
134.8
Position on chr10 (Mb)

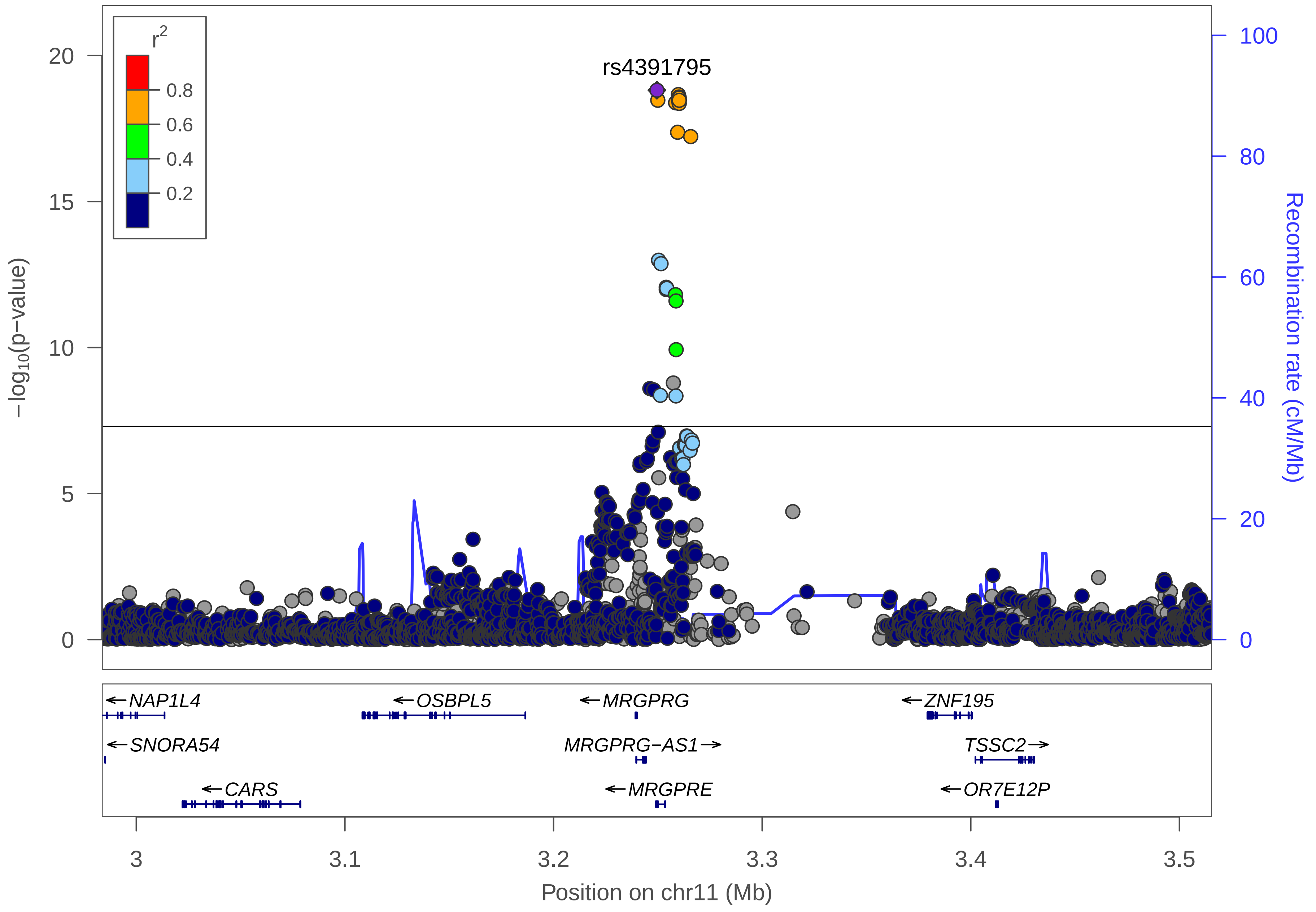

rs4391795
r2
0.8
0.6
0.4
0.2
−log10(p−value)
Recombination rate (cM/Mb)
20
15
10
5
0
100
80
60
40
20
0
NAP1L4
SNORA54
CARS
OSBPL5
MRGPRG
MRGPRG−AS1
MRGPRE
ZNF195
TSSC2
OR7E12P
3
3.1
3.2
3.3
3.4
3.5
Position on chr11 (Mb)

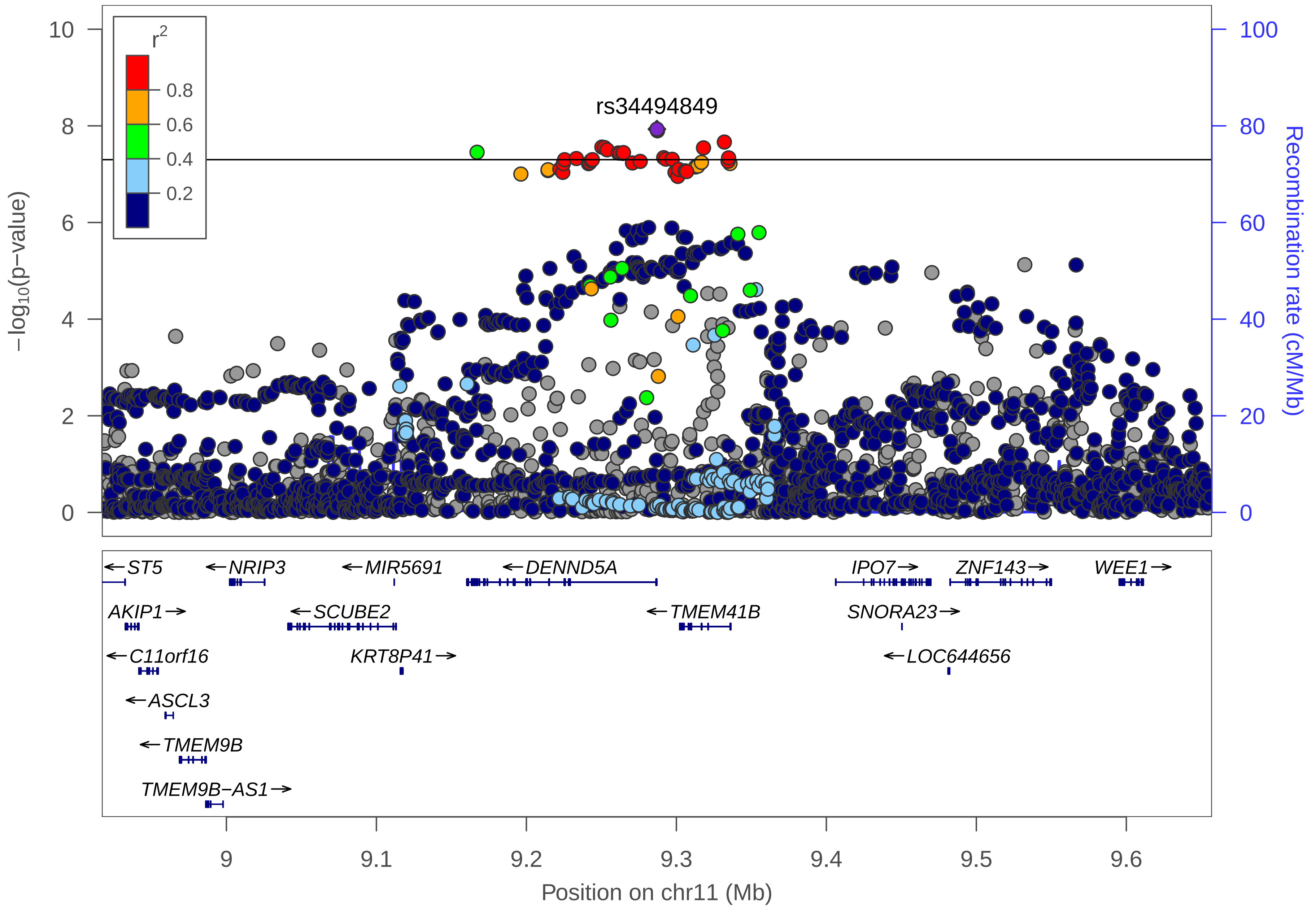

r2
0.8
0.6
0.4
0.2
rs34494849
−log10(p−value)
Recombination rate (cM/Mb)
0
2
4
6
8
10
0
20
40
60
80
100
ST5
NRIP3
MIR5691
DENND5A
IPO7
ZNF143
WEE1
AKIP1
SCUBE2
TMEM41B
SNORA23
C11orf16
KRT8P41
LOC644656
ASCL3
TMEM9B
TMEM9B−AS1
9
9.1
9.2
9.3
9.4
9.5
9.6
Position on chr11 (Mb)

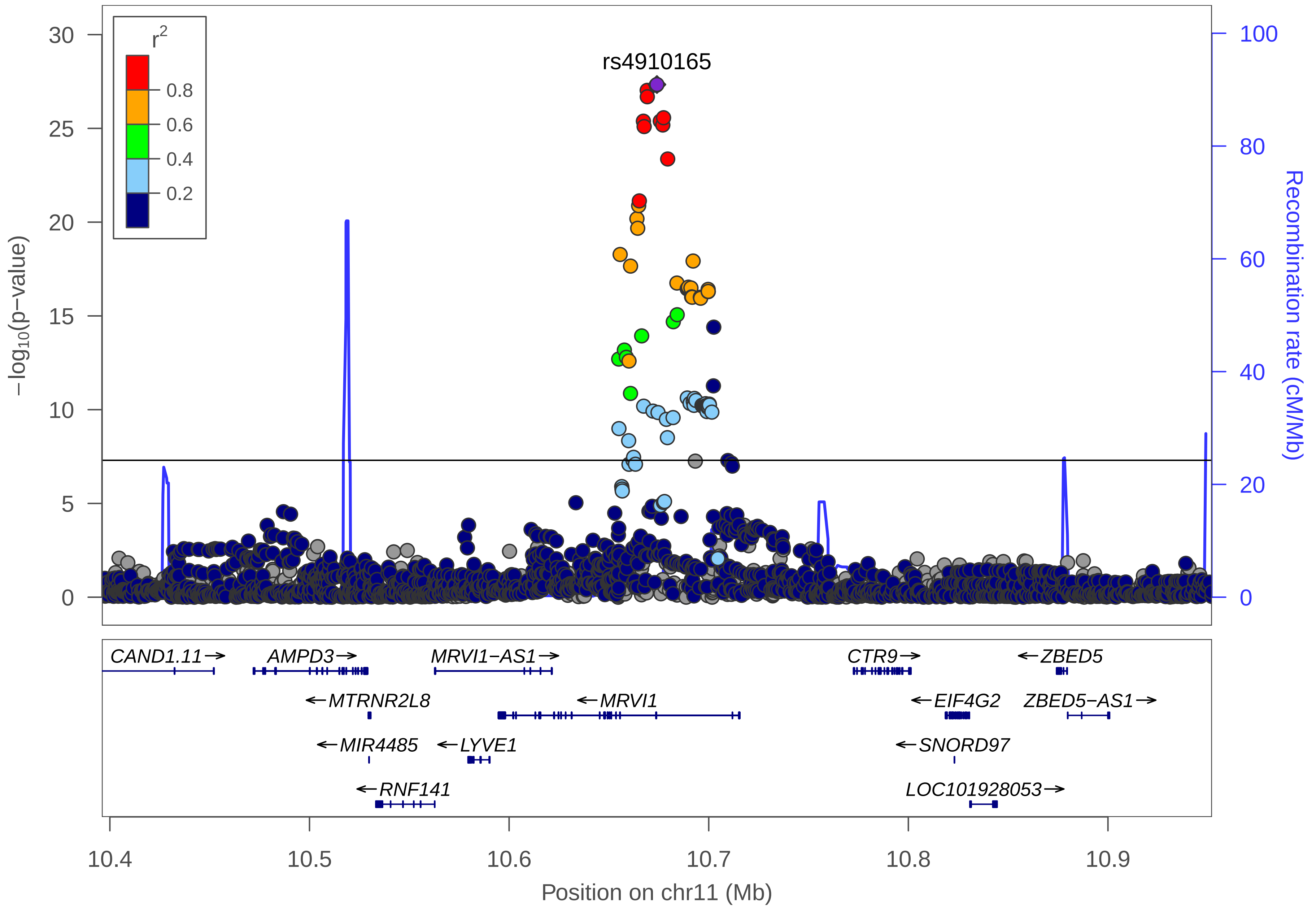

rs4910165
r2
0.8
0.6
0.4
0.2
30
25
20
15
10
5
0
−log10(p−value)
100
80
60
40
20
0
Recombination rate (cM/Mb)
CAND1.11→
AMPD3→
MRVI1−AS1→
CTR9→
←ZBED5
←MTRNR2L8
←MRVI1
←EIF4G2
ZBED5−AS1→
←MIR4485
←LYVE1
←SNORD97
←RNF141
LOC101928053→
10.4
10.5
10.6
10.7
10.8
10.9
Position on chr11 (Mb)

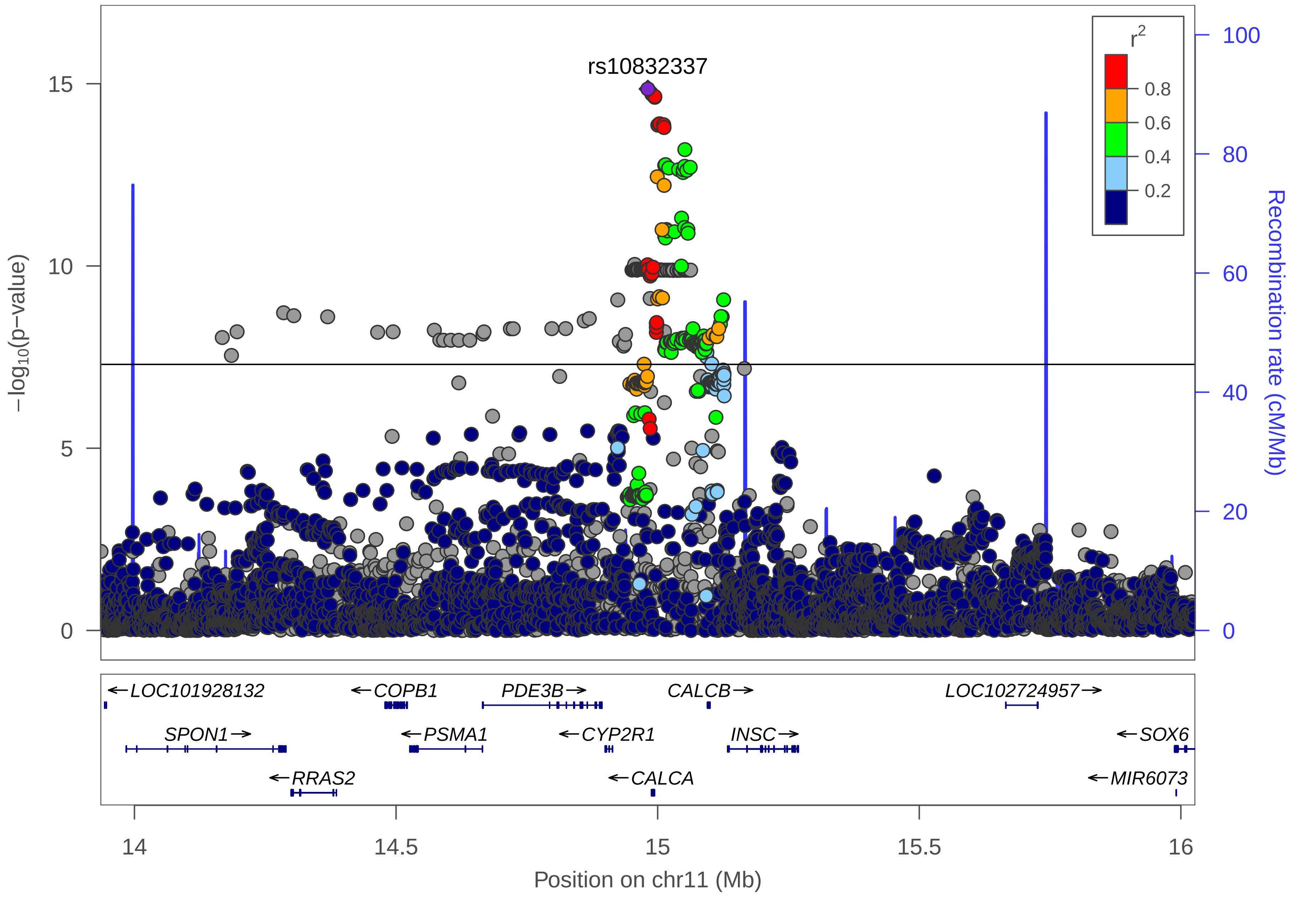

rs10832337
r2
0.8
0.6
0.4
0.2
−log10(p−value)
Recombination rate (cM/Mb)
LOC101928132
COPB1
PDE3B
CALCB
LOC102724957
SPON1
PSMA1
CYP2R1
INSC
SOX6
RRAS2
CALCA
MIR6073
Position on chr11 (Mb)

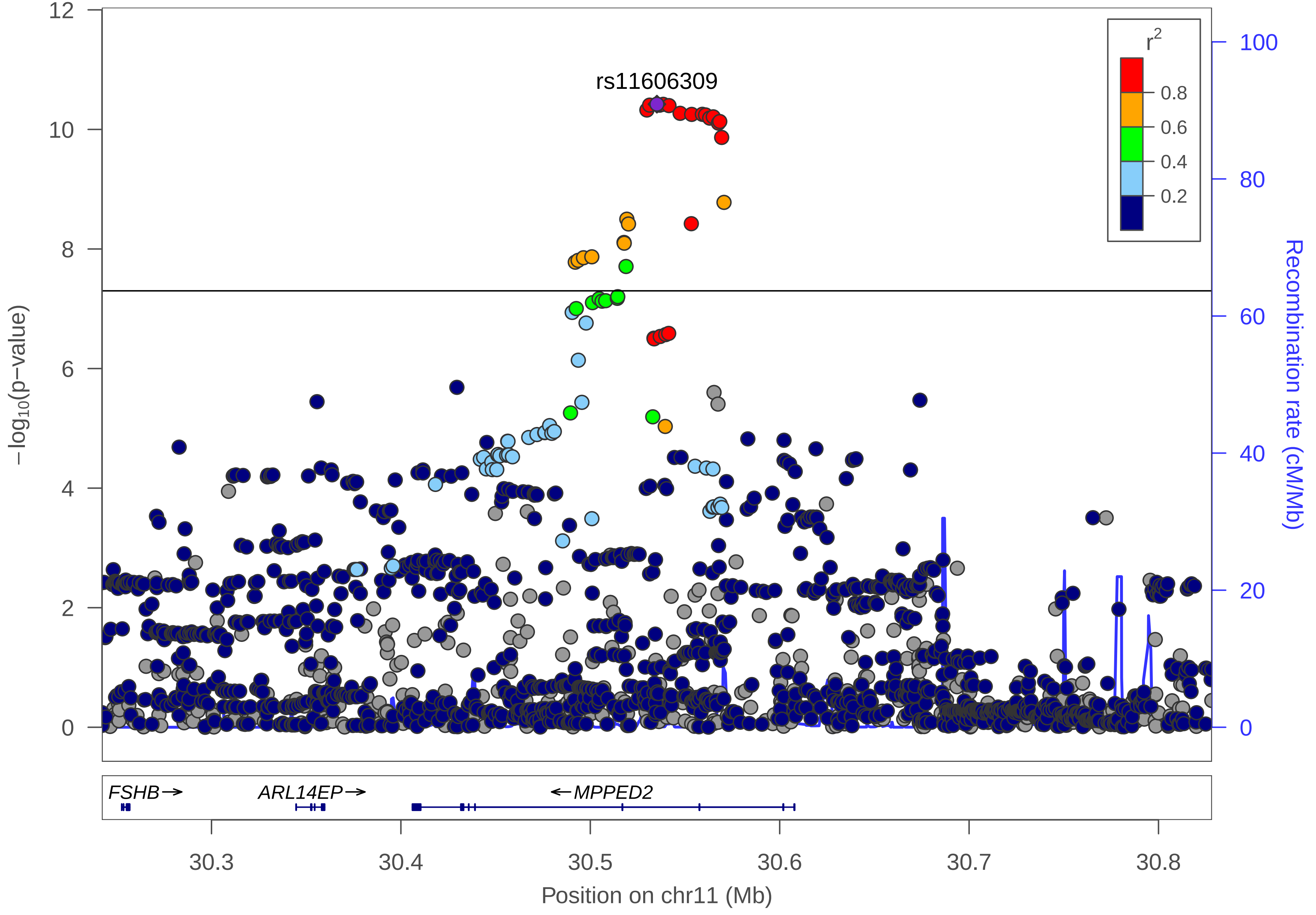
rs11606309
r2
0.8
0.6
0.4
0.2
−log10(p−value)
Recombination rate (cM/Mb)
12
10
8
6
4
2
0
100
80
60
40
20
0
FSHB→
ARL14EP→
←MPPED2
30.3
30.4
30.5
30.6
30.7
30.8
Position on chr11 (Mb)

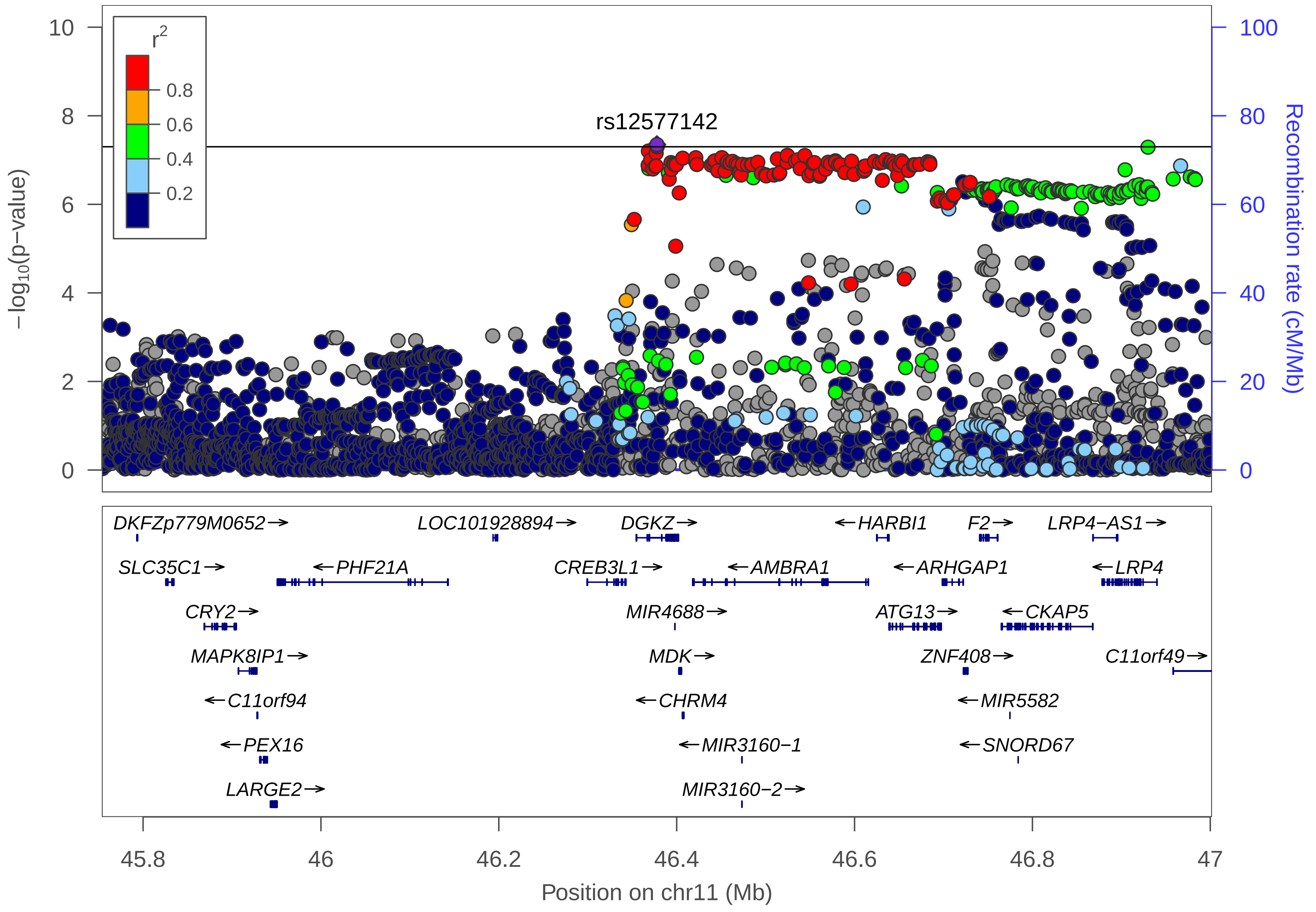

r2
0.8
0.6
0.4
0.2
rs12577142
−log10(p−value)
Recombination rate (cM/Mb)
10
8
6
4
2
0
100
80
60
40
20
0
DKFZp779M0652→
LOC101928894→
DGKZ→
←HARBI1
F2→
LRP4−AS1→
SLC35C1→
←PHF21A
CREB3L1→
←AMBRA1
←ARHGAP1
←LRP4
CRY2→
MIR4688→
ATG13→
←CKAP5
MAPK8IP1→
MDK→
ZNF408→
C11orf49→
←C11orf94
←CHRM4
←MIR5582
←PEX16
←MIR3160−1
←SNORD67
LARGE2→
MIR3160−2→
45.8
46
46.2
46.4
46.6
46.8
47
Position on chr11 (Mb)

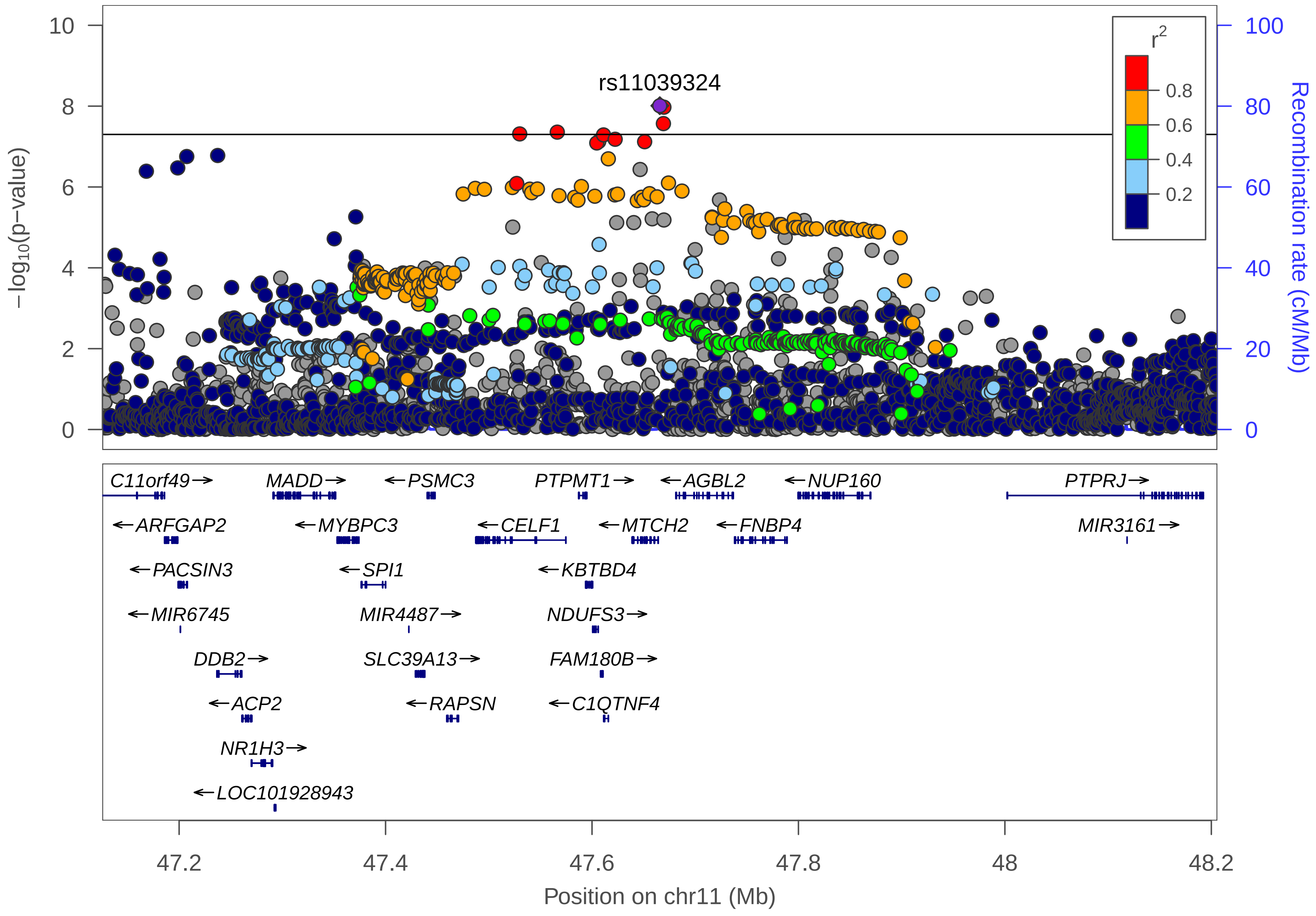

rs11039324
−log10(p−value)
Recombination rate (cM/Mb)
r2
0.8
0.6
0.4
0.2
C11orf49→
MADD→
←PSMC3
PTPMT1→
←AGBL2
←NUP160
PTPRJ→
←ARFGAP2
←MYBPC3
←CELF1
←MTCH2
←FNBP4
MIR3161→
←PACSIN3
←SPI1
←KBTBD4
←MIR6745
MIR4487→
NDUFS3→
DDB2→
SLC39A13→
FAM180B→
←ACP2
←RAPSN
←C1QTNF4
NR1H3→
←LOC101928943
47.2
47.4
47.6
47.8
48
48.2
Position on chr11 (Mb)

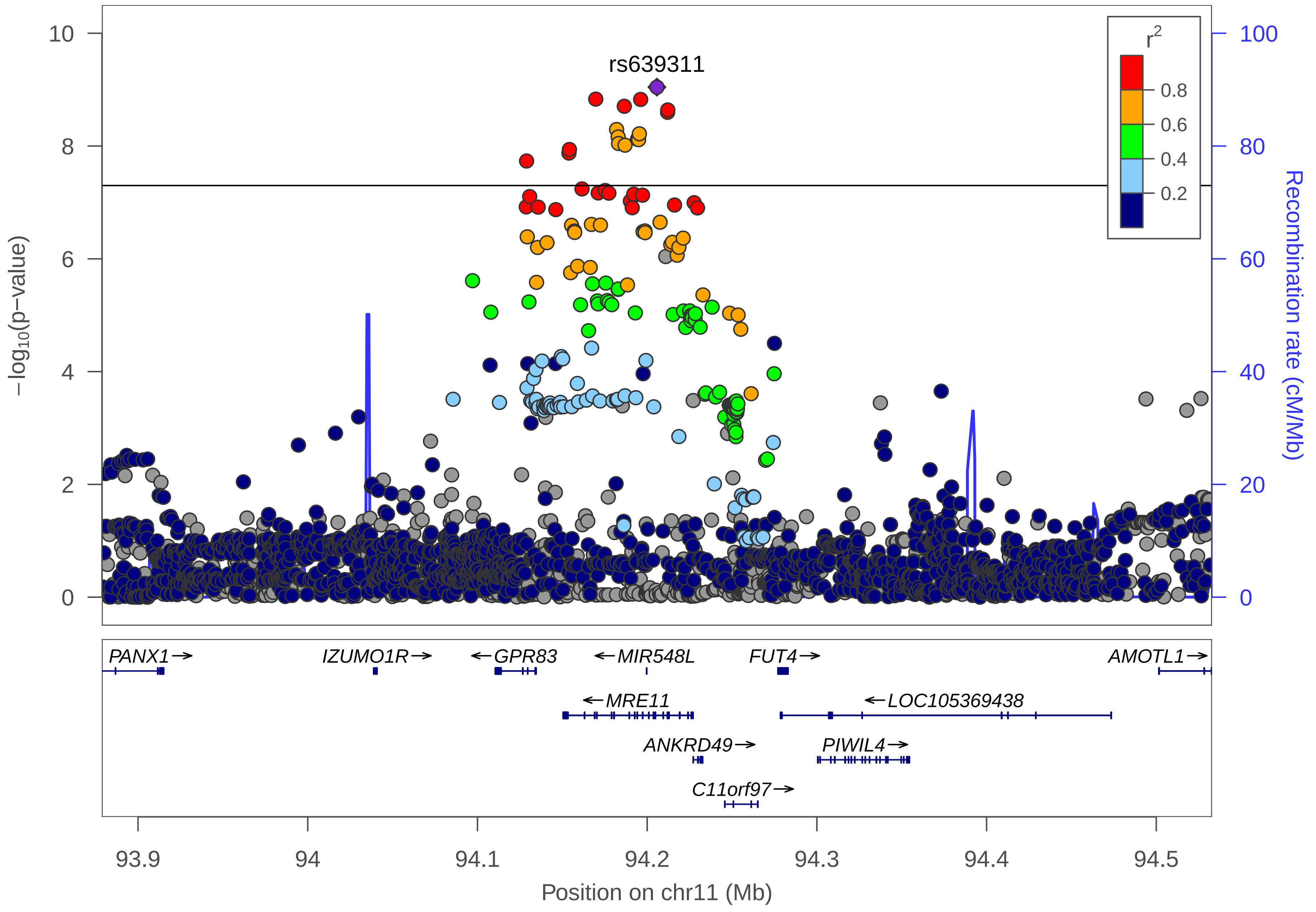

rs639311
r2
0.8
0.6
0.4
0.2
10
8
6
4
2
0
100
80
60
40
20
0
−log10(p−value)
Recombination rate (cM/Mb)
PANX1→
IZUMO1R→
←GPR83
←MIR548L
FUT4→
AMOTL1→
←MRE11
←LOC105369438
ANKRD49→
PIWIL4→
C11orf97→
93.9
94
94.1
94.2
94.3
94.4
94.5
Position on chr11 (Mb)

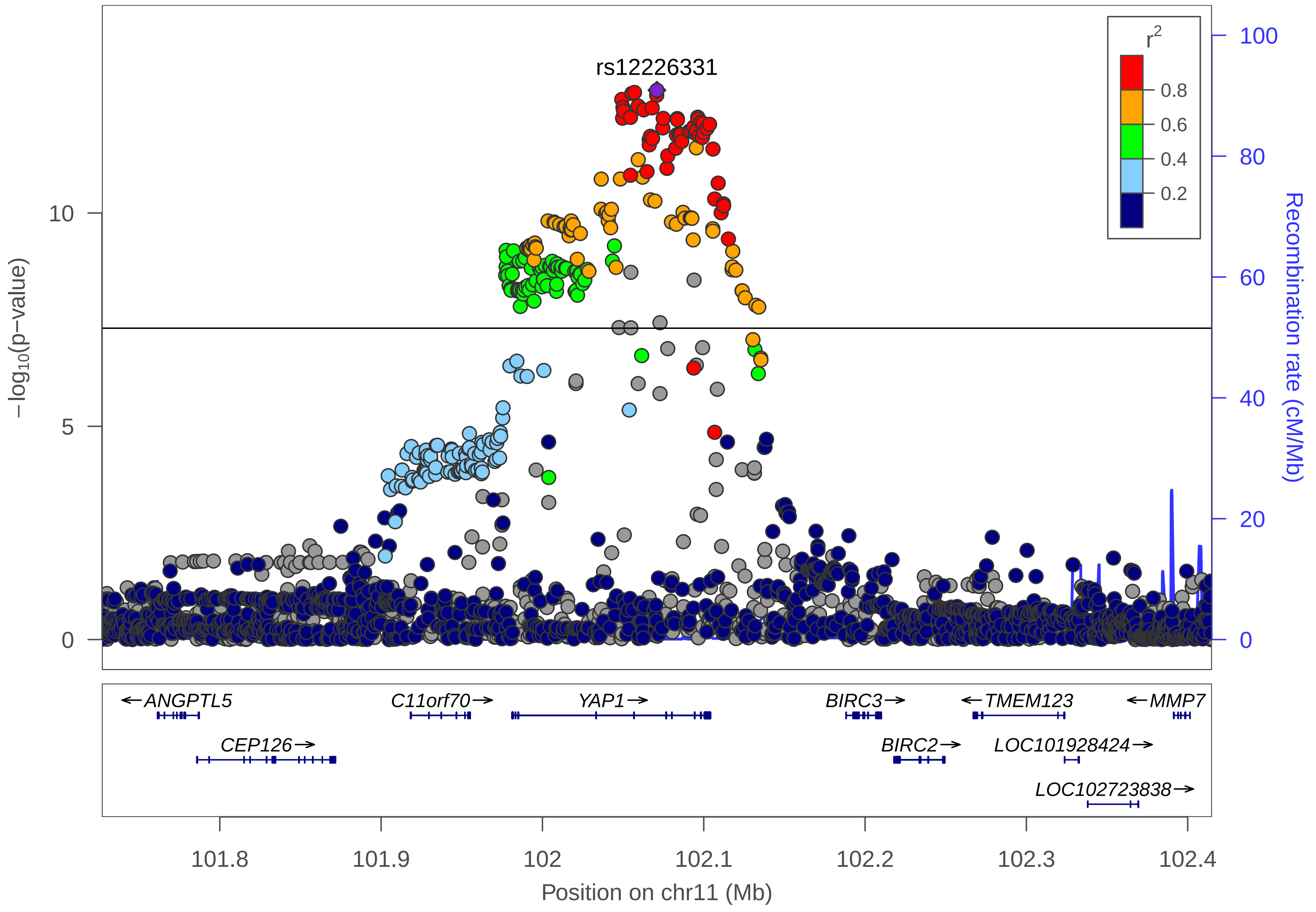

rs12226331
r2
0.8
0.6
0.4
0.2
−log10(p−value)
10
5
0
Recombination rate (cM/Mb)
100
80
60
40
20
0
ANGPTL5
CEP126
C11orf70
YAP1
BIRC3
BIRC2
TMEM123
LOC101928424
LOC102723838
MMP7
101.8
101.9
102
102.1
102.2
102.3
102.4
Position on chr11 (Mb)

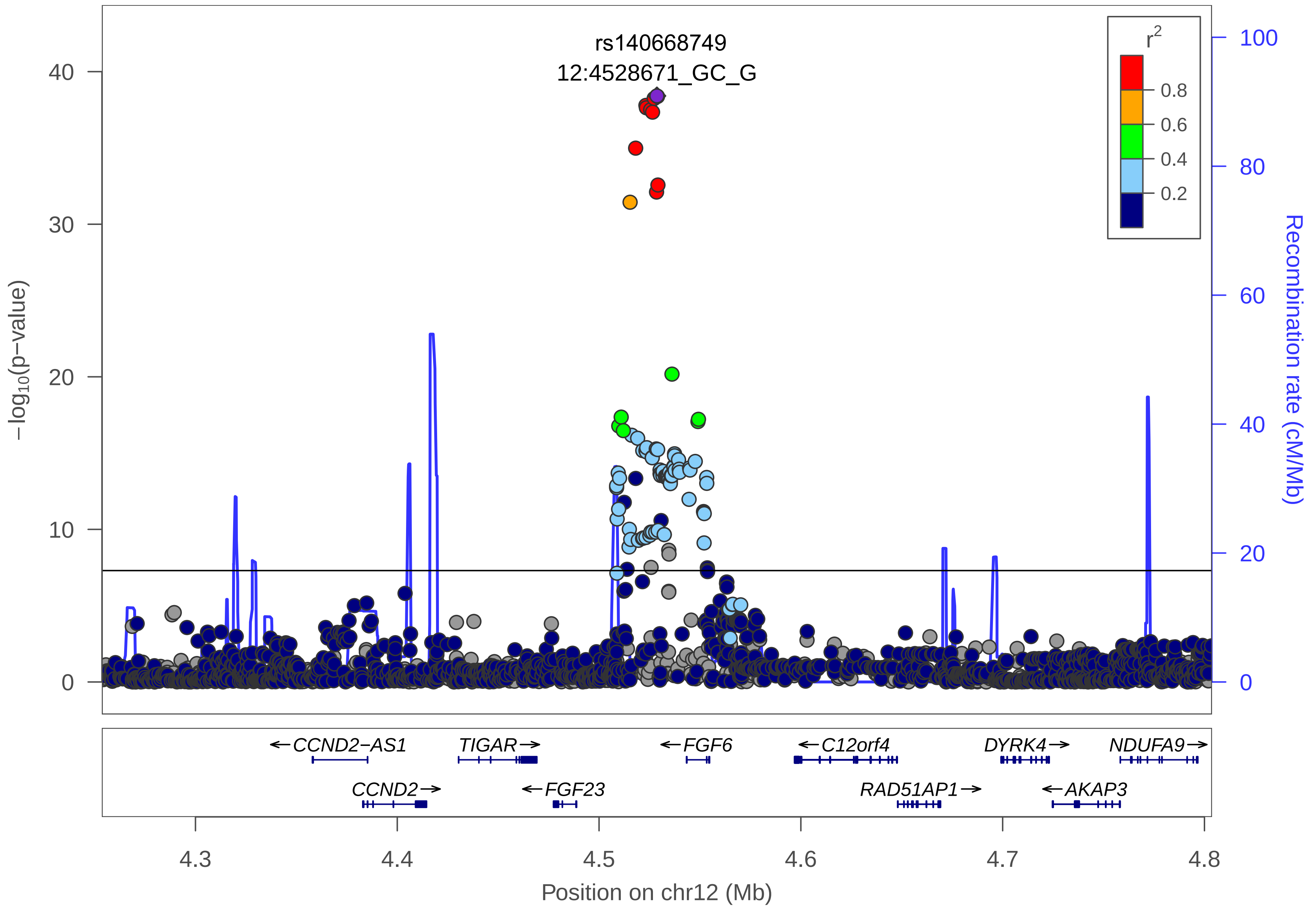

rs140668749
12:4528671_GC_G
$r^2$
0.8
0.6
0.4
0.2
$-\log_{10}$(p-value)
40
30
20
10
0
Recombination rate (cM/Mb)
100
80
60
40
20
0
←CCND2−AS1
TIGAR→
←FGF6
←C12orf4
DYRK4→
NDUFA9→
CCND2→
←FGF23
RAD51AP1→
←AKAP3
4.3
4.4
4.5
4.6
4.7
4.8
Position on chr12 (Mb)

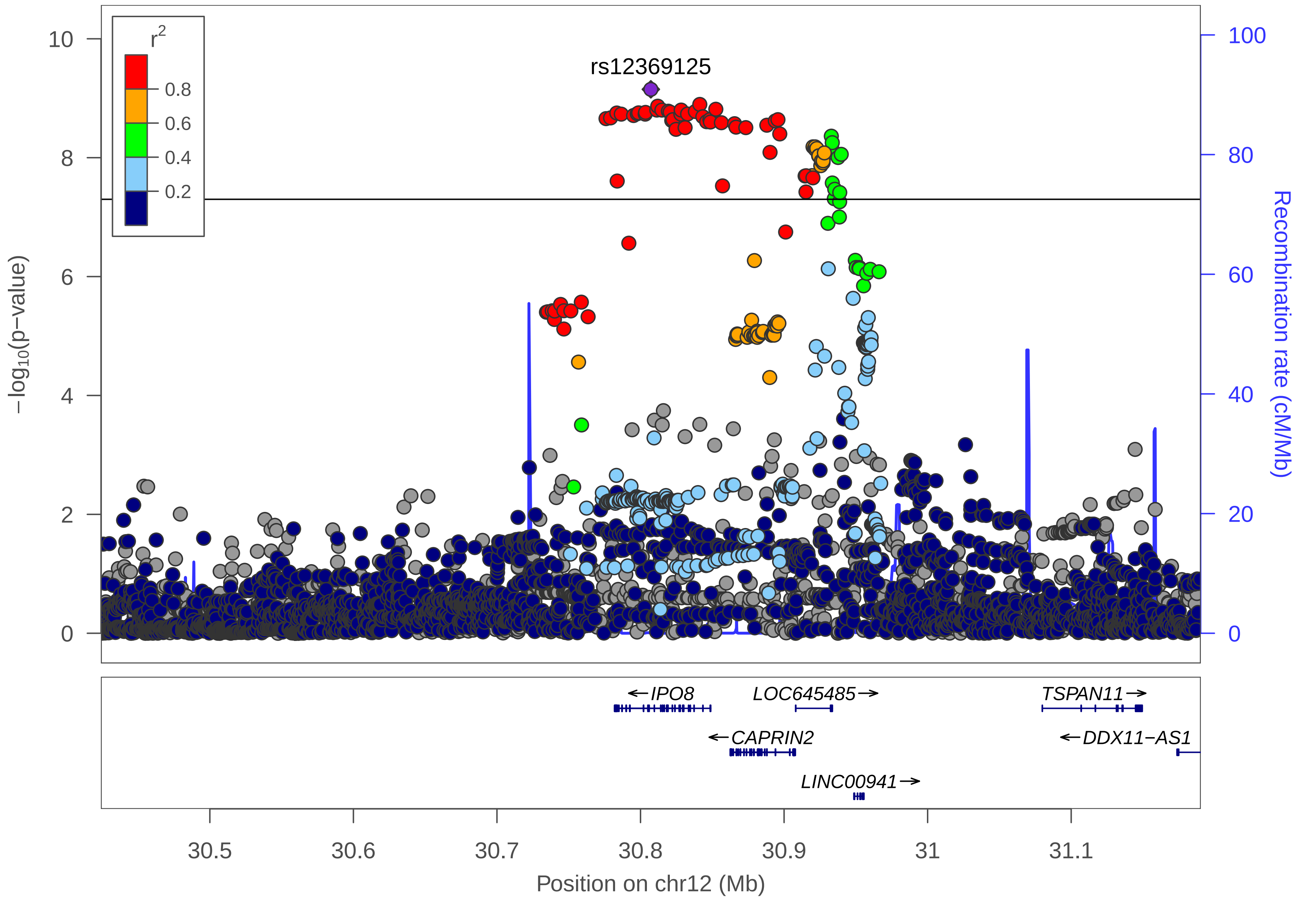

rs12369125
r2
0.8
0.6
0.4
0.2
−log10(p−value)
Recombination rate (cM/Mb)
IPO8
LOC645485
TSPAN11
CAPRIN2
DDX11−AS1
LINC00941
30.5
30.6
30.7
30.8
30.9
31
31.1
Position on chr12 (Mb)

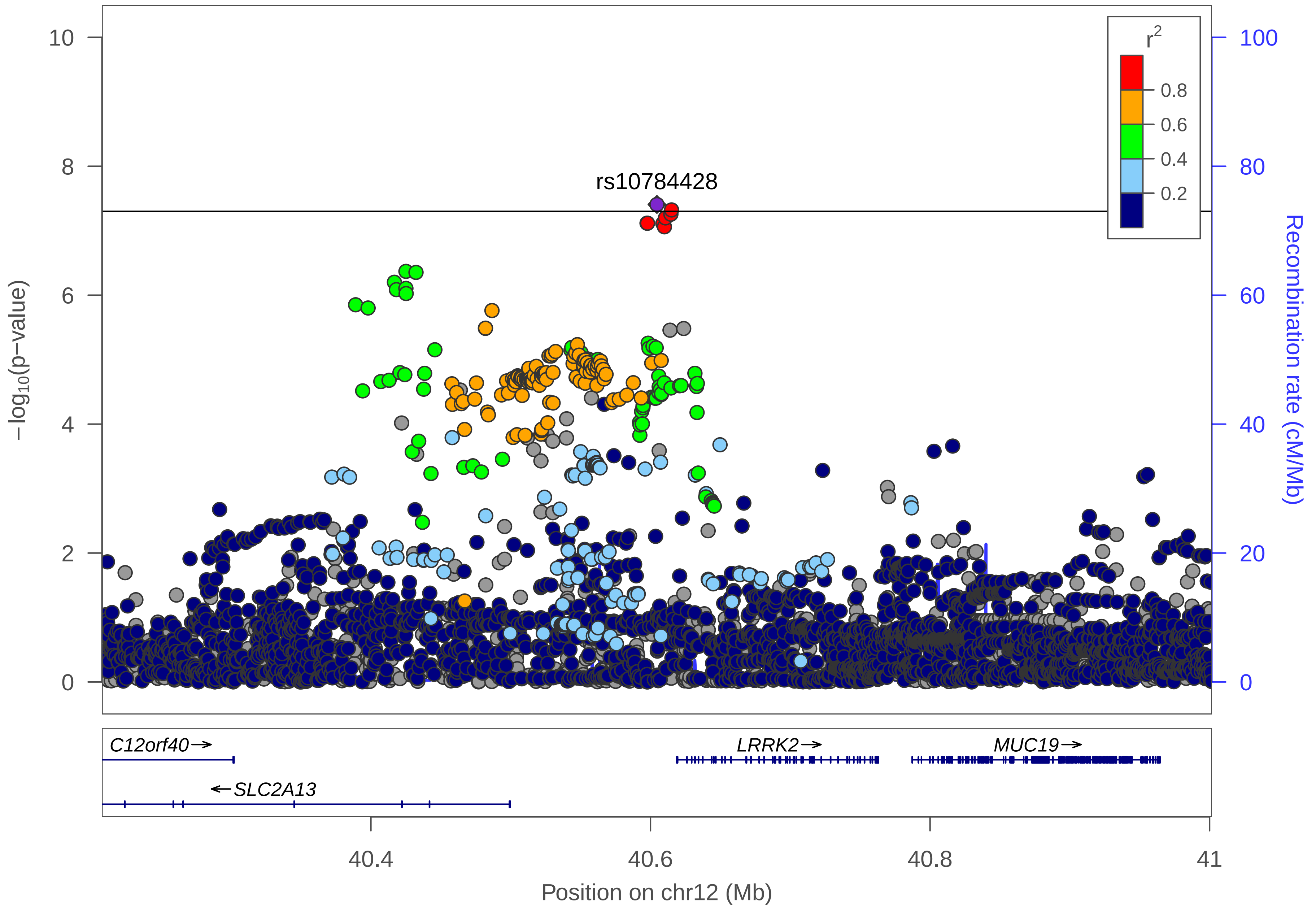
rs10784428
r2
0.8
0.6
0.4
0.2
−log10(p−value)
Recombination rate (cM/Mb)
0
2
4
6
8
10
0
20
40
60
80
100
C12orf40→
LRRK2→
MUC19→
←SLC2A13
40.4
40.6
40.8
41
Position on chr12 (Mb)

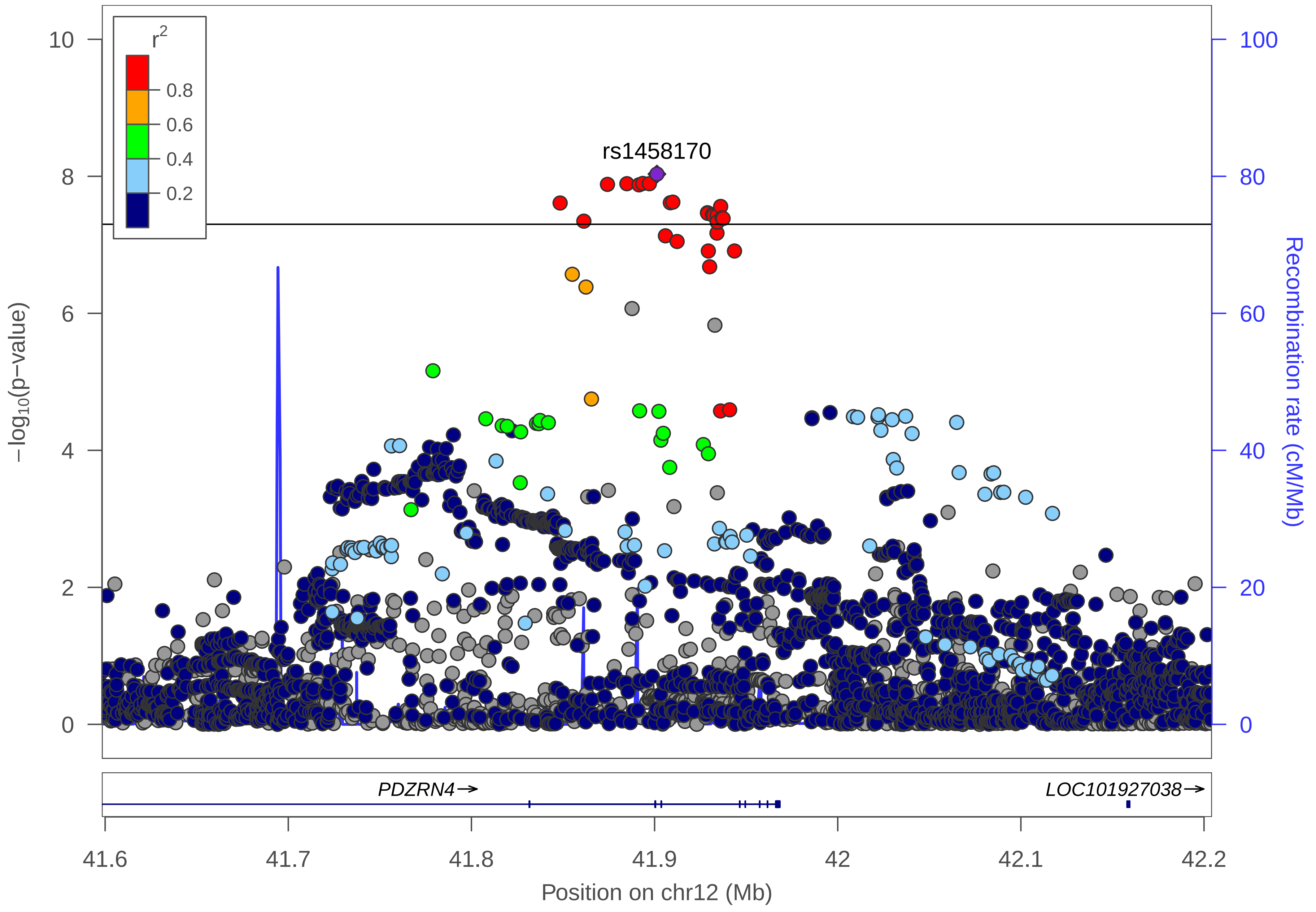

r2
0.8
0.6
0.4
0.2
rs1458170
−log10(p−value)
Recombination rate (cM/Mb)
0
2
4
6
8
10
0
20
40
60
80
100
PDZRN4→
LOC101927038→
41.6
41.7
41.8
41.9
42
42.1
42.2
Position on chr12 (Mb)

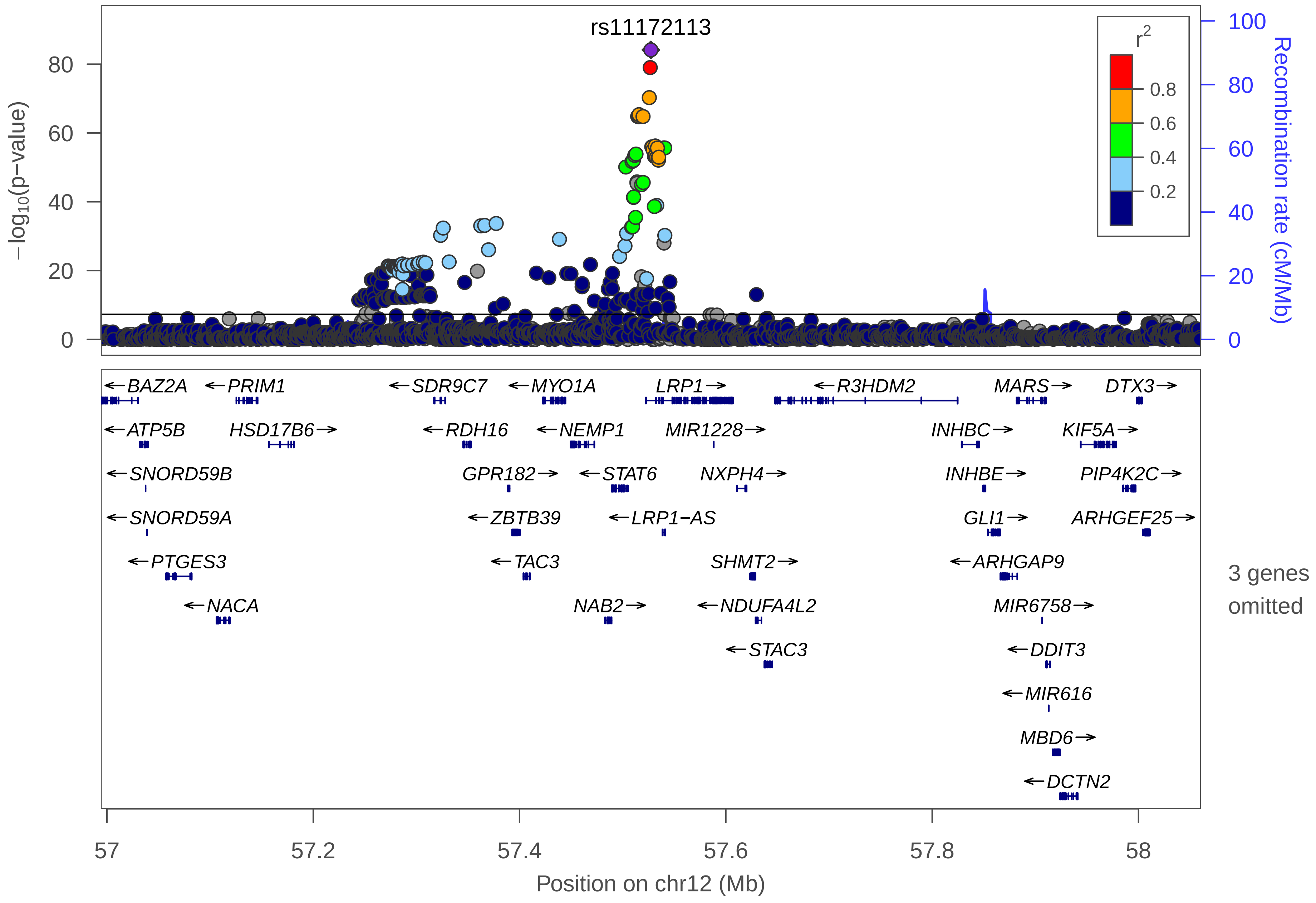

rs11172113
r2
0.8
0.6
0.4
0.2
−log10(p−value)
Recombination rate (cM/Mb)
BAZ2A
PRIM1
SDR9C7
MYO1A
LRP1
R3HDM2
MARS
DTX3
ATP5B
HSD17B6
RDH16
NEMP1
MIR1228
INHBC
KIF5A
SNORD59B
GPR182
STAT6
NXPH4
INHBE
PIP4K2C
SNORD59A
ZBTB39
LRP1−AS
GLI1
ARHGEF25
PTGES3
TAC3
SHMT2
ARHGAP9
NACA
NAB2
NDUFA4L2
MIR6758
STAC3
DDIT3
MIR616
MBD6
DCTN2
3 genes omitted
57
57.2
57.4
57.6
57.8
58
Position on chr12 (Mb)

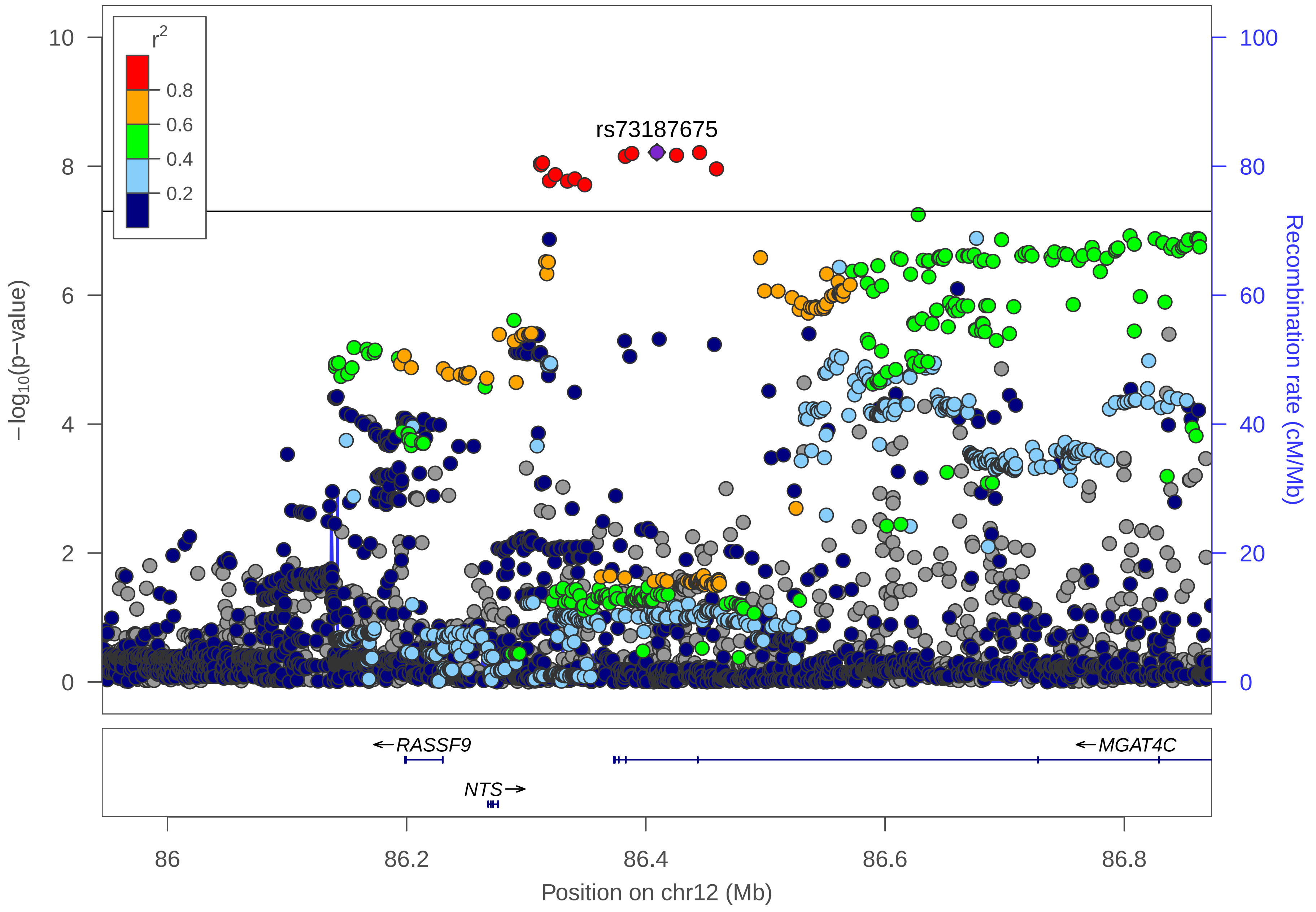
r2
0.8
0.6
0.4
0.2
rs73187675
−log10(p−value)
Recombination rate (cM/Mb)
RASSF9
MGAT4C
NTS
86
86.2
86.4
86.6
86.8
Position on chr12 (Mb)
0
2
4
6
8
10
0
20
40
60
80
100

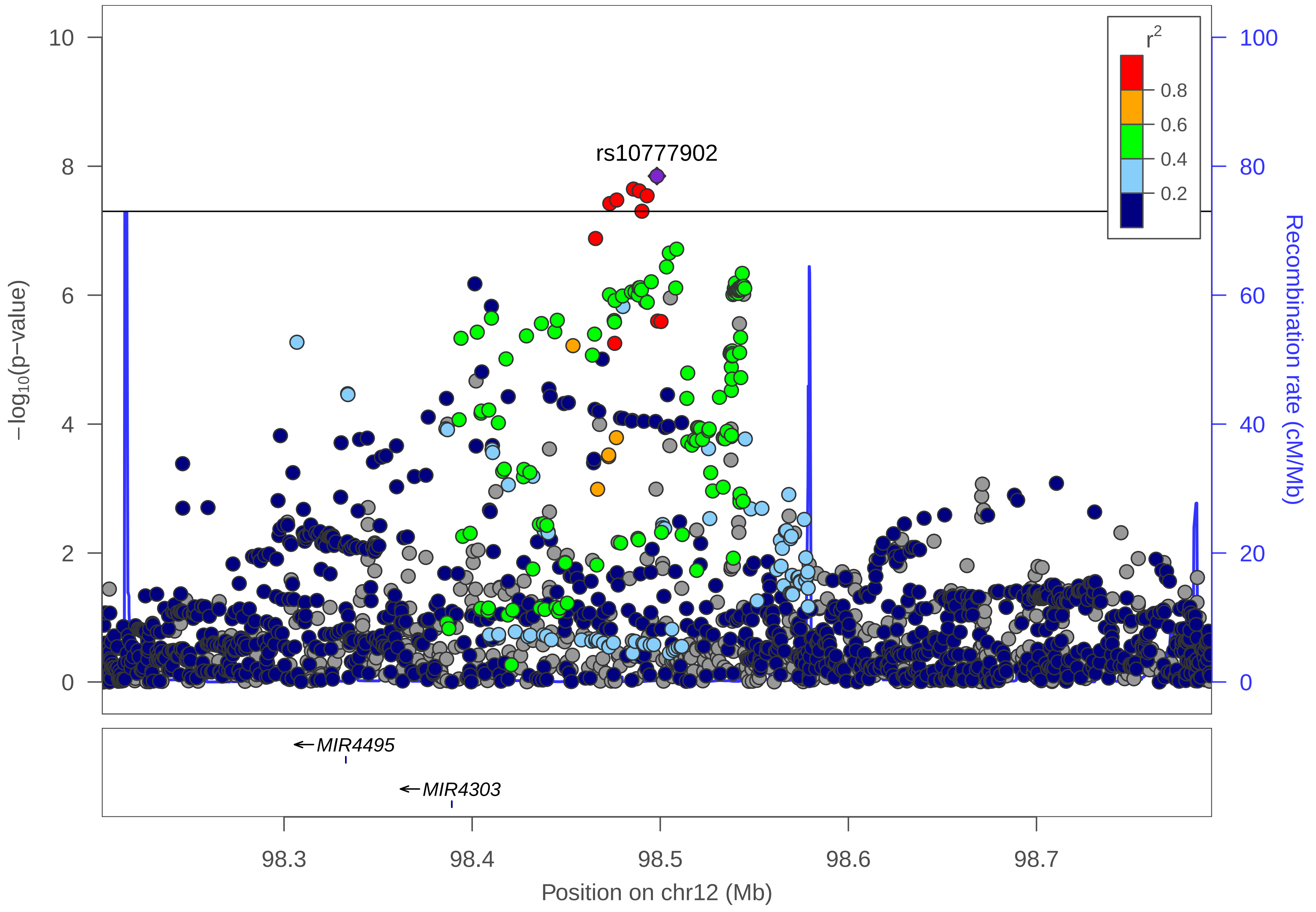
rs10777902
r^2
0.8
0.6
0.4
0.2
−log10(p−value)
Recombination rate (cM/Mb)
10
8
6
4
2
0
100
80
60
40
20
0
MIR4495
MIR4303
98.3
98.4
98.5
98.6
98.7
Position on chr12 (Mb)

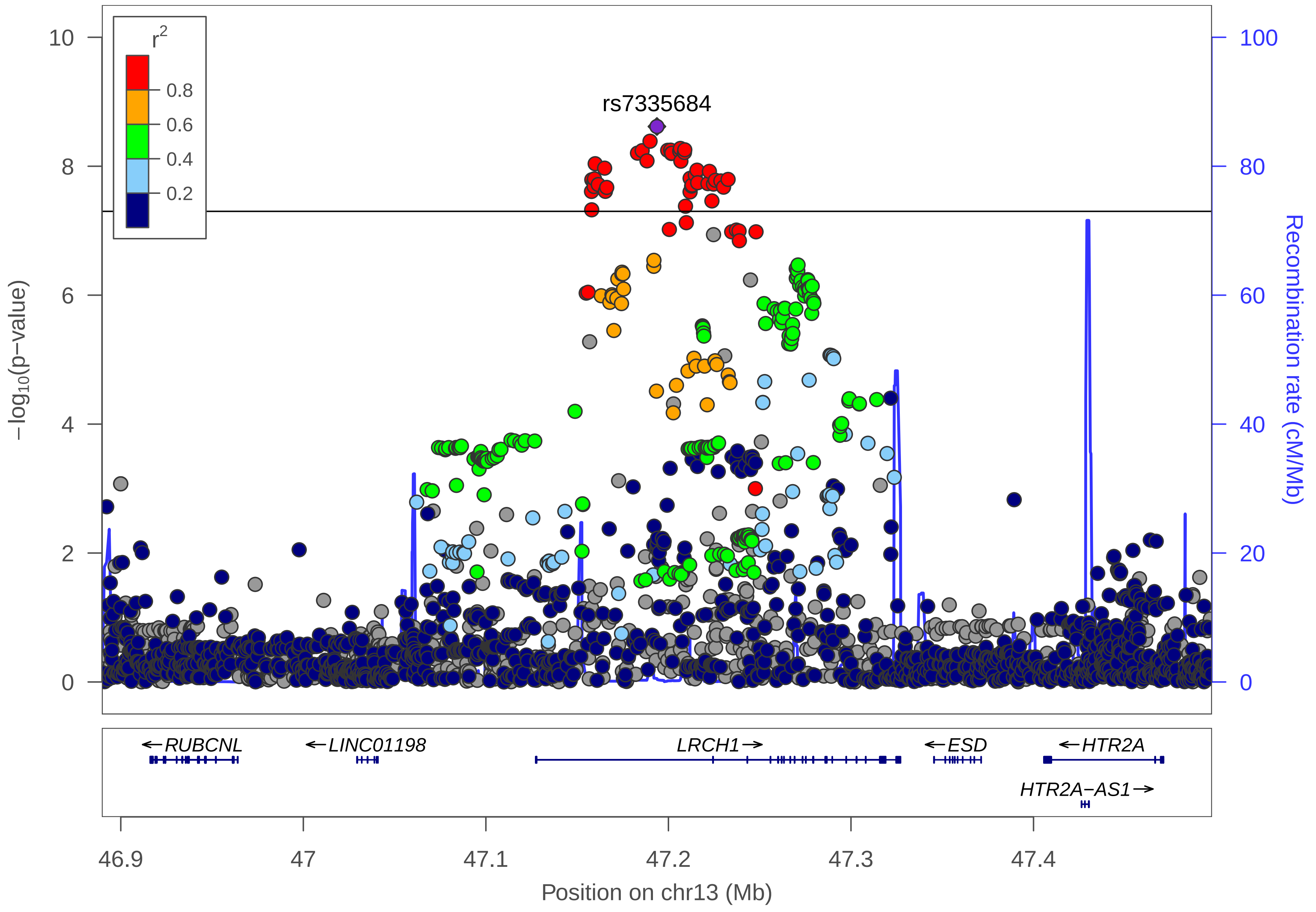

r2
0.8
0.6
0.4
0.2
rs7335684
10
8
6
4
2
0
−log10(p−value)
100
80
60
40
20
0
Recombination rate (cM/Mb)
←RUBCNL
←LINC01198
LRCH1→
←ESD
←HTR2A
HTR2A−AS1→
46.9
47
47.1
47.2
47.3
47.4
Position on chr13 (Mb)

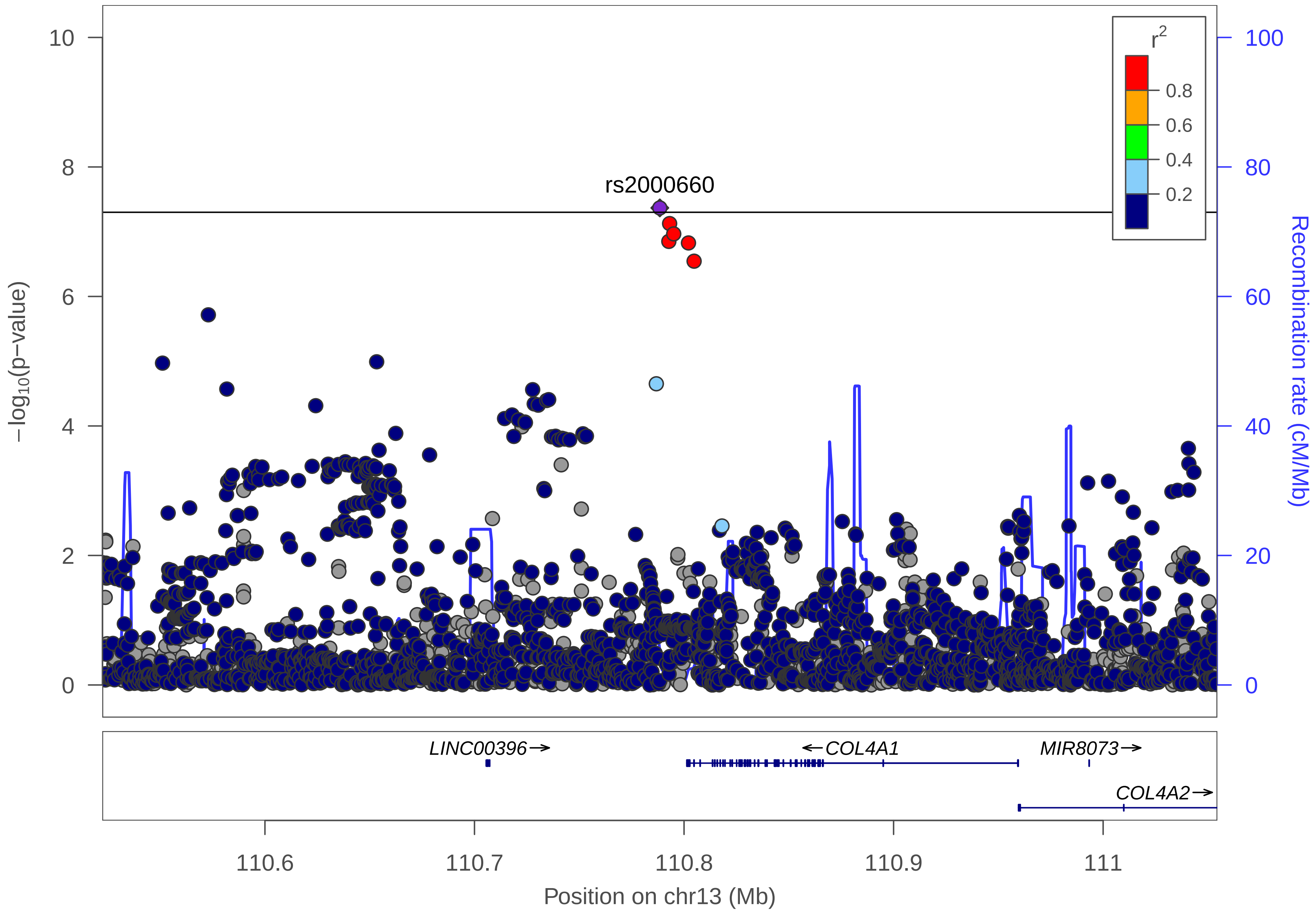

rs2000660
r2
0.8
0.6
0.4
0.2
−log10(p−value)
Recombination rate (cM/Mb)
10
8
6
4
2
0
100
80
60
40
20
0
LINC00396→
←COL4A1
MIR8073→
COL4A2→
110.6
110.7
110.8
110.9
111
Position on chr13 (Mb)

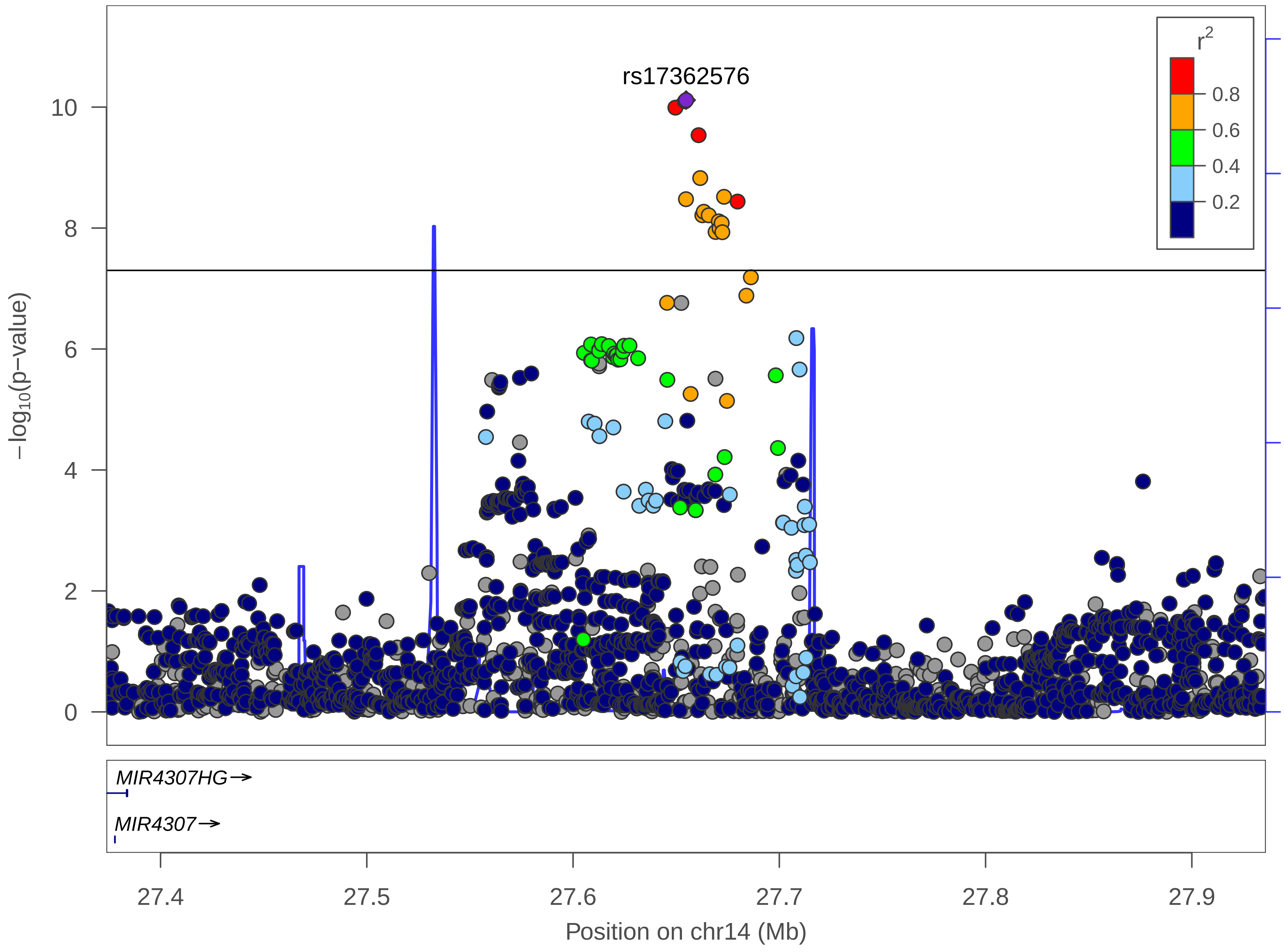

rs17362576
r^2
0.8
0.6
0.4
0.2
−log10(p−value)
10
8
6
4
2
0
Recombination rate (cM/Mb)
100
80
60
40
20
0
MIR4307HG→
MIR4307→
27.4
27.5
27.6
27.7
27.8
27.9
Position on chr14 (Mb)

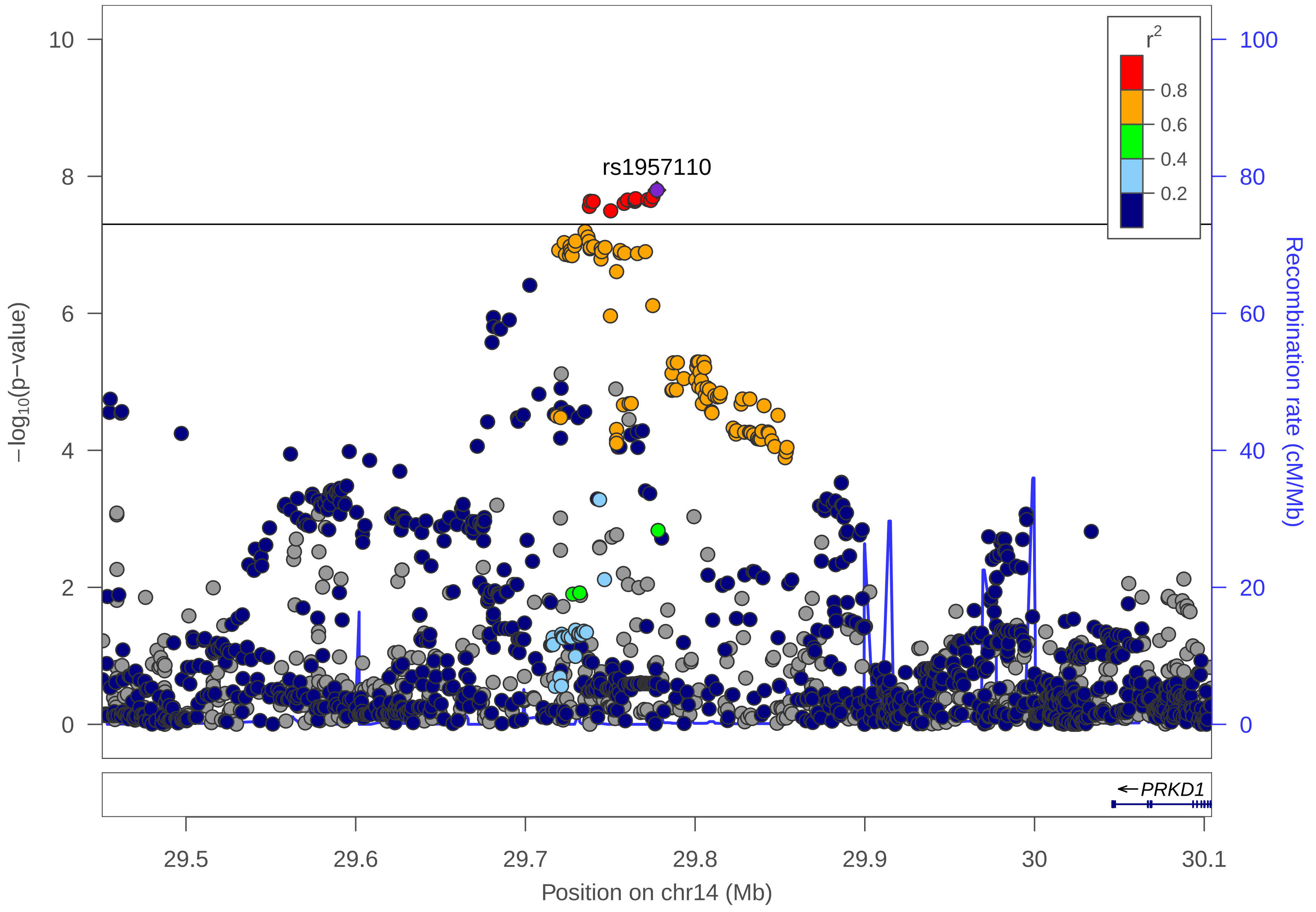

rs1957110
r2
0.8
0.6
0.4
0.2
−log10(p−value)
Recombination rate (cM/Mb)
0
2
4
6
8
10
0
20
40
60
80
100
PRKD1
29.5
29.6
29.7
29.8
29.9
30
30.1
Position on chr14 (Mb)

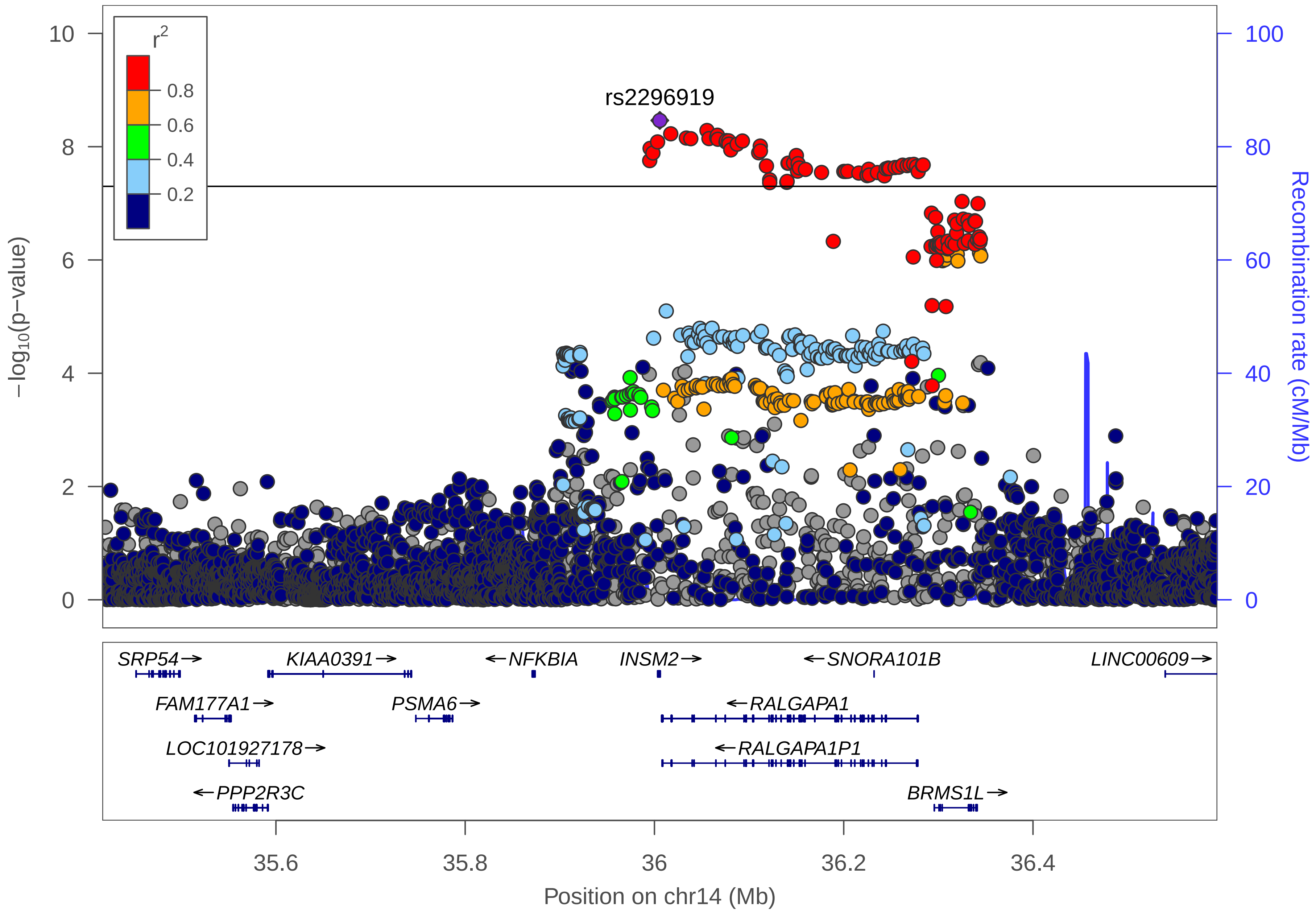

r2
0.8
0.6
0.4
0.2
rs2296919
−log10(p−value)
Recombination rate (cM/Mb)
SRP54→
KIAA0391→
←NFKBIA
INSM2→
←SNORA101B
LINC00609→
FAM177A1→
PSMA6→
←RALGAPA1
LOC101927178→
←RALGAPA1P1
←PPP2R3C
BRMS1L→
35.6
35.8
36
36.2
36.4
Position on chr14 (Mb)

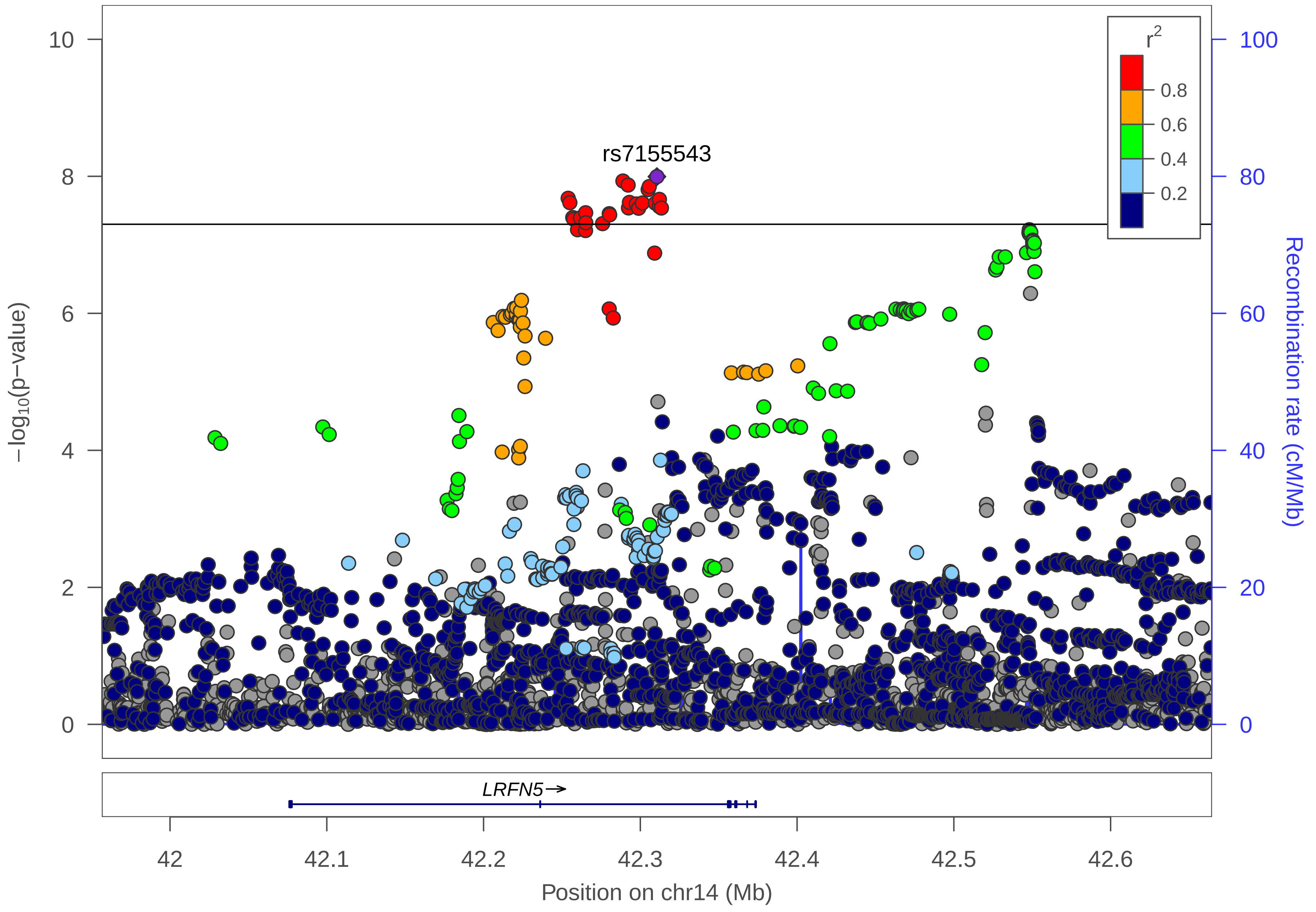

rs7155543
r²
0.8
0.6
0.4
0.2
10
8
6
4
2
0
100
80
60
40
20
0
−log10(p−value)
Recombination rate (cM/Mb)
LRFN5→
42
42.1
42.2
42.3
42.4
42.5
42.6
Position on chr14 (Mb)

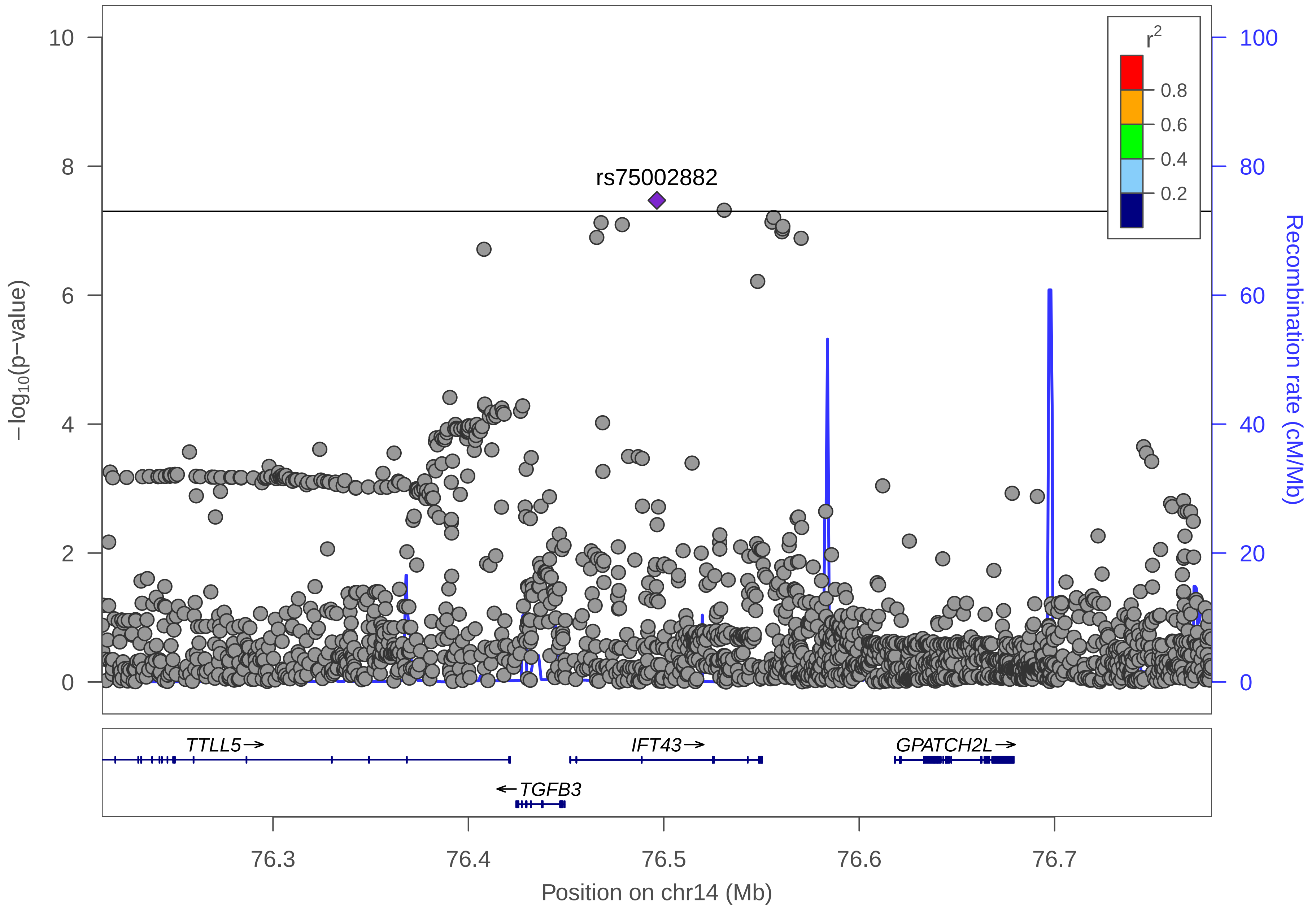

rs75002882
r2
0.8
0.6
0.4
0.2
−log10(p−value)
Recombination rate (cM/Mb)
10
8
6
4
2
0
100
80
60
40
20
0
TTLL5→
IFT43→
GPATCH2L→
←TGFB3
76.3
76.4
76.5
76.6
76.7
Position on chr14 (Mb)

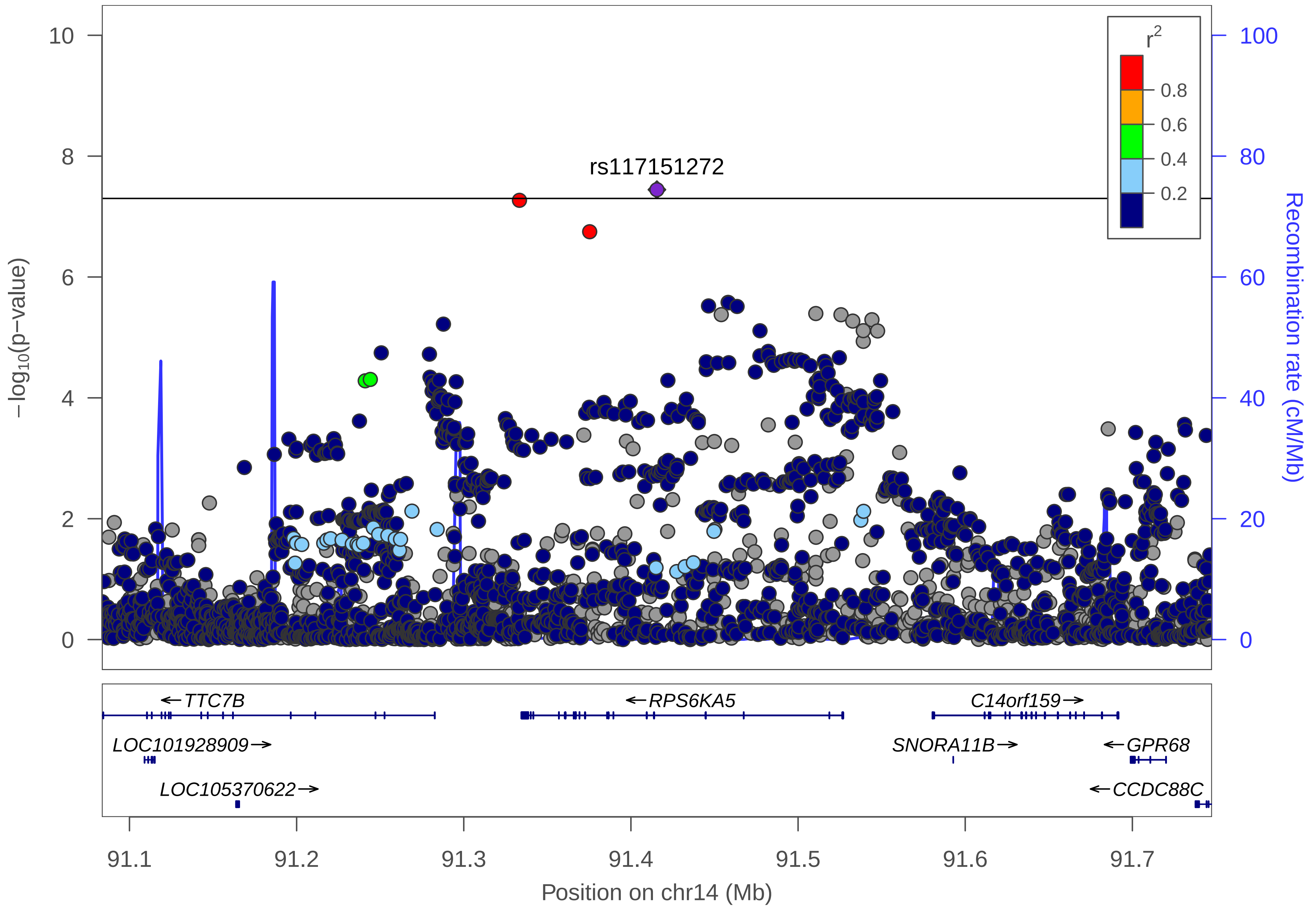

rs117151272
r2
0.8
0.6
0.4
0.2
−log10(p−value)
Recombination rate (cM/Mb)
0
2
4
6
8
10
0
20
40
60
80
100
←TTC7B
←RPS6KA5
C14orf159→
LOC101928909→
SNORA11B→
←GPR68
LOC105370622→
←CCDC88C
91.1
91.2
91.3
91.4
91.5
91.6
91.7
Position on chr14 (Mb)

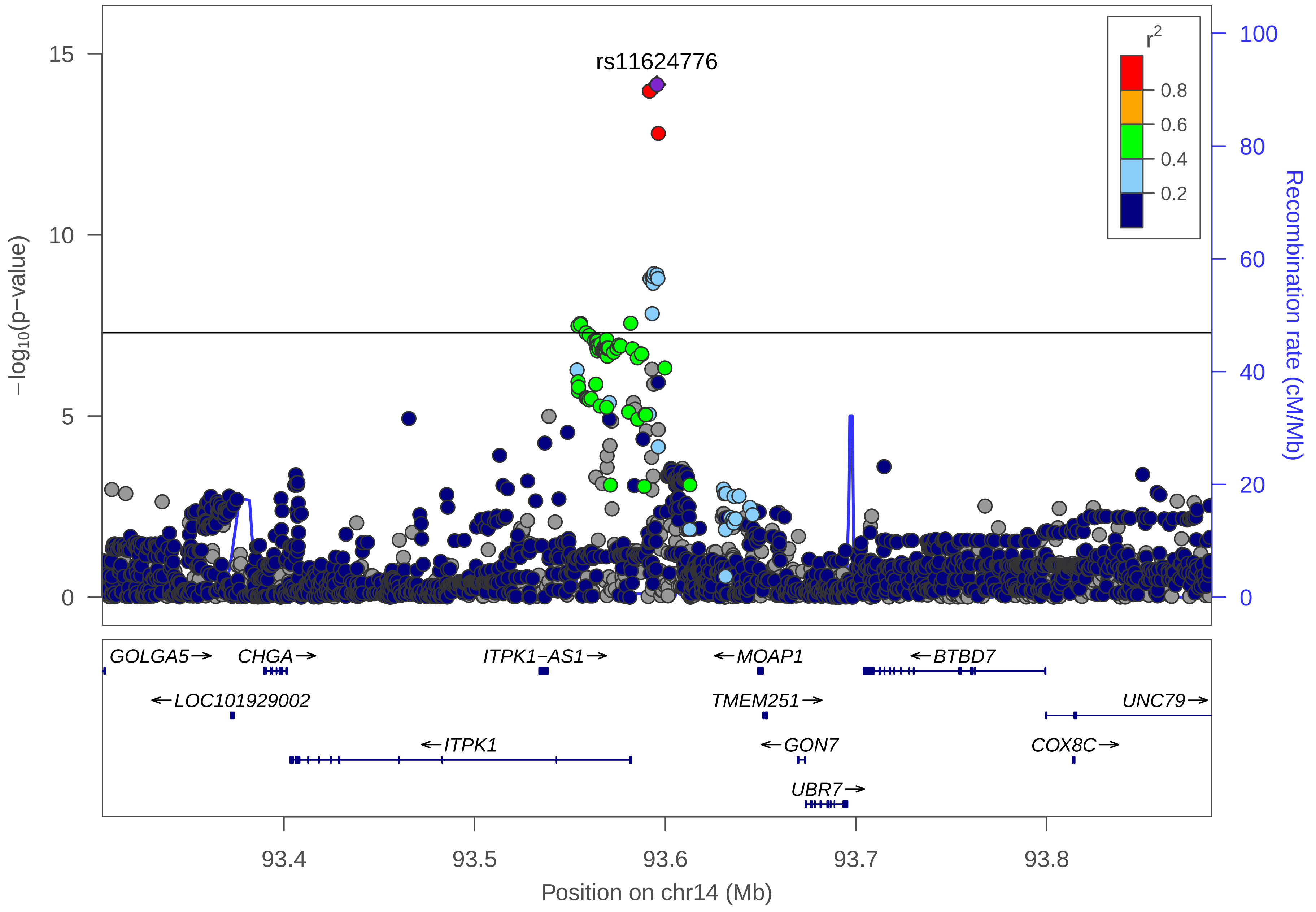
rs11624776
r2
0.8
0.6
0.4
0.2
−log10(p−value)
Recombination rate (cM/Mb)
15
10
5
0
100
80
60
40
20
0
GOLGA5→
CHGA→
ITPK1−AS1→
←MOAP1
←BTBD7
←LOC101929002
TMEM251→
UNC79→
←ITPK1
←GON7
COX8C→
UBR7→
93.4
93.5
93.6
93.7
93.8
Position on chr14 (Mb)

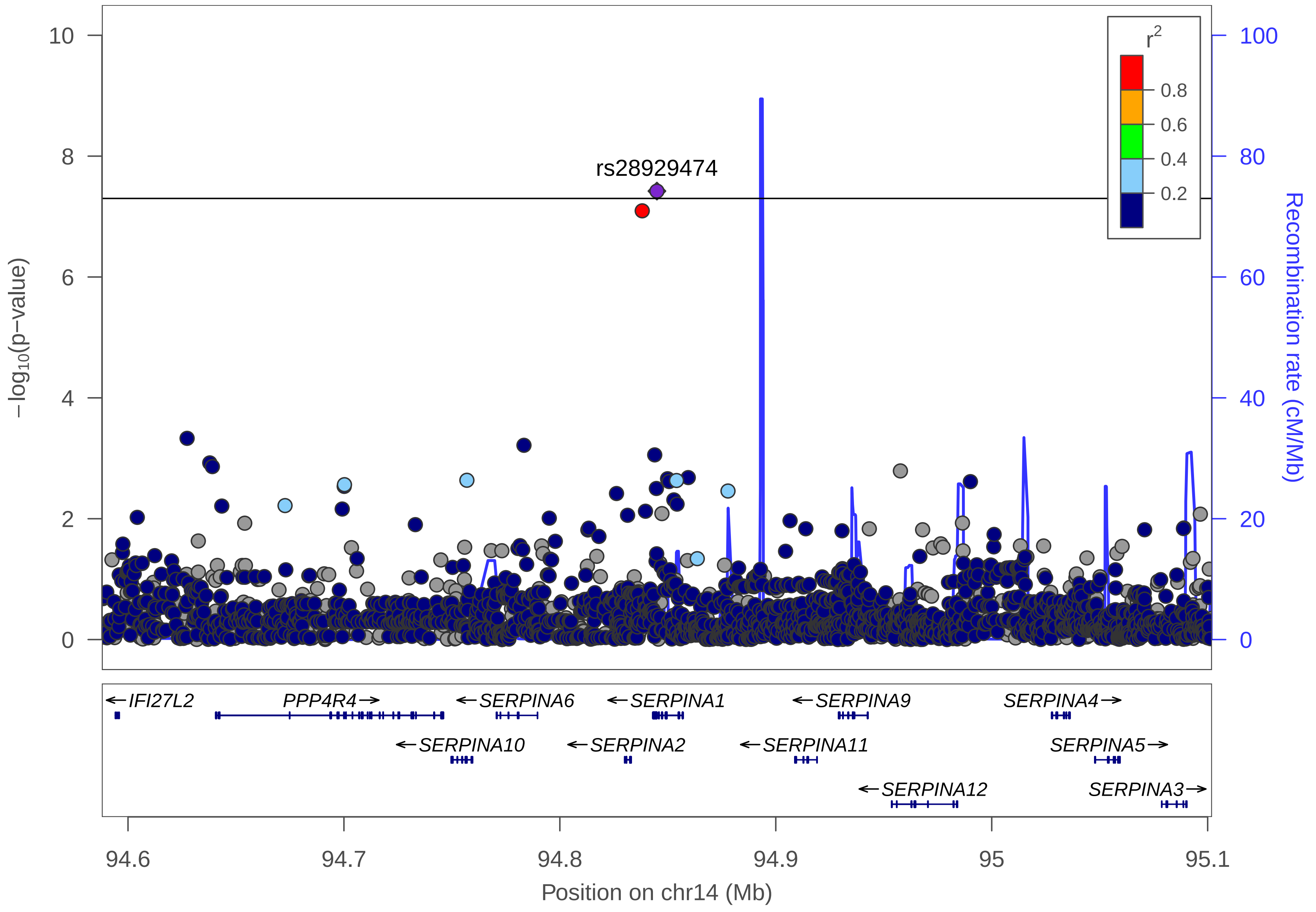

rs28929474
−log10(p−value)
Recombination rate (cM/Mb)
r2
0.8
0.6
0.4
0.2
0
2
4
6
8
10
20
40
60
80
100
IFI27L2
PPP4R4
SERPINA6
SERPINA1
SERPINA9
SERPINA4
SERPINA10
SERPINA2
SERPINA11
SERPINA5
SERPINA12
SERPINA3
94.6
94.7
94.8
94.9
95
95.1
Position on chr14 (Mb)

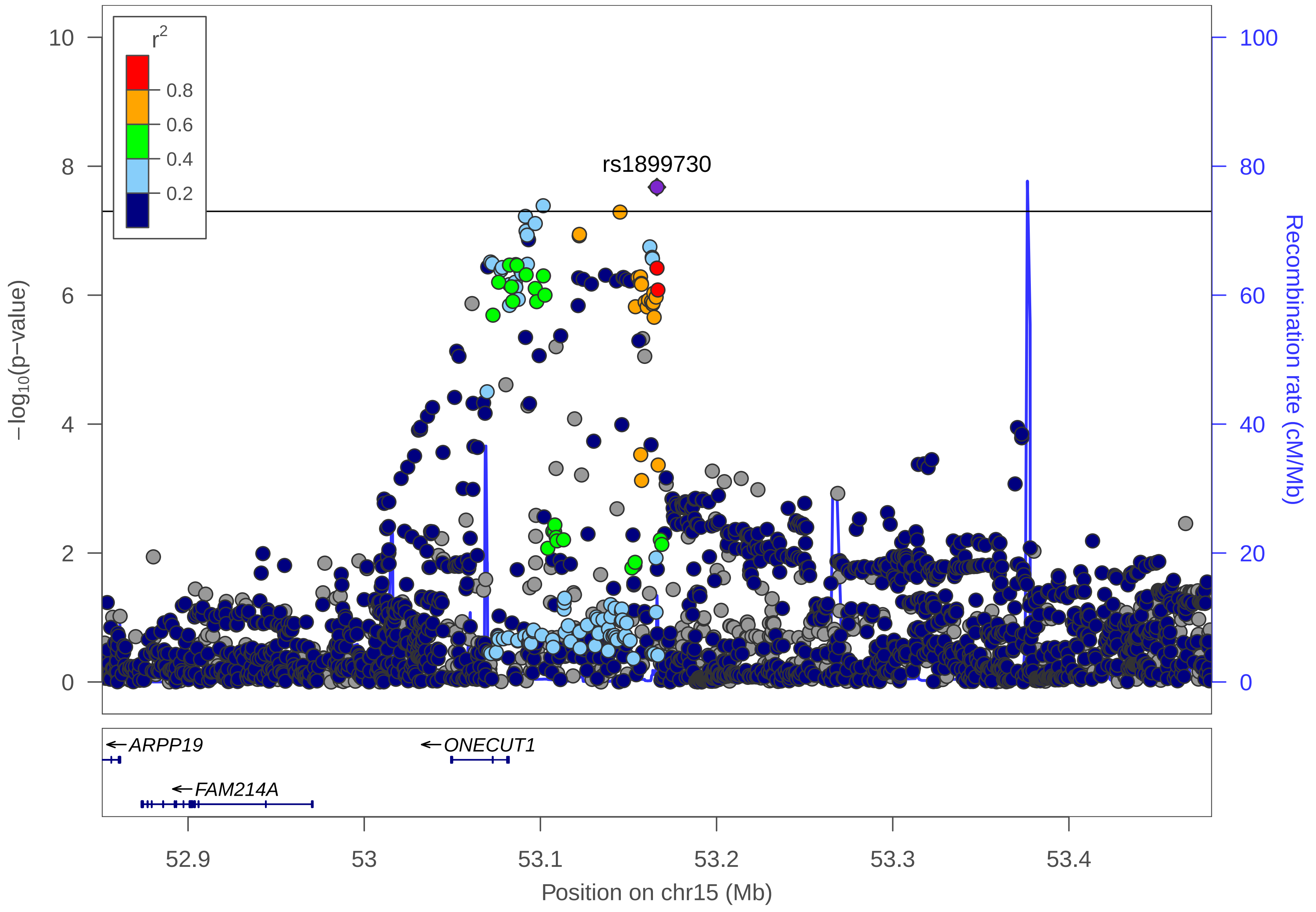

r²
0.8
0.6
0.4
0.2
rs1899730
10
8
6
4
2
0
−log10(p−value)
100
80
60
40
20
0
Recombination rate (cM/Mb)
ARPP19
ONECUT1
FAM214A
52.9
53
53.1
53.2
53.3
53.4
Position on chr15 (Mb)

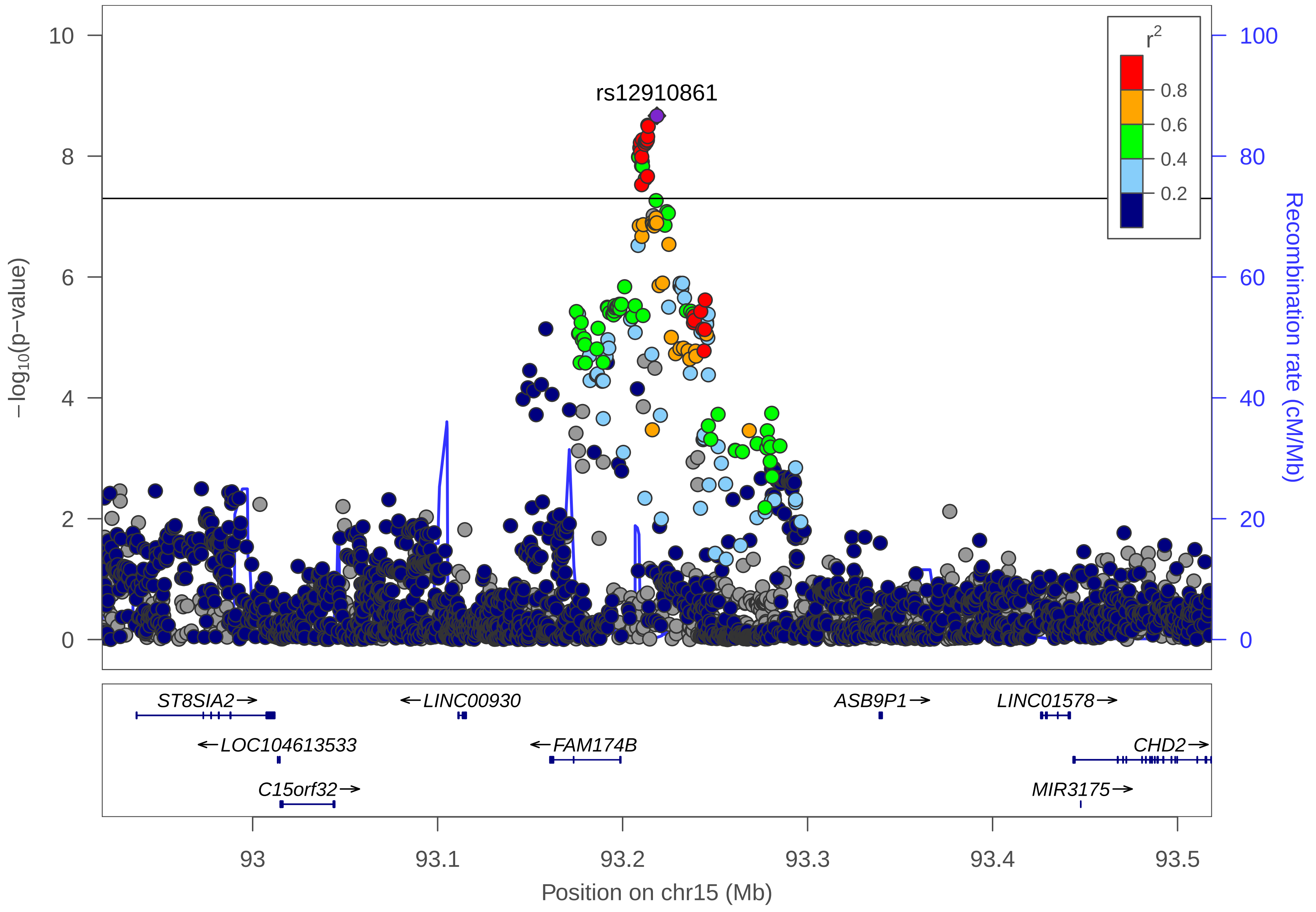

rs12910861
r2
0.8
0.6
0.4
0.2
−log10(p−value)
Recombination rate (cM/Mb)
0
2
4
6
8
10
0
20
40
60
80
100
ST8SIA2→
←LINC00930
ASB9P1→
LINC01578→
←LOC104613533
←FAM174B
CHD2→
C15orf32→
MIR3175→
93
93.1
93.2
93.3
93.4
93.5
Position on chr15 (Mb)

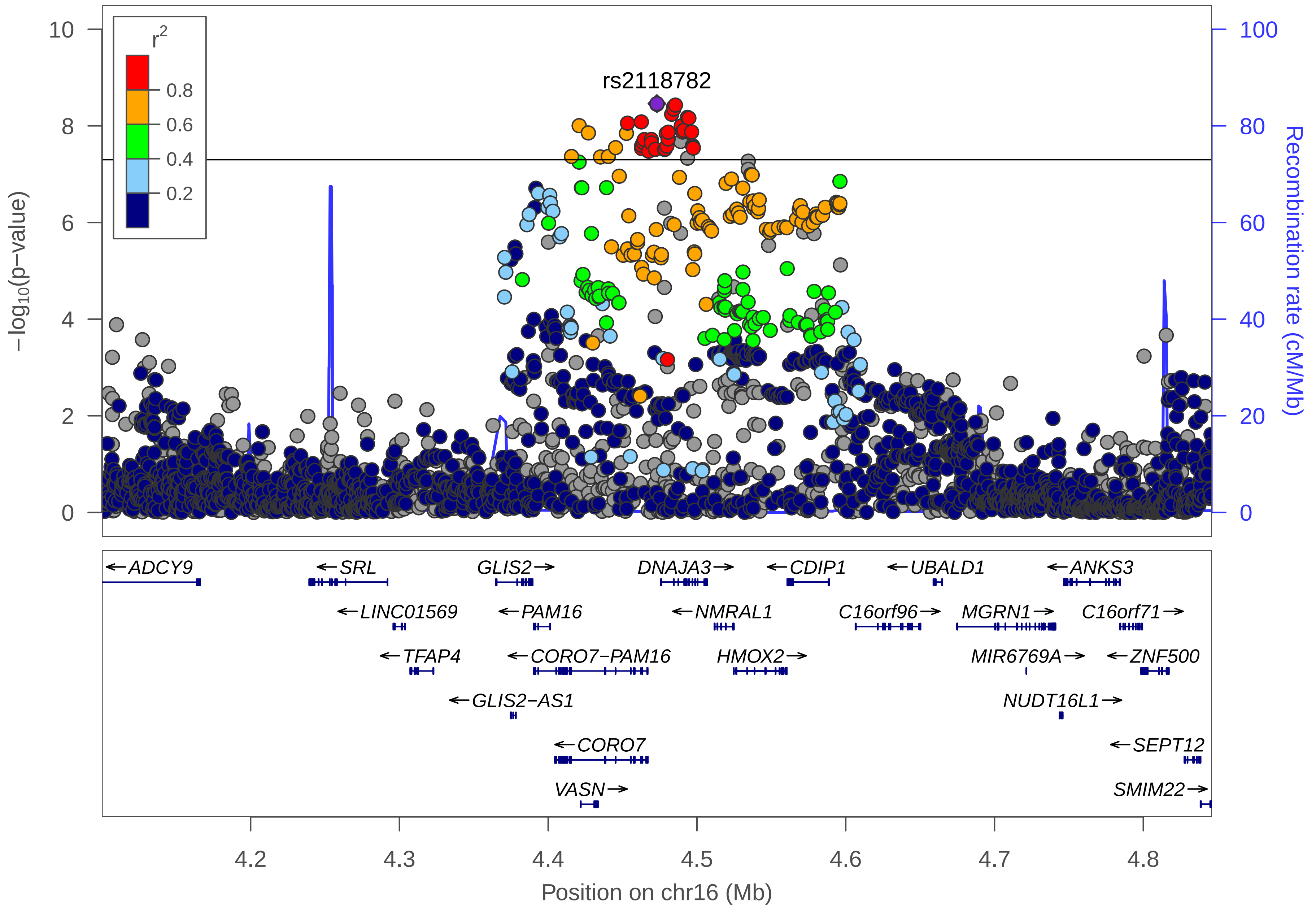

rs2118782
r2
0.8
0.6
0.4
0.2
−log10(p−value)
Recombination rate (cM/Mb)
0
2
4
6
8
10
0
20
40
60
80
100
ADCY9
SRL
GLIS2
DNAJA3
CDIP1
UBALD1
ANKS3
LINC01569
PAM16
NMRAL1
C16orf96
MGRN1
C16orf71
TFAP4
CORO7−PAM16
HMOX2
MIR6769A
ZNF500
GLIS2−AS1
NUDT16L1
CORO7
SEPT12
VASN
SMIM22
4.2
4.3
4.4
4.5
4.6
4.7
4.8
Position on chr16 (Mb)

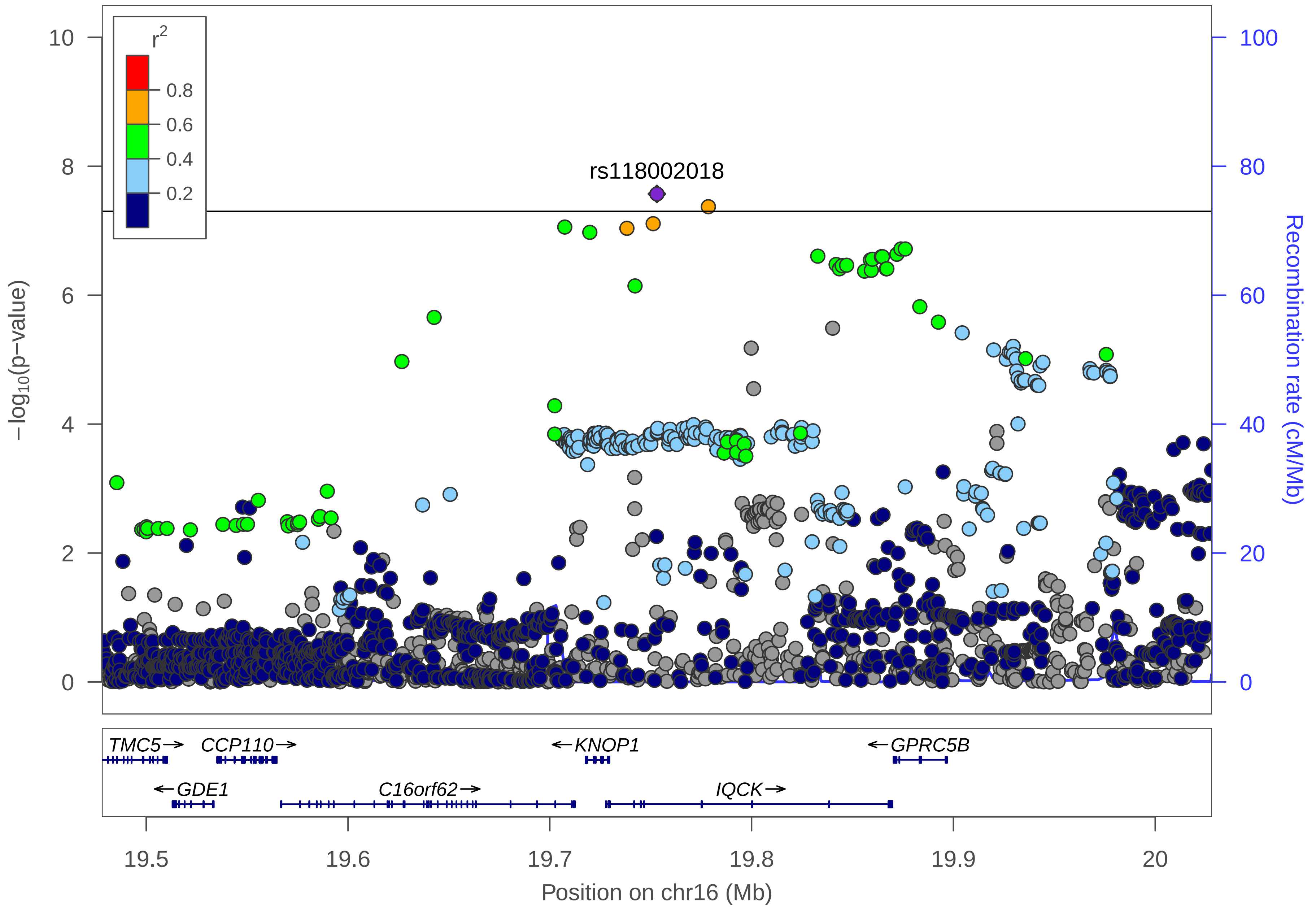

r²
0.8
0.6
0.4
0.2
rs118002018
10
8
6
4
2
0
100
80
60
40
20
0
−log10(p−value)
Recombination rate (cM/Mb)
TMC5→
CCP110→
←KNOP1
←GPRC5B
←GDE1
C16orf62→
IQCK→
19.5
19.6
19.7
19.8
19.9
20
Position on chr16 (Mb)

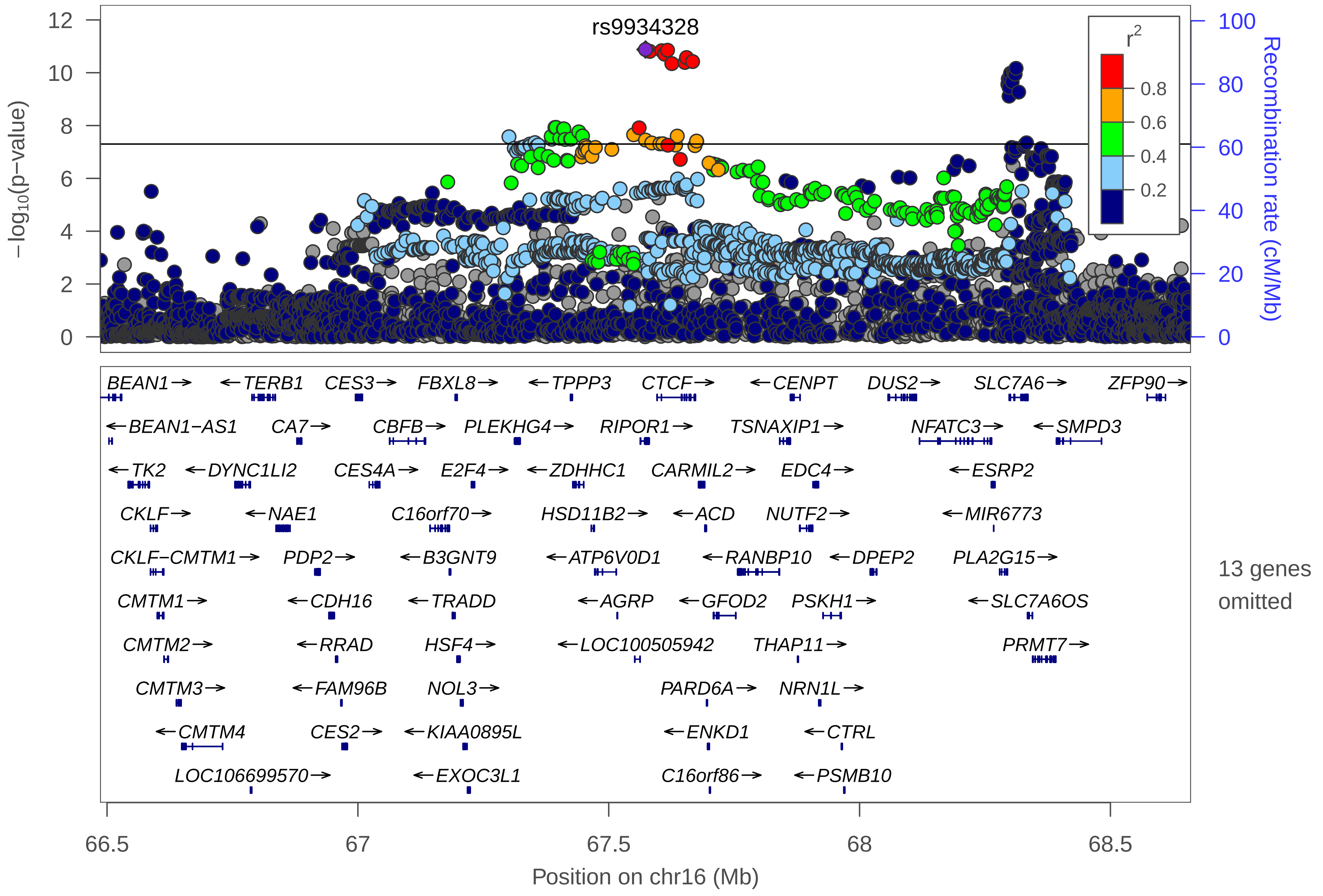
rs9934328
r2
0.8
0.6
0.4
0.2
−log10(p−value)
Recombination rate (cM/Mb)
BEAN1→
←TERB1
CES3→
FBXL8→
←TPPP3
CTCF→
←CENPT
DUS2→
SLC7A6→
ZFP90→
←BEAN1−AS1
CA7→
CBFB→
PLEKHG4→
RIPOR1→
TSNAXIP1→
NFATC3→
←SMPD3
←TK2
←DYNC1LI2
CES4A→
E2F4→
←ZDHHC1
CARMIL2→
EDC4→
←ESRP2
CKLF→
←NAE1
C16orf70→
HSD11B2→
←ACD
NUTF2→
←MIR6773
CKLF−CMTM1→
PDP2→
←B3GNT9
←ATP6V0D1
←RANBP10
←DPEP2
PLA2G15→
CMTM1→
←CDH16
←TRADD
←AGRP
←GFOD2
PSKH1→
←SLC7A6OS
CMTM2→
←RRAD
HSF4→
←LOC100505942
THAP11→
PRMT7→
CMTM3→
←FAM96B
NOL3→
PARD6A→
NRN1L→
←CMTM4
CES2→
←KIAA0895L
←ENKD1
←CTRL
LOC106699570→
←EXOC3L1
C16orf86→
←PSMB10
13 genes omitted
Position on chr16 (Mb)

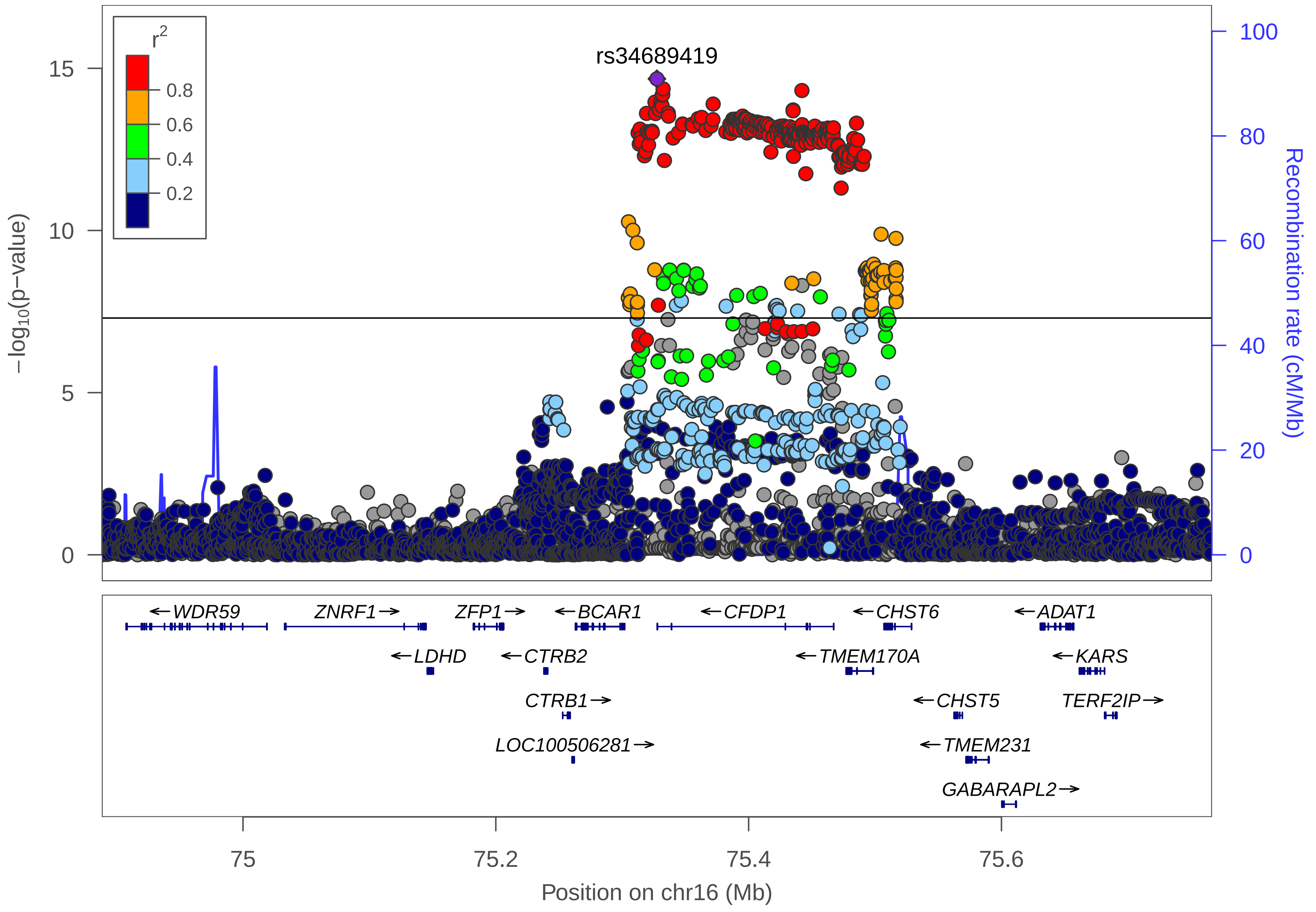

rs34689419
r2
0.8
0.6
0.4
0.2
−log10(p−value)
15
10
5
0
100
80
60
40
20
0
Recombination rate (cM/Mb)
←WDR59
ZNRF1→
ZFP1→
←BCAR1
←CFDP1
←CHST6
←ADAT1
←LDHD
←CTRB2
←TMEM170A
←KARS
CTRB1→
←CHST5
TERF2IP→
LOC100506281→
←TMEM231
GABARAPL2→
75
75.2
75.4
75.6
Position on chr16 (Mb)

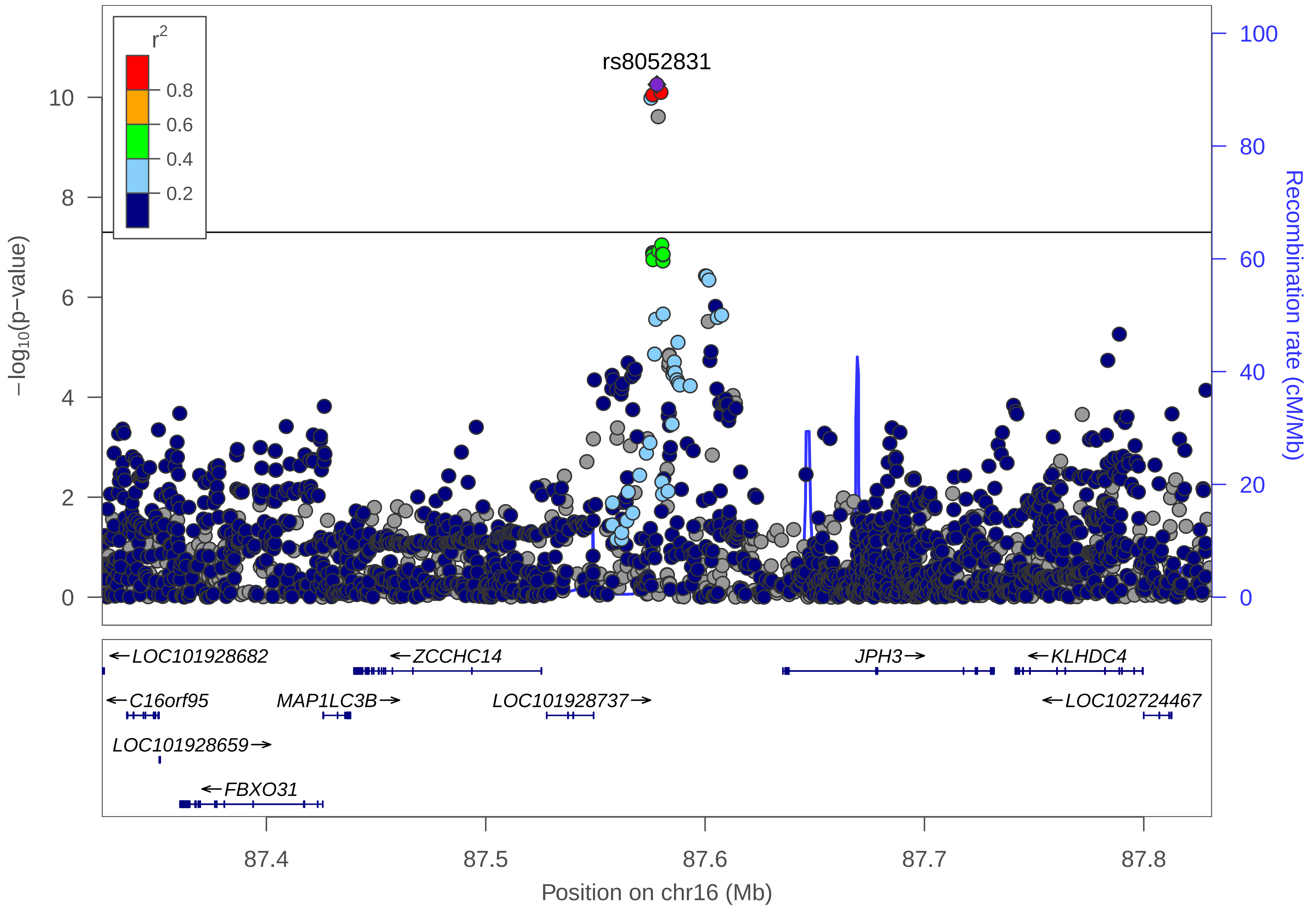

r2
0.8
0.6
0.4
0.2
rs8052831
10
8
6
4
2
0
−log10(p−value)
100
80
60
40
20
0
Recombination rate (cM/Mb)
←LOC101928682
←ZCCHC14
JPH3→
←KLHDC4
←C16orf95
MAP1LC3B→
LOC101928737→
←LOC102724467
LOC101928659→
←FBXO31
87.4
87.5
87.6
87.7
87.8
Position on chr16 (Mb)

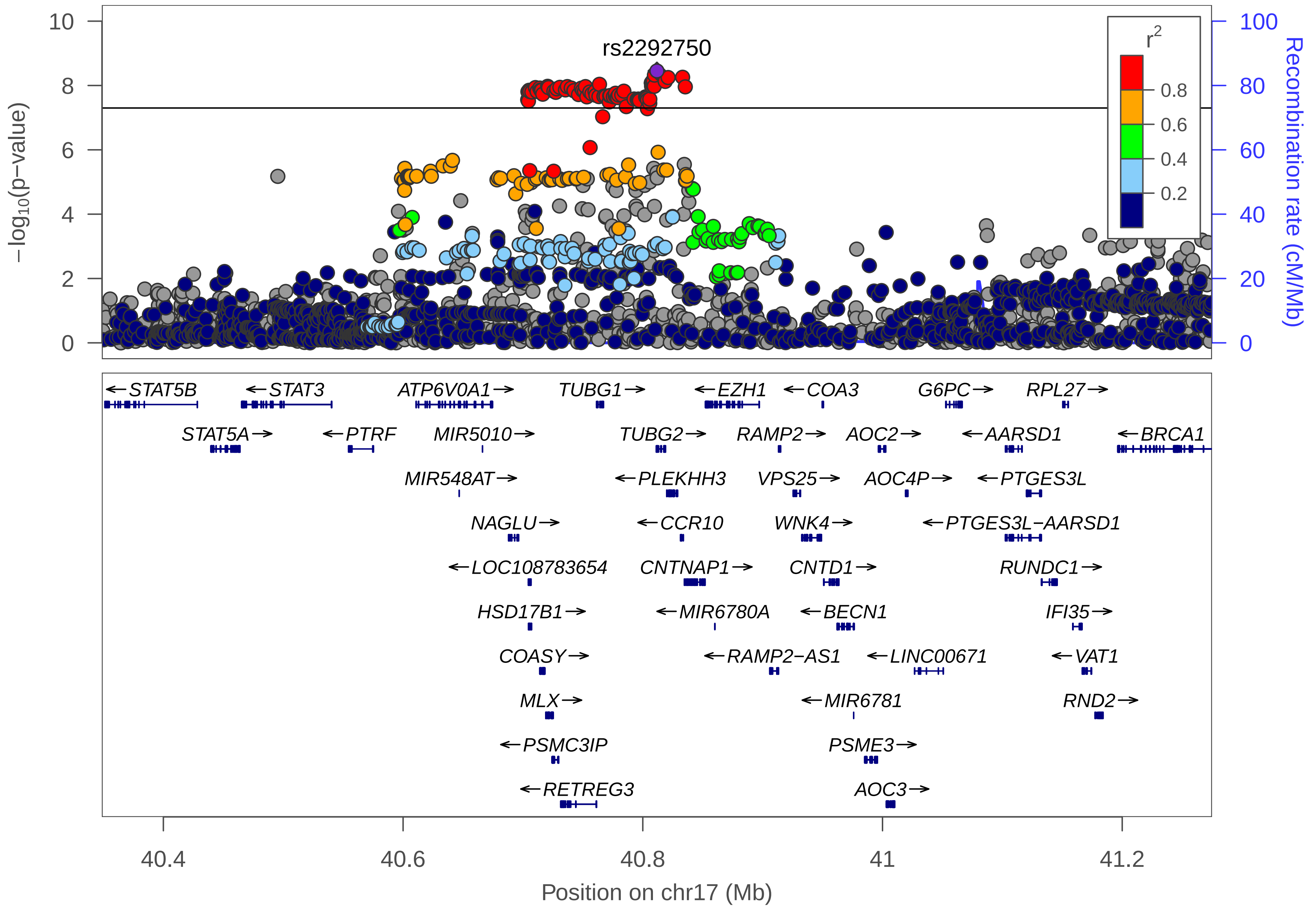

rs2292750
r2
0.8
0.6
0.4
0.2
−log10(p−value)
Recombination rate (cM/Mb)
10
8
6
4
2
0
100
80
60
40
20
0
STAT5B
STAT3
ATP6V0A1
TUBG1
EZH1
COA3
G6PC
RPL27
STAT5A
PTRF
MIR5010
TUBG2
RAMP2
AOC2
AARSD1
BRCA1
MIR548AT
PLEKHH3
VPS25
AOC4P
PTGES3L
NAGLU
CCR10
WNK4
PTGES3L−AARSD1
LOC108783654
CNTNAP1
CNTD1
RUNDC1
HSD17B1
MIR6780A
BECN1
IFI35
COASY
RAMP2−AS1
LINC00671
VAT1
MLX
MIR6781
RND2
PSMC3IP
PSME3
RETREG3
AOC3
40.4
40.6
40.8
41
41.2
Position on chr17 (Mb)

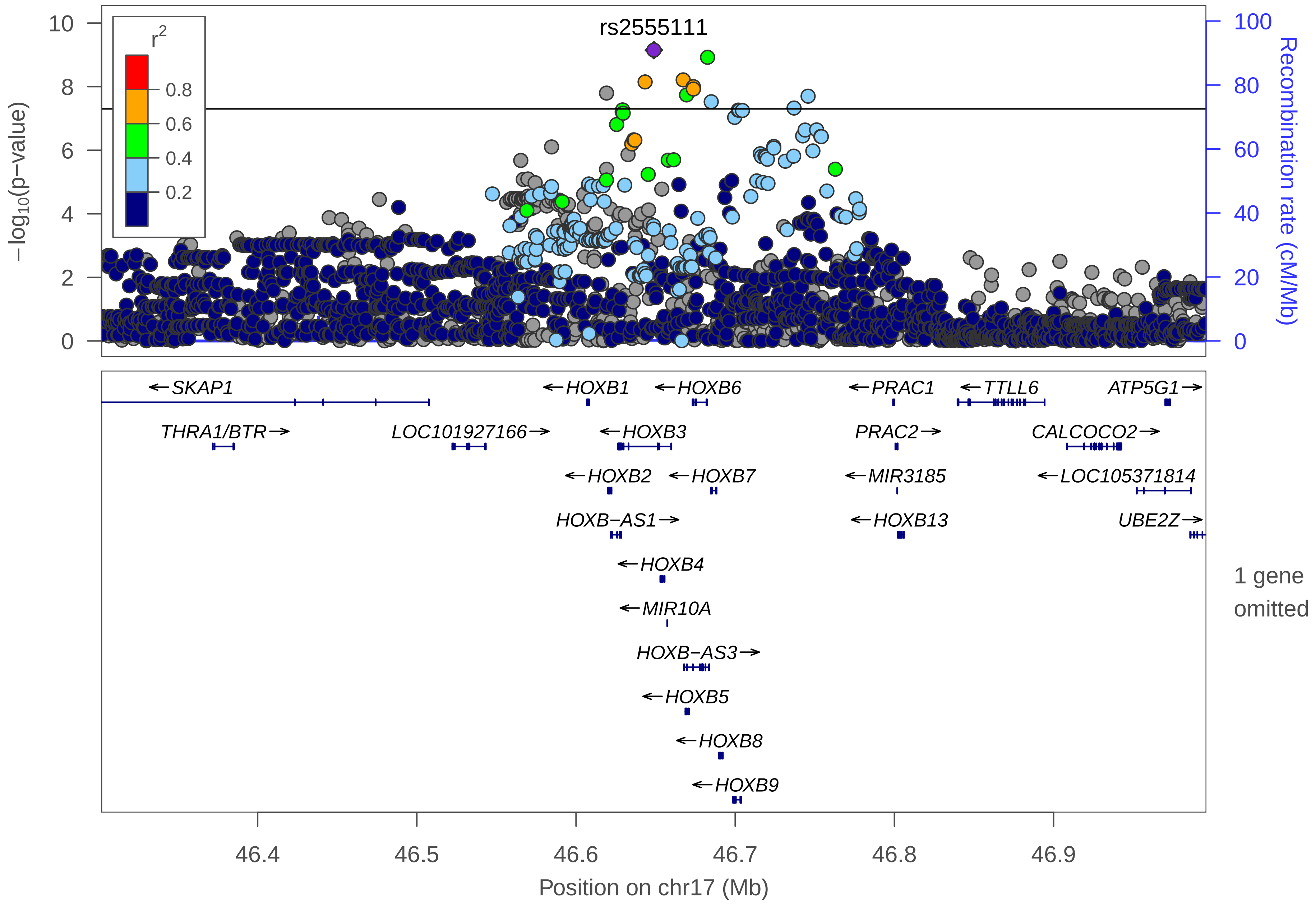

rs2555111
r2
0.8
0.6
0.4
0.2
10
8
6
4
2
0
−log10(p−value)
100
80
60
40
20
0
Recombination rate (cM/Mb)
SKAP1
HOXB1
HOXB6
PRAC1
TTLL6
ATP5G1
THRA1/BTR
LOC101927166
HOXB3
PRAC2
CALCOCO2
HOXB2
HOXB7
MIR3185
LOC105371814
HOXB−AS1
HOXB13
UBE2Z
HOXB4
1 gene omitted
MIR10A
HOXB−AS3
HOXB5
HOXB8
HOXB9
46.4
46.5
46.6
46.7
46.8
46.9
Position on chr17 (Mb)

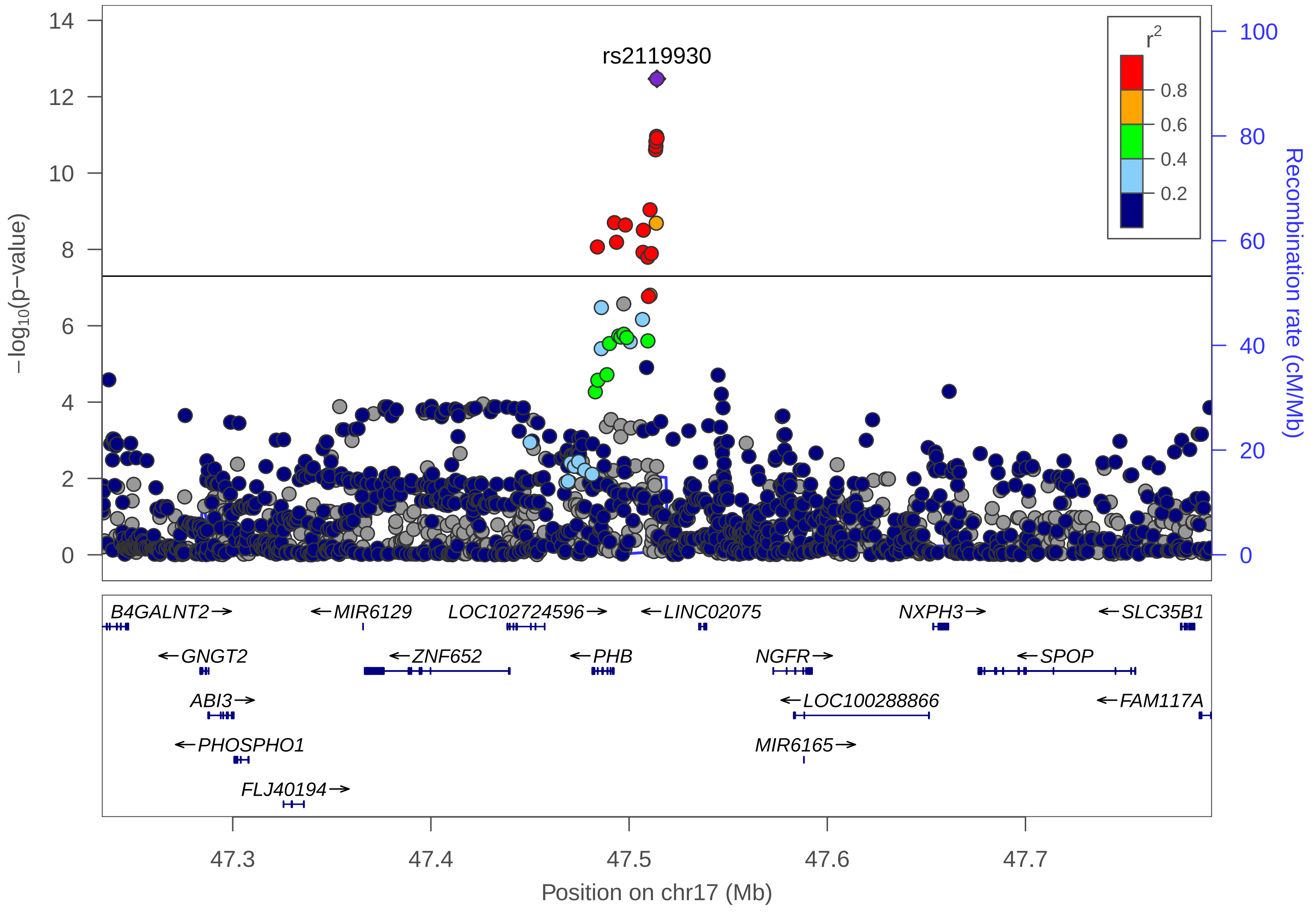
rs2119930
r2
0.8
0.6
0.4
0.2
−log10(p−value)
Recombination rate (cM/Mb)
B4GALNT2→
←MIR6129
LOC102724596→
←LINC02075
NXPH3→
←SLC35B1
←GNGT2
←ZNF652
←PHB
NGFR→
←SPOP
ABI3→
←LOC100288866
←FAM117A
←PHOSPHO1
MIR6165→
FLJ40194→
47.3
47.4
47.5
47.6
47.7
Position on chr17 (Mb)

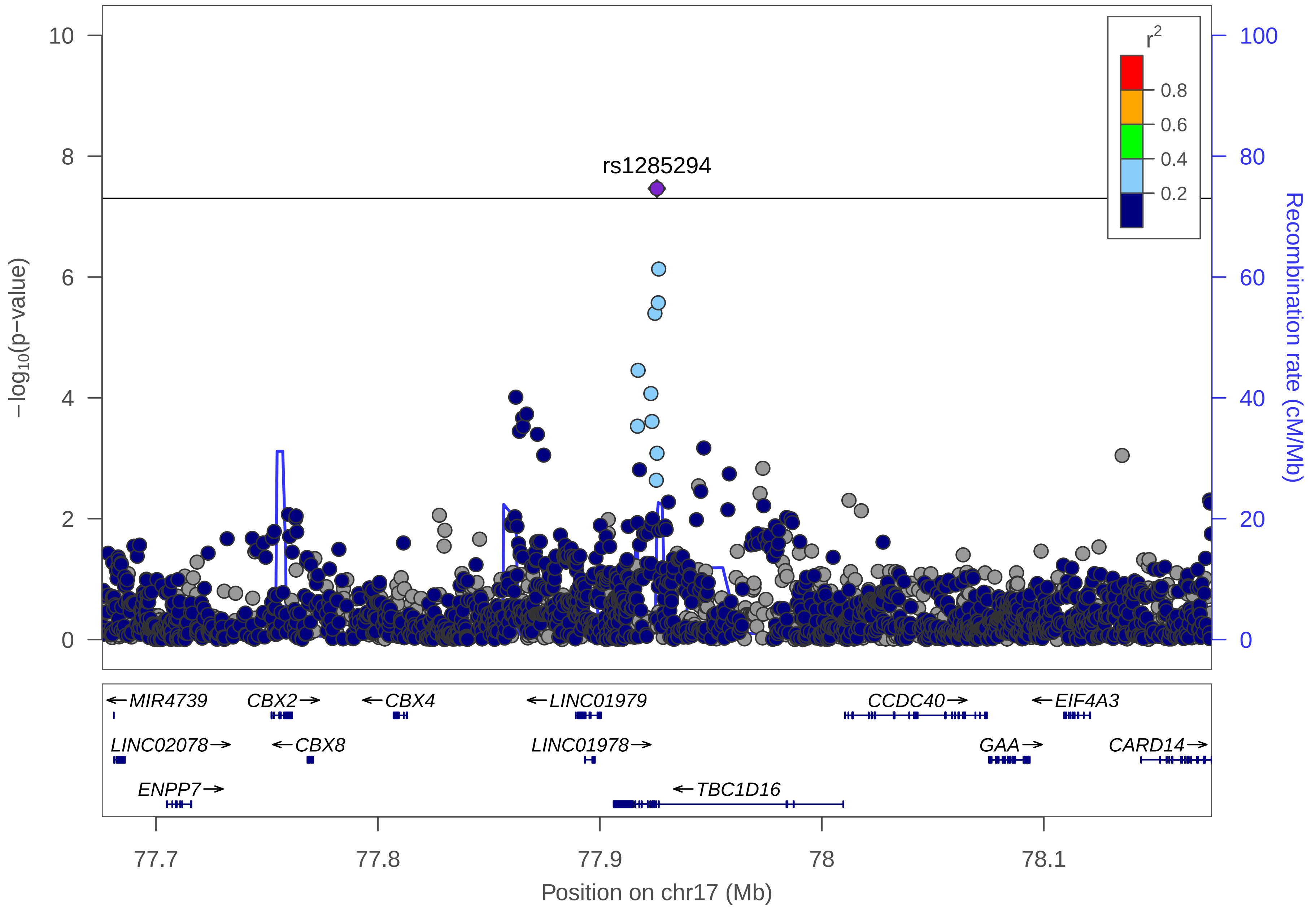

rs1285294
−log10(p−value)
Recombination rate (cM/Mb)
r2
0.8
0.6
0.4
0.2
0
2
4
6
8
10
0
20
40
60
80
100
MIR4739
CBX2
CBX4
LINC01979
CCDC40
EIF4A3
LINC02078
CBX8
LINC01978
GAA
CARD14
ENPP7
TBC1D16
77.7
77.8
77.9
78
78.1
Position on chr17 (Mb)

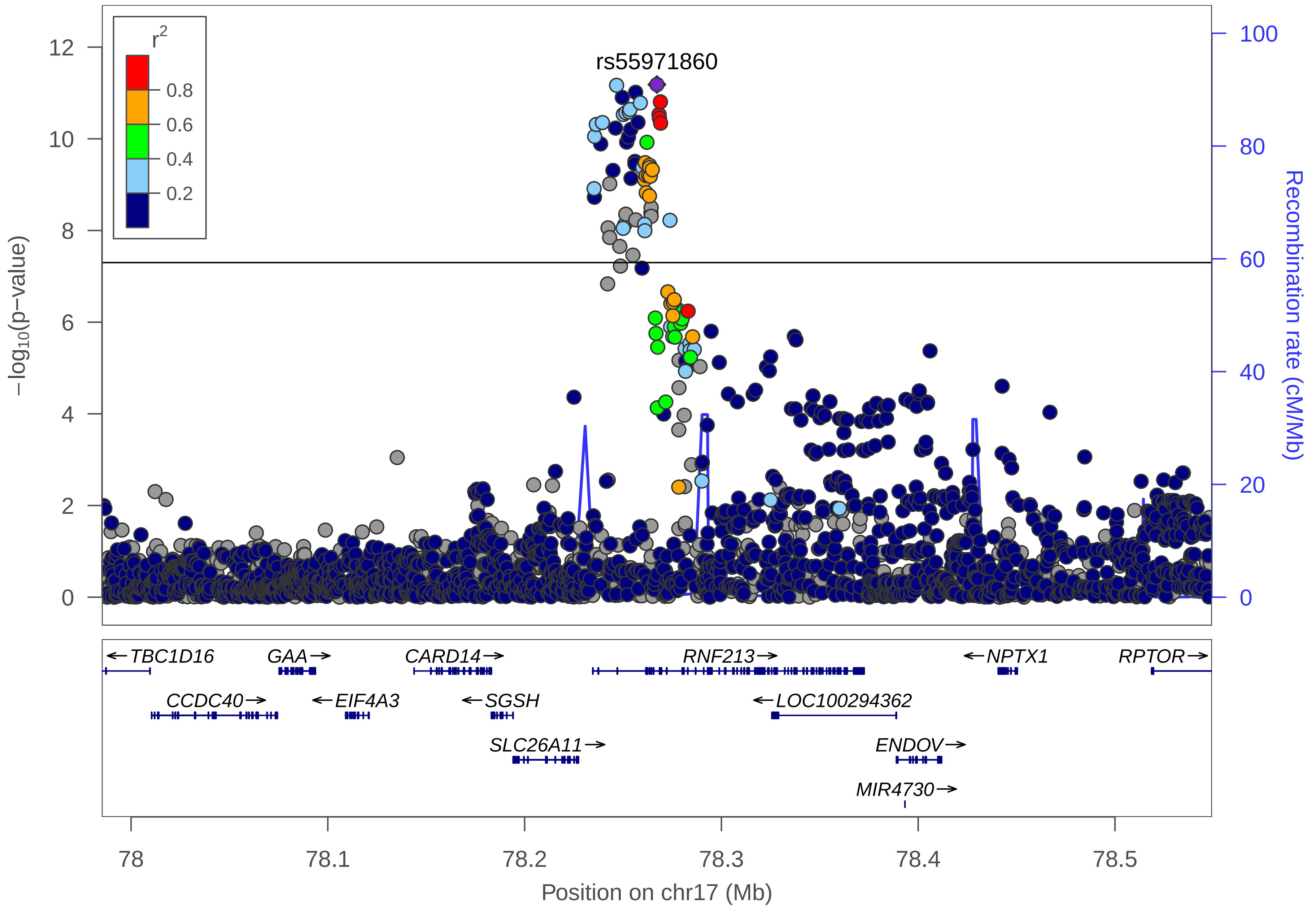

r2
0.8
0.6
0.4
0.2
rs55971860
12
10
8
6
4
2
0
−log10(p−value)
100
80
60
40
20
0
Recombination rate (cM/Mb)
←TBC1D16
GAA→
CARD14→
RNF213→
←NPTX1
RPTOR→
CCDC40→
←EIF4A3
←SGSH
←LOC100294362
SLC26A11→
ENDOV→
MIR4730→
78
78.1
78.2
78.3
78.4
78.5
Position on chr17 (Mb)

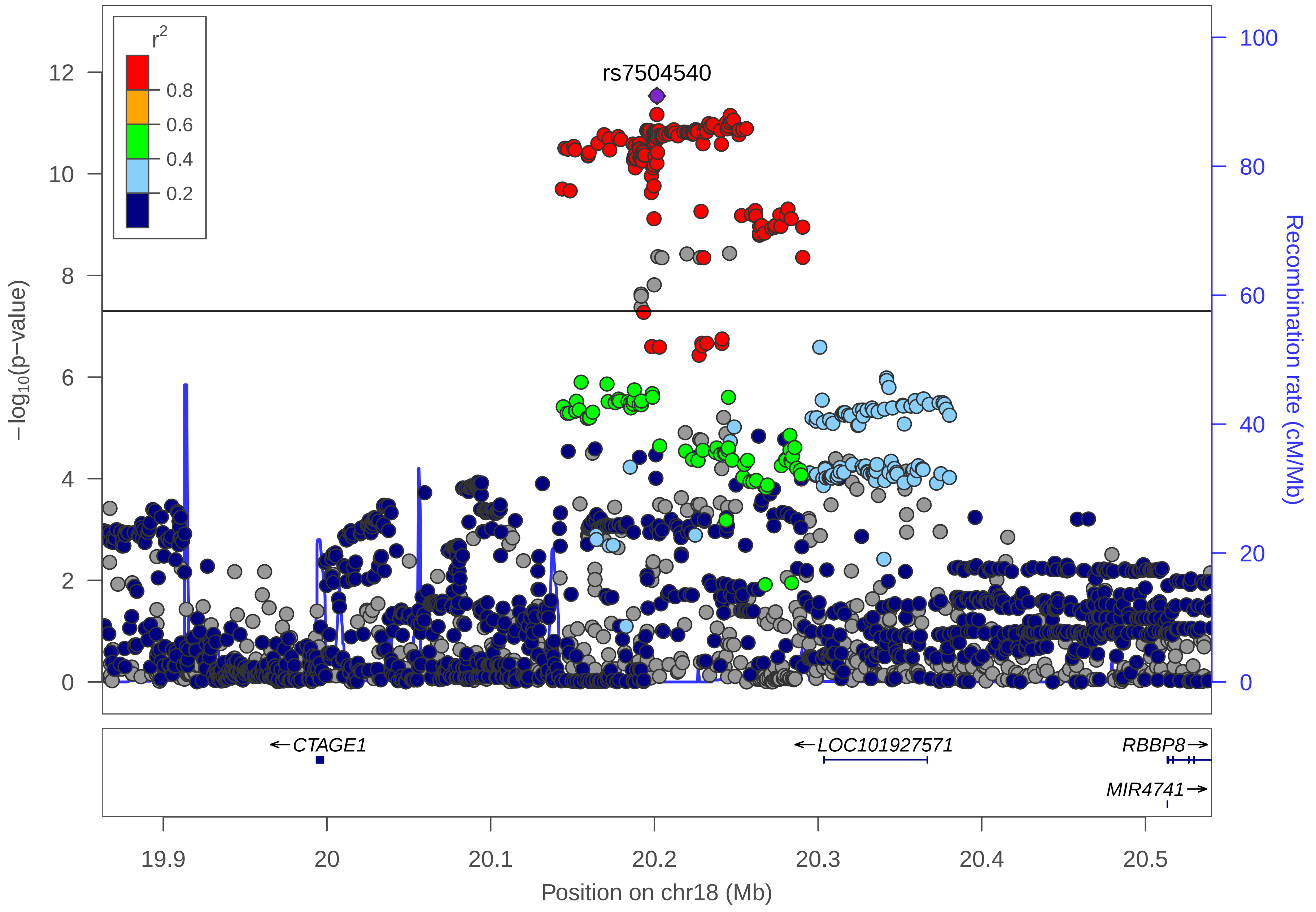

r2
0.8
0.6
0.4
0.2
rs7504540
12
10
8
6
4
2
0
−log10(p−value)
100
80
60
40
20
0
Recombination rate (cM/Mb)
←CTAGE1
←LOC101927571
RBBP8→
MIR4741→
19.9
20
20.1
20.2
20.3
20.4
20.5
Position on chr18 (Mb)

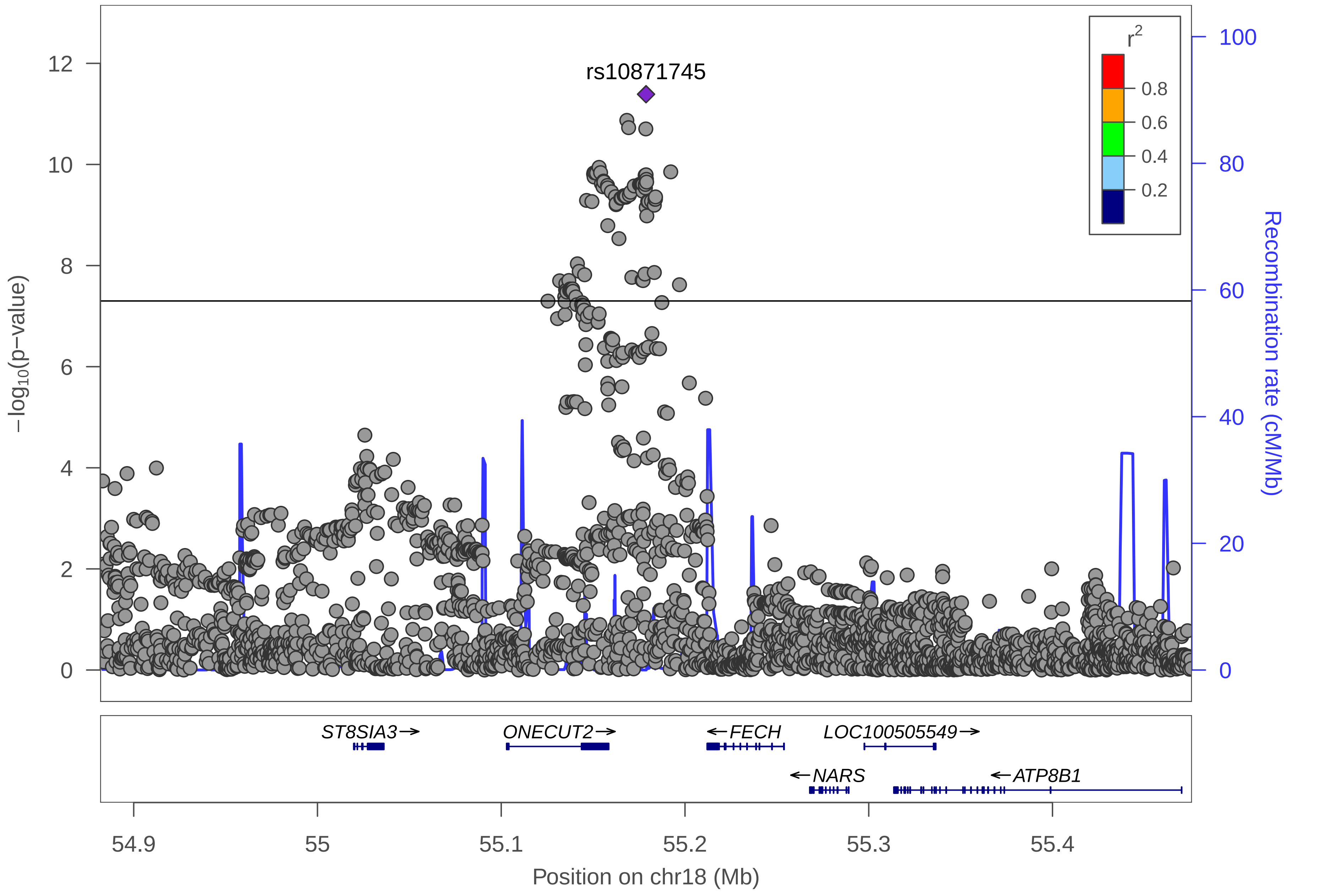

rs10871745
r2
0.8
0.6
0.4
0.2
−log10(p−value)
Recombination rate (cM/Mb)
0
2
4
6
8
10
12
0
20
40
60
80
100
ST8SIA3→
ONCUT2→
←FECH
LOC100505549→
←NARS
←ATP8B1
54.9
55
55.1
55.2
55.3
55.4
Position on chr18 (Mb)

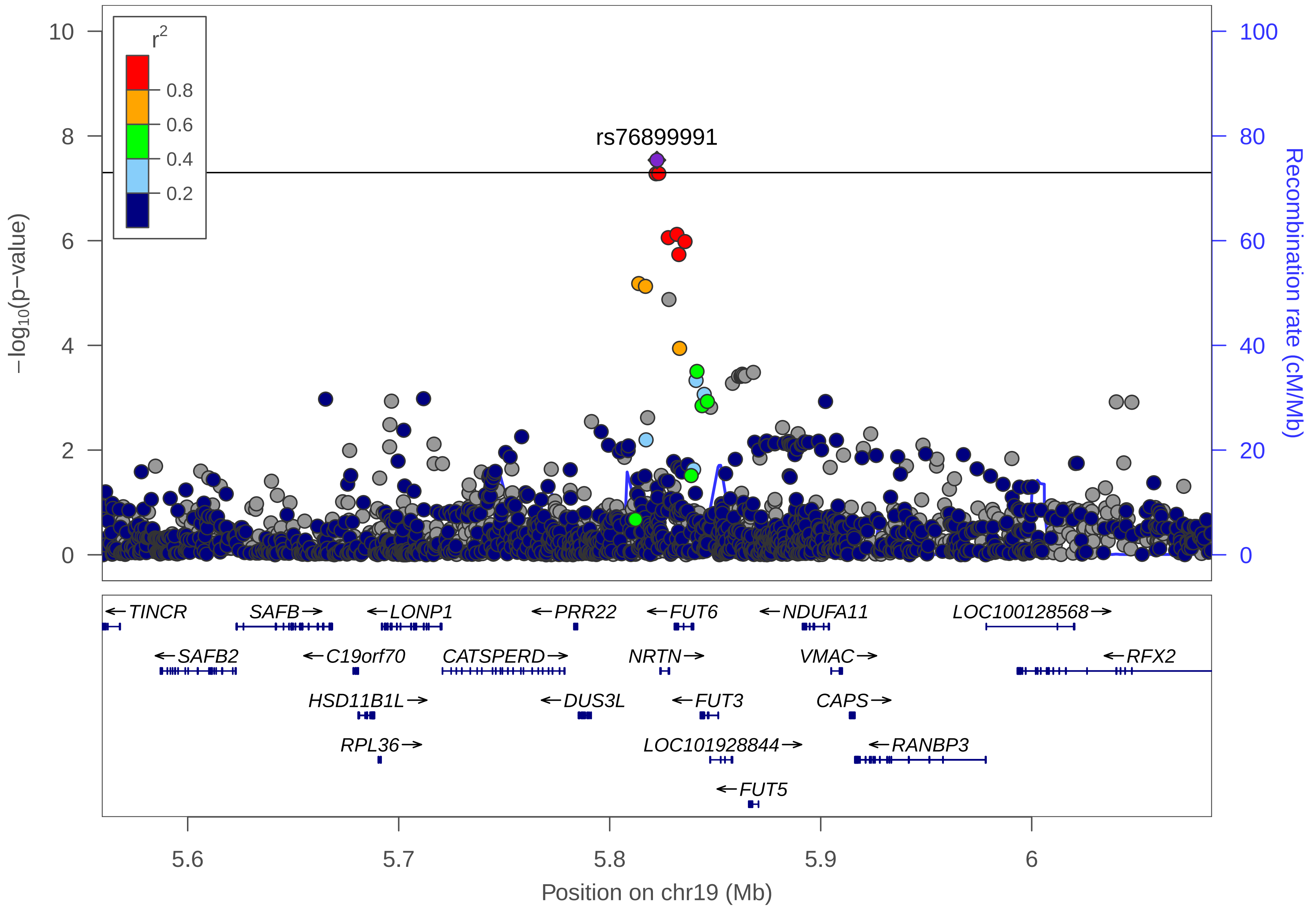

r2
0.8
0.6
0.4
0.2
rs76899991
10
8
6
4
2
0
−log10(p−value)
100
80
60
40
20
0
Recombination rate (cM/Mb)
←TINCR
SAFB→
←LONP1
←PRR22
←FUT6
←NDUFA11
LOC100128568→
←SAFB2
←C19orf70
CATSPERD→
NRTN→
VMAC→
←RFX2
HSD11B1L→
←DUS3L
←FUT3
CAPS→
RPL36→
LOC101928844→
←RANBP3
←FUT5
5.6
5.7
5.8
5.9
6
Position on chr19 (Mb)

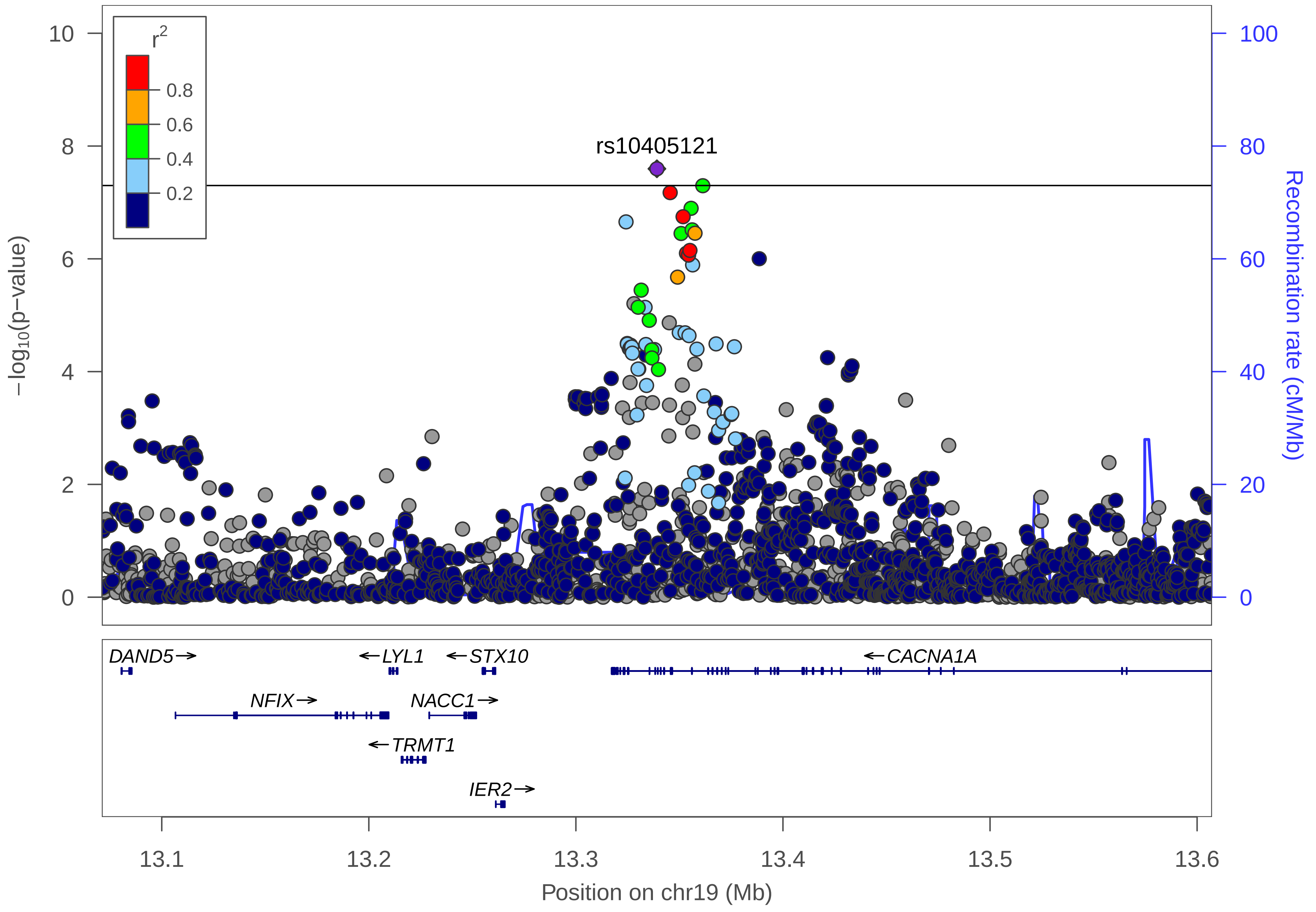

rs10405121
r2
0.8
0.6
0.4
0.2
−log10(p−value)
0
2
4
6
8
10
Recombination rate (cM/Mb)
0
20
40
60
80
100
DAND5
LYL1
STX10
CACNA1A
NFIX
NACC1
TRMT1
IER2
13.1
13.2
13.3
13.4
13.5
13.6
Position on chr19 (Mb)

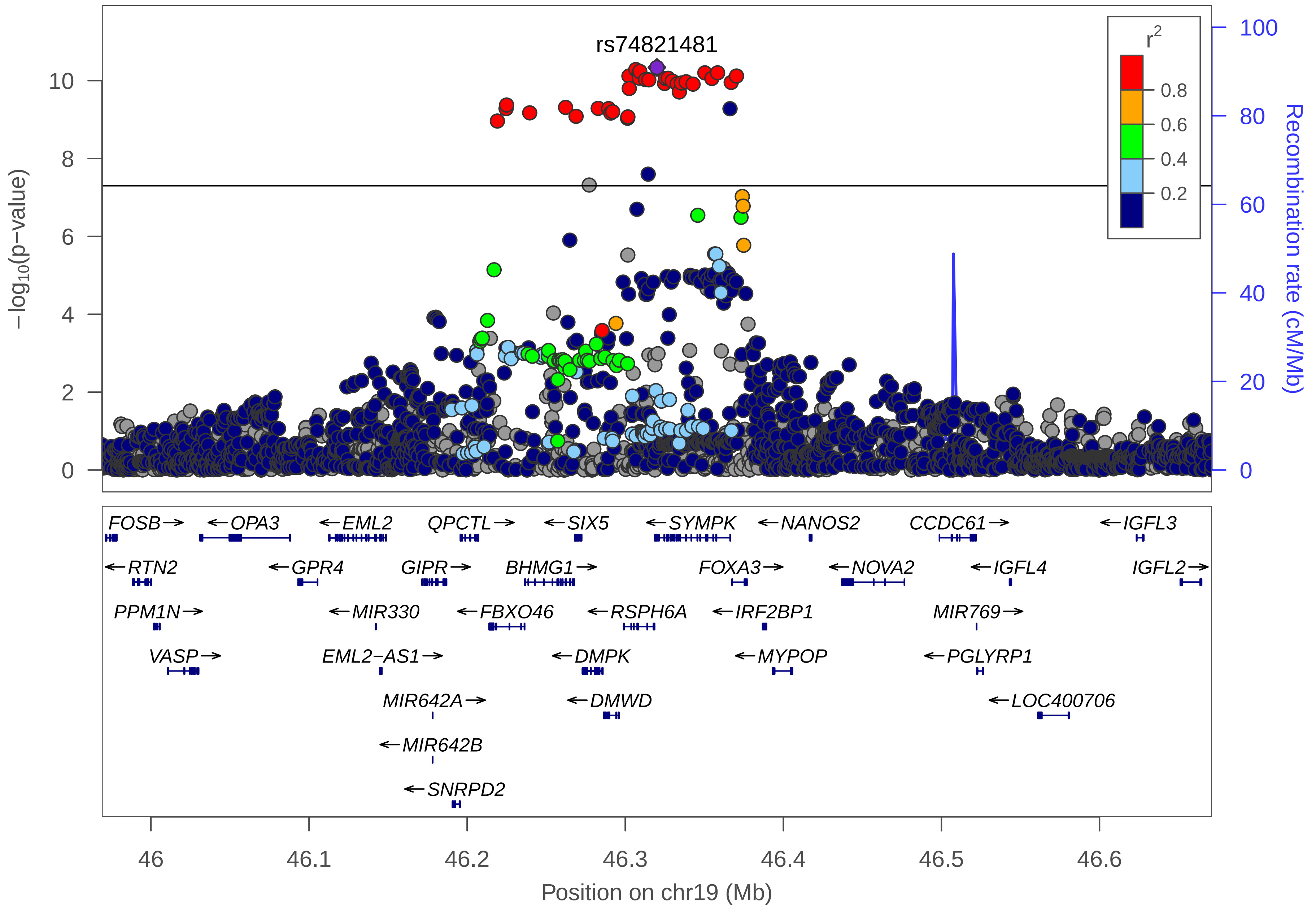

rs74821481
r²
0.8
0.6
0.4
0.2
−log10(p−value)
Recombination rate (cM/Mb)
0
2
4
6
8
10
0
20
40
60
80
100
FOSB→
←OPA3
←EML2
QPCTL→
←SIX5
←SYMPK
←NANOS2
CCDC61→
←IGFL3
←RTN2
←GPR4
GIPR→
BHMG1→
FOXA3→
←NOVA2
←IGFL4
IGFL2→
PPM1N→
←MIR330
←FBXO46
←RSPH6A
←IRF2BP1
MIR769→
VASP→
EML2−AS1→
←DMPK
←MYPOP
←PGLYRP1
MIR642A→
←DMWD
←LOC400706
←MIR642B
←SNRPD2
46
46.1
46.2
46.3
46.4
46.5
46.6
Position on chr19 (Mb)

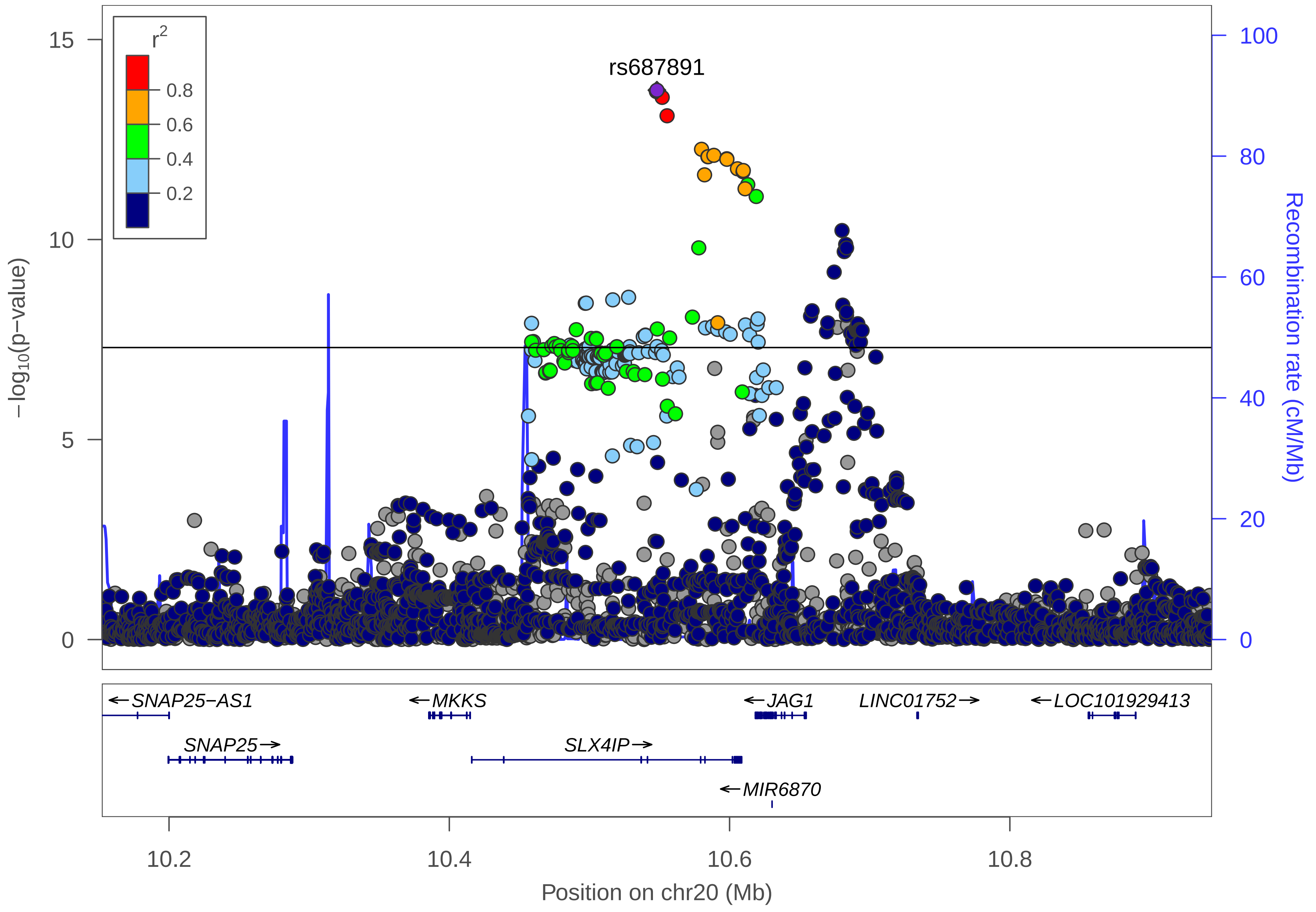

r2
0.8
0.6
0.4
0.2
rs687891
15
10
5
0
−log10(p−value)
100
80
60
40
20
0
Recombination rate (cM/Mb)
←SNAP25−AS1
←MKKS
←JAG1
LINC01752→
←LOC101929413
SNAP25→
SLX4IP→
←MIR6870
10.2
10.4
10.6
10.8
Position on chr20 (Mb)

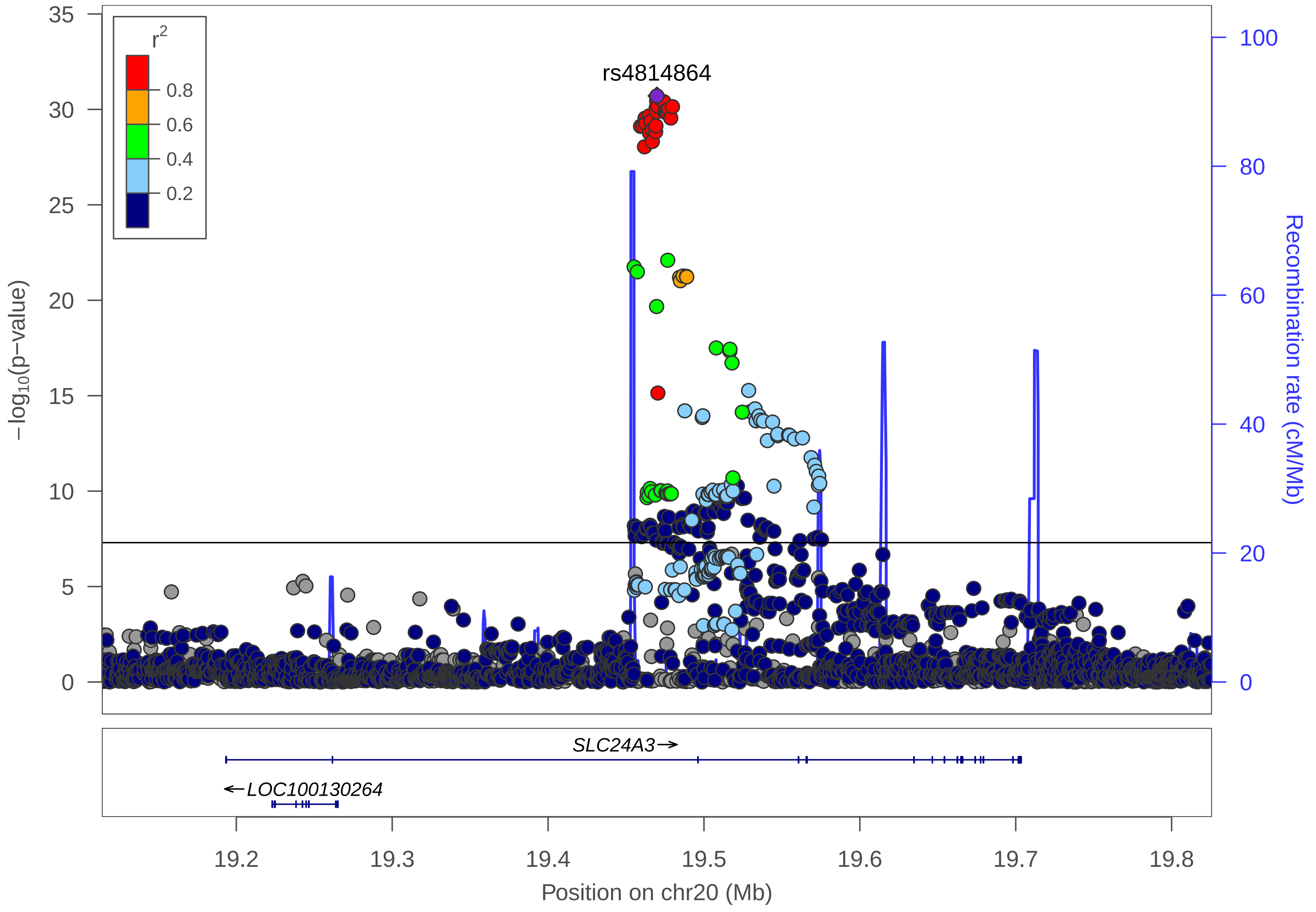

r2
0.8
0.6
0.4
0.2
rs4814864
35
30
25
20
15
10
5
0
−log10(p−value)
100
80
60
40
20
0
Recombination rate (cM/Mb)
SLC24A3→
←LOC100130264
19.2
19.3
19.4
19.5
19.6
19.7
19.8
Position on chr20 (Mb)

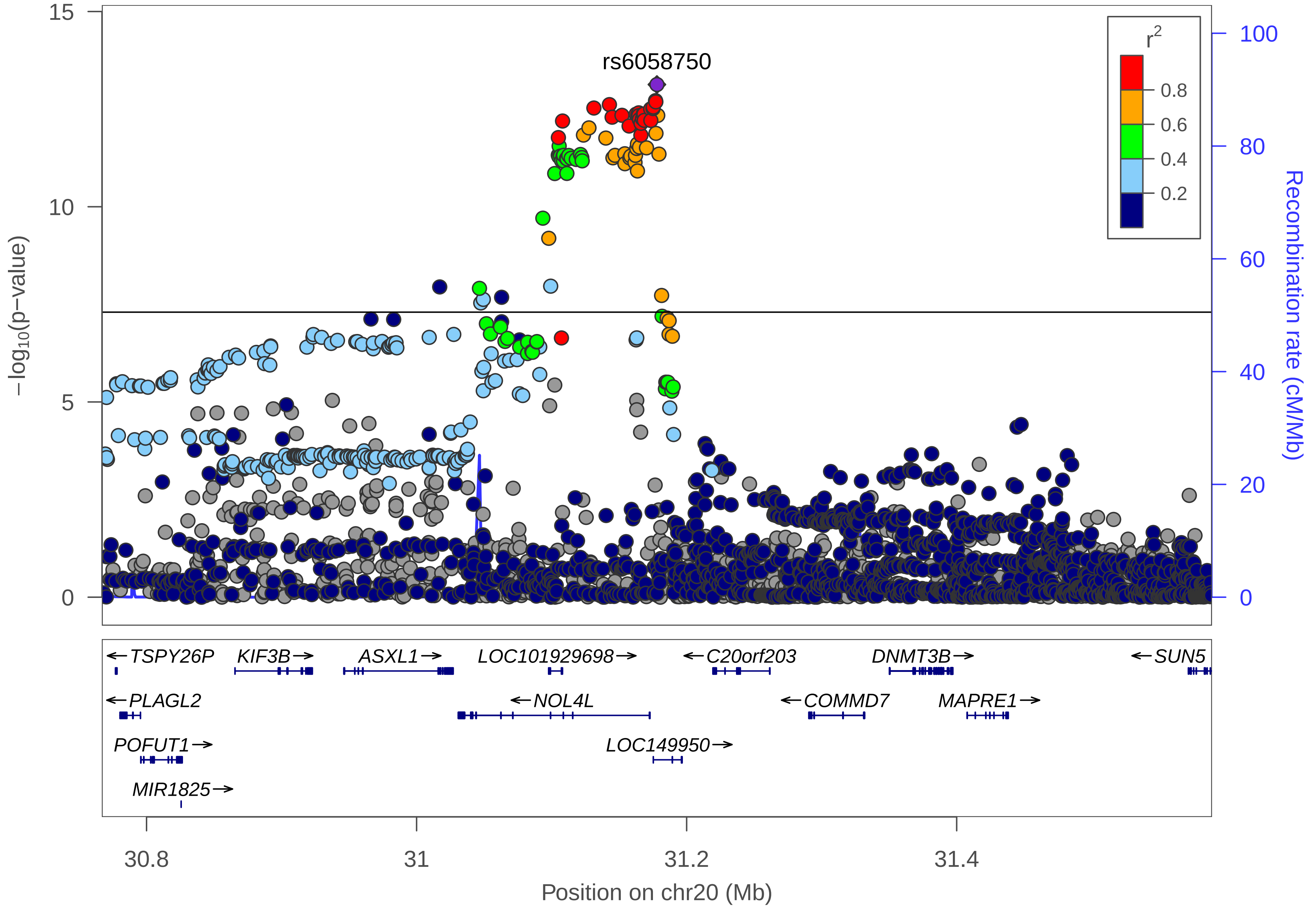

rs6058750
r²
0.8
0.6
0.4
0.2
15
10
5
0
−log10(p−value)
100
80
60
40
20
0
Recombination rate (cM/Mb)
←TSPY26P
KIF3B→
ASXL1→
LOC101929698→
←C20orf203
DNMT3B→
←SUN5
←PLAGL2
←NOL4L
←COMMD7
MAPRE1→
POFUT1→
LOC149950→
MIR1825→
30.8
31
31.2
31.4
Position on chr20 (Mb)

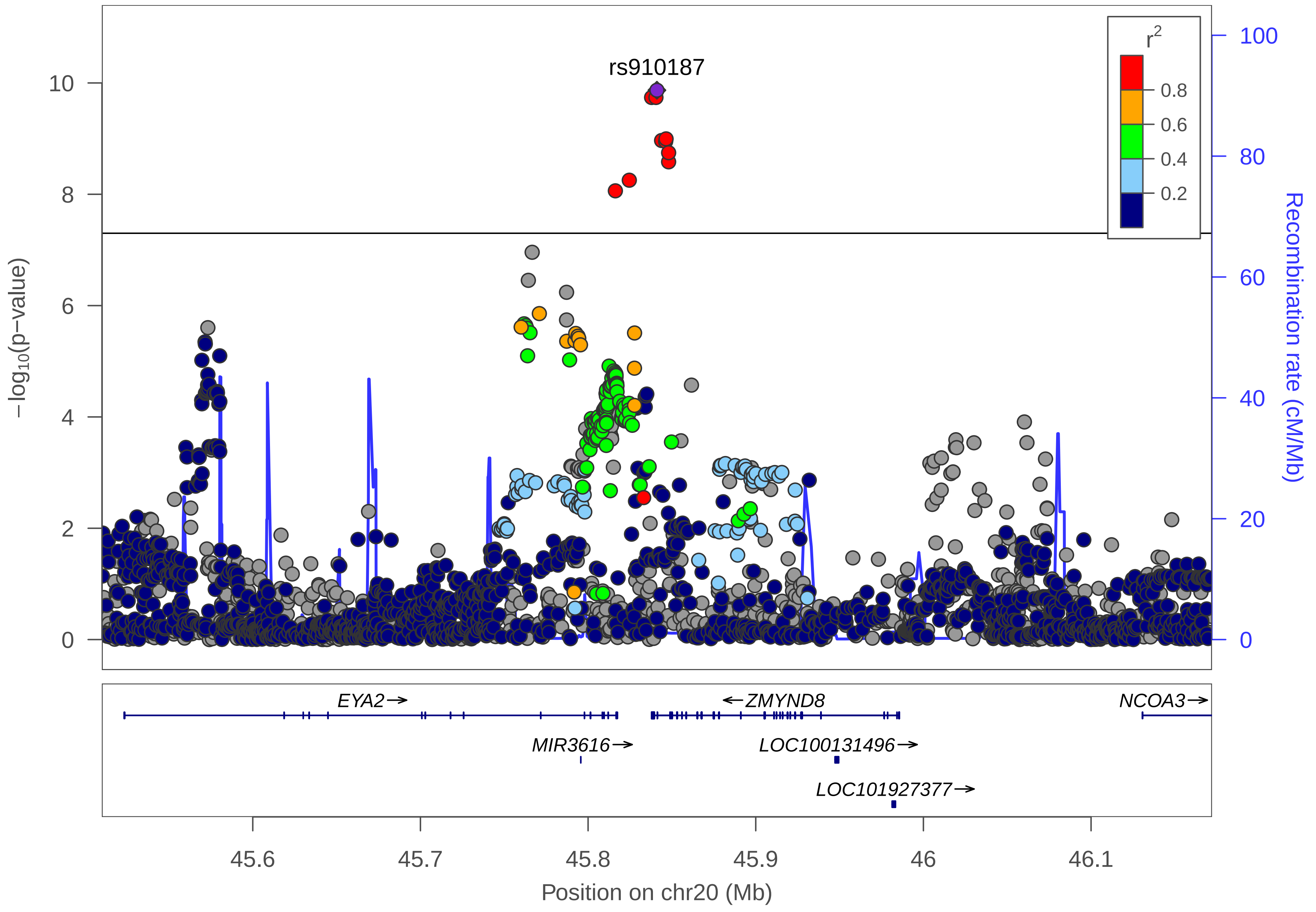

rs910187
r^2
0.8
0.6
0.4
0.2
−log10(p−value)
Recombination rate (cM/Mb)
10
8
6
4
2
0
100
80
60
40
20
0
EYA2→
←ZMYND8
NCOA3→
MIR3616→
LOC100131496→
LOC101927377→
45.6
45.7
45.8
45.9
46
46.1
Position on chr20 (Mb)

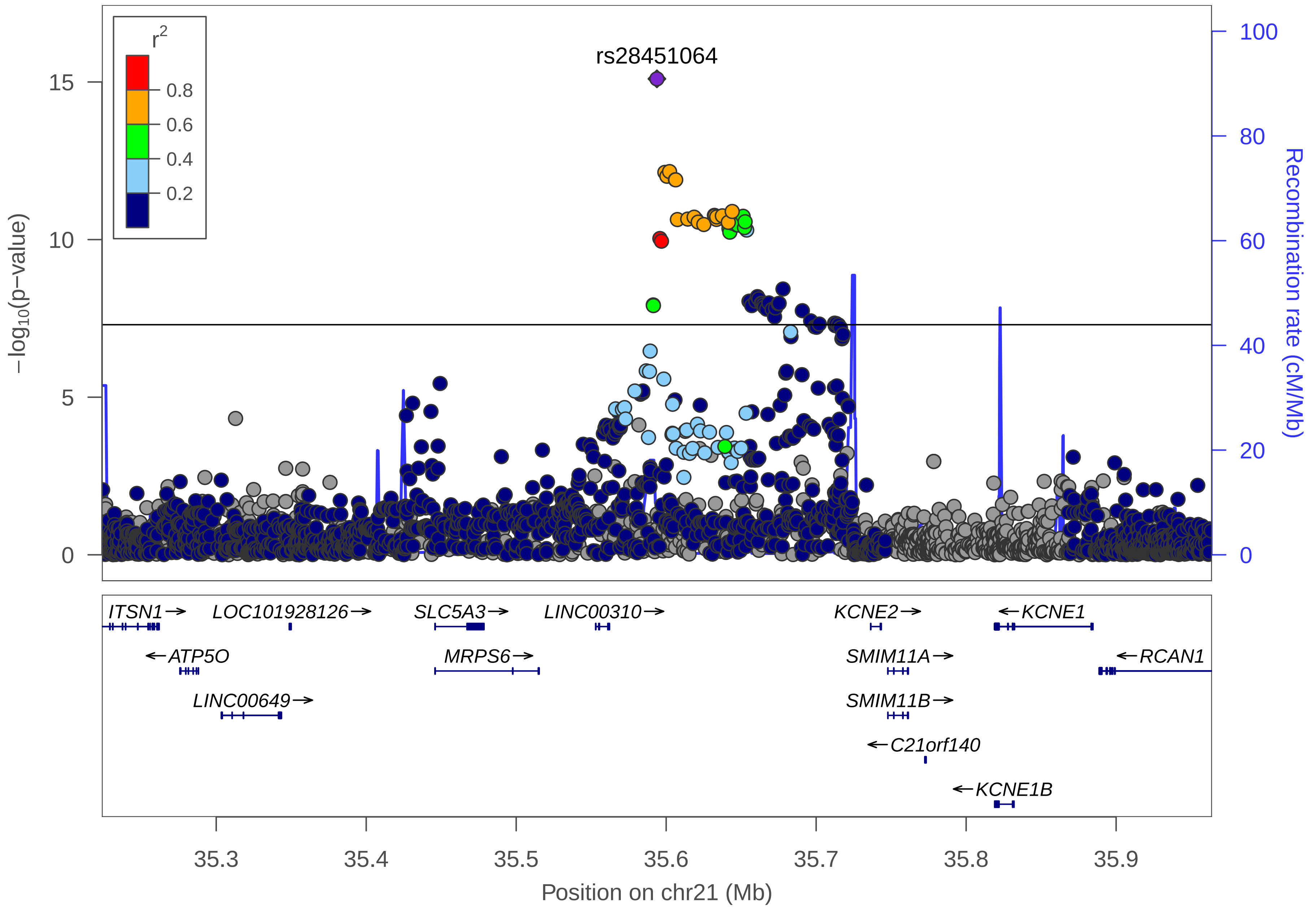
rs28451064
r2
0.8
0.6
0.4
0.2
−log10(p−value)
15
10
5
0
100
80
60
40
20
0
Recombination rate (cM/Mb)
ITSN1→
LOC101928126→
SLC5A3→
LINC00310→
KCNE2→
←KCNE1
←ATP5O
MRPS6→
SMIM11A→
←RCAN1
LINC00649→
SMIM11B→
←C21orf140
←KCNE1B
35.3
35.4
35.5
35.6
35.7
35.8
35.9
Position on chr21 (Mb)

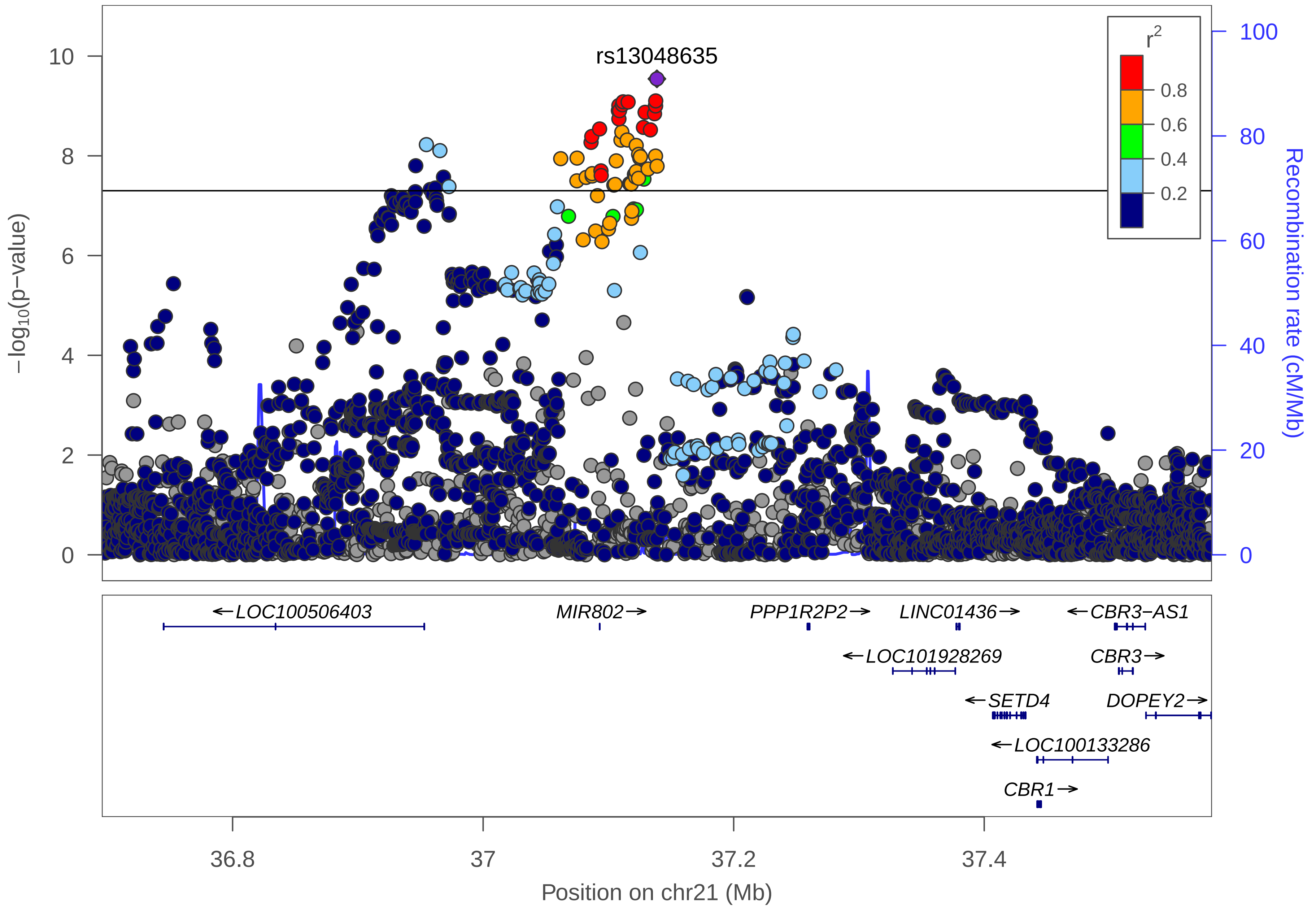

rs13048635
r2
0.8
0.6
0.4
0.2
−log10(p−value)
Recombination rate (cM/Mb)
10
8
6
4
2
0
100
80
60
40
20
0
←LOC100506403
MIR802→
PPP1R2P2→
LINC01436→
←CBR3−AS1
←LOC101928269
CBR3→
←SETD4
DOPEY2→
←LOC100133286
CBR1→
36.8
37
37.2
37.4
Position on chr21 (Mb)

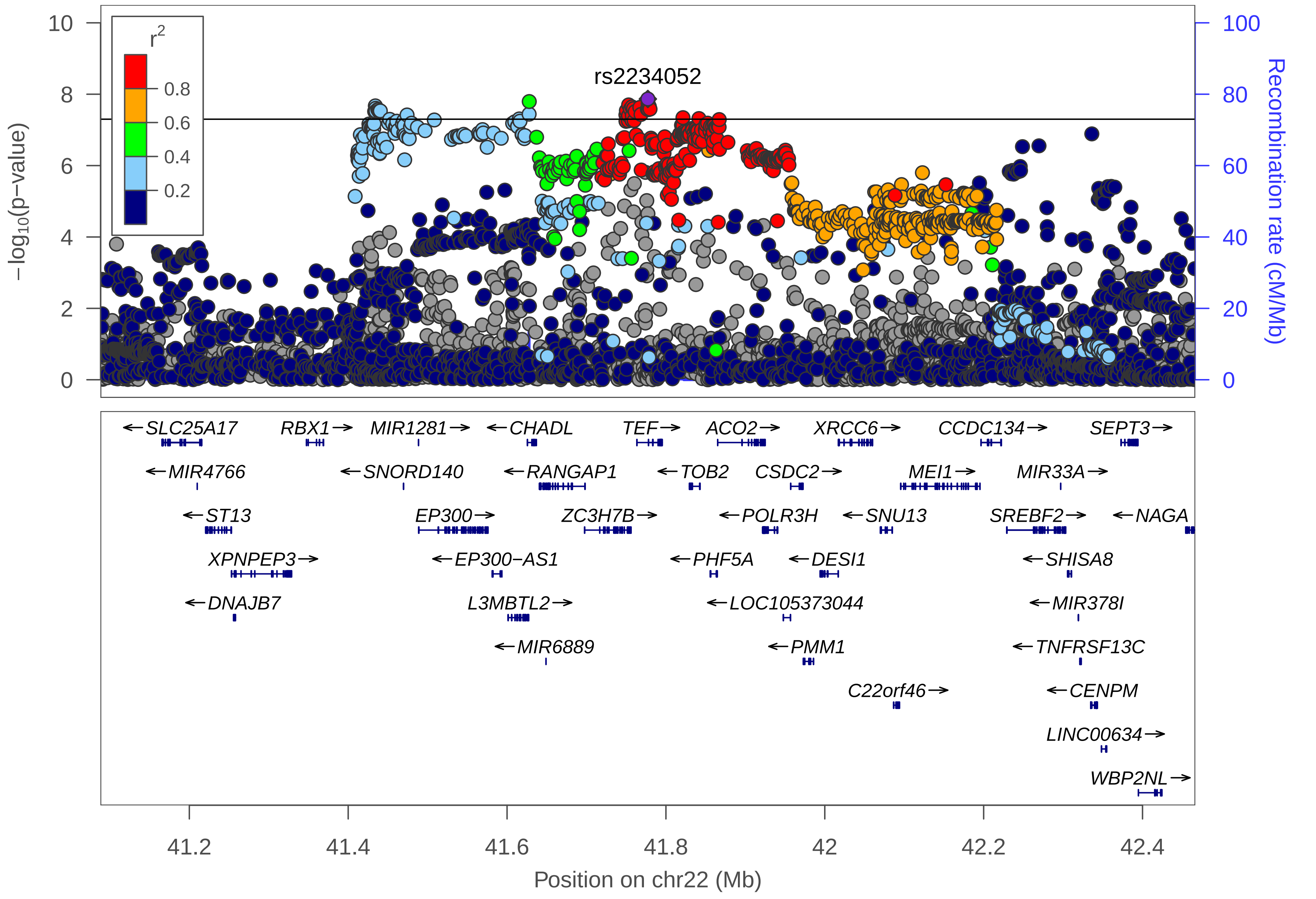

rs2234052
r2
0.8
0.6
0.4
0.2
−log10(p−value)
Recombination rate (cM/Mb)
10
8
6
4
2
0
100
80
60
40
20
0
SLC25A17
RBX1
MIR1281
CHADL
TEF
ACO2
XRCC6
CCDC134
SEPT3
MIR4766
SNORD140
RANGAP1
TOB2
CSDC2
MEI1
MIR33A
ST13
EP300
ZC3H7B
POLR3H
SNU13
SREBF2
NAGA
XPNPEP3
EP300−AS1
PHF5A
DESI1
SHISA8
DNAJB7
L3MBTL2
LOC105373044
MIR378I
MIR6889
PMM1
TNFRSF13C
C22orf46
CENPM
LINC00634
WBP2NL
41.2
41.4
41.6
41.8
42
42.2
42.4
Position on chr22 (Mb)

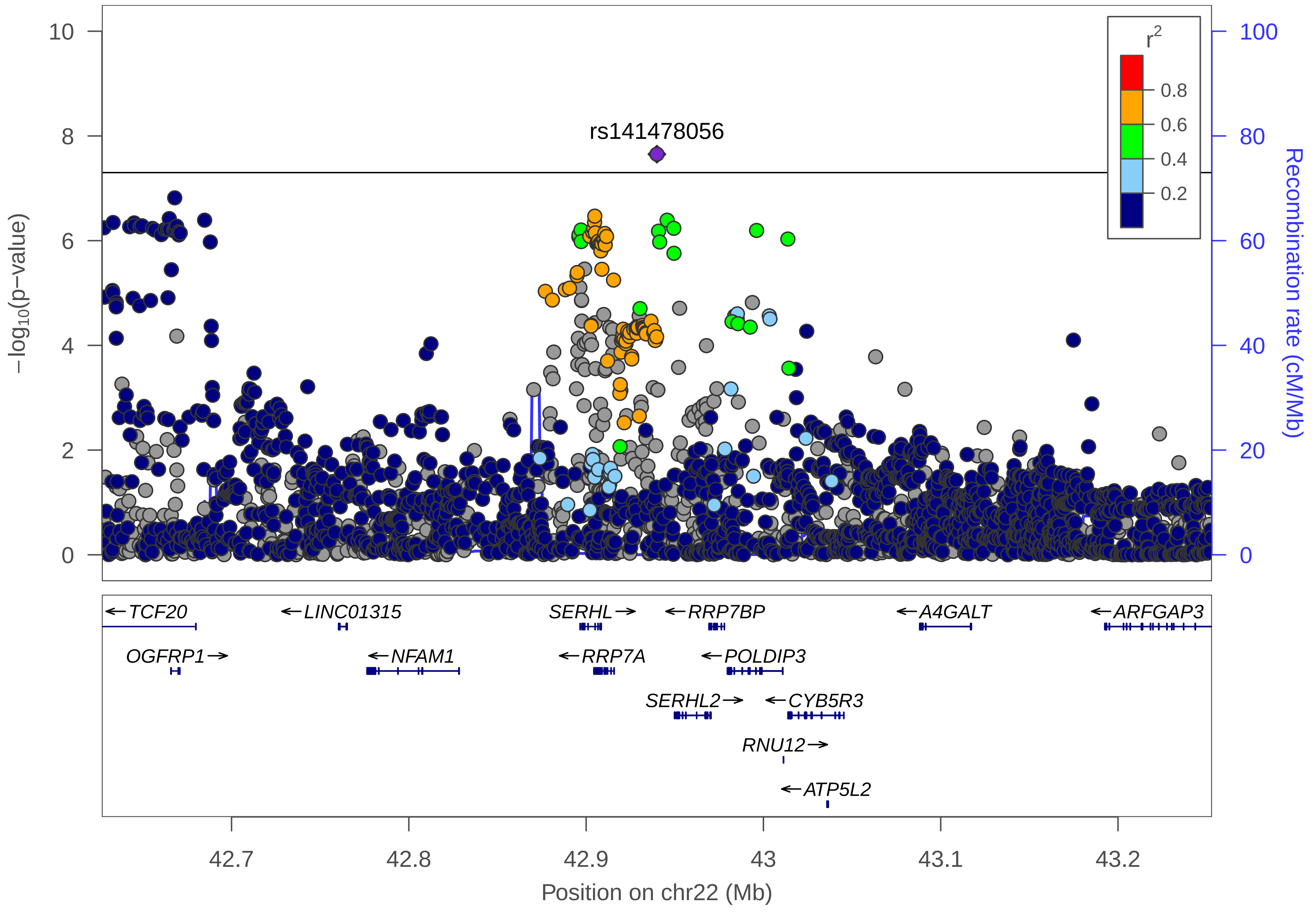

rs141478056
r2
0.8
0.6
0.4
0.2
−log10(p−value)
Recombination rate (cM/Mb)
10
8
6
4
2
0
100
80
60
40
20
0
←TCF20
←LINC01315
SERHL→
←RRP7BP
←A4GALT
←ARFGAP3
OGFRP1→
←NFAM1
←RRP7A
←POLDIP3
SERHL2→
←CYB5R3
RNU12→
←ATP5L2
42.7
42.8
42.9
43
43.1
43.2
Position on chr22 (Mb)

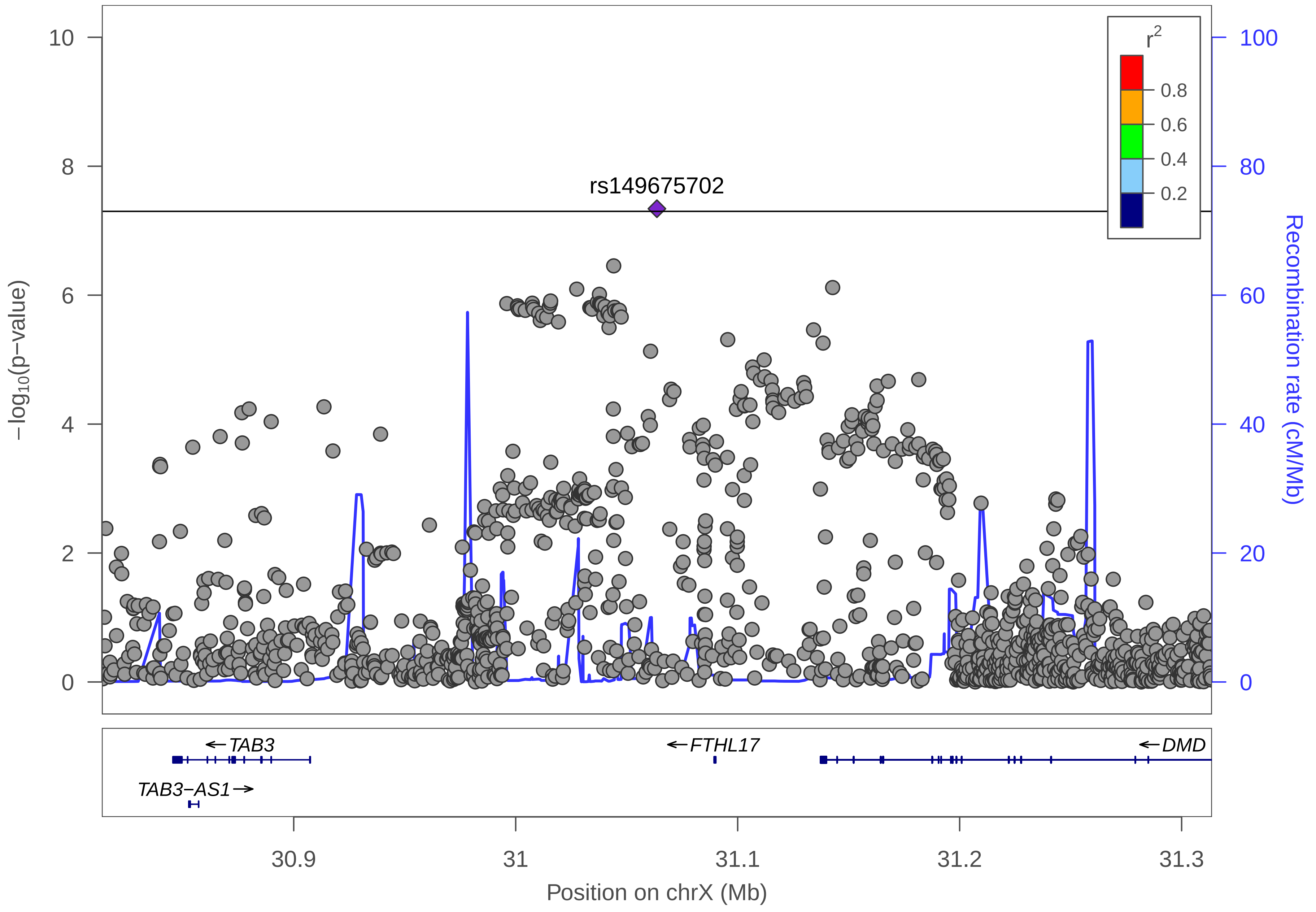

rs149675702
r$^2$
0.8
0.6
0.4
0.2
−log10(p−value)
Recombination rate (cM/Mb)
10
8
6
4
2
0
100
80
60
40
20
0
TAB3
TAB3−AS1
FTHL17
DMD
30.9
31
31.1
31.2
31.3
Position on chrX (Mb)
