## Supplementary Figures 1,2, and 5, Supplementary Figure Legends 3 and 4 for "Fine-mapping a genome-wide meta-analysis of 98,374 migraine cases identifies 181 sets of candidate causal variants"

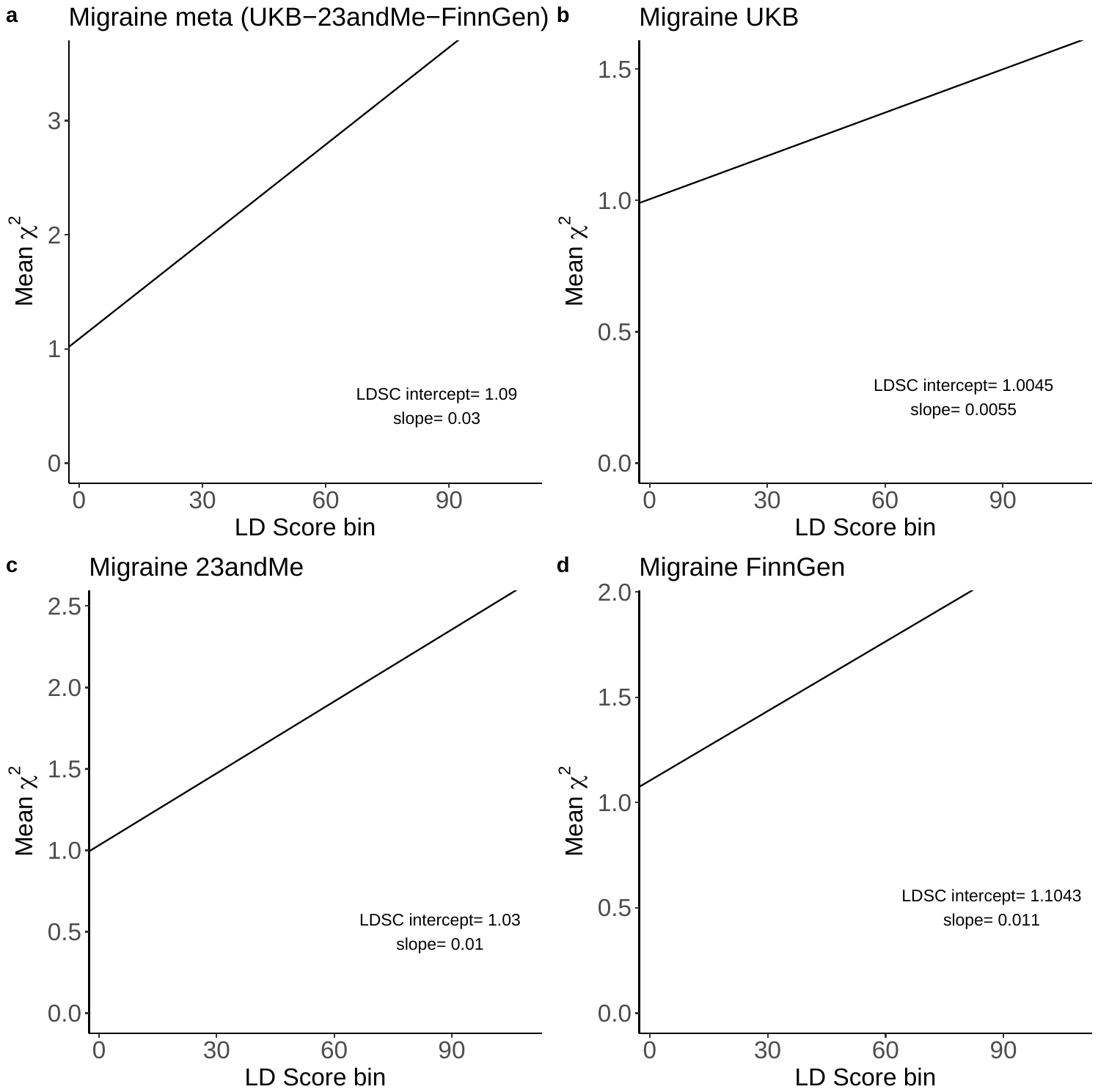

**Supplementary Figure 1.** **LD Score plots of GWAS results from a) the meta-analysis, and separately from b) UK Biobank c) 23andMe , and d) FinnGen.** X-axes show LD Score bins and y-axes corresponding mean $\chi^{2}$-statistics. LDSC intercept measures confounding inflation of the GWAS summary statistics due to cryptic relatedness, population stratification and model misspecification, and values close to 1 indicate only small inflation. Heritability can be estimated by scaling the regression slope.

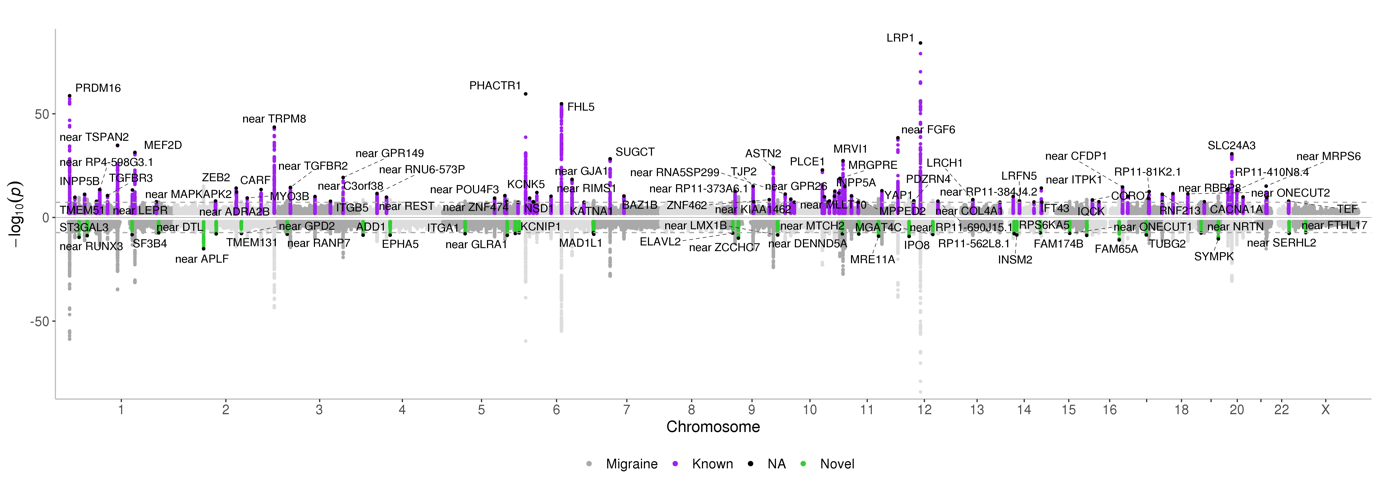

**Supplementary Figure 2. Miami plot of the inverse-variance weighted fixed effects meta-analysis including 98,374 migraine cases and 869,160 controls.** X-axis presents the chromosomal location and y-axis the -log10(p-value). The upper panel shows the previously known loci highlighted in purple, and the lower panel shows the new loci in green.

**Supplementary Figure 3.** **Forest plots of the 122 lead migraine variants from the inverse-variance weighted fixed-effect meta-analysis of the discovery (98,374 cases and 869,160 controls) and replication data (34,807 cases and 193,475 controls).** For each variant, the log-odds-ratio estimate (BETA) with its 95%-confidence intervals (green) are shown from each of the three studies included in the discovery meta-analysis and from the two studies included in the replication meta-analysis, and the combined estimates of the inverse-variance weighted fixed-effect meta-analyses (blue diamonds). The sample sizes of each study are presented as grey squares. Additionally, the lead variant and effect allele, two-sided *P*-value by the inverse-variance weighted fixed-effect meta-analysis and heterogeneity index ($I^{2}$) are displayed.

**Supplementary Figure 4**. **LocusZoom-plots of the 122 LD-independent migraine risk loci identified from the meta-analysis (N = 967,534; 98,374 cases and 869,160 controls).** X-axis shows the chromosomal location, and Y-axis shows the strength of the association as two-sided -log10 *P*-value from the inverse-variance weighted fixed-effects meta-analysis. Black horizontal line corresponds to *P* = 5 × 10^−8^ and blue line shows the recombination rate. The squared correlation to the lead variant is shown by colors based on the combined UK Biobank and FinnGen data.

**Supplementary Figure 5. Comparison of two reference LD panels in fine-mapping of 26 migraine risk loci with in-sample LD available.** Y-axis shows the max$\Delta$ which is a maximum difference of a variant-specific posterior inclusion probabilities between the reference LD and in-sample LD from fine-mapping. X-axis shows a) posterior expectation of the number of causal variants (PENC) from FINEMAP, or, from the top variant(s) of the credible set(s), b) the maximum pairwise squared correlation, c) the maximum marginal P-value from the inverse-variance weighted fixed-effect meta-analysis, and d) the minimum INFO-value. The results using UKB LD reference panel are shown by blue circles, and using combined UKB-FG reference panel by orange crosses.
